# Global SARS-CoV-2 seroprevalence: a systematic review and meta-analysis of standardized population-based studies from Jan 2020-May 2022

**DOI:** 10.1101/2021.12.14.21267791

**Authors:** Isabel Bergeri, Mairead Whelan, Harriet Ware, Lorenzo Subissi, Anthony Nardone, Hannah C Lewis, Zihan Li, Xiaomeng Ma, Marta Valenciano, Brianna Cheng, Lubna Al Ariqi, Arash Rashidian, Joseph Okeibunor, Tasnim Azim, Pushpa Wijesinghe, Linh-Vi Le, Aisling Vaughan, Richard Pebody, Andrea Vicari, Tingting Yan, Mercedes Yanes-Lane, Christian Cao, David A. Clifton, Matthew P Cheng, Jesse Papenburg, David Buckeridge, Niklas Bobrovitz, Rahul K Arora, Maria D Van Kerkhove, the Unity Studies Collaborator Group

**Affiliations:** World Health Organization, Geneva, Switzerland; Centre for Health Informatics, Cumming School of Medicine, University of Calgary, Canada; World Health Organization, Regional Office for Africa, Brazzaville, Congo; Faculty of Engineering, University of Waterloo, Canada; Institute of Health Policy Management and Evaluation, University of Toronto, Toronto, ON, Canada; Epiconcept, Paris, France; School of Population and Global Health, McGill University, Montreal, Quebec, Canada; World Health Organization, Regional Office for the Eastern Mediterranean, Cairo, Egypt; World Health Organization, Regional Office for South-East Asia, New Delhi, India; World Health Organization, Regional Office for the Western Pacific, Manila, Philippines; World Health Organization Regional Office for Europe, Copenhagen, Denmark; World Health Organization, Regional Office for the Americas (Pan American Health Organization), Washington DC, United States of America; Temerty Faculty of Medicine, University of Toronto, Toronto, Canada; COVID-19 Immunity Task Force Secretariat, McGill University, Montreal, Canada; Institute of Biomedical Engineering, University of Oxford, UK; Division of Infectious Diseases and Medical Microbiology, McGill University Health Centre, Montreal, Quebec, Canada; Department of Epidemiology, Biostatistics and Occupational Health, McGill University, Montreal, Canada; Department of Critical Care Medicine, University of Calgary, Canada

## Abstract

**Background:** Our understanding of the global scale of SARS-CoV-2 infection remains incomplete: routine surveillance data underestimates infection and cannot infer on population immunity, there is a predominance of asymptomatic infections, and uneven access to diagnostics. We meta-analyzed SARS-CoV-2 seroprevalence studies, standardized to those described in WHO’s Unity protocol for general population seroepidemiological studies, two years into the pandemic, to estimate the extent of population infection and remaining susceptibility.

**Methods and Findings:** We conducted a systematic review and meta-analysis, searching MEDLINE, Embase, Web of Science, preprints, and grey literature for SARS-CoV-2 seroprevalence published between 2020-01-01 and 2022-05-20. The review protocol is registered with PROSPERO, (CRD42020183634). We included general population cross-sectional and cohort studies meeting an assay quality threshold (90% sensitivity, 97% specificity; exceptions for humanitarian settings). We excluded studies with an unclear or closed population sample frame. Eligible studies - those aligned with the WHO Unity protocol - were extracted and critically appraised in duplicate, with Risk of Bias evaluated using a modified Joanna Briggs Institute checklist. We meta-analyzed seroprevalence by country and month, pooling to estimate regional and global seroprevalence over time; compared seroprevalence from infection to confirmed cases to estimate under-ascertainment; meta-analyzed differences in seroprevalence between demographic subgroups such as age and sex; and identified national factors associated with seroprevalence using meta-regression. The main limitations of our methodology include that some estimates were driven by certain countries or populations being over-represented. We identified 513 full texts reporting 965 distinct seroprevalence studies (41% LMIC) sampling 5,346,069 participants between January 2020 and April 2022, including 459 low/moderate risk of bias studies with national/sub-national scope in further analysis. By September 2021, global SARS-CoV-2 seroprevalence from infection or vaccination was 59.2%, 95% CI [56.1-62.2%]. Overall seroprevalence rose steeply in 2021 due to infection in some regions (e.g., 26.6% [24.6-28.8] to 86.7% [84.6-88.5%] in Africa in December 2021) and vaccination and infection in others (e.g., 9.6% [8.3-11.0%] to 95.9% [92.6-97.8%] in Europe high-income countries in December 2021). After the emergence of Omicron, infection-induced seroprevalence rose to 47.9% [41.0-54.9%] in EUR HIC and 33.7% [31.6-36.0%] in AMR HIC in March 2022. In 2021 Quarter Three (July to September), median seroprevalence to cumulative incidence ratios ranged from around 2:1 in the Americas and Europe HICs to over 100:1 in Africa (LMICs). Children 0-9 years and adults 60+ were at lower risk of seropositivity than adults 20-29 (*p*<0.0001 and *p*=0.005, respectively). In a multivariable model using pre-vaccination data, stringent public health and social measures were associated with lower seroprevalence (*p*=0.02).

**Conclusions:** In this study, we observed that global seroprevalence has risen considerably over time and with regional variation, however around 40 % of the global population remains susceptible to SARS-CoV-2 infection. Our estimates of infections based on seroprevalence far exceed reported COVID-19 cases. Quality and standardized seroprevalence studies are essential to inform COVID-19 response, particularly in resource-limited regions.

## Introduction

The COVID-19 pandemic, caused by the SARS-CoV-2 virus, continues to severely impact population health and health care systems. The 394 million cases and 5.7 million deaths reported as of 7 February 2022 [1]. underestimate the global burden of this pandemic, particularly in low- and middle-income countries (LMICs) with limited capacity for contact tracing, diagnostic testing, and surveillance [2].

Seroprevalence studies estimate the prevalence of SARS-CoV-2 antibodies. These studies are crucial to understand the true extent of infection overall, by demographic group, and by geographic area, as well as to estimate case under-ascertainment. As anti-SARS-CoV-2 antibodies are highly predictive of immune protection [3,4], seroprevalence studies are also indicative of population levels of humoral immunity, and therefore important to inform scenario modeling, public health planning, and national policies in response to the pandemic. Although seroprevalence provides crucial information on population-level infection dynamics, it is important to note that it does not imply protection against infection and therefore is not an appropriate measure to gauge progress towards herd immunity.

During 2021, many regions have experienced third and fourth waves of SARS-CoV-2 infection [1]; concurrently, some countries have vaccinated most residents, while others remain unable to achieve high vaccine coverage due to challenges with supply and uptake [5]. A new wave of well-conducted seroprevalence studies, including many in LMICs, provides robust estimates of seroprevalence in late 2020 and into 2021 [6–8]. Synthesizing these studies is crucial to understand the shifting global dynamics and true extent of SARS-CoV-2 infection, humoral immunity, and population susceptibility. While previous global systematic reviews of seroprevalence have been conducted [9–12], these have included only studies that sampled participants in 2020 and pooled seroprevalence across all time points. These meta-analyses also highlight the importance of improved standardization and study quality to enable more robust analysis [9–11].

Estimates of seroprevalence can be difficult to compare systematically across different settings due to variations in design aspects including sampled populations, testing and analytical methods, timing in relation to waves of infection, and study quality and reporting. The World Health Organization’s Unity Initiative (henceforth “WHO Unity”) aims to help produce harmonized and representative seroprevalence study results in accordance with global equity principles [2]. The WHO Unity population-based, age-stratified seroepidemiological investigation protocol (the SEROPREV protocol) [2] provides a standard study design and laboratory approach to general population seroprevalence studies. WHO Unity and its partners have supported the implementation of SEROPREV by providing financial and technical resources, including a well-performing serologic assay. SEROPREV has been implemented in 74 countries globally and in 51 LMICs as of September 2021 [2]. Synthesizing results aligned with the standard SEROPREV protocol improves study comparability, enabling further analysis of these comparable studies to answer key questions about the progress of the pandemic globally.

This systematic review and meta-analysis synthesized seroprevalence studies worldwide aligned with the SEROPREV protocol, regardless of whether the study received support from WHO. Our objectives were to: (i) estimate changes in global and regional seroprevalence over time by WHO region and country income level; (ii) assess the level of undetected infection, by global and regional case ascertainment over time by calculating the ratio of seroprevalence to cumulative incidence of confirmed cases; and (iii) identify factors associated with seropositivity including demographic differences by 10-year age band and sex through meta-analysis, and study design and country-level differences through meta-regression.

## Methods

### Search strategy and study selection

We conducted a systematic review of seroprevalence studies (hereafter “studies”) published from 1 January 2020 to 20 May 2022. We designed a primary search strategy in consultation with a health sciences librarian in MEDLINE, Embase, Web of Science, and Europe PMC using key terms such as SARS-COV-2, COVID-19, seroprevalence, and serology. We attempted to mitigate possible publication bias by including both published articles and unpublished literature such as grey literature, preprints, institutional reports, and media reports (full strategy in Supplementary file S.3.1). For our secondary search and article capture strategy, we invited submissions to our database through the open-access SeroTracker platform and recommendations from international experts, including literature compiled through the WHO Unity studies initiative. In order to access timely evidence and mitigate challenges with publication delay, we also contacted WHO Unity study collaborators that had not yet made results available to the general public prior to our inclusion dates, to upload their aggregate results to the open access Zenodo research data repository [13]. We accepted these templates up to May 20th 2022 in line with our primary search strategy, and screened them according to the same criteria as other sources captured in our primary search. This systematic review and meta-analysis protocol was registered with PROSPERO (CRD42020183634) prior to the conduct of the review (Supplement S6) [14], reported according to the Preferred Reporting Items Systematic review and Meta-Analyses (PRISMA) guideline [15] (S1 Checklists), and searches and extractions conducted per the previously established SeroTracker protocol [16].

Studies were screened, data extracted, and critically appraised in duplicate, with these tasks shared by a team of 13 study authors (listed in Research Contributions section under data curation). We have study team members proficient in English, French, Portuguese, Spanish, and Cyrillic languages - articles in all other languages were translated using Google Translate where possible. Conflicts were resolved by consensus. Inclusion and exclusion criteria aligned with the SEROPREV standardized protocol for general population seroprevalence to minimize possible bias introduced by inter-study heterogeneity and other measures of study quality such as poor assay performance and/or sampling methods (full protocol criteria described in Supplementary File S2.2 and S2.3). We included cross-sectional or longitudinal cohort studies with the objective of estimating SARS-CoV-2 seroprevalence in the general population. Restricting inclusion to direct population samples such as household surveys would have led to very little data in some regions and times, as these studies are expensive and difficult to conduct. Thus, household and community samples were included, as well as studies where a robust sampling frame was described that approximates to a wider population, such as individuals attending medical services (blood donors, pregnant mothers, primary care attendees) or residual sera taken from patients for a variety of other investigations. Finally, we also included people residing in slum dwellings, and some patient populations in humanitarian settings where the patient population in question was extensive enough to be considered a proxy sample frame (evaluated on a case-by-case basis). Both random and non-random (i.e., convenience, sequential, quota) sampling methods were included. Convenience samples must have a clear and defined sampling frame, i.e., studies recruiting volunteers were not included.

Studies had to use serological assays with at least 90% sensitivity and 97% specificity as reported by the manufacturer or study authors through an independent evaluation of the test used (Supplementary file S2.1), unless conducted in vulnerable countries as defined in the Global Humanitarian Response Plan (HRP) [17] We employed exception criteria for HRP countries by including all assays despite those with sometimes lower performance as sometimes rapid diagnostic tests were the only operationally possible option due to ongoing humanitarian emergencies. Studies employing dried blood spots as a specimen type must meet this threshold as determined through study author-conducted sensitivity and specificity validation using dried blood spots. Multi-assay testing algorithms were included if the combined sensitivity and specificity met these performance thresholds, using standard formulas for parallel and serial testing [18]. Complex multiple testing strategies (3+ tests used) were reviewed on a case-by-case basis by two study members. To accommodate these limitations and ensure study inclusion equity, we included all assay types from HRP countries regardless of their reported performance values, as long as the authors reported an assay that was independently validated from either an in-house evaluation or a WHO-approved head-to-head evaluation [19-21]. Finally, algorithms employing a commercial or author-designed binding assay followed by confirmatory testing by virus neutralization assay were included as they constitute the gold standard in serological evaluation [22].

We excluded studies sampling specific closed populations (such as prisons, care homes, or other single-institution populations), recruiting participants without a clear sampling frame approximating the target population or testing strategy, and studies that excluded people previously diagnosed with or vaccinated against COVID-19 after initial sampling.

### Data extraction, synthesis, and analysis

From each study, we extracted seroprevalence estimates for the overall sample, and stratified by age, sex, vaccination status, and timing of specimen collection according to the prespecified protocol. We extracted information on study population, laboratory assay used, any corrections made in estimating seroprevalence (e.g., for population or assay performance), seroprevalence, and denominator. Standardized results uploaded to Zenodo by Unity study collaborators additionally included information on the proportion of asymptomatic seropositive individuals.

Our procedure for the standardized, aggregate early data results submitted by Unity collaborators was to direct study authors to input their results into a formulated standard Excel template designed to match the same data extracted during routine published study extraction. A blank version of the Excel template is available for reference [23]. These templates were uploaded directly into R for analysis in tandem with other studies included in the meta-analysis. Templates were verified by two independent reviewers and we conducted follow-up to complete information with study investigators where needed. In instances where data from early reporting templates we had received were published prior to May 20th 2022 or a partial dataset was previously published, we ensured to de-duplicate this data for analysis. We evaluated cases of duplicated results on a case-by-case basis, prioritizing the authors published version by default but made exceptions where data was more complete, robust, or up-to-date in the submitted templates. Once authors published or preprinted their results, a link to the full source was added to the Zenodo repository.

We critically appraised all studies using a modified version of the Joanna Briggs Institute (JBI) checklist for prevalence studies [24]. To assess risk of bias, a decision rule assigned a rating of low, moderate, or high risk of bias based on the specific combination of JBI checklist ratings for that study [25]. This decision rule was developed based on guidance on estimating disease prevalence [26,27] and was validated against overall risk of bias assessments derived manually by two independent reviewers for previously collected seroprevalence studies in the SeroTracker database, showing good agreement with manual review (intraclass correlation 0.77, 95% CI 0.74-0.80; n = 2070 studies) [25]. Early results from templates were screened and evaluated for risk of bias using the same criteria as studies captured through routine screening processes.

We classified seroprevalence studies by geographical scope (local [i.e. cities, counties], sub-national [i.e. provinces or states], or national), sample frame, sampling method, and type of serological assay (Supplement S2.1 Table S1, Table 5). Where an article or source material contains multiple, methodologically distinct serosurveys we split the article into multiple ‘studies’ - for the purpose of this review ‘study’ means a distinct estimate. Where multiple summary estimates were available per study, we prioritized estimates based on estimate adjustment, antibody isotypes measured, test type used, and antibody targets measured (full details: Supplement S3.1). We included multiple estimates per study when broken down by time frame in our analysis over time.

Countries were classified according to WHO region [28], vulnerability via HRP status [17], and World Bank income level [29].

We anchored each estimate to the date halfway between sampling start and end (“sampling midpoint date”) to best reflect the time period of the study. To select the most representative and high quality studies for analysis, we used only sub-national or national studies rated low or moderate risk of bias to estimate seroprevalence in the general population over time and identify factors associated with seroprevalence (sub-dataset 1). We used only national studies rated low or moderate risk of bias to estimate case ascertainment (sub-dataset 2).

To explore possible causes of heterogeneity among study results, we constructed a Poisson generalized linear mixed-effects model with log link function using the glmer function from the lme4 package in R [30–32]. Independent predictors were defined *a priori* as WHO region, income group, geographic scope, sample frame, pandemic timing, age, cumulative confirmed cases, and average public health and social measure (PHSM) stringency index [33]. To focus on factors associated with seroprevalence from infection, we included studies where less than 5% of the national population was vaccinated two weeks before the sampling midpoint date. We included all *a priori* predictors in the final model, and to evaluate the importance of each relevant predictor, we compared the Akaike Information Criterion (AIC) of the final model to all models dropping a single predictor at a time (full details on the model and predictor definitions: Supplement S3.2).

To estimate seroprevalence in the general population, we first produced monthly country-level estimates by meta-analyzing seroprevalence in each country, grouping studies in a 12-week rolling window considering the infrequent availability of seroprevalence studies in most countries (rma.glmm from R package metafor)[34,35]. We then produced monthly regional estimates by taking weighted averages of country estimates by population, ensuring that country contributions to these estimates are proportional to country population. We stratified these estimates by the expected key sources of heterogeneity among study results: region, income class, and time. We pooled HIC and LMIC together in the Eastern Mediterranean (EMR) and Western Pacific regions (WPR) due to the lower number of studies, and in the Africa (AFR) and South-East Asia regions (SEAR) (the only two HIC in these regions had no studies).

We produced monthly global estimates where estimates were available for a majority of regions, calculating global estimates as a population-weighted average of regional estimates to ensure regional representation (full details: Supplement S3.2). We produced 95% confidence intervals for the mean seroprevalence estimate, reflecting uncertainty in the summary effect size [36], and 95% prediction intervals to give a range for the predicted parameter value in a new study. All numerical results presented are from this stage. To visualize the trend in regional and global estimates over time, we fit a smooth curve to these estimates using non-parametric regression (gam from R package mgcv) [37]. We also summarized the relevant variant genome frequency in each region shared via the GISAID initiative [38].

We also estimated to what extent laboratory confirmed SARS-CoV-2 cases [39] underestimated the full extent of infections based on seroprevalence. For studies that sampled participants in 2021, we used national seroprevalence estimates and vaccination rates [40] to calculate seroprevalence attributable to infection only. In countries administering only vaccines using Spike (S) protein antigens (e.g., mRNA), we calculated the ascertainment ratio using only studies that detected anti-nucleocapsid (N) seroprevalence. In countries administering inactivated vaccines that may generate both anti-S and anti-N responses, we adjusted the reported seroprevalence using a standard formula [41]. We then produced regional and global estimates of seroprevalence using the two-stage process described above, and computed the ratio to the corresponding cumulative incidence of confirmed SARS-CoV-2 cases in the region or globally. We stratified by HIC vs. LMIC in all regions.

Aggregated results shared by Unity collaborators reported the proportion of seropositives that were symptomatic at some time point prior to sampling, summarized using the median and interquartile range, and tested for differences in distribution across age and sex groups using analysis of variance (ANOVA).

To quantify population differences in SARS-CoV-2 seroprevalence, we identified studies with seroprevalence estimates for sex and age subgroups. We calculated the ratio in seroprevalence between groups within each study, comparing each age group to adults 20-29 and males to females. We then aggregated the ratios across studies using inverse variance-weighted random-effects meta-analysis. The amount of variation attributable to between-study heterogeneity vs. within-study variance was quantified using the I^2^ statistic.

Our main analysis used seroprevalence estimates uncorrected for test characteristics. As a sensitivity analysis, we also produced global and regional estimates adjusting for test characteristics through Bayesian measurement error models, with binomial sensitivity and specificity distributions. The sensitivity and specificity values for correction were prioritized from the WHO SARS-CoV-2 Test Kit Comparative Study conducted at the NRL Australia [19], followed by a multicentre evaluation of 47 commercial SARS-CoV-2 immunoassays by 41 Dutch laboratories [42], and from independent evaluations by study authors where author-designed assays were used.

Data was analyzed using R statistical software version 4.1.2.[32]

## Results

### Study characteristics

We identified 173,430 titles and abstracts in our search spanning from 2020-01-01 to 2022-20-05 (Figure 1). Of these, 5,281 full text articles were included in full text screening. 513 seroprevalence data sources containing studies aligned with the SEROPREV protocol were identified, 480 published (94%) and 33 aggregated results from collaborators (6%), of which 12 sources were not in one of our main languages and translated via Google. The 513 sources contained a total of 965 unique seroprevalence studies (detailed references and information available in Supplement, S4.1-S4.3, S5). Over 1,500 full-text articles were excluded due to not containing studies compatible with the SEROPREV protocol; the main reasons for articles’ exclusion at this stage was having an incorrect sample frame for this analyses’ scope (i.e., we focussed on seroprevalence in the general population and therefore excluded 1,073 articles of exclusively health care workers, close contacts of confirmed cases, or other specific closed populations) or not meeting our pre-defined assay quality performance threshold (374 articles).

**Figure 1.**
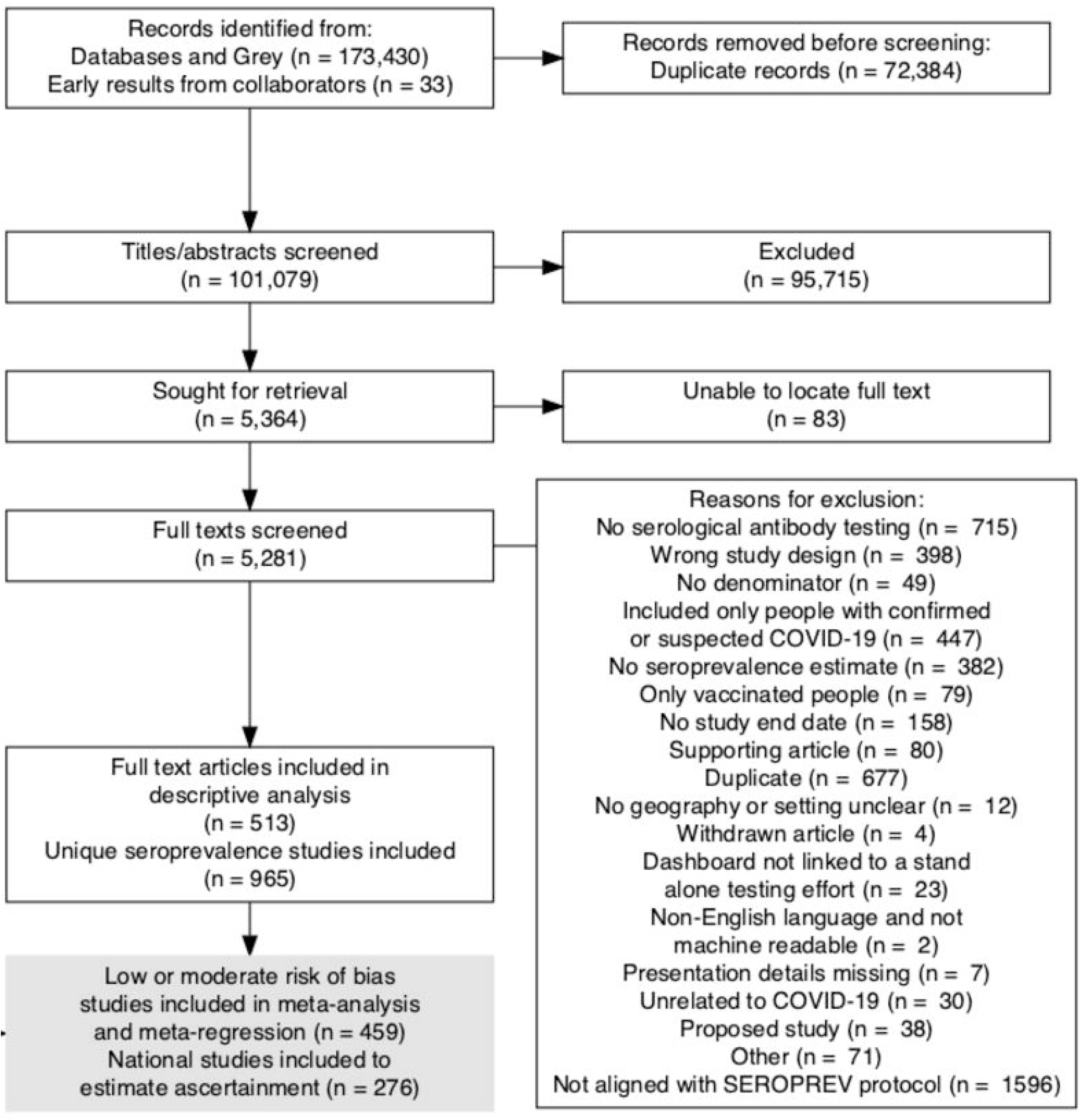
PRISMA Flow Diagram of Inclusion. In cases where sources contained multiple primary estimates of seroprevalence (i.e. in non-overlapping populations, separate methodological seroprevalence studies reported in the same article, etc), the source (full text) was split into multiple individual studies included in the analysis. For this reason, we report more unique seroprevalence studies than original full text articles included.

A total of 52% (100/194) of WHO Member States (MS) and four WHO Countries, areas and territories, across all 6 WHO regions, were represented among the seroprevalence studies included in the descriptive analysis (Figure S1). Twenty three of 47 MS were represented in AFR; 11 of 21 MS and one territory in EMR; 13 of 35 MS and one territory in AMR; 39 of 53 MS and two territories in EUR; 6 of 11 MS in SEAR; and 8 of 27 MS in WPR (Figure S1). Data from 61 of 134 LMICs and from 36 of 63 vulnerable HRP countries were included. A large proportion of studies included in the descriptive analysis were conducted in LMIC (397/965, 41%) and in vulnerable HRP countries (206/965, 21%) (Table 1). Of studies included in the meta-analysis and meta-regression, these proportions were 30% (137/459) and 14% (66/459), respectively. 20 of the 66 (30%) meta-analyzed studies in HRP countries had test performance values below 90% sensitivity or 97% specificity and were included due to exception criteria.

**Table 1.**
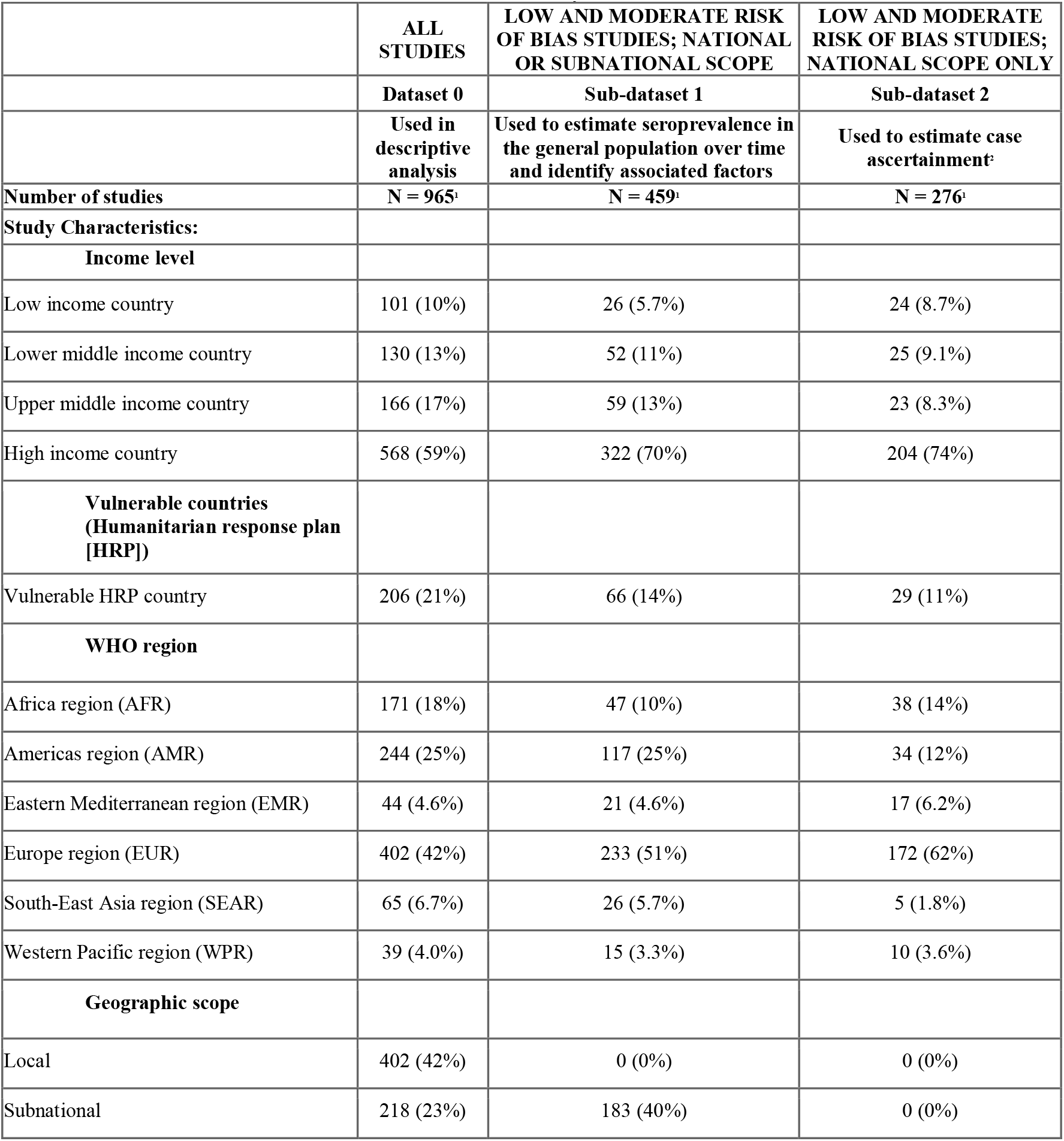
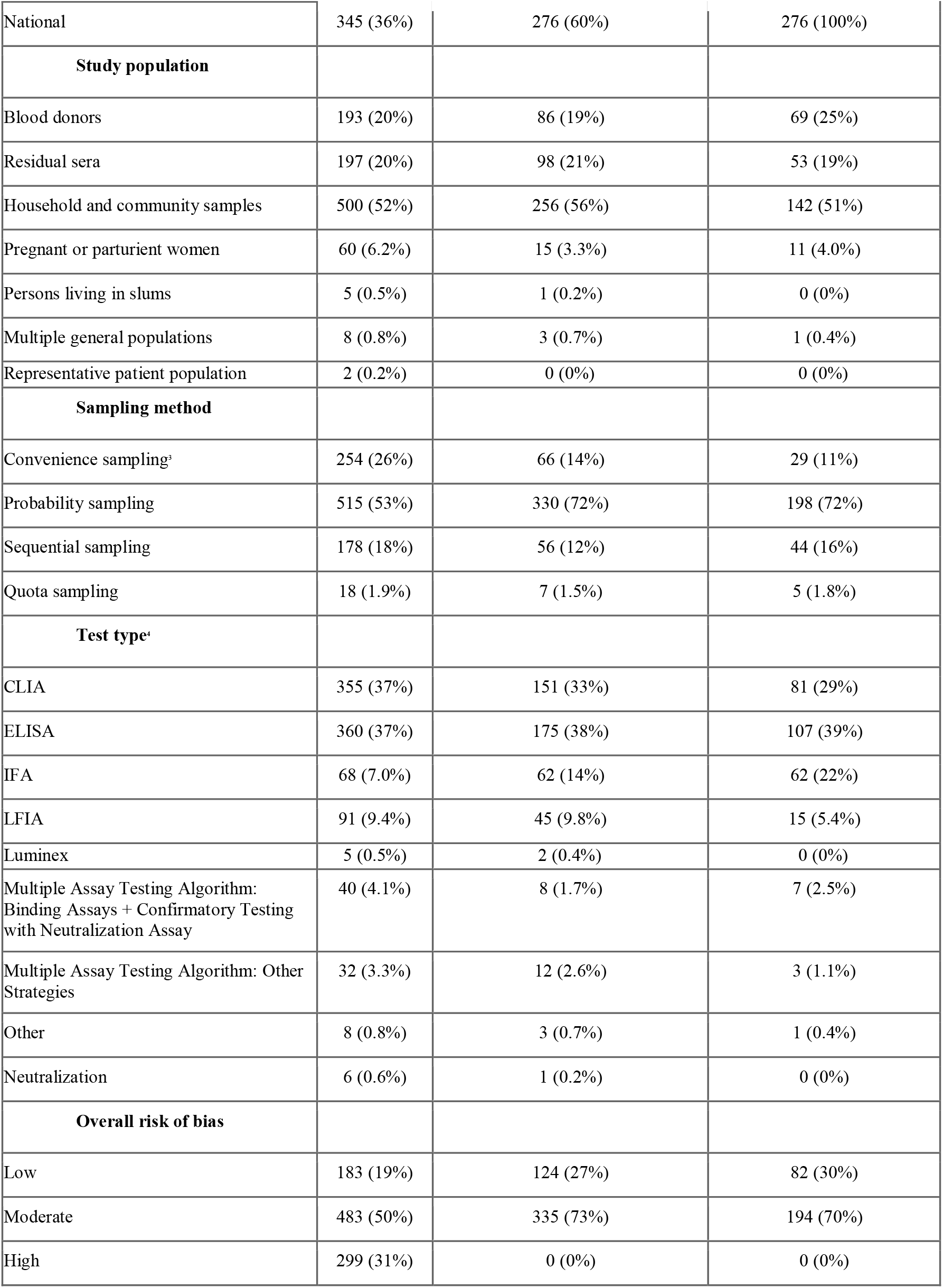
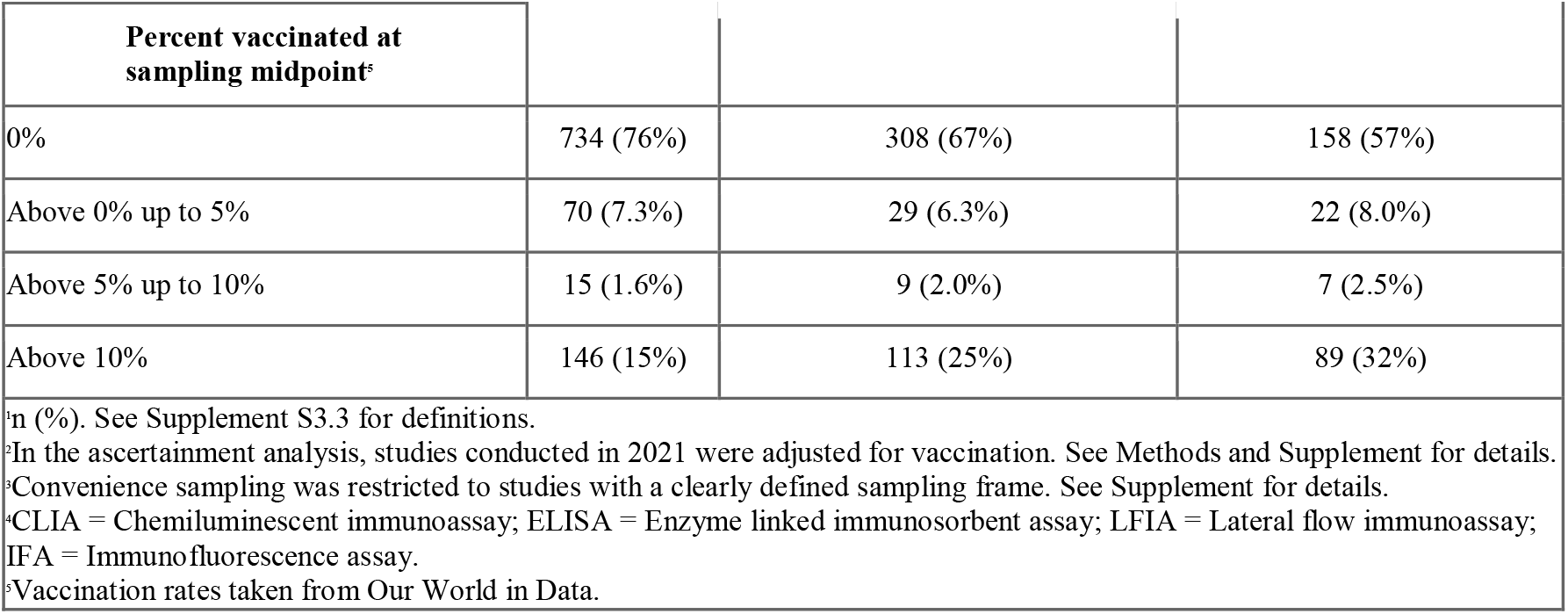
Characteristics of Included Studies, Jan 2021-May 2022.

Among the 965 studies included in the descriptive analysis, 42% (402/965) reported results at a local level, 36% (345/965) at a national level, and 23% (218/965) at a sub-national level. The most common sampling frame and method was household and communities (52%, 500/965) and probability sampling (53%, 515/965), respectively. Within household-based samples, only 86/500 studies (17.2%) used convenience sampling. Among the testing strategies used to measure seroprevalence, most studies used ELISA (37%, 360/965) or CLIA assays (37%, 355/965) and few studies used a lateral flow immunoassay (9.4%, 91/965) or multiple assay testing algorithm (7.4%, 72/965). The majority of studies (734/965, 76%) had no vaccination at the sampling midpoint date in the country of the study (Table 1). Very few studies (14/965, 1.5%) in 4 countries (Canada, Japan, United Kingdom, United States of America) sampled participants during 2022.

Most (50%, 483/965) studies were rated moderate risk of bias. A summary of overall risk of bias ratings and breakdown of each risk of bias indicator for all studies is available (Figure S2 and Table S8, respectively). Sub-national and national studies at low or moderate risk were included in the subsequent results.

### Meta-regression

In the multivariable meta-regression, 329 studies remained after applying our inclusion criteria for pre-vaccination studies. The full model is reported here (model comparison and diagnostics: Table S11). Sub-national studies reported higher seroprevalence estimates compared to national studies (PR 1.27 [1.02-1.59], *p*=0.03). Compared to HIC, higher seroprevalence estimates were reported by low income (PR 7.33 [3.49-15.41], *p*<0.0001), lower-middle income (PR 7.33 [3.49-15.41], *p*<0.0001), and upper middle income countries (PR 3.97 [2.88-5.49], *p*<0.0001). Higher cumulative incidence of reported cases was associated with higher seroprevalence (PR 1.39 [1.30-1.49], *p*<0.0001), while more stringent PHSM measures up to the sampling midpoint date, continuous from 0 to 10, were associated with lower seroprevalence (PR 0.89 [0.81-0.98], *p*=0.02). Much of the heterogeneity in effect sizes was explained by WHO region, income class, and cumulative confirmed cases. By contrast, sample frame was the least important predictor based on the AIC criterion (Table S11) and compared to studies that sampled households and communities, there were no differences between seroprevalence in studies that sampled blood donors (PR 1.04 [0.77-1.40], *p*=0.79) nor residual sera (PR 1.08 [0.83-1.41], *p=*0.55).

### Overall and infection-induced seroprevalence by month, region, and income class

We estimated weighted seroprevalence in a series of separate meta-analyses each month and found in September 2021, global seroprevalence from infection or vaccination (overall seroprevalence) was 59.2%, [95% CI 56.1-62.2%, 95% prediction interval 51.2-66.7%] - an 6.6 fold increase since the June 2020 estimate of 7.7% [CI 5.7-10.3%, prediction interval 4.2-13.8] (Table S9). In September 2021, global seroprevalence attributable to infection was 35.9% [CI 29.5-42.7%, prediction interval 22.8-51.4%] (Figure 3, Table S9).

**Figure 2.**
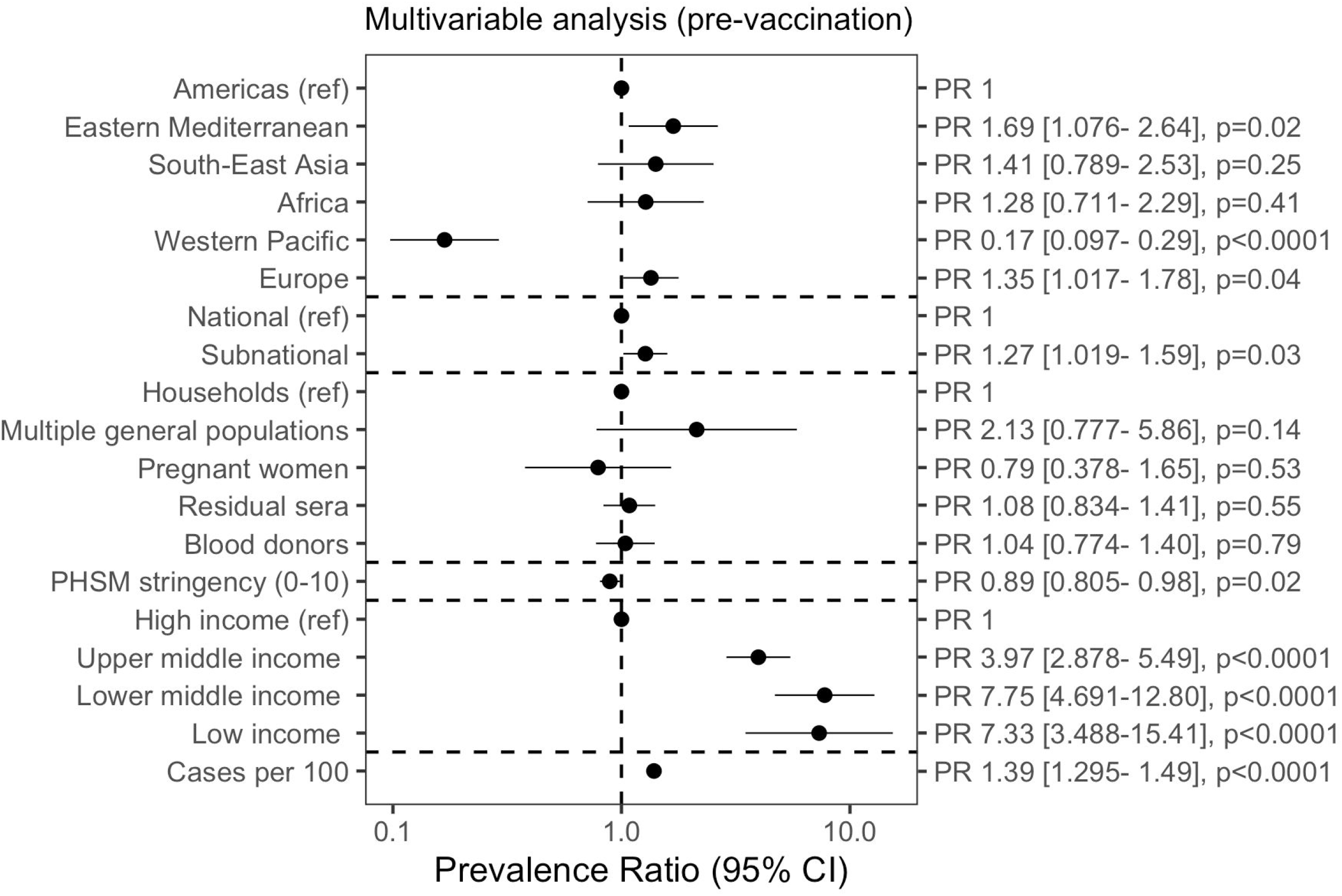
Meta-regression of seroprevalence (pre-vaccination) to identify study design and country factors associated with seroprevalence. We fit a log-Poisson generalized linear mixed-effects model, including studies where less than 5% of the national population was vaccinated two weeks before the sampling midpoint date. We performed model comparison using the AIC criterion (Table S11). Public health and social measures (PHSM) data was taken from the London School of Hygiene and Tropical Medicine global dataset. The PHSM index scale ranged from 0 (least stringent) to 10 (most stringent) (see Supplement S3.2). k = 329; □^2^(95% CI) = 0.74 (0.63-0.87). The marginal R2, or variation between studies explained only by fixed effects, was 62.9%. Multivariable analysis included additional controls for transmission phase and age group not shown in figure.

**Figure 3.**
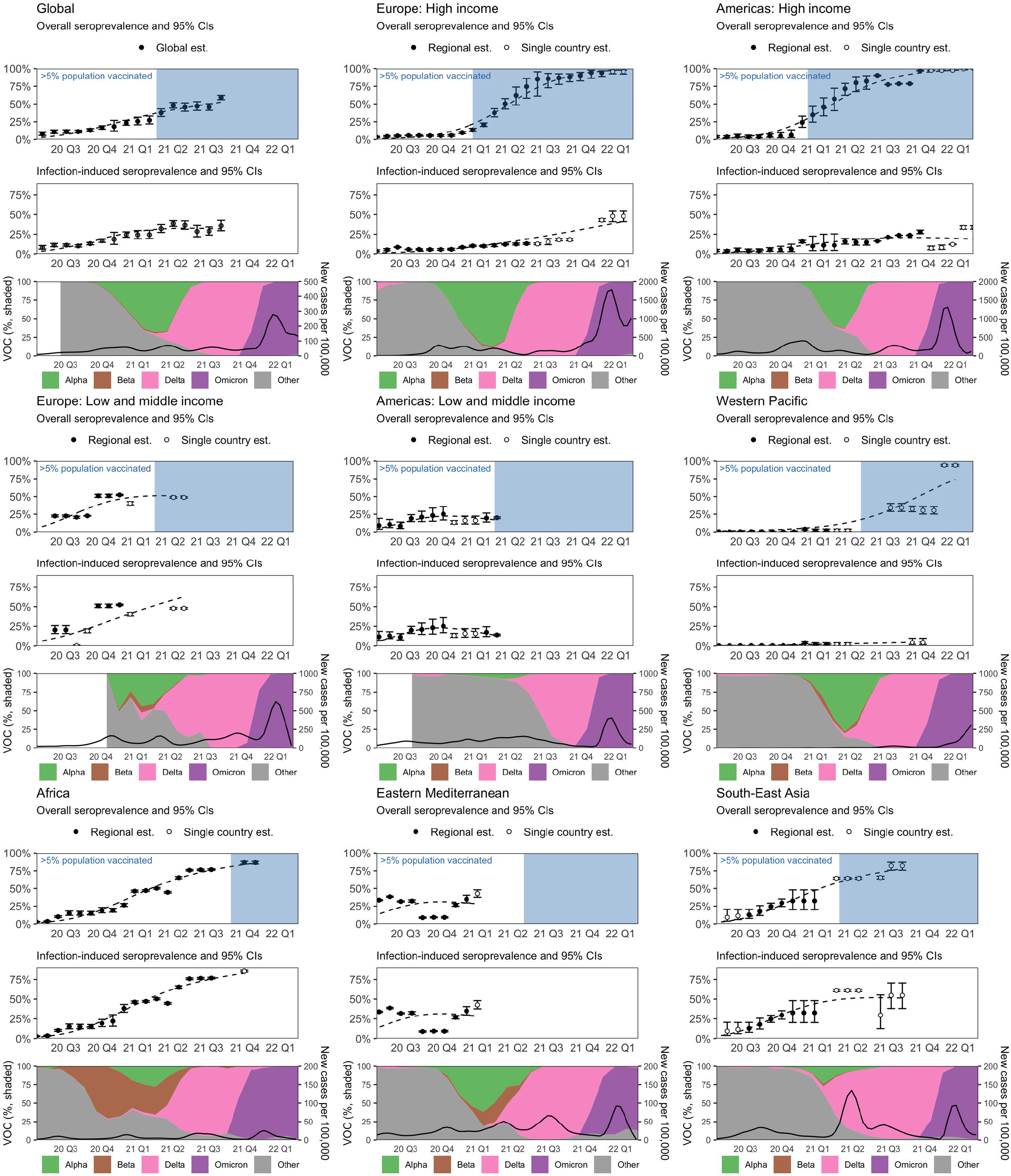
Estimated seroprevalence globally and by WHO region from January 2020 to March 2022. Est: Estimate; CI: Confidence Interval; VOC: Variant of Concern. **Top and middle panel:** We produced weighted point estimates and 95% CIs of overall (top) and infection-induced (middle) seroprevalence by meta-analyzing studies in 12-week rolling windows. To visualize the trend in seroprevalence in each WHO region and globally, we fit a flexible, smooth function of time (dashed line) to the point estimates using non-parametric regression (full details: Supplement S3.2). Countries included in each region-month estimate are in Table S9. **Bottom panel, left axis:** Shaded areas represent the relative frequency of major VOCs circulating, based on weekly counts of hCoV-19 genomes submitted to GISAID we have aggregated by month. Weeks with fewer than 10 total submissions in a given country were excluded from the analysis. **Bottom panel, right axis:** New confirmed cases per 100,000 people, smoothed using local regression (LOESS).

Regional analyses began in January 2020 and ended in February 2021-March 2022 depending on when seroprevalence studies in each region sampled participants. Overall seroprevalence in February 2021 was 42.7% [37.6-48.0%] in EMR (1.3x since June 2020). In April 2021, overall seroprevalence was 20.0% [18.8-21.2%] in AMR LMIC (2.3x since June 2020). In June 2021, overall seroprevalence was 48.7% [47.7-49.7%] in EUR LMIC, a 2.2x increase since July 2020. In September 2021, overall seroprevalence was 82.2% [75.9-87.2%] in SEAR (8.9x since June 2020). In December 2021, overall seroprevalence was 86.7% [84.6-88.5%] in AFR (25x since June 2020) and 30.3% [25.3-35.9%] in WPR (135x since June 2020). Finally, in March 2022, overall seroprevalence was 95.9% [92.3-97.8%] in EUR HIC (22x since June 2020), and 99.8% [99.7-99.9%] in AMR (HIC) (27x since June 2020). (Figure 3, middle panel and Table S9). Infection-induced seroprevalence is reported in Table S9; for example, 47.9% [41.0-54.9%] of the population in EUR HIC (United Kingdom studies only) and 33.7% [31.6-36.0%] of the population in AMR HIC (Canada studies only) had infection-induced antibodies in March 2022. In the meta-analyses by country with at least 2 studies, 75% (188/250) showed considerable heterogeneity from 75% to 100%.(36)

### Ratios of seroprevalence to cumulative incidence

Snapshots of seroprevalence to confirmed case ratios, based on estimated weighted seroprevalence using national studies, are shown in Table 2. Globally, the median ratio was 51.3:1 in 2020 Quarter Three and 10.5:1 in 2021 Quarter Three. In 2020 Quarter Three, the median ratio ranged from 3.4:1 in AMR (HIC) to 219.6:1 in EMR. In 2021 Quarter Three, this ranged from 1.8:1 in AMR (HIC) to 176.7:1 in AFR (Table 2).

**Table 2:** Median estimated seroprevalence to cumulative incidence ratios by WHO region, World Bank income level, and quarter using national studies. NA = national studies not available. Seroprevalence studies that sampled participants in 2021 were adjusted for antibody target and vaccination rate to calculate seroprevalence attributable to infection (full details: Methods and Supplement S3.2). **\***There are no high income countries in the WHO South-East Asia region; the two high-income countries in the WHO Africa region, Mauritius and Seychelles, both have no seroprevalence studies and were hence not included in this analysis.

|  |  | Estimated seroprevalence to case ratios: Median [Range] |  |  |  |  |
| --- | --- | --- | --- | --- | --- | --- |
| WHO region | Income level* | 2020 Q3 (July to September) | 2020 Q4 (October to December) | 2021 Q1 (January to March) | 2021 Q2 (April to June) | 2021 Q3 (July to September) |
| Africa (AFR) | Low-middle income (LMIC) | 82.2 [43-104.9] | 152.9 [115.5-156.9] | 154.4 [145.9-211.2] | 185.5 [131.2-226.1] | 176.7 [164.1-189.2] |
| Americas (AMR) | Low-middle income | 22 [6.1-27.5] | 18.3 [18.3-18.3] | NA | NA | NA |
| Americas | High income (HIC) | 4.6 [2.5-6.5] | 2.6 [2-2.9] | 2.3 [2.2-2.4] | 2 [1.9-2.2] | 1.8 [1.8-1.8] |
| Eastern Mediterranean (EMR) | Low-middle income | 219.6 [36.9-425.2] | 28.8 [20.8-56.7] | 59.3 [56.8-61.8] | NA | NA |
| Eastern Mediterranean | High income | 8.8 [8.5-27.2] | 26.2 [25.3-27] | NA | NA | NA |
| Europe (EUR) | Low-middle income | 84.5 [71.8-97.2] | 44.4 [35.5-53.3] | 25.8 [25.8-25.8] | 11.5 [11.3-11.8] | NA |
| Europe | High income | 8.1 [5.5-15.4] | 1.9 [1.5-4.3] | 2.1 [1.8-2.1] | 1.6 [1.6-1.8] | 1.9 [1.7-2.1] |
| South-East Asia (SEAR) | Low-middle income | 30.1 [22.4-37.8] | 40.3 [33.5-48.3] | 37.5 [37.5-37.5] | NA | NA |
| Western Pacific (WPR) | Low-middle income | 45.4 [29.4-338.9] | 44.3 [38.7-51.6] | 34.5 [31.2-37.9] | NA | NA |
| Western Pacific | High income | 3.4 [2.9-4] | 3.1 [3-3.2] | NA | NA | NA |
| **Global** | **All** | **51.3 [39.6-62.9]** | **22.7 [20.9-24.5]** | **17.9 [15-20.7]** | **13.5 [13.5-13.5]** | **10.5 [9.3-11.7]** |

### Subgroup analysis

Asymptomatic seroprevalence by age and sex subgroups for studies reporting subgroups on symptoms are shown in Supplementary Figure S3. Median asymptomatic prevalence was similar across age groups (ANOVA *p* = 0.28). Median asymptomatic prevalence in males was 64.6% compared to 58.6% in females (ANOVA *p* = 0.47).

Within studies, compared to the reference category of 20-29 years old, seroprevalence was significantly lower for children 0-9 years (prevalence ratio 0.75, 95% CI [0.67-0.84], *p*<0.0001) and adults 60+ years (0.87 [0.80-0.96], *p*=0.005). There were no differences between other age groups nor between males and females. (Full results: Figure 4)

**Figure 4.**
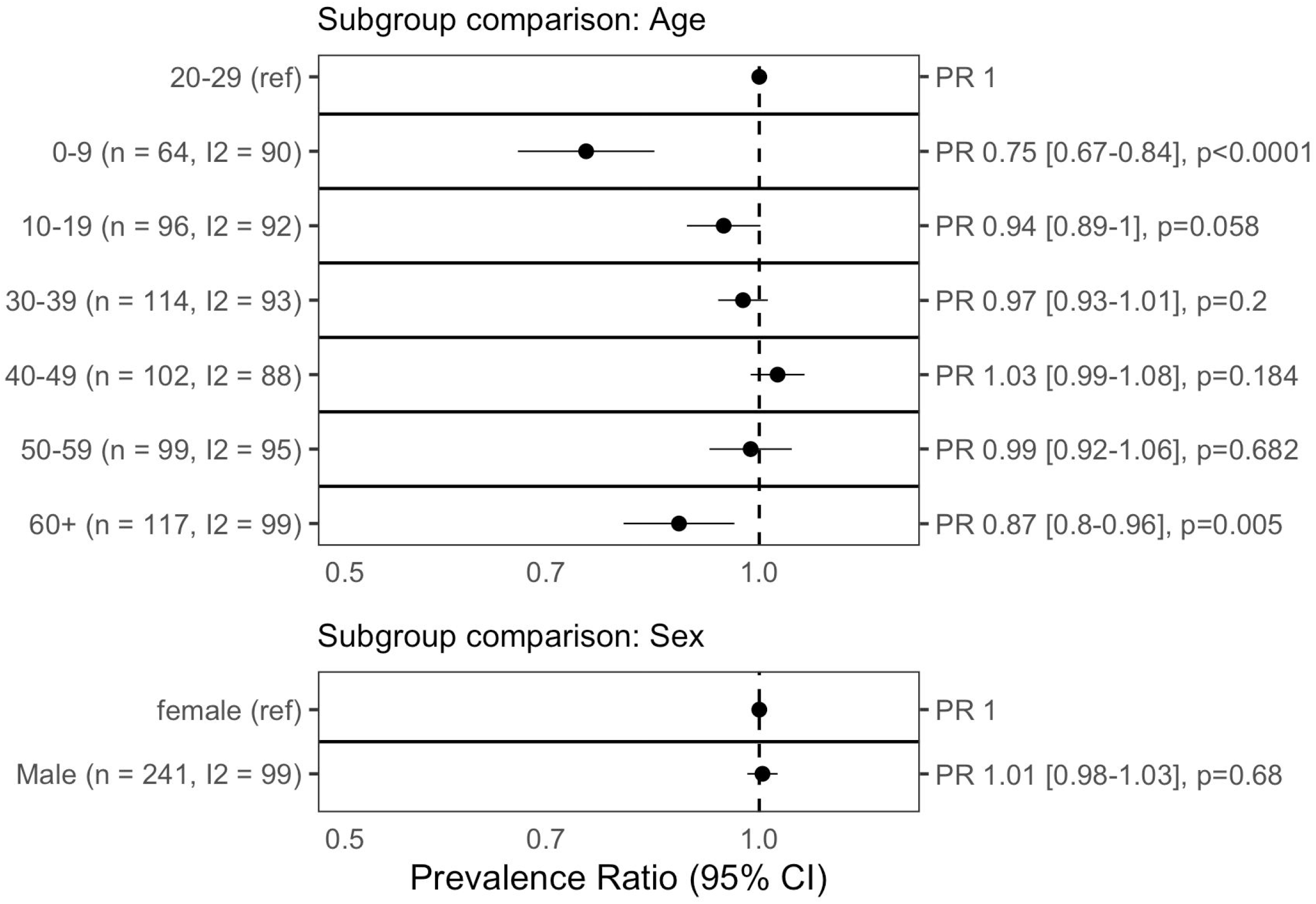
Meta-analysis of seroprevalence differences by demographic groups. We calculated the ratio in prevalence between subgroups within each study then aggregated the ratios across studies using inverse variance-weighted random-effects meta-analysis. Heterogeneity was quantified using the I2 statistic. Each row represents a separate meta-analysis.

### Sensitivity analysis

Regional and global estimates of seroprevalence accounting for serological test performance from independent test kit evaluations showed no qualitative differences from the primary results (Table S10). For example, overall global seroprevalence in September 2021 using corrected estimates was 61.5% [56.7-66.1%], compared to 59.2% [56.1-62.2%] using uncorrected estimates.

## Discussion

### Summary

We synthesized data from over 900 seroprevalence studies worldwide (43% from LMICs) published up to December 2021 (search dates: 2020-01-01 to 2022-20-05), providing global and regional estimates of SARS-CoV-2 seroprevalence over time with substantial representation of regions with limited available seroprevalence data. We estimate that approximately 59.2% of the global population had antibodies against SARS-CoV-2 in September 2021 with 35.9% of the global population attributable to infection (when excluding vaccination). Global seroprevalence has risen considerably over time, from 7.7% more than a year before, in June 2020.

### Results in context

Our findings provide evidence of regional and temporal variation in the overall seroprevalence, over 80% in SEAR and AFR in late 2021 and over 90% in AMR HIC and EUR HIC in early 2022. In WPR, there was a paucity of high-quality population-based studies in 2021 and estimated seroprevalence was as low as 30.3% in December 2021, though one study in Japan suggests this has increased to over 90% in February 2022 [43]. Regional variation is driven by differences in the extent of SARS-CoV-2 infection and vaccination. This is exemplified by our monthly timeline of seroprevalence by region, 2020-2021, which provides estimates of evolving temporal changes of the global pandemic. We observed increases in seroprevalence following the emergence of variants in regions with available data (e.g., 6% (July 2020) to 41% (April 2021) in AFR following the Beta variant and 12% (February 2021) to 75% (August 2021) in SEAR following the Delta variant), demonstrating the substantial number of infections caused by more transmissible variants. In HIC regions, the increases in overall seroprevalence were driven by increased vaccine coverage in early 2021 (e.g., 6% (January 2021) to 95% (August 2021) in AMR HIC and 7% (January 2021) to 72% (August 2021) in EUR HIC), while we also observed increases in infection-induced seroprevalence following the Omicron variant (e.g., 7% (December 2021) to 34% (March 2022) in AMR HIC and 18% (October 2021) to 48% (March 2022) in EUR HIC). Another possibility for regional variation is the potential cross-reactivity of antibodies against P. falciparum or other common cold coronaviruses which has been remarked upon in the literature [44–46], which may impact seroprevalence estimates in areas of Africa or elsewhere with a high incidence of malaria. Our results add global representation and principled estimation of changes in seroprevalence over time as compared to previous evidence syntheses [9,11,12]. These estimates are similar to estimates of true infections by global epidemiological models. For example, our global estimate of seroprevalence attributable to infection (35.9%) is similar to the Institute of Health Metrics and Evaluation cumulative infection incidence estimate of 42.8% on 15 September 2021 [47]. Our analysis provides an orthogonal estimate based solely on robust seroprevalence data, using a method that has the added value of being easily interpretable and with fewer assumed parameters.

Our results provide evidence of considerable case under-ascertainment, indicating that many cases of SARS-CoV-2, including subclinical cases, are not captured by surveillance systems which in many countries are based on testing of symptomatic patients, have varying sensitivity in their definitions of positive cases, or simply have limited access to testing [48]. There was wide variation in under-ascertainment (as estimated through seroprevalence-case ratios) in all regions, income groups and over time, with higher ratios consistently observed in LMICs compared to HICs. Our ratios of seroprevalence to reported cases in late 2020 were comparable to other studies for AMR, EUR, and SEAR [9–12]. Our estimates of seroprevalence to cumulative incidence ratios for AFR, WPR, and EMR are novel, with no other analyses we found having systematically estimated ascertainment through seroprevalence in these regions; moreover, estimates of true infections from epidemiological models suggest the high levels of under-ascertainment suggested by this study are plausible [47].

We also provide more granular evidence of significant variation in infection by age by 10-year band. Children aged <10 years, but not children aged 10-19, were less likely to be seropositive compared to adults aged 20-29 years; similarly, adults aged >60 years, but not those aged 30-39, 40-49, or 50-59, were less likely to be seropositive than adults 20-29. These findings add nuance and granularity to differences in seroprevalence by age observed by other studies [10]. Lower seroprevalence in adults 60+ could be explained by immunosenescence that can lead to quicker seroreversion [49] higher mortality and hence a lower proportion of individuals with evidence of past infection, gaps in vaccine access, or more cautious behaviour resulting in fewer infections in this age group. There are several possible explanations for lower seroprevalence in children: milder infections, which are generally associated with lower antibody titers [50] school closures; and ineligibility for vaccination.

Our multivariable model suggests higher seroprevalence estimates were reported by low and lower-middle income countries compared to high-income countries, with the highest seroprevalence in lower-middle income countries (pre-vaccination). Potential explanations for this result are multifaceted and include weaker health system functionality and performance, lower capacity to isolate, and less stringent use of and ability to effectively implement PHSM. This is also consistent with findings by Rostami et al. [11] Our results suggest that an increase in overall PHSM stringency was associated with lower seroprevalence. This and other work has shown that the use of PHSM was associated with reduced SARS-CoV-2 infections, especially when implemented early and limiting population mobility [51–53]. Our model also found that sub-national studies have higher estimates than national studies; one hypothesis for this is that sub-national studies are often concentrated on cities or areas with denser populations, which may contribute to increased transmission of the virus. Further research is needed to validate this hypothesis. Finally, our results suggest that blood donors and residual sera studies may be good proxies for the general population, as there was no statistical difference between seroprevalence estimates in these sample frames compared to household and community samples.

In line with the equity principles of the Unity initiative, our dataset had global coverage, including a broad range of LMICs (one third of studies included in our dataset 1, n=177) and vulnerable HRP countries (14% of included studies). Related other global meta-analyses of seroprevalence had 23% and 35% LMIC coverage respectively [9,11]. Unity study collaborators shared timely evidence by uploading their aggregated and standardized early results to an open data repository, enabling geographic coverage and reducing publication bias.

### Strengths and limitations

Our regional and global meta-analysis estimates are timely, robust and geographically diverse with estimates from all WHO regions. The laboratory and epidemiological standardization enabled by the Unity Studies framework, as well as the analysis of only studies assessed to have low or moderate risk of bias using a validated risk of bias tool [25], enabled high-quality and comparable data. Despite this effort, there are still methodological differences between the meta-analyzed studies that may reduce their comparability. For example, 14% of studies in our analysis dataset (66/459, 18 of which were household-based) were convenience samples, which are less representative than population-based probability samples. To limit this bias, we required Unity-Aligned convenience samples to have a clearly defined sample frame (i.e., sampling of volunteers excluded). Our risk of bias evaluation also included subjective review of the demographic breakdown in the study, coverage of subgroup estimates, and author comments on representativeness of the sample, such that the most non-representative studies were rated high risk of bias and excluded from analysis.

A few limitations should be described. First, although we conducted meta-regression to explore heterogeneity of the included studies, there remained some residual heterogeneity that could not be explained quantitatively — likely driven by differences in disease transmission in the different countries and time points that serosurveys were conducted. Second, we did not account for waning of population immunity, so the present work likely underestimates the extent of past infection and case ascertainment. Thirdly, seroprevalence studies are cumulative, meaning that results reflect all COVID-19 countermeasures implemented up to the time of participant sampling and, thus, we cannot isolate the contributions of particular PHSM. Fourthly, while we screened study eligibility based on high assay performance criteria, different serological assays may yield varying results which should be taken into account when interpreting seroprevalence data. Some argue against combining studies using different assays, because assay performances can vary considerably leading to potential bias in the results. With the moderate seroprevalence values generally observed in our results (roughly 20% to 80%), we expect limited bias to be introduced by the different assays. Nevertheless, we conducted a sensitivity analysis adjusting estimates from individual studies with assay performance whenever available, and found that global and regional estimates remained similar. Finally, at certain points in time, our meta-analysis estimates were driven by studies from specific countries — either very populous countries (i.e. SEAR: India, AMR HIC: USA, AMR LMIC: Brazil, WPR: China), or countries in regions with scarce data during the time in question (e.g. EMR: 2 countries in early 2021). We also could not produce global estimates for late 2021/early 2022 due to the delays between when studies conducted their sampling (we extracted from the ‘sampling midpoint’), and when these results were later published or released within our search dates.

### Implications and next steps

Population-based seroprevalence studies primarily give a reliable estimate of the exposure to infection. In cases where antibodies can be measured quantitatively, it may also be possible to use them to assess the level of protection in a population, although there is currently no consensus on antibody-based correlates of protection for SARS-CoV-2 [4]. While antibodies persist in most infected individuals for up to a year (with early evidence pointing at up to 18 months) [54–57] the reinfection risk with the immune-escaping Omicron variant, is reported to be much higher than in previous VOCs in both vaccinated and previously infected individuals, indicating that the presence of antibodies is less indicative of a level of protection against infection. However, seroprevalence estimates remain indicative of protection against severe disease and death, as cellular immunity is unlikely to be disrupted even with an immune escaping VOCs.

Seroprevalence studies have been invaluable throughout the COVID-19 pandemic to understand the true extent and dynamics over time of SARS-CoV-2 infection and, to some extent, immunity. Serosurveillance provides key epidemiologic information that crucially supplements other routine data sources in populations. In populations with reported high vaccine coverage, seroprevalence studies provide a supplement to vaccine coverage data and are an important tool for the evaluation of vaccination programs. In populations with low vaccine coverage, it provides an estimate of cumulative incidence of past SARS-CoV-2 infection (including asymptomatic and mild disease), true case fatality ratio, and avoid many of the limitations of passive disease reporting systems which can be unreliable due to under-diagnosis and under-notification. Seroprevalence data can be used to compare seroprevalence between different groups (e.g. age, sex geography, etc.) to identify susceptible populations and thus inform decisions regarding the implementation of counter measures such as vaccination programs and PHSM [58]. A key challenge in implementing serosurveillance has been timeliness of study implementation, data analysis and reporting - as such, it will be important for public health decision makers to prioritize investment and establish emergency-mode procedures to facilitate timely study implementation early on in future outbreak or variant emergence responses as part of overall surveillance strategy [59,60]. There is also a need to continue to build national capacity with WHO and other partners to rapidly enable high quality study implementation and communication of findings in a format friendly to decision makers. The pandemic persists in large because of inequitable access to countermeasures tools such as vaccines; emphasizing the importance of equitable vaccine deployment globally, the strengthening of health systems and of tailored PHSM to mitigate disease transmission until high population protection is achieved. Globally standardized and quality seroprevalence studies continue to be essential to inform health policy decision-making around COVID-19 control measures, particularly in capacity-limited regions with low testing capacity and vaccination rates.

## Conclusion

In conclusion, our results show that seroprevalence has increased considerably over time, particularly in past months, due to infection in some regions and vaccination in others. Nevertheless, there is regional variation and 40 % of the global population remains susceptible to SARS-CoV-2. As our understanding of SARS-CoV-2 develops, the role of seroprevalence studies may change including the adaptation of study objectives and methodology to the situation. Currently, our global estimates of infections based on seroprevalence far exceed reported cases captured by surveillance systems. As we enter the third year of the COVID-19 pandemic, implementation of a global system for targeted, continuous, multi-pathogen, and standardized quality serosurveillance [59,60] is a crucial next step to monitor the COVID-19 pandemic and contribute to readiness for other emerging respiratory pathogens.

## Data Availability

Standardized results uploaded to Zenodo by UNITY study collaborators supporting the findings of this study are available at this community and DOIs for each included Zenodo study are cited in the Supplementary File (S5) https://zenodo.org/communities/unity-sero-2021?page=1&size=20.
Detailed information on each study is available in Supplement S4. Other relevant data are available in a data repository (doi:10.5281/zenodo.5773152) and/or available from the Zenodo community upon reasonable request.

## Supplementary material

Supplementary material file is attached.

## Contributors

Conceptualization: IB, LS, TN, MVK, RKA, NB.

Data curation: HCL, AnV, LA, JO, TA, LL, AiV, MW, XM, ZL, NB, HW, CC, MYL, ML, MS, GRD, NI, CZ, SP, HPR, TY, KCN, DK, SAA, ND, CD, NAD, EL, RKI, ASB, ELB, AS, JC.

Formal analysis: HW, RKA, DB, JP, MPC.

Funding acquisition: IB, MVK, RKA, NB, TY.

Investigation: IB, LS, HCL, AN, MV, BC, AnV, LA, AR, JO,TA, PW, LL, AiV, PM, KV, EB, SM, OMK, LS, CZ, EC, KKS, OAN, SEQ, JDN, SO, DB, PCM, MS, AFM, TMW, SN, BC, ED, TSD, SAA, LF, AC, TK, RS, AZM, SAS, MO, DAM, RKI, TS, VT, ML, AR, ITT, ASB, NT, SI, JCK, ASB, EAI, RAA, SB, TGH, PJL, HM, AC.

Methodology: HW, RKA, DB, NB, ML, HQ, CPY, TW, ND, CD, TE.

Project administration: IB, MVK, MW, NB, RKA, TY, PM, HQ, OMK, EC, OAN, IYR, EAA, SO, AB, MS, PC, SAA, ZN, AR, GSF, RKI, TS, FI, AR, ECB, ASB, BLH.

Resources: LS, BC, HCL, AnV, LA, AR, JO, TA, LL, AiV.

Supervision: IB, AnV, LA, AR, JO, TA, PRW, LL, RP, MVK, DB, MPC, JP, RKA, NB, TY.

Writing – original draft: IB, LS, HCL, AN, MW, HW, RKA.

Writing – review & editing: all authors.

All authors debated, discussed, edited, and approved the final manuscript. All authors had full access to the full data in the study and accepted responsibility to submit for publication.

## Declaration of interests

### Competing interests

The corresponding authors had full access to all the data in the study and had final responsibility for the decision to submit for publication. Authors were not precluded from accessing data in the study, and they accept responsibility to submit for publication.

The authors have read the journal’s policy and the authors of this manuscript have the following competing interests: RKA, MW, HW, ZL, XM, CC, MYL, DB, JP, MPC, ML, MS, GRD, NI, CZ, SP, HPR, TY, KCN, DK, SAA, ND, CD, NAD, EL, RKI, ASB, ELB, AS, JC and NB report grants from Canada’s COVID-19 Immunity Task Force through the Public Health Agency of Canada, and the Canadian Medical Association Joule Innovation Fund. RKA, MW, HW, ZL, CC, MYL, NB also report grants from the World Health Organization and the Robert Koch Institute. RKA reports personal fees from the Public Health Agency of Canada and the Bill and Melinda Gates Foundation Strategic Investment Fund, as well as equity in Alethea Medical (Outside the submitted work). MPC reports grants from McGill Interdisciplinary Initiative in Infection and Immunity and Canadian Institute of Health Research, and personal fees from GEn1E Lifesciences (Outside the submitted work), nplex biosciences (Outside the submitted work), Kanvas biosciences (Outside the submitted work). JP reports grants from MedImmune (Outside the submitted work) and Sanofi-Pasteur (Outside the submitted work), grants and personal fees from Merck (Outside the submitted work) and AbbVie (Outside the submitted work), and personal fees from AstraZeneca (Outside the submitted work). DB reports grants from the World Health Organization, Canadian Institutes of Health Research, Natural Sciences and Engineering Council of Canada (Outside the submitted work), Institute national d’excellence en santé et service sociaux (Outside the submitted work), and personal fees from McGill University Health Centre (Outside the submitted work) and Public Health Agency of Canada (Outside the submitted work). CC reports funding from Sanofi Pasteur (Outside of the submitted work). TY reports working for Health Canada as a part-time Senior Policy Analyst with the COVID-19 Testing and Screening Expert Panel, from Nov 2020-Jun 2021 (Outside of the submitted work).

## Data availability

Standardized results uploaded to Zenodo by Unity study collaborators supporting the findings of this study are available at this community and DOIs for each included Zenodo study are cited in the Supplementary File (S5) https://zenodo.org/communities/unity-sero-2021?page=1&size=20 (13). Detailed information on each study is available in Supplement S4. All relevant data are available in a public data repository (doi:10.5281/zenodo.5773152) and/or available from the Zenodo community.

## Acknowledgements

This work was supported by WHO through funding from the WHO COVID-19 Solidarity Response Fund and the German Federal Ministry of Health COVID-19 Research and Development Fund. IB, LS, AnV, LA, AR, JO, TA, PW, LL, AiV, RP, MVK are employed by WHO, and HCL, AN, MV, BC are WHO consultants. The designations employed and the presentation of the material in this publication do not imply the expression of any opinion whatsoever on the part of WHO concerning the legal status of any country, territory, city or area or of its authorities, or concerning the delimitation of its frontiers or boundaries. Dotted and dashed lines on maps represent approximate border lines for which there may not yet be full agreement.

SeroTracker (led by RKA, including MW, HW, ZL, XM, TY, CC, MYL, JP, MPC, DB, ML, MS, GRD, NI, CZ, SP, HPR, TY, KCN, DK, SAA, ND, CD, NAD, EL, RKI, ASB, ELB, AS, JC) is grateful for support from WHO, Canada’s COVID-19 Immunity Task Force through the Public Health Agency of Canada, the Robert Koch Institute, and the Canadian Medical Association Joule Innovation Fund. RKA additionally thanks the Rhodes Trust for its support.

We thank colleagues at partner organizations including WHO/Dubai Logistics Team; Myrna Charles, Kathleen Gallagher, Amen Ben Hamida, Christopher Murrill, Toni Whistler, Venkatachalam Udhayakumar (US Centers for Disease Control and Prevention); Eeva Broberg, Erika Duffell, Maria Keramarou and Pasi Penttinen (European Centre for Disease Prevention and Control), Vincent Richard (Institut Pasteur and the Institut Pasteur International Network); WHO regional offices (Jacob Barnor, Alina Guseinova, Jason M Mwenda, Dmitriy Pereyaslov, Harimahefa Razafimandimby, and WHO HQ (Michael Ryan). We also thank team members from Gabon: Rafiou Adamou, Ayola A Adegnika, Samira Z Assoumou, Rosemary A Audu, Paulin E Ndong, Paulin N Essone, Edgard B Ngoungou.

We would especially like to thank all WHO Unity Studies members, who are also named co-authors of this paper and listed in the section below. These and other collaborators, in all the countries who embarked in this global response effort to COVID-19, are recognized on a dedicated webpage on the Unity and WHO website,(61), as well as all individuals who supported, conducted or participated in each of the studies supported.

## Unity Studies Collaborator Group

**Unity Studies Collaborator Group: Ximena Aguilera^19^, Sheikh Al-Shoteri^20^, Eman A Aly^8^, Mauricio Apablaza^21^, Rosemary A Audu^22^, Amal Barakat^8,23^, Abdulla S Bin-Ghouth^24^, Enyew Birru^25^, Dejan Bokonjic^26^, Shelly Bolotin^27,28^, Henry K Bosa^29,30^, Emily L Boucher^31^, Elma Catovic-Baralija^32^, Alexei Ceban^33^, Annie Chauma-Mwale^34^, Judy Chen^35^, Battogtokh Chimeddorj^36^, Pui Shan Chung^10^, Cheryl Cohen^37^, Tienhan S Dabakuyo-Yonli^38^, Gabriel R Deveaux^31^, Boly Diop^39^, Titus H Divala^40^, Emily K Dokubo^41^, Irene O Donkor^42^, Claire Donnici^31^, Nathan Duarte^43^, Natalie A Duarte^44^, Timothy G Evans^7, 14^, Lee Fairlie^45^, Ousmane Faye^46^, Gudrun S Freidl^11^, Claudia González^19^, Tiffany G Harris^47,48^, Belinda L Herring^3^, Sopon Iamsirithaworn^49^, Gloria Icaza^50^, Rhoda Ila^51^, Natasha Ilincic^44^, Elsie A Ilori^52^, Francis Y Inbanathan^9^, Vicki Indenbaum^53^, John Kaldor^54^, Dayoung Kim^31^, Olatunji M Kolawole^55^, Jambo C Kondwani^56,57^, Tatiana Kuchuk^58^, Pritesh J Lalwani^59^, Moses Laman^60^, Evelyn Lavu^60^‡, Juliana Leite^12^, Michael Liu^61^, Emma Loeschnik^62^, Kristine Macartney^63^, Dorothy A Machalek^54,64^, Sheila Makiala-Mandanda^65,66^, Henri-Pierre Mallet^67^, Alexandre Manirakiza^68^, Pilly Mapira^51^, Pinyi N Mawien^69^, Puneet Misra^70^, Sanjin Musa^71,72^, Portia C Mutevedzi^73,74^, Osama A Najjar^75^, Sutthichai Nakphook^49^, Kim C Noel^16^, Zuridin Nurmatov^76^, Maria Ome-Kaius^60^, Eric M Osoro^77, 78^, Krishna P Paudel^79^, Sara Perlman-Arrow^7^, Sharif E Qaddomi^8,80^, Hude Quan^81^, Alissar Rady^82^, Hannah P Rahim^83^, Muriel Ramírez-Santana^84^, Izzat Y Rayyan^8,80^, Angel Rodriguez^12^, Karampreet Sachathep^85,86^, Mitchell Segal^13^, Anabel Selemon^31^, Tahmina Shirin^87^, Kristen A Stafford^88,89^, Laura Steinhardt^41^, Vanessa Tran^27,90^, Isidore T Traore^91,92^, Pablo Vial^93^, Tri Yunis M Wahyono^94^, Tyler Williamson^2,81^, Cedric P Yansouni^95,96^, Caseng Zhang^97^, Chong Zhuo Lin^98^ Didier Koumavii^99^

^19^Centro de Epidemiología y Políticas de Salud, Facultad de Medicina Clínica Alemana, Universidad del Desarrollo, Chile

^20^Aden University, Yemen

^21^Facultad de Gobierno, Universidad del Desarrollo, Chile

^22^Nigerian Institute of Medical Research, Nigeria

^23^Infectious Hazard Preparedness, WHO Health Emergencies Programme

^24^Hadhramout University, Al Mukalla, Yemen

^25^Ethiopian Public Health Institute, Addis Ababa, Ethiopia

^26^University of East Sarajevo Faculty of Medicine Foča, Bosnia and Herzegovina (Republic of Srpska)

^27^Public Health Ontario, Toronto, Ontario, Canada

^28^Dalla Lana School of Public Health, University of Toronto, Ontario, Canada

^29^Ministry of Health, Uganda

^30^Kellogg College, University of Oxford, England

^31^Cumming School of Medicine, University of Calgary, Calgary, Alberta, Canada

^32^Department for Blood Transmissible Disease Testing, Institute of Transfusion Medicine of the Federation of Bosnia and Herzegovina, Sarajevo, Bosnia and Herzegovina (Federation)

^33^World Health Organization, Country Office in the Republic of Moldova

^34^Public Health Institute of Malawi, Ministry of Health, Malawi

^35^Faculty of Medicine and Health Sciences, McGill University, Montreal, Quebec, Canada

^36^Department of Microbiology and Infection Prevention Control, School of Biomedicine, Mongolian National University of Medical Sciences, Ulaanbaatar, Mongolia

^37^Center for Respiratory Disease and Meningitis, National Institute for Communicable Diseases, Johanessburg, South Africa ^38^Epidemiology and Quality of Life Research Unit, INSERM U1231, Georges François Leclerc Centre – UNICANCER, Dijon, France ^39^Surveillance Division, Prevention Directorate, Ministry of Health and Social Action, Dakar, Senegal

^40^Kamuzu University of Health Sciences, University in Blantyre, Malawi

^41^U.S. Centers for Disease Control and Prevention, Atlanta, United States of America

^42^Noguchi Memorial Institute for Medical Research, Accra, Ghana

^43^Faculty of Engineering, McGill University, Quebec, Canada

^44^University of Toronto, Toronto, Ontario, Canada

^45^WITS Reproductive Health and HIV Institute, Faculty of Health Sciences, University of the Witwatersrand, South Africa

^46^Virology Department, Institut Pasteur de Dakar, Dakar, Senegal

^47^The International Center for AIDS Care and Treatment Programs (ICAP), Department of Epidemiology, Mailman School of Public Health, Columbia University, New York, United States of America

^48^Department of Epidemiology, Mailman School of Public Health, Columbia University

^49^Department of Disease Control, Ministry of Public Health, Thailand

^50^Instituto de Matemáticas, Universidad de Talca, Chile

^51^School of Medicine and Health Sciences, University of Papua New Guinea

^52^Nigeria Centre for Disease Control, Nigeria

^53^Central Virology Laboratory and Sheba Medical Center, Ministry of Health, Tel-Hashomer, Israel

^54^The Kirby Institute, University of New South Wales, Kensington, Australia

^55^Department of Microbiology, Faculty of Life Sciences, University of Ilorin, Ilorin, Nigeria

^56^Malawi-Liverpool-Wellcome Clinical Research Programme

^57^Liverpool School of Tropical Medicine

^58^Research and Production Center “Preventive Medicine” of the Ministry of Health of the Kyrgyz Republic, Kyrgyzstan

^59^Instituto Leônidas e Maria Deane (ILMD), Fiocruz Amazônia, Manaus, Amazonas, Brazil

^60^Papua New Guinea Institute of Medical Research, Goroka, Papua New Guinea

^61^Harvard Medical School, Boston, Massachusetts

^62^Department of Epidemiology and Biostatistics, Schulich School of Medicine and Dentistry, Western University, Ontario, Canada

^63^National Centre for Immunisation Research and Surveillance, Sydney, Australia

^64^Centre for Women’s Infectious Diseases, The Royal Women’s Hospital, Victoria, Australia

^65^Institut National de Recherche Biomédicale, Kinshasa, Democratic Republic of the Congo

^66^Université de Kinshasa, Democratic Republic of Congo

^67^Agence de Régulation de l’Action Sanitaire et Sociale de Polynésie française

^68^Institut Pasteur of Bangui, Central African Republic

^69^Preventive Health Services, Ministry of Health, Juba, South Sudan

^70^All India Institute of Medical Sciences, New Delhi, India

^71^Department of Epidemiology, Institute for Public Health of the Federation of Bosnia and Herzegovina, Bosnia and Herzegovina

^72^Sarajevo Medical School, University Sarajevo School of Science and Technology, Bosnia and Herzegovina

^73^South African Medical Research Council Vaccines and Infectious Diseases Analytics Research Unit, Faculty of Health Sciences, University of the Witwatersrand, Johannesburg, South Africa

^74^School of Pathology, Faculty of Health Sciences, University of Witwatersrand, South Africa

^75^Palestinian National Ministry of Health, Occupied Palestine Territory

^76^Scientific and Production Association for Preventive Medicine, Ministry of Health of the Kyrgyz Republic, Kyrgyzstan

^77^Washington State University, Global Health Kenya, Nairobi, Kenya

^78^Paul G. Allen School of Global Health, Washington State University, United States of America

^79^Epidemiology and Disease Control Division, Ministry of Health and Population, Nepal

^80^The Palestinian National Institute for Public Health, Ramallah, Occupied Palestine Territory

^81^Department of Community Health Sciences, University of Calgary, Calgary, Alberta, Canada

^82^World Health Organization, Country Office Beirut, Lebanon

^83^Boston Consulting Group

^84^Public Health Department, Facultad de Medicina, Universidad Católica del Norte, Coquimbo, Chile

^85^The International Center for AIDS Care and Treatment Programs (ICAP), Columbia University, New York, United States of America

^86^Department of Population and Family Health, Mailman School of Public Health, New York, United States of America

^87^Institute of Epidemiology, Disease Control and Research (IEDCR), Bangladesh

^88^Center for International Health, Education, and Biosecurity, University of Maryland School of Medicine, Baltimore, United States of America

^89^Division of Epidemiology and Prevention, Institute of Human Virology, University of Maryland School of Medicine

^90^Department of Laboratory Medicine and Pathobiology, University of Toronto, Toronto, Canada

^91^Programme de Recherche sur les maladies infectieuses, Centre MURAZ, Bobo-Dioulasso, Burkina Faso

^92^Institut Supérieur des Sciences de la Santé, Université Nazi Boni, Bobo-Dioulasso, Burkina Faso

^93^Instituto de Ciencias e Innovación en Medicina, Facultad de Medicina Clínica Alemana, Universidad del Desarrollo, Chile

^94^Department of Epidemiology, Faculty of Public Health, University of Indonesia, Depok, Indonesia

^95^J.D. MacLean Centre for Tropical Diseases, McGill University Health Centre, Quebec, Canada

^96^Divisions of Infectious Diseases and Medical Microbiology, McGill University Health Centre, Quebec, Canada

^97^Faculty of Health Sciences, McMaster University, Ontario, Canada

^98^Institute for Public Health, National Institutes of Health, Ministry of Health, Malaysia

^99^ Faculty of Public Health, University of Lomé

‡Deceased

## Supporting Files

### Supplementary Materials

Section S1: PRISMA Checklist. Section S2: Supplementary Methods. Section S.2.1 Additional Methods Description. Table S1: Study and Estimate-level Classification Decisions. Section S.2.2: Criteria for WHO Unity-alignment of sero-epidemiological investigations. Table S2: Unity-aligned vs. not unity-aligned sero-epidemiological studies. S.2.3: SeroTracker inclusion and exclusion criteria. Table S3: Criteria for including evidence. Table S4: Criteria for excluding evidence. Section S.2.4: Search Strategy. Section S.2.5: Risk of Bias Tool Breakdown. Section S3: Supplementary analysis information. Section S.3.1: Estimate Prioritization. Section S.3.2: Additional analysis description. Section S4: Supplementary Results. Section S.4.1: Studies included for descriptive analysis, meta-analysis, and ascertainment estimation. Table S5: Studies used in ascertainment analysis. Table S6: Additional studies used in meta-analysis and meta-regression. Table S7: Additional studies used in descriptive analysis. Section S.4.2: Risk of bias breakdown for all studies. Table S8: Risk of bias breakdown for all studies. Section S.4.3: Additional figures and results. Figure S1: WHO Member States with seroprevalence data identified. Figure S2: Risk of Bias Assessment for Included Studies. Figure S3: Asymptomatic Prevalence for Included Studies. Table S9: Estimated seroprevalence over time by reion and globally (uncorrected for test characteristics). Table S10: Estimated seroprevalence over time by region and globally (corrected for test characteristics). Table S11: Meta-regression results and model comparison. Section S5: References for all included studies. Section S6: PROSPERO Protocol Registration. Section S7: References cited in supplementary file.

## Notes

### Funding Statement

This work was supported by WHO (WHO COVID-19 Solidarity Response Fund, to IB;
German Federal Ministry of Health COVID-19 Research and Development Fund, to IB;
World Health Organisation funding, to RKA), the Public Health Agency of Canada
(Canada's COVID-19 Immunity Task Force through the Public Health Agency of
Canada, to RKA), the Canadian Medical Association (Joule Innovation Fund, to RKA),
and the Robert Koch Institute (funding to RKA). IB, LS, AnV, LA, AR, JO, TA, PW, LL,
AiV, RP, MVK are employed and receive salaries from WHO (one of the funders of this
study), and AN, MV, BC and HCL are WHO consultants.
Authors who are members of the SeroTracker Group (led by RKA, including MW, HW,
ZL, XM, TY, CC, MYL, JP, MPC, DB, ML, MS, GRD, NI, CZ, SP, HPR, TY, KCN, DK,
SAA, ND, CD, NAD, EL, RKI, ASB, ELB, AS, JC) were supported through the
aforementioned grants from WHO, Canada's COVID-19 Immunity Task Force through
the Public Health Agency of Canada, the Robert Koch Institute, and the Canadian
Medical Association Joule Innovation Fund.
WHO had a role in the study design, data collection, data analysis, data interpretation,
and the writing of the report. No other funders had any such role.

### Summary of Updates

Extended search, initially was from Jan 2020 to Oct 2021 in v1, then covering the full first two years of the pandemic (Jan 2020-Dec 2021) in v2, and now covering Jan 2020-May 2022 search dates in v3.

## References

1. World Health Organization. WHO Coronavirus Disease (COVID-19) Dashboard. 2021 [cited 2021 Jun 11]. Available from: https://covid19.who.int/

2. Bergeri I, Lewis HC, Subissi L, Nardone A, Valenciano M, Cheng B, et al. Early epidemiological investigations: World Health Organization UNITY protocols provide a standardized and timely international investigation framework during the COVID-19 pandemic. Influenza Other Respir Viruses. 2021 Oct 5 [cited 2021 Nov 4];n/a(n/a). Available from: https://onlinelibrary.wiley.com/doi/abs/10.1111/irv.12915

3. Khoury DS, Cromer D, Reynaldi A, Schlub TE, Wheatley AK, Juno JA, et al. Neutralizing antibody levels are highly predictive of immune protection from symptomatic SARS-CoV-2 infection. Nat Med. 2021 Jul;27(7):1205–11.

4. Earle KA, Ambrosino DM, Fiore-Gartland A, Goldblatt D, Gilbert PB, Siber GR, et al. Evidence for antibody as a protective correlate for COVID-19 vaccines. Vaccine. 2021 Jul;39(32):4423–8.

5. Mathieu E, Ritchie H, Ortiz-Ospina E, Roser M, Hasell J, Appel C, et al. A global database of COVID-19 vaccinations. Nat Hum Behav. 2021 Jul 1;5(7):947–53.

6. Barrie MB, Lakoh S, Kelly JD, Kanu JS, Squire J, Koroma Z, et al. SARS-CoV-2 antibody prevalence in Sierra Leone, March 2021: a cross-sectional, nationally representative, age-stratified serosurvey. medRxiv. 2021 Jan 1;2021.06.27.21259271.

7. Murhekar MV, Bhatnagar T, Thangaraj JWV, Saravanakumar V, Kumar MS, Selvaraju S, et al. SARS-CoV-2 seroprevalence among the general population and healthcare workers in India, December 2020-January 2021. Int J Infect Dis IJID Off Publ Int Soc Infect Dis. 2021 Jul;108:145–55.

8. Al-Abri SS, Al-Wahaibi A, Al-Kindi H, Kurup PJ, Al-Maqbali A, Al-Mayahi Z, et al. SARS-COV-2 antibody seroprevalence in the general population of Oman: results from four successive nationwide seroepidemiological surveys. Int J Infect Dis. 2021 Nov 1;112:269–77.

9. Bobrovitz N, Arora RK, Cao C, Boucher E, Liu M, Rahim H, et al. Global seroprevalence of SARS-CoV-2 antibodies: a systematic review and meta-analysis. PLoS ONE. 2021 Jun 23 [cited 2021 Mar 7];16(6). Available from: https://doi.org/10.1371/journal.pone.0252617

10. Chen X, Chen Z, Azman AS, Deng X, Sun R, Zhao Z, et al. Serological evidence of human infection with SARS-CoV-2: a systematic review and meta-analysis. Lancet Glob Health. 2021 Mar 8 [cited 2021 Mar 16];0(0). Available from: https://www.thelancet.com/journals/langlo/article/PIIS2214-109X(21)00026-7/abstract

11. Rostami A, Sepidarkish M, Fazlzadeh A, Mokdad AH, Sattarnezhad A, Esfandyari S, et al. Update on SARS-CoV-2 seroprevalence – Regional and worldwide. Clin Microbiol Infect. 2021 Sep 25 [cited 2021 Oct 7];0(0). Available from: https://www.clinicalmicrobiologyandinfection.com/article/S1198-743X(21)00539-5/fulltext

12. Byambasuren O, Dobler CC, Bell K, Rojas DP, Clark J, McLaws ML, et al. Comparison of seroprevalence of SARS-CoV-2 infections with cumulative and imputed COVID-19 cases: Systematic review. Darlix JLE, editor. PLoS ONE. 2021 Apr 2;16(4):e0248946.

13. WHO Unity Global SARS-CoV-2 Seroepidemiological Investigations. Zenodo Community. 2021 [cited 2021 Nov 5]. Available from: https://zenodo.org/communities/unity-sero-2021?page=1&size=20

14. Bobrovitz N, Arora RK, Boucher E, Yan T, Rahim H, Van Wyk J, et al. A systematic review and meta-analysis of SARS-CoV-2 seroprevalence studies aligned with the WHO population-based sero-epidemiological ‘Unity’ protocol CRD42020183634. PROSPERO; 2021. Available from: https://www.crd.york.ac.uk/prospero/display_record.php?ID=CRD42020183634

15. Moher D, Liberati A, Tetzlaff J, Altman DG. Preferred reporting items for systematic reviews and meta-analyses: the PRISMA statement. BMJ. 2009 Jul 21;339:b2535.

16. SeroTracker Group. SeroTracker: Our Data (Our Protocol). Airtable. 2021 [cited 2021 Nov 22]. Available from: serotracker.com/en/Data

17. COVID-19 Global Humanitarian Response Plan. United Nations Office for the Coordination of Humanitarian Affairs; 2020 Mar [cited 2021 Nov 4]. Available from: https://www.humanitarianresponse.info/en/programme-cycle/space/document/covid-19-global-humanitarian-response-plan

18. Morgan M, Luijkx T. Sensitivity and specificity of multiple tests. In: Radiopaedia.org. [cited 2021 Nov 19]. Available from: https://doi.org/10.53347/rID-34868

19. NRL. WHO COVID Evaluations Summary of Results. nrlquality. 2021 [cited 2021 Nov 22]. Available from: https://www.nrlquality.org.au/who-covid-evaluations-summary-of-results

20. Aus AGD of HTG. Post market review of COVID-19 point-of-care tests. Therapeutic Goods Administration (TGA). Australian Government Department of Health; 2021 [cited 2021 Nov 22]. Available from: https://www.tga.gov.au/post-market-review-covid-19-point-care-tests

21. FIND. FIND evaluation of SARS-CoV-2 antibody (Ab) detection tests. FIND: Diagnosis for All. 2021 [cited 2021 Nov 22]. Available from: https://www.finddx.org/sarscov2-eval-antibody/

22. Bewley KR, Coombes NS, Gagnon L, McInroy L, Baker N, Shaik I, et al. Quantification of SARS-CoV-2 neutralizing antibody by wild-type plaque reduction neutralization, microneutralization and pseudotyped virus neutralization assays. Nat Protoc. 2021 Jun;16(6):3114–40.

23. Ware H, Whelan M, Li Z, Cheng B, Valenciano, Lewis HC, et al. WHO Unity Sero-Epidemiological Studies Early Results Submission Template - Blank. 2021 Nov 23 [cited 2021 Nov 22]; Available from: https://zenodo.org/record/5719533

24. Munn Z, Moola S, Lisy K, Riitano D, Tufanaru C. Methodological guidance for systematic reviews of observational epidemiological studies reporting prevalence and cumulative incidence data. Int J Evid Based Healthc. 2015 Sep;13(3):147–53.

25. Bobrovitz N, Noel KC, Li Z, Cao C, Deveaux G, Selemon A, et al. SeroTracker-ROB: reproducible decision rules for risk of bias assessment of seroprevalence studies. Epidemiology; 2021 Nov [cited 2021 Nov 21]. Available from: http://medrxiv.org/lookup/doi/10.1101/2021.11.17.21266471

26. Migliavaca CB, Stein C, Colpani V, Munn Z, Falavigna M. Quality assessment of prevalence studies: a systematic review. J Clin Epidemiol. 2020 Nov;127:59–68.

27. Borges Migliavaca C, Stein C, Colpani V, Barker TH, Munn Z, Falavigna M, et al. How are systematic reviews of prevalence conducted? A methodological study. BMC Med Res Methodol. 2020 Apr 26;20(1):96.

28. World Health Organization. Regional offices. 2021 [cited 2021 Nov 19]. Available from: https://www.who.int/about/who-we-are/regional-offices

29. The World Bank, Van Rompaey C, Metreau E. World Bank Country and Lending Groups. 2021 [cited 2021 Nov 9]. Available from: https://datahelpdesk.worldbank.org/knowledgebase/articles/906519-world-bank-country-and-lending-groups

30. Balduzzi S, Rücker G, Schwarzer G. How to perform a meta-analysis with R: a practical tutorial. (“meta” package). Evid Based Ment Health. 2019;(22):153–60.

31. Bates D, Mächler M, Bolker B, Walker S. Fitting Linear Mixed-Effects Models Using lme4. J Stat Softw. 2015 [cited 2021 Nov 21];67(1). Available from: http://www.jstatsoft.org/v67/i01/

32. R Core Team. R: A language and environment for statistical computing. R Foundation for Statistical Computing; 2021. Available from: https://www.R-project.org/.

33. World Health Organization, London School of Hygiene and Tropical Medicine. Public Health and Social Measures. 2021 [cited 2021 Nov 9]. Available from: https://www.who.int/emergencies/diseases/novel-coronavirus-2019/phsm

34. World Population Prospects - Population Division - United Nations. [cited 2021 Nov 9]. Available from: https://population.un.org/wpp/

35. Viechtbauer W. Conducting meta-analyses in R with the metafor package. J Stat Softw. 2010;36(3):1–48.

36. Higgins J, Thomas J, Chandler J, Cumpston M, Page M, Welch V. Cochrane Handbook for Systematic Reviews of Interventions (version 6.2). Cochrane; 2021. Available from: www.training.cochrane.org/handbook

37. Wood SN. Fast stable restricted maximum likelihood and marginal likelihood estimation of semiparametric generalized linear models. J R Stat Soc Ser C Appl Stat. 2011;73(1):3–36.

38. GISAID. GISAID EpiCov Database. 2021 [cited 2022 Jan 17]. Available from: https://www.gisaid.org/hcov19-variants/

39. WHO Headquarters (HQ). WHO COVID-19 Case definition. World Health Organization; 2020 [cited 2021 Nov 5]. Available from: https://www.who.int/publications-detail-redirect/WHO-2019-nCoV-Surveillance_Case_Definition-2020.2

40. Ritchie H, Mathieu E, Rodés-Guirao L, Appel C, Giattino EOO, Hasell J, et al. Coronavirus Pandemic (COVID-19). Our World in Data. 2020 [cited 2021 Nov 4]. Available from: https://ourworldindata.org/coronavirus

41. Institute for Health metrics and Evaluation (IHME). COVID-19: Estimating the historical time series of infections. Seattle, Washington, USA: IHME, University of Washington; 2021. Available from: http://www.healthdata.org/special-analysis/covid-19-estimating-historical-infections-time-series

42. Beld MJC van den, Murk JL, Kluytmans J, Koopmans MPG, Reimerink J, Loo IHM van, et al. Increasing the Efficiency of a National Laboratory Response to COVID-19: a Nationwide Multicenter Evaluation of 47 Commercial SARS-CoV-2 Immunoassays by 41 Laboratories. J Clin Microbiol. 2021;59(9):e00767–21.

43. Ren Z, Nishimura M, Tjan LH, Furukawa K, Kurahashi Y, Sutandhio S, et al. Large-scale serosurveillance of COVID-19 in Japan: Acquisition of neutralizing antibodies for Delta but not for Omicron and requirement of booster vaccination to overcome the Omicron’s outbreak. PLoS ONE. 2022 Apr 5;17(4):e0266270.

44. Tso FY, Lidenge SJ, Peña PB, Clegg AA, Ngowi JR, Mwaiselage J, et al. High prevalence of pre-existing serological cross-reactivity against severe acute respiratory syndrome coronavirus-2 (SARS-CoV-2) in sub-Saharan Africa. Int J Infect Dis IJID Off Publ Int Soc Infect Dis. 2021 Jan;102:577–83.

45. Steinhardt LC, Ige F, Iriemenam NC, Greby SM, Hamada Y, Uwandu M, et al. Cross-Reactivity of Two SARS-CoV-2 Serological Assays in a Setting Where Malaria Is Endemic. J Clin Microbiol. 2021 Apr 14 [cited 2022 Jan 25]; Available from: https://journals.asm.org/doi/abs/10.1128/JCM.00514-21

46. Iesa MAM, Osman MEM, Hassan MA, Dirar AIA, Abuzeid N, Mancuso JJ, et al. SARS-CoV-2 and Plasmodium falciparum common immunodominant regions may explain low COVID-19 incidence in the malaria-endemic belt. New Microbes New Infect. 2020 Nov 1;38:100817.

47. Institute for Health metrics and Evaluation (IHME). COVID-19 Estimates. Seattle, Washington, USA: IHME, University of Washington; 2021. Available from: http://www.healthdata.org/covid/data-downloads

48. Suthar AB, Schubert S, Garon J, Couture A, Brown AM, Charania S. Coronavirus Disease Case Definitions, Diagnostic Testing Criteria, and Surveillance in 25 Countries with Highest Reported Case Counts. Emerg Infect Dis. 2022 Jan;28(1):148–56.

49. Hansen CH, Michlmayr D, Gubbels SM, Mølbak K, Ethelberg S. Assessment of protection against reinfection with SARS-CoV-2 among 4 million PCR-tested individuals in Denmark in 2020: a population-level observational study. The Lancet. 2021 Mar;397(10280):1204–12.

50. Seow J, Graham C, Merrick B, Acors S, Pickering S, Steel KJA, et al. Longitudinal observation and decline of neutralizing antibody responses in the three months following SARS-CoV-2 infection in humans. Nat Microbiol. 2020 Dec;5(12):1598–607.

51. Brauner JM, Mindermann S, Sharma M, Johnston D, Salvatier J, Gavenciak T, et al. Inferring the effectiveness of government interventions against COVID-19. Science. 2021;371(6531):eabd9338.

52. Deb P, Furceri D, Ostry J, Tawk N. The Effect of Containment Measures on the COVID-19 Pandemic. IMF Work Pap. 2020 Aug 7 [cited 2021 Nov 9];20(159). Available from: https://elibrary.imf.org/view/journals/001/2020/159/001.2020.issue-159-en.xml

53. Talic S, Shah S, Wild H, Gasevic D, Maharaj A, Ademi Z, et al. Effectiveness of public health measures in reducing the incidence of covid-19, SARS-CoV-2 transmission, and covid-19 mortality: systematic review and meta-analysis. BMJ. 2021 Nov 18;375:e068302.

54. Laing ED, Epsi NJ, Richard SA, Samuels EC, Wang W, Vassell R, et al. SARS-CoV-2 antibodies remain detectable 12 months after infection and antibody magnitude is associated with age and COVID-19 severity. Infectious Diseases (except HIV/AIDS); 2021 May [cited 2022 Feb 4]. Available from: http://medrxiv.org/lookup/doi/10.1101/2021.04.27.21256207

55. Puya-Dehgani-Mobaraki, Wang C, Floridi A, Floridi E, Zaidi AK. Long-Term Persistence of IgG Antibodies in recovered COVID-19 individuals at 18 months and the impact of two-dose BNT162b2 (Pfizer-BioNTech) mRNA vaccination on the antibody response. Infectious Diseases (except HIV/AIDS); 2022 Jan [cited 2022 Feb 4]. Available from: http://medrxiv.org/lookup/doi/10.1101/2022.01.18.22269349

56. Vitale J, Mumoli N, Clerici P, De Paschale M, Evangelista I, Cei M, et al. Assessment of SARS-CoV-2 Reinfection 1 Year After Primary Infection in a Population in Lombardy, Italy. JAMA Intern Med. 2021 Oct 1;181(10):1407.

57. Wei J, Matthews PC, Stoesser N, Maddox T, Lorenzi L, Studley R, et al. Anti-spike antibody response to natural SARS-CoV-2 infection in the general population. Nat Commun. 2021 Dec;12(1):6250.

58. Duarte N, Yanes-Lane M, Arora RK, Bobrovitz N, Liu M, Bego MG, et al. Adapting Serosurveys for the SARS-CoV-2 Vaccine Era. Open Forum Infect Dis. 2022 Feb 1;9(2):ofab632.

59. Metcalf CJE, Mina MJ, Winter AK, Grenfell BT. Opportunities and challenges of a world serum bank – authors’ reply. The Lancet. 2017;389(10066):252.

60. Mina MJ, Metcalf CJE, McDermott AB, Douek DC, Farrar J, Grenfell BT. A Global lmmunological Observatory to meet a time of pandemics. eLife. 2020 Jun 8;9:e58989.

61. World Health Organization. Unity Studies: Early Investigation Protocols. 2021 [cited 2021 Nov 22]. Available from: https://www.who.int/emergencies/diseases/novel-coronavirus-2019/technical-guidance/early-investigations

