## Supplementary Materials for "Global SARS-CoV-2 seroprevalence: a systematic review and meta-analysis of standardized population-based studies from Jan 2020-May 2022"

### S1: PRISMA Checklist

PRISMA Checklist [1]

| **Section and Topic** | **Item #** | **Checklist item** | **Location where item is reported** |
| --- | --- | --- | --- |
| **TITLE** | | |  |
| Title | 1 | Identify the report as a systematic review. | Title, lines 1-2 |
| **ABSTRACT** | | |  |
| Abstract | 2 | See the PRISMA 2020 for Abstracts checklist. | Abstract, lines 41-82 |
| **INTRODUCTION** | | |  |
| Rationale | 3 | Describe the rationale for the review in the context of existing knowledge. | Introduction, lines 111-144 |
| Objectives | 4 | Provide an explicit statement of the objective(s) or question(s) the review addresses. | Introduction, lines 146-152 |
| **METHODS** | | |  |
| Eligibility criteria | 5 | Specify the inclusion and exclusion criteria for the review and how studies were grouped for the syntheses. | Methods, lines 178-213; Supplement S2.2, S2.3 |
| Information sources | 6 | Specify all databases, registers, websites, organisations, reference lists and other sources searched or consulted to identify studies. Specify the date when each source was last searched or consulted. | Methods, lines 157-168; Supplement S2.4 |
| Search strategy | 7 | Present the full search strategies for all databases, registers and websites, including any filters and limits used. | Supplement S2.4 |
| Selection process | 8 | Specify the methods used to decide whether a study met the inclusion criteria of the review, including how many reviewers screened each record and each report retrieved, whether they worked independently, and if applicable, details of automation tools used in the process. | Methods, lines 174-178 |
| Data collection process | 9 | Specify the methods used to collect data from reports, including how many reviewers collected data from each report, whether they worked independently, any processes for obtaining or confirming data from study investigators, and if applicable, details of automation tools used in the process. | Methods, lines 174-178, lines 217-234 |
| Data items | 10a | List and define all outcomes for which data were sought. Specify whether all results that were compatible with each outcome domain in each study were sought (e.g. for all measures, time points, analyses), and if not, the methods used to decide which results to collect. | Methods, lines 217-222 |
|  | 10b | List and define all other variables for which data were sought (e.g. participant and intervention characteristics, funding sources). Describe any assumptions made about any missing or unclear information. | Methods, lines 217-222, lines 255-256; Supplement Table S1, S3.2 |
| Study risk of bias assessment | 11 | Specify the methods used to assess risk of bias in the included studies, including details of the tool(s) used, how many reviewers assessed each study and whether they worked independently, and if applicable, details of automation tools used in the process. | Methods, lines 236-244; Supplement S2.5 |
| Effect measures | 12 | Specify for each outcome the effect measure(s) (e.g. risk ratio, mean difference) used in the synthesis or presentation of results. | Methods, lines 277-309 |
| Synthesis methods | 13a | Describe the processes used to decide which studies were eligible for each synthesis (e.g. tabulating the study intervention characteristics and comparing against the planned groups for each synthesis (item #5)). | Methods, lines 259-263 |
|  | 13b | Describe any methods required to prepare the data for presentation or synthesis, such as handling of missing summary statistics, or data conversions. | Methods, lines 246-253, lines 259-263; Supplement S3.1 |
|  | 13c | Describe any methods used to tabulate or visually display results of individual studies and syntheses. | Methods, lines 291-294 |
|  | 13d | Describe any methods used to synthesize results and provide a rationale for the choice(s). If meta-analysis was performed, describe the model(s), method(s) to identify the presence and extent of statistical heterogeneity, and software package(s) used. | Methods, lines 266-316 |
|  | 13e | Describe any methods used to explore possible causes of heterogeneity among study results (e.g. subgroup analysis, meta-regression). | Methods, lines 266-275, lines 311-316 |
|  | 13f | Describe any sensitivity analyses conducted to assess robustness of the synthesized results. | Methods, lines 318-324; Supplement S3.2 |
| Reporting bias assessment | 14 | Describe any methods used to assess risk of bias due to missing results in a synthesis (arising from reporting biases). | N/A |
| Certainty assessment | 15 | Describe any methods used to assess certainty (or confidence) in the body of evidence for an outcome. | Methods, lines 289-291 |
| **RESULTS** | | |  |
| Study selection | 16a | Describe the results of the search and selection process, from the number of records identified in the search to the number of studies included in the review, ideally using a flow diagram. | Results, lines 329-349, Figure 1 |
|  | 16b | Cite studies that might appear to meet the inclusion criteria, but which were excluded, and explain why they were excluded. | N/A |
| Study characteristics | 17 | Cite each included study and present its characteristics. | Supplement S4.1 |
| Risk of bias in studies | 18 | Present assessments of risk of bias for each included study. | Supplement S4.2 |
| Results of individual studies | 19 | For all outcomes, present, for each study: (a) summary statistics for each group (where appropriate) and (b) an effect estimate and its precision (e.g. confidence/credible interval), ideally using structured tables or plots. | Supplement S4.1 |
| Results of syntheses | 20a | For each synthesis, briefly summarise the characteristics and risk of bias among contributing studies. | Results, lines 351-376, Table 1; Supplement Figure S2 |
|  | 20b | Present results of all statistical syntheses conducted. If meta-analysis was done, present for each the summary estimate and its precision (e.g. confidence/credible interval) and measures of statistical heterogeneity. If comparing groups, describe the direction of the effect. | Results, lines 406-471, Figure 3, Figure 4, Table 2; Supplement Figure S3, Table S9 |
|  | 20c | Present results of all investigations of possible causes of heterogeneity among study results. | Results, lines 383-405, Figure 2; Supplement Table S11 |
|  | 20d | Present results of all sensitivity analyses conducted to assess the robustness of the synthesized results. | Results, lines 473-476; Table S10 |
| Reporting biases | 21 | Present assessments of risk of bias due to missing results (arising from reporting biases) for each synthesis assessed. | N/A |
| Certainty of evidence | 22 | Present assessments of certainty (or confidence) in the body of evidence for each outcome assessed. | Results Figure 2, Figure 3, Figure 4; Supplement Table S9, Table S11 |
| **DISCUSSION** | | |  |
| Discussion | 23a | Provide a general interpretation of the results in the context of other evidence. | Discussion, lines 486-551 |
|  | 23b | Discuss any limitations of the evidence included in the review. | Discussion, lines 552-564 |
|  | 23c | Discuss any limitations of the review processes used. | Discussion, lines 556-585 |
|  | 23d | Discuss implications of the results for practice, policy, and future research. | Discussion, lines 586-628 |
| **OTHER INFORMATION** | | |  |
| Registration and protocol | 24a | Provide registration information for the review, including register name and registration number, or state that the review was not registered. | Methods, lines 168-172; Supplement S6 |
|  | 24b | Indicate where the review protocol can be accessed, or state that a protocol was not prepared. | Methods, lines 168-172; Supplement S6 |
|  | 24c | Describe and explain any amendments to information provided at registration or in the protocol. | N/A |
| Support | 25 | Describe sources of financial or non-financial support for the review, and the role of the funders or sponsors in the review. | Acknowledgements, lines 685-690; Declaration of interests, lines 650-655 |
| Competing interests | 26 | Declare any competing interests of review authors. | Declaration of interests, lines 650-675 |
| Availability of data, code and other materials | 27 | Report which of the following are publicly available and where they can be found: template data collection forms; data extracted from included studies; data used for all analyses; analytic code; any other materials used in the review. | Data availability, lines 677-682 |

### S2: Supplementary Methods

#### S.2.1: Additional methods description

**Search strategy, screening, extraction protocol**

Preliminary screening was conducted using the SeroTracker screening criteria [[2]](https://www.zotero.org/google-docs/?tDhCCq) for seroprevalence studies and then further refined by the ‘Unity’ aligned criteria (below, and S2.3), using Covidence software. Studies were then uploaded to AirTable for extraction of summary-level estimates and study factors.

Notably, published studies meeting the inclusion criteria, or otherwise deemed “Unity Aligned” beyond the May 20th cutoff date for this analysis, are extracted and updated weekly on an interactive and open-access dashboard and data platform, SeroTracker.com/Unity [[3]](https://www.zotero.org/google-docs/?wIId0q).

##### Table S1: Study- and Estimate-level Classification Decisions

| **Geographic scope** | **Definitions** |
| --- | --- |
| National | Research was conducted across multiple regions within a country |
| Sub-national | Research was conducted across multiple sub-regions or municipalities within the same state or province. |
| Local | Research was conducted in a single county, municipality, or neighborhood. |
| **Sample Frames** | **Definitions** |
| Household and community samples/samples from existing surveillance programs | Population based studies with community or house-hold based surveys derived from nonspecific members of the general population (including members selected from previously established population surveys) |
| Blood donors | Retention samples that have already been taken from banked blood derived from blood donation practices; or samples that are taken from individuals who have donated blood. |
| Residual sera | Samples that have already been taken in health care settings for a variety of reasons other than COVID serological testing; or general groups of non-COVID patients attending healthcare that do not solely belong to a specific patient population |
| Pregnant or parturient women | Samples that have already been taken from pregnant women during routine pregnancy trimester screening; or samples taken from or parturient women. |
| Persons living in slums | Samples taken from people residing in slum dwellings. |
| Representative patient population | Patient population groups where the health condition is common/endemic enough for it to constitute a meaningful sample of the general population (Malaria, HIV) |
| Multiple general populations | Samples that are composed of multiple distinct above-described populations with a single pooled seroprevalence estimate. |
| **Sampling method** | **Definitions** |
| Convenience | Non-probability sampling based on convenient sources of specimens. The sampling method should still have a clear and defined sample frame (i.e., advertisements in public spaces are not included) |
| Sequential | Non-probability sampling based on entire sampling performed within a self-determined time interval. The sampling method should still have a clear and defined sample frame, and is often drawn from convenience samples. |
| Stratified non-probability (Quota) | Non-probability sampling based on groupings of similar units. Groupings are then sampled in a non-randomized fashion, wherein nonresponders may be also ignored and replaced |
| Probability | Probability sampling based on randomized sampling methods wherein all individuals have an equal probability of being selected. |
| **Serological assays** | **Definitions ^^**[**^9^****^]^** |
| Multiplex | Assay that simultaneously detects presence of antibodies against multiple viral antigens (i.e., spike, nucleocapsid, envelope). |
| Rapid diagnostic test | Point-of-care diagnostic platform that can be used and evaluated directly at the testing site. Typical features include short turnaround time (<15 minutes), low cost, easy interpretation of results (typically binary), and ease of use with minimal training. |
| ELISA | Enzyme linked immunosorbent assay: Wells coated with the viral antigen of interest. After the formation of immune complexes, an enzyme-linked secondary antibody followed by the enzyme’s substrate are added, which results in chemical modification of the substrate in the presence of immune complexes, which is measured colorimetrically through spectroscopy. The relative light absorption at a particular wavelength is directly proportional to the amount of labeled complexes present. |
| CLIA | Chemiluminescent immunoassay (includes chemiluminescent microparticle immunoassays [CMIA]): Stationary solid particles or wells coated with the viral antigen of interest. After the formation of immune complexes, substrate is added which results in generation of light, the intensity of which is directly proportional to the amount of labeled complexes present. |
| IFA | Immunofluorescence assay. Antibodies bind to the SARS-CoV-2 protein of interest. Fluorescent dyes are coupled to these immune complexes in order to visualize the protein of interest using microscopy or flow cytometry |
| LFIA | Lateral flow immunoassay: A liquid sample containing the analyte of interest moves without the assistance of external forces (capillary action) through various zones of polymeric strips. Antibodies of interest can bind to immobilized antigen of interest in various zones. A secondary reporter antibody (typically functionalized with gold nanoparticles) then binds to these immune complexes resulting in visual band formation for interpretation. |
| Neutralization assay | Heat treated, inactivated serum is added to cell culture (typically Vero E6 cells). Presence of neutralizing antibodies against SARS-CoV-2 is measured by the extent of binding inhibition in cell culture inoculated with the virus . Quantification is determined by the serum dilutions needed to prevent cytopathic effects in the cell monolayer. |

#### S2.2: Criteria for WHO Unity-alignment of sero-epidemiological investigations

The primary aim of the Unity Studies is to contribute to global standardization of sero-epidemiological studies, which in turn will facilitate international comparison of data. The aim is to collect estimates at population level rather than in specific populations. To achieve this, the study protocol “[Population-based age-stratified seroepidemiological investigation protocol for coronavirus 2019 (COVID-19) infection](https://www.who.int/publications/i/item/WHO-2019-nCoV-Seroepidemiology-2020.2)” contains criteria outlining methodological approaches related to study design, data collection and analysis. The following key criteria should be adhered to:

- **Study population:** General population, any age group. Age stratification is not required. For studies not providing age-stratification, authors will be requested to provide that data if available.
- **Study design**: A one-time cross-sectional; or repeat cross sectional study; or a longitudinal cohort study. There is no sample size requirement.
- **Geographic scope**: national, sub-national, or local (ideally representative of overall burden of infection)
- **Sampling**: Both random sampling or convenience sampling is acceptable
- ***Random Sampling***: Individuals can be approached to participate in investigations using a variety of probability sampling techniques.
- ***Convenience Sampling***: Where a robust sampling frame can be described that approximates to a wider population, convenience samples are included as UNITY studies. Thus, for example, individuals attending medical services (e.g. blood donors, pregnant mothers, primary care attendees, etc.) or residual sera taken from patients for other a variety of other investigations can be included. Sampling of specific (e.g. healthcare workers) or closed (e.g. residents of institutions such as boarding schools or prisons) populations or recruited without a sampling frame (e.g. through advertising or volunteers at a venue) are not included.
- There may be circumstances where a study makes use of a patient population for a prevalence disease (e.g. some HIV clinics) as a proxy for the general population, subject to evaluation by the Unity team for inclusion on a case-by-case basis
- **Recruitment**: Study participants can be recruited through existing studies (e.g. general health surveys, research population, or patient cohorts (non-COVID-19 patient cohorts, etc.) or by establishing new studies.
- **Inclusion criteria**: All individuals identified for recruitment into the investigation, irrespective of acute or prior COVID-19 infection. All attempts should be made to include participants over a range of ages.
- **Exclusion criteria**: Refusal to give informed consent, or contraindication to venipuncture.

Suspected or confirmed acute or prior COVID-19 infection should not be excluded, with an exception for excluding active COVID-19 cases (for research staff safety reasons or local isolation and public health protocols).

- **Serological testing method**: Serological testing may be carried out using any testing method (and may use multiple assays), whether enzyme linked immunosorbent assay (ELISA), immunofluorescence assay (IFA), or rapid diagnostic test (RDT), **as long as they meet the specified sensitivity and specificity thresholds as per below.**
  - Test should be validated for the specimen type being used
  - Test must meet 90% sensitivity, 97% specificity or higher as reported by the manufacturer (or author-evaluated in-house test, if manufacturer data is not present)
- **An exception regarding the use of assays below a certain sensitivity and specificity can be made for studies conducted in** **humanitarian settings** (63 countries).
  - We will include all assay types from HRP countries regardless of their reported sensitivity and specificity, as long as the authors report an assay which has been independently validated from either an in-house evaluation or a WHO-approved head-to-head evaluation
- **Ethical clearance**: Availability of ethical approval/exception

Sero-epidemiological investigations that are conducted in line with the key criteria outlined above can be considered as **‘Unity-aligned’**.

While the criteria listed above are broad and inclusive, accounting for various circumstances and offering many possibilities for investigators to collect data, there are several methodological aspects that are **not acceptable** for ‘Unity-alignment’:

- There may be circumstances where probability sampling techniques cannot be used and therefore non-probability sampling can be justified. In this case it is pivotal to make sure that the sampling frame clearly corresponds to the study population. For example, while it is acceptable to approach a clearly defined number of individuals attending specific medical services within a pre-defined time frame, it is not acceptable to call for participants/volunteers where it is not possible to establish representativeness of the study population at large.
- Studies for which robust sampling frames that do not approximate to wider populations (e.g. through advertising or volunteers at a venue) or that sample specific (e.g. healthcare workers) or closed (e.g. residents of institutions such as boarding schools or prisons) populations.
- Use of serological assays below a certain sensitivity (90%) and specificity (97%) threshold, unless countries meet the HRP exception criteria explained above.
- Use of multiple assays with a non-overlapping algorithm (study uses two or more different tests for different subsets of samples)

##### Table S2. Unity-aligned vs not unity-aligned sero-epidemiological studies

|  | **Unity-aligned** | **Not Unity-aligned** |
| --- | --- | --- |
| **Study population** | Age-stratified sample from general population | Populations with substantial comorbidities or elevated exposure to COVID-19 |
| **Potential output and analysis** | - Seroprevalence of antibodies to COVID-19   Cumulative incidence of infection/Infection attack rates   - Fraction of asymptomatic infection - (Case fatality ratio) | Any other outcomes |
| **Study design** | Population-based, age-stratified prospective study   - One-time cross-sectional investigation - Repeated cross-sectional investigation in the same geographic area (but not sampling the same individuals) - Longitudinal cohort investigation with serial sampling of the same individuals each time |  |
| **Geographic scope** | - National - Sub-national - Local |  |
| **Sampling** | - Random sampling - Convenience sampling | Absence of sampling framework that approximates to a wider population |
| **Recruitment** | - Through new studies - Through existing studies |  |
| **Inclusion criteria** | - General population - All individuals identified for recruitment into the investigation, irrespective of age and irrespective of acute or prior COVID-19 infection. - All attempts should be made to include participants over a range of ages. | High-risk group i.e. those with substantial comorbidities or elevated exposure (e.g. healthcare workers with immediate exposure to COVID-19 patients) |
| **Exclusion criteria** | - Suspected or confirmed acute or prior COVID-19 infection should **not** be excluded | - Exclusion of study participants with suspected or confirmed acute or prior COVID-19 infection |
| **Testing method** | - ELISA - IFA - CLIA/CMIA - Multiplex immunoassay - Other comprehensively validated test | - Use of serological assays below sensitivity/specificity threshold.   (**except in humanitarian settings** reporting assays that have been independently validated from either an in-house evaluation or a WHO-approved head-to-head evaluation) |
| **Ethical considerations** | - Ethical clearance (i.e. ethical approval or exemption) prior to study implementation - Informed consent | - No ethical clearance (including the absence of ethical exemption) - Absence of informed consent |

#### S2.3 SeroTracker inclusion and exclusion criteria

##### Table S3: Criteria for including evidence (must meet all the criteria to be included)

| **Characteristics** | **Criteria for inclusion** |
| --- | --- |
| Population | - Humans of any age   - If it’s a general population sample, it may include COVID-19 antigen positive persons and those with suspected disease. |
| Study design | - Sero-surveys – defined as the collection and testing of serum (or proxy such as oral fluid) specimens from a sample of a defined population over a specified period of time to estimate the prevalence of antibodies against SARS-CoV-2 as an indicator of immunity - Cross-sectional, repeated cross sectional, and cohort study designs, with serology measurements at single time points or repeated at multiple time points |
| *Special design | - Include systematic reviews and meta-analysis of seroprevalence studies for the purpose of tracking evidence synthesis efforts |
| Sampling | - Any sampling method |
| Types of evidence | - Published or unpublished academic literature, grey literature (government or institutional reports), or media reports. Slide deck presentations were included if we could identify the person giving the presentation and the date of the presentation |
| Outcome measures | - Reports a seroprevalence estimate (proportion of the population with detectable antibodies) - Reports the number of participants enrolled in the study (denominator) - Reports study sampling end date/week - Reports the locations at which the study took places such that they could be categorized as neighbourhood, city, state/province/territory, or country |
| Languages | - Any |

##### Table S4: Criteria for excluding evidence (if any met then exclude)

| **Characteristics** | **Criteria for exclusion** |
| --- | --- |
| Population | - Non-human (e.g., *in silico*, animal, *in vitro*) - The study only included individuals with suspected, active, or previously diagnosed with COVID-19 using PCR, antigen testing, clinical assessment, or self-assessment - The study only included individuals vaccinated against SARS-CoV2 |
| Study design | - Study designs other than cross-sectional or cohort design: case reports, case-control studies, evaluations of serological tests, study protocols, |
| Sampling | - N/A |
| Types of evidence | - Multimedia sources of data (audio clips, video clips) were excluded due to the feasibility of extracting. Slide deck presentations were excluded if we could not identify the person giving the presentation and the date of the presentation - Dashboards not associated with a defined serology study |
| Outcome measures | - Only reports incidence or prevalence of SARS-CoV-2 antigen (as opposed to antibody) - Does not report study sampling end date/week - Does not report the number of participants included in the study (sample denominator) - Does not report the location at which the study took place |
| Language | - N/A |

We defined a seroprevalence study as the collection and testing of serum, plasma, whole blood, dried blood specimens, or saliva from a sample of a defined population over a specified period of time to estimate the proportion of the population with antibodies against SARS-CoV-2 [[4–6]](https://www.zotero.org/google-docs/?uwAmRr).

If a study sampled people with confirmed or suspected SARS-CoV-2 for the reason of obtaining estimates that were representative of the general population then the study was included. We included studies examining seroprevalence among close contacts of patients with confirmed infection. Each cluster was considered one unit, and therefore, studies of one unit (i.e., one cluster around a patient) were considered case reports and excluded.

Multimedia sources of data (audio clips, video clips) were excluded due to the feasibility of extracting. However, slide deck presentations were included if we could identify the person giving the presentation, the date of the presentation, and the location where the presentation was given.

Systematic reviews and meta-analysis of seroprevalence studies were included for the purposes of tracking evidence synthesis efforts however, these data will not be quantitatively analysed.

Assay validation studies that test the effectiveness of the assay on a sample of the population irrespective of PCR diagnostics status were included.

Articles not in English or French were included if they could be extracted in full using machine translation [[15]](https://www.zotero.org/google-docs/?cWtr1R).

#### S2.4: Search Strategy

Database: Ovid MEDLINE(R) and Epub Ahead of Print, In-Process & Other Non-Indexed Citations and Daily

Dates: January 1, 2020 to October 29, 2021

Notes: Covid-19 search terms were adapted from Ovid Expert Searches

| **#** | **Search terms** |
| --- | --- |
| 1 | exp Coronavirus/ |
| 2 | exp Coronavirus Infections/ |
| 3 | (coronavirus* or corona virus* or OC43 or NL63 or 229E or HKU1 or HCoV* or ncov* or covid* or sars-cov* or sarscov* or Sars-coronavirus* or Severe Acute Respiratory Syndrome Coronavirus*).tw,kf.[EB2] |
| 4 | or/1-3 |
| 5 | 4 not ((MERS or MERS-CoV or Middle East respiratory syndrome or camel* or dromedar* or equine or coronary or coronal or covidence* or covidien or influenza virus or HIV or bovine or calves or TGEV or feline or porcine or BCoV or PED or PEDV or PDCoV or FIPV or FCoV or SADS-CoV or canine or CCov or zoonotic or avian influenza or H1N1 or H5N1 or H5N6 or IBV).mp. or (animals/ not humans/)) |
| 6 | ((pneumonia or covid* or coronavirus* or corona virus* or ncov* or 2019-ncov or sars* or virus).tw,kf. or exp pneumonia/) and Wuhan.tw,kf. |
| 7 | (2019-ncov* or 2019nCov* or ncov19 or ncov-19 or 2019-novel CoV or sars-cov2* or sars-cov-2* or sarscov2* or sarscov-2* or Sars-coronavirus2 or Sars-coronavirus-2 or SARS-like coronavirus* or coronavirus 2 or coronavirus2* or corona or coronavirus-19 or covid19 or covid-19 or covid 2019 or ((novel or new or nouveau) adj2 (CoV or nCoV or covid or coronavirus* or corona virus or Pandemi*2)) or ((covid or covid19* or covid-19) and pandemic*2) or (coronavirus* and pneumonia)).tw,kf. |
| 8 | COVID-19.rx,px,ox. or severe acute respiratory syndrome coronavirus 2.os. |
| 9 | or/6-8 |
| 10 | 5 or 9 |
| 11 | immunoglobulins/ or antibodies/ or antibodies, blocking/ or exp antibodies, neutralizing/ or antibodies, viral/ or antigen-antibody complex/ or immune sera/ or exp immunoglobulin isotypes/ or immunoglobulin a/ or immunoglobulin d/ or immunoglobulin e/ or immunoglobulin g/ or immunoglobulin m/ |
| 12 | serologic tests/ or complement fixation tests/ or hemagglutination inhibition tests/ or neutralization tests/ |
| 13 | immunoassay/ or fluoroimmunoassay/ or exp immunoblotting/ or immunoenzyme techniques/ or exp enzyme-linked immunosorbent assay/ or exp enzyme-linked immunospot assay/ or immunosorbent techniques/ or serologic tests/ or complement fixation tests/ or hemagglutination inhibition tests/ or neutralization tests/ or Serology/di |
| 14 | (enzyme linked immunosorbent or enzyme-linked immunosorbent or ELISA or immunofluorescence or complement fixation or hemagglutination inhibition or immunoblot or western blot or neutrali*).tw,kf. |
| 15 | (antibod* or immunoglobulin* or immune globulin* or titer* or isotype* or IgG or IgM or IgA or neutrali* or sera or serum or serolog* or saliva).tw,kf. |
| 16 | or/11-14 |
| 17 | seroepidemiologic studies/ |
| 18 | incidence/ or prevalence/ |
| 19 | (seroconver* or seroprevalence or sero-prevalence or seroincidence or sero-incidence or seroepidemiolog* or sero-epidemiolog*).mp. |
| 20 | (inciden* or prevalen* or count* or rate*).mp. |
| 21 | (serosurvey or sero-survey or screen* or diagnostic).mp. |
| 22 | (seroconver* or seroprevalence or sero-prevalence or seroincidence or sero-incidence or seroepidemiolog* or sero-epidemiolog* or inciden* or prevalen* or silent or asymptomatic or serosurvey or sero-survey).tw,kf. |
| 23 | or/17-21 |
| 24 | 10 and (16 and 23) |
| 25 | 10 and 15 |
| 26 | 10 and 22 |
| 27 | or/24-26 |
| 28 | limit 27 to yr="2020-Current" |
| 29 | remove duplicates from 28 |

#### S2.5 Risk of Bias Tool Breakdown

To assess risk of bias, a decision rule assigned a rating of low, moderate, or high risk of bias to each study based on the specific combination of JBI checklist ratings for that study [[16]](https://www.zotero.org/google-docs/?e6PLHh). This decision rule was developed based on guidance on estimating disease prevalence [[17,18]](https://www.zotero.org/google-docs/?Oc2SJT) and was validated against assessments derived manually by two independent reviewers for 2,070 seroprevalence studies in the SeroTracker database, showing good agreement with manual review (intraclass correlation 0.77, 95% CI 0.74-0.80) in a recent preprint [[16]](https://www.zotero.org/google-docs/?HgfqVa).

| **Item 1: Was the sample frame appropriate to address the target population?** | |
| --- | --- |
| Yes | Sample frame described and approximated the target population |
| No | Sample frame did not approximate the target population (e.g., blood donors do not represent general population, doctors do not represent all health care providers) |
| Exclude | Sample frame not described |
| *Notes | The term “target population” should not be taken to infer every individual from everywhere or with similar disease or exposure characteristics. Instead, give consideration to specific population characteristics in the study, including age range, gender, morbidities, medications, and other potentially influential factors. For example, a sample frame may not be appropriate to address the target population if a certain group has been used (such as those working for one organisation, or one profession) and the results then inferred to the target population (i.e. working adults). A sample frame may be appropriate when it includes almost all the members of the target population (i.e. a census, or a complete list of participants or complete registry data). |

| **Item 2: Were study participants recruited in an appropriate way?** | |
| --- | --- |
| Yes | Probability sampling method (simple or stratified random) or entire sample (e.g., an entire town) was used |
| No | Non-probability sampling |
| Exclude | Sampling method not reported |

| **Item 3: Was the sample size adequate?** | |
| --- | --- |
| Yes | ≥599 |
| No | <599 |
| Exclude | Sample size not reported |
| *Notes | To calculate the required sample size we used an assumed prevalence of 2.5%, which was the global average estimated by the WHO in April, 2020.^1^ Based on guidance by the Joanna Briggs Institute and published medical statistical recommendations we selected a precision value that was half the assumed prevalence (1.25%) [2,3]. We calculated a minimum sample size of 599 using these inputs:  Sample size calculation:  Where n = sample size;  Z = Z statistic for level of confidence (95%);  P = expected prevalence (2.5% WHO global estimate);  d = precision (1.25%)    In cases where the sample size calculation was provided and the required sample for 80% power was below our threshold (n<599) but was met by the study, this item was marked as yes. |

| **Item 4: Were the study subjects and setting described in detail?** | |
| --- | --- |
| Yes | Average age and distribution of gender/sex provided |
| No | Neither age or gender/sex is provided, or only one of age and gender/sex is provided |

| **Item 5: Was data analysis conducted with sufficient coverage of the identified sample?** | |
| --- | --- |
| Yes | The demographic characteristics (gender/sex, age, and ethnicity) of the sample are at least somewhat representative of the population in both the main and sub-group analyses |
| No | The demographic characteristics (gender/sex, age, and ethnicity) of the sample are not representative of the population in both the main and sub-group analyses |
| Unclear | Information is not provided about demographic characteristics of the sample (gender/sex, age, and ethnicity) |

| **Item 6: Were valid methods used for the identification of the condition?** | |
| --- | --- |
| Yes | - The serological test used met the WHO Unity Study Criteria for serological tests: sensitivity minimum 90%, specificity minimum 97% - Two test algorithms with combined sensitivity >=90% and combined specificity >=97%. Calculation varies based on whether parallel or series testing was utilized - Two test algorithms using a commercial or in-house binding assay with confirmatory testing using virus neutralization assay irrespective of combined sensitivity/specificity - Multiple testing algorithms employing three or more tests using robust strategies and well performing tests including:   - Parallel testing with two tests followed by a third confirmatory test for discordant results. ⅔ positive is defined as true positive   - Series testing with 3 tests (⅔ positive for true seropositive)   - Parallel testing with 3 tests (At least one positive tests and no negative tests) |
| No | - The serological test used did not meet the WHO Unity Study Criteria for serological tests: sensitivity minimum 90%, specificity minimum 97% - The combined sensitivity and specificity for the two test algorithm is below the threshold - Complex multiple test algorithms without a rational or robust strategy |
| Exclude | Test sensitivity and specificity not reported |

| **Item 7: Was the condition measured in a standard, reliable way for all participants?** | |
| --- | --- |
| Yes | The same serology test was used for all participants |
| No | Different serology tests were used for participants |
| Unclear | No details were provided about which participants received which serology tests |

| **Item 8: Was there appropriate statistical analysis?** | |
| --- | --- |
| Yes | Does all of the following: corrects for test characteristics, corrects for population characteristics or the sample is somewhat representative of the population (probability sampling), and provides the information necessary to determine the numerator, denominator, prevalence estimate, and confidence interval. |
| No | Does not correct for population characteristics and the sample is not likely representative of the population (non-probability sampling), does not correct for test or provide the information necessary to correct for test characteristics, or does not provide the information necessary to determine the numerator, denominator, prevalence estimate, and confidence interval. |

| **Item 9: Was the response rate adequate, and if not, was the low response rate managed appropriately?** | |
| --- | --- |
| Yes | Response rate > 60% or the demographics of the sample were a reasonable match to those of the target population [5] |
| No | Response rate < 60% and the demographics of the sample were not a reasonable match to those of the target population |
| Unclear | Response rate not provided and it was unclear if the demographics of the sample differed from the target population |
| N/A | Response rate not applicable (e.g., blood donors, residual sera) |

| **Item 10: Overall risk of bias** | |
| --- | --- |
| Low | The estimates are very likely correct for the target population. To obtain a low risk of bias classification, all criteria must be met or departures from the criteria must be minimal and unlikely to impact on the validity and reliability of the prevalence estimate. These include sample sizes that are just below the threshold when all other criteria are met, reporting only some of characteristics of the sample, test characteristics below the threshold but corrections for the test performance, and response rates that are just below the threshold in the context of probability based sampling of an appropriate sampling frame with population weighted seroprevalence estimates. |
| Moderate | The estimates are likely correct for the target population. To obtain a moderate risk of bias classification, most criteria must be met and departures from the criteria are likely to have only a small impact on the validity and reliability of the prevalence estimates. |
| High | The estimates are not likely correct for the target population. To obtain a high risk of bias, many criteria must not be met or departures from criteria are likely to have a major impact on the validity and reliability of the prevalence estimates. |
| Unclear | There was insufficient information to assess the risk of bias. |

### S3 Supplementary analysis information

#### S3.1 Estimate Prioritization

Where multiple summary estimates were available or multiple estimates available for one subgroup, we prioritized estimates based on estimate adjustment (population adjustment, test adjustment), antibody isotypes measured (Total Ab, IgG, IgM, other), test type used (Neutralization, CLIA, ELISA, LFIA, other), and antibody targets measured (Multiple Ab targets, Spike, Nucleocapsid).

When multiple estimate adjustments were available, we prioritized as follows:

1. Test unadjusted and Population adjusted
2. Both unadjusted
3. Both adjusted
4. Test adjusted and Population unadjusted

When multiple isotypes were available, we prioritized as follows:

1. Total antibodies
2. IgG OR other antibodies
3. IgG only
4. IgM OR other antibodies
5. IgG AND other antibodies
6. IgM only
7. IgM AND other antibodies
8. Other

When multiple test types were available (i.e. from use of different tests), we prioritized as follows:

1. Neutralization
2. CLIA
3. ELISA
4. LFIA
5. Other

When multiple antibody targets were available (i.e. from use of different tests), we prioritized as follows:

1. Multiple antibody targets
2. Anti - Spike
3. Anti - Nucleocapsid

If the above defined prioritization criteria led to estimates left on equal terms, we further introduced test sensitivity and specificity (highest sensitivity and specificity) prioritization criteria. If multiple estimates were present (e.g. an extraction only contained equivalent subgroup data), we further pooled estimate information to generate a single summary estimate.

**The Python code used for our automated estimate prioritization is available on GitHub:** https://github.com/serotracker/iit-backend/blob/8059e9b905395de997f28a1a2dff5def795276ad/app/utils/estimate_prioritization/estimate_prioritization.py

#### S3.2 Additional analysis description

We drew confirmed case data from the WHO COVID-19 dashboard matched to nine days before the study mid-date to account for time from infection to seroconversion [[24,25]](https://www.zotero.org/google-docs/?oYGbBL). We used data from Our World in Data dashboard to pull vaccination data, timed fourteen days before the study’s mid date [19]. We also calculated the average global public health and social measures (PHSM) stringency index value on a scale from 1 to 10 in the country of the study between the date of the 100th reported case and 14 days before the study mid date. The PHSM data was taken from a global dataset created in collaboration between the London School of Hygiene and Tropical Medicine (LSHTM) and WHO, which aggregates six indicators including masks, international travel, gatherings, schools, workplaces and businesses, and movement into one overall stringency index [[20]](https://www.zotero.org/google-docs/?3qSxOQ). Finally, we defined the transmission phase in the country of the study using the seven-day rolling average of daily confirmed cases from the WHO dashboard. We smoothed the case series using smoothing splines and identified the peaks of the series using local maxima. We then classified the study mid date as before the peak of 1st SARS-CoV-2 wave, after the peak of 1st SARS-CoV-2 wave, or after the peak of 2nd SARS-CoV-2 wave in the country of the study.

We produced estimates of seroprevalence over time by first meta-analyzing studies by country in 12-week rolling windows. For each month starting January 1, 2020, we first identified the sub-national and national studies in each region within a 12-week moving window centred on that week. If there was a single study in the window, we specified seroprevalence to be equal to that study’s estimate. If there were multiple studies in the window, we pooled the studies using a random intercept logistic regression model (rma.glmm from R package metafor) to calculate a pooled seroprevalence for that week. We then advanced the 12-week window to the next week and repeated the process.

We then produced monthly regional estimates by taking weighted averages of country estimates by population, ensuring that country contributions to these estimates are proportional to country population. The regional estimates were calculated as

$p_{r}=\Sigma w_{c}\times p_{c}$,

Where weights $w_{c}$ are weights for each country, normalized to sum to 1, and $p_{c}$ are the seroprevalence proportions in each region computed via random-effects meta-analysis (‘metaprop’). The standard error of the global estimates was calculated using the laws of variance as

$${se}_{r}=\sqrt{\Sigma w_{c}^{2}\times{se}_{c}^{2}},$$

where ${se}_{c}$ are the standard errors of the seroprevalence proportions in each country computed via metaprop on the logit scale. The confidence intervals of the regional estimates were calculated using the normal distribution as $p_{r}\pm2\times{se}_{r}.$ The prediction intervals were calculated similarly using the standard errors for prediction intervals in each country computed via metaprop on the logit scale.

We computed global estimates as the population-weighted average of the regional estimates, carrying forward the latest regional estimates to September 2021 to represent all regions.

To capture the trend in seroprevalence in each WHO region and globally, we took the weighted point estimates and fitted a flexible, smooth function of time using non-parametric regression. To anchor the trend line in the early pandemic period before the first serosurveys were conducted, we specified seroprevalence to be less than 0.1 percent before the 500th confirmed case of COVID-19 in each region.

We repeated the previous step using only national studies and used the results to compute the ratio between regional and global seroprevalence and the corresponding cumulative incidence of confirmed SARS-CoV-2 cases. This analysis measured to what extent confirmed SARS-CoV-2 cases underestimated seroprevalence, as reported by the WHO’s case definition [[21]](https://www.zotero.org/google-docs/?P1FBxl).

To produce estimates of ascertainment, we adjusted 2021 seroprevalence data for antibody target and vaccination to generate estimates of seroprevalence attributable to infection. In countries administering only vaccines using Spike (S) protein antigens (e.g., mRNA, viral vector), we calculated the ascertainment ratio from January to April 2021 using only studies that detected anti-nucleocapsid (N) seroprevalence, as a proxy for infection-induced seroprevalence. In countries administering inactivated vaccines that may generate both anti-S and anti-N responses (e.g., Sinopharm, Bharat Biotech), we adjusted the reported seroprevalence for studies between January and April 2021 using the formula

${SP}_{inf}=({SP}_{obs}-v)/(1-v)$,

Where ${SP}_{obs}$ is observed seroprevalence, $v$ is the vaccination rate in adults, and ${SP}_{inf}$is a proxy for infection-induced seroprevalence [[22]](https://www.zotero.org/google-docs/?uJ2NwI).

To examine study and geographic factors affecting seroprevalence estimates, we constructed a Poisson generalized linear mixed-effects model with log link function using the glmer function from the lme4 package in R [23]. Categorical covariates in the meta-regression, coded as indicator variables, included country WHO region (reference: Americas), country World Bank income class (reference: High income), study geographic scope (reference: national), study sample frame (reference: household and community samples), and transmission phase in the country of the study at the time of the study (reference: before peak of 1st wave). Continuous covariates included cumulative incidence of reported cases per hundred in the country of the study and LSHTM global index tracking PHSM in the country of the study. The PHSM index scale ranged from 0 (least stringent) to 10 (most stringent). We performed model comparison using all possible models that could be constructed by dropping a single *a priori* specified predictor, i.e., using $p-1$ predictors where there were $p$ predictors in total.

In a sensitivity analysis, we adjusted seroprevalence estimates for test characteristics through Bayesian measurement error models, with binomial sensitivity and specificity distributions. The sensitivity and specificity values for correction were prioritized from the WHO SARS-CoV-2 Test Kit Comparative Study conducted at the NRL Australia [5], followed by a multicentre evaluation of 47 commercial SARS-CoV-2 immunoassays by 41 Dutch laboratories [7], and from independent evaluations by study authors where author-designed assays were used.

**The Python code used for our test adjustment is available on GitHub:**

https://github.com/serotracker/iit-backend/blob/011fdb721d82abea449206ac5c84c86684b0867a/app/namespaces/test_adjustment/test_adjustment_service.py

### S4 Supplementary Results

#### S4.1 Studies included for descriptive analysis, meta-analysis, and ascertainment estimation

##### Table S5. Studies used in ascertainment analysis.

###### Africa region

| Country (Location) | Author (Organization) | Sampling Dates (YMD) | Geographic scope | Overall risk of bias | Sample size | Age | Sex | Sampling method | Sample frame | Seroprevalence (95% CI) | Test Manufacturers - Isotypes - Names | Test Sens, Spec | Sens, Spec Source |
| --- | --- | --- | --- | --- | --- | --- | --- | --- | --- | --- | --- | --- | --- |
| Cameroon - HRP | Tiffany Harris (ICAP at Columbia University)[1] | 2020-10-20 - 2020-12-16 | National | Moderate | 9,332 | Multiple groups | All | Convenience | Household and community samples | 10.5% (95% CI 9.1-12%) | Abbott Laboratories - IgG - Abbott Architect SARS-CoV-2 IgG,Beijing Wantai Biological - IgG IgM IgA - Wantai SARS-CoV-2 Total Ab ELISA | 0.957, 1 | Complex test algorithm: Manual calculation |
| Ethiopia - HRP | Enyew Birru Tadesse (Federal Ministry of Health)[2] | 2020-06-24 - 2020-07-08 | National | Low | 16,932 | Multiple groups | All | Probability | Household and community samples | 1.9% | Abbott Laboratories - IgG - Abbott Architect SARS-CoV-2 IgG | 0.99, 1 | Validated by manufacturer |
| Gabon | Dr Paulin ESSONE NDONG, Pr Edgar Brice NGOUNGOU, Pr Ayola Akim ADEGNIKA (Laboratoire National de Santé Publique, CIRMF, CERMEL, Laboratoire CHUMEFJE, Laboratoire Bactério-virologie USS)[3] | 2021-11-28 - 2021-12-20 | National | Moderate | 3,659 | Multiple groups | All | Probability | Household and community samples | 86.8% (95% CI 85.6-87.9%) | Beijing Wantai Biological - IgG IgM IgA - Wantai SARS-CoV-2 Total Ab ELISA | 0.967, 0.975 | Validated by manufacturer |
| Ghana | Irene Owusu Donkor (Noguchi Memorial Institute for Medical Rsearch)[4] | 2021-12-12 - 2021-12-19 | National | Moderate | 1,027 | Multiple groups | All | Probability | Household and community samples | 86.7% (95% CI 84.4-88.7%) | Beijing Wantai Biological - IgG IgM IgA - Wantai SARS-CoV-2 Total Ab ELISA | 0.967, 0.975 | Validated by manufacturer |
| Ghana | Irene Owusu Donkor (Noguchi Memorial Institute for Medical Rsearch)[4] | 2021-07-01 - 2021-07-29 | National | Moderate | 2,275 | Multiple groups | All | Probability | Household and community samples | 66.9% (95% CI 64.9-68.8%) | Beijing Wantai Biological - IgG IgM IgA - Wantai SARS-CoV-2 Total Ab ELISA | 0.967, 0.975 | Validated by manufacturer |
| Ghana | Irene Owusu Donkor (Noguchi Memorial Institute for Medical Rsearch)[4] | 2021-05-03 - 2021-06-30 | National | Moderate | 2,234 | Multiple groups | All | Probability | Household and community samples | 65.9% (95% CI 63.9-67.9%) | Beijing Wantai Biological - IgG IgM IgA - Wantai SARS-CoV-2 Total Ab ELISA | 0.967, 0.975 | Validated by manufacturer |
| Ghana | Irene Owusu Donkor (Noguchi Memorial Institute for Medical Rsearch)[4] | 2021-02-13 - 2021-02-23 | National | Moderate | 401 | Multiple groups | All | Probability | Household and community samples | 29.4% (95% CI 25-34.2%) | Beijing Wantai Biological - IgG IgM IgA - Wantai SARS-CoV-2 Total Ab ELISA | 0.967, 0.975 | Validated by manufacturer |
| Kenya - HRP | Sophie Uyoga (KEMRI-Wellcome Trust Research Programme)[5] | 2021-01-03 - 2021-03-15 | National | Moderate | 3,018 | Multiple groups | All | Convenience | Blood donors | 45.3% | NA - IgG - Author designed (ELISA) -Spike | 0.927, 0.99 | Validated by independent authors/third party/non-developers |
| Kenya - HRP | Ifedayo M. O. Adetifa (KEMRI-Wellcome Trust Research Programme)[6] | 2020-08-20 - 2020-09-30 | National | Moderate | 3,723 | Adults (18-64 years) | All | Sequential | Blood donors | 9.1% | NA - IgG - Author designed (ELISA) -Spike | 0.927, 0.99 | Validated by independent authors/third party/non-developers |
| Malawi | Kondwani Jambo (Malawi-Liverpool-Wellcome Trust Clinical Research programme, Liverpool School of Tropical Medicine)[7] | 2021-07-01 - 2021-07-31 | National | Low | 288 | Multiple groups | All | Probability | Blood donors | 70.1% | Beijing Wantai Biological - IgG IgM IgA - Wantai SARS-CoV-2 Total Ab ELISA,Omega diagnostics - IgG - COVID-19 IgG ELISA | 0.928, 1 | Complex test algorithm: Manual calculation |
| Malawi | Kondwani Jambo (Malawi-Liverpool-Wellcome Trust Clinical Research programme, Liverpool School of Tropical Medicine)[7] | 2021-06-01 - 2021-06-30 | National | Low | 319 | Multiple groups | All | Probability | Blood donors | 61.8% | Beijing Wantai Biological - IgG IgM IgA - Wantai SARS-CoV-2 Total Ab ELISA,Omega diagnostics - IgG - COVID-19 IgG ELISA | 0.928, 1 | Complex test algorithm: Manual calculation |
| Malawi | Kondwani Jambo (Malawi-Liverpool-Wellcome Trust Clinical Research programme, Liverpool School of Tropical Medicine)[7] | 2021-05-01 - 2021-05-31 | National | Low | 298 | Multiple groups | All | Probability | Blood donors | 63.6% | Beijing Wantai Biological - IgG IgM IgA - Wantai SARS-CoV-2 Total Ab ELISA,Omega diagnostics - IgG - COVID-19 IgG ELISA | 0.928, 1 | Complex test algorithm: Manual calculation |
| Malawi | Kondwani Jambo (Malawi-Liverpool-Wellcome Trust Clinical Research programme, Liverpool School of Tropical Medicine)[7] | 2021-04-01 - 2021-04-30 | National | Low | 296 | Multiple groups | All | Probability | Blood donors | 64.7% | Beijing Wantai Biological - IgG IgM IgA - Wantai SARS-CoV-2 Total Ab ELISA,Omega diagnostics - IgG - COVID-19 IgG ELISA | 0.928, 1 | Complex test algorithm: Manual calculation |
| Malawi | Kondwani Jambo (Malawi-Liverpool-Wellcome Trust Clinical Research programme, Liverpool School of Tropical Medicine)[7] | 2021-03-01 - 2021-03-31 | National | Low | 298 | Multiple groups | All | Probability | Blood donors | 53.1% | Beijing Wantai Biological - IgG IgM IgA - Wantai SARS-CoV-2 Total Ab ELISA,Omega diagnostics - IgG - COVID-19 IgG ELISA | 0.928, 1 | Complex test algorithm: Manual calculation |
| Malawi | Kondwani Jambo (Malawi-Liverpool-Wellcome Trust Clinical Research programme, Liverpool School of Tropical Medicine)[7] | 2021-02-01 - 2021-02-28 | National | Low | 365 | Multiple groups | All | Probability | Blood donors | 53.1% | Beijing Wantai Biological - IgG IgM IgA - Wantai SARS-CoV-2 Total Ab ELISA,Omega diagnostics - IgG - COVID-19 IgG ELISA | 0.928, 1 | Complex test algorithm: Manual calculation |
| Malawi | Kondwani Jambo (Malawi-Liverpool-Wellcome Trust Clinical Research programme, Liverpool School of Tropical Medicine)[7] | 2021-01-01 - 2021-01-31 | National | Low | 303 | Multiple groups | All | Probability | Blood donors | 26.7% | Beijing Wantai Biological - IgG IgM IgA - Wantai SARS-CoV-2 Total Ab ELISA,Omega diagnostics - IgG - COVID-19 IgG ELISA | 0.928, 1 | Complex test algorithm: Manual calculation |
| Malawi | Kondwani Jambo (Malawi-Liverpool-Wellcome Trust Clinical Research programme, Liverpool School of Tropical Medicine)[7] | 2020-12-01 - 2020-12-31 | National | Low | 263 | Multiple groups | All | Probability | Blood donors | 11% | Beijing Wantai Biological - IgG IgM IgA - Wantai SARS-CoV-2 Total Ab ELISA,Omega diagnostics - IgG - COVID-19 IgG ELISA | 0.928, 1 | Complex test algorithm: Manual calculation |
| Malawi | Kondwani Jambo (Malawi-Liverpool-Wellcome Trust Clinical Research programme, Liverpool School of Tropical Medicine)[7] | 2020-11-01 - 2020-11-30 | National | Low | 365 | Multiple groups | All | Probability | Blood donors | 10.5% | Beijing Wantai Biological - IgG IgM IgA - Wantai SARS-CoV-2 Total Ab ELISA,Omega diagnostics - IgG - COVID-19 IgG ELISA | 0.928, 1 | Complex test algorithm: Manual calculation |
| Malawi | Matthews Kagoli/Joe Theu (Public Health Institute of Malawi(PHIM) MOH)[8] | 2020-10-14 - 2020-12-08 | National | Low | 4,261 | Multiple groups | All | Probability | Household and community samples | 7.8% | Beijing Wantai Biological - IgG IgM IgA - Wantai SARS-CoV-2 Total Ab ELISA | 0.967, 0.975 | Validated by manufacturer |
| Malawi | Kondwani Jambo (Malawi-Liverpool-Wellcome Trust Clinical Research programme, Liverpool School of Tropical Medicine)[7] | 2020-10-01 - 2020-10-31 | National | Low | 324 | Multiple groups | All | Probability | Blood donors | 21.1% | Beijing Wantai Biological - IgG IgM IgA - Wantai SARS-CoV-2 Total Ab ELISA,Omega diagnostics - IgG - COVID-19 IgG ELISA | 0.928, 1 | Complex test algorithm: Manual calculation |
| Malawi | Kondwani Jambo (Malawi-Liverpool-Wellcome Trust Clinical Research programme, Liverpool School of Tropical Medicine)[7] | 2020-09-01 - 2020-09-30 | National | Low | 309 | Multiple groups | All | Probability | Blood donors | 13% | Beijing Wantai Biological - IgG IgM IgA - Wantai SARS-CoV-2 Total Ab ELISA,Omega diagnostics - IgG - COVID-19 IgG ELISA | 0.928, 1 | Complex test algorithm: Manual calculation |
| Malawi | Kondwani Jambo (Malawi-Liverpool-Wellcome Trust Clinical Research programme, Liverpool School of Tropical Medicine)[7] | 2020-08-01 - 2020-08-31 | National | Low | 192 | Multiple groups | All | Probability | Blood donors | 9.9% | Beijing Wantai Biological - IgG IgM IgA - Wantai SARS-CoV-2 Total Ab ELISA,Omega diagnostics - IgG - COVID-19 IgG ELISA | 0.928, 1 | Complex test algorithm: Manual calculation |
| Malawi | Kondwani Jambo (Malawi-Liverpool-Wellcome Trust Clinical Research programme, Liverpool School of Tropical Medicine)[7] | 2020-07-01 - 2020-07-31 | National | Low | 204 | Multiple groups | All | Probability | Blood donors | 8.2% | Beijing Wantai Biological - IgG IgM IgA - Wantai SARS-CoV-2 Total Ab ELISA,Omega diagnostics - IgG - COVID-19 IgG ELISA | 0.928, 1 | Complex test algorithm: Manual calculation |
| Malawi | Kondwani Jambo (Malawi-Liverpool-Wellcome Trust Clinical Research programme, Liverpool School of Tropical Medicine)[7] | 2020-06-01 - 2020-06-30 | National | Low | 242 | Multiple groups | All | Probability | Blood donors | 1.4% | Beijing Wantai Biological - IgG IgM IgA - Wantai SARS-CoV-2 Total Ab ELISA,Omega diagnostics - IgG - COVID-19 IgG ELISA | 0.928, 1 | Complex test algorithm: Manual calculation |
| Malawi | Kondwani Jambo (Malawi-Liverpool-Wellcome Trust Clinical Research programme, Liverpool School of Tropical Medicine)[7] | 2020-05-01 - 2020-05-31 | National | Low | 194 | Multiple groups | All | Probability | Blood donors | 1.1% | Beijing Wantai Biological - IgG IgM IgA - Wantai SARS-CoV-2 Total Ab ELISA,Omega diagnostics - IgG - COVID-19 IgG ELISA | 0.928, 1 | Complex test algorithm: Manual calculation |
| Malawi | Kondwani Jambo (Malawi-Liverpool-Wellcome Trust Clinical Research programme, Liverpool School of Tropical Medicine)[7] | 2020-04-01 - 2020-04-30 | National | Low | 205 | Multiple groups | All | Probability | Blood donors | 0.5% | Beijing Wantai Biological - IgG IgM IgA - Wantai SARS-CoV-2 Total Ab ELISA,Omega diagnostics - IgG - COVID-19 IgG ELISA | 0.928, 1 | Complex test algorithm: Manual calculation |
| Malawi | Kondwani Jambo (Malawi-Liverpool-Wellcome Trust Clinical Research programme, Liverpool School of Tropical Medicine)[7] | 2020-01-01 - 2020-01-31 | National | Low | 227 | Multiple groups | All | Probability | Blood donors | 0% | Beijing Wantai Biological - IgG IgM IgA - Wantai SARS-CoV-2 Total Ab ELISA,Omega diagnostics - IgG - COVID-19 IgG ELISA | 0.928, 1 | Complex test algorithm: Manual calculation |
| Malawi | Kondwani Jambo (Malawi-Liverpool-Wellcome Trust Clinical Research programme, Liverpool School of Tropical Medicine)[7] | 2020-02-01 - 2020-02-29 | National | Low | 194 | Multiple groups | All | Probability | Blood donors | 0.3% | Beijing Wantai Biological - IgG IgM IgA - Wantai SARS-CoV-2 Total Ab ELISA,Omega diagnostics - IgG - COVID-19 IgG ELISA | 0.928, 1 | Complex test algorithm: Manual calculation |
| Malawi | Kondwani Jambo (Malawi-Liverpool-Wellcome Trust Clinical Research programme, Liverpool School of Tropical Medicine)[7] | 2020-03-01 - 2020-03-31 | National | Low | 199 | Multiple groups | All | Probability | Blood donors | 0.5% | Beijing Wantai Biological - IgG IgM IgA - Wantai SARS-CoV-2 Total Ab ELISA,Omega diagnostics - IgG - COVID-19 IgG ELISA | 0.928, 1 | Complex test algorithm: Manual calculation |
| Nigeria - HRP | Olatunji Kolawole (Ministerial Expert Advisory Committee on COVID-19- Health Sector Response)[9] | 2021-06-29 - 2021-08-31 | National | Moderate | 4,904 | Multiple groups | All | Probability | Residual sera | 78.9% (95% CI 77.7-80%) | Beijing Wantai Biological - IgG IgM IgA - Wantai SARS-CoV-2 Total Ab ELISA | 0.967, 0.975 | Validated by manufacturer |
| Senegal | Cheikh Talla (Institut Pasteur de Dakar)[10] | 2020-10-25 - 2020-11-26 | National | Low | 1,422 | Multiple groups | All | Probability | Household and community samples | 27.9% | Omega diagnostics - IgG - COVID-19 IgG ELISA,ID.Vet - IgG - ID Screen,Beijing Wantai Biological - IgG, IgM, IgA - Wantai SARS-CoV-2 Total Ab ELISA | N/A | Complex test algorithm: Manual review |
| Sierra Leone - HRP | Mohamed Bailor Barrie (Harvard Medical School)[11] | 2021-03-01 - 2021-03-31 | National | Low | 1,893 | Multiple groups | All | Probability | Household and community samples | 2.8% | Hangzhou Biotest Biotech Co. Ltd - IgG, IgM - RightSign COVID-19 IgG/IgM Rapid Test Cassette | 0.914, 1 | Validated by manufacturer |
| South Africa | Nicole Wolter (National Health Laboratory Service)[12] | 2021-04-01 - 2021-04-30 | National | Moderate | 1,849 | Multiple groups | All | Probability | Multiple general populations | 47.2% | Beijing Wantai Biological - IgG, IgM, IgA - Wantai SARS-CoV-2 Total Ab ELISA,Roche Diagnostics - IgG, IgM, IgA - Elecsys® Anti‐SARS‐CoV‐2 (N) | 1, 0.97 | Complex test algorithm: Manual calculation |
| South Africa | Marion Vermeulen (South African National Blood Service)[13] | 2021-01-15 - 2021-05-15 | National | Moderate | 16,762 | Multiple groups | All | Sequential | Blood donors | 47.4% | Roche Diagnostics - IgG, IgM, IgA - Elecsys® Anti‐SARS‐CoV‐2 (N) | 1, 0.995 | Validated by manufacturer |
| Togo - HRP | Didier K Ekouevi (Université de Lomé)[14] | 2021-05-05 - 2021-06-30 | National | Low | 7,593 | Multiple groups | All | Probability | Household and community samples | 61.8% | Beijing Wantai Biological - IgG IgM IgA - Wantai SARS-CoV-2 Total Ab ELISA | 0.967, 0.975 | Validated by manufacturer |
| Uganda - HRP | Dr. Henry Kyobe Bosa (Ministry of Health/ UPDF)[15] | 2021-03-14 - 2021-03-28 | National | Moderate | 7,029 | Multiple groups | All | Probability | Household and community samples | 15.6% (95% CI 14.7-16.4%) | Hangzhou Biotest Biotech Co. Ltd - IgG IgM - RightSign COVID-19 IgG/IgM Rapid Test Cassette | 0.914, 1 | Validated by manufacturer |
| Zambia - HRP | Jonas Z Hines (US Centers for Disease Control and Prevention)[16] | 2020-07-02 - 2020-07-31 | National | Moderate | 1,657 | Multiple groups | All | Convenience | Residual sera | 5.3% | EUROIMMUN - IgG - Anti-SARS-CoV-2 ELISA IgG | 0.9, 1 | Validated by manufacturer |
| Zambia - HRP | Lloyd B Mulenga (Zambia Ministry of Health)[17] | 2020-07-04 - 2020-07-27 | National | Low | 2,704 | Multiple groups | All | Probability | Household and community samples | 2.1% | EUROIMMUN - IgG - Anti-SARS-CoV-2 ELISA IgG | 0.9, 1 | Validated by manufacturer |

###### Americas region

| Country (Location) | Author (Organization) | Sampling Dates (YMD) | Geographic scope | Overall risk of bias | Sample size | Age | Sex | Sampling method | Sample frame | Seroprevalence (95% CI) | Test Manufacturers - Isotypes - Names | Test Sens, Spec | Sens, Spec Source |
| --- | --- | --- | --- | --- | --- | --- | --- | --- | --- | --- | --- | --- | --- |
| Brazil - HRP | Fernando C Barros (Universidade Federal de Pelotas)[18] | 2020-05-14 - 2020-06-23 | National | Moderate | 89,362 | Multiple groups | All | Probability | Household and community samples | 2.3% | Guangzhou Wondfo Biotech Co. Ltd - IgG, IgM, IgA - Wondfo SARS-CoV-2 Antibody Test | 0.864, 0.996 | Validated by manufacturer |
| Colombia - HRP | Marcela Mercado-Reye (Instituto Nacional de Salud)[19] | 2020-09-21 - 2020-12-11 | National | Moderate | 17,863 | Multiple groups | All | Probability | Household and community samples | 32.5% | Siemens - IgG, IgM, IgA - ADVIA Centaur Immunoassay System | 0.86, 0.99 | Validated by independent authors/third party/non-developers |
| Dominican Republic - HRP | Robert Paulino Ramirez (Universidad Iberoamericana)[20] | 2020-04-15 - 2020-06-15 | National | Moderate | 12,897 | Multiple groups | All | Probability | Household and community samples | 5.5% | GenBody Inc. - IgG, IgM - GenBody COVID-19 IgM/IgG | 0.893, 0.959 | Validated by manufacturer |
| Honduras | Mario Rene Mejia Nunez (Colegio Médico)[21] | 2020-06-16 - 2020-06-23 | National | Moderate | 792 | Multiple groups | All | Probability | Household and community samples | 6.2% | NA - - Not reported/ Unable to specify | 0.934, 0.977 | Validated by manufacturers |
| Mexico - HRP | Miguel Fernández-Rojas (Salud Digna A.C.)[22] | 2020-07-05 - 2020-12-31 | National | Moderate | 522,690 | Multiple groups | All | Sequential | Residual sera | 32.8% | Roche Diagnostics - IgG, IgM, IgA - Elecsys® Anti‐SARS‐CoV‐2 (Type Unknown) | 0.995, 0.998 | Validated by manufacturers |
| Mexico - HRP | Francisco Canto-Osorio (Instituto Nacional de Salud Públic)[23] | 2020-08-15 - 2020-11-15 | National | Low | 944 | Children and Youth (0-17 years) | All | Probability | Household and community samples | 18.7% | Roche Diagnostics - IgG, IgM, IgA - Elecsys® Anti‐SARS‐CoV‐2 (N) | 1, 0.995 | Validated by manufacturer |
| Mexico - HRP | Francisco Canto-Osorio (Instituto Nacional de Salud Públic)[23] | 2020-08-15 - 2020-11-15 | National | Low | 858 | Children and Youth (0-17 years) | All | Probability | Household and community samples | 26.7% | Roche Diagnostics - IgG, IgM, IgA - Elecsys® Anti‐SARS‐CoV‐2 (N) | 1, 0.995 | Validated by manufacturer |
| Mexico - HRP | Ana Basto-Abreu (National Institute of Public Health)[24] | 2020-08-15 - 2020-11-15 | National | Moderate | 9,463 | Multiple groups | All | Probability | Household and community samples | 24.9% | Roche Diagnostics - IgG, IgM, IgA - Elecsys® Anti‐SARS‐CoV‐2 (N) | 1, 0.995 | Validated by manufacturer |
| Canada | Canadian Blood Service (Canadian Blood Services )[25] | 2022-04-08 - 2022-04-15 | National | Moderate | 7,908 | Multiple groups | All | Sequential | Blood donors | 35.3% | Roche Diagnostics - IgG, IgM, IgA - Elecsys® Anti‐SARS‐CoV‐2 (N) | 1, 0.995 | Validated by manufacturer |
| Canada | Canadian Blood Services (Canadian Blood Services )[26] | 2022-01-01 - 2022-01-31 | National | Moderate | 32,505 | Multiple groups | All | Sequential | Blood donors | 97.7% | Roche Diagnostics - IgG, IgM, IgA - Elecsys® Anti‐SARS‐CoV‐2 (S) | 0.988, 1 | Validated by manufacturer |
| Canada | Canadian Blood Services (Canadian Blood Services )[27] | 2021-12-14 - 2021-12-30 | National | Moderate | 16,816 | Multiple groups | All | Sequential | Blood donors | 97.3% | Roche Diagnostics - IgG, IgM, IgA - Elecsys® Anti‐SARS‐CoV‐2 (S) | 0.988, 1 | Validated by manufacturer |
| Canada | Canadian Blood Services (Canadian Blood Services )[28] | 2021-11-13 - 2021-11-24 | National | Low | 9,018 | Multiple groups | All | Probability | Blood donors | 97.3% | Roche Diagnostics - IgG, IgM, IgA - Elecsys® Anti‐SARS‐CoV‐2 (S) | 0.988, 1 | Validated by manufacturer |
| Canada | Canadian Blood Services (Canadian Blood Services )[29] | 2021-10-14 - 2021-10-23 | National | Low | 9,627 | Multiple groups | All | Probability | Blood donors | 96.6% | Roche Diagnostics - IgG, IgM, IgA - Elecsys® Anti‐SARS‐CoV‐2 (S) | 0.988, 1 | Validated by manufacturer |
| Canada | Canadian Blood Services (Canadian Blood Services )[30] | 2021-09-14 - 2021-09-24 | National | Low | 9,363 | Multiple groups | All | Probability | Blood donors | 95.9% | Roche Diagnostics - IgG, IgM, IgA - Elecsys® Anti‐SARS‐CoV‐2 (S) | 0.988, 1 | Validated by manufacturer |
| Canada | Canadian Blood Services (Canadian Blood Services )[31] | 2021-08-15 - 2021-08-26 | National | Low | 9,109 | Multiple groups | All | Probability | Blood donors | 95% | Roche Diagnostics - IgG, IgM, IgA - Elecsys® Anti‐SARS‐CoV‐2 (S) | 0.988, 1 | Validated by manufacturer |
| Canada | Canadian Blood Services (Canadian Blood Services )[32] | 2021-07-14 - 2021-07-23 | National | Low | 8,457 | Multiple groups | All | Probability | Blood donors | 93.6% | Roche Diagnostics - IgG, IgM, IgA - Elecsys® Anti‐SARS‐CoV‐2 (S) | 0.988, 1 | Validated by manufacturer |
| Canada | Canadian Blood Services (Canadian Blood Services)[33] | 2021-06-14 - 2021-06-29 | National | Moderate | 16,884 | Multiple groups | All | Sequential | Blood donors | 89.7% | Roche Diagnostics - IgG, IgM, IgA - Elecsys® Anti‐SARS‐CoV‐2 (S) | 0.988, 1 | Validated by manufacturer |
| Canada | Canadian Blood Services (Canadian Blood Services)[34] | 2021-05-08 - 2021-06-04 | National | Moderate | 17,001 | Multiple groups | All | Sequential | Blood donors | 63.3% | Roche Diagnostics - IgG, IgM, IgA - Elecsys® Anti‐SARS‐CoV‐2 (S) | 0.988, 1 | Validated by manufacturer |
| Canada | Canadian Blood Services (Canadian Blood Services)[35] | 2021-04-13 - 2021-04-30 | National | Moderate | 16,931 | Multiple groups | All | Sequential | Blood donors | 26.9% | Roche Diagnostics - IgG, IgM, IgA - Elecsys® Anti‐SARS‐CoV‐2 (S) | 0.988, 1 | Validated by manufacturer |
| Canada | Canadian Blood Services (Canadian Blood Services)[36] | 2021-02-27 - 2021-03-13 | National | Moderate | 16,873 | Multiple groups | All | Sequential | Blood donors | 10.1% | Roche Diagnostics - IgG, IgM, IgA - Elecsys® Anti‐SARS‐CoV‐2 (S) | 0.988, 1 | Validated by manufacturer |
| Canada | Canadian Blood Services (Canadian Blood Services)[37] | 2021-01-01 - 2021-01-27 | National | Moderate | 33,400 | Multiple groups | All | Sequential | Blood donors | 3.2% | Roche Diagnostics - IgG, IgM, IgA - Elecsys® Anti‐SARS‐CoV‐2 (S) | 0.988, 1 | Validated by manufacturer |
| Canada | Canadian Blood Services (Canadian Blood Services)[38] | 2020-12-10 - 2020-12-23 | National | Moderate | 16,961 | Multiple groups | All | Sequential | Blood donors | 1.4% | Abbott Laboratories - IgG - Abbott Architect SARS-CoV-2 IgG | 0.99, 1 | Validated by manufacturer |
| Canada | Canadian Blood Services (Canadian Blood Services)[38] | 2020-11-07 - 2020-11-25 | National | Moderate | 17,049 | Multiple groups | All | Sequential | Blood donors | 1.5% | Abbott Laboratories - IgG - Abbott Architect SARS-CoV-2 IgG | 0.99, 1 | Validated by manufacturer |
| Canada | Canadian Blood Services (Canadian Blood Services)[38] | 2020-10-12 - 2020-10-31 | National | Moderate | 16,811 | Multiple groups | All | Sequential | Blood donors | 0.9% | Abbott Laboratories - IgG - Abbott Architect SARS-CoV-2 IgG | 0.99, 1 | Validated by manufacturer |
| Canada | Sahar Saeed (Canadian Blood Services)[39] | 2020-05-09 - 2020-07-21 | National | Moderate | 74,642 | Multiple groups | All | Probability | Blood donors | 0.7% | Abbott Laboratories - IgG - Abbott Architect SARS-CoV-2 IgG | 0.99, 1 | Validated by manufacturer |
| Chile - HRP | Pablo Vial (Universidad del Desarrollo)[40] | 2020-09-25 - 2020-11-25 | National | Low | 2,493 | Multiple groups | All | Probability | Household and community samples | 10.4% | Roche Diagnostics - IgG, IgM, IgA - Elecsys® Anti‐SARS‐CoV‐2 (S),Zhuhai Livzon Diagnostics Inc - IgG, IgM - 2019-nCoV IgG/IgM Antibody Detection Kit | 0.999, 0.992 | Complex test algorithm: Manual calculation |
| United States of America | Roger Dodd (American Red Cross)[41] | 2021-06-18 - 2021-06-25 | National | Moderate | 45,966 | Multiple groups | All | Convenience | Blood donors | 90.1% | Ortho Clinical Diagnostics Inc. - IgG, IgM, IgA - VITROS Immunodiagnostic Products Anti-SARS-CoV-2 Total | 1, 1 | Validated by manufacturer |
| United States of America | Jefferson Jones (Vitalant Research Institute)[42] | 2021-05-01 - 2021-05-31 | National | Moderate | 134,949 | Multiple groups | All | Probability | Blood donors | 83.3% | Ortho Clinical Diagnostics Inc. - IgG, IgM, IgA - VITROS Immunodiagnostic Products Anti-SARS-CoV-2 Total,Roche Diagnostics - IgG, IgM, IgA - Elecsys® Anti‐SARS‐CoV‐2 (N) | 1, 0.995 | Complex test algorithm: Manual calculation |
| United States of America | Jefferson Jones (Vitalant Research Institute)[42] | 2021-04-01 - 2021-04-30 | National | Moderate | 134,510 | Multiple groups | All | Probability | Blood donors | 71.4% | Ortho Clinical Diagnostics Inc. - IgG, IgM, IgA - VITROS Immunodiagnostic Products Anti-SARS-CoV-2 Total,Roche Diagnostics - IgG, IgM, IgA - Elecsys® Anti‐SARS‐CoV‐2 (N) | 1, 0.995 | Complex test algorithm: Manual calculation |
| United States of America | Jefferson Jones (Vitalant Research Institute)[42] | 2021-03-01 - 2021-03-31 | National | Moderate | 134,557 | Multiple groups | All | Probability | Blood donors | 49.2% | Ortho Clinical Diagnostics Inc. - IgG, IgM, IgA - VITROS Immunodiagnostic Products Anti-SARS-CoV-2 Total,Roche Diagnostics - IgG, IgM, IgA - Elecsys® Anti‐SARS‐CoV‐2 (N) | 1, 0.995 | Complex test algorithm: Manual calculation |
| United States of America | Jefferson Jones (Vitalant Research Institute)[42] | 2021-02-01 - 2021-02-28 | National | Moderate | 133,288 | Multiple groups | All | Probability | Blood donors | 33.5% | Ortho Clinical Diagnostics Inc. - IgG, IgM, IgA - VITROS Immunodiagnostic Products Anti-SARS-CoV-2 Total,Roche Diagnostics - IgG, IgM, IgA - Elecsys® Anti‐SARS‐CoV‐2 (N) | 1, 0.995 | Complex test algorithm: Manual calculation |
| United States of America | Jefferson Jones (Vitalant Research Institute)[42] | 2021-01-01 - 2021-01-31 | National | Moderate | 134,632 | Multiple groups | All | Probability | Blood donors | 20.5% | Ortho Clinical Diagnostics Inc. - IgG, IgM, IgA - VITROS Immunodiagnostic Products Anti-SARS-CoV-2 Total,Roche Diagnostics - IgG, IgM, IgA - Elecsys® Anti‐SARS‐CoV‐2 (N) | 1, 0.995 | Complex test algorithm: Manual calculation |
| United States of America | Patrick Sullivan (Emory University)[43] | 2020-11-01 - 2020-12-08 | National | Low | 2,241 | Multiple groups | All | Probability | Household and community samples | 7.2% | Bio-rad - IgG, IgM, IgA - Platelia SARS-CoV-2 Total Ab assay | 0.993, 0.98 | Validated by manufacturer |
| United States of America | Shuchi Anand (Stanford University)[44] | 2020-07-01 - 2020-07-31 | National | Low | 28,503 | Multiple groups | All | Probability | Residual sera | 9.3% | Siemens - IgG, IgM, IgA - ADVIA Centaur Immunoassay System | 1, 0.998 | Validated by manufacturers |

###### Eastern Mediterranean region

| Country (Location) | Author (Organization) | Sampling Dates (YMD) | Geographic scope | Overall risk of bias | Sample size | Age | Sex | Sampling method | Sample frame | Seroprevalence (95% CI) | Test Manufacturers - Isotypes - Names | Test Sens, Spec | Sens, Spec Source |
| --- | --- | --- | --- | --- | --- | --- | --- | --- | --- | --- | --- | --- | --- |
| Iran (Islamic Republic of) - HRP | Kazem Khalagi (Tehran University of Medical Sciences)[45] | 2020-08-03 - 2020-10-31 | National | Low | 11,256 | Multiple groups | All | Probability | Household and community samples | 11.8% | Pishtaz Diagnostics Iran - IgG, IgM - ELISA kit Pishtaz Teb Diagnostics | 0.941, 0.983 | Validated by manufacturer |
| Iran (Islamic Republic of) - HRP | Hossein Poustchi (Tehran University of Medical Sciences)[46] | 2020-04-17 - 2020-06-02 | National | Low | 3,530 | Multiple groups | All | Probability | Household and community samples | 12.9% | Pishtaz Diagnostics Iran - IgG, IgM - ELISA kit Pishtaz Teb Diagnostics | 0.941, 0.983 | Validated by manufacturer |
| Jordan - HRP | Saverio Bellizzi (World Health Organization)[47] | 2020-12-27 - 2021-01-06 | National | Moderate | 5,044 | Multiple groups | All | Probability | Household and community samples | 34.2% | Beijing Wantai Biological - IgM - Wantai SARS-CoV-2 IgM ELISA | 0.868, 1 | Validated by manufacturer |
| Jordan - HRP | Saverio Bellizzi (World Health Organization)[47] | 2020-10-01 - 2020-10-31 | National | Moderate | 5,470 | Multiple groups | All | Probability | Household and community samples | 7% | Beijing Wantai Biological - IgM - Wantai SARS-CoV-2 IgM ELISA | 0.868, 1 | Validated by manufacturer |
| Jordan - HRP | Saverio Bellizzi (World Health Organization)[47] | 2020-08-01 - 2020-08-31 | National | Moderate | 4,704 | Multiple groups | All | Probability | Household and community samples | 0.3% | Beijing Wantai Biological - IgM - Wantai SARS-CoV-2 IgM ELISA | 0.868, 1 | Validated by manufacturer |
| Lebanon - HRP | Dr Alissar Rady (WHO)[@] | 2021-02-15 - 2021-03-22 | National | Low | 1,252 | Multiple groups | All | Probability | Household and community samples | 39.2% | Beijing Wantai Biological - IgG IgM IgA - Wantai SARS-CoV-2 Total Ab ELISA | 0.967, 0.975 | Validated by manufacturer |
| Lebanon - HRP | Dr Alissar Rady (WHO)[@] | 2021-02-15 - 2021-03-22 | National | Moderate | 755 | Multiple groups | All | Probability | Household and community samples | 46.7% | Beijing Wantai Biological - IgG IgM IgA - Wantai SARS-CoV-2 Total Ab ELISA | 0.967, 0.975 | Validated by manufacturer |
| Lebanon - HRP | Abbas Hoballah (Lebanese University)[48] | 2020-12-07 - 2021-01-15 | National | Low | 2,058 | Multiple groups | All | Probability | Household and community samples | 15.9% | Technogenetics - IgG, IgM - nCOVID-19 IgG & IgM POCT | 0.946, 0.993 | Validated by manufacturer |
| occupied Palestinian territory, including east Jerusalem - HRP | Sharif E. Qaddomi (PNIPH)[@] | 2020-12-01 - 2020-12-31 | National | Moderate | 6,154 | Multiple groups | All | Probability | Household and community samples | 39.7% (95% CI 38.5-40.9%) | Roche Diagnostics - IgG IgM IgA - Elecsys® Anti‐SARS‐CoV‐2 (S) | 0.988, 1 | Validated by manufacturer |
| Oman | Seif Al-Abri (Ministry of Health (Oman))[49] | 2020-11-08 - 2020-11-19 | National | Moderate | 4,064 | Multiple groups | All | Probability | Household and community samples | 22% | DiaSorin - IgG - Liaison SARS-CoV-2 S1/S2 IgG | 0.974, 0.985 | Validated by manufacturer |
| Oman | Seif Al-Abri (Ministry of Health (Oman))[49] | 2020-09-13 - 2020-09-24 | National | Moderate | 4,780 | Multiple groups | All | Probability | Household and community samples | 16.4% | DiaSorin - IgG - Liaison SARS-CoV-2 S1/S2 IgG | 0.974, 0.985 | Validated by manufacturer |
| Oman | Seif Al-Abri (Ministry of Health (Oman))[49] | 2020-08-16 - 2020-08-27 | National | Moderate | 4,403 | Multiple groups | All | Probability | Household and community samples | 9.7% | DiaSorin - IgG - Liaison SARS-CoV-2 S1/S2 IgG | 0.974, 0.985 | Validated by manufacturer |
| Oman | Seif Al-Abri (Ministry of Health (Oman))[49] | 2020-07-12 - 2020-07-23 | National | Moderate | 4,210 | Multiple groups | All | Probability | Household and community samples | 5.5% | DiaSorin - IgG - Liaison SARS-CoV-2 S1/S2 IgG | 0.974, 0.985 | Validated by manufacturer |
| Pakistan - HRP | Ahsan Ahmad (Ministry of National Health Services Regulations and Coordination)[50] | 2020-10-21 - 2020-11-08 | National | Moderate | 4,998 | Multiple groups | All | Probability | Household and community samples | 7% | Bioperfectus - IgG, IgM - Novel Corona Virus (SARS-CoV-2) IgM/IgG Rapid Test Kit | 0.9, 0.965 | Validated by manufacturer |
| Pakistan - HRP | Mohsina Haq (Riphah International University )[51] | 2020-07-15 - 2020-07-31 | National | Moderate | 15,390 | Multiple groups | All | Probability | Household and community samples | 42.4% | Roche Diagnostics - IgG, IgM, IgA - Elecsys® Anti‐SARS‐CoV‐2 (N) | 1, 0.995 | Validated by manufacturer |
| Qatar | Peter Coyle (Cornell University)[52] | 2020-10-10 - 2020-10-21 | National | Moderate | 709 | Multiple groups | All | Convenience | Residual sera | 71.9% | Roche Diagnostics - IgG, IgM, IgA - Elecsys® Anti‐SARS‐CoV‐2 (N) | 1, 0.995 | Validated by manufacturer |
| Qatar | Peter Coyle (Cornell University)[53] | 2020-05-12 - 2020-09-09 | National | Moderate | 112,941 | Multiple groups | All | Sequential | Residual sera | 13.3% | Roche Diagnostics - IgG, IgM, IgA - Elecsys® Anti‐SARS‐CoV‐2 (N) | 1, 0.995 | Validated by manufacturer |

###### Europe region

| Country (Location) | Author (Organization) | Sampling Dates (YMD) | Geographic scope | Overall risk of bias | Sample size | Age | Sex | Sampling method | Sample frame | Seroprevalence (95% CI) | Test Manufacturers - Isotypes - Names | Test Sens, Spec | Sens, Spec Source |
| --- | --- | --- | --- | --- | --- | --- | --- | --- | --- | --- | --- | --- | --- |
| Kazakhstan | Manar Smangul (Ministry of Health of the Republic of Kazakhstan)[54] | 2020-10-24 - 2021-01-11 | National | Moderate | 6,891 | Multiple groups | All | Probability | Household and community samples | 57% (95% CI 55.8-58.2%) | Beijing Wantai Biological - IgG IgM IgA - Wantai SARS-CoV-2 Total Ab ELISA | 0.967, 0.975 | Validated by manufacturer |
| Kyrgyzstan | Anna Popova (Saint Petersburg Pasteur Institute)[55] | 2021-06-28 - 2021-07-03 | National | Moderate | 9,471 | Multiple groups | All | Probability | Household and community samples | 48.7% | NA - - Author designed (ELISA) - Nucleocapsid | NA, NA | Validated by independent authors/third party/non-developers |
| Kyrgyzstan | Nurmatov Zuridin, Kuchuk Tatyana (Scientific Production Association “Preventive medicine”)[56] | 2020-07-04 - 2020-08-12 | National | Moderate | 4,540 | Multiple groups | All | Convenience | Household and community samples | 32.5% (95% CI 31.1-33.9%) | Beijing Wantai Biological - IgG IgM IgA - Wantai SARS-CoV-2 Total Ab ELISA | 0.967, 0.975 | Validated by manufacturer |
| Republic of Moldova | Alexei Ceban Stela Gheorghita, WHO CO (National Agency for Public Health)[57] | 2020-11-17 - 2021-01-15 | National | Moderate | 5,656 | Multiple groups | All | Probability | Residual sera | 48.4% (95% CI 47.1-49.7%) | Beijing Wantai Biological - IgG IgM IgA - Wantai SARS-CoV-2 Total Ab ELISA | 0.967, 0.975 | Validated by manufacturer |
| Uzbekistan | R A Rakhimov (Research Institute of Virology)[58] | 2020-08-27 - 2020-09-11 | National | Moderate | 86,879 | Multiple groups | All | Convenience | Residual sera | 23.1% | Beijing Wantai Biological - IgG, IgM, IgA - Wantai Rapid Test for Total Antibody to SARS-CoV-2 | 0.947, 0.989 | Validated by manufacturer |
| Andorra | Cristina Royo Cebrecos (ISGlobal)[59] | 2020-05-04 - 2020-05-28 | National | Moderate | 72,964 | Multiple groups | All | Probability | Household and community samples | 11% | Zhuhai Livzon Diagnostics Inc - IgG, IgM - 2019-nCoV IgG/IgM Antibody Detection Kit | 0.906, 0.992 | Validated by manufacturer |
| Belgium | Sereina Herzog (University of Antwerp)[60] | 2020-10-12 - 2020-10-17 | National | Low | 2,966 | Multiple groups | All | Probability | Residual sera | 4.2% | EUROIMMUN - IgG - Anti-SARS-CoV-2 ELISA IgG | 0.9, 1 | Validated by manufacturer |
| Belgium | Sereina Herzog (University of Antwerp)[60] | 2020-09-07 - 2020-09-12 | National | Low | 3,047 | Multiple groups | All | Probability | Residual sera | 3.3% | EUROIMMUN - IgG - Anti-SARS-CoV-2 ELISA IgG | 0.9, 1 | Validated by manufacturer |
| Belgium | Sereina Herzog (University of Antwerp)[60] | 2020-06-29 - 2020-07-04 | National | Low | 3,023 | Multiple groups | All | Probability | Residual sera | 3.7% | EUROIMMUN - IgG - Anti-SARS-CoV-2 ELISA IgG | 0.9, 1 | Validated by manufacturer |
| Belgium | Sereina Herzog (University of Antwerp)[60] | 2020-06-08 - 2020-06-13 | National | Low | 2,960 | Multiple groups | All | Probability | Residual sera | 4.7% | EUROIMMUN - IgG - Anti-SARS-CoV-2 ELISA IgG | 0.9, 1 | Validated by manufacturer |
| Belgium | Sereina Herzog (University of Antwerp)[60] | 2020-05-18 - 2020-05-25 | National | Low | 3,242 | Multiple groups | All | Probability | Residual sera | 6.2% | EUROIMMUN - IgG - Anti-SARS-CoV-2 ELISA IgG | 0.9, 1 | Validated by manufacturer |
| Belgium | Sereina Herzog (University of Antwerp)[60] | 2020-04-20 - 2020-04-26 | National | Low | 3,397 | Multiple groups | All | Probability | Residual sera | 5.2% | EUROIMMUN - IgG - Anti-SARS-CoV-2 ELISA IgG | 0.9, 1 | Validated by manufacturer |
| Belgium | Sereina Herzog (University of Antwerp)[60] | 2020-03-30 - 2020-04-05 | National | Low | 3,910 | Multiple groups | All | Probability | Residual sera | 1.8% | EUROIMMUN - IgG - Anti-SARS-CoV-2 ELISA IgG | 0.9, 1 | Validated by manufacturer |
| Denmark | Kamille Fogh (Copenhagen University Hospital)[61] | 2021-01-08 - 2021-01-31 | National | Moderate | 22,677 | Multiple groups | All | Probability | Blood donors | 5.8% | Beijing Wantai Biological - IgG, IgM, IgA - Wantai SARS-CoV-2 Total Ab ELISA | 0.967, 0.975 | Validated by manufacturer |
| Denmark | Laura Espenhain (Statens Serum Institut)[62] | 2020-12-01 - 2020-12-31 | National | Low | 4,044 | Multiple groups | All | Probability | Household and community samples | 4.1% | Beijing Wantai Biological - IgG, IgM, IgA - Wantai SARS-CoV-2 Total Ab ELISA | 0.967, 0.975 | Validated by manufacturer |
| Denmark | Laura Espenhain (Statens Serum Institut)[62] | 2020-10-01 - 2020-11-30 | National | Low | 9,654 | Multiple groups | All | Probability | Household and community samples | 2.2% | Beijing Wantai Biological - IgG, IgM, IgA - Wantai SARS-CoV-2 Total Ab ELISA | 0.967, 0.975 | Validated by manufacturer |
| Denmark | Kamille Fogh (Copenhagen University Hospital)[63] | 2020-10-02 - 2020-10-11 | National | Moderate | 318,552 | Multiple groups | All | Probability | Household and community samples | 0.8% | Zhuhai Livzon Diagnostics Inc - IgG, IgM - 2019-nCoV IgG/IgM Antibody Detection Kit | 0.906, 0.992 | Validated by manufacturer |
| Denmark | Laura Espenhain (Statens Serum Institut)[62] | 2020-08-15 - 2020-09-30 | National | Low | 11,478 | Multiple groups | All | Probability | Household and community samples | 2.1% | Beijing Wantai Biological - IgG, IgM, IgA - Wantai SARS-CoV-2 Total Ab ELISA | 0.967, 0.975 | Validated by manufacturer |
| Denmark | Ole Birger Pedersen (Copenhagen University Hospital Rigshospitalet)[64] | 2020-06-02 - 2020-06-19 | National | Low | 1,200 | Seniors (65+ years) | All | Probability | Blood donors | 1.8% | Beijing Wantai Biological - IgG, IgM, IgA - Wantai SARS-CoV-2 Total Ab ELISA | 0.967, 0.975 | Validated by manufacturer |
| Denmark | Laura Espenhain (Statens Serum Institut)[62] | 2020-05-05 - 2020-05-15 | National | Low | 2,512 | Multiple groups | All | Probability | Household and community samples | 1.2% | Beijing Wantai Biological - IgG, IgM, IgA - Wantai SARS-CoV-2 Total Ab ELISA | 0.967, 0.975 | Validated by manufacturer |
| Denmark | Christian Erikstrup (Copenhagen University Hospital)[65] | 2020-04-06 - 2020-05-03 | National | Moderate | 20,640 | Adults (18-64 years) | All | Probability | Blood donors | 2% | Zhuhai Livzon Diagnostics Inc - IgG, IgM - 2019-nCoV IgG/IgM Antibody Detection Kit | 0.906, 0.992 | Validated by manufacturer |
| Finland | Finnish institute for health and welfare (Finnish Institute for Health and Welfare)[66] | 2021-12-19 - 2021-12-25 | National | Moderate | 2 | Adults (18-64 years) | All | Probability | Household and community samples | 50% | NA - IgG, IgM - Author designed (IFA) - MULTIPLEXED | 1, 1 | Validated by developers |
| Finland | Finnish institute for health and welfare (Finnish Institute for Health and Welfare)[66] | 2021-12-12 - 2021-12-18 | National | Moderate | 1 | Adults (18-64 years) | All | Probability | Household and community samples | 100% | NA - IgG, IgM - Author designed (IFA) - MULTIPLEXED | 1, 1 | Validated by developers |
| Finland | Finnish institute for health and welfare (Finnish Institute for Health and Welfare)[66] | 2021-12-05 - 2021-12-11 | National | Moderate | 4 | Adults (18-64 years) | All | Probability | Household and community samples | 100% | NA - IgG, IgM - Author designed (IFA) - MULTIPLEXED | 1, 1 | Validated by developers |
| Finland | Finnish institute for health and welfare (Finnish Institute for Health and Welfare)[66] | 2021-11-28 - 2021-12-04 | National | Moderate | 4 | Adults (18-64 years) | All | Probability | Household and community samples | 75% | NA - IgG, IgM - Author designed (IFA) - MULTIPLEXED | 1, 1 | Validated by developers |
| Finland | Finnish institute for health and welfare (Finnish Institute for Health and Welfare)[66] | 2021-11-21 - 2021-11-27 | National | Moderate | 8 | Adults (18-64 years) | All | Probability | Household and community samples | 87.5% | NA - IgG, IgM - Author designed (IFA) - MULTIPLEXED | 1, 1 | Validated by developers |
| Finland | Finnish institute for health and welfare (Finnish Institute for Health and Welfare)[66] | 2021-11-14 - 2021-11-20 | National | Moderate | 13 | Adults (18-64 years) | All | Probability | Household and community samples | 92.3% | NA - IgG, IgM - Author designed (IFA) - MULTIPLEXED | 1, 1 | Validated by developers |
| Finland | Finnish institute for health and welfare (Finnish Institute for Health and Welfare)[66] | 2021-11-07 - 2021-11-13 | National | Moderate | 30 | Adults (18-64 years) | All | Probability | Household and community samples | 93.3% | NA - IgG, IgM - Author designed (IFA) - MULTIPLEXED | 1, 1 | Validated by developers |
| Finland | Finnish institute for health and welfare (Finnish Institute for Health and Welfare)[66] | 2021-10-31 - 2021-11-06 | National | Moderate | 79 | Adults (18-64 years) | All | Probability | Household and community samples | 91.1% | NA - IgG, IgM - Author designed (IFA) - MULTIPLEXED | 1, 1 | Validated by developers |
| Finland | Finnish institute for health and welfare (Finnish Institute for Health and Welfare)[66] | 2021-10-24 - 2021-10-30 | National | Moderate | 11 | Adults (18-64 years) | All | Probability | Household and community samples | 90.9% | NA - IgG, IgM - Author designed (IFA) - MULTIPLEXED | 1, 1 | Validated by developers |
| Finland | Finnish institute for health and welfare (Finnish Institute for Health and Welfare)[66] | 2021-10-17 - 2021-10-23 | National | Moderate | 1 | Adults (18-64 years) | All | Probability | Household and community samples | 0% | NA - IgG, IgM - Author designed (IFA) - MULTIPLEXED | 1, 1 | Validated by developers |
| Finland | Finnish institute for health and welfare (Finnish Institute for Health and Welfare)[66] | 2021-09-26 - 2021-10-02 | National | Moderate | 2 | Adults (18-64 years) | All | Probability | Household and community samples | 50% | NA - IgG, IgM - Author designed (IFA) - MULTIPLEXED | 1, 1 | Validated by developers |
| Finland | Finnish institute for health and welfare (Finnish Institute for Health and Welfare)[66] | 2021-09-12 - 2021-09-18 | National | Moderate | 1 | Adults (18-64 years) | All | Probability | Household and community samples | 100% | NA - IgG, IgM - Author designed (IFA) - MULTIPLEXED | 1, 1 | Validated by developers |
| Finland | Finnish institute for health and welfare (Finnish Institute for Health and Welfare)[66] | 2021-09-05 - 2021-09-11 | National | Moderate | 1 | Adults (18-64 years) | All | Probability | Household and community samples | 100% | NA - IgG, IgM - Author designed (IFA) - MULTIPLEXED | 1, 1 | Validated by developers |
| Finland | Finnish institute for health and welfare (Finnish Institute for Health and Welfare)[66] | 2021-08-29 - 2021-09-04 | National | Moderate | 1 | Adults (18-64 years) | All | Probability | Household and community samples | 0% | NA - IgG, IgM - Author designed (IFA) - MULTIPLEXED | 1, 1 | Validated by developers |
| Finland | Finnish institute for health and welfare (Finnish Institute for Health and Welfare)[66] | 2021-08-22 - 2021-08-28 | National | Moderate | 8 | Adults (18-64 years) | All | Probability | Household and community samples | 75% | NA - IgG, IgM - Author designed (IFA) - MULTIPLEXED | 1, 1 | Validated by developers |
| Finland | Finnish institute for health and welfare (Finnish Institute for Health and Welfare)[66] | 2021-08-15 - 2021-08-21 | National | Moderate | 7 | Adults (18-64 years) | All | Probability | Household and community samples | 85.7% | NA - IgG, IgM - Author designed (IFA) - MULTIPLEXED | 1, 1 | Validated by developers |
| Finland | Finnish institute for health and welfare (Finnish Institute for Health and Welfare)[66] | 2021-08-08 - 2021-08-14 | National | Moderate | 25 | Adults (18-64 years) | All | Probability | Household and community samples | 84% | NA - IgG, IgM - Author designed (IFA) - MULTIPLEXED | 1, 1 | Validated by developers |
| Finland | Finnish institute for health and welfare (Finnish Institute for Health and Welfare)[66] | 2021-08-01 - 2021-08-07 | National | Moderate | 42 | Adults (18-64 years) | All | Probability | Household and community samples | 92.9% | NA - IgG, IgM - Author designed (IFA) - MULTIPLEXED | 1, 1 | Validated by developers |
| Finland | Finnish institute for health and welfare (Finnish Institute for Health and Welfare)[66] | 2021-07-25 - 2021-07-31 | National | Moderate | 11 | Adults (18-64 years) | All | Probability | Household and community samples | 81.8% | NA - IgG, IgM - Author designed (IFA) - MULTIPLEXED | 1, 1 | Validated by developers |
| Finland | Finnish institute for health and welfare (Finnish Institute for Health and Welfare)[66] | 2021-07-18 - 2021-07-24 | National | Moderate | 2 | Adults (18-64 years) | All | Probability | Household and community samples | 100% | NA - IgG, IgM - Author designed (IFA) - MULTIPLEXED | 1, 1 | Validated by developers |
| Finland | Finnish institute for health and welfare (Finnish Institute for Health and Welfare)[66] | 2021-07-11 - 2021-07-17 | National | Moderate | 2 | Adults (18-64 years) | All | Probability | Household and community samples | 100% | NA - IgG, IgM - Author designed (IFA) - MULTIPLEXED | 1, 1 | Validated by developers |
| Finland | Finnish institute for health and welfare (Finnish Institute for Health and Welfare)[66] | 2021-07-04 - 2021-07-10 | National | Moderate | 12 | Adults (18-64 years) | All | Probability | Household and community samples | 58.3% | NA - IgG, IgM - Author designed (IFA) - MULTIPLEXED | 1, 1 | Validated by developers |
| Finland | Finnish institute for health and welfare (Finnish Institute for Health and Welfare)[66] | 2021-06-27 - 2021-07-03 | National | Moderate | 48 | Adults (18-64 years) | All | Probability | Household and community samples | 68.8% | NA - IgG, IgM - Author designed (IFA) - MULTIPLEXED | 1, 1 | Validated by developers |
| Finland | Finnish institute for health and welfare (Finnish Institute for Health and Welfare)[66] | 2021-06-20 - 2021-06-26 | National | Moderate | 44 | Adults (18-64 years) | All | Probability | Household and community samples | 72.7% | NA - IgG, IgM - Author designed (IFA) - MULTIPLEXED | 1, 1 | Validated by developers |
| Finland | Finnish institute for health and welfare (Finnish Institute for Health and Welfare)[66] | 2021-06-13 - 2021-06-19 | National | Moderate | 47 | Adults (18-64 years) | All | Probability | Household and community samples | 57.5% | NA - IgG, IgM - Author designed (IFA) - MULTIPLEXED | 1, 1 | Validated by developers |
| Finland | Finnish institute for health and welfare (Finnish Institute for Health and Welfare)[66] | 2021-06-06 - 2021-06-12 | National | Moderate | 68 | Adults (18-64 years) | All | Probability | Household and community samples | 60.3% | NA - IgG, IgM - Author designed (IFA) - MULTIPLEXED | 1, 1 | Validated by developers |
| Finland | Finnish institute for health and welfare (Finnish Institute for Health and Welfare)[66] | 2021-05-30 - 2021-06-05 | National | Moderate | 39 | Adults (18-64 years) | All | Probability | Household and community samples | 48.7% | NA - IgG, IgM - Author designed (IFA) - MULTIPLEXED | 1, 1 | Validated by developers |
| Finland | Finnish institute for health and welfare (Finnish Institute for Health and Welfare)[66] | 2021-05-23 - 2021-05-29 | National | Moderate | 93 | Adults (18-64 years) | All | Probability | Household and community samples | 49.5% | NA - IgG, IgM - Author designed (IFA) - MULTIPLEXED | 1, 1 | Validated by developers |
| Finland | Finnish institute for health and welfare (Finnish Institute for Health and Welfare)[66] | 2021-05-16 - 2021-05-22 | National | Moderate | 86 | Adults (18-64 years) | All | Probability | Household and community samples | 44.2% | NA - IgG, IgM - Author designed (IFA) - MULTIPLEXED | 1, 1 | Validated by developers |
| Finland | Finnish institute for health and welfare (Finnish Institute for Health and Welfare)[66] | 2021-05-09 - 2021-05-15 | National | Moderate | 52 | Adults (18-64 years) | All | Probability | Household and community samples | 26.9% | NA - IgG, IgM - Author designed (IFA) - MULTIPLEXED | 1, 1 | Validated by developers |
| Finland | Finnish institute for health and welfare (Finnish Institute for Health and Welfare)[66] | 2021-05-02 - 2021-05-08 | National | Moderate | 74 | Adults (18-64 years) | All | Probability | Household and community samples | 28.4% | NA - IgG, IgM - Author designed (IFA) - MULTIPLEXED | 1, 1 | Validated by developers |
| Finland | Finnish institute for health and welfare (Finnish Institute for Health and Welfare)[66] | 2021-04-25 - 2021-05-01 | National | Moderate | 109 | Adults (18-64 years) | All | Probability | Household and community samples | 22.9% | NA - IgG, IgM - Author designed (IFA) - MULTIPLEXED | 1, 1 | Validated by developers |
| Finland | Finnish institute for health and welfare (Finnish Institute for Health and Welfare)[66] | 2021-04-18 - 2021-04-24 | National | Moderate | 102 | Adults (18-64 years) | All | Probability | Household and community samples | 18.6% | NA - IgG, IgM - Author designed (IFA) - MULTIPLEXED | 1, 1 | Validated by developers |
| Finland | Finnish institute for health and welfare (Finnish Institute for Health and Welfare)[66] | 2021-04-11 - 2021-04-17 | National | Moderate | 29 | Adults (18-64 years) | All | Probability | Household and community samples | 13.8% | NA - IgG, IgM - Author designed (IFA) - MULTIPLEXED | 1, 1 | Validated by developers |
| Finland | Finnish institute for health and welfare (Finnish Institute for Health and Welfare)[66] | 2021-04-04 - 2021-04-10 | National | Moderate | 59 | Adults (18-64 years) | All | Probability | Household and community samples | 6.8% | NA - IgG, IgM - Author designed (IFA) - MULTIPLEXED | 1, 1 | Validated by developers |
| Finland | Finnish institute for health and welfare (Finnish Institute for Health and Welfare)[66] | 2021-03-28 - 2021-04-03 | National | Moderate | 60 | Adults (18-64 years) | All | Probability | Household and community samples | 8.3% | NA - IgG, IgM - Author designed (IFA) - MULTIPLEXED | 1, 1 | Validated by developers |
| Finland | Finnish institute for health and welfare (Finnish Institute for Health and Welfare)[66] | 2021-03-21 - 2021-03-27 | National | Moderate | 73 | Adults (18-64 years) | All | Probability | Household and community samples | 10.6% | NA - IgG, IgM - Author designed (IFA) - MULTIPLEXED | 1, 1 | Validated by developers |
| Finland | Finnish institute for health and welfare (Finnish Institute for Health and Welfare)[66] | 2021-03-14 - 2021-03-20 | National | Moderate | 104 | Adults (18-64 years) | All | Probability | Household and community samples | 6.7% | NA - IgG, IgM - Author designed (IFA) - MULTIPLEXED | 1, 1 | Validated by developers |
| Finland | Finnish institute for health and welfare (Finnish Institute for Health and Welfare)[66] | 2021-03-07 - 2021-03-13 | National | Moderate | 64 | Adults (18-64 years) | All | Probability | Household and community samples | 3.1% | NA - IgG, IgM - Author designed (IFA) - MULTIPLEXED | 1, 1 | Validated by developers |
| Finland | Finnish institute for health and welfare (Finnish Institute for Health and Welfare)[66] | 2021-02-28 - 2021-03-06 | National | Moderate | 132 | Adults (18-64 years) | All | Probability | Household and community samples | 2.3% | NA - IgG, IgM - Author designed (IFA) - MULTIPLEXED | 1, 1 | Validated by developers |
| Finland | Finnish institute for health and welfare (Finnish Institute for Health and Welfare)[66] | 2021-02-21 - 2021-02-27 | National | Moderate | 123 | Adults (18-64 years) | All | Probability | Household and community samples | 6.5% | NA - IgG, IgM - Author designed (IFA) - MULTIPLEXED | 1, 1 | Validated by developers |
| Finland | Finnish institute for health and welfare (Finnish Institute for Health and Welfare)[66] | 2021-02-14 - 2021-02-20 | National | Moderate | 69 | Adults (18-64 years) | All | Probability | Household and community samples | 7.2% | NA - IgG, IgM - Author designed (IFA) - MULTIPLEXED | 1, 1 | Validated by developers |
| Finland | Finnish institute for health and welfare (Finnish Institute for Health and Welfare)[66] | 2021-02-07 - 2021-02-13 | National | Moderate | 111 | Adults (18-64 years) | All | Probability | Household and community samples | 5.4% | NA - IgG, IgM - Author designed (IFA) - MULTIPLEXED | 1, 1 | Validated by developers |
| Finland | Finnish institute for health and welfare (Finnish Institute for Health and Welfare)[66] | 2021-01-31 - 2021-02-06 | National | Moderate | 110 | Adults (18-64 years) | All | Probability | Household and community samples | 2.7% | NA - IgG, IgM - Author designed (IFA) - MULTIPLEXED | 1, 1 | Validated by developers |
| Finland | Finnish institute for health and welfare (Finnish Institute for Health and Welfare)[66] | 2021-01-24 - 2021-01-30 | National | Moderate | 30 | Adults (18-64 years) | All | Probability | Household and community samples | 0% | NA - IgG, IgM - Author designed (IFA) - MULTIPLEXED | 1, 1 | Validated by developers |
| Finland | Finnish institute for health and welfare (Finnish Institute for Health and Welfare)[66] | 2021-01-17 - 2021-01-23 | National | Moderate | 5 | Adults (18-64 years) | All | Probability | Household and community samples | 0% | NA - IgG, IgM - Author designed (IFA) - MULTIPLEXED | 1, 1 | Validated by developers |
| Finland | Finnish institute for health and welfare (Finnish Institute for Health and Welfare)[66] | 2021-01-10 - 2021-01-16 | National | Moderate | 6 | Adults (18-64 years) | All | Probability | Household and community samples | 0% | NA - IgG, IgM - Author designed (IFA) - MULTIPLEXED | 1, 1 | Validated by developers |
| Finland | Finnish institute for health and welfare (Finnish Institute for Health and Welfare)[66] | 2021-01-03 - 2021-01-09 | National | Moderate | 12 | Adults (18-64 years) | All | Probability | Household and community samples | 0% | NA - IgG, IgM - Author designed (IFA) - MULTIPLEXED | 1, 1 | Validated by developers |
| Finland | Finnish institute for health and welfare (Finnish Institute for Health and Welfare)[66] | 2020-12-27 - 2021-01-02 | National | Moderate | 26 | Adults (18-64 years) | All | Probability | Household and community samples | 0% | NA - IgG, IgM - Author designed (IFA) - MULTIPLEXED | 1, 1 | Validated by developers |
| Finland | Finnish institute for health and welfare (Finnish Institute for Health and Welfare)[66] | 2020-12-20 - 2020-12-26 | National | Moderate | 65 | Adults (18-64 years) | All | Probability | Household and community samples | 0% | NA - IgG, IgM - Author designed (IFA) - MULTIPLEXED | 1, 1 | Validated by developers |
| Finland | Finnish institute for health and welfare (Finnish Institute for Health and Welfare)[66] | 2020-12-13 - 2020-12-19 | National | Moderate | 115 | Adults (18-64 years) | All | Probability | Household and community samples | 0.9% | NA - IgG, IgM - Author designed (IFA) - MULTIPLEXED | 1, 1 | Validated by developers |
| Finland | Finnish institute for health and welfare (Finnish Institute for Health and Welfare)[66] | 2020-12-06 - 2020-12-12 | National | Moderate | 106 | Adults (18-64 years) | All | Probability | Household and community samples | 0.9% | NA - IgG, IgM - Author designed (IFA) - MULTIPLEXED | 1, 1 | Validated by developers |
| Finland | Finnish institute for health and welfare (Finnish Institute for Health and Welfare)[66] | 2020-11-29 - 2020-12-05 | National | Moderate | 80 | Adults (18-64 years) | All | Probability | Household and community samples | 2.5% | NA - IgG, IgM - Author designed (IFA) - MULTIPLEXED | 1, 1 | Validated by developers |
| Finland | Finnish institute for health and welfare (Finnish Institute for Health and Welfare)[66] | 2020-11-22 - 2020-11-28 | National | Moderate | 85 | Adults (18-64 years) | All | Probability | Household and community samples | 3.5% | NA - IgG, IgM - Author designed (IFA) - MULTIPLEXED | 1, 1 | Validated by developers |
| Finland | Finnish institute for health and welfare (Finnish Institute for Health and Welfare)[66] | 2020-11-15 - 2020-11-21 | National | Moderate | 64 | Adults (18-64 years) | All | Probability | Household and community samples | 3.1% | NA - IgG, IgM - Author designed (IFA) - MULTIPLEXED | 1, 1 | Validated by developers |
| Finland | Finnish institute for health and welfare (Finnish Institute for Health and Welfare)[66] | 2020-11-08 - 2020-11-14 | National | Moderate | 52 | Adults (18-64 years) | All | Probability | Household and community samples | 1.9% | NA - IgG, IgM - Author designed (IFA) - MULTIPLEXED | 1, 1 | Validated by developers |
| Finland | Finnish institute for health and welfare (Finnish Institute for Health and Welfare)[66] | 2020-11-01 - 2020-11-07 | National | Moderate | 55 | Adults (18-64 years) | All | Probability | Household and community samples | 5.4% | NA - IgG, IgM - Author designed (IFA) - MULTIPLEXED | 1, 1 | Validated by developers |
| Finland | Finnish institute for health and welfare (Finnish Institute for Health and Welfare)[66] | 2020-10-25 - 2020-10-31 | National | Moderate | 56 | Adults (18-64 years) | All | Probability | Household and community samples | 1.8% | NA - IgG, IgM - Author designed (IFA) - MULTIPLEXED | 1, 1 | Validated by developers |
| Finland | Finnish institute for health and welfare (Finnish Institute for Health and Welfare)[66] | 2020-10-18 - 2020-10-24 | National | Moderate | 55 | Adults (18-64 years) | All | Probability | Household and community samples | 0% | NA - IgG, IgM - Author designed (IFA) - MULTIPLEXED | 1, 1 | Validated by developers |
| Finland | Finnish institute for health and welfare (Finnish Institute for Health and Welfare)[66] | 2020-10-11 - 2020-10-17 | National | Moderate | 59 | Adults (18-64 years) | All | Probability | Household and community samples | 1.7% | NA - IgG, IgM - Author designed (IFA) - MULTIPLEXED | 1, 1 | Validated by developers |
| Finland | Finnish institute for health and welfare (Finnish Institute for Health and Welfare)[66] | 2020-10-04 - 2020-10-10 | National | Moderate | 51 | Adults (18-64 years) | All | Probability | Household and community samples | 0% | NA - IgG, IgM - Author designed (IFA) - MULTIPLEXED | 1, 1 | Validated by developers |
| Finland | Finnish institute for health and welfare (Finnish Institute for Health and Welfare)[66] | 2020-09-27 - 2020-10-03 | National | Moderate | 60 | Adults (18-64 years) | All | Probability | Household and community samples | 1.7% | NA - IgG, IgM - Author designed (IFA) - MULTIPLEXED | 1, 1 | Validated by developers |
| Finland | Finnish institute for health and welfare (Finnish Institute for Health and Welfare)[66] | 2020-04-19 - 2020-04-25 | National | Moderate | 674 | Adults (18-64 years) | All | Probability | Household and community samples | 0.3% | NA - - Author designed (Neutralization Assay),NA - IgG, IgM - Author designed (IFA) - MULTIPLEXED | N/A | Complex test algorithm: Manual review |
| France | Josiane Warszawski (University of Paris-Saclay)[67] | 2020-05-02 - 2020-06-02 | National | Moderate | 12,114 | Multiple groups | All | Stratified non-probability | Household and community samples | 6.5% | EUROIMMUN - IgG - Anti-SARS-CoV-2 ELISA IgG | 0.9, 1 | Validated by manufacturer |
| France | Stephane Le Vu (Santé publique France)[68] | 2020-05-11 - 2020-05-17 | National | Low | 3,592 | Multiple groups | All | Probability | Residual sera | 5% | NA - - Author designed (Neutralization Assay),NA - IgG - Author designed (ELISA) -Spike,NA - IgG - Author designed (ELISA) - Nucleocapsid | N/A | Complex test algorithm: Manual review |
| France | Stephane Le Vu (Santé publique France)[68] | 2020-04-06 - 2020-04-12 | National | Low | 3,595 | Multiple groups | All | Probability | Residual sera | 4.3% | NA - - Author designed (Neutralization Assay),NA - IgG - Author designed (ELISA) -Spike,NA - IgG - Author designed (ELISA) - Nucleocapsid | N/A | Complex test algorithm: Manual review |
| France | Stephane Le Vu (Santé publique France)[68] | 2020-03-09 - 2020-03-15 | National | Low | 3,834 | Multiple groups | All | Probability | Residual sera | 0.7% | NA - - Author designed (Neutralization Assay),NA - IgG - Author designed (ELISA) -Spike,NA - IgG - Author designed (ELISA) - Nucleocapsid | N/A | Complex test algorithm: Manual review |
| Germany | Clemens Fuest (ifo Institute)[69] | 2020-10-26 - 2020-11-18 | National | Moderate | 9,929 | Multiple groups | All | Convenience | Household and community samples | 1.1% | Roche Diagnostics - IgG, IgM, IgA - Elecsys® Anti‐SARS‐CoV‐2 (N) | 1, 0.995 | Validated by manufacturer |
| Germany | Daniela Gornyk (Helmholtz Centre for Infection Research)[70] | 2020-07-15 - 2020-12-15 | National | Moderate | 13,405 | Multiple groups | All | Probability | Household and community samples | 2% | EUROIMMUN - IgG - Anti-SARS-CoV-2 ELISA IgG | 0.9, 1 | Validated by manufacturer |
| Greece | Michalis Koureas (University of Thessaly)[71] | 2020-12-01 - 2020-12-31 | National | Moderate | 6,231 | Multiple groups | All | Probability | Residual sera | 7.9% | Abbott Laboratories - IgG - Abbott Architect SARS-CoV-2 IgG | 0.99, 1 | Validated by manufacturer |
| Holy See | Massimo Ralli (Università Cattolica del Sacro Cuore)[72] | 2021-01-01 - 2021-01-04 | National | Moderate | 17 | Multiple groups | All | Convenience | Household and community samples | 5.9% | Siemens - IgG, IgM, IgA - Atellica® IM SARS-CoV-2 Total (COV2T) | 0.985, 0.998 | Validated by manufacturer |
| Hungary | Bela Merkely (Semmelweis University)[73] | 2020-05-01 - 2020-05-16 | National | Low | 10,474 | Multiple groups | All | Probability | Household and community samples | 0.7% | Abbott Laboratories - IgG - Abbott Architect SARS-CoV-2 IgG | 0.99, 1 | Validated by manufacturer |
| Israel | Victoria Indenbaum (Tel Aviv University)[74] | 2020-01-01 - 2021-03-31 | National | Moderate | 2,765 | Children and Youth (0-17 years) | All | Convenience | Residual sera | 5.6% | NA - - Author designed (ELISA) -Spike,NA - - Author designed (Neutralization Assay) | N/A | Complex test algorithm: Manual review |
| Israel | Shay Reicher (Ministry of Health)[75] | 2020-06-28 - 2020-09-14 | National | Moderate | 54,357 | Multiple groups | All | Convenience | Household and community samples | 4.6% | Abbott Laboratories - IgG - Abbott Architect SARS-CoV-2 IgG,DiaSorin - IgG - Liaison SARS-CoV-2 S1/S2 IgG | 0.964, 1 | Complex test algorithm: Manual calculation |
| Italy | Instituto Nazionale di Statistica (Italian Ministry of Health)[76] | 2020-05-25 - 2020-07-15 | National | Moderate | 64,660 | Multiple groups | All | Stratified non-probability | Household and community samples | 2.5% | Abbott Laboratories - IgG - Abbott Architect SARS-CoV-2 IgG | 0.99, 1 | Validated by manufacturer |
| Jersey | Statistics Jersey (Statistics Jersey)[77] | 2020-06-21 - 2020-06-27 | National | Low | 1,386 | Multiple groups | All | Probability | Household and community samples | 4.2% | CTK Biotech, Inc. - IgG, IgM - OnSite COVID-19 IgG/IgM Rapid Test | 0.969, 0.994 | Validated by manufacturer |
| Lithuania | Kastytis Smigelskas ( Lithuanian University of Health Sciences)[78] | 2020-08-10 - 2020-09-10 | National | Moderate | 3,089 | Multiple groups | All | Probability | Household and community samples | 1.9% | AMEDA Labordiagnostik GmbH - IgG, IgM - AMP Rapid Test SARS-CoV-2 IgG/IgM | 1, 0.975 | Validated by manufacturer |
| Luxembourg | Chantal Snoeck (Luxembourg Institute of Health)[79] | 2020-04-15 - 2020-05-05 | National | Low | 1,820 | Multiple groups | All | Probability | Household and community samples | 2.1% | EUROIMMUN - IgG - Anti-SARS-CoV-2 ELISA IgG | 0.9, 1 | Validated by manufacturer |
| Norway | Gro Tunheim (Norwegian Institute of Public Health)[80] | 2020-12-27 - 2021-02-13 | National | Moderate | 1,912 | Multiple groups | All | Convenience | Residual sera | 3.2% | NA - - Author designed (type unknown) | 0.96, 0.998 | Validated by independent authors/third party/non-developers |
| Portugal | ISN COVID-19 Group (National Institute of Health)[81] | 2021-09-28 - 2021-11-19 | National | Moderate | 4,545 | Multiple groups | All | Stratified non-probability | Household and community samples | 86.4% | Abbott Laboratories - IgG - Abbott Architect SARS-CoV-2 IgG,Abbott Laboratories - IgG - SARS-CoV-2 IgG II Quant | 1, 0.996 | Complex test algorithm: Manual calculation |
| Portugal | Luisa Canto e Castro (Los Alamos National Laboratory)[82] | 2021-03-01 - 2021-03-17 | National | Low | 2,435 | Multiple groups | All | Probability | Household and community samples | 17.3% | Siemens - IgG, IgM, IgA - Atellica® IM SARS-CoV-2 Total (COV2T) | 0.985, 0.998 | Validated by manufacturer |
| Portugal | Luisa Castro (Los Alamos National Laboratory)[83] | 2020-09-08 - 2020-10-14 | National | Moderate | 13,398 | Multiple groups | All | Convenience | Household and community samples | 2.2% | Siemens - IgG, IgM, IgA - ADVIA Centaur Immunoassay System | 0.981, 0.999 | Validated by independent authors/third party/non-developers |
| Portugal | Irina Kislaya ( Instituto Nacional de Saúde Doutor Ricardo Jorge)[84] | 2020-05-21 - 2020-07-08 | National | Moderate | 2,301 | Multiple groups | All | Stratified non-probability | Residual sera | 2.9% | EUROIMMUN - IgG - Anti-SARS-CoV-2 ELISA IgG,Beijing Wantai Biological - IgM - Wantai SARS-CoV-2 IgM ELISA | 0.987, 1 | Complex test algorithm: Manual calculation |
| Slovenia | Mario Poljak (University of Ljubljana)[85] | 2020-10-17 - 2020-11-10 | National | Low | 1,211 | Multiple groups | All | Probability | Household and community samples | 4.3% | Roche Diagnostics - IgG, IgM, IgA - Elecsys® Anti‐SARS‐CoV‐2 (N),Roche Diagnostics - IgG, IgM, IgA - Elecsys® Anti‐SARS‐CoV‐2 (S) | 1, 0.995 | Complex test algorithm: Manual calculation |
| Slovenia | Mario Poljak (University of Ljubljana)[85] | 2020-04-20 - 2020-05-01 | National | Low | 1,316 | Multiple groups | All | Probability | Household and community samples | 3.1% | EUROIMMUN - IgG - Anti-SARS-CoV-2 ELISA IgG | 0.9, 1 | Validated by manufacturer |
| Spain | Mayte Perez-Olmeda (Instituto de Salud Carlos III)[86] | 2020-11-16 - 2020-11-29 | National | Moderate | 5,827 | Multiple groups | All | Probability | Household and community samples | 5.4% | Beckman Coulter - IgG - Access SARS-CoV-2 IgG Antibody Test | 1, 0.998 | Validated by manufacturer |
| Spain | Ministry of Health (Instituto de Salud Carlos III)[87] | 2020-06-08 - 2020-06-22 | National | Low | 62,167 | Multiple groups | All | Probability | Household and community samples | 5.2% | Zhejiang Orient Gene Biotech - IgG, IgM - COVID-19 IgG/IgM Rapid Test | 0.972, 1 | Validated by manufacturer |
| Spain | Nicolas Prados (IVIRMA Global Headquarters)[88] | 2020-04-27 - 2020-06-26 | National | Moderate | 6,140 | Adults (18-64 years) | All | Convenience | Residual sera | 0.7% | Epitope Diagnostics, Inc. - IgM - EDI™ Novel Coronavirus COVID-19 IgM ELISA Kit | 1, 1 | Validated by manufacturer |
| Sweden | Swedish Public Health Agency (Swedish Public Health Agency)[89] | 2021-09-20 - 2021-10-03 | National | Moderate | 2,959 | Multiple groups | All | Convenience | Residual sera | 75.6% | Sci LifeLab / KTH - - Not available | 0.989, 0.994 | Validated by independent authors/third party/non-developers |
| Sweden | Swedish Public Health Agency (Swedish Public Health Agency)[90] | 2021-05-24 - 2021-06-04 | National | Moderate | 3,949 | Multiple groups | All | Stratified non-probability | Residual sera | 52.2% | Sci LifeLab / KTH - - Not available | 0.989, 0.994 | Validated by independent authors/third party/non-developers |
| Sweden | Swedish Public Health Agency (The Swedish Public Health Agency (Folkhälsomyndigheten))[91] | 2020-11-23 - 2020-12-04 | National | Moderate | 3,183 | Multiple groups | All | Convenience | Blood donors | 7% | Sci LifeLab / KTH - - Not available | 0.994, 0.989 | Validated by manufacturers |
| The United Kingdom | Public Health Scotland (Public Health Scotland)[92] | 2022-02-28 - 2022-04-03 | National | Moderate | 2,499 | Multiple groups | All | Sequential | Blood donors | 98.6% | EUROIMMUN - IgG - Anti-SARS-CoV-2 ELISA IgG | 0.9, 1 | Validated by manufacturer |
| The United Kingdom | Public Health Scotland (Public Health Scotland)[92] | 2022-02-28 - 2022-04-03 | National | Moderate | 3,181 | Adults (18-64 years) | Female | Sequential | Pregnant or parturient women | 95% | Roche Diagnostics - IgG, IgM, IgA - Elecsys® Anti‐SARS‐CoV‐2 (S) | 0.988, 1 | Validated by manufacturer |
| The United Kingdom | Public Health Scotland (Public Health Scotland)[92] | 2022-02-28 - 2022-04-03 | National | Low | 2,885 | Multiple groups | All | Probability | Residual sera | 91.8% | DiaSorin - IgG - Liaison SARS-CoV-2 S1/S2 IgG | 0.974, 0.985 | Validated by manufacturer |
| The United Kingdom | Public Health Scotland (Public Health Scotland)[93] | 2022-01-24 - 2022-02-27 | National | Moderate | 2,499 | Multiple groups | All | Sequential | Blood donors | 98.6% | EUROIMMUN - IgG - Anti-SARS-CoV-2 ELISA IgG | 0.9, 1 | Validated by manufacturer |
| The United Kingdom | Public Health Scotland (Public Health Scotland)[93] | 2022-01-24 - 2022-02-27 | National | Moderate | 3,217 | Adults (18-64 years) | Female | Sequential | Pregnant or parturient women | 94% | Roche Diagnostics - IgG, IgM, IgA - Elecsys® Anti‐SARS‐CoV‐2 (S) | 0.988, 1 | Validated by manufacturer |
| The United Kingdom | Public Health Scotland (Public Health Scotland)[93] | 2022-01-24 - 2022-02-27 | National | Low | 2,884 | Multiple groups | All | Probability | Residual sera | 90.7% | DiaSorin - IgG - Liaison SARS-CoV-2 S1/S2 IgG | 0.974, 0.985 | Validated by manufacturer |
| The United Kingdom | Public Health Scotland (Public Health Scotland)[94] | 2021-12-20 - 2022-01-23 | National | Moderate | 2,499 | Multiple groups | All | Sequential | Blood donors | 98.7% | EUROIMMUN - IgG - Anti-SARS-CoV-2 ELISA IgG | 0.9, 1 | Validated by manufacturer |
| The United Kingdom | Public Health Scotland (Public Health Scotland)[94] | 2021-12-20 - 2022-01-23 | National | Low | 2,863 | Multiple groups | All | Probability | Residual sera | 88.9% | DiaSorin - IgG - Liaison SARS-CoV-2 S1/S2 IgG | 0.974, 0.985 | Validated by manufacturer |
| The United Kingdom | Public Health Scotland (Public Health Scotland)[95] | 2021-11-15 - 2021-12-19 | National | Moderate | 2,493 | Multiple groups | All | Sequential | Blood donors | 96.3% | EUROIMMUN - IgG - Anti-SARS-CoV-2 ELISA IgG | 0.9, 1 | Validated by manufacturer |
| The United Kingdom | Public Health Scotland (Public Health Scotland)[95] | 2021-11-15 - 2021-12-19 | National | Low | 2,815 | Multiple groups | All | Probability | Residual sera | 85.3% | DiaSorin - IgG - Liaison SARS-CoV-2 S1/S2 IgG | 0.974, 0.985 | Validated by manufacturer |
| The United Kingdom | Public Health Scotland (Public Health Scotland)[96] | 2021-10-11 - 2021-11-14 | National | Moderate | 2,496 | Multiple groups | All | Sequential | Blood donors | 94.4% | EUROIMMUN - IgG - Anti-SARS-CoV-2 ELISA IgG | 0.9, 1 | Validated by manufacturer |
| The United Kingdom | Public Health Scotland (Public Health Scotland)[96] | 2021-10-11 - 2021-11-14 | National | Low | 2,882 | Multiple groups | All | Probability | Residual sera | 78.1% | DiaSorin - IgG - Liaison SARS-CoV-2 S1/S2 IgG | 0.974, 0.985 | Validated by manufacturer |
| The United Kingdom | Clarissa Oeser (UK Health Security Agency (UKHSA, formerly PHE))[97] | 2021-09-01 - 2021-10-31 | National | Moderate | 600 | Children and Youth (0-17 years) | All | Convenience | Residual sera | 54.5% | Roche Diagnostics - IgG, IgM, IgA - Elecsys® Anti‐SARS‐CoV‐2 (S) | 0.988, 1 | Validated by manufacturer |
| The United Kingdom | Public Health Scotland (Public Health Scotland)[98] | 2021-09-06 - 2021-10-10 | National | Moderate | 2,494 | Multiple groups | All | Sequential | Blood donors | 95.3% | EUROIMMUN - IgG - Anti-SARS-CoV-2 ELISA IgG | 0.9, 1 | Validated by manufacturer |
| The United Kingdom | Public Health Scotland (Public Health Scotland)[98] | 2021-09-06 - 2021-10-10 | National | Moderate | 3,023 | Adults (18-64 years) | Female | Sequential | Pregnant or parturient women | 18.1% | Roche Diagnostics - IgG, IgM, IgA - Elecsys® Anti‐SARS‐CoV‐2 (N) | 1, 0.995 | Validated by manufacturer |
| The United Kingdom | Public Health Scotland (Public Health Scotland)[98] | 2021-09-06 - 2021-10-10 | National | Low | 3,132 | Multiple groups | All | Probability | Residual sera | 74.3% | DiaSorin - IgG - Liaison SARS-CoV-2 S1/S2 IgG | 0.974, 0.985 | Validated by manufacturer |
| The United Kingdom | Clarissa Oeser (UK Health Security Agency (UKHSA, formerly PHE))[97] | 2021-07-01 - 2021-08-31 | National | Moderate | 685 | Children and Youth (0-17 years) | All | Convenience | Residual sera | 29.5% | Roche Diagnostics - IgG, IgM, IgA - Elecsys® Anti‐SARS‐CoV‐2 (S) | 0.988, 1 | Validated by manufacturer |
| The United Kingdom | Public Health Scotland (Public Health Scotland)[99] | 2021-06-28 - 2021-08-01 | National | Moderate | 2,493 | Multiple groups | All | Sequential | Blood donors | 94.2% | EUROIMMUN - IgG - Anti-SARS-CoV-2 ELISA IgG | 0.9, 1 | Validated by manufacturer |
| The United Kingdom | Public Health Scotland (Public Health Scotland)[99] | 2021-06-28 - 2021-08-01 | National | Moderate | 2,990 | Adults (18-64 years) | Female | Sequential | Pregnant or parturient women | 12.7% | Roche Diagnostics - IgG, IgM, IgA - Elecsys® Anti‐SARS‐CoV‐2 (N) | 1, 0.995 | Validated by manufacturer |
| The United Kingdom | Public Health Scotland (Public Health Scotland)[99] | 2021-06-28 - 2021-08-01 | National | Low | 3,427 | Multiple groups | All | Probability | Residual sera | 69.4% | DiaSorin - IgG - Liaison SARS-CoV-2 S1/S2 IgG | 0.974, 0.985 | Validated by manufacturer |
| The United Kingdom | Public Health Scotland (Public Health Scotland)[100] | 2021-05-24 - 2021-06-27 | National | Moderate | 2,494 | Multiple groups | All | Sequential | Blood donors | 76.8% | EUROIMMUN - IgG - Anti-SARS-CoV-2 ELISA IgG | 0.9, 1 | Validated by manufacturer |
| The United Kingdom | Public Health Scotland (Public Health Scotland)[100] | 2021-05-24 - 2021-06-27 | National | Moderate | 3,052 | Adults (18-64 years) | Female | Sequential | Pregnant or parturient women | 11.2% | Roche Diagnostics - IgG, IgM, IgA - Elecsys® Anti‐SARS‐CoV‐2 (N) | 1, 0.995 | Validated by manufacturer |
| The United Kingdom | Public Health Scotland (Public Health Scotland)[100] | 2021-05-24 - 2021-06-27 | National | Low | 3,554 | Multiple groups | All | Probability | Residual sera | 59.3% | DiaSorin - IgG - Liaison SARS-CoV-2 S1/S2 IgG | 0.974, 0.985 | Validated by manufacturer |
| The United Kingdom | Clarissa Oeser (UK Health Security Agency (UKHSA, formerly PHE))[97] | 2021-05-01 - 2021-06-30 | National | Moderate | 1,186 | Children and Youth (0-17 years) | All | Convenience | Residual sera | 27.7% | Roche Diagnostics - IgG, IgM, IgA - Elecsys® Anti‐SARS‐CoV‐2 (S) | 0.988, 1 | Validated by manufacturer |
| The United Kingdom | Public Health Scotland (Public Health Scotland)[101] | 2021-04-19 - 2021-05-23 | National | Moderate | 2,494 | Multiple groups | All | Sequential | Blood donors | 58.8% | EUROIMMUN - IgG - Anti-SARS-CoV-2 ELISA IgG | 0.9, 1 | Validated by manufacturer |
| The United Kingdom | Public Health Scotland (Public Health Scotland)[101] | 2021-04-19 - 2021-05-23 | National | Moderate | 3,333 | Adults (18-64 years) | Female | Sequential | Pregnant or parturient women | 10% | Roche Diagnostics - IgG, IgM, IgA - Elecsys® Anti‐SARS‐CoV‐2 (N) | 1, 0.995 | Validated by manufacturer |
| The United Kingdom | Public Health Scotland (Public Health Scotland)[101] | 2021-05-03 - 2021-05-09 | National | Low | 713 | Multiple groups | All | Probability | Residual sera | 47.5% | DiaSorin - IgG - Liaison SARS-CoV-2 S1/S2 IgG | 0.974, 0.985 | Validated by manufacturer |
| The United Kingdom | Public Health Scotland (Public Health Scotland)[102] | 2021-04-12 - 2021-04-18 | National | Low | 708 | Multiple groups | All | Probability | Residual sera | 42.8% | DiaSorin - IgG - Liaison SARS-CoV-2 S1/S2 IgG | 0.974, 0.985 | Validated by manufacturer |
| The United Kingdom | Public Health Scotland (Public Health Scotland)[102] | 2021-03-15 - 2021-04-18 | National | Moderate | 2,493 | Multiple groups | All | Sequential | Blood donors | 39.7% | EUROIMMUN - IgG - Anti-SARS-CoV-2 ELISA IgG | 0.9, 1 | Validated by manufacturer |
| The United Kingdom | Public Health Scotland (Public Health Scotland)[102] | 2021-03-15 - 2021-04-18 | National | Moderate | 3,326 | Adults (18-64 years) | Female | Sequential | Pregnant or parturient women | 11.4% | Roche Diagnostics - IgG, IgM, IgA - Elecsys® Anti‐SARS‐CoV‐2 (N) | 1, 0.995 | Validated by manufacturer |
| The United Kingdom | Clarissa Oeser (UK Health Security Agency (UKHSA, formerly PHE))[97] | 2021-03-01 - 2021-04-30 | National | Moderate | 925 | Children and Youth (0-17 years) | All | Convenience | Residual sera | 31.1% | Roche Diagnostics - IgG, IgM, IgA - Elecsys® Anti‐SARS‐CoV‐2 (S) | 0.988, 1 | Validated by manufacturer |
| The United Kingdom | Robert W Aldridge (University College London)[103] | 2021-02-01 - 2021-05-03 | National | Moderate | 10,330 | Multiple groups | All | Convenience | Household and community samples | 8.2% | Roche Diagnostics - IgG, IgM, IgA - Elecsys® Anti‐SARS‐CoV‐2 (N) | 1, 0.995 | Validated by manufacturer |
| The United Kingdom | Heather J Whitaker (Public Health England)[104] | 2021-02-22 - 2021-03-21 | National | Moderate | 7,720 | Multiple groups | All | Convenience | Blood donors | 46.4% | Roche Diagnostics - IgG, IgM, IgA - Elecsys® Anti‐SARS‐CoV‐2 (S) | 0.988, 1 | Validated by manufacturer |
| The United Kingdom | Public Health Scotland (Public Health Scotland)[105] | 2021-02-08 - 2021-03-14 | National | Moderate | 2,492 | Multiple groups | All | Sequential | Blood donors | 22.1% | EUROIMMUN - IgG - Anti-SARS-CoV-2 ELISA IgG | 0.9, 1 | Validated by manufacturer |
| The United Kingdom | Public Health Scotland (Public Health Scotland)[105] | 2021-02-08 - 2021-03-14 | National | Moderate | 3,355 | Adults (18-64 years) | Female | Sequential | Pregnant or parturient women | 8.9% | Roche Diagnostics - IgG, IgM, IgA - Elecsys® Anti‐SARS‐CoV‐2 (N) | 1, 0.995 | Validated by manufacturer |
| The United Kingdom | Clarissa Oeser (UK Health Security Agency (UKHSA, formerly PHE))[97] | 2021-01-01 - 2021-02-28 | National | Moderate | 970 | Children and Youth (0-17 years) | All | Convenience | Residual sera | 20.7% | Roche Diagnostics - IgG, IgM, IgA - Elecsys® Anti‐SARS‐CoV‐2 (S) | 0.988, 1 | Validated by manufacturer |
| The United Kingdom | Public Health Scotland (Public Health Scotland)[106] | 2021-01-04 - 2021-02-07 | National | Moderate | 2,497 | Multiple groups | All | Sequential | Blood donors | 12.1% | EUROIMMUN - IgG - Anti-SARS-CoV-2 ELISA IgG | 0.9, 1 | Validated by manufacturer |
| The United Kingdom | Public Health Scotland (Public Health Scotland)[106] | 2021-01-04 - 2021-02-07 | National | Moderate | 3,478 | Adults (18-64 years) | Female | Sequential | Pregnant or parturient women | 6.9% | Roche Diagnostics - IgG, IgM, IgA - Elecsys® Anti‐SARS‐CoV‐2 (N) | 1, 0.995 | Validated by manufacturer |
| The United Kingdom | Public Health Scotland (Public Health Scotland)[107] | 2020-11-30 - 2021-01-03 | National | Moderate | 2,499 | Multiple groups | All | Sequential | Blood donors | 4.2% | EUROIMMUN - IgG - Anti-SARS-CoV-2 ELISA IgG | 0.9, 1 | Validated by manufacturer |
| The United Kingdom | Public Health Scotland (Public Health Scotland)[107] | 2020-11-30 - 2021-01-03 | National | Moderate | 3,052 | Adults (18-64 years) | Female | Sequential | Pregnant or parturient women | 5.3% | Roche Diagnostics - IgG, IgM, IgA - Elecsys® Anti‐SARS‐CoV‐2 (N) | 1, 0.995 | Validated by manufacturer |
| The United Kingdom | Clarissa Oeser (UK Health Security Agency (UKHSA, formerly PHE))[97] | 2020-11-01 - 2020-12-31 | National | Moderate | 711 | Children and Youth (0-17 years) | All | Convenience | Residual sera | 13.2% | Roche Diagnostics - IgG, IgM, IgA - Elecsys® Anti‐SARS‐CoV‐2 (S) | 0.988, 1 | Validated by manufacturer |
| The United Kingdom | Heather Whitaker (UK Health Security Agency)[108] | 2020-11-01 - 2020-12-31 | National | Moderate | 4,934 | Multiple groups | All | Sequential | Residual sera | 6.6% | EUROIMMUN - IgG - Anti-SARS-CoV-2 ELISA IgG | 0.9, 1 | Validated by manufacturer |
| The United Kingdom | Shamez Ladhani (Public Health England)[109] | 2020-11-17 - 2020-11-30 | National | Moderate | 1,216 | Multiple groups | All | Probability | Household and community samples | 10.4% | Abbott Laboratories - IgG - Abbott Architect SARS-CoV-2 IgG | 0.99, 1 | Validated by manufacturer |
| The United Kingdom | Public Health Scotland (Public Health Scotland)[110] | 2020-10-26 - 2020-11-29 | National | Moderate | 2,499 | Multiple groups | All | Sequential | Blood donors | 4.2% | EUROIMMUN - IgG - Anti-SARS-CoV-2 ELISA IgG | 0.9, 1 | Validated by manufacturer |
| The United Kingdom | Heather Whitaker (UK Health Security Agency)[108] | 2020-09-01 - 2020-10-01 | National | Moderate | 5,478 | Multiple groups | All | Sequential | Residual sera | 5.5% | EUROIMMUN - IgG - Anti-SARS-CoV-2 ELISA IgG | 0.9, 1 | Validated by manufacturer |
| The United Kingdom | Public Health England (Public Health England)[111] | 2020-08-19 - 2020-09-13 | National | Low | 7,888 | Multiple groups | All | Probability | Blood donors | 6.1% | EUROIMMUN - IgG - Anti-SARS-CoV-2 ELISA IgG | 0.9, 1 | Validated by manufacturer |
| The United Kingdom | Public Health England (Public Health England)[111] | 2020-08-13 - 2020-09-06 | National | Low | 7,899 | Multiple groups | All | Probability | Blood donors | 5.5% | EUROIMMUN - IgG - Anti-SARS-CoV-2 ELISA IgG | 0.9, 1 | Validated by manufacturer |
| The United Kingdom | Heather Whitaker (UK Health Security Agency)[108] | 2020-07-01 - 2020-08-31 | National | Moderate | 5,781 | Multiple groups | All | Sequential | Residual sera | 5.2% | EUROIMMUN - IgG - Anti-SARS-CoV-2 ELISA IgG | 0.9, 1 | Validated by manufacturer |
| The United Kingdom | Public Health England (Public Health England)[111] | 2020-07-13 - 2020-08-07 | National | Low | 8,538 | Multiple groups | All | Probability | Blood donors | 5.4% | EUROIMMUN - IgG - Anti-SARS-CoV-2 ELISA IgG | 0.9, 1 | Validated by manufacturer |
| The United Kingdom | Public Health England (Public Health England)[111] | 2020-07-01 - 2020-08-14 | National | Low | 2,153 | Seniors (65+ years) | All | Probability | Residual sera | 3.4% | Abbott Laboratories - IgG - Abbott Architect SARS-CoV-2 IgG | 0.99, 1 | Validated by manufacturer |
| The United Kingdom | Public Health England (Public Health England)[111] | 2020-06-01 - 2020-08-02 | National | Moderate | 285 | Children and Youth (0-17 years) | All | Probability | Household and community samples | 4.9% | Abbott Laboratories - IgG - Abbott Architect SARS-CoV-2 IgG | 0.99, 1 | Validated by manufacturer |
| The United Kingdom | Public Health England (Public Health England)[111] | 2020-06-01 - 2020-08-02 | National | Low | 2,062 | Children and Youth (0-17 years) | All | Probability | Residual sera | 2.4% | Abbott Laboratories - IgG - Abbott Architect SARS-CoV-2 IgG | 0.99, 1 | Validated by manufacturer |
| The United Kingdom | Public Health England (Public Health England)[111] | 2020-06-08 - 2020-07-06 | National | Low | 11,385 | Multiple groups | All | Probability | Blood donors | 7.1% | EUROIMMUN - IgG - Anti-SARS-CoV-2 ELISA IgG | 0.9, 1 | Validated by manufacturer |
| The United Kingdom | Heather Whitaker (UK Health Security Agency)[108] | 2020-05-01 - 2020-06-30 | National | Moderate | 5,455 | Multiple groups | All | Sequential | Residual sera | 4.1% | EUROIMMUN - IgG - Anti-SARS-CoV-2 ELISA IgG | 0.9, 1 | Validated by manufacturer |
| The United Kingdom | Public Health England (Public Health England)[111] | 2020-05-13 - 2020-06-07 | National | Low | 7,777 | Adults (18-64 years) | All | Probability | Blood donors | 8.2% | EUROIMMUN - IgG - Anti-SARS-CoV-2 ELISA IgG | 0.9, 1 | Validated by manufacturer |
| The United Kingdom | Public Health England (Public Health England)[111] | 2020-05-01 - 2020-05-31 | National | Moderate | 1,061 | Children and Youth (0-17 years) | All | Probability | Residual sera | 7.8% | Abbott Laboratories - IgG - Abbott Architect SARS-CoV-2 IgG | 0.99, 1 | Validated by manufacturer |
| The United Kingdom | Public Health England (Public Health England)[111] | 2020-03-16 - 2020-06-30 | National | Low | 6,000 | Seniors (65+ years) | All | Probability | Residual sera | 3.9% | EUROIMMUN - IgG - Anti-SARS-CoV-2 ELISA IgG | 0.9, 1 | Validated by manufacturer |
| The United Kingdom | Public Health England (Public Health England)[111] | 2020-04-01 - 2020-04-30 | National | Moderate | 647 | Children and Youth (0-17 years) | All | Probability | Residual sera | 6.6% | Abbott Laboratories - IgG - Abbott Architect SARS-CoV-2 IgG | 0.99, 1 | Validated by manufacturer |
| The United Kingdom | Public Health England (Public Health England)[111] | 2020-04-01 - 2020-04-30 | National | Moderate | 201 | Children and Youth (0-17 years) | All | Probability | Household and community samples | 4.5% | Abbott Laboratories - IgG - Abbott Architect SARS-CoV-2 IgG | 0.99, 1 | Validated by manufacturer |
| The United Kingdom | Public Health England (Public Health England)[111] | 2020-02-01 - 2020-03-31 | National | Moderate | 106 | Children and Youth (0-17 years) | All | Probability | Household and community samples | 1.4% | Abbott Laboratories - IgG - Abbott Architect SARS-CoV-2 IgG | 0.99, 1 | Validated by manufacturer |

###### South-East Asia region

| Country (Location) | Author (Organization) | Sampling Dates (YMD) | Geographic scope | Overall risk of bias | Sample size | Age | Sex | Sampling method | Sample frame | Seroprevalence (95% CI) | Test Manufacturers - Isotypes - Names | Test Sens, Spec | Sens, Spec Source |
| --- | --- | --- | --- | --- | --- | --- | --- | --- | --- | --- | --- | --- | --- |
| India | Manoj V Murhekar (Indian Council of Medical Research)[112] | 2020-12-18 - 2021-01-06 | National | Low | 28,598 | Multiple groups | All | Probability | Household and community samples | 24.6% | Siemens - IgG, IgM, IgA - Atellica® IM SARS-CoV-2 Total (COV2T),Abbott Laboratories - IgG - Abbott Architect SARS-CoV-2 IgG | 1, 0.998 | Complex test algorithm: Manual calculation |
| India | Manoj V Murhekar (The Indian Council of Medical Research (ICMR))[112] | 2020-08-18 - 2020-09-20 | National | Low | 29,082 | Multiple groups | All | Probability | Household and community samples | 7% | Abbott Laboratories - IgG - Abbott Architect SARS-CoV-2 IgG | 0.99, 1 | Validated by manufacturer |
| Indonesia | Dr. dr. Tri Yunis Miko Wahyono, M.Sc (University of Indonesia)[113] | 2020-12-15 - 2021-03-03 | National | Moderate | 10,161 | Multiple groups | All | Probability | Household and community samples | 20.5% (95% CI 19.7-21.3%) | Beijing Wantai Biological - IgG IgM IgA - Wantai SARS-CoV-2 Total Ab ELISA | 0.967, 0.975 | Validated by manufacturer |
| Nepal | Government of Nepal (Government of Nepal)[114] | 2020-10-09 - 2020-10-22 | National | Low | 3,040 | Multiple groups | All | Probability | Household and community samples | 14.4% | Beijing Wantai Biological - IgG, IgM, IgA - Wantai SARS-CoV-2 Total Ab ELISA | 0.967, 0.975 | Validated by manufacturer |
| Thailand | Sutthichai Nakphook, MD (Ministry of Public Health)[115] | 2020-12-18 - 2021-02-02 | National | Moderate | 1,200 | Multiple groups | All | Probability | Household and community samples | 0.4% (95% CI 0.1-1%) | Beijing Wantai Biological - IgG IgM IgA - Wantai SARS-CoV-2 Total Ab ELISA | 0.967, 0.975 | Validated by manufacturer |

###### Western Pacific region

| Country (Location) | Author (Organization) | Sampling Dates (YMD) | Geographic scope | Overall risk of bias | Sample size | Age | Sex | Sampling method | Sample frame | Seroprevalence (95% CI) | Test Manufacturers - Isotypes - Names | Test Sens, Spec | Sens, Spec Source |
| --- | --- | --- | --- | --- | --- | --- | --- | --- | --- | --- | --- | --- | --- |
| Australia | Kaitlyn Vette (University of Sydney)[116] | 2020-06-19 - 2020-08-06 | National | Moderate | 2,972 | Adults (18-64 years) | Female | Convenience | Pregnant or parturient women | 0.5% | Beijing Wantai Biological - IgG, IgM, IgA - Wantai SARS-CoV-2 Total Ab ELISA | 0.967, 0.975 | Validated by manufacturer |
| Australia | Nicholas Coatsworth (Australian National University)[117] | 2020-06-02 - 2020-07-17 | National | Moderate | 2,991 | Multiple groups | All | Sequential | Residual sera | 0.2% | NA - - Author designed (ELISA) - MULTIPLEXED | 1, 0.989 | Validated by independent authors/third party/non-developers |
| China | Zhongjie Li (Chinese Center for Disease Control and Prevention)[118] | 2020-04-10 - 2020-04-18 | National | Moderate | 34,857 | Multiple groups | All | Probability | Household and community samples | 1.2% | Guangzhou Wondfo Biotech Co. Ltd - IgG, IgM - Finecare SARS-CoV-2 Antibody test,Innovita Biological Technology Co. Ltd - IgG - 2019-nCoV IgG Rapid Test,Bioscience Diagnostics - IgG, IgM - Magnetic Chemiluminescence Enzyme Immunoassay Kit,NA - - Author designed (Neutralization Assay) | N/A | Complex test algorithm: Manual review |
| Japan | Takashi Yoshiyama (Government of Japan)[119] | 2020-06-01 - 2020-06-07 | National | Moderate | 7,950 | Multiple groups | All | Probability | Household and community samples | 0.1% | Roche Diagnostics - IgG, IgM, IgA - Elecsys® Anti‐SARS‐CoV‐2 (N),Abbott Laboratories - IgG - Abbott Architect SARS-CoV-2 IgG | 0.99, 1 | Complex test algorithm: Manual calculation |
| Malaysia | Chong Zhuo Lin (Institute for Public Health)[120] | 2020-08-07 - 2020-10-06 | National | Moderate | 5,131 | Multiple groups | All | Probability | Household and community samples | 0.6% (95% CI 0.4-0.8%) | Beijing Wantai Biological - IgG IgM IgA - Wantai SARS-CoV-2 Total Ab ELISA,GenScript - IgG IgM - cPass SARS-CoV-2 Surrogate Virus Neutralization Test ELISA Kit | 0.907, 1 | Complex test algorithm: Manual calculation |
| Mongolia | Battogtokh Chimeddorj (Mongolian National University of Medical Sciences (MNUMS))[121] | 2021-01-18 - 2021-03-15 | National | Moderate | 4,530 | Multiple groups | All | Convenience | Household and community samples | 1.6% (95% CI 1.3-2%) | Beijing Wantai Biological - IgG IgM IgA - Wantai SARS-CoV-2 Total Ab ELISA,Kantaro Biosciences LLC - IgG - COVID-SeroKlir, Kantaro Semi-Quantitative SARS-CoV-2 IgG Antibody Kit | 0.955, 1 | Complex test algorithm: Manual calculation |
| Mongolia | Battogtokh Chimeddorj (Mongolian National University of Medical Sciences (MNUMS))[121] | 2020-10-18 - 2020-12-05 | National | Moderate | 5,000 | Multiple groups | All | Convenience | Household and community samples | 1.4% (95% CI 1.1-1.8%) | Beijing Wantai Biological - IgG IgM IgA - Wantai SARS-CoV-2 Total Ab ELISA,Kantaro Biosciences LLC - IgG - COVID-SeroKlir, Kantaro Semi-Quantitative SARS-CoV-2 IgG Antibody Kit | 0.955, 1 | Complex test algorithm: Manual calculation |
| Papua New Guinea | Moses Laman (Papua New Guinea Institute of Medical Research)[122] | 2020-05-01 - 2020-06-30 | National | Moderate | 12,177 | Multiple groups | All | Convenience | Household and community samples | 2.6% (95% CI 2.4-2.9%) | Hanghzhou AllTest Biotech Co. Ltd - IgG IgM - 2019-nCoV IgG/IgM Rapid Test Cassette | 1, 0.962 | Validated by manufacturer |
| Republic of Korea | Ah-Ra Kim (National Institute of Infectious Diseases)[123] | 2020-11-01 - 2020-12-12 | National | Moderate | 910 | Multiple groups | All | Convenience | Residual sera | 0.1% | Roche Diagnostics - IgG, IgM, IgA - Elecsys® Anti‐SARS‐CoV‐2 (N),Abbott Laboratories - IgG - Abbott Architect SARS-CoV-2 IgG,Beckman Coulter - IgG - Access SARS-CoV-2 IgG Antibody Test,SG medical, Inc. - - R-FIND COVID-19 ELISA,Sugentech, Inc. - IgG, IgM - SGTi-flex COVID-19 IgM/IgG (manual),NA - - Author designed (Neutralization Assay) | N/A | Complex test algorithm: Manual review |
| Republic of Korea | Eun Hee Nah (Korea Association of Health Promotion)[124] | 2020-09-24 - 2020-12-09 | National | Moderate | 4,085 | Multiple groups | All | Probability | Residual sera | 0.4% | Roche Diagnostics - IgG, IgM, IgA - Elecsys® Anti‐SARS‐CoV‐2 (N) | 1, 0.995 | Validated by manufacturer |

##### Table S6. Additional studies used in meta-analysis and meta-regression.

###### Africa region

| Country (Location) | Author (Organization) | Sampling Dates (YMD) | Geographic scope | Overall risk of bias | Sample size | Age | Sex | Sampling method | Sample frame | Seroprevalence (95% CI) | Test Manufacturers - Isotypes - Names | Test Sens, Spec | Sens, Spec Source |
| --- | --- | --- | --- | --- | --- | --- | --- | --- | --- | --- | --- | --- | --- |
| Democratic Republic of the Congo - HRP (All the 35 health districts of Kinshasa) | Makiala Mandanda Sheila (Institut National de Recherche Biomédicale)[125] | 2021-03-06 - 2021-03-14 | Subnational | Moderate | 2,476 | Multiple groups | All | Probability | Household and community samples | 72.1% (95% CI 70.3-73.9%) | Beijing Wantai Biological - IgG IgM IgA - Wantai SARS-CoV-2 Total Ab ELISA | 0.967, 0.975 | Validated by manufacturer |
| Ethiopia - HRP (Oromia, Harari) | Nega Assefa (The London School of Hygiene & Tropical Medicine)[126] | 2020-04-01 - 2021-03-31 | Subnational | Moderate | 1,447 | Adults (18-64 years) | Female | Probability | Pregnant or parturient women | 5.7% | Beijing Wantai Biological - IgG, IgM, IgA - Wantai Rapid Test for Total Antibody to SARS-CoV-2 | 0.947, 0.989 | Validated by manufacturer |
| Nigeria - HRP (Anambra State) | OgoChukwu Vincent Okpala (Anambra State Ministry of Health)[127] | 2020-12-08 - 2020-12-15 | Subnational | Moderate | 3,142 | Multiple groups | All | Probability | Household and community samples | 17.6% | Hangzhou Realy Tech Co. Ltd - IgG, IgM - Realy-Tech 2019 nCOV/COVID-19 IgG/IgM Rapid Test | 0.967, 0.937 | Validated by manufacturer |
| Nigeria - HRP (Enugu State) | Kristen A. Stafford (UMB); Laura Steinhardt (CDC); Elsie Ilori (NCDC); Rosemary Audu (NIMR) (University of Maryland, Baltimore)[128] | 2020-09-21 - 2020-10-27 | Subnational | Low | 2,147 | Multiple groups | All | Probability | Household and community samples | 25.2% | Abbott Laboratories - IgG - Abbott Architect SARS-CoV-2 IgG,EUROIMMUN AG - IgG - SARS-CoV-2-NCP-IgG ELISA | N/A | Complex test algorithm: Manual review |
| Nigeria - HRP (Gombe State) | Kristen A. Stafford (UMB); Laura Steinhardt (CDC); Elsie Ilori (NCDC); Rosemary Audu (NIMR) (University of Maryland, Baltimore)[128] | 2020-09-21 - 2020-10-27 | Subnational | Low | 3,432 | Multiple groups | All | Probability | Household and community samples | 9.3% | Abbott Laboratories - IgG - Abbott Architect SARS-CoV-2 IgG,EUROIMMUN AG - IgG - SARS-CoV-2-NCP-IgG ELISA | N/A | Complex test algorithm: Manual review |
| Nigeria - HRP (Nasarawa State) | Kristen A. Stafford (UMB); Laura Steinhardt (CDC); Elsie Ilori (NCDC); Rosemary Audu (NIMR) (University of Maryland, Baltimore)[128] | 2020-09-21 - 2020-10-27 | Subnational | Low | 2,657 | Multiple groups | All | Probability | Household and community samples | 18% | Abbott Laboratories - IgG - Abbott Architect SARS-CoV-2 IgG,EUROIMMUN AG - IgG - SARS-CoV-2-NCP-IgG ELISA | N/A | Complex test algorithm: Manual review |
| Nigeria - HRP (Lagos State) | Kristen A. Stafford (UMB); Laura Steinhardt (CDC); Elsie Ilori (NCDC); Rosemary Audu (NIMR) (Nigeria Institute for Medical Research)[128] | 2020-09-21 - 2020-10-27 | Subnational | Low | 2,393 | Multiple groups | All | Probability | Household and community samples | 23.2% | Abbott Laboratories - IgG - Abbott Architect SARS-CoV-2 IgG,EUROIMMUN AG - IgG - SARS-CoV-2-NCP-IgG ELISA | N/A | Complex test algorithm: Manual review |
| Nigeria - HRP (Niger State) | Hussaini Majiya (Ibrahim Badamasi Babangida University)[129] | 2020-06-26 - 2020-06-30 | Subnational | Moderate | 185 | Multiple groups | All | Probability | Household and community samples | 25.4% | Cambridge Network - IgG, IgM - COVID-19 IgG and IgM Rapid Test | 1, 1 | Validated by independent authors/third party/non-developers |
| South Africa (Gauteng) | Jaya George (National Health Laboratory Service)[130] | 2020-10-01 - 2020-10-31 | Subnational | Moderate | 1,664 | Multiple groups | All | Convenience | Residual sera | 28.2% | Roche Diagnostics - IgG, IgM, IgA - Elecsys® Anti‐SARS‐CoV‐2 (N) | 1, 0.995 | Validated by manufacturer |

###### Americas region

| Country (Location) | Author (Organization) | Sampling Dates (YMD) | Geographic scope | Overall risk of bias | Sample size | Age | Sex | Sampling method | Sample frame | Seroprevalence (95% CI) | Test Manufacturers - Isotypes - Names | Test Sens, Spec | Sens, Spec Source |
| --- | --- | --- | --- | --- | --- | --- | --- | --- | --- | --- | --- | --- | --- |
| Brazil - HRP (Rio Grande do Sul) | Aluisio Barros (Universidade Federal de Pelotas)[131] | 2021-04-09 - 2021-04-11 | Subnational | Moderate | 4,499 | Multiple groups | All | Probability | Household and community samples | 18.2% | NA - IgG - Author designed (ELISA) -Spike | 0.95, 0.986 | Validated by developers |
| Brazil - HRP (São Paulo) | Nayara Vale (Universidade Federal de São Paulo)[132] | 2021-02-01 - 2021-02-28 | Subnational | Moderate | 940 | Adults (18-64 years) | All | Probability | Blood donors | 30.9% | Roche Diagnostics - IgG, IgM, IgA - Elecsys® Anti‐SARS‐CoV‐2 (N) | 1, 0.995 | Validated by manufacturer |
| Brazil - HRP (Rio Grande do Sul) | Aluisio Barros (Universidade Federal de Pelotas)[131] | 2021-02-05 - 2021-02-07 | Subnational | Moderate | 4,501 | Multiple groups | All | Probability | Household and community samples | 10% | NA - IgG - Author designed (ELISA) -Spike | 0.95, 0.986 | Validated by developers |
| Brazil - HRP (Greater São Paulo) | José Olimpio Moura de Albuquerque (Secretaria Municipal de Saude de Sao Paulo)[133] | 2021-02-02 - 2021-02-04 | Subnational | Moderate | 1,742 | Multiple groups | All | Probability | Household and community samples | 16% | Celer Technologies Inc. - IgG, IgM - One Step COVID-19 Test | 0.848, 0.99 | Validated by independent authors/third party/non-developers |
| Brazil - HRP (Greater São Paulo) | José Olimpio Moura de Albuquerque (Secretaria Municipal de Saude de Sao Paulo)[133] | 2021-01-19 - 2021-01-21 | Subnational | Moderate | 1,795 | Multiple groups | All | Probability | Household and community samples | 13.9% | Celer Technologies Inc. - IgG, IgM - One Step COVID-19 Test | 0.848, 0.99 | Validated by independent authors/third party/non-developers |
| Brazil - HRP (Greater São Paulo) | José Olimpio Moura de Albuquerque (Secretaria Municipal de Saude de Sao Paulo)[133] | 2021-01-05 - 2021-01-07 | Subnational | Moderate | 1,908 | Multiple groups | All | Probability | Household and community samples | 14.1% | Celer Technologies Inc. - IgG, IgM - One Step COVID-19 Test | 0.848, 0.99 | Validated by independent authors/third party/non-developers |
| Brazil - HRP (Maranhão) | Antonio da Silva (Universidade Federal do Maranhao)[134] | 2020-10-19 - 2020-10-30 | Subnational | Low | 4,563 | Multiple groups | All | Probability | Household and community samples | 38.1% | Roche Diagnostics - IgG, IgM, IgA - Elecsys® Anti‐SARS‐CoV‐2 (N) | 1, 0.995 | Validated by manufacturer |
| Brazil - HRP (São Paulo) | Nayara Vale (Universidade Federal de São Paulo)[132] | 2020-10-01 - 2020-10-31 | Subnational | Moderate | 940 | Adults (18-64 years) | All | Probability | Blood donors | 22% | Roche Diagnostics - IgG, IgM, IgA - Elecsys® Anti‐SARS‐CoV‐2 (N) | 1, 0.995 | Validated by manufacturer |
| Brazil - HRP (Vale do Rio Pardo) | Ana Schneider (Universidade de Santa Cruz do Sul)[135] | 2020-10-04 - 2020-10-17 | Subnational | Moderate | 1,063 | Multiple groups | All | Probability | Household and community samples | 3.4% | Beijing Lepu Medical Technology - IgG, IgM - Leccurate SARS-CoV-2 Antibody Rapid Test Kit (Colloidal Gold Immunochromatography) | 0.989, 0.976 | Validated by manufacturer |
| Brazil - HRP (Greater São Paulo) | José Olimpio Moura de Albuquerque (Secretaria Municipal de Saude de Sao Paulo)[133] | 2020-09-22 - 2020-09-24 | Subnational | Moderate | 2,012 | Multiple groups | All | Probability | Household and community samples | 13.6% | Celer Technologies Inc. - IgG, IgM - One Step COVID-19 Test | 0.848, 0.99 | Validated by independent authors/third party/non-developers |
| Brazil - HRP (Greater São Paulo) | José Olimpio Moura de Albuquerque (Secretaria Municipal de Saude de Sao Paulo)[133] | 2020-09-08 - 2020-09-10 | Subnational | Moderate | 2,125 | Multiple groups | All | Probability | Household and community samples | 11.9% | Celer Technologies Inc. - IgG, IgM - One Step COVID-19 Test | 0.848, 0.99 | Validated by independent authors/third party/non-developers |
| Brazil - HRP (Rio Grande do Sul) | Pedro C. Hallal (Universidade Federal de Pelotas )[136] | 2020-09-04 - 2020-09-06 | Subnational | Moderate | 4,500 | Multiple groups | All | Probability | Household and community samples | 1.6% | Guangzhou Wondfo Biotech Co. Ltd - IgG, IgM, IgA - Wondfo SARS-CoV-2 Antibody Test | 0.864, 0.996 | Validated by manufacturer |
| Brazil - HRP (Greater São Paulo) | José Olimpio Moura de Albuquerque (Secretaria Municipal de Saude de Sao Paulo)[133] | 2020-08-25 - 2020-08-27 | Subnational | Moderate | 2,225 | Multiple groups | All | Probability | Household and community samples | 13.9% | Celer Technologies Inc. - IgG, IgM - One Step COVID-19 Test | 0.848, 0.99 | Validated by independent authors/third party/non-developers |
| Brazil - HRP (Greater São Paulo) | José Olimpio Moura de Albuquerque (Secretaria Municipal de Saude de Sao Paulo)[133] | 2020-08-11 - 2020-08-13 | Subnational | Moderate | 2,447 | Multiple groups | All | Probability | Household and community samples | 11% | Celer Technologies Inc. - IgG, IgM - One Step COVID-19 Test | 0.848, 0.99 | Validated by independent authors/third party/non-developers |
| Brazil - HRP (Minas Gerais) | Daniel Chaves (Fundacao Hemominas)[137] | 2020-03-01 - 2020-12-31 | Subnational | Moderate | 7,837 | Multiple groups | All | Probability | Blood donors | 5.6% | Abbott Laboratories - IgG - Abbott Architect SARS-CoV-2 IgG | 0.99, 1 | Validated by manufacturer |
| Brazil - HRP (Greater São Paulo) | José Olimpio Moura de Albuquerque (Secretaria Municipal de Saude de Sao Paulo)[133] | 2020-07-28 - 2020-07-30 | Subnational | Moderate | 2,529 | Multiple groups | All | Probability | Household and community samples | 10.9% | Celer Technologies Inc. - IgG, IgM - One Step COVID-19 Test | 0.848, 0.99 | Validated by independent authors/third party/non-developers |
| Brazil - HRP (Greater São Paulo) | José Olimpio Moura de Albuquerque (Secretaria Municipal de Saude de Sao Paulo)[133] | 2020-07-13 - 2020-07-16 | Subnational | Moderate | 2,323 | Multiple groups | All | Probability | Household and community samples | 11.1% | Celer Technologies Inc. - IgG, IgM - One Step COVID-19 Test | 0.848, 0.99 | Validated by independent authors/third party/non-developers |
| Brazil - HRP (Greater São Paulo) | José Olimpio Moura de Albuquerque (Secretaria Municipal de Saude de Sao Paulo)[133] | 2020-06-29 - 2020-07-02 | Subnational | Moderate | 2,481 | Multiple groups | All | Probability | Household and community samples | 9.7% | Celer Technologies Inc. - IgG, IgM - One Step COVID-19 Test | 0.848, 0.99 | Validated by independent authors/third party/non-developers |
| Brazil - HRP (São Paulo) | Nayara Vale (Universidade Federal de São Paulo)[132] | 2020-06-01 - 2020-06-30 | Subnational | Moderate | 926 | Adults (18-64 years) | All | Probability | Blood donors | 11.4% | Roche Diagnostics - IgG, IgM, IgA - Elecsys® Anti‐SARS‐CoV‐2 (N) | 1, 0.995 | Validated by manufacturer |
| Brazil - HRP (Greater São Paulo) | José Olimpio Moura de Albuquerque (Secretaria Municipal de Saude de Sao Paulo)[133] | 2020-06-10 - 2020-06-17 | Subnational | Moderate | 2,645 | Multiple groups | All | Probability | Household and community samples | 9.5% | Celer Technologies Inc. - IgG, IgM - One Step COVID-19 Test | 0.848, 0.99 | Validated by independent authors/third party/non-developers |
| Brazil - HRP (Espirito Santo) | Ethel Leonor Noia Maciel (Laboratorio de Epidemiologia)[138] | 2020-05-10 - 2020-06-21 | Subnational | Moderate | 1,693 | Children and Youth (0-17 years) | All | Probability | Household and community samples | 6.1% | Celer Technologies Inc. - IgG, IgM - One Step COVID-19 Test | 0.864, 0.996 | Validated by independent authors/third party/non-developers |
| Colombia - HRP (Department of Cesar, Department of La Guajira) | Gustavo Concha (Medical Mission Institute)[139] | 2021-03-28 - 2021-04-26 | Subnational | Moderate | 452 | Multiple groups | All | Convenience | Household and community samples | 27.7% | PharmACT - IgG, IgM - BELTEST-IT COV-2 Rapid Test | 0.982, 0.997 | Validated by manufacturer |
| Colombia - HRP (Cordoba Department) | Evelin Garay (Colombia-Universidad de Cordoba)[140] | 2020-07-15 - 2020-11-15 | Subnational | Moderate | 2,564 | Multiple groups | All | Stratified non-probability | Household and community samples | 40.8% | Eurofins Ingenasa - IgG, IgM, IgA - INgezim COVID 19 DR | 0.983, 0.992 | Validated by manufacturer |
| Colombia - HRP (Cordoba) | Nelson Alvis Guzma (Universidad de Cordoba)[141] | 2020-03-15 - 2020-10-02 | Subnational | Moderate | 2,447 | Multiple groups | All | Probability | Household and community samples | 40.8% | Eurofins Ingenasa - IgG, IgM, IgA - INgezim COVID 19 DR | 0.983, 0.992 | Validated by manufacturer |
| Peru - HRP (Cusco Region) | Charles Huamani (Universidad Andina del Cusco)[142] | 2020-09-12 - 2020-09-27 | Subnational | Moderate | 1,924 | Multiple groups | All | Probability | Household and community samples | 33.1% | Roche Diagnostics - IgG, IgM, IgA - Elecsys® Anti‐SARS‐CoV‐2 (N) | 1, 0.995 | Validated by manufacturer |
| Canada (Quebec) | Antoine Lewin (Hema Quebec)[143] | 2021-01-25 - 2021-03-11 | Subnational | Moderate | 7,924 | Multiple groups | All | Sequential | Blood donors | 15% | NA - IgG, IgM - Author designed (ELISA) -Spike | 0.989, 0.985 | Validated by developers |
| Canada (Alberta) | Carmen Charlton (Alberta Precision Laboratories)[144] | 2021-01-11 - 2021-01-14 | Subnational | Moderate | 11,049 | Multiple groups | All | Sequential | Residual sera | 3.6% | Abbott Laboratories - IgG - Abbott Architect SARS-CoV-2 IgG,DiaSorin - IgG - Liaison SARS-CoV-2 S1/S2 IgG | 0.964, 1 | Complex test algorithm: Manual calculation |
| Canada (Alberta) | Carmen Charlton (Alberta Precision Laboratories)[144] | 2020-12-07 - 2020-12-10 | Subnational | Moderate | 12,076 | Multiple groups | All | Sequential | Residual sera | 2% | Abbott Laboratories - IgG - Abbott Architect SARS-CoV-2 IgG,DiaSorin - IgG - Liaison SARS-CoV-2 S1/S2 IgG | 0.964, 1 | Complex test algorithm: Manual calculation |
| Canada (Alberta) | Carmen Charlton (Alberta Precision Laboratories)[144] | 2020-11-09 - 2020-11-13 | Subnational | Moderate | 11,471 | Multiple groups | All | Sequential | Residual sera | 1.2% | Abbott Laboratories - IgG - Abbott Architect SARS-CoV-2 IgG,DiaSorin - IgG - Liaison SARS-CoV-2 S1/S2 IgG | 0.964, 1 | Complex test algorithm: Manual calculation |
| Canada (Ontario) | Shelly Bolotin, Vanessa Tran (Public Health Ontario)[145] | 2020-10-01 - 2020-10-30 | Subnational | Moderate | 7,107 | Multiple groups | All | Convenience | Residual sera | 1.2% | Abbott Laboratories - IgG - Abbott Architect SARS-CoV-2 IgG,Ortho Clinical Diagnostics Inc. - IgG - VITROS Immunodiagnostic Products Anti-SARS-CoV-2 IgG | 0.904, 1 | Complex test algorithm: Manual calculation |
| Canada (Alberta) | Carmen Charlton (Alberta Precision Laboratories)[144] | 2020-10-05 - 2020-10-09 | Subnational | Moderate | 13,490 | Multiple groups | All | Sequential | Residual sera | 0.9% | Abbott Laboratories - IgG - Abbott Architect SARS-CoV-2 IgG,DiaSorin - IgG - Liaison SARS-CoV-2 S1/S2 IgG | 0.964, 1 | Complex test algorithm: Manual calculation |
| Canada (Ontario) | Shelly Bolotin, Vanessa Tran (Public Health Ontario)[145] | 2020-09-03 - 2020-09-30 | Subnational | Moderate | 4,901 | Multiple groups | All | Convenience | Residual sera | 0.7% | Abbott Laboratories - IgG - Abbott Architect SARS-CoV-2 IgG,Ortho Clinical Diagnostics Inc. - IgG - VITROS Immunodiagnostic Products Anti-SARS-CoV-2 IgG | 0.904, 1 | Complex test algorithm: Manual calculation |
| Canada (Alberta) | Carmen Charlton (Alberta Precision Laboratories)[144] | 2020-09-07 - 2020-09-11 | Subnational | Moderate | 12,006 | Multiple groups | All | Sequential | Residual sera | 1% | Abbott Laboratories - IgG - Abbott Architect SARS-CoV-2 IgG,DiaSorin - IgG - Liaison SARS-CoV-2 S1/S2 IgG | 0.964, 1 | Complex test algorithm: Manual calculation |
| Canada (Ontario) | Shelly Bolotin, Vanessa Tran (Public Health Ontario)[145] | 2020-08-01 - 2020-08-31 | Subnational | Moderate | 6,789 | Multiple groups | All | Convenience | Residual sera | 1.1% | Abbott Laboratories - IgG - Abbott Architect SARS-CoV-2 IgG,Ortho Clinical Diagnostics Inc. - IgG - VITROS Immunodiagnostic Products Anti-SARS-CoV-2 IgG | 0.904, 1 | Complex test algorithm: Manual calculation |
| Canada (Alberta) | Carmen Charlton (Alberta Precision Laboratories)[144] | 2020-08-04 - 2020-08-11 | Subnational | Moderate | 12,284 | Multiple groups | All | Sequential | Residual sera | 0.7% | Abbott Laboratories - IgG - Abbott Architect SARS-CoV-2 IgG,DiaSorin - IgG - Liaison SARS-CoV-2 S1/S2 IgG | 0.964, 1 | Complex test algorithm: Manual calculation |
| Canada (Ontario) | Shelly Bolotin, Vanessa Tran (Public Health Ontario)[145] | 2020-07-04 - 2020-07-31 | Subnational | Moderate | 7,001 | Multiple groups | All | Convenience | Residual sera | 1.1% | Abbott Laboratories - IgG - Abbott Architect SARS-CoV-2 IgG,Ortho Clinical Diagnostics Inc. - IgG - VITROS Immunodiagnostic Products Anti-SARS-CoV-2 IgG | 0.904, 1 | Complex test algorithm: Manual calculation |
| Canada (Alberta) | Carmen Charlton (Alberta Precision Laboratories)[144] | 2020-07-06 - 2020-07-10 | Subnational | Moderate | 13,175 | Multiple groups | All | Sequential | Residual sera | 0.5% | Abbott Laboratories - IgG - Abbott Architect SARS-CoV-2 IgG,DiaSorin - IgG - Liaison SARS-CoV-2 S1/S2 IgG | 0.964, 1 | Complex test algorithm: Manual calculation |
| Canada (Ontario) | Shelly Bolotin, Vanessa Tran (Public Health Ontario)[145] | 2020-06-05 - 2020-06-30 | Subnational | Moderate | 7,014 | Multiple groups | All | Convenience | Residual sera | 1.1% | Abbott Laboratories - IgG - Abbott Architect SARS-CoV-2 IgG,Ortho Clinical Diagnostics Inc. - IgG - VITROS Immunodiagnostic Products Anti-SARS-CoV-2 IgG | 0.904, 1 | Complex test algorithm: Manual calculation |
| Canada (Alberta) | Carmen Charlton (Alberta Precision Laboratories)[144] | 2020-06-01 - 2020-06-05 | Subnational | Moderate | 8,442 | Multiple groups | All | Sequential | Residual sera | 0.9% | Abbott Laboratories - IgG - Abbott Architect SARS-CoV-2 IgG,DiaSorin - IgG - Liaison SARS-CoV-2 S1/S2 IgG | 0.964, 1 | Complex test algorithm: Manual calculation |
| Canada (Ontario) | Shelly Bolotin, Vanessa Tran (Public Health Ontario)[145] | 2020-05-26 - 2020-05-31 | Subnational | Moderate | 1,061 | Multiple groups | All | Convenience | Residual sera | 1.5% | Abbott Laboratories - IgG - Abbott Architect SARS-CoV-2 IgG,Ortho Clinical Diagnostics Inc. - IgG - VITROS Immunodiagnostic Products Anti-SARS-CoV-2 IgG | 0.904, 1 | Complex test algorithm: Manual calculation |
| Canada (British Columbia) | Danuta Skowronski (British Columbia Centre for Disease Control)[146] | 2020-05-15 - 2020-05-27 | Subnational | Moderate | 870 | Multiple groups | All | Probability | Residual sera | 0.8% | Ortho Clinical Diagnostics Inc. - IgG, IgM, IgA - VITROS Immunodiagnostic Products Anti-SARS-CoV-2 Total | 1, 1 | Validated by manufacturer |
| Canada (Ontario) | Shelly Bolotin, Vanessa Tran (Public Health Ontario)[145] | 2020-03-27 - 2020-04-30 | Subnational | Moderate | 827 | Multiple groups | All | Convenience | Residual sera | 0.5% | Abbott Laboratories - IgG - Abbott Architect SARS-CoV-2 IgG,Ortho Clinical Diagnostics Inc. - IgG - VITROS Immunodiagnostic Products Anti-SARS-CoV-2 IgG | 0.904, 1 | Complex test algorithm: Manual calculation |
| Canada (British Columbia) | Danuta Skowronski (British Columbia Centre for Disease Control)[146] | 2020-03-05 - 2020-03-13 | Subnational | Moderate | 870 | Multiple groups | All | Probability | Residual sera | 0.8% | Ortho Clinical Diagnostics Inc. - IgG, IgM, IgA - VITROS Immunodiagnostic Products Anti-SARS-CoV-2 Total | 1, 1 | Validated by manufacturer |
| Chile - HRP (Cities: Santiago (34 municipalities), Coquimbo-La serena (2 Municipalities) and Talca (1 Municipality)) | Ximena Aguilera (Universidad del Desarrollo, Centro de Epidemiología y Políticas de Salud)[147] | 2021-10-05 - 2021-11-25 | Subnational | Moderate | 2,198 | Multiple groups | All | Probability | Household and community samples | 97.3% (95% CI 96.6-98%) | Beijing Wantai Biological - IgG IgM IgA - Wantai Rapid Test for Total Antibody to SARS-CoV-2 | 0.947, 0.989 | Validated by manufacturer |
| United States of America (Texas) | The University of Texas Health Science Center at Houston (University of Texas Health Science Center at Houston)[148] | 2021-09-20 - 2021-10-04 | Subnational | Moderate | 88,091 | Multiple groups | All | Convenience | Household and community samples | 75.5% | Roche Diagnostics - IgG, IgM, IgA - Elecsys® Anti‐SARS‐CoV‐2 (S) | 0.988, 1 | Validated by manufacturer |
| United States of America (California) | Megha Mehrotra (California Department of Public Health)[149] | 2021-04-20 - 2021-08-01 | Subnational | Low | 7,483 | Multiple groups | All | Probability | Household and community samples | 67% | Enable Biosciences - IgG, IgM, IgA - Enable’s ADAP SARS-CoV-2 total antibody assay | 1, 1 | Validated by manufacturer |
| United States of America (California) | Megha Mehrotra (California Department of Public Health)[149] | 2021-04-20 - 2021-06-16 | Subnational | Low | 1,375 | Children and Youth (0-17 years) | All | Probability | Household and community samples | 41% | Enable Biosciences - IgG, IgM, IgA - Enable’s ADAP SARS-CoV-2 total antibody assay | 1, 1 | Validated by manufacturer |
| United States of America (California) | Alexia Couture (Centers for Disease Control and Prevention)[150] | 2021-05-01 - 2021-05-31 | Subnational | Moderate | 651 | Children and Youth (0-17 years) | All | Convenience | Residual sera | 43.7% | Roche Diagnostics - IgG, IgM, IgA - Elecsys® Anti‐SARS‐CoV‐2 (N) | 1, 0.995 | Validated by manufacturer |
| United States of America (Illinois) | Alexia Couture (Centers for Disease Control and Prevention)[150] | 2021-05-01 - 2021-05-31 | Subnational | Moderate | 942 | Children and Youth (0-17 years) | All | Convenience | Residual sera | 41.5% | Roche Diagnostics - IgG, IgM, IgA - Elecsys® Anti‐SARS‐CoV‐2 (N) | 1, 0.995 | Validated by manufacturer |
| United States of America (North Carolina) | Alexia Couture (Centers for Disease Control and Prevention)[150] | 2021-05-01 - 2021-05-31 | Subnational | Moderate | 648 | Children and Youth (0-17 years) | All | Convenience | Residual sera | 26.3% | Roche Diagnostics - IgG, IgM, IgA - Elecsys® Anti‐SARS‐CoV‐2 (N) | 1, 0.995 | Validated by manufacturer |
| United States of America (Ohio) | Alexia Couture (Centers for Disease Control and Prevention)[150] | 2021-05-01 - 2021-05-31 | Subnational | Moderate | 627 | Children and Youth (0-17 years) | All | Convenience | Residual sera | 37% | Roche Diagnostics - IgG, IgM, IgA - Elecsys® Anti‐SARS‐CoV‐2 (N) | 1, 0.995 | Validated by manufacturer |
| United States of America (South Carolina) | Alexia Couture (Centers for Disease Control and Prevention)[150] | 2021-05-01 - 2021-05-31 | Subnational | Moderate | 618 | Children and Youth (0-17 years) | All | Convenience | Residual sera | 34.8% | Roche Diagnostics - IgG, IgM, IgA - Elecsys® Anti‐SARS‐CoV‐2 (N) | 1, 0.995 | Validated by manufacturer |
| United States of America (California) | Marcus Wong (University of California)[151] | 2021-02-15 - 2021-04-15 | Subnational | Moderate | 4,641 | Multiple groups | All | Convenience | Household and community samples | 31.3% | Ortho Clinical Diagnostics Inc. - IgG, IgM, IgA - VITROS Immunodiagnostic Products Anti-SARS-CoV-2 Total,NA - - Author designed (ELISA) -Spike | N/A | Complex test algorithm: Manual review |
| United States of America (Arkansas) | Joshua Kennedy (University of Arkansas for Medical Sciences)[152] | 2020-11-07 - 2020-12-19 | Subnational | Moderate | 2,405 | Multiple groups | All | Convenience | Residual sera | 7.4% | Beckman Coulter - IgG - Access SARS-CoV-2 IgG Antibody Test,NA - - Author designed (ELISA) - Unknown | N/A | Complex test algorithm: Manual review |
| United States of America (Arkansas) | Victor M. Cardenas (University of Arkansas for Medical Sciences)[153] | 2020-11-01 - 2020-12-15 | Subnational | Moderate | 342 | Multiple groups | All | Probability | Household and community samples | 12.9% | Beckman Coulter - IgG - Access SARS-CoV-2 IgG Antibody Test | 1, 0.998 | Validated by manufacturer |
| United States of America (California) | Alice Pressman (Sutter Health)[154] | 2020-10-01 - 2020-12-31 | Subnational | Moderate | 4,500 | Adults (18-64 years) | Female | Convenience | Pregnant or parturient women | 8.1% | Siemens - IgG, IgM, IgA - Atellica® IM SARS-CoV-2 Total (COV2T) | 0.985, 0.998 | Validated by manufacturer |
| United States of America (California) | Marcus Wong (University of California)[151] | 2020-10-15 - 2020-12-15 | Subnational | Moderate | 5,308 | Multiple groups | All | Convenience | Household and community samples | 0.6% | Ortho Clinical Diagnostics Inc. - IgG, IgM, IgA - VITROS Immunodiagnostic Products Anti-SARS-CoV-2 Total | N/A | Complex test algorithm: Manual review |
| United States of America (Arkansas) | Joshua Kennedy (University of Arkansas for Medical Sciences)[152] | 2020-09-12 - 2020-10-24 | Subnational | Moderate | 2,098 | Multiple groups | All | Convenience | Residual sera | 4.1% | Beckman Coulter - IgG - Access SARS-CoV-2 IgG Antibody Test,NA - - Author designed (ELISA) - Unknown | N/A | Complex test algorithm: Manual review |
| United States of America (Virginia) | Rebecca Levorson (Inova Health System)[155] | 2020-07-31 - 2020-10-13 | Subnational | Moderate | 1,038 | Children and Youth (0-17 years) | All | Convenience | Multiple general populations | 8.5% | Ortho Clinical Diagnostics Inc. - IgG - VITROS Immunodiagnostic Products Anti-SARS-CoV-2 IgG | 1, 1 | Validated by manufacturer |
| United States of America (North Carolina) | Cesar Lopez (University of North Carolina)[156] | 2020-08-09 - 2020-10-03 | Subnational | Moderate | 2,633 | Multiple groups | All | Convenience | Residual sera | 10.8% | NA - IgG - Author designed (ELISA) -Spike | 0.98, 1 | Validated by developers |
| United States of America (Arkansas) | Joshua Kennedy (University of Arkansas for Medical Sciences)[152] | 2020-08-15 - 2020-09-05 | Subnational | Moderate | 1,301 | Multiple groups | All | Convenience | Residual sera | 2.6% | Beckman Coulter - IgG - Access SARS-CoV-2 IgG Antibody Test,NA - - Author designed (ELISA) - Unknown | N/A | Complex test algorithm: Manual review |
| United States of America (California) | Marcus Wong (University of California)[151] | 2020-07-15 - 2020-09-15 | Subnational | Moderate | 4,670 | Multiple groups | All | Convenience | Household and community samples | 0.6% | Ortho Clinical Diagnostics Inc. - IgG, IgM, IgA - VITROS Immunodiagnostic Products Anti-SARS-CoV-2 Total | 1, 1 | Validated by manufacturer |
| United States of America (California) | Cameron Adams (University of California, Berkeley)[157] | 2020-07-04 - 2020-09-01 | Subnational | Moderate | 4,760 | Multiple groups | All | Stratified non-probability | Household and community samples | 1% | Ortho Clinical Diagnostics Inc. - IgG, IgM, IgA - VITROS Immunodiagnostic Products Anti-SARS-CoV-2 Total | 1, 1 | Validated by manufacturer |
| United States of America (Colorado) | Kiersten Kugeler (Centers for Disease Control and Prevention)[158] | 2020-07-22 - 2020-08-08 | Subnational | Moderate | 1,598 | Multiple groups | All | Convenience | Residual sera | 6.8% | Ortho Clinical Diagnostics Inc. - IgG, IgM, IgA - VITROS Immunodiagnostic Products Anti-SARS-CoV-2 Total | 1, 1 | Validated by manufacturer |
| United States of America (Colorado) | Kiersten Kugeler (Centers for Disease Control and Prevention)[158] | 2020-07-22 - 2020-08-08 | Subnational | Moderate | 381 | Multiple groups | All | Probability | Household and community samples | 8% | Ortho Clinical Diagnostics Inc. - IgG, IgM, IgA - VITROS Immunodiagnostic Products Anti-SARS-CoV-2 Total | 1, 1 | Validated by manufacturer |
| United States of America (North Carolina) | Cesar Lopez (University of North Carolina)[156] | 2020-06-14 - 2020-08-08 | Subnational | Moderate | 3,528 | Multiple groups | All | Convenience | Residual sera | 10.5% | NA - IgG - Author designed (ELISA) -Spike | 0.98, 1 | Validated by developers |
| United States of America (Virginia) | Elizabeth T Rogawski McQuade (University of Virginia )[159] | 2020-06-01 - 2020-08-14 | Subnational | Moderate | 4,675 | Multiple groups | All | Convenience | Residual sera | 2.2% | Abbott Laboratories - IgG - Abbott Architect SARS-CoV-2 IgG | 0.99, 1 | Validated by manufacturer |
| United States of America (Connecticut) | Shiwani Mahajan (Yale School of Medicine)[160] | 2020-06-23 - 2020-07-22 | Subnational | Moderate | 148 | Multiple groups | All | Probability | Household and community samples | 6.4% | Ortho Clinical Diagnostics Inc. - IgG - VITROS Immunodiagnostic Products Anti-SARS-CoV-2 IgG | 1, 1 | Validated by manufacturer |
| United States of America (Connecticut) | Shiwani Mahajan (Yale School of Medicine)[160] | 2020-06-23 - 2020-07-22 | Subnational | Moderate | 171 | Multiple groups | All | Probability | Household and community samples | 19.9% | Ortho Clinical Diagnostics Inc. - IgG - VITROS Immunodiagnostic Products Anti-SARS-CoV-2 IgG | 1, 1 | Validated by manufacturer |
| United States of America (New York) | Lylach Haizler Cohen (Donald and Barbara Zucker School of Medicine)[161] | 2020-05-27 - 2020-07-24 | Subnational | Moderate | 1,671 | Adults (18-64 years) | Female | Convenience | Pregnant or parturient women | 16.1% | Roche Diagnostics - IgG, IgM, IgA - Elecsys® Anti‐SARS‐CoV‐2 (N) | 1, 0.995 | Validated by manufacturer |
| United States of America (Connecticut) | Shiwani Mahajan (Yale School of Medicine)[160] | 2020-06-04 - 2020-06-23 | Subnational | Moderate | 567 | Multiple groups | All | Probability | Household and community samples | 4% | Ortho Clinical Diagnostics Inc. - IgG - VITROS Immunodiagnostic Products Anti-SARS-CoV-2 IgG | 1, 1 | Validated by manufacturer |
| United States of America (Connecticut) | Travis Lim (Centers for Disease and Control )[162] | 2020-03-23 - 2020-08-14 | Subnational | Low | 8,929 | Multiple groups | All | Probability | Residual sera | 5.7% | NA - IgG, IgM, IgA - Author designed (ELISA) -Spike | 0.96, 0.993 | Validated by developers |
| United States of America (Florida) | Travis Lim (Centers for Disease and Control )[162] | 2020-03-23 - 2020-08-14 | Subnational | Low | 7,787 | Multiple groups | All | Probability | Residual sera | 4.8% | NA - IgG, IgM, IgA - Author designed (ELISA) -Spike | 0.96, 0.993 | Validated by developers |
| United States of America (Louisiana) | Travis Lim (Centers for Disease and Control )[162] | 2020-03-23 - 2020-08-14 | Subnational | Low | 2,849 | Multiple groups | All | Probability | Residual sera | 7.7% | NA - IgG, IgM, IgA - Author designed (ELISA) -Spike | 0.96, 0.993 | Validated by developers |
| United States of America (Minnesota) | Travis Lim (Centers for Disease and Control )[162] | 2020-03-23 - 2020-08-14 | Subnational | Low | 7,944 | Multiple groups | All | Probability | Residual sera | 4.9% | NA - IgG, IgM, IgA - Author designed (ELISA) -Spike | 0.96, 0.993 | Validated by developers |
| United States of America (Missouri) | Travis Lim (Centers for Disease and Control )[162] | 2020-03-23 - 2020-08-14 | Subnational | Low | 9,356 | Multiple groups | All | Probability | Residual sera | 2.5% | NA - IgG, IgM, IgA - Author designed (ELISA) -Spike | 0.96, 0.993 | Validated by developers |
| United States of America (Utah) | Travis Lim (Centers for Disease and Control )[162] | 2020-03-23 - 2020-08-14 | Subnational | Low | 8,769 | Multiple groups | All | Probability | Residual sera | 2.7% | NA - IgG, IgM, IgA - Author designed (ELISA) -Spike | 0.96, 0.993 | Validated by developers |
| United States of America (Washington) | Travis Lim (Centers for Disease and Control )[162] | 2020-03-23 - 2020-08-14 | Subnational | Low | 7,544 | Multiple groups | All | Probability | Residual sera | 2% | NA - IgG, IgM, IgA - Author designed (ELISA) -Spike | 0.96, 0.993 | Validated by developers |
| United States of America (Utah) | Matthew Samore (University of Utah)[163] | 2020-05-04 - 2020-06-30 | Subnational | Low | 8,108 | Multiple groups | All | Probability | Household and community samples | 1.1% | Abbott Laboratories - IgG - Abbott Architect SARS-CoV-2 IgG | 0.99, 1 | Validated by manufacturer |
| United States of America (Oregon) | Melissa Sutton (Oregon Health Authority)[164] | 2020-05-11 - 2020-06-15 | Subnational | Moderate | 897 | Multiple groups | All | Probability | Residual sera | 1% | Abbott Laboratories - IgG - Abbott Architect SARS-CoV-2 IgG | 0.99, 1 | Validated by manufacturer |
| United States of America (North Carolina) | Cesar Lopez (University of North Carolina)[156] | 2020-04-19 - 2020-06-13 | Subnational | Moderate | 3,463 | Multiple groups | All | Convenience | Residual sera | 5.3% | NA - IgG - Author designed (ELISA) -Spike | 0.98, 1 | Validated by developers |
| United States of America (Rhode Island) | Philip A Chan (Brown University)[165] | 2020-05-05 - 2020-05-22 | Subnational | Low | 1,032 | Multiple groups | All | Probability | Household and community samples | 2.9% | Abbott Laboratories - IgG - Abbott Architect SARS-CoV-2 IgG | 0.99, 1 | Validated by manufacturer |
| United States of America (Indiana) | Nir Menachemi (Indiana University Fairbanks School of Public Health)[166] | 2020-04-25 - 2020-04-29 | Subnational | Low | 3,518 | Multiple groups | All | Probability | Household and community samples | 1.1% | NA - IgG - Author designed (ELISA) -Unknown | 1, 0.996 |  |

###### Eastern Mediterranean region

| Country (Location) | Author (Organization) | Sampling Dates (YMD) | Geographic scope | Overall risk of bias | Sample size | Age | Sex | Sampling method | Sample frame | Seroprevalence (95% CI) | Test Manufacturers - Isotypes - Names | Test Sens, Spec | Sens, Spec Source |
| --- | --- | --- | --- | --- | --- | --- | --- | --- | --- | --- | --- | --- | --- |
| Iran (Islamic Republic of) - HRP (Guilan) | Maryam Shakiba (Tehran University of Medical Sciences)[167] | 2020-04-11 - 2020-04-19 | Subnational | Moderate | 528 | Multiple groups | All | Probability | Household and community samples | 22.1% | VivaChek Biotech (Hangzhou) Co. Ltd - IgG, IgM - VivaDiag COVID-19 IgM/IgG Rapid Test | 0.971, 1 | Validated by manufacturer |
| Iran (Islamic Republic of) - HRP (Mazandaran) | Seyed Abbas Mousavi (Mazandaran University of Medical Sciences)[168] | 2020-03-15 - 2020-05-15 | Subnational | Low | 1,588 | Multiple groups | All | Probability | Household and community samples | 17.3% | Pishtaz Diagnostics Iran - IgG, IgM - ELISA kit Pishtaz Teb Diagnostics | 0.941, 0.983 | Validated by manufacturer |
| occupied Palestinian territory, including east Jerusalem - HRP (West Bank) | Beesan Maraqa (An-Najah National University)[169] | 2020-11-01 - 2020-12-31 | Subnational | Moderate | 1,269 | Multiple groups | All | Probability | Residual sera | 24% | Roche Diagnostics - IgG, IgM, IgA - Elecsys® Anti‐SARS‐CoV‐2 (N) | 1, 0.995 | Validated by manufacturer |
| United Arab Emirates (Emirate of Abu Dhabi) | Ahmed R Alsuwaidi (Abu Dhabi Public Health Center)[170] | 2020-07-19 - 2020-08-14 | Subnational | Moderate | 8,831 | Multiple groups | All | Probability | Household and community samples | 11.3% | DiaSorin - IgG - Liaison SARS-CoV-2 S1/S2 IgG,Roche Diagnostics - IgG, IgM, IgA - Elecsys® Anti‐SARS‐CoV‐2 (N) | 0.974, 1 | Complex test algorithm: Manual calculation |

###### Europe region

| Country (Location) | Author (Organization) | Sampling Dates (YMD) | Geographic scope | Overall risk of bias | Sample size | Age | Sex | Sampling method | Sample frame | Seroprevalence (95% CI) | Test Manufacturers - Isotypes - Names | Test Sens, Spec | Sens, Spec Source |
| --- | --- | --- | --- | --- | --- | --- | --- | --- | --- | --- | --- | --- | --- |
| Bosnia and Herzegovina (Republic of Srpska) | Biljana Mijovic (University of Banja Luka)[171] | 2020-12-01 - 2021-01-15 | Subnational | Moderate | 1,855 | Multiple groups | All | Probability | Household and community samples | 40.3% | Beijing Wantai Biological - IgG, IgM, IgA - Wantai SARS-CoV-2 Total Ab ELISA | 0.967, 0.975 | Validated by manufacturer |
| Bosnia and Herzegovina (District Sarajevo) | Sanjin Musa (Institute for Public Health of the Federation of Bosnia and Herzegovina)[172] | 2020-11-02 - 2020-12-07 | Subnational | Moderate | 1,015 | Multiple groups | All | Probability | Blood donors | 19.1% (95% CI 16.6-21.6%) | Abbott Laboratories - IgG - Abbott Architect SARS-CoV-2 IgG,Abbott Laboratories - IgG - SARS-CoV-2 IgG II Quant | 0.984, 1 | Complex test algorithm: Manual calculation |
| Georgia (4 Districts:4 highly touristic districts of Georgia (Borjomi, Martvili, Khelvachauri and Kobuleti)) | Khatuna Zakhashvili (National Center for Disease Control and Public Health)[173] | 2020-12-01 - 2020-12-14 | Subnational | Moderate | 1,219 | Multiple groups | All | Probability | Household and community samples | 44.1% (95% CI 41.2-46.9%) | Beijing Wantai Biological - IgG IgM IgA - Wantai Rapid Test for Total Antibody to SARS-CoV-2 | 0.947, 0.989 | Validated by manufacturer |
| Georgia (4 Districts:4 highly touristic districts of Georgia (Borjomi, Martvili, Khelvachauri and Kobuleti)) | Khatuna Zakhashvili (National Center for Disease Control and Public Health)[173] | 2020-08-01 - 2020-08-14 | Subnational | Moderate | 1,218 | Multiple groups | All | Probability | Household and community samples | 0.5% (95% CI 0.2-1.1%) | Beijing Wantai Biological - IgG IgM IgA - Wantai Rapid Test for Total Antibody to SARS-CoV-2 | 0.947, 0.989 | Validated by manufacturer |
| Denmark (Capital Region and Region Zealand) | Maren J H Rytter (Slagelse Sygehus)[174] | 2020-06-22 - 2020-07-03 | Subnational | Moderate | 750 | Adults (18-64 years) | All | Probability | Blood donors | 2% | Zhuhai Livzon Diagnostics Inc - IgG, IgM - 2019-nCoV IgG/IgM Antibody Detection Kit | 0.906, 0.992 | Validated by manufacturer |
| Denmark (Capital Region of Denmark) | Kasper Iversen (University of Copenhagen)[175] | 2020-04-15 - 2020-04-22 | Subnational | Moderate | 4,672 | Adults (18-64 years) | All | Sequential | Blood donors | 3% | Zhuhai Livzon Diagnostics Inc - IgG, IgM - 2019-nCoV IgG/IgM Antibody Detection Kit | 0.906, 0.992 | Validated by manufacturer |
| Faroe Islands (Faroe Islands) | Maria Skaalum Petersen (University of the Faroe Islands)[176] | 2020-04-27 - 2020-05-01 | Subnational | Moderate | 1,075 | Adults (18-64 years) | All | Probability | Household and community samples | 0.6% | Beijing Wantai Biological - IgG, IgM, IgA - Wantai SARS-CoV-2 Total Ab ELISA | 0.967, 0.975 | Validated by manufacturer |
| France (Corsica) | Lisandru Capai (Université de Corse)[177] | 2021-02-01 - 2021-02-12 | Subnational | Moderate | 1,254 | Multiple groups | All | Convenience | Residual sera | 8.5% | EUROIMMUN - IgG - Anti-SARS-CoV-2 ELISA IgG | 0.9, 1 | Validated by manufacturer |
| Germany (North Rhine-Westphalia) | Insa Backhaus (Heinrich Heine University)[178] | 2020-11-02 - 2020-11-27 | Subnational | Moderate | 2,186 | Adults (18-64 years) | All | Probability | Household and community samples | 3.3% | Roche Diagnostics - IgG, IgM, IgA - Elecsys® Anti‐SARS‐CoV‐2 (N) | 1, 0.995 | Validated by manufacturer |
| Germany (Bavaria) | Otto Laub (University of Regensburg)[179] | 2020-05-22 - 2020-07-22 | Subnational | Moderate | 2,832 | Children and Youth (0-17 years) | All | Probability | Multiple general populations | 4.8% | NA - IgG - Author designed (ELISA) -Spike,Roche Diagnostics - IgG, IgM, IgA - Elecsys® Anti‐SARS‐CoV‐2 (N),NA - - Author designed (Neutralization Assay) | N/A | Complex test algorithm: Manual review |
| Germany (North Rhine-Westphalia, Lower Saxony, Hesse) | Bastian Fischer (Heart and Diabetes Center NRW)[180] | 2020-03-09 - 2020-06-03 | Subnational | Moderate | 3,186 | Multiple groups | All | Sequential | Residual sera | 0.9% | Abbott Laboratories - IgG - Abbott Architect SARS-CoV-2 IgG,EUROIMMUN - IgG - Anti-SARS-CoV-2 ELISA IgG,DiaSorin - IgG - Liaison SARS-CoV-2 S1/S2 IgG | N/A | Complex test algorithm: Manual review |
| Italy (Campania (Southern Italy)) | Federica Calo (University of Campania)[181] | 2020-06-01 - 2020-06-10 | Subnational | Moderate | 143 | Multiple groups | All | Sequential | Blood donors | 2.8% | Beijing Wantai Biological - IgG, IgM, IgA - Wantai SARS-CoV-2 Total Ab ELISA | 0.967, 0.975 | Validated by manufacturer |
| Italy (Trento) | Paola Stefanelli (Azienda Provinciale per i Servizi Sanitari)[182] | 2020-05-05 - 2020-05-15 | Subnational | Moderate | 6,075 | Multiple groups | All | Convenience | Household and community samples | 23.1% | Abbott Laboratories - IgG - Abbott Architect SARS-CoV-2 IgG | 0.99, 1 | Validated by manufacturer |
| Jersey (Jersey) | Statistics Jersey (Government of Jersey)[183] | 2020-04-29 - 2020-05-05 | Subnational | Low | 855 | Multiple groups | All | Probability | Household and community samples | 2.6% | Healgen - IgG, IgM - COVID-19 IgG/IgM Rapid Test Cassette | 0.938, 0.985 | Validated by manufacturer |
| Jersey (Jersey) | Statistics Jersey (Statistics Jersey)[184] | 2020-05-15 - 2020-05-27 | Subnational | Low | 1,062 | Multiple groups | All | Probability | Household and community samples | 4.4% | CTK Biotech, Inc. - IgG, IgM - OnSite COVID-19 IgG/IgM Rapid Test | 0.969, 0.994 | Validated by manufacturer |
| Spain (Catalonia) | Marianna Karachaliou (Barcelona Institute For Global Health)[185] | 2020-06-30 - 2020-11-15 | Subnational | Moderate | 4,740 | Multiple groups | All | Convenience | Household and community samples | 15.3% | NA - - Author designed (Multiplex) | N/A | Complex test algorithm: Manual review |
| Spain (Cantabria) | Paula Iruzubieta (University of Cantabria)[186] | 2020-04-27 - 2020-05-29 | Subnational | Moderate | 1,022 | Multiple groups | All | Probability | Household and community samples | 3.1% | Hangzhou Clongene Biotech Co. Ltd - IgG, IgM - Lungene 2019-nCoV IgG/IgM Rapid Test | 0.911, 0.965 | Validated by manufacturer |
| Switzerland (Ticino) | Ottavio Beretta (Ufficio del medico cantonale)[187] | 2021-05-03 - 2021-06-13 | Subnational | Moderate | 722 | Multiple groups | All | Probability | Household and community samples | 22.4% | Technogenetics - IgG, IgM - Rapid Test SARS-CoV-2 IgM/IgG Gold | 0.951, 0.989 | Validated by manufacturer |
| Switzerland (Canton of Fribourg) | Emna El May (Université de Fribourg)[188] | 2020-11-30 - 2021-02-05 | Subnational | Moderate | 449 | Multiple groups | All | Probability | Household and community samples | 19% | NA - - Author designed (Luminex) | 0.966, 0.997 | Validated by developers |
| Switzerland (Ticino) | Ottavio Beretta (Ufficio del medico cantonale)[187] | 2020-11-02 - 2020-12-27 | Subnational | Moderate | 870 | Multiple groups | All | Probability | Household and community samples | 14.4% | Technogenetics - IgG, IgM - Rapid Test SARS-CoV-2 IgM/IgG Gold | 0.951, 0.989 | Validated by manufacturer |
| Switzerland (Canton of Fribourg) | Daniela Anker (Université de Fribourg)[189] | 2020-07-08 - 2020-10-14 | Subnational | Moderate | 418 | Multiple groups | All | Probability | Household and community samples | 8% | NA - - Author designed (Luminex) | 0.966, 0.997 | Validated by developers |
| Switzerland (Ticino) | Ottavio Beretta (Ufficio del medico cantonale)[187] | 2020-08-03 - 2020-08-03 | Subnational | Moderate | 905 | Multiple groups | All | Probability | Household and community samples | 8.9% | Technogenetics - IgG, IgM - Rapid Test SARS-CoV-2 IgM/IgG Gold | 0.951, 0.989 | Validated by manufacturer |
| Switzerland (Canton of Zurich) | Erin West (University of Zurich)[190] | 2020-06-17 - 2020-09-14 | Subnational | Moderate | 472 | Adults (18-64 years) | All | Probability | Household and community samples | 4% | Swiss Federal Institute of Technology - IgG, IgA - Sensitive Anti-SARS-CoV-2 Spike Trimer Immunoglobulin Serological (SenASTrIS) | 0.966, 0.997 | Validated by manufacturers |
| Switzerland (Canton of Geneva) | Aude Richard (Geneva University Hospitals)[191] | 2020-06-01 - 2020-06-30 | Subnational | Low | 3,407 | Multiple groups | All | Probability | Household and community samples | 6.3% | EUROIMMUN - IgG - Anti-SARS-CoV-2 ELISA IgG | 0.9, 1 | Validated by manufacturer |
| Switzerland (Ticino) | Ottavio Beretta (Ufficio del medico cantonale)[187] | 2020-05-11 - 2020-06-28 | Subnational | Moderate | 934 | Multiple groups | All | Probability | Household and community samples | 9.6% | Technogenetics - IgG, IgM - Rapid Test SARS-CoV-2 IgM/IgG Gold | 0.951, 0.989 | Validated by manufacturer |
| Switzerland (Vaud) | Craig Fenwick (Lausanne University Hospital and University of Lausanne)[192] | 2020-05-04 - 2020-06-27 | Subnational | Moderate | 311 | Multiple groups | All | Probability | Household and community samples | 3.9% | Roche Diagnostics - IgG, IgM, IgA - Elecsys® Anti‐SARS‐CoV‐2 (N) | 1, 0.995 | Validated by manufacturer |
| Switzerland (Geneva) | Silvia Stringhini (Geneva University Hospitals)[193] | 2020-05-04 - 2020-05-09 | Subnational | Moderate | 775 | Multiple groups | All | Probability | Household and community samples | 10.8% | EUROIMMUN - IgG - Anti-SARS-CoV-2 ELISA IgG | 0.9, 1 | Validated by manufacturer |
| The United Kingdom (England) | Office for National Statistics (Office For National Statistics)[194] | 2021-02-04 - 2021-03-03 | Subnational | Low | 34,623 | Multiple groups | All | Probability | Household and community samples | 34.6% | NA - IgG - Author designed (ELISA) -Spike | 0.99, 0.991 | Validated by independent authors/third party/non-developers |
| The United Kingdom (Northern Ireland) | Office for National Statistics (Office For National Statistics)[194] | 2021-02-04 - 2021-03-03 | Subnational | Low | 632 | Multiple groups | All | Probability | Household and community samples | 31.2% | NA - IgG - Author designed (ELISA) -Spike | 0.99, 0.991 | Validated by independent authors/third party/non-developers |
| The United Kingdom (Scotland) | Office for National Statistics (Office For National Statistics)[194] | 2021-02-04 - 2021-03-03 | Subnational | Low | 2,737 | Multiple groups | All | Probability | Household and community samples | 22.3% | NA - IgG - Author designed (ELISA) -Spike | 0.99, 0.991 | Validated by independent authors/third party/non-developers |
| The United Kingdom (Wales) | Office for National Statistics (Office For National Statistics)[194] | 2021-02-04 - 2021-03-03 | Subnational | Low | 1,224 | Multiple groups | All | Probability | Household and community samples | 30.5% | NA - IgG - Author designed (ELISA) -Spike | 0.99, 0.991 | Validated by independent authors/third party/non-developers |
| The United Kingdom (England) | Office for National Statistics (Office of National Statistics)[195] | 2020-12-18 - 2021-01-14 | Subnational | Moderate | 24,150 | Multiple groups | All | Probability | Household and community samples | 15.3% | NA - IgG - Author designed (ELISA) -Spike | 0.99, 0.991 | Validated by independent authors/third party/non-developers |
| The United Kingdom (Northern Ireland) | Office for National Statistics (Office of National Statistics)[195] | 2020-12-18 - 2021-01-14 | Subnational | Moderate | 313 | Multiple groups | All | Probability | Household and community samples | 12% | NA - IgG - Author designed (ELISA) -Spike | 0.99, 0.991 | Validated by independent authors/third party/non-developers |
| The United Kingdom (Scotland) | Office for National Statistics (Office of National Statistics)[195] | 2020-12-18 - 2021-01-14 | Subnational | Moderate | 1,832 | Multiple groups | All | Probability | Household and community samples | 10.7% | NA - IgG - Author designed (ELISA) -Spike | 0.99, 0.991 | Validated by independent authors/third party/non-developers |
| The United Kingdom (Wales) | Office for National Statistics (Office of National Statistics)[195] | 2020-12-18 - 2021-01-14 | Subnational | Moderate | 732 | Multiple groups | All | Probability | Household and community samples | 11.5% | NA - IgG - Author designed (ELISA) -Spike | 0.99, 0.991 | Validated by independent authors/third party/non-developers |
| The United Kingdom (England) | Office for National Statistics (Office of National Statistics)[195] | 2020-11-20 - 2020-12-17 | Subnational | Moderate | 21,684 | Multiple groups | All | Probability | Household and community samples | 10.1% | NA - IgG - Author designed (ELISA) -Spike | 0.99, 0.991 | Validated by independent authors/third party/non-developers |
| The United Kingdom (Northern Ireland) | Office for National Statistics (Office of National Statistics)[195] | 2020-11-20 - 2020-12-17 | Subnational | Moderate | 248 | Multiple groups | All | Probability | Household and community samples | 3.7% | NA - IgG - Author designed (ELISA) -Spike | 0.99, 0.991 | Validated by independent authors/third party/non-developers |
| The United Kingdom (Scotland) | Office for National Statistics (Office of National Statistics)[195] | 2020-11-20 - 2020-12-17 | Subnational | Moderate | 1,034 | Multiple groups | All | Probability | Household and community samples | 5.9% | NA - IgG - Author designed (ELISA) -Spike | 0.99, 0.991 | Validated by independent authors/third party/non-developers |
| The United Kingdom (Wales) | Office for National Statistics (Office of National Statistics)[195] | 2020-11-20 - 2020-12-17 | Subnational | Moderate | 631 | Multiple groups | All | Probability | Household and community samples | 7.2% | NA - IgG - Author designed (ELISA) -Spike | 0.99, 0.991 | Validated by independent authors/third party/non-developers |
| The United Kingdom (Scotland) | Office for National Statistics (Office of National Statistics)[195] | 2020-10-23 - 2020-11-19 | Subnational | Moderate | 397 | Multiple groups | All | Probability | Household and community samples | 7.8% | NA - IgG - Author designed (ELISA) -Spike | 0.99, 0.991 | Validated by independent authors/third party/non-developers |
| The United Kingdom (Northern Ireland) | Office for National Statistics (Office of National Statistics)[195] | 2020-10-23 - 2020-11-19 | Subnational | Moderate | 333 | Multiple groups | All | Probability | Household and community samples | 3.5% | NA - IgG - Author designed (ELISA) -Spike | 0.99, 0.991 | Validated by independent authors/third party/non-developers |
| The United Kingdom (Wales) | Office for National Statistics (Office of National Statistics)[195] | 2020-10-23 - 2020-11-19 | Subnational | Moderate | 497 | Multiple groups | All | Probability | Household and community samples | 6.1% | NA - IgG - Author designed (ELISA) -Spike | 0.99, 0.991 | Validated by independent authors/third party/non-developers |
| The United Kingdom (England) | Office for National Statistics (Office of National Statistics)[195] | 2020-10-23 - 2020-11-19 | Subnational | Moderate | 15,834 | Multiple groups | All | Probability | Household and community samples | 8.7% | NA - IgG - Author designed (ELISA) -Spike | 0.99, 0.991 | Validated by independent authors/third party/non-developers |
| The United Kingdom (England) | Office for National Statistics (Office of National Statistics)[195] | 2020-09-25 - 2020-10-22 | Subnational | Moderate | 15,687 | Multiple groups | All | Probability | Household and community samples | 6.9% | NA - IgG - Author designed (ELISA) -Spike | 0.99, 0.991 | Validated by independent authors/third party/non-developers |
| The United Kingdom (Northern Ireland) | Office for National Statistics (Office of National Statistics)[195] | 2020-09-25 - 2020-10-22 | Subnational | Moderate | 249 | Multiple groups | All | Probability | Household and community samples | 2.8% | NA - IgG - Author designed (ELISA) -Spike | 0.99, 0.991 | Validated by independent authors/third party/non-developers |
| The United Kingdom (Scotland) | Office for National Statistics (Office of National Statistics)[195] | 2020-09-25 - 2020-10-22 | Subnational | Moderate | 383 | Multiple groups | All | Probability | Household and community samples | 6.8% | NA - IgG - Author designed (ELISA) -Spike | 0.99, 0.991 | Validated by independent authors/third party/non-developers |
| The United Kingdom (Wales) | Office for National Statistics (Office of National Statistics)[195] | 2020-09-25 - 2020-10-22 | Subnational | Moderate | 473 | Multiple groups | All | Probability | Household and community samples | 4.7% | NA - IgG - Author designed (ELISA) -Spike | 0.99, 0.991 | Validated by independent authors/third party/non-developers |
| The United Kingdom (Wales) | Office for National Statistics (Office of National Statistics)[195] | 2020-08-28 - 2020-09-24 | Subnational | Moderate | 236 | Multiple groups | All | Probability | Household and community samples | 4.7% | NA - IgG - Author designed (ELISA) -Spike | 0.99, 0.991 | Validated by independent authors/third party/non-developers |
| The United Kingdom (England) | Office for National Statistics (Office of National Statistics)[195] | 2020-08-28 - 2020-09-24 | Subnational | Moderate | 8,742 | Multiple groups | All | Probability | Household and community samples | 5.8% | NA - IgG - Author designed (ELISA) -Spike | 0.99, 0.991 | Validated by independent authors/third party/non-developers |
| The United Kingdom (England) | Office for National Statistics (Office of National Statistics)[195] | 2020-07-31 - 2020-08-27 | Subnational | Moderate | 6,189 | Multiple groups | All | Probability | Household and community samples | 5.6% | NA - IgG - Author designed (ELISA) -Spike | 0.99, 0.991 | Validated by independent authors/third party/non-developers |
| The United Kingdom (England) | Office for National Statistics (Office of National Statistics)[195] | 2020-07-03 - 2020-07-30 | Subnational | Moderate | 3,236 | Multiple groups | All | Probability | Household and community samples | 6.5% | NA - IgG - Author designed (ELISA) -Spike | 0.99, 0.991 | Validated by independent authors/third party/non-developers |
| The United Kingdom (England) | Office for National Statistics (Office of National Statistics)[195] | 2020-06-05 - 2020-07-02 | Subnational | Moderate | 2,392 | Multiple groups | All | Probability | Household and community samples | 4.2% | NA - - Author designed (ELISA) -Spike | 0.99, 0.991 | Validated by independent authors/third party/non-developers |
| The United Kingdom (England) | Office for National Statistics (Office of National Statistics)[195] | 2020-05-08 - 2020-06-04 | Subnational | Low | 1,575 | Multiple groups | All | Probability | Household and community samples | 6.3% | NA - IgG - Author designed (ELISA) -Spike | 0.99, 0.991 | Validated by independent authors/third party/non-developers |
| The United Kingdom (England) | Public Health England (Public Health England)[111] | 2020-04-24 - 2020-04-26 | Subnational | Low | 865 | Adults (18-64 years) | All | Probability | Blood donors | 5% | EUROIMMUN - IgG - Anti-SARS-CoV-2 ELISA IgG | 0.9, 1 | Validated by manufacturer |
| The United Kingdom (England) | Public Health England (Public Health England)[111] | 2020-04-23 - 2020-04-24 | Subnational | Low | 1,043 | Adults (18-64 years) | All | Probability | Blood donors | 7.3% | EUROIMMUN - IgG - Anti-SARS-CoV-2 ELISA IgG | 0.9, 1 | Validated by manufacturer |
| The United Kingdom (England) | Public Health England (Public Health England)[111] | 2020-04-15 - 2020-04-20 | Subnational | Low | 936 | Adults (18-64 years) | All | Probability | Blood donors | 6.2% | EUROIMMUN - IgG - Anti-SARS-CoV-2 ELISA IgG | 0.9, 1 | Validated by manufacturer |
| The United Kingdom (Scotland) | Craig P Thompson (University of Oxford)[196] | 2020-03-17 - 2020-05-18 | Subnational | Moderate | 3,500 | Adults (18-64 years) | All | Probability | Blood donors | 3.2% | NA - - Author designed (Neutralization Assay) | 0.94, 1 | Validated by developers |
| The United Kingdom (England) | Public Health England (Public Health England)[111] | 2020-04-14 - 2020-04-16 | Subnational | Low | 1,017 | Adults (18-64 years) | All | Probability | Blood donors | 4.6% | EUROIMMUN - IgG - Anti-SARS-CoV-2 ELISA IgG | 0.9, 1 | Validated by manufacturer |
| The United Kingdom (England) | Public Health England (Public Health England)[111] | 2020-04-09 - 2020-04-13 | Subnational | Low | 1,085 | Adults (18-64 years) | All | Probability | Blood donors | 11.1% | EUROIMMUN - IgG - Anti-SARS-CoV-2 ELISA IgG | 0.9, 1 | Validated by manufacturer |
| The United Kingdom (England) | Public Health England (Public Health England)[111] | 2020-04-02 - 2020-04-03 | Subnational | Low | 916 | Adults (18-64 years) | All | Probability | Blood donors | 2.4% | EUROIMMUN - IgG - Anti-SARS-CoV-2 ELISA IgG | 0.9, 1 | Validated by manufacturer |
| The United Kingdom (England) | Public Health England (Public Health England)[111] | 2020-03-26 - 2020-03-27 | Subnational | Low | 757 | Adults (18-64 years) | All | Probability | Blood donors | 2.6% | EUROIMMUN - IgG - Anti-SARS-CoV-2 ELISA IgG | 0.9, 1 | Validated by manufacturer |

###### South-East Asia region

| Country (Location) | Author (Organization) | Sampling Dates (YMD) | Geographic scope | Overall risk of bias | Sample size | Age | Sex | Sampling method | Sample frame | Seroprevalence (95% CI) | Test Manufacturers - Isotypes - Names | Test Sens, Spec | Sens, Spec Source |
| --- | --- | --- | --- | --- | --- | --- | --- | --- | --- | --- | --- | --- | --- |
| India (Delhi) | Pragya Sharma (Maulana Azad Medical College)[197] | 2021-09-24 - 2021-10-14 | Subnational | Low | 27,811 | Multiple groups | All | Probability | Household and community samples | 88% | Ortho Clinical Diagnostics Inc. - IgG - VITROS Immunodiagnostic Products Anti-SARS-CoV-2 IgG | 1, 1 | Validated by manufacturer |
| India (Kerala) | Government of Kerala (Government of Kerala Department of Health)[198] | 2021-09-01 - 2021-09-30 | Subnational | Moderate | 1,459 | Children and Youth (0-17 years) | All | Probability | Household and community samples | 40.2% | Siemens - IgG, IgM, IgA - Atellica® IM SARS-CoV-2 Total (COV2T),Abbott Laboratories - IgG - Abbott Architect SARS-CoV-2 IgG | 1, 0.998 | Complex test algorithm: Manual calculation |
| India (Kerala) | Government of Kerala (Government of Kerala Department of Health)[198] | 2021-09-01 - 2021-09-30 | Subnational | Moderate | 1,476 | Multiple groups | All | Probability | Household and community samples | 87.7% | Siemens - IgG, IgM, IgA - Atellica® IM SARS-CoV-2 Total (COV2T),Abbott Laboratories - IgG - Abbott Architect SARS-CoV-2 IgG | 1, 0.998 | Complex test algorithm: Manual calculation |
| India (Kerala) | Government of Kerala (Government of Kerala Department of Health)[198] | 2021-09-01 - 2021-09-30 | Subnational | Moderate | 4,429 | Multiple groups | All | Probability | Household and community samples | 82.6% | Siemens - IgG, IgM, IgA - Atellica® IM SARS-CoV-2 Total (COV2T),Abbott Laboratories - IgG - Abbott Architect SARS-CoV-2 IgG | 1, 0.998 | Complex test algorithm: Manual calculation |
| India (Kerala) | Government of Kerala (Government of Kerala Department of Health)[198] | 2021-09-01 - 2021-09-30 | Subnational | Moderate | 1,706 | Multiple groups | All | Probability | Persons living in slums | 85.3% | Siemens - IgG, IgM, IgA - Atellica® IM SARS-CoV-2 Total (COV2T),Abbott Laboratories - IgG - Abbott Architect SARS-CoV-2 IgG | 1, 0.998 | Complex test algorithm: Manual calculation |
| India (Kerala) | Government of Kerala (Government of Kerala Department of Health)[198] | 2021-09-01 - 2021-09-30 | Subnational | Moderate | 1,521 | Multiple groups | All | Probability | Household and community samples | 78.2% | Siemens - IgG, IgM, IgA - Atellica® IM SARS-CoV-2 Total (COV2T),Abbott Laboratories - IgG - Abbott Architect SARS-CoV-2 IgG | 1, 0.998 | Complex test algorithm: Manual calculation |
| India (Kerala) | Government of Kerala (Government of Kerala Department of Health)[198] | 2021-08-15 - 2021-09-15 | Subnational | Moderate | 2,274 | Adults (18-64 years) | Female | Convenience | Pregnant or parturient women | 65.4% | Siemens - IgG, IgM, IgA - Atellica® IM SARS-CoV-2 Total (COV2T),Abbott Laboratories - IgG - Abbott Architect SARS-CoV-2 IgG | 1, 0.998 | Complex test algorithm: Manual calculation |
| India (1. National Capital Region (New Delhi & Faridabad) 2.Gorakhpur, Uttar Pradesh 3. Bhubaneshwar, Odisha 4. Pondicherry 5.Agartala, Tripura) | Dr Puneet Misra (All India Institute of Medical Sciences, New Delhi)[199] | 2021-03-15 - 2021-06-17 | Subnational | Moderate | 5,123 | Multiple groups | All | Probability | Household and community samples | 64.4% (95% CI 63.1-65.7%) | Beijing Wantai Biological - IgG IgM IgA - Wantai SARS-CoV-2 Total Ab ELISA | 0.967, 0.975 | Validated by manufacturer |
| India (Delhi) | Nandini Sharma (Maulana Azad Medical College)[200] | 2021-01-11 - 2021-01-22 | Subnational | Low | 27,436 | Multiple groups | All | Probability | Household and community samples | 50.5% | Ortho Clinical Diagnostics Inc. - IgG - VITROS Immunodiagnostic Products Anti-SARS-CoV-2 IgG | 1, 1 | Validated by manufacturer |
| India (Kashmir) | Muhammad Khan (Government Medical College Srinagar)[201] | 2020-10-17 - 2020-11-04 | Subnational | Low | 6,230 | Multiple groups | All | Probability | Household and community samples | 36.9% | Abbott Laboratories - IgG - Abbott Architect SARS-CoV-2 IgG | 0.99, 1 | Validated by manufacturer |
| India (NCT Delhi) | Nandini Sharma (Government of National Capital Territory of Delhi)[202] | 2020-10-15 - 2020-10-21 | Subnational | Low | 14,905 | Multiple groups | All | Probability | Household and community samples | 25.2% | Calbiotech Inc. - IgG - ErbaLisa COVID-19 IgG ELISA Kit | 0.983, 0.981 | Validated by manufacturer |
| India (Puducherry) | Sitanshu Sekhar Kar (Jawaharlal Institute)[203] | 2020-10-12 - 2020-10-16 | Subnational | Moderate | 900 | Multiple groups | All | Probability | Household and community samples | 34.5% | Roche Diagnostics - IgG, IgM, IgA - Elecsys® Anti‐SARS‐CoV‐2 (N) | 1, 0.995 | Validated by manufacturer |
| India (Puducherry) | Sitanshu Sekhar Kar (Jawaharlal Institute)[203] | 2020-09-10 - 2020-09-16 | Subnational | Moderate | 898 | Multiple groups | All | Probability | Household and community samples | 20.7% | Roche Diagnostics - IgG, IgM, IgA - Elecsys® Anti‐SARS‐CoV‐2 (N) | 1, 0.995 | Validated by manufacturer |
| India (Uttar Pradesh) | Vasanthakumar Namasivayam (Government of Uttar Pradesh)[204] | 2020-09-04 - 2020-09-10 | Subnational | Low | 16,012 | Multiple groups | All | Probability | Household and community samples | 22.1% | Zydus Diagnostics - IgG - Covid Kavach™ Anti-SARS CoV-2 IgG Antibody Detection ELISA | 0.987, 1 | Validated by manufacturer |
| India (NCT Delhi) | Nandini Sharma (Government of National Capital Territory of Delhi)[202] | 2020-09-01 - 2020-09-07 | Subnational | Low | 16,953 | Multiple groups | All | Probability | Household and community samples | 24.6% | Calbiotech Inc. - IgG - ErbaLisa COVID-19 IgG ELISA Kit | 0.983, 0.981 | Validated by manufacturer |
| India (Puducherry) | Sitanshu Sekhar Kar (Jawaharlal Institute)[203] | 2020-08-11 - 2020-08-16 | Subnational | Moderate | 869 | Multiple groups | All | Probability | Household and community samples | 4.9% | Roche Diagnostics - IgG, IgM, IgA - Elecsys® Anti‐SARS‐CoV‐2 (N) | 1, 0.995 | Validated by manufacturer |
| India (NCT Delhi) | Nandini Sharma (Government of National Capital Territory of Delhi)[202] | 2020-08-01 - 2020-08-07 | Subnational | Low | 14,779 | Multiple groups | All | Probability | Household and community samples | 28.8% | J. Mitra & Co. Pvt. Ltd - IgG - COVID Kawach IgG Microlisa | 0.921, 0.977 | Validated by developers |
| India (West Bengal) | Parthasarathi Satpati (Medical College Kolkata)[205] | 2020-07-26 - 2020-08-08 | Subnational | Moderate | 458 | Multiple groups | All | Probability | Household and community samples | 4.2% | Calbiotech Inc. - IgG - ErbaLisa COVID-19 IgG ELISA Kit | 0.983, 0.981 | Validated by manufacturer |
| India (Srinagar) | S Muhammad Salim Khan (Government Medical College Srinagar)[206] | 2020-07-01 - 2020-07-15 | Subnational | Moderate | 2,906 | Multiple groups | All | Convenience | Residual sera | 3.6% | Abbott Laboratories - IgG - Abbott Architect SARS-CoV-2 IgG | 0.99, 1 | Validated by manufacturer |
| India (NCT Delhi) | Ministry of Health and Family Welfare (Indian Center for Disease Control)[207] | 2020-06-27 - 2020-07-10 | Subnational | Moderate | 21,387 | Multiple groups | All | Probability | Household and community samples | 23.5% | Zydus Diagnostics - IgG - Covid Kavach™ Anti-SARS CoV-2 IgG Antibody Detection ELISA | 0.987, 1 | Validated by manufacturer |
| Indonesia (East Java) | Ni Luh Ayu Megasari (Kobe University)[208] | 2020-06-15 - 2020-12-15 | Subnational | Moderate | 1,819 | Multiple groups | All | Convenience | Household and community samples | 11.4% | Nanjing Vazyme Medical Technology Co. Ltd - IgG, IgM - 2019-Novel Coronavirus (2019-nCoV) IgG/IgM Detection Kit | 0.967, 0.688 | Validated by manufacturer |

###### Western Pacific region

| Country (Location) | Author (Organization) | Sampling Dates (YMD) | Geographic scope | Overall risk of bias | Sample size | Age | Sex | Sampling method | Sample frame | Seroprevalence (95% CI) | Test Manufacturers - Isotypes - Names | Test Sens, Spec | Sens, Spec Source |
| --- | --- | --- | --- | --- | --- | --- | --- | --- | --- | --- | --- | --- | --- |
| China (Hong Kong) | Lin-Lei Chen (The University of Hong Kong)[209] | 2021-11-15 - 2021-12-15 | Subnational | Moderate | 300 | Multiple groups | All | Convenience | Residual sera | 30.3% | Shenzhen Yhlo Biotech Co. Ltd - IgG - iFlash-SARS-CoV-2 S1 IgG | 0.9, 0.98 | Validated by manufacturer |
| China (Wuhan) | Zhenyu He (Chinese Academy of Medical Sciences & Peking Union Medical College)[210] | 2020-04-14 - 2020-04-15 | Subnational | Low | 9,542 | Multiple groups | All | Probability | Household and community samples | 6.9% | Roche Diagnostics - IgG, IgM, IgA - Elecsys® Anti‐SARS‐CoV‐2 (N) | 1, 0.995 | Validated by manufacturer |
| Japan (Kanagawa Prefecture) | Atsushi Goto (Yokohama City University)[211] | 2022-01-30 - 2022-02-28 | Subnational | Low | 1,277 | Multiple groups | All | Probability | Household and community samples | 94.2% | TOSOH - IgG - AIA-CL SARS-CoV-2 SP-IgG | 1, 1 | Validated by manufacturer |
| Japan (Hiroshima Prefecture) | Prefecture of Hiroshima (Hiroshima University )[212] | 2020-10-15 - 2020-11-15 | Subnational | Moderate | 2,396 | Multiple groups | All | Probability | Household and community samples | 0.2% | Roche Diagnostics - IgG, IgM, IgA - Elecsys® Anti‐SARS‐CoV‐2 (N) | 1, 0.995 | Validated by manufacturer |
| Malaysia (Selangor) | I-Ching Sam (University of Malaya)[213] | 2020-12-15 - 2021-04-15 | Subnational | Moderate | 653 | Multiple groups | All | Convenience | Residual sera | 4.1% | Beijing Wantai Biological - IgG, IgM, IgA - Wantai SARS-CoV-2 Total Ab ELISA | 0.967, 0.975 | Validated by manufacturer |

##### Table S7. Additional studies used in descriptive analysis.

###### Africa region

| Country (Location) | Author (Organization) | Sampling Dates (YMD) | Geographic scope | Overall risk of bias | Sample size | Age | Sex | Sampling method | Sample frame | Seroprevalence (95% CI) | Test Manufacturers - Isotypes - Names | Test Sens, Spec | Sens, Spec Source |
| --- | --- | --- | --- | --- | --- | --- | --- | --- | --- | --- | --- | --- | --- |
| Angola - HRP (Luanda) | Cruz Sebastiao (Instituto Nacional de Investigação em Saúde)[214] | 2020-09-01 - 2020-09-30 | Local | High | 55 | Adults (18-64 years) | All | Convenience | Blood donors | 20% | Biomerieux - IgG, IgM, IgA - VIDAS Multiparametric immunoassay system for medium throughput | 1, 1 | Validated by manufacturer |
| Burkina Faso - HRP (Ouagadougou & Bobo-Dioulasso) | Dr Isidore Traore (National Institute of Public Health)[215] | 2021-04-03 - 2021-04-16 | Local | Moderate | 5,240 | Multiple groups | All | Probability | Household and community samples | 67.8% (95% CI 66.5-69.1%) | Beijing Wantai Biological - IgG IgM IgA - Wantai SARS-CoV-2 Total Ab ELISA | 0.967, 0.975 | Validated by manufacturer |
| Cameroon - HRP (Yaoundé) | Kene Nwosu (Hopital Central de Yaounde)[216] | 2020-10-14 - 2020-11-26 | Local | Low | 971 | Multiple groups | All | Probability | Household and community samples | 31.3% | Abbott Laboratories - IgG, IgM - Panbio COVID-19 IgG/IgM rapid test device | 0.978, 0.928 | Validated by manufacturer |
| Central African Republic - HRP (Bangui, the capital of the Central African Republic) | Alexandre Manirakiza (Institut Pasteur of Bangui)[217] | 2021-07-12 - 2021-08-20 | Local | Moderate | 799 | Multiple groups | All | Probability | Household and community samples | 75.5% (95% CI 72.3-78.4%) | Beijing Wantai Biological - IgG IgM IgA - Wantai SARS-CoV-2 Total Ab ELISA | 0.967, 0.975 | Validated by manufacturer |
| Congo - HRP | Novy Bonguili (Laboratoire National de Santé Publique)[218] | 2020-01-01 - 2020-02-15 | National | High | 484 | Multiple groups | All | Convenience | Representative patient population | 5% | NA - IgG, IgM - Author designed (IFA) - NON-MULTIPLEXED | 1, 0.975 | Validated by independent authors/third party/non-developers |
| Democratic Republic of the Congo - HRP (Kinshasa) | Antoine Nkuba (Institut National de Recherche Biomédicale)[219] | 2020-10-22 - 2020-11-08 | Local | Low | 1,080 | Multiple groups | All | Probability | Household and community samples | 16.6% | NA - - Author designed (Luminex) | 1, 0.997 | Validated by developers |
| Ethiopia - HRP (Addis Ababa) | Esayas Gudina (University of Munich)[220] | 2021-04-08 - 2021-04-10 | Local | High | 176 | Multiple groups | All | Convenience | Household and community samples | 72.7% | Roche Diagnostics - IgG, IgM, IgA - Elecsys® Anti‐SARS‐CoV‐2 (N) | 1, 0.995 | Validated by manufacturer |
| Ethiopia - HRP (Addis Ababa) | Esayas Gudina (University of Munich)[220] | 2021-04-01 - 2021-04-03 | Local | High | 188 | Multiple groups | All | Convenience | Household and community samples | 54.8% | Roche Diagnostics - IgG, IgM, IgA - Elecsys® Anti‐SARS‐CoV‐2 (N) | 1, 0.995 | Validated by manufacturer |
| Ethiopia - HRP (Jimma) | Esayas Gudina (University of Munich)[220] | 2021-02-04 - 2021-03-16 | Local | High | 100 | Multiple groups | All | Convenience | Household and community samples | 31% | Roche Diagnostics - IgG, IgM, IgA - Elecsys® Anti‐SARS‐CoV‐2 (N) | 1, 0.995 | Validated by manufacturer |
| Ethiopia - HRP (Jimma) | Esayas Gudina (University of Munich)[220] | 2021-02-02 - 2021-03-15 | Local | High | 166 | Multiple groups | All | Convenience | Household and community samples | 45.2% | Roche Diagnostics - IgG, IgM, IgA - Elecsys® Anti‐SARS‐CoV‐2 (N) | 1, 0.995 | Validated by manufacturer |
| Ethiopia - HRP (Addis Ababa) | Esayas Gudina (University of Munich)[220] | 2021-02-11 - 2021-02-26 | Local | High | 151 | Multiple groups | All | Convenience | Household and community samples | 58.2% | Roche Diagnostics - IgG, IgM, IgA - Elecsys® Anti‐SARS‐CoV‐2 (N) | 1, 0.995 | Validated by manufacturer |
| Ethiopia - HRP (Addis Ababa) | Esayas Gudina (University of Munich)[220] | 2021-02-01 - 2021-03-07 | Local | High | 149 | Multiple groups | All | Convenience | Household and community samples | 45.7% | Roche Diagnostics - IgG, IgM, IgA - Elecsys® Anti‐SARS‐CoV‐2 (N) | 1, 0.995 | Validated by manufacturer |
| Ethiopia - HRP (Jimma) | Esayas Gudina (University of Munich)[220] | 2021-01-21 - 2021-02-25 | Local | High | 133 | Multiple groups | All | Convenience | Household and community samples | 26.3% | Roche Diagnostics - IgG, IgM, IgA - Elecsys® Anti‐SARS‐CoV‐2 (N) | 1, 0.995 | Validated by manufacturer |
| Ethiopia - HRP (Jimma) | Esayas Gudina (University of Munich)[220] | 2021-01-02 - 2021-02-04 | Local | High | 191 | Multiple groups | All | Convenience | Household and community samples | 40.8% | Roche Diagnostics - IgG, IgM, IgA - Elecsys® Anti‐SARS‐CoV‐2 (N) | 1, 0.995 | Validated by manufacturer |
| Ethiopia - HRP (Addis Ababa) | Esayas Gudina (University of Munich)[220] | 2021-01-02 - 2021-01-22 | Local | High | 218 | Multiple groups | All | Convenience | Household and community samples | 54.2% | Roche Diagnostics - IgG, IgM, IgA - Elecsys® Anti‐SARS‐CoV‐2 (N) | 1, 0.995 | Validated by manufacturer |
| Ethiopia - HRP (Addis Ababa) | Esayas Gudina (University of Munich)[220] | 2020-12-05 - 2021-02-04 | Local | High | 224 | Multiple groups | All | Convenience | Household and community samples | 39.7% | Roche Diagnostics - IgG, IgM, IgA - Elecsys® Anti‐SARS‐CoV‐2 (N) | 1, 0.995 | Validated by manufacturer |
| Ethiopia - HRP (Jimma) | Esayas Gudina (University of Munich)[220] | 2020-12-01 - 2021-02-01 | Local | High | 297 | Multiple groups | All | Convenience | Household and community samples | 32.3% | Roche Diagnostics - IgG, IgM, IgA - Elecsys® Anti‐SARS‐CoV‐2 (N) | 1, 0.995 | Validated by manufacturer |
| Ethiopia - HRP (Jimma) | Esayas Gudina (University of Munich)[220] | 2020-12-03 - 2021-01-27 | Local | High | 238 | Multiple groups | All | Convenience | Household and community samples | 18% | Roche Diagnostics - IgG, IgM, IgA - Elecsys® Anti‐SARS‐CoV‐2 (N) | 1, 0.995 | Validated by manufacturer |
| Ethiopia - HRP (Addis Ababa) | Saro Abdella (Ethiopian Public Health Institute)[221] | 2020-07-22 - 2020-09-02 | Local | Low | 956 | Multiple groups | All | Probability | Household and community samples | 4.7% | Core Technology Co. Ltd - IgG, IgM - Coretests COVID-19 IgM/IgG Ab Test | 0.939, 0.981 | Validated by manufacturer |
| Ethiopia - HRP (Jimma) | Saro Abdella (Ethiopian Public Health Institute)[221] | 2020-07-22 - 2020-09-02 | Local | Low | 900 | Multiple groups | All | Probability | Household and community samples | 1.2% | Core Technology Co. Ltd - IgG, IgM - Coretests COVID-19 IgM/IgG Ab Test | 0.939, 0.981 | Validated by manufacturer |
| Ethiopia - HRP (Dire Dawa city) | Tamrat Shaweno (Jimma University Institute of Health)[222] | 2020-06-15 - 2020-07-30 | Local | Moderate | 684 | Multiple groups | All | Probability | Household and community samples | 3.2% | Abbott Laboratories - IgG - Abbott Architect SARS-CoV-2 IgG | 0.99, 1 | Validated by manufacturer |
| Ethiopia - HRP (Addis Ababa) | John Kempen (Kuwait University)[223] | 2020-05-18 - 2020-05-21 | Local | High | 99 | Multiple groups | All | Convenience | Residual sera | 3% | Abbott Laboratories - IgG - Abbott Architect SARS-CoV-2 IgG | 0.99, 1 | Validated by manufacturer |
| Gabon (Libreville) | Amandine Mveang Nzoghe (Centre Hospitalier Universitaire (CHU) ‐ Mère‐ Enfant)[224] | 2020-07-15 - 2020-10-15 | Local | High | 1,495 | Multiple groups | All | Convenience | Residual sera | 36.2% | Biomerieux - IgG, IgM, IgA - VIDAS Multiparametric immunoassay system for medium throughput,Roche Diagnostics - IgG, IgM, IgA - Elecsys® Anti‐SARS‐CoV‐2 (N) | 1, 0.995 | Complex test algorithm: Manual calculation |
| Kenya - HRP (Busia) | R Lucinde (KEMRI-Wellcome Trust Research Programme)[225] | 2021-09-20 - 2021-10-22 | Local | High | 297 | Adults (18-64 years) | Female | Sequential | Pregnant or parturient women | 65.7% | NA - - Author designed (ELISA) -Spike | 0.927, 0.99 | Validated by developers |
| Kenya - HRP (Kilifi) | R Lucinde (KEMRI-Wellcome Trust Research Programme)[225] | 2021-09-01 - 2021-10-31 | Local | High | 305 | Adults (18-64 years) | Female | Sequential | Pregnant or parturient women | 58.4% | NA - - Author designed (ELISA) -Spike | 0.927, 0.99 | Validated by developers |
| Kenya - HRP (Nairobi) | R Lucinde (KEMRI-Wellcome Trust Research Programme)[225] | 2021-09-07 - 2021-10-19 | Local | High | 245 | Adults (18-64 years) | Female | Sequential | Pregnant or parturient women | 78.8% | NA - - Author designed (ELISA) -Spike | 0.927, 0.99 | Validated by developers |
| Kenya - HRP | James Nyagwange (KEMRI-Wellcome Trust Research Programme)[226] | 2021-08-01 - 2021-08-31 | National | High | 176 | Adults (18-64 years) | All | Convenience | Blood donors | 83% | Beijing Wantai Biological - IgG, IgM, IgA - Wantai SARS-CoV-2 Total Ab ELISA | 0.967, 0.975 | Validated by manufacturer |
| Kenya - HRP (Kilifi) | R Lucinde (KEMRI-Wellcome Trust Research Programme)[225] | 2021-07-01 - 2021-08-31 | Local | High | 260 | Adults (18-64 years) | Female | Sequential | Pregnant or parturient women | 56.9% | NA - - Author designed (ELISA) -Spike | 0.927, 0.99 | Validated by developers |
| Kenya - HRP (Kilifi) | R Lucinde (KEMRI-Wellcome Trust Research Programme)[225] | 2021-05-01 - 2021-06-30 | Local | High | 382 | Adults (18-64 years) | Female | Sequential | Pregnant or parturient women | 27.2% | NA - - Author designed (ELISA) -Spike | 0.927, 0.99 | Validated by developers |
| Kenya - HRP (Kisumu) | Anthony Etyang (KEMRI-Wellcome Trust Research Programme)[227] | 2021-05-01 - 2021-05-31 | Local | Low | 217 | Multiple groups | All | Probability | Household and community samples | 39.2% | NA - - Author designed (ELISA) -Spike | 0.927, 0.99 | Validated by independent authors/third party/non-developers |
| Kenya - HRP (Nairobi) | Anthony Etyang (KEMRI-Wellcome Trust Research Programme)[227] | 2021-05-01 - 2021-05-31 | Local | Low | 113 | Multiple groups | All | Probability | Household and community samples | 47.8% | NA - - Author designed (ELISA) -Spike | 0.927, 0.99 | Validated by independent authors/third party/non-developers |
| Kenya - HRP (Busia) | R Lucinde (KEMRI-Wellcome Trust Research Programme)[225] | 2021-04-15 - 2021-05-21 | Local | High | 270 | Adults (18-64 years) | Female | Sequential | Pregnant or parturient women | 28.9% | NA - - Author designed (ELISA) -Spike | 0.927, 0.99 | Validated by developers |
| Kenya - HRP (Kilifi) | Anthony Etyang (KEMRI-Wellcome Trust Research Programme)[227] | 2021-04-01 - 2021-04-30 | Local | Low | 145 | Multiple groups | All | Probability | Household and community samples | 24.1% | NA - - Author designed (ELISA) -Spike | 0.927, 0.99 | Validated by independent authors/third party/non-developers |
| Kenya - HRP (Kilifi) | R Lucinde (KEMRI-Wellcome Trust Research Programme)[225] | 2021-03-01 - 2021-04-30 | Local | High | 260 | Adults (18-64 years) | Female | Sequential | Pregnant or parturient women | 21.1% | NA - - Author designed (ELISA) -Spike | 0.927, 0.99 | Validated by developers |
| Kenya - HRP (Nairobi) | R Lucinde (KEMRI-Wellcome Trust Research Programme)[225] | 2021-01-27 - 2021-03-11 | Local | High | 265 | Adults (18-64 years) | Female | Sequential | Pregnant or parturient women | 30.2% | NA - - Author designed (ELISA) -Spike | 0.927, 0.99 | Validated by developers |
| Kenya - HRP (Nairobi) | Patrick Munywoki (Kenya Medical Research Institute)[228] | 2020-11-27 - 2020-12-05 | Local | Moderate | 511 | Multiple groups | All | Probability | Household and community samples | 43.3% | Beijing Wantai Biological - IgG, IgM, IgA - Wantai SARS-CoV-2 Total Ab ELISA | 0.967, 0.975 | Validated by manufacturer |
| Kenya - HRP (Kilifi) | R Lucinde (KEMRI-Wellcome Trust Research Programme)[225] | 2020-11-01 - 2020-12-31 | Local | High | 236 | Adults (18-64 years) | Female | Sequential | Pregnant or parturient women | 13.6% | NA - - Author designed (ELISA) -Spike | 0.927, 0.99 | Validated by developers |
| Kenya - HRP (Kilifi) | R Lucinde (KEMRI-Wellcome Trust)[229] | 2020-11-01 - 2020-11-24 | Local | High | 154 | Adults (18-64 years) | Female | Sequential | Pregnant or parturient women | 10.4% | NA - IgG - Author designed (ELISA) -Spike | 0.927, 0.99 | Validated by independent authors/third party/non-developers |
| Kenya - HRP | Isaac Ngere (Washington State University)[230] | 2020-11-02 - 2020-11-23 | Local | Low | 1,164 | Multiple groups | All | Probability | Household and community samples | 32.7% | Beijing Wantai Biological - IgG, IgM, IgA - Wantai SARS-CoV-2 Total Ab ELISA | 0.967, 0.975 | Validated by manufacturer |
| Kenya - HRP (Kilifi) | R Lucinde (KEMRI-Wellcome Trust)[229] | 2020-10-01 - 2020-10-31 | Local | High | 183 | Adults (18-64 years) | Female | Sequential | Pregnant or parturient women | 1.6% | NA - IgG - Author designed (ELISA) -Spike | 0.927, 0.99 | Validated by independent authors/third party/non-developers |
| Kenya - HRP (Kilifi) | R Lucinde (KEMRI-Wellcome Trust)[229] | 2020-09-18 - 2020-09-30 | Local | High | 82 | Adults (18-64 years) | Female | Sequential | Pregnant or parturient women | 0% | NA - IgG - Author designed (ELISA) -Spike | 0.927, 0.99 | Validated by independent authors/third party/non-developers |
| Kenya - HRP (Nairobi) | R Lucinde (KEMRI-Wellcome Trust)[229] | 2020-07-30 - 2020-08-25 | Local | High | 196 | Adults (18-64 years) | Female | Sequential | Pregnant or parturient women | 46.4% | NA - IgG - Author designed (ELISA) -Spike | 0.927, 0.99 | Validated by independent authors/third party/non-developers |
| Kenya - HRP | James Nyagwange (KEMRI-Wellcome Trust Research Programme)[226] | 2020-05-01 - 2020-05-31 | National | High | 176 | Adults (18-64 years) | All | Convenience | Blood donors | 2.3% | Beijing Wantai Biological - IgG, IgM, IgA - Wantai SARS-CoV-2 Total Ab ELISA | 0.967, 0.975 | Validated by manufacturer |
| Kenya - HRP (Western Region) | Trevor Crowell (Walter Reed Army Institute of Research)[231] | 2020-01-01 - 2020-03-19 | Subnational | High | 582 | Adults (18-64 years) | All | Convenience | Residual sera | 3.3% | Bio-rad - IgG, IgM, IgA - Platelia SARS-CoV-2 Total Ab assay | 0.993, 0.98 | Validated by manufacturer |
| Madagascar (Antananarivo) | Solohery Razafimahatratra (Institut Pasteur de Madagascar)[232] | 2021-05-01 - 2021-05-26 | Local | High | 500 | Adults (18-64 years) | All | Sequential | Blood donors | 64.8% | Beijing Wantai Biological - IgG, IgM, IgA - Wantai SARS-CoV-2 Total Ab ELISA | 0.967, 0.975 | Validated by manufacturer |
| Madagascar (Antananarivo) | Solohery Razafimahatratra (Institut Pasteur de Madagascar)[232] | 2021-04-01 - 2021-04-30 | Local | High | 493 | Adults (18-64 years) | All | Sequential | Blood donors | 58.4% | Beijing Wantai Biological - IgG, IgM, IgA - Wantai SARS-CoV-2 Total Ab ELISA | 0.967, 0.975 | Validated by manufacturer |
| Madagascar (Antananarivo) | Solohery Razafimahatratra (Institut Pasteur de Madagascar)[232] | 2021-03-01 - 2021-03-31 | Local | High | 497 | Adults (18-64 years) | All | Sequential | Blood donors | 49.9% | Beijing Wantai Biological - IgG, IgM, IgA - Wantai SARS-CoV-2 Total Ab ELISA | 0.967, 0.975 | Validated by manufacturer |
| Madagascar (Antananarivo) | Solohery Razafimahatratra (Institut Pasteur de Madagascar)[232] | 2021-02-01 - 2021-02-28 | Local | High | 319 | Adults (18-64 years) | All | Sequential | Blood donors | 50.5% | Beijing Wantai Biological - IgG, IgM, IgA - Wantai SARS-CoV-2 Total Ab ELISA | 0.967, 0.975 | Validated by manufacturer |
| Madagascar (Antananarivo) | Solohery Razafimahatratra (Institut Pasteur de Madagascar)[232] | 2021-01-01 - 2021-01-31 | Local | High | 389 | Adults (18-64 years) | All | Sequential | Blood donors | 44.7% | Beijing Wantai Biological - IgG, IgM, IgA - Wantai SARS-CoV-2 Total Ab ELISA | 0.967, 0.975 | Validated by manufacturer |
| Madagascar (Antananarivo) | Solohery Razafimahatratra (Institut Pasteur de Madagascar)[232] | 2020-12-01 - 2020-12-31 | Local | High | 177 | Adults (18-64 years) | All | Sequential | Blood donors | 22% | ID.Vet - IgG - ID Screen | 0.952, 0.998 | Validated by manufacturer |
| Madagascar (Antananarivo) | Matthieu Schoenhals (Institut Pasteur de Madagascar)[233] | 2020-11-01 - 2020-11-30 | Local | High | 502 | Adults (18-64 years) | All | Convenience | Blood donors | 31.3% | ID.Vet - IgG - ID Screen | 0.952, 0.998 | Validated by manufacturer |
| Madagascar (Fianarantsoa) | Matthieu Schoenhals (Institut Pasteur de Madagascar)[233] | 2020-11-01 - 2020-11-30 | Local | High | 172 | Adults (18-64 years) | All | Convenience | Blood donors | 12.8% | ID.Vet - IgG - ID Screen | 0.952, 0.998 | Validated by manufacturer |
| Madagascar (Mahajanga) | Matthieu Schoenhals (Institut Pasteur de Madagascar)[233] | 2020-11-01 - 2020-11-30 | Local | High | 30 | Adults (18-64 years) | All | Convenience | Blood donors | 16.7% | ID.Vet - IgG - ID Screen | 0.952, 0.998 | Validated by manufacturer |
| Madagascar (Toamasina) | Matthieu Schoenhals (Institut Pasteur de Madagascar)[233] | 2020-11-01 - 2020-11-30 | Local | High | 282 | Adults (18-64 years) | All | Convenience | Blood donors | 18.4% | ID.Vet - IgG - ID Screen | 0.952, 0.998 | Validated by manufacturer |
| Madagascar (Toliara) | Matthieu Schoenhals (Institut Pasteur de Madagascar)[233] | 2020-11-01 - 2020-11-30 | Local | High | 199 | Adults (18-64 years) | All | Convenience | Blood donors | 18.1% | ID.Vet - IgG - ID Screen | 0.952, 0.998 | Validated by manufacturer |
| Madagascar (Antananarivo) | Solohery Razafimahatratra (Institut Pasteur de Madagascar)[232] | 2020-11-01 - 2020-11-30 | Local | High | 503 | Adults (18-64 years) | All | Sequential | Blood donors | 31.2% | ID.Vet - IgG - ID Screen | 0.952, 0.998 | Validated by manufacturer |
| Madagascar (Antananarivo) | Matthieu Schoenhals (Institut Pasteur de Madagascar)[233] | 2020-10-01 - 2020-10-31 | Local | High | 502 | Adults (18-64 years) | All | Convenience | Blood donors | 36% | ID.Vet - IgG - ID Screen | 0.952, 0.998 | Validated by manufacturer |
| Madagascar (Fianarantsoa) | Matthieu Schoenhals (Institut Pasteur de Madagascar)[233] | 2020-10-01 - 2020-10-31 | Local | High | 152 | Adults (18-64 years) | All | Convenience | Blood donors | 15.1% | ID.Vet - IgG - ID Screen | 0.952, 0.998 | Validated by manufacturer |
| Madagascar (Mahajanga) | Matthieu Schoenhals (Institut Pasteur de Madagascar)[233] | 2020-10-01 - 2020-10-31 | Local | High | 26 | Adults (18-64 years) | All | Convenience | Blood donors | 26.9% | ID.Vet - IgG - ID Screen | 0.952, 0.998 | Validated by manufacturer |
| Madagascar (Toamasina) | Matthieu Schoenhals (Institut Pasteur de Madagascar)[233] | 2020-10-01 - 2020-10-31 | Local | High | 199 | Adults (18-64 years) | All | Convenience | Blood donors | 26.1% | ID.Vet - IgG - ID Screen | 0.952, 0.998 | Validated by manufacturer |
| Madagascar (Toliara) | Matthieu Schoenhals (Institut Pasteur de Madagascar)[233] | 2020-10-01 - 2020-10-31 | Local | High | 139 | Adults (18-64 years) | All | Convenience | Blood donors | 22.3% | ID.Vet - IgG - ID Screen | 0.952, 0.998 | Validated by manufacturer |
| Madagascar (Antananarivo) | Solohery Razafimahatratra (Institut Pasteur de Madagascar)[232] | 2020-10-01 - 2020-10-31 | Local | High | 497 | Adults (18-64 years) | All | Sequential | Blood donors | 35.8% | ID.Vet - IgG - ID Screen | 0.952, 0.998 | Validated by manufacturer |
| Madagascar (Antananarivo) | Matthieu Schoenhals (Institut Pasteur de Madagascar)[233] | 2020-09-01 - 2020-09-30 | Local | High | 481 | Adults (18-64 years) | All | Convenience | Blood donors | 38.5% | ID.Vet - IgG - ID Screen | 0.952, 0.998 | Validated by manufacturer |
| Madagascar (Fianarantsoa) | Matthieu Schoenhals (Institut Pasteur de Madagascar)[233] | 2020-09-01 - 2020-09-30 | Local | High | 173 | Adults (18-64 years) | All | Convenience | Blood donors | 18.5% | ID.Vet - IgG - ID Screen | 0.952, 0.998 | Validated by manufacturer |
| Madagascar (Mahajanga) | Matthieu Schoenhals (Institut Pasteur de Madagascar)[233] | 2020-09-01 - 2020-09-30 | Local | High | 53 | Adults (18-64 years) | All | Convenience | Blood donors | 37.7% | ID.Vet - IgG - ID Screen | 0.952, 0.998 | Validated by manufacturer |
| Madagascar (Toamasina) | Matthieu Schoenhals (Institut Pasteur de Madagascar)[233] | 2020-09-01 - 2020-09-30 | Local | High | 213 | Adults (18-64 years) | All | Convenience | Blood donors | 29.1% | ID.Vet - IgG - ID Screen | 0.952, 0.998 | Validated by manufacturer |
| Madagascar (Toliara) | Matthieu Schoenhals (Institut Pasteur de Madagascar)[233] | 2020-09-01 - 2020-09-30 | Local | High | 185 | Adults (18-64 years) | All | Convenience | Blood donors | 20.5% | ID.Vet - IgG - ID Screen | 0.952, 0.998 | Validated by manufacturer |
| Madagascar (Fianarantsoa) | Matthieu Schoenhals (Institut Pasteur de Madagascar)[233] | 2020-08-01 - 2020-08-31 | Local | High | 120 | Adults (18-64 years) | All | Convenience | Blood donors | 9.2% | ID.Vet - IgG - ID Screen | 0.952, 0.998 | Validated by manufacturer |
| Madagascar (Mahajanga) | Matthieu Schoenhals (Institut Pasteur de Madagascar)[233] | 2020-08-01 - 2020-08-31 | Local | High | 107 | Adults (18-64 years) | All | Convenience | Blood donors | 35.5% | ID.Vet - IgG - ID Screen | 0.952, 0.998 | Validated by manufacturer |
| Madagascar (Toamasina) | Matthieu Schoenhals (Institut Pasteur de Madagascar)[233] | 2020-08-01 - 2020-08-31 | Local | High | 227 | Adults (18-64 years) | All | Convenience | Blood donors | 40.1% | ID.Vet - IgG - ID Screen | 0.952, 0.998 | Validated by manufacturer |
| Madagascar (Toliara) | Matthieu Schoenhals (Institut Pasteur de Madagascar)[233] | 2020-08-01 - 2020-08-31 | Local | High | 167 | Adults (18-64 years) | All | Convenience | Blood donors | 7.2% | ID.Vet - IgG - ID Screen | 0.952, 0.998 | Validated by manufacturer |
| Madagascar (Antananarivo) | Matthieu Schoenhals (Institut Pasteur de Madagascar)[233] | 2020-08-01 - 2020-08-30 | Local | High | 432 | Adults (18-64 years) | All | Convenience | Blood donors | 35.2% | ID.Vet - IgG - ID Screen | 0.952, 0.998 | Validated by manufacturer |
| Madagascar (Antananarivo) | Matthieu Schoenhals (Institut Pasteur de Madagascar)[233] | 2020-07-01 - 2020-07-31 | Local | High | 428 | Adults (18-64 years) | All | Convenience | Blood donors | 29% | ID.Vet - IgG - ID Screen | 0.952, 0.998 | Validated by manufacturer |
| Madagascar (Mahajanga) | Matthieu Schoenhals (Institut Pasteur de Madagascar)[233] | 2020-07-01 - 2020-07-31 | Local | High | 98 | Adults (18-64 years) | All | Convenience | Blood donors | 1% | ID.Vet - IgG - ID Screen | 0.952, 0.998 | Validated by manufacturer |
| Madagascar (Toamasina) | Matthieu Schoenhals (Institut Pasteur de Madagascar)[233] | 2020-07-01 - 2020-07-31 | Local | High | 182 | Adults (18-64 years) | All | Convenience | Blood donors | 43.4% | ID.Vet - IgG - ID Screen | 0.952, 0.998 | Validated by manufacturer |
| Madagascar (Toliara) | Matthieu Schoenhals (Institut Pasteur de Madagascar)[233] | 2020-07-01 - 2020-07-31 | Local | High | 147 | Adults (18-64 years) | All | Convenience | Blood donors | 0.7% | ID.Vet - IgG - ID Screen | 0.952, 0.998 | Validated by manufacturer |
| Madagascar (Fianarantsoa) | Matthieu Schoenhals (Institut Pasteur de Madagascar)[233] | 2020-07-01 - 2020-07-30 | Local | High | 142 | Adults (18-64 years) | All | Convenience | Blood donors | 0% | ID.Vet - IgG - ID Screen | 0.952, 0.998 | Validated by manufacturer |
| Madagascar (Antananarivo) | Matthieu Schoenhals (Institut Pasteur de Madagascar)[233] | 2020-06-01 - 2020-06-30 | Local | High | 308 | Adults (18-64 years) | All | Convenience | Blood donors | 3.3% | ID.Vet - IgG - ID Screen | 0.952, 0.998 | Validated by manufacturer |
| Madagascar (Fianarantsoa) | Matthieu Schoenhals (Institut Pasteur de Madagascar)[233] | 2020-06-01 - 2020-06-30 | Local | High | 129 | Adults (18-64 years) | All | Convenience | Blood donors | 0.8% | ID.Vet - IgG - ID Screen | 0.952, 0.998 | Validated by manufacturer |
| Madagascar (Mahajanga) | Matthieu Schoenhals (Institut Pasteur de Madagascar)[233] | 2020-06-01 - 2020-06-30 | Local | High | 50 | Adults (18-64 years) | All | Convenience | Blood donors | 0% | ID.Vet - IgG - ID Screen | 0.952, 0.998 | Validated by manufacturer |
| Madagascar (Toamasina) | Matthieu Schoenhals (Institut Pasteur de Madagascar)[233] | 2020-06-01 - 2020-06-30 | Local | High | 82 | Adults (18-64 years) | All | Convenience | Blood donors | 39% | ID.Vet - IgG - ID Screen | 0.952, 0.998 | Validated by manufacturer |
| Madagascar (Toliara) | Matthieu Schoenhals (Institut Pasteur de Madagascar)[233] | 2020-06-01 - 2020-06-30 | Local | High | 87 | Adults (18-64 years) | All | Convenience | Blood donors | 1.2% | ID.Vet - IgG - ID Screen | 0.952, 0.998 | Validated by manufacturer |
| Madagascar (Antananarivo) | Matthieu Schoenhals (Institut Pasteur de Madagascar)[233] | 2020-05-01 - 2020-05-31 | Local | High | 598 | Adults (18-64 years) | All | Convenience | Blood donors | 0.2% | ID.Vet - IgG - ID Screen | 0.952, 0.998 | Validated by manufacturer |
| Madagascar (Antananarivo) | Matthieu Schoenhals (Institut Pasteur de Madagascar)[233] | 2020-04-01 - 2020-04-30 | Local | High | 157 | Adults (18-64 years) | All | Convenience | Blood donors | 0% | ID.Vet - IgG - ID Screen | 0.952, 0.998 | Validated by manufacturer |
| Madagascar (Antananarivo) | Matthieu Schoenhals (Institut Pasteur de Madagascar)[233] | 2020-03-01 - 2020-03-30 | Local | High | 496 | Adults (18-64 years) | All | Convenience | Blood donors | 0.8% | ID.Vet - IgG - ID Screen | 0.952, 0.998 | Validated by manufacturer |
| Mali - HRP (Bancoumana) | Issaka Sagara (National Institutes of Health)[234] | 2020-12-28 - 2021-01-29 | Local | Moderate | 904 | Multiple groups | All | Convenience | Household and community samples | 35.5% | NA - - Author designed (ELISA) -Spike | 0.739, 0.994 | Validated by independent authors/third party/non-developers |
| Mali - HRP (Bamako) | Issaka Sagara (National Institutes of Health)[234] | 2020-12-21 - 2021-01-26 | Local | Moderate | 528 | Multiple groups | All | Convenience | Household and community samples | 44.9% | NA - - Author designed (ELISA) -Spike | 0.739, 0.994 | Validated by independent authors/third party/non-developers |
| Mali - HRP (Doneguebougou) | Issaka Sagara (National Institutes of Health)[234] | 2020-12-14 - 2021-01-15 | Local | Moderate | 1,088 | Multiple groups | All | Convenience | Household and community samples | 25.8% | NA - - Author designed (ELISA) -Spike | 0.739, 0.994 | Validated by independent authors/third party/non-developers |
| Mali - HRP (Bamako) | Issaka Sagara (National Institutes of Health)[234] | 2020-07-29 - 2020-10-16 | Local | Moderate | 587 | Multiple groups | All | Convenience | Household and community samples | 13.1% | NA - - Author designed (ELISA) -Spike | 0.739, 0.994 | Validated by independent authors/third party/non-developers |
| Mali - HRP (Bancoumana) | Issaka Sagara (National Institutes of Health)[234] | 2020-07-29 - 2020-09-24 | Local | Moderate | 963 | Multiple groups | All | Convenience | Household and community samples | 5.3% | NA - - Author designed (ELISA) -Spike | 0.739, 0.994 | Validated by independent authors/third party/non-developers |
| Mali - HRP (Doneguebougou) | Issaka Sagara (National Institutes of Health)[234] | 2020-07-28 - 2020-08-27 | Local | Moderate | 1,109 | Multiple groups | All | Convenience | Household and community samples | 4.1% | NA - - Author designed (ELISA) -Spike | 0.739, 0.994 | Validated by independent authors/third party/non-developers |
| Mozambique - HRP (Massinga) | Mussagy Mahomed (Republica de Mocambique Ministerio da Saude)[235] | 2020-11-26 - 2020-12-03 | Local | Moderate | 1,577 | Multiple groups | All | Probability | Household and community samples | 7.4% | Qingdao Hightop Biotech Co. Ltd - IgG, IgM - Hightop COVID-19 IgM/IgG Ab Rapid Test Kit | 0.942, 0.939 | Validated by manufacturer |
| Mozambique - HRP (Maxixe) | Mussagy Mahomed (Republica de Mocambique Ministerio da Saude)[235] | 2020-11-07 - 2020-11-21 | Local | Moderate | 3,974 | Multiple groups | All | Probability | Household and community samples | 5.5% | Qingdao Hightop Biotech Co. Ltd - IgG, IgM - Hightop COVID-19 IgM/IgG Ab Rapid Test Kit | 0.942, 0.939 | Validated by manufacturer |
| Mozambique - HRP (Chimoio) | Arlete Mahumane (Republica de Mocambique Ministerio da Saude)[236] | 2020-11-02 - 2020-11-17 | Local | Moderate | 9,756 | Multiple groups | All | Probability | Household and community samples | 1.4% | Qingdao Hightop Biotech Co. Ltd - IgG, IgM - Hightop COVID-19 IgM/IgG Ab Rapid Test Kit | 0.942, 0.939 | Validated by manufacturer |
| Mozambique - HRP (Lichinga) | Jeronimo Langa (Republica de Mocambique Ministerio da Saude)[237] | 2020-09-28 - 2020-10-09 | Local | Moderate | 1,635 | Multiple groups | All | Probability | Household and community samples | 0.3% | Qingdao Hightop Biotech Co. Ltd - IgG, IgM - Hightop COVID-19 IgM/IgG Ab Rapid Test Kit | 0.942, 0.939 | Validated by manufacturer |
| Mozambique - HRP (Tete) | Mussagy Mahomed (Republica de Mocambique Ministerio da Saude)[238] | 2020-08-31 - 2020-10-12 | Local | Moderate | 1,946 | Multiple groups | All | Probability | Household and community samples | 0.7% | Qingdao Hightop Biotech Co. Ltd - IgG, IgM - Hightop COVID-19 IgM/IgG Ab Rapid Test Kit | 0.942, 0.939 | Validated by manufacturer |
| Mozambique - HRP (Pemba) | Paulo Arnaldo (República de Moçambique Ministério da Saúde)[239] | 2020-07-06 - 2020-07-13 | Local | Moderate | 1,360 | Multiple groups | All | Probability | Household and community samples | 2.5% | Qingdao Hightop Biotech Co. Ltd - IgG, IgM - Hightop COVID-19 IgM/IgG Ab Rapid Test Kit | 0.942, 0.939 | Validated by manufacturer |
| Mozambique - HRP (Nampula) | Eduardo Samo Gudo (Republica de Mocambique Ministerio da Saude)[240] | 2020-06-17 - 2020-06-30 | Local | Moderate | 1,769 | Multiple groups | All | Probability | Household and community samples | 5% | SD Biosensor - IgG, IgM - Standard Q COVID-19 IgM/IgG Duo rapid immunochromatography test kit | 0.991, 0.951 | Validated by manufacturer |
| Nigeria - HRP (Abuja, Enugu) | Ijeoma Ifeorah (Molecular Pathology Institute)[241] | 2020-08-01 - 2020-08-31 | Local | High | 113 | Adults (18-64 years) | All | Convenience | Blood donors | 42% | NovaTec Immundiagnostics GmbH - IgG, IgM - NovaLisa® SARS-CoV-2 IgG/IgM | 1, 0.98 | Validated by manufacturer |
| South Africa (Bushbuckridge Municipality) | Jackie Kleynhans (National Health Laboratory Service)[242] | 2021-11-15 - 2021-11-27 | Local | Moderate | 550 | Multiple groups | All | Probability | Household and community samples | 59.8% | Roche Diagnostics - IgG, IgM, IgA - Elecsys® Anti‐SARS‐CoV‐2 (N) | 1, 0.995 | Validated by manufacturer |
| South Africa (Matlosana Municipality) | Jackie Kleynhans (National Health Laboratory Service)[242] | 2021-11-15 - 2021-11-27 | Local | Moderate | 479 | Multiple groups | All | Probability | Household and community samples | 69.9% | Roche Diagnostics - IgG, IgM, IgA - Elecsys® Anti‐SARS‐CoV‐2 (N) | 1, 0.995 | Validated by manufacturer |
| South Africa | Russel Cable (Stellenbosch University)[243] | 2021-11-08 - 2021-11-12 | National | High | 3,395 | Multiple groups | All | Convenience | Blood donors | 71.1% | Roche Diagnostics - IgG, IgM, IgA - Elecsys® Anti‐SARS‐CoV‐2 (N) | 1, 0.995 | Validated by manufacturer |
| South Africa (Bushbuckridge Municipality) | Jackie Kleynhans (National Health Laboratory Service)[242] | 2021-09-13 - 2021-09-25 | Local | Moderate | 548 | Multiple groups | All | Probability | Household and community samples | 59.3% | Roche Diagnostics - IgG, IgM, IgA - Elecsys® Anti‐SARS‐CoV‐2 (N) | 1, 0.995 | Validated by manufacturer |
| South Africa (Matlosana Municipality) | Jackie Kleynhans (National Health Laboratory Service)[242] | 2021-09-13 - 2021-09-25 | Local | Moderate | 493 | Multiple groups | All | Probability | Household and community samples | 68.8% | Roche Diagnostics - IgG, IgM, IgA - Elecsys® Anti‐SARS‐CoV‐2 (N) | 1, 0.995 | Validated by manufacturer |
| South Africa (Bushbuckridge Municipality) | Jackie Kleynhans (National Health Laboratory Service)[242] | 2021-07-19 - 2021-08-05 | Local | Moderate | 578 | Multiple groups | All | Probability | Household and community samples | 38.9% | Roche Diagnostics - IgG, IgM, IgA - Elecsys® Anti‐SARS‐CoV‐2 (N) | 1, 0.995 | Validated by manufacturer |
| South Africa (Matlosana Municipality) | Jackie Kleynhans (National Health Laboratory Service)[242] | 2021-07-19 - 2021-08-05 | Local | Moderate | 499 | Multiple groups | All | Probability | Household and community samples | 55.7% | Roche Diagnostics - IgG, IgM, IgA - Elecsys® Anti‐SARS‐CoV‐2 (N) | 1, 0.995 | Validated by manufacturer |
| South Africa (Bushbuckridge Municipality) | Jackie Kleynhans (National Health Laboratory Service)[242] | 2021-05-20 - 2021-06-09 | Local | Moderate | 579 | Multiple groups | All | Probability | Household and community samples | 26.1% | Roche Diagnostics - IgG, IgM, IgA - Elecsys® Anti‐SARS‐CoV‐2 (N) | 1, 0.995 | Validated by manufacturer |
| South Africa (Matlosana Municipality) | Jackie Kleynhans (National Health Laboratory Service)[242] | 2021-05-20 - 2021-06-09 | Local | Moderate | 505 | Multiple groups | All | Probability | Household and community samples | 48.9% | Roche Diagnostics - IgG, IgM, IgA - Elecsys® Anti‐SARS‐CoV‐2 (N) | 1, 0.995 | Validated by manufacturer |
| South Africa (Hillbrow Community Healthcare Centre and Esselen Street Clinic, Region F, City of Johannesburg, Gauteng, South Africa) | Lee Fairlie (Wits Reproductive Health and HIV Institute)[244] | 2021-03-17 - 2021-06-09 | Local | High | 500 | Multiple groups | All | Convenience | Pregnant or parturient women | 64% (95% CI 59.6-68.2%) | Beijing Wantai Biological - IgG IgM IgA - Wantai SARS-CoV-2 Total Ab ELISA,Roche Diagnostics - IgG IgM IgA - Elecsys® Anti‐SARS‐CoV‐2 (N) | 1, 0.97 | Complex test algorithm: Manual calculation |
| South Africa (Agincourt) | Jackie Kleynhans (National Institute for Communicable Diseases)[245] | 2021-03-22 - 2021-04-11 | Local | Moderate | 586 | Multiple groups | All | Probability | Household and community samples | 25.3% (95% CI 21.8-29%) | Roche Diagnostics - IgG IgM IgA - Elecsys® Anti‐SARS‐CoV‐2 (N) | 1, 0.995 | Validated by manufacturer |
| South Africa (Klerksdorp) | Jackie Kleynhans (National Institute for Communicable Diseases)[245] | 2021-03-22 - 2021-04-11 | Local | Moderate | 505 | Multiple groups | All | Probability | Household and community samples | 40.4% (95% CI 36.1-44.8%) | Roche Diagnostics - IgG IgM IgA - Elecsys® Anti‐SARS‐CoV‐2 (N) | 1, 0.995 | Validated by manufacturer |
| South Africa (Agincourt) | Jackie Kleynhans (National Institute for Communicable Diseases)[245] | 2021-01-25 - 2021-02-21 | Local | Moderate | 586 | Multiple groups | All | Probability | Household and community samples | 21.5% (95% CI 18.1-24.9%) | Roche Diagnostics - IgG IgM IgA - Elecsys® Anti‐SARS‐CoV‐2 (N) | 1, 0.995 | Validated by manufacturer |
| South Africa (Klerksdorp) | Jackie Kleynhans (National Institute for Communicable Diseases)[245] | 2021-01-25 - 2021-02-21 | Local | Moderate | 509 | Multiple groups | All | Probability | Household and community samples | 35.4% (95% CI 31-39.5%) | Roche Diagnostics - IgG IgM IgA - Elecsys® Anti‐SARS‐CoV‐2 (N) | 1, 0.995 | Validated by manufacturer |
| South Africa (Agincourt) | Jackie Kleynhans (National Institute for Communicable Diseases)[245] | 2020-11-23 - 2020-12-12 | Local | Moderate | 566 | Multiple groups | All | Probability | Household and community samples | 7.1% (95% CI 5.1-9.5%) | Roche Diagnostics - IgG IgM IgA - Elecsys® Anti‐SARS‐CoV‐2 (N) | 1, 0.995 | Validated by manufacturer |
| South Africa (Klerksdorp) | Jackie Kleynhans (National Institute for Communicable Diseases)[245] | 2020-11-23 - 2020-12-12 | Local | Moderate | 523 | Multiple groups | All | Probability | Household and community samples | 27% (95% CI 23.2-31%) | Roche Diagnostics - IgG IgM IgA - Elecsys® Anti‐SARS‐CoV‐2 (N) | 1, 0.995 | Validated by manufacturer |
| South Africa (Agincourt) | Jackie Kleynhans (National Institute for Communicable Diseases)[245] | 2020-09-21 - 2020-10-10 | Local | Moderate | 490 | Multiple groups | All | Probability | Household and community samples | 4.9% (95% CI 3.2-7.2%) | Roche Diagnostics - IgG IgM IgA - Elecsys® Anti‐SARS‐CoV‐2 (N) | 1, 0.995 | Validated by manufacturer |
| South Africa (Klerksdorp) | Jackie Kleynhans (National Institute for Communicable Diseases)[245] | 2020-09-21 - 2020-10-10 | Local | Moderate | 498 | Multiple groups | All | Probability | Household and community samples | 21.7% (95% CI 18.1-25.6%) | Roche Diagnostics - IgG IgM IgA - Elecsys® Anti‐SARS‐CoV‐2 (N) | 1, 0.995 | Validated by manufacturer |
| South Africa (Agincourt) | Jackie Kleynhans (National Institute for Communicable Diseases)[245] | 2020-07-20 - 2020-09-17 | Local | Moderate | 439 | Multiple groups | All | Probability | Household and community samples | 1.1% (95% CI 0.4-2.6%) | Roche Diagnostics - IgG IgM IgA - Elecsys® Anti‐SARS‐CoV‐2 (N) | 1, 0.995 | Validated by manufacturer |
| South Africa (Klerksdorp) | Jackie Kleynhans (National Institute for Communicable Diseases)[245] | 2020-07-20 - 2020-09-17 | Local | Moderate | 501 | Multiple groups | All | Probability | Household and community samples | 14.4% (95% CI 11.4-17.8%) | Roche Diagnostics - IgG IgM IgA - Elecsys® Anti‐SARS‐CoV‐2 (N) | 1, 0.995 | Validated by manufacturer |
| South Africa (Cape Town) | Marvin Hsiao ( University of Cape Town)[246] | 2020-06-15 - 2020-08-07 | Local | High | 2,791 | Adults (18-64 years) | All | Sequential | Residual sera | 40.2% | Roche Diagnostics - IgG, IgM, IgA - Elecsys® Anti‐SARS‐CoV‐2 (N) | 1, 0.995 | Validated by manufacturer |
| South Sudan - HRP (Juba) | Kirsten E Wiens (World Health Organization)[247] | 2020-08-10 - 2020-09-11 | Local | Low | 2,214 | Multiple groups | All | Probability | Household and community samples | 22.3% | NA - IgG - Author designed (ELISA) -Spike | 0.655, 1 | Validated by developers |
| Zambia - HRP (Zambian Central Province) | Kwame Shanaube (Zambart)[248] | 2020-10-15 - 2021-03-15 | Local | Moderate | 2,977 | Multiple groups | All | Probability | Household and community samples | 13.5% | Abbott Laboratories - IgG - Abbott Architect SARS-CoV-2 IgG | 0.99, 1 | Validated by manufacturer |
| Zimbabwe - HRP (Harare) | Arun Fryatt (Biomedical Research and Training Institute)[249] | 2021-02-10 - 2021-04-17 | Local | Moderate | 1,530 | Multiple groups | All | Probability | Household and community samples | 53% | NA - - Author designed (type unknown) | 0.992, 0.9865 | Validated by developers |
| Zimbabwe - HRP (Harare) | Arun Fryatt (Biomedical Research and Training Institute)[249] | 2020-11-20 - 2020-12-20 | Local | Moderate | 620 | Multiple groups | All | Probability | Household and community samples | 19% | NA - - Author designed (type unknown) | 0.992, 0.9865 | Validated by developers |

###### Americas region

| Country (Location) | Author (Organization) | Sampling Dates (YMD) | Geographic scope | Overall risk of bias | Sample size | Age | Sex | Sampling method | Sample frame | Seroprevalence (95% CI) | Test Manufacturers - Isotypes - Names | Test Sens, Spec | Sens, Spec Source |
| --- | --- | --- | --- | --- | --- | --- | --- | --- | --- | --- | --- | --- | --- |
| Argentina - HRP (Quilmes) | Laura Munoz (Ministerio de Salud de la Provincia de Buenos Aires)[250] | 2020-07-15 - 2020-07-16 | Local | High | 284 | Multiple groups | All | Stratified non-probability | Household and community samples | 14.8% | COVIDAR - IgG - SEROKIT-ELISA COVIDAR IgG | 0.95, 1 | Validated by manufacturers |
| Argentina - HRP (Buenos Aires) | Silvana Figar (Ministry of Health of the City of Buenos Aires)[251] | 2020-06-10 - 2020-06-26 | Local | Low | 873 | Multiple groups | All | Probability | Persons living in slums | 53.4% | CONICET - IgG, IgM - COVIDAR IgM and IgG ELISA | 0.9, 1 | Validated by independent authors/third party/non-developers |
| Brazil - HRP (São Paulo) | Beatriz Tess (Inteligência em Pesquisa e Consultoria)[252] | 2021-04-22 - 2021-05-01 | Local | Moderate | 1,187 | Multiple groups | All | Probability | Household and community samples | 41.6% | Abbott Laboratories - IgG - Abbott Architect SARS-CoV-2 IgG,Roche Diagnostics - IgG, IgM, IgA - Elecsys® Anti‐SARS‐CoV‐2 (N) | 0.99, 1 | Complex test algorithm: Manual calculation |
| Brazil - HRP (Para) | Keise Pereira (Universidade Federal do Para)[253] | 2021-04-01 - 2021-04-30 | Local | High | 140 | Multiple groups | All | Convenience | Household and community samples | 52.1% | EUROIMMUN - IgG - Anti-SARS-CoV-2 ELISA IgG | 0.9, 1 | Validated by manufacturer |
| Brazil - HRP (São Paulo) | City of Sao Paulo (City of São Paulo)[254] | 2021-02-16 - 2021-02-18 | Local | Moderate | 1,793 | Multiple groups | All | Probability | Household and community samples | 25% | Guangzhou Wondfo Biotech Co. Ltd - IgG, IgM - Finecare SARS-CoV-2 Antibody test,EUROIMMUN - IgG - Anti-SARS-CoV-2 ELISA IgG | 0.987, 0.991 | Complex test algorithm: Manual calculation |
| Brazil - HRP (Salvador) | Mariam Fofana (Yale School of Public Health)[255] | 2020-11-18 - 2021-02-26 | Local | Moderate | 2,035 | Multiple groups | All | Probability | Persons living in slums | 46.4% | EUROIMMUN - IgG - Anti-SARS-CoV-2 ELISA IgG | 0.9, 1 | Validated by manufacturer |
| Brazil - HRP (Belém) | Hilton Pereira da Silva (Universidade Federal do Pará)[256] | 2020-09-15 - 2021-01-15 | Local | Moderate | 101 | Multiple groups | All | Probability | Household and community samples | 83.2% | EUROIMMUN - IgG - Anti-SARS-CoV-2 ELISA IgG | 0.9, 1 | Validated by manufacturer |
| Brazil - HRP (São Paulo) | Joao Luiz Miraglia (Hospital Israelita Albert Einstein)[257] | 2020-09-15 - 2020-12-15 | Local | Moderate | 272 | Multiple groups | All | Probability | Household and community samples | 45.2% | Roche Diagnostics - IgG, IgM, IgA - Elecsys® Anti‐SARS‐CoV‐2 (N) | 1, 0.995 | Validated by manufacturer |
| Brazil - HRP (Manaus) | Gemilson Soares Pontes (Instituto Nacional de Pesquisas da Amazônia)[258] | 2020-10-10 - 2020-11-14 | Local | High | 280 | Multiple groups | All | Sequential | Household and community samples | 64.6% | EUROIMMUN - IgA - Anti-SARS-CoV-2 ELISA IgA,EUROIMMUN - IgG - Anti-SARS-CoV-2 ELISA IgG | 0.997, 0.983 | Complex test algorithm: Manual calculation |
| Brazil - HRP (Sao Paulo) | Lewis Buss (Universidade de Sao Paulo)[259] | 2020-10-12 - 2020-10-24 | Local | Moderate | 877 | Multiple groups | All | Sequential | Blood donors | 19.5% | Abbott Laboratories - IgG - Abbott Architect SARS-CoV-2 IgG | 0.99, 1 | Validated by manufacturer |
| Brazil - HRP (Manaus) | Lewis Buss (Universidade de Sao Paulo)[259] | 2020-10-10 - 2020-10-17 | Local | Moderate | 882 | Multiple groups | All | Sequential | Blood donors | 43.1% | Abbott Laboratories - IgG - Abbott Architect SARS-CoV-2 IgG | 0.99, 1 | Validated by manufacturer |
| Brazil - HRP (Sao Paulo) | Lewis Buss (Universidade de Sao Paulo)[259] | 2020-09-07 - 2020-09-29 | Local | Moderate | 933 | Multiple groups | All | Sequential | Blood donors | 16.7% | Abbott Laboratories - IgG - Abbott Architect SARS-CoV-2 IgG | 0.99, 1 | Validated by manufacturer |
| Brazil - HRP (Manaus) | Lewis Buss (Universidade de Sao Paulo)[259] | 2020-09-05 - 2020-09-14 | Local | Moderate | 868 | Multiple groups | All | Sequential | Blood donors | 45.4% | Abbott Laboratories - IgG - Abbott Architect SARS-CoV-2 IgG | 0.99, 1 | Validated by manufacturer |
| Brazil - HRP (Victória) | Carolina Ferrugini (Federal University of Espírito Santo)[260] | 2020-07-15 - 2020-10-15 | Local | High | 261 | Adults (18-64 years) | Female | Sequential | Pregnant or parturient women | 27.2% | ECO Diagnostica LTDA - IgG, IgM - COVID-19 IgG/IgM ECO Test | 0.971, 0.987 | Validated by manufacturer |
| Brazil - HRP (Sao Paulo) | Lewis Buss (Universidade de Sao Paulo)[259] | 2020-08-10 - 2020-08-29 | Local | Moderate | 906 | Multiple groups | All | Sequential | Blood donors | 17.2% | Abbott Laboratories - IgG - Abbott Architect SARS-CoV-2 IgG | 0.99, 1 | Validated by manufacturer |
| Brazil - HRP (Manaus) | Lewis Buss (Universidade de Sao Paulo)[259] | 2020-08-08 - 2020-08-19 | Local | Moderate | 881 | Multiple groups | All | Sequential | Blood donors | 44% | Abbott Laboratories - IgG - Abbott Architect SARS-CoV-2 IgG | 0.99, 1 | Validated by manufacturer |
| Brazil - HRP (Rio de Janeiro) | Pamella Lugon (Oswaldo Cruz Foundation)[261] | 2020-05-18 - 2020-09-24 | Local | High | 515 | Multiple groups | All | Convenience | Multiple general populations | 29.3% | Abbott Laboratories - IgG - Abbott Architect SARS-CoV-2 IgG | 0.99, 1 | Validated by manufacturer |
| Brazil - HRP (Sao Paulo) | Lewis Buss (Universidade de Sao Paulo)[259] | 2020-07-13 - 2020-07-25 | Local | Moderate | 879 | Multiple groups | All | Sequential | Blood donors | 12.8% | Abbott Laboratories - IgG - Abbott Architect SARS-CoV-2 IgG | 0.99, 1 | Validated by manufacturer |
| Brazil - HRP (Sao Paulo) | Luanda Mara da Silva Oliveira (Universidade de São Paulo)[262] | 2020-06-30 - 2020-08-04 | Local | High | 439 | Multiple groups | All | Convenience | Residual sera | 13.9% | DiaSorin - IgG - Liaison SARS-CoV-2 S1/S2 IgG,Roche Diagnostics - IgG, IgM, IgA - Elecsys® Anti‐SARS‐CoV‐2 (N) | 0.974, 1 | Complex test algorithm: Manual calculation |
| Brazil - HRP (Betim) | Ana Silva (Universidade Federal de Minas Gerais)[263] | 2020-07-13 - 2020-07-15 | Local | Moderate | 1,080 | Multiple groups | All | Probability | Household and community samples | 2.6% | Guangzhou Wondfo Biotech Co. Ltd - IgG, IgM - Finecare SARS-CoV-2 Antibody test | 0.872, 0.991 | Validated by manufacturer |
| Brazil - HRP (Joinville) | Henrique Diegoli (City Hall of Joinville)[264] | 2020-06-15 - 2020-08-07 | Local | Moderate | 4,403 | Multiple groups | All | Probability | Household and community samples | 4.8% | Celer Technologies Inc. - IgG, IgM - One Step COVID-19 Test | 0.8643, 0.9957 | Validated by independent authors/third party/non-developers |
| Brazil - HRP (Manaus) | Lewis Buss (Universidade de Sao Paulo)[259] | 2020-07-06 - 2020-07-15 | Local | Moderate | 1,147 | Multiple groups | All | Sequential | Blood donors | 46.9% | Abbott Laboratories - IgG - Abbott Architect SARS-CoV-2 IgG | 0.99, 1 | Validated by manufacturer |
| Brazil - HRP (Rio de Janeiro) | Alanna Calheiros Santos (Oswaldo Cruz Foundation)[265] | 2020-01-07 - 2021-01-11 | Local | High | 31 | Adults (18-64 years) | Female | Convenience | Pregnant or parturient women | 19.4% | Roche Diagnostics - IgG, IgM, IgA - Elecsys® Anti‐SARS‐CoV‐2 (N) | 1, 0.995 | Validated by manufacturer |
| Brazil - HRP (Carmópolis) | Breno Bernardes-Souza (Federal University of Ouro Preto)[266] | 2020-06-27 - 2020-06-28 | Local | Moderate | 400 | Multiple groups | All | Probability | Household and community samples | 1.8% | Qingdao Hightop Biotech Co. Ltd - IgG, IgM - Hightop COVID-19 IgM/IgG Ab Rapid Test Kit | 0.942, 0.939 | Validated by manufacturer |
| Brazil - HRP (Sao Paulo) | Lewis Buss (Universidade de Sao Paulo)[259] | 2020-06-08 - 2020-06-20 | Local | Moderate | 880 | Multiple groups | All | Sequential | Blood donors | 15% | Abbott Laboratories - IgG - Abbott Architect SARS-CoV-2 IgG | 0.99, 1 | Validated by manufacturer |
| Brazil - HRP (Ribeirão Preto (RP)) | Edson Zangiacomi Martinez (Universidade de São Paulo)[267] | 2020-06-11 - 2020-06-14 | Local | Moderate | 646 | Multiple groups | All | Probability | Household and community samples | 2.8% | Guangzhou Wondfo Biotech Co. Ltd - IgG, IgM, IgA - Wondfo SARS-CoV-2 Antibody Test | 0.864, 0.996 | Validated by manufacturer |
| Brazil - HRP (Manaus) | Lewis Buss (Universidade de Sao Paulo)[259] | 2020-06-05 - 2020-06-15 | Local | Moderate | 911 | Multiple groups | All | Sequential | Blood donors | 52.7% | Abbott Laboratories - IgG - Abbott Architect SARS-CoV-2 IgG | 0.99, 1 | Validated by manufacturer |
| Brazil - HRP (Nepomuceno) | Breno Bernardes-Souza (Federal University of Ouro Preto)[266] | 2020-05-30 - 2020-05-31 | Local | Moderate | 400 | Multiple groups | All | Probability | Household and community samples | 0.5% | Qingdao Hightop Biotech Co. Ltd - IgG, IgM - Hightop COVID-19 IgM/IgG Ab Rapid Test Kit | 0.942, 0.939 | Validated by manufacturer |
| Brazil - HRP (Sao Paulo) | Lewis Buss (Universidade de Sao Paulo)[259] | 2020-05-08 - 2020-05-21 | Local | Moderate | 826 | Multiple groups | All | Sequential | Blood donors | 7.5% | Abbott Laboratories - IgG - Abbott Architect SARS-CoV-2 IgG | 0.99, 1 | Validated by manufacturer |
| Brazil - HRP (Manaus) | Lewis Buss (Universidade de Sao Paulo)[259] | 2020-05-05 - 2020-05-14 | Local | Moderate | 901 | Multiple groups | All | Sequential | Blood donors | 42.1% | Abbott Laboratories - IgG - Abbott Architect SARS-CoV-2 IgG | 0.99, 1 | Validated by manufacturer |
| Brazil - HRP | Beatriz Tess (Universidade de Sao Paulo)[268] | 2020-05-04 - 2020-05-12 | Subnational | High | 463 | Multiple groups | All | Stratified non-probability | Multiple general populations | 6% | Snibe Co. Ltd (Shenzhen New Industries Biomedical Engineering Co. Ltd) - IgG, IgM - MAGLUMI 2019-nCoV IgM/IgG,Roche Diagnostics - IgG, IgM, IgA - Elecsys® Anti‐SARS‐CoV‐2 (N) | 1, 0.982 | Complex test algorithm: Manual calculation |
| Brazil - HRP (Ribeirão Preto (RP)) | Edson Zangiacomi Martinez (Universidade de São Paulo)[267] | 2020-05-01 - 2020-05-03 | Local | Moderate | 709 | Multiple groups | All | Probability | Household and community samples | 1.3% | Guangzhou Wondfo Biotech Co. Ltd - IgG, IgM, IgA - Wondfo SARS-CoV-2 Antibody Test | 0.864, 0.996 | Validated by manufacturer |
| Brazil - HRP (Sao Paulo) | Lewis Buss (Universidade de Sao Paulo)[259] | 2020-04-08 - 2020-04-30 | Local | Moderate | 900 | Multiple groups | All | Sequential | Blood donors | 6.8% | Abbott Laboratories - IgG - Abbott Architect SARS-CoV-2 IgG | 0.99, 1 | Validated by manufacturer |
| Brazil - HRP (Manaus) | Lewis Buss (Universidade de Sao Paulo)[259] | 2020-04-06 - 2020-04-17 | Local | Moderate | 829 | Multiple groups | All | Sequential | Blood donors | 7.7% | Abbott Laboratories - IgG - Abbott Architect SARS-CoV-2 IgG | 0.99, 1 | Validated by manufacturer |
| Brazil - HRP (Sao Paulo) | Lewis Buss (Universidade de Sao Paulo)[259] | 2020-03-09 - 2020-03-21 | Local | Moderate | 2,454 | Multiple groups | All | Sequential | Blood donors | 5.8% | Abbott Laboratories - IgG - Abbott Architect SARS-CoV-2 IgG | 0.99, 1 | Validated by manufacturer |
| Brazil - HRP (Manaus) | Lewis Buss (Universidade de Sao Paulo)[259] | 2020-03-06 - 2020-03-12 | Local | Moderate | 832 | Multiple groups | All | Sequential | Blood donors | 2.6% | Abbott Laboratories - IgG - Abbott Architect SARS-CoV-2 IgG | 0.99, 1 | Validated by manufacturer |
| Brazil - HRP (Aracaju) | Ricardo Queiroz Gurgel (Federal University of Sergipe)[269] | 2020-01-15 - 2020-04-15 | Local | High | 987 | Multiple groups | All | Sequential | Residual sera | 1.6% | NA - - Not reported/ Unable to specify,Boditech Med Inc. - IgG, IgM - Ichroma COVID-19 Ab Test | N/A | Complex test algorithm: Manual review |
| Brazil - HRP (Manaus) | Lewis Buss (Universidade de Sao Paulo)[259] | 2020-02-07 - 2020-02-13 | Local | Moderate | 821 | Multiple groups | All | Sequential | Blood donors | 3.6% | Abbott Laboratories - IgG - Abbott Architect SARS-CoV-2 IgG | 0.99, 1 | Validated by manufacturer |
| Brazil - HRP (Sao Paulo) | Lewis Buss (Universidade de Sao Paulo)[259] | 2020-02-08 - 2020-02-29 | Local | Moderate | 799 | Multiple groups | All | Sequential | Blood donors | 4.2% | Abbott Laboratories - IgG - Abbott Architect SARS-CoV-2 IgG | 0.99, 1 | Validated by manufacturer |
| Colombia - HRP (Mitú) | Hector Serrano-Coll (Universidad de Córdoba)[270] | 2020-12-01 - 2020-12-31 | Local | High | 589 | Multiple groups | All | Convenience | Household and community samples | 57.6% | Eurofins Ingenasa - IgG, IgM, IgA - INgezim COVID 19 DR | 0.983, 0.992 | Validated by manufacturer |
| Colombia - HRP (Monteria) | Salim Mattar (Universidad de Córdoba)[271] | 2020-08-04 - 2020-09-01 | Local | Low | 1,368 | Multiple groups | All | Probability | Household and community samples | 55.3% | Eurofins Ingenasa - IgG, IgM, IgA - INgezim COVID 19 DR | 0.983, 0.992 | Validated by manufacturer |
| Ecuador - HRP (Atahualpa) | Oscar Del Brutto (Universidad Espíritu Santo)[272] | 2020-05-01 - 2021-04-30 | Local | High | 673 | Adults (18-64 years) | All | Convenience | Household and community samples | 63.7% | Biohit Health Care Ltd - IgG, IgM - BIOHIT SARS-CoV-2 antibody test | 0.975, 1 | Validated by independent authors/third party/non-developers |
| Ecuador - HRP (Azuay) | David Acurio Paez (University of Antwerp)[273] | 2020-08-11 - 2020-11-01 | Local | Moderate | 2,457 | Multiple groups | All | Probability | Household and community samples | 13.2% | SD Biosensor - IgG, IgM - Standard Q COVID-19 IgM/IgG Duo rapid immunochromatography test kit | 0.991, 0.951 | Validated by manufacturer |
| French Guiana | Claude Flamand (Institut Pasteur in French Guiana)[274] | 2020-07-15 - 2020-07-23 | National | High | 480 | Multiple groups | All | Convenience | Residual sera | 13.1% | EUROIMMUN - IgG - Anti-SARS-CoV-2 ELISA IgG | 0.9, 1 | Validated by manufacturer |
| Jamaica (Kingston) | TeAnne Chisolm (University of the West Indies)[275] | 2020-11-01 - 2020-11-30 | Local | High | 77 | Adults (18-64 years) | Female | Sequential | Pregnant or parturient women | 24% | Abbott Laboratories - IgG - Abbott Architect SARS-CoV-2 IgG | 0.99, 1 | Validated by manufacturer |
| Mexico - HRP (Campeche) | Victor Monteon (Universidad Autonoma Campeche)[276] | 2021-08-15 - 2021-09-15 | Subnational | High | 479 | Multiple groups | All | Sequential | Blood donors | 69.1% | NA - - Author designed (ELISA) -Spike | 0.94, 1 | Validated by developers |
| Mexico - HRP | Jose Munoz-Medina (Instituto de Salud para el Bienestar)[277] | 2020-12-01 - 2020-12-31 | National | High | 5,359 | Multiple groups | All | Stratified non-probability | Residual sera | 30.6% | DiaSorin - IgG - Liaison SARS-CoV-2 S1/S2 IgG,GenScript - IgG, IgM - cPass SARS-CoV-2 Surrogate Virus Neutralization Test ELISA Kit | 0.914, 1 | Complex test algorithm: Manual calculation |
| Mexico - HRP (Veracruz City) | Jose Maria Remes-Troche (Universidad Veracruzana)[278] | 2020-06-01 - 2020-07-31 | Local | High | 2,174 | Multiple groups | All | Sequential | Residual sera | 29.5% | Abbott Laboratories - IgG - Abbott Architect SARS-CoV-2 IgG | 0.99, 1 | Validated by manufacturer |
| Mexico - HRP (Nuevo Leon) | Natalia Martinez-Acu a (Autonomous University of Nuevo Leon)[279] | 2020-01-01 - 2020-12-15 | Subnational | High | 2,068 | Adults (18-64 years) | All | Sequential | Blood donors | 4.8% | Abbott Laboratories - IgG - Abbott Architect SARS-CoV-2 IgG | 0.99, 1 | Validated by manufacturer |
| Peru - HRP (La Riconanda) | Benoit Champigneulle (Grenoble Alpes University)[280] | 2020-10-11 - 2020-10-18 | Local | High | 159 | Multiple groups | All | Sequential | Household and community samples | 48.4% | AAZ LMB - IgG, IgM - COVID-PRESTO® | 1, 1 | Validated by independent authors/third party/non-developers |
| Peru - HRP (Jaen) | Yordi Tarazona-Castro (Universidad Peruana de Ciencias Aplicadas)[281] | 2020-04-15 - 2021-03-15 | Local | High | 464 | Multiple groups | All | Sequential | Representative patient population | 35.8% | ImmunoDiagnostics - IgG, IgM - SARS-CoV-2 NP IgG/IgM ELISA Kit | 0.925, 0.933 | Validated by manufacturer |
| Peru - HRP (Iquitos) | Carlos Alvarez-Antonio (Universidad de Ingeniería y Tecnología)[282] | 2020-08-13 - 2020-08-18 | Local | Low | 621 | Multiple groups | All | Probability | Household and community samples | 68% | Zhejiang Orient Gene Biotech - IgG, IgM - COVID-19 IgG/IgM Rapid Test | 0.972, 1 | Validated by manufacturer |
| Peru - HRP (Iquitos) | Carlos Alvarez-Antonio (Universidad de Ingeniería y Tecnología)[282] | 2020-07-13 - 2020-07-18 | Local | Low | 716 | Multiple groups | All | Probability | Household and community samples | 73.7% | Zhejiang Orient Gene Biotech - IgG, IgM - COVID-19 IgG/IgM Rapid Test | 0.972, 1 | Validated by manufacturer |
| Peru - HRP (Lima metropolitan area) | Mary F Reyes Vega (Peruvian Ministry of Health)[283] | 2020-06-28 - 2020-07-09 | Local | Low | 3,212 | Multiple groups | All | Probability | Household and community samples | 21.4% | SD Biosensor - IgG, IgM - Standard Q COVID-19 IgM/IgG Duo rapid immunochromatography test kit | 0.991, 0.951 | Validated by manufacturer |
| Peru - HRP (Lima) | Enrique Guevara Rios (National Maternal Perinatal Institut)[284] | 2020-04-15 - 2020-05-15 | Local | Moderate | 2,419 | Adults (18-64 years) | Female | Sequential | Pregnant or parturient women | 7% | NA - - Not reported/ Unable to specify | 0.976, 0.988 | Validated by manufacturers |
| Canada (Vancouver) | David Goldfarb (University of British Colombia)[285] | 2021-01-01 - 2021-05-31 | Local | High | 5,417 | Adults (18-64 years) | All | Convenience | Blood donors | 2.6% | Roche Diagnostics - IgG, IgM, IgA - Elecsys® Anti‐SARS‐CoV‐2 (N) | 1, 0.995 | Validated by manufacturer |
| Canada | Xuyang Tang (University of Toronto)[286] | 2020-12-01 - 2021-03-31 | National | High | 6,955 | Multiple groups | All | Convenience | Household and community samples | 6.5% | NA - IgG - Author designed (ELISA) -Spike,NA - IgG - Author designed (ELISA) - Nucleocapsid | N/A | Complex test algorithm: Manual review |
| Canada | Ashleigh Tuite (Canadian Blood Services)[287] | 2020-04-15 - 2021-03-15 | National | High | 17,999 | Multiple groups | All | Convenience | Blood donors | 2.1% | Abbott Laboratories - IgG - Abbott Architect SARS-CoV-2 IgG,NA - IgG - Author designed (ELISA) - Nucleocapsid,NA - IgG - Author designed (ELISA) -Spike | N/A | Complex test algorithm: Manual review |
| Canada (Edmonton) | Emilie Manny (University of Alberta)[288] | 2020-08-14 - 2020-10-23 | Local | High | 565 | Children and Youth (0-17 years) | All | Convenience | Household and community samples | 1.6% | Abbott Laboratories - IgG - Abbott Architect SARS-CoV-2 IgG | 0.99, 1 | Validated by manufacturer |
| Canada (Québec) | Hema-Quebec (Héma-Québec)[289] | 2020-05-25 - 2020-07-09 | Subnational | High | 7,691 | Adults (18-64 years) | All | Convenience | Blood donors | 2.2% | NA - IgG, IgM, IgA - Author designed (ELISA) -Spike | 0.989, 0.985 | Validated by developers |
| Canada (British Columbia) | Guadalein Tanunliong (University of British Columbia)[290] | 2020-03-15 - 2020-05-31 | Subnational | High | 488 | Multiple groups | All | Probability | Residual sera | 1.8% | Meso Scale Discovery - IgG - Meso Scale Discovery multiplex assay | 1, 1 | Validated by independent authors/third party/non-developers |
| Panama - HRP (Panama City, Colon) | Alcibiades Villarreal (Instituto de Investigaciones Científicas y Servicios de Alta Tecnología)[291] | 2020-04-30 - 2020-07-07 | Local | High | 255 | Multiple groups | All | Convenience | Blood donors | 13.3% | NA - IgG, IgM - Author designed (ELISA) - Nucleocapsid | 0.8701, 0.9889 | Validated by developers |
| United States of America | Kristie Clarke (Centers for Disease Control and Prevention (CDC))[292] | 2022-02-01 - 2022-02-28 | National | High | 45,810 | Multiple groups | All | Convenience | Household and community samples | 57.7% | Roche Diagnostics - IgG, IgM, IgA - Elecsys® Anti‐SARS‐CoV‐2 (N) | 1, 0.995 | Validated by manufacturer |
| United States of America | Kristie Clarke (US Center for Disease Control)[293] | 2021-09-15 - 2022-02-15 | National | High | 71,490 | Children and Youth (0-17 years) | All | Stratified non-probability | Residual sera | 52% | Roche Diagnostics - IgG, IgM, IgA - Elecsys® Anti‐SARS‐CoV‐2 (N) | 1, 0.995 | Validated by manufacturer |
| United States of America (New Jersey) | Alexia Couture (Centers for Disease Control and Prevention)[150] | 2021-05-01 - 2021-05-31 | Subnational | High | 393 | Children and Youth (0-17 years) | All | Convenience | Residual sera | 44% | Roche Diagnostics - IgG, IgM, IgA - Elecsys® Anti‐SARS‐CoV‐2 (N) | 1, 0.995 | Validated by manufacturer |
| United States of America (Nevada) | Alexia Couture (Centers for Disease Control and Prevention)[150] | 2021-05-01 - 2021-05-31 | Subnational | High | 237 | Children and Youth (0-17 years) | All | Convenience | Residual sera | 30.4% | Roche Diagnostics - IgG, IgM, IgA - Elecsys® Anti‐SARS‐CoV‐2 (N) | 1, 0.995 | Validated by manufacturer |
| United States of America (Tennessee) | Alexia Couture (Centers for Disease Control and Prevention)[150] | 2021-05-01 - 2021-05-31 | Subnational | High | 192 | Children and Youth (0-17 years) | All | Convenience | Residual sera | 36.5% | Roche Diagnostics - IgG, IgM, IgA - Elecsys® Anti‐SARS‐CoV‐2 (N) | 1, 0.995 | Validated by manufacturer |
| United States of America (California) | Neeraj Sood (University of Southern California)[294] | 2021-04-09 - 2021-04-25 | Local | Low | 1,335 | Multiple groups | All | Probability | Household and community samples | 66.9% | Luminex Corporation - IgG - xMAP® SARS‑CoV‑2 Multi-Antigen IgG Assay | 0.981, 0.992 | Validated by manufacturer |
| United States of America (San Francisco) | Isobel Routledge (University of California San Francisco)[295] | 2021-02-04 - 2021-02-17 | Local | High | 915 | Multiple groups | All | Stratified non-probability | Residual sera | 28.4% | Ortho Clinical Diagnostics Inc. - IgG, IgM, IgA - VITROS Immunodiagnostic Products Anti-SARS-CoV-2 Total | 1, 1 | Validated by manufacturer |
| United States of America (Pittsburgh) | Lingqing Xu (University of Pittsburgh)[296] | 2021-02-01 - 2021-02-05 | Local | High | 194 | Multiple groups | All | Sequential | Residual sera | 16.5% | NA - - Author designed (Neutralization Assay),NA - IgG - Author designed (ELISA) - Nucleocapsid | N/A | Complex test algorithm: Manual review |
| United States of America (St Louis) | Brittany K. Smith (Washington University School of Medicine)[297] | 2020-12-24 - 2021-01-21 | Local | High | 1,001 | Multiple groups | All | Convenience | Residual sera | 20.7% | NA - IgG - Author designed (ELISA) -Spike | 0.982, 0.987 | Validated by developers |
| United States of America (Seattle) | Sylvia LaCourse (University of Washington)[298] | 2020-09-09 - 2021-05-07 | Local | Moderate | 1,304 | Adults (18-64 years) | Female | Convenience | Pregnant or parturient women | 4.8% | Abbott Laboratories - IgG - Abbott Architect SARS-CoV-2 IgG | 0.99, 1 | Validated by manufacturer |
| United States of America (Illinois) | Ryan Kilpatrick (University of Texas Medical Branch)[299] | 2020-11-15 - 2021-01-15 | Local | High | 1,008 | Multiple groups | All | Convenience | Household and community samples | 4.5% | Abbott Laboratories - IgG - Abbott Architect SARS-CoV-2 IgG | 0.99, 1 | Validated by manufacturer |
| United States of America | Robert Stout (Clinical Reference Laboratory Inc.)[300] | 2020-12-01 - 2020-12-06 | National | High | 14,605 | Multiple groups | All | Convenience | Residual sera | 10.4% | Roche Diagnostics - IgG, IgM, IgA - Elecsys® Anti‐SARS‐CoV‐2 (N) | 1, 0.995 | Validated by manufacturer |
| United States of America | Roger Dodd (American Red Cross)[301] | 2020-11-29 - 2020-11-30 | National | High | 13,268 | Multiple groups | All | Sequential | Blood donors | 9.7% | Ortho Clinical Diagnostics Inc. - IgG, IgM, IgA - VITROS Immunodiagnostic Products Anti-SARS-CoV-2 Total | 1, 1 | Validated by manufacturer |
| United States of America (St Louis) | Brittany K. Smith (Washington University School of Medicine)[297] | 2020-10-27 - 2020-11-30 | Local | High | 1,013 | Multiple groups | All | Convenience | Residual sera | 11.7% | NA - IgG - Author designed (ELISA) -Spike | 0.982, 0.987 | Validated by developers |
| United States of America (New York) | Cara Staszewski (North Shore University Hospital)[302] | 2020-10-15 - 2020-11-15 | Local | High | 149 | Adults (18-64 years) | Female | Sequential | Pregnant or parturient women | 14.1% | Roche Diagnostics - IgG, IgM, IgA - Elecsys® Anti‐SARS‐CoV‐2 (S) | 0.988, 1 | Validated by manufacturer |
| United States of America (Pennsylvania) | Callum Arnold (Pennsylvania State University)[303] | 2020-08-07 - 2020-12-20 | Local | High | 345 | Adults (18-64 years) | All | Convenience | Household and community samples | 8.2% | NA - IgG - Author designed (ELISA) -Spike | 0.933, 0.99 | Validated by independent authors/third party/non-developers |
| United States of America (Arizona) | Megan Jehn (Arizona State University)[304] | 2020-09-12 - 2020-10-01 | Local | Moderate | 260 | Multiple groups | All | Probability | Household and community samples | 11.5% | Roche Diagnostics - IgG, IgM, IgA - Elecsys® Anti‐SARS‐CoV‐2 (S) | 0.988, 1 | Validated by manufacturer |
| United States of America (New Mexico) | Fabrizio Bonelli (Dia Sorin Inc)[305] | 2020-09-01 - 2020-09-30 | Subnational | High | 1,931 | Multiple groups | All | Sequential | Residual sera | 2% | DiaSorin - IgG - Liaison SARS-CoV-2 S1/S2 IgG,Beckman Coulter - IgG - Access SARS-CoV-2 IgG Antibody Test | 1, 0.983 | Complex test algorithm: Manual calculation |
| United States of America (New York) | Fabrizio Bonelli (Dia Sorin Inc)[305] | 2020-09-01 - 2020-09-30 | Subnational | High | 1,491 | Multiple groups | All | Sequential | Residual sera | 24.4% | Siemens - IgG, IgM, IgA - ADVIA Centaur Immunoassay System,DiaSorin - IgG - Liaison SARS-CoV-2 S1/S2 IgG | N/A | Complex test algorithm: Manual review |
| United States of America | Robert Stout (Clinical Reference Laboratory Inc)[306] | 2020-09-01 - 2020-09-30 | National | High | 61,910 | Multiple groups | All | Sequential | Household and community samples | 6.6% | Roche Diagnostics - IgG, IgM, IgA - Elecsys® Anti‐SARS‐CoV‐2 (N) | 1, 0.995 | Validated by manufacturer |
| United States of America (St. Louis) | Sumanth Gandra (Washington University School of Medicine)[307] | 2020-09-01 - 2020-09-30 | Local | High | 226 | Multiple groups | All | Convenience | Residual sera | 8.4% | Abbott Laboratories - IgG - Abbott Architect SARS-CoV-2 IgG | 0.99, 1 | Validated by manufacturer |
| United States of America (Houston) | Elaine Symanski (Houston Health Department)[308] | 2020-09-01 - 2020-09-19 | Local | Low | 685 | Multiple groups | All | Probability | Household and community samples | 11.5% | Bio-rad - IgG, IgM, IgA - Platelia SARS-CoV-2 Total Ab assay,NA - IgG - Author designed (ELISA) -Spike | N/A | Complex test algorithm: Manual review |
| United States of America (Washington D.C) | Burak Bahar (Children’s National Hospital)[309] | 2020-07-07 - 2020-10-29 | Local | High | 385 | Children and Youth (0-17 years) | All | Convenience | Residual sera | 9.9% | DiaSorin - IgG - Liaison SARS-CoV-2 S1/S2 IgG | 0.974, 0.985 | Validated by manufacturer |
| United States of America (Denver) | Xiaofan Jia (University of Colorado)[310] | 2020-07-08 - 2020-10-23 | Local | High | 102 | Adults (18-64 years) | All | Convenience | Household and community samples | 3.9% | NA - - Author designed (CLIA) | 1, 0.999 | Validated by independent authors/third party/non-developers |
| United States of America (Denver) | Xiaofan Jia (University of Colorado)[310] | 2020-07-08 - 2020-10-23 | Local | High | 562 | Children and Youth (0-17 years) | All | Convenience | Household and community samples | 2.8% | NA - - Author designed (CLIA) | 1, 0.999 | Validated by independent authors/third party/non-developers |
| United States of America (New York City) | Teresa Janevic (University of Pennsylvania)[311] | 2020-04-15 - 2020-12-15 | Local | Moderate | 967 | Adults (18-64 years) | Female | Convenience | Pregnant or parturient women | 20% | NA - - Author designed (ELISA) -Spike | 0.95, 1 | Validated by developers |
| United States of America (Washington DC) | Adrienne Sherman (Government of the District of Columbia)[312] | 2020-07-27 - 2020-08-21 | Local | High | 671 | Multiple groups | All | Convenience | Household and community samples | 7.6% | DiaSorin - IgG - Liaison SARS-CoV-2 S1/S2 IgG | 0.974, 0.985 | Validated by manufacturer |
| United States of America (New York City) | Jannae Parrott (New York City Department of Health and Mental Hygiene)[313] | 2020-06-01 - 2020-10-09 | Local | Low | 1,074 | Multiple groups | All | Probability | Household and community samples | 24.3% | DiaSorin - IgG - Liaison SARS-CoV-2 S1/S2 IgG | 0.974, 0.985 | Validated by manufacturer |
| United States of America (St Louis) | Brittany K. Smith (Washington University School of Medicine)[297] | 2020-07-22 - 2020-08-16 | Local | High | 1,052 | Multiple groups | All | Convenience | Residual sera | 5.9% | NA - IgG - Author designed (ELISA) -Spike | 0.982, 0.987 | Validated by developers |
| United States of America (California) | Tim Bruckner (University of California Irvine)[314] | 2020-07-10 - 2020-08-16 | Local | Low | 2,979 | Multiple groups | All | Probability | Household and community samples | 11.5% | NA - IgG, IgM - Author designed (IFA) - MULTIPLEXED | 0.94, 1 | Validated by independent authors/third party/non-developers |
| United States of America (Baton Rouge) | Amy K Feehan (Louisiana State University)[315] | 2020-07-15 - 2020-07-31 | Local | Low | 2,138 | Multiple groups | All | Probability | Household and community samples | 3.6% | Abbott Laboratories - IgG - Abbott Architect SARS-CoV-2 IgG | 0.99, 1 | Validated by manufacturer |
| United States of America (Fullerton) | Jason Yamaki (St. Jude Medical Center)[316] | 2020-07-15 - 2020-07-27 | Local | High | 865 | Multiple groups | All | Sequential | Residual sera | 9.4% | Abbott Laboratories - IgG - Abbott Architect SARS-CoV-2 IgG | 0.99, 1 | Validated by manufacturer |
| United States of America (Lantana, West Palm Beach, Delray Beach, Belle Glade) | Charles Gonik (Wayne State University School of Medicine)[317] | 2020-06-29 - 2020-08-05 | Local | Moderate | 618 | Adults (18-64 years) | Female | Convenience | Pregnant or parturient women | 26.4% | Abbott Laboratories - IgG - Abbott Architect SARS-CoV-2 IgG | 0.99, 1 | Validated by manufacturer |
| United States of America (Iowa) | Haley Steffen (University of Iowa)[318] | 2020-05-01 - 2020-09-22 | Local | Moderate | 1,000 | Adults (18-64 years) | Female | Sequential | Pregnant or parturient women | 5.8% | DiaSorin - IgG - Liaison SARS-CoV-2 S1/S2 IgG,Roche Diagnostics - IgG, IgM, IgA - Elecsys® Anti‐SARS‐CoV‐2 (N),EUROIMMUN - IgG - Anti-SARS-CoV-2 ELISA IgG | N/A | Complex test algorithm: Manual review |
| United States of America (New York City) | Nina Molenaar ( Icahn School of Medicine)[319] | 2020-04-20 - 2020-09-22 | Local | High | 708 | Adults (18-64 years) | Female | Convenience | Pregnant or parturient women | 16.4% | NA - - Author designed (ELISA) -Spike | 0.95, 1 | Validated by developers |
| United States of America (New York City) | Daniel Stadlbauer (Icahn School of Medicine at Mount Sinai)[320] | 2020-06-29 - 2020-07-05 | Local | High | 311 | Multiple groups | All | Sequential | Residual sera | 22.2% | NA - IgG - Author designed (ELISA) -Spike | 0.95, 1 | Validated by developers |
| United States of America (New York City) | Daniel Stadlbauer (Icahn School of Medicine at Mount Sinai)[320] | 2020-06-29 - 2020-07-05 | Local | High | 182 | Multiple groups | All | Sequential | Residual sera | 23.1% | NA - IgG - Author designed (ELISA) -Spike | 0.95, 1 | Validated by developers |
| United States of America | McKaylee Robertson (City University of New York)[321] | 2020-05-15 - 2020-08-15 | National | High | 4,459 | Multiple groups | All | Convenience | Household and community samples | 6.8% | Bio-rad - IgG, IgM, IgA - Platelia SARS-CoV-2 Total Ab assay | 0.993, 0.98 | Validated by manufacturer |
| United States of America (New York City) | Kathy Kamath (Serimmune)[322] | 2020-06-24 - 2020-07-07 | Local | High | 69 | Multiple groups | All | Sequential | Blood donors | 11.6% | Serimmune - IgG, IgM, IgA - Serum Epitope Repertoire Analysis (SERA) | 0.91, 0.993 | Validated by manufacturers |
| United States of America (pennsylvania) | Glenn J. Rapsinski (UPMC Children’s Hospital of Pittsburgh)[323] | 2020-06-22 - 2020-07-03 | Subnational | High | 1,196 | Children and Youth (0-17 years) | All | Sequential | Residual sera | 0.9% | EUROIMMUN - IgG - Anti-SARS-CoV-2 ELISA IgG,Beckman Coulter - IgG - Access SARS-CoV-2 IgG Antibody Test,Siemens - IgG, IgM, IgA - ADVIA Centaur Immunoassay System | N/A | Complex test algorithm: Manual review |
| United States of America (Long Island) | Olga Morozova (Stony Brook University)[324] | 2020-03-25 - 2020-09-18 | Local | High | 1,042 | Multiple groups | All | Convenience | Household and community samples | 21% | Abbott Laboratories - IgG - Abbott Architect SARS-CoV-2 IgG | 0.99, 1 | Validated by manufacturer |
| United States of America (California) | Travis Lim (Centers for Disease and Control )[162] | 2020-03-23 - 2020-08-14 | Local | Low | 4,980 | Multiple groups | All | Probability | Residual sera | 1.9% | NA - IgG - Author designed (ELISA) -Spike | 0.96, 0.993 | Validated by developers |
| United States of America (New York City) | Travis Lim (Centers for Disease and Control )[162] | 2020-03-23 - 2020-08-14 | Local | Low | 7,746 | Multiple groups | All | Probability | Residual sera | 17.2% | NA - IgG, IgM, IgA - Author designed (ELISA) -Spike | 0.96, 0.993 | Validated by developers |
| United States of America (Philadelphia) | Travis Lim (Centers for Disease and Control )[162] | 2020-03-23 - 2020-08-14 | Local | Low | 8,657 | Multiple groups | All | Probability | Residual sera | 4.3% | NA - IgG, IgM, IgA - Author designed (ELISA) -Spike | 0.96, 0.993 | Validated by developers |
| United States of America (North Carolina) | Amir Barzin (University of North Carolina Chapel Hill)[325] | 2020-06-01 - 2020-06-04 | Subnational | High | 1,449 | Adults (18-64 years) | All | Convenience | Residual sera | 0% | Abbott Laboratories - IgG - Abbott Architect SARS-CoV-2 IgG | 0.99, 1 | Validated by manufacturer |
| United States of America (Pittsburgh) | Daniel Geisler (University of Pittsburgh School of Medicine)[326] | 2020-04-27 - 2020-07-04 | Local | High | 2,338 | Children and Youth (0-17 years) | All | Sequential | Residual sera | 3.6% | EUROIMMUN - IgG - Anti-SARS-CoV-2 ELISA IgG | 0.9, 1 | Validated by manufacturer |
| United States of America (New York) | Josh Reifer (Sherman Abrams Laboratory )[327] | 2020-05-15 - 2020-06-15 | Local | High | 28,523 | Multiple groups | All | Convenience | Residual sera | 44% | DiaSorin - IgG - Liaison SARS-CoV-2 S1/S2 IgG | 0.974, 0.985 | Validated by manufacturer |
| United States of America (New York) | Cara Staszewski (North Shore University Hospital)[302] | 2020-05-15 - 2020-06-15 | Local | High | 149 | Adults (18-64 years) | Female | Sequential | Pregnant or parturient women | 16.1% | Gold Standard Diagnostic - IgG - SARS-CoV-2 IgG ELISA Assay | 1, 1 | Validated by manufacturer |
| United States of America | Robert Stout (Clinical Reference Laboratory Inc.)[300] | 2020-05-12 - 2020-06-15 | National | High | 50,072 | Multiple groups | All | Convenience | Residual sera | 3% | Roche Diagnostics - IgG, IgM, IgA - Elecsys® Anti‐SARS‐CoV‐2 (N) | 1, 0.995 | Validated by manufacturer |
| United States of America (Long Island) | Minesh Khatri (New York University Long Island School of Medicine)[328] | 2020-02-01 - 2020-08-25 | Local | High | 343 | Multiple groups | All | Convenience | Residual sera | 25.7% | Abbott Laboratories - IgG - Abbott Architect SARS-CoV-2 IgG | 0.99, 1 | Validated by manufacturer |
| United States of America (Louisiana) | Amy K Feehan (Oschner Clinic Foundation)[329] | 2020-05-09 - 2020-05-15 | Local | Low | 2,640 | Adults (18-64 years) | All | Probability | Household and community samples | 6.9% | Abbott Laboratories - IgG - Abbott Architect SARS-CoV-2 IgG | 0.99, 1 | Validated by manufacturer |
| United States of America (San Francisco) | Isobel Routledge (University of California San Francisco)[330] | 2020-03-28 - 2020-06-26 | Local | Moderate | 4,735 | Multiple groups | All | Stratified non-probability | Residual sera | 4.1% | NA - - Author designed (Luminex),NA - IgG - Author designed (ELISA) -Unknown | N/A | Complex test algorithm: Manual review |
| United States of America (pennsylvania) | Glenn J. Rapsinski (UPMC Children’s Hospital of Pittsburgh)[323] | 2020-04-27 - 2020-05-19 | Subnational | High | 1,142 | Children and Youth (0-17 years) | All | Sequential | Residual sera | 0.5% | EUROIMMUN - IgG - Anti-SARS-CoV-2 ELISA IgG,Beckman Coulter - IgG - Access SARS-CoV-2 IgG Antibody Test,Siemens - IgG, IgM, IgA - ADVIA Centaur Immunoassay System | N/A | Complex test algorithm: Manual review |
| United States of America (Philadelphia) | Dustin Flannery (University of Pennsylvania)[331] | 2020-04-04 - 2020-06-03 | Local | Moderate | 1,293 | Adults (18-64 years) | Female | Sequential | Pregnant or parturient women | 6.2% | NA - IgG, IgM - Author designed (ELISA) -Spike | 1, 0.989 | Validated by independent authors/third party/non-developers |
| United States of America (St Louis) | Brittany Smith (Washington University School of Medicine)[332] | 2020-04-27 - 2020-05-12 | Local | High | 503 | Multiple groups | All | Sequential | Residual sera | 4.2% | NA - IgG - Author designed (ELISA) -Spike | 0.982, 0.987 | Validated by independent authors/third party/non-developers |
| United States of America (Rhode Island) | Daniel Nesbitt (Lindsley F Kimball Research Institute)[333] | 2020-04-27 - 2020-05-11 | Subnational | High | 1,996 | Multiple groups | All | Sequential | Blood donors | 0.7% | Ortho Clinical Diagnostics Inc. - IgG - VITROS Immunodiagnostic Products Anti-SARS-CoV-2 IgG | 1, 1 | Validated by manufacturer |
| United States of America (Georgia) | Holly Biggs (CDC COVID-19 Response Team)[334] | 2020-04-28 - 2020-05-03 | Local | Low | 696 | Multiple groups | All | Probability | Household and community samples | 2.7% | Ortho Clinical Diagnostics Inc. - IgG, IgM, IgA - VITROS Immunodiagnostic Products Anti-SARS-CoV-2 Total | 1, 1 | Validated by manufacturer |
| United States of America (Atlanta) | Kristina Bajema (Centers for Disease Control and Prevention)[335] | 2020-04-27 - 2020-05-01 | Local | High | 1,343 | Multiple groups | All | Convenience | Residual sera | 4.2% | NA - IgG, IgM, IgA - Author designed (ELISA) -Spike | 0.96, 0.993 | Validated by developers |
| United States of America (St Louis) | Brittany Smith (Washington University School of Medicine)[332] | 2020-04-14 - 2020-05-08 | Local | High | 555 | Children and Youth (0-17 years) | All | Sequential | Residual sera | 2.7% | NA - IgG - Author designed (ELISA) -Spike | 0.982, 0.987 | Validated by independent authors/third party/non-developers |
| United States of America (Baltimore) | Oliver Laeyendecker (Johns Hopkins University )[336] | 2020-03-16 - 2020-05-31 | Local | High | 3,255 | Multiple groups | All | Sequential | Residual sera | 3.5% | Hangzhou Biotest Biotech Co. Ltd - IgG, IgM - RightSign COVID-19 IgG/IgM Rapid Test Cassette,EUROIMMUN - IgG - Anti-SARS-CoV-2 ELISA IgG | 0.991, 1 | Complex test algorithm: Manual calculation |
| United States of America (California) | Neeraj Sood (Stanford University School of Medicine)[337] | 2020-04-10 - 2020-04-14 | Local | Low | 863 | Multiple groups | All | Probability | Household and community samples | 4.3% | Premier Biotech - IgG, IgM - COVID-19 IgG/IgM Rapid Test Cassette | 1, 1 | Validated by manufacturer |
| United States of America (Seattle) | Jennifer Rathe (University of Washington)[338] | 2020-03-26 - 2020-04-15 | Local | High | 100 | Multiple groups | All | Probability | Household and community samples | 4% | NA - - Author designed (Neutralization Assay),Abbott Laboratories - IgG - Abbott Architect SARS-CoV-2 IgG,NA - IgG, IgM, IgA - Author designed (ELISA) -Spike | N/A | Complex test algorithm: Manual review |
| United States of America (California) | Eran Bendavid (Stanford University School of Medicine)[339] | 2020-04-03 - 2020-04-04 | Local | High | 3,328 | Multiple groups | All | Stratified non-probability | Household and community samples | 1.5% | Premier Biotech - IgG, IgM - COVID-19 IgG/IgM Rapid Test Cassette | 1, 1 | Validated by manufacturer |
| United States of America (Seattle) | Adam S Dingens (Fred Hutchinson Cancer Research Center)[340] | 2020-03-03 - 2020-04-24 | Local | High | 1,076 | Children and Youth (0-17 years) | All | Convenience | Residual sera | 0.9% | Abbott Laboratories - IgG - Abbott Architect SARS-CoV-2 IgG,NA - IgG - Author designed (ELISA) -Spike | N/A | Complex test algorithm: Manual review |
| United States of America (San Francisco) | Dianna Ng (University of California, San Francisco)[341] | 2020-03-01 - 2020-03-31 | Local | High | 1,000 | Multiple groups | All | Sequential | Blood donors | 0.1% | Abbott Laboratories - IgG - Abbott Architect SARS-CoV-2 IgG,Ortho Clinical Diagnostics Inc. - IgG - VITROS Immunodiagnostic Products Anti-SARS-CoV-2 IgG,NA - - Author designed (Neutralization Assay) | N/A | Complex test algorithm: Manual review |

###### Eastern Mediterranean region

| Country (Location) | Author (Organization) | Sampling Dates (YMD) | Geographic scope | Overall risk of bias | Sample size | Age | Sex | Sampling method | Sample frame | Seroprevalence (95% CI) | Test Manufacturers - Isotypes - Names | Test Sens, Spec | Sens, Spec Source |
| --- | --- | --- | --- | --- | --- | --- | --- | --- | --- | --- | --- | --- | --- |
| Egypt - HRP (Cairo) | Sahar Abdelmaksoud (Ain-Shams University)[342] | 2020-09-06 - 2020-10-31 | Local | High | 100 | Adults (18-64 years) | All | Probability | Blood donors | 38% | AMEDA Labordiagnostik GmbH - IgG, IgM - AMP Rapid Test SARS-CoV-2 IgG/IgM | 1, 0.975 | Validated by manufacturer |
| Egypt - HRP (Cairo) | Samia Girgis (Ain-Shams University)[343] | 2020-05-05 - 2020-10-31 | Local | High | 2,927 | Multiple groups | All | Sequential | Residual sera | 30% | Roche Diagnostics - IgG, IgM, IgA - Elecsys® Anti‐SARS‐CoV‐2 (N) | 1, 0.995 | Validated by manufacturer |
| Iran (Islamic Republic of) - HRP (Zahedan) | Zahra Mohammadi (Tehran University of Medical Sciences)[344] | 2020-07-01 - 2020-07-31 | Local | High | 134 | Adults (18-64 years) | Male | Probability | Household and community samples | 4.8% | Pishtaz Diagnostics Iran - IgG, IgM - ELISA kit Pishtaz Teb Diagnostics | 0.941, 0.983 | Validated by manufacturer |
| Iran (Islamic Republic of) - HRP (Ardabil) | Hamed Zandian (Ardabil University of Medical Sciences)[345] | 2020-05-20 - 2020-06-07 | Local | Moderate | 1,013 | Multiple groups | All | Probability | Household and community samples | 12.1% | Pishtaz Diagnostics Iran - IgG, IgM - ELISA kit Pishtaz Teb Diagnostics | 0.941, 0.983 | Validated by manufacturer |
| Iraq - HRP (Duhok) | Nawfal Hussein (University of Duhok)[346] | 2021-01-10 - 2021-01-30 | Local | Moderate | 743 | Multiple groups | All | Probability | Household and community samples | 62.6% | Roche Diagnostics - IgG, IgM, IgA - Elecsys® Anti‐SARS‐CoV‐2 (N) | 1, 0.995 | Validated by manufacturer |
| Jordan - HRP | Maher Sughayer (King Hussein Cancer Center)[347] | 2021-06-01 - 2021-06-30 | National | High | 536 | Adults (18-64 years) | All | Sequential | Blood donors | 74.4% | Roche Diagnostics - IgG, IgM, IgA - Elecsys® Anti‐SARS‐CoV‐2 (S) | 0.988, 1 | Validated by manufacturer |
| Jordan - HRP | Maher Sughayer (King Hussein Cancer Center)[348] | 2021-01-28 - 2021-02-05 | National | High | 292 | Adults (18-64 years) | All | Sequential | Blood donors | 27.4% | Roche Diagnostics - IgG, IgM, IgA - Elecsys® Anti‐SARS‐CoV‐2 (N) | 1, 0.995 | Validated by manufacturer |
| Jordan - HRP | Maher Sughayer (King Hussein Cancer Center)[348] | 2020-09-01 - 2020-09-15 | National | High | 348 | Adults (18-64 years) | All | Sequential | Blood donors | 0% | Roche Diagnostics - IgG, IgM, IgA - Elecsys® Anti‐SARS‐CoV‐2 (N) | 1, 0.995 | Validated by manufacturer |
| Jordan - HRP | Maher Sughayer (King Hussein Cancer Center)[348] | 2020-01-15 - 2020-06-15 | National | High | 736 | Adults (18-64 years) | All | Sequential | Blood donors | 0% | Roche Diagnostics - IgG, IgM, IgA - Elecsys® Anti‐SARS‐CoV‐2 (N) | 1, 0.995 | Validated by manufacturer |
| occupied Palestinian territory, including east Jerusalem - HRP (West Bank) | Nouar Qutob (Arab American University )[349] | 2020-06-15 - 2020-06-30 | Local | Low | 1,355 | Multiple groups | All | Probability | Household and community samples | 0% | Roche Diagnostics - IgG, IgM, IgA - Elecsys® Anti‐SARS‐CoV‐2 (N) | 1, 0.995 | Validated by manufacturer |
| occupied Palestinian territory, including east Jerusalem - HRP (West Bank) | Nouar Qutob (Arab American University )[349] | 2020-05-01 - 2020-07-09 | Local | High | 1,136 | Multiple groups | All | Convenience | Residual sera | 0.3% | Roche Diagnostics - IgG, IgM, IgA - Elecsys® Anti‐SARS‐CoV‐2 (N) | 1, 0.995 | Validated by manufacturer |
| Pakistan - HRP (District East) | Muhammed Imran Nisar (Aga Khan University)[350] | 2020-08-17 - 2020-08-22 | Local | Moderate | 500 | Multiple groups | All | Probability | Household and community samples | 24% | Roche Diagnostics - IgG, IgM, IgA - Elecsys® Anti‐SARS‐CoV‐2 (N) | 1, 0.995 | Validated by manufacturer |
| Pakistan - HRP (District Malir) | Muhammed Imran Nisar (Aga Khan University)[350] | 2020-08-17 - 2020-08-22 | Local | Moderate | 501 | Multiple groups | All | Probability | Household and community samples | 16% | Roche Diagnostics - IgG, IgM, IgA - Elecsys® Anti‐SARS‐CoV‐2 (N) | 1, 0.995 | Validated by manufacturer |
| Pakistan - HRP | Sheeba Ansari (Liaquat University)[351] | 2020-06-15 - 2020-08-15 | Local | High | 728 | Multiple groups | All | Convenience | Residual sera | 10.8% | NA - - Not reported/ Unable to specify | 1, 0.998 | Validated by manufacturers |
| Pakistan - HRP (District East) | Muhammed Imran Nisar (Aga Khan University)[350] | 2020-06-25 - 2020-07-11 | Local | Moderate | 500 | Multiple groups | All | Probability | Household and community samples | 20% | Roche Diagnostics - IgG, IgM, IgA - Elecsys® Anti‐SARS‐CoV‐2 (N) | 1, 0.995 | Validated by manufacturer |
| Pakistan - HRP (District Malir) | Muhammed Imran Nisar (Aga Khan University)[350] | 2020-06-25 - 2020-07-11 | Local | Moderate | 504 | Multiple groups | All | Probability | Household and community samples | 12.7% | Roche Diagnostics - IgG, IgM, IgA - Elecsys® Anti‐SARS‐CoV‐2 (N) | 1, 0.995 | Validated by manufacturer |
| Pakistan - HRP | Wajiha Javed (Getz Pharma)[352] | 2020-06-22 - 2020-07-06 | Subnational | High | 24,210 | Adults (18-64 years) | All | Probability | Household and community samples | 17.5% | Getz Pharma - IgG, IgM - IgG/IgM Test Kit (Colloidal gold) | 0.953, 0.987 | Validated by manufacturer |
| Pakistan - HRP (District East) | Muhammed Imran Nisar (Aga Khan University)[350] | 2020-04-15 - 2020-04-25 | Local | Moderate | 500 | Multiple groups | All | Probability | Household and community samples | 0.4% | Roche Diagnostics - IgG, IgM, IgA - Elecsys® Anti‐SARS‐CoV‐2 (N) | 1, 0.995 | Validated by manufacturer |
| Pakistan - HRP (District Malir) | Muhammed Imran Nisar (Aga Khan University)[350] | 2020-04-15 - 2020-04-25 | Local | Moderate | 500 | Multiple groups | All | Probability | Household and community samples | 0% | Roche Diagnostics - IgG, IgM, IgA - Elecsys® Anti‐SARS‐CoV‐2 (N) | 1, 0.995 | Validated by manufacturer |
| Saudi Arabia (Riyadh) | Mohammed Alosaimi (King Saud University)[353] | 2020-11-22 - 2020-12-17 | Local | High | 515 | Multiple groups | All | Convenience | Blood donors | 12.2% | Abbott Laboratories - IgG - Abbott Architect SARS-CoV-2 IgG | 0.99, 1 | Validated by manufacturer |
| Saudi Arabia | Sami Almudarra (Saudi Ministry of Health)[354] | 2020-08-01 - 2020-08-31 | Local | Moderate | 5,629 | Multiple groups | All | Probability | Household and community samples | 50.2% | Roche Diagnostics - IgG, IgM, IgA - Elecsys® Anti‐SARS‐CoV‐2 (N) | 1, 0.995 | Validated by manufacturer |
| United Arab Emirates (Dubai) | May Raouf (Dubai Health Authority)[355] | 2020-08-15 - 2020-12-15 | Local | Moderate | 1,418 | Adults (18-64 years) | All | Probability | Blood donors | 13.5% | Abbott Laboratories - IgG - Abbott Architect SARS-CoV-2 IgG | 0.99, 1 | Validated by manufacturer |
| Yemen - HRP (Aden) | Abdulla Bin-Ghouth (World Health Organization Country Office, Yemen)[356] | 2020-11-15 - 2020-12-15 | Local | Moderate | 2,001 | Multiple groups | All | Probability | Household and community samples | 27.4% | Healgen - IgG, IgM - COVID-19 IgG/IgM Rapid Test Cassette,Beijing Wantai Biological - IgG, IgM, IgA - Wantai SARS-CoV-2 Total Ab ELISA | 0.907, 1 | Complex test algorithm: Manual calculation |

###### Europe region

| Country (Location) | Author (Organization) | Sampling Dates (YMD) | Geographic scope | Overall risk of bias | Sample size | Age | Sex | Sampling method | Sample frame | Seroprevalence (95% CI) | Test Manufacturers - Isotypes - Names | Test Sens, Spec | Sens, Spec Source |
| --- | --- | --- | --- | --- | --- | --- | --- | --- | --- | --- | --- | --- | --- |
| Albania (Tirana) | Genc Sulcebe (University of Medicine and University Hospital Center of Tirana)[357] | 2020-12-21 - 2020-12-28 | Local | Moderate | 815 | Adults (18-64 years) | All | Probability | Household and community samples | 48.2% | EUROIMMUN - IgG - Anti-SARS-CoV-2 ELISA IgG | 0.9, 1 | Validated by manufacturer |
| Albania (Tirana) | Genc Sulcebe (University of Medicine and University Hospital Center of Tirana)[357] | 2020-06-27 - 2020-07-03 | Local | High | 266 | Adults (18-64 years) | All | Probability | Household and community samples | 7.5% | EUROIMMUN - IgG - Anti-SARS-CoV-2 ELISA IgG | 0.9, 1 | Validated by manufacturer |
| Bosnia and Herzegovina (Sarajevo) | Adna Asic (International Burch University)[358] | 2020-04-21 - 2020-07-17 | Local | High | 1,121 | Multiple groups | All | Convenience | Residual sera | 4.3% | Dynamiker Biotechnology (Tianjin) Co. Ltd - IgG, IgM - 2019-nCoV IgG/IgM Rapid Test | 0.957, 0.991 | Validated by manufacturer |
| Bosnia and Herzegovina (Sarajevo) | Adna Asic (International Burch University)[358] | 2020-04-15 - 2020-05-15 | Local | High | 1,720 | Multiple groups | All | Convenience | Residual sera | 3.4% | Artron Laboratories Inc. - IgG, IgM - One Step Novel Coronavirus (COVID-19) IgM/IgG Antibody Test | 0.972, 0.978 | Validated by manufacturer |
| Bulgaria (Varna) | Denitsa Tsaneva-Damyanova (Medical Diagnostic Laboratory “STATUS” )[359] | 2020-03-26 - 2020-04-20 | Local | High | 586 | Multiple groups | All | Convenience | Residual sera | 4.8% | Zhejiang Orient Gene Biotech - IgG, IgM - COVID-19 IgG/IgM Rapid Test | 0.972, 1 | Validated by manufacturer |
| Kazakhstan | Yuliya Semenova (Nazarbayev University School of Medicine)[360] | 2020-07-16 - 2021-07-07 | National | High | 85,346 | Multiple groups | All | Sequential | Residual sera | 63% | Vector BEST - IgG, IgM - SARS- CoV-2-IgG-EIA-BEST and SARS-CoV-2- IgM-EIA-BEST | 0.9, 1 | Validated by independent authors/third party/non-developers |
| Kazakhstan (Almaty) | Manar Smagul (Kazakh National Medical University)[361] | 2020-10-24 - 2021-01-11 | Local | Low | 4,461 | Multiple groups | All | Probability | Household and community samples | 57% | Beijing Wantai Biological - IgG, IgM, IgA - Wantai SARS-CoV-2 Total Ab ELISA | 0.967, 0.975 | Validated by manufacturer |
| Kazakhstan (Kostanay) | Manar Smagul (Kazakh National Medical University)[361] | 2020-10-24 - 2021-01-11 | Local | Low | 614 | Multiple groups | All | Probability | Household and community samples | 39.4% | Beijing Wantai Biological - IgG, IgM, IgA - Wantai SARS-CoV-2 Total Ab ELISA | 0.967, 0.975 | Validated by manufacturer |
| Kazakhstan (Oskemen) | Manar Smagul (Kazakh National Medical University)[361] | 2020-10-24 - 2021-01-11 | Local | Low | 664 | Multiple groups | All | Probability | Household and community samples | 60.7% | Beijing Wantai Biological - IgG, IgM, IgA - Wantai SARS-CoV-2 Total Ab ELISA | 0.967, 0.975 | Validated by manufacturer |
| North Macedonia (Skopje) | Tatjana Makarovska Bojadjieva ( Institute for Transfusion Medicine)[362] | 2020-11-15 - 2021-03-15 | Local | High | 9,773 | Multiple groups | All | Sequential | Blood donors | 25% | Abbott Laboratories - IgG - Abbott Architect SARS-CoV-2 IgG | 0.99, 1 | Validated by manufacturer |
| Russian Federation (Krasnodar) | Vladimir Gorodin (Kuban State Medical University)[363] | 2020-10-26 - 2020-12-30 | Local | High | 806 | Adults (18-64 years) | All | Convenience | Household and community samples | 13.2% | Beckman Coulter - IgG - Access SARS-CoV-2 IgG Antibody Test | 1, 0.998 | Validated by manufacturer |
| Russian Federation (Saint Petersburg) | Anton Barchuk (Smorodinova Research Institute of Influenza)[364] | 2020-10-12 - 2020-12-06 | Local | Low | 1,322 | Multiple groups | All | Probability | Household and community samples | 21% | Genetico - IgG, IgM, IgA - CoronaPass total antibodies test | 0.911, 0.9 | Validated by manufacturer |
| Russian Federation (Saint Petersburg) | Anton Barchuk (Smorodinova Research Institute of Influenza)[364] | 2020-07-20 - 2020-08-08 | Local | Moderate | 474 | Multiple groups | All | Probability | Household and community samples | 12.2% | Genetico - IgG, IgM, IgA - CoronaPass total antibodies test | 0.911, 0.9 | Validated by manufacturer |
| Russian Federation (Saint Petersburg) | Anton Barchuk (European University at St. Petersburg )[365] | 2020-05-27 - 2020-06-26 | Local | Low | 990 | Multiple groups | All | Probability | Household and community samples | 7.4% | Abbott Laboratories - IgG - Abbott Architect SARS-CoV-2 IgG | 0.99, 1 | Validated by manufacturer |
| Turkey - HRP (Ankara) | Ozge Yucel Celik (Etlik Zübeyde Hanim Women’s Health Care, Training, and Research Hospital)[366] | 2020-06-01 - 2020-07-31 | Local | High | 230 | Adults (18-64 years) | All | Sequential | Pregnant or parturient women | 0% | Assure Tech. (Hangzhou) Co. Ltd - IgG, IgM - COVID-19 IgG/IgM Rapid Test Device | 1, 0.988 | Validated by manufacturer |
| Turkey - HRP (Ankara) | Ozge Yucel Celik (Etlik Zübeyde Hanim Women’s Health Care, Training, and Research Hospital)[366] | 2020-06-01 - 2020-07-31 | Local | High | 205 | Adults (18-64 years) | All | Sequential | Pregnant or parturient women | 2.9% | Assure Tech. (Hangzhou) Co. Ltd - IgG, IgM - COVID-19 IgG/IgM Rapid Test Device | 1, 0.988 | Validated by manufacturer |
| Austria (Tyrol) | Anita Siller (Medical University of Innsbruck )[367] | 2021-03-19 - 2021-09-30 | Subnational | High | 19,792 | Multiple groups | All | Sequential | Blood donors | 63.2% | Abbott Laboratories - IgG - SARS-CoV-2 IgG II Quant | 0.994, 0.996 | Validated by manufacturer |
| Austria (Vienna) | M. Riesenhuber (Universitätsklinik für Innere Medizin II )[368] | 2021-02-08 - 2021-02-22 | Local | High | 3,082 | Multiple groups | All | Sequential | Residual sera | 9.3% | Roche Diagnostics - IgG, IgM, IgA - Elecsys® Anti‐SARS‐CoV‐2 (N) | 1, 0.995 | Validated by manufacturer |
| Austria (Weißenkirchen in der Wachau) | Heike Rebholz (Danube Private University)[369] | 2021-02-13 - 2021-02-13 | Local | High | 552 | Multiple groups | All | Convenience | Household and community samples | 25% | EUROIMMUN - IgG - Anti-SARS-CoV-2 ELISA IgG | 0.9, 1 | Validated by manufacturer |
| Austria | Lisa Weidner ( University Hospital of Salzburg)[370] | 2020-11-05 - 2020-12-04 | National | High | 3,311 | Multiple groups | All | Convenience | Blood donors | 5.4% | Roche Diagnostics - IgG, IgM, IgA - Elecsys® Anti‐SARS‐CoV‐2 (N) | 1, 0.995 | Validated by manufacturer |
| Austria (Ischgl) | Wegene Borena (Medical University Of Innsbruck)[371] | 2020-11-02 - 2020-11-08 | Local | Moderate | 801 | Adults (18-64 years) | All | Convenience | Household and community samples | 45.4% | EUROIMMUN - IgA - Anti-SARS-CoV-2 ELISA IgA,EUROIMMUN - IgG - Anti-SARS-CoV-2 ELISA IgG,Abbott Laboratories - IgG - Abbott Architect SARS-CoV-2 IgG,Roche Diagnostics - IgG, IgM, IgA - Elecsys® Anti‐SARS‐CoV‐2 (N),NA - - Author designed (Neutralization Assay) | N/A | Complex test algorithm: Manual review |
| Austria (Weißenkirchen) | Dennis Ladage (Danube Private University)[372] | 2020-06-01 - 2020-06-30 | Local | Moderate | 835 | Multiple groups | All | Convenience | Household and community samples | 8.5% | EUROIMMUN - IgG - Anti-SARS-CoV-2 ELISA IgG | 0.9, 1 | Validated by manufacturer |
| Austria (Ischgl) | Jens Lehmann ( Medical University of Innsbruck)[373] | 2020-04-21 - 2020-04-27 | Local | High | 451 | Multiple groups | All | Probability | Household and community samples | 43.7% | Abbott Laboratories - IgG - Abbott Architect SARS-CoV-2 IgG,EUROIMMUN - IgA - Anti-SARS-CoV-2 ELISA IgA,EUROIMMUN - IgG - Anti-SARS-CoV-2 ELISA IgG,NA - - Author designed (Neutralization Assay) | N/A | Complex test algorithm: Manual review |
| Belgium (Ghent) | Laura Heireman (Ghent University Hospital)[374] | 2020-01-15 - 2020-11-15 | Local | High | 265 | Adults (18-64 years) | Female | Convenience | Pregnant or parturient women | 1.5% | EUROIMMUN - IgG - Anti-SARS-CoV-2 ELISA IgG,Epitope Diagnostics, Inc. - IgG - EDI™ Novel Coronavirus COVID-19 IgG ELISA Kit | 0.998, 0.998 | Complex test algorithm: Manual calculation |
| Belgium | Peter van Dam (Antwerp University Hospital)[375] | 2020-03-24 - 2020-05-31 | Local | High | 90 | Multiple groups | All | Sequential | Residual sera | 5.6% | DiaSorin - IgG - Liaison SARS-CoV-2 S1/S2 IgG | 0.974, 0.985 | Validated by manufacturer |
| Croatia | Tatjana Vilibic-Cavlek (Croatian Institute of Public Health)[376] | 2020-12-15 - 2021-02-15 | National | High | 1,436 | Multiple groups | All | Convenience | Multiple general populations | 25.1% | Vircell S.L. - IgG - COVID-19 ELISA IgG,NA - - Author designed (Neutralization Assay) | N/A | Complex test algorithm: Manual review |
| Croatia | Tatjana Vilibic-Cavlek (Croatian Institute of Public Health)[376] | 2020-05-15 - 2020-07-15 | National | High | 1,088 | Multiple groups | All | Convenience | Multiple general populations | 2.2% | Vircell S.L. - IgG - COVID-19 ELISA IgG,NA - - Author designed (Neutralization Assay) | N/A | Complex test algorithm: Manual review |
| Croatia (Zagreb) | Jasna Lenicek Krleza (Children’s Hospital Zagreb)[377] | 2020-05-13 - 2020-05-29 | Local | High | 240 | Children and Youth (0-17 years) | All | Convenience | Residual sera | 2.9% | Vircell S.L. - IgG - COVID-19 ELISA IgG,Vircell S.L. - IgM, IgA - COVID-19 ELISA IgM/IgA,NA - - Author designed (Neutralization Assay) | N/A | Complex test algorithm: Manual review |
| Czechia | Pavel Piler (Masaryk University)[378] | 2021-02-01 - 2021-03-31 | National | High | 19,548 | Multiple groups | All | Convenience | Household and community samples | 51.4% | DiaSorin - IgG - Liaison SARS-CoV-2 S1/S2 IgG,Abbott Laboratories - IgG - SARS-CoV-2 IgG II Quant | 1, 0.981 | Complex test algorithm: Manual calculation |
| Czechia | Pavel Piler (Masaryk University)[378] | 2020-12-01 - 2021-01-31 | National | High | 6,880 | Multiple groups | All | Convenience | Household and community samples | 43.4% | DiaSorin - IgG - Liaison SARS-CoV-2 S1/S2 IgG,Abbott Laboratories - IgG - SARS-CoV-2 IgG II Quant | 1, 0.981 | Complex test algorithm: Manual calculation |
| Czechia | Pavel Piler (Masaryk University)[378] | 2020-10-01 - 2020-11-30 | National | High | 3,626 | Multiple groups | All | Convenience | Household and community samples | 28.3% | DiaSorin - IgG - Liaison SARS-CoV-2 S1/S2 IgG,Abbott Laboratories - IgG - SARS-CoV-2 IgG II Quant | 1, 0.981 | Complex test algorithm: Manual calculation |
| Czechia (Prague) | Marketa Bloomfield (Charles University and Thomayer’s Hospital)[379] | 2020-07-03 - 2020-08-19 | Local | High | 200 | Children and Youth (0-17 years) | All | Sequential | Residual sera | 0% | Roche Diagnostics - IgG, IgM, IgA - Elecsys® Anti‐SARS‐CoV‐2 (N) | 1, 0.995 | Validated by manufacturer |
| Denmark (Capital Region and Region Zealand) | Maren J H Rytter (Slagelse Sygehus)[174] | 2020-06-22 - 2020-07-03 | Subnational | High | 1,033 | Children and Youth (0-17 years) | All | Convenience | Residual sera | 1.6% | Zhuhai Livzon Diagnostics Inc - IgG, IgM - 2019-nCoV IgG/IgM Antibody Detection Kit | 0.906, 0.992 | Validated by manufacturer |
| Denmark (Eastern Central Denmark Region) | Senne Jespersen (Aarhus University Hospital)[380] | 2020-06-17 - 2020-06-30 | Subnational | High | 180 | Adults (18-64 years) | All | Convenience | Blood donors | 0.6% | Beijing Wantai Biological - IgG, IgM, IgA - Wantai Rapid Test for Total Antibody to SARS-CoV-2 | 0.947, 0.989 | Validated by manufacturer |
| Denmark (Western Central Denmark Region) | Senne Jespersen (Aarhus University Hospital)[380] | 2020-06-17 - 2020-06-30 | Subnational | High | 180 | Adults (18-64 years) | All | Convenience | Blood donors | 1.2% | Beijing Wantai Biological - IgG, IgM, IgA - Wantai Rapid Test for Total Antibody to SARS-CoV-2 | 0.947, 0.989 | Validated by manufacturer |
| Denmark | Ole Birger Pedersen (Copenhagen University Hospital Rigshospitalet)[64] | 2020-06-01 - 2020-06-26 | National | High | 1,110 | Adults (18-64 years) | All | Sequential | Blood donors | 3% | Beijing Wantai Biological - IgG, IgM, IgA - Wantai SARS-CoV-2 Total Ab ELISA | 0.967, 0.975 | Validated by manufacturer |
| Denmark (Copenhagen) | Pia Egerup (Copenhagen University Hospital Hvidovre)[381] | 2020-04-04 - 2020-07-03 | Local | Moderate | 1,313 | Adults (18-64 years) | Female | Sequential | Pregnant or parturient women | 2.1% | Shenzhen Yhlo Biotech Co. Ltd - IgG, IgM - iFlash-SARS-CoV-2 IgM/IgG | 0.94, 0.993 | Validated by independent authors/third party/non-developers |
| Estonia | Hiie Soeorg (University of Tartu)[382] | 2021-02-08 - 2021-03-25 | National | High | 2,517 | Multiple groups | All | Stratified non-probability | Residual sera | 20.1% | Abbott Laboratories - IgG - SARS-CoV-2 IgG II Quant | 0.994, 0.996 | Validated by manufacturer |
| Estonia | Piret Veerus (University of Tartu)[383] | 2020-05-04 - 2020-06-10 | National | High | 433 | Adults (18-64 years) | Female | Convenience | Pregnant or parturient women | 0.5% | Abbott Laboratories - IgG - Abbott Architect SARS-CoV-2 IgG | 0.99, 1 | Validated by manufacturer |
| Finland | Finnish institute for health and welfare (Finnish Institute for Health and Welfare)[66] | 2020-09-20 - 2020-09-26 | National | High | 56 | Adults (18-64 years) | All | Probability | Household and community samples | 1.8% | NA - - Author designed (Neutralization Assay),NA - - Author designed (Multiplex) | N/A | Complex test algorithm: Manual review |
| Finland | Finnish institute for health and welfare (Finnish Institute for Health and Welfare)[66] | 2020-09-13 - 2020-09-19 | National | High | 48 | Adults (18-64 years) | All | Probability | Household and community samples | 2.1% | NA - - Author designed (Neutralization Assay),NA - - Author designed (Multiplex) | N/A | Complex test algorithm: Manual review |
| Finland | Finnish institute for health and welfare (Finnish Institute for Health and Welfare)[66] | 2020-09-06 - 2020-09-12 | National | High | 54 | Adults (18-64 years) | All | Probability | Household and community samples | 0% | NA - - Author designed (Neutralization Assay),NA - - Author designed (Multiplex) | N/A | Complex test algorithm: Manual review |
| Finland | Finnish institute for health and welfare (Finnish Institute for Health and Welfare)[66] | 2020-08-30 - 2020-09-05 | National | High | 63 | Adults (18-64 years) | All | Probability | Household and community samples | 0% | NA - - Author designed (Neutralization Assay),NA - - Author designed (Multiplex) | N/A | Complex test algorithm: Manual review |
| Finland | Finnish institute for health and welfare (Finnish Institute for Health and Welfare)[66] | 2020-08-23 - 2020-08-29 | National | High | 48 | Adults (18-64 years) | All | Probability | Household and community samples | 2.1% | NA - - Author designed (Neutralization Assay),NA - - Author designed (Multiplex) | N/A | Complex test algorithm: Manual review |
| Finland | Finnish institute for health and welfare (Finnish Institute for Health and Welfare)[66] | 2020-08-16 - 2020-08-22 | National | High | 63 | Adults (18-64 years) | All | Probability | Household and community samples | 0% | NA - - Author designed (Neutralization Assay),NA - - Author designed (Multiplex) | N/A | Complex test algorithm: Manual review |
| Finland | Finnish institute for health and welfare (Finnish Institute for Health and Welfare)[66] | 2020-08-09 - 2020-08-15 | National | High | 64 | Adults (18-64 years) | All | Probability | Household and community samples | 0% | NA - - Author designed (Neutralization Assay),NA - - Author designed (Multiplex) | N/A | Complex test algorithm: Manual review |
| Finland | Finnish institute for health and welfare (Finnish Institute for Health and Welfare)[66] | 2020-08-02 - 2020-08-08 | National | High | 49 | Adults (18-64 years) | All | Probability | Household and community samples | 2% | NA - - Author designed (Neutralization Assay),NA - - Author designed (Multiplex) | N/A | Complex test algorithm: Manual review |
| Finland | Finnish institute for health and welfare (Finnish Institute for Health and Welfare)[66] | 2020-07-26 - 2020-08-01 | National | High | 75 | Adults (18-64 years) | All | Probability | Household and community samples | 1.3% | NA - - Author designed (Neutralization Assay),NA - - Author designed (Multiplex) | N/A | Complex test algorithm: Manual review |
| Finland | Finnish institute for health and welfare (Finnish Institute for Health and Welfare)[66] | 2020-07-19 - 2020-07-25 | National | High | 9 | Adults (18-64 years) | All | Probability | Household and community samples | 11.1% | NA - - Author designed (Neutralization Assay),NA - - Author designed (Multiplex) | N/A | Complex test algorithm: Manual review |
| Finland | Finnish institute for health and welfare (Finnish Institute for Health and Welfare)[66] | 2020-07-12 - 2020-07-18 | National | High | 7 | Adults (18-64 years) | All | Probability | Household and community samples | 0% | NA - - Author designed (Neutralization Assay),NA - - Author designed (Multiplex) | N/A | Complex test algorithm: Manual review |
| Finland | Finnish institute for health and welfare (Finnish Institute for Health and Welfare)[66] | 2020-07-05 - 2020-07-11 | National | High | 51 | Adults (18-64 years) | All | Probability | Household and community samples | 0% | NA - - Author designed (Neutralization Assay),NA - - Author designed (Multiplex) | N/A | Complex test algorithm: Manual review |
| Finland | Finnish institute for health and welfare (Finnish Institute for Health and Welfare)[66] | 2020-06-28 - 2020-07-04 | National | High | 85 | Adults (18-64 years) | All | Probability | Household and community samples | 0% | NA - - Author designed (Neutralization Assay),NA - - Author designed (Multiplex) | N/A | Complex test algorithm: Manual review |
| Finland | Finnish institute for health and welfare (Finnish Institute for Health and Welfare)[66] | 2020-06-21 - 2020-06-27 | National | High | 32 | Adults (18-64 years) | All | Probability | Household and community samples | 0% | NA - - Author designed (Neutralization Assay),NA - - Author designed (Multiplex) | N/A | Complex test algorithm: Manual review |
| Finland | Finnish institute for health and welfare (Finnish Institute for Health and Welfare)[66] | 2020-06-14 - 2020-06-20 | National | High | 78 | Adults (18-64 years) | All | Probability | Household and community samples | 0% | NA - - Author designed (Neutralization Assay),NA - - Author designed (Multiplex) | N/A | Complex test algorithm: Manual review |
| Finland | Finnish institute for health and welfare (Finnish Institute for Health and Welfare)[66] | 2020-06-07 - 2020-06-13 | National | High | 174 | Adults (18-64 years) | All | Probability | Household and community samples | 0% | NA - - Author designed (Neutralization Assay),NA - - Author designed (Multiplex) | N/A | Complex test algorithm: Manual review |
| Finland | Finnish institute for health and welfare (Finnish Institute for Health and Welfare)[66] | 2020-05-31 - 2020-06-06 | National | High | 214 | Adults (18-64 years) | All | Probability | Household and community samples | 0.5% | NA - - Author designed (Neutralization Assay),NA - - Author designed (Multiplex) | N/A | Complex test algorithm: Manual review |
| Finland | Finnish institute for health and welfare (Finnish Institute for Health and Welfare)[66] | 2020-05-24 - 2020-05-30 | National | High | 178 | Adults (18-64 years) | All | Probability | Household and community samples | 0% | NA - - Author designed (Neutralization Assay),NA - IgG, IgM - Author designed (IFA) - MULTIPLEXED | N/A | Complex test algorithm: Manual review |
| Finland | Finnish institute for health and welfare (Finnish Institute for Health and Welfare)[66] | 2020-05-17 - 2020-05-23 | National | High | 210 | Adults (18-64 years) | All | Probability | Household and community samples | 0.5% | NA - - Author designed (Neutralization Assay),NA - IgG, IgM - Author designed (IFA) - MULTIPLEXED | N/A | Complex test algorithm: Manual review |
| Finland | Finnish institute for health and welfare (Finnish Institute for Health and Welfare)[66] | 2020-05-10 - 2020-05-16 | National | High | 401 | Adults (18-64 years) | All | Probability | Household and community samples | 0.2% | NA - - Author designed (Neutralization Assay),NA - IgG, IgM - Author designed (IFA) - MULTIPLEXED | N/A | Complex test algorithm: Manual review |
| Finland | Finnish institute for health and welfare (Finnish Institute for Health and Welfare)[66] | 2020-05-03 - 2020-05-09 | National | High | 514 | Adults (18-64 years) | All | Probability | Household and community samples | 0% | NA - - Author designed (Neutralization Assay),NA - IgG, IgM - Author designed (IFA) - MULTIPLEXED | N/A | Complex test algorithm: Manual review |
| Finland | Finnish institute for health and welfare (Finnish Institute for Health and Welfare)[66] | 2020-04-26 - 2020-05-02 | National | High | 426 | Adults (18-64 years) | All | Probability | Household and community samples | 0.5% | NA - - Author designed (Neutralization Assay),NA - IgG, IgM - Author designed (IFA) - MULTIPLEXED | N/A | Complex test algorithm: Manual review |
| Finland | Finnish institute for health and welfare (Finnish Institute for Health and Welfare)[66] | 2020-04-12 - 2020-04-18 | National | High | 362 | Adults (18-64 years) | All | Probability | Household and community samples | 0.3% | NA - - Author designed (Neutralization Assay),NA - IgG, IgM - Author designed (IFA) - MULTIPLEXED | N/A | Complex test algorithm: Manual review |
| France (Paris) | Anael Ayrolles (Paris University)[384] | 2020-12-15 - 2021-04-15 | Local | High | 52 | Children and Youth (0-17 years) | All | Convenience | Residual sera | 34.6% | Abbott Laboratories - IgG - AdviseDX SARS-CoV-2 anti-IgG II assay | 0.981, 0.996 | Validated by manufacturer |
| France | Fabrice Carrat (Unité des Virus Émergents)[385] | 2020-09-01 - 2020-09-30 | National | High | 5,770 | Multiple groups | All | Convenience | Household and community samples | 3.9% | EUROIMMUN - IgG - Anti-SARS-CoV-2 ELISA IgG | 0.9, 1 | Validated by manufacturer |
| France (Nancy) | Anne Petit (Université de Lorraine)[386] | 2020-06-26 - 2020-07-24 | Local | Moderate | 2,006 | Multiple groups | All | Convenience | Household and community samples | 2.1% | Bio-rad - IgG, IgM, IgA - Platelia SARS-CoV-2 Total Ab assay,Biosynex - IgG, IgM - Biosynex COVID-19 BSS assay,EUROIMMUN - IgA - Anti-SARS-CoV-2 ELISA IgA | N/A | Complex test algorithm: Manual review |
| France (Perpignan) | Adeline Beaumont (Perpignan Hospital Center)[387] | 2020-06-29 - 2020-07-17 | Local | Moderate | 700 | Multiple groups | All | Probability | Household and community samples | 35.4% | Roche Diagnostics - IgG, IgM, IgA - Elecsys® Anti‐SARS‐CoV‐2 (S) | 0.988, 1 | Validated by manufacturer |
| France | Tom Woudenberg (Institut Pasteur)[388] | 2020-05-01 - 2020-08-15 | National | High | 1,306 | Multiple groups | All | Convenience | Residual sera | 8.5% | NA - - Author designed (Luminex) | 0.98, 0.99 | Validated by developers |
| France (Paris) | Vassilis Tsatsarisa (Université Paris-Saclay)[389] | 2020-04-29 - 2020-06-26 | Local | High | 529 | Adults (18-64 years) | Female | Sequential | Pregnant or parturient women | 4.7% | Abbott Laboratories - IgG - Abbott Architect SARS-CoV-2 IgG | 0.99, 1 | Validated by manufacturer |
| France (Paris) | Jeremie Mattern (Paris Saclay University)[390] | 2020-05-04 - 2020-05-31 | Local | High | 249 | Adults (18-64 years) | Female | Sequential | Pregnant or parturient women | 8% | Abbott Laboratories - IgG - Abbott Architect SARS-CoV-2 IgG | 0.99, 1 | Validated by manufacturer |
| France (Corsica) | Lisandru Capai (Université de Corse)[391] | 2020-04-16 - 2020-06-15 | Subnational | High | 1,973 | Multiple groups | All | Sequential | Residual sera | 5.5% | EUROIMMUN - IgG - Anti-SARS-CoV-2 ELISA IgG | 0.9, 1 | Validated by manufacturer |
| France (Paris) | Robert Cohen (Centre Hospitalier Intercommunal de Créteil)[392] | 2020-04-14 - 2020-05-12 | Local | High | 605 | Children and Youth (0-17 years) | All | Sequential | Residual sera | 10.7% | Biosynex - IgG, IgM - Biosynex COVID-19 BSS assay | 0.975, 1 | Validated by manufacturer |
| Germany (Erlangen) | Felix Wachter (Friedrich Alexander University)[393] | 2021-06-15 - 2021-07-15 | Local | High | 946 | Children and Youth (0-17 years) | All | Sequential | Residual sera | 14.6% | Roche Diagnostics - IgG, IgM, IgA - Elecsys® Anti‐SARS‐CoV‐2 (S) | 0.988, 1 | Validated by manufacturer |
| Germany | Bastian Fischer (Institut fur Laboratoriums- und Transfusionsmedizin)[394] | 2021-06-01 - 2021-06-30 | National | High | 996 | Adults (18-64 years) | All | Convenience | Blood donors | 2.3% | EUROIMMUN - IgG - Anti-SARS-CoV-2 ELISA IgG,EUROIMMUN - IgG - Anti-SARS-CoV-2 NCP ELISA (IgG) | 0.851, 1 | Complex test algorithm: Manual calculation |
| Germany (Gangelt) | Enrico Richter (German Center for Infection Research)[395] | 2021-04-01 - 2021-04-30 | Local | Moderate | 406 | Multiple groups | All | Probability | Household and community samples | 35.9% | EUROIMMUN - IgG - Anti-SARS-CoV-2 ELISA IgG | 0.9, 1 | Validated by manufacturer |
| Germany (Erlangen) | Felix Wachter (Friedrich Alexander University)[393] | 2021-04-01 - 2021-04-30 | Local | High | 808 | Children and Youth (0-17 years) | All | Sequential | Residual sera | 9.2% | Roche Diagnostics - IgG, IgM, IgA - Elecsys® Anti‐SARS‐CoV‐2 (S) | 0.988, 1 | Validated by manufacturer |
| Germany (Gangelt) | Enrico Richter (German Center for Infection Research)[395] | 2021-01-01 - 2021-01-31 | Local | Moderate | 488 | Multiple groups | All | Probability | Household and community samples | 21% | EUROIMMUN - IgG - Anti-SARS-CoV-2 ELISA IgG | 0.9, 1 | Validated by manufacturer |
| Germany (Munich) | Katja Radon (German Research Center for Environmental Health)[396] | 2020-11-02 - 2021-01-31 | Local | Low | 4,433 | Multiple groups | All | Probability | Household and community samples | 3.1% | Roche Diagnostics - IgG, IgM, IgA - Elecsys® Anti‐SARS‐CoV‐2 (N) | 1, 0.995 | Validated by manufacturer |
| Germany (Berlin-Mitte) | Robert Koch Institut (Robert Koch Institut)[397] | 2020-11-17 - 2020-12-05 | Local | Low | 2,287 | Multiple groups | All | Probability | Household and community samples | 2.4% | EUROIMMUN - IgG - Anti-SARS-CoV-2 NCP ELISA (IgG),NA - - Author designed (Neutralization Assay) | N/A | Complex test algorithm: Manual review |
| Germany (Erlangen) | Felix Wachter (Friedrich Alexander University)[393] | 2020-10-15 - 2020-11-15 | Local | High | 872 | Children and Youth (0-17 years) | All | Sequential | Residual sera | 1.4% | Roche Diagnostics - IgG, IgM, IgA - Elecsys® Anti‐SARS‐CoV‐2 (S) | 0.988, 1 | Validated by manufacturer |
| Germany (Gangelt) | Enrico Richter (German Center for Infection Research)[395] | 2020-10-01 - 2020-10-31 | Local | Moderate | 587 | Multiple groups | All | Probability | Household and community samples | 18.1% | EUROIMMUN - IgG - Anti-SARS-CoV-2 ELISA IgG | 0.9, 1 | Validated by manufacturer |
| Germany (Ruhr) | Folke Brinkmann (Ruhr University Bochum)[398] | 2020-06-15 - 2021-02-15 | Subnational | High | 2,145 | Children and Youth (0-17 years) | All | Convenience | Residual sera | 2.6% | Roche Diagnostics - IgG, IgM, IgA - Elecsys® Anti‐SARS‐CoV‐2 (N) | 1, 0.995 | Validated by manufacturer |
| Germany (Straubing) | Robert Koch Institute (Robert Koch Institute)[399] | 2020-09-08 - 2020-09-26 | Local | Low | 2,361 | Multiple groups | All | Probability | Household and community samples | 1.8% | EUROIMMUN - IgG - Anti-SARS-CoV-2 ELISA IgG,NA - - Author designed (Neutralization Assay) | N/A | Complex test algorithm: Manual review |
| Germany (Würzburg) | Felizitas Eichner (Universität Würzburg)[400] | 2020-06-01 - 2020-10-31 | Local | Moderate | 3,034 | Multiple groups | All | Stratified non-probability | Household and community samples | 1.1% | Roche Diagnostics - IgG, IgM, IgA - Elecsys® Anti‐SARS‐CoV‐2 (N) | 1, 0.995 | Validated by manufacturer |
| Germany (Bavaria) | Markus Hippich ( German Research Center for Environmental Health)[401] | 2020-07-01 - 2020-07-31 | Subnational | High | 2,746 | Children and Youth (0-17 years) | All | Convenience | Residual sera | 0.9% | NA - - Author designed (CLIA) | 0.973, 1 | Validated by independent authors/third party/non-developers |
| Germany (Bavaria) | Ralf Wagner (Friedrich-Alexander University)[402] | 2020-06-28 - 2020-07-10 | Local | Moderate | 4,201 | Multiple groups | All | Probability | Household and community samples | 8.6% | Shenzhen Yhlo Biotech Co. Ltd - IgG, IgM - iFlash-SARS-CoV-2 IgM/IgG,Roche Diagnostics - IgG, IgM, IgA - Elecsys® Anti‐SARS‐CoV‐2 (N),NA - IgG - Author designed (ELISA) -Spike | N/A | Complex test algorithm: Manual review |
| Germany (Hamburg) | Gabor Dunay (University Medical Center Hamburg-Eppendorf)[403] | 2020-05-11 - 2020-06-30 | Local | High | 4,657 | Children and Youth (0-17 years) | All | Convenience | Household and community samples | 1.4% | Roche Diagnostics - IgG, IgM, IgA - Elecsys® Anti‐SARS‐CoV‐2 (N),DiaSorin - IgG - Liaison SARS-CoV-2 S1/S2 IgG | 0.974, 1 | Complex test algorithm: Manual calculation |
| Germany (Kupferzell) | Claudia Santos Hovener (Robert Koch Institute)[404] | 2020-05-20 - 2020-06-09 | Local | Moderate | 2,203 | Multiple groups | All | Probability | Household and community samples | 11.3% | EUROIMMUN - IgG - Anti-SARS-CoV-2 ELISA IgG | 0.9, 1 | Validated by manufacturer |
| Germany (Bonn) | N Ahmad Aziz (University of Bonn)[405] | 2020-04-24 - 2020-06-30 | Local | High | 4,755 | Multiple groups | All | Convenience | Household and community samples | 1% | EUROIMMUN - IgG - Anti-SARS-CoV-2 ELISA IgG | 0.9, 1 | Validated by manufacturer |
| Germany (Munich) | Michael Pritsch (University Hospital LMU Munich)[406] | 2020-04-05 - 2020-06-12 | Local | Low | 5,313 | Multiple groups | All | Probability | Household and community samples | 1.9% | Roche Diagnostics - IgG, IgM, IgA - Elecsys® Anti‐SARS‐CoV‐2 (N) | 1, 0.995 | Validated by manufacturer |
| Germany (Southwest) | Stefan Runkel (University Medical Center of The Johannes Gutenberg-University Mainz)[407] | 2020-03-15 - 2020-06-15 | Subnational | High | 3,880 | Multiple groups | All | Convenience | Blood donors | 0.9% | Abbott Laboratories - IgG - Abbott Architect SARS-CoV-2 IgG | 0.99, 1 | Validated by manufacturer |
| Germany (Jena) | Janine Zollkau (Universitätsklinikum Jena)[408] | 2020-04-06 - 2020-05-13 | Local | High | 180 | Multiple groups | Female | Sequential | Pregnant or parturient women | 0.6% | Epitope Diagnostics, Inc. - IgG - EDI™ Novel Coronavirus COVID-19 IgG ELISA Kit | 0.984, 0.998 | Validated by manufacturer |
| Germany (Regensberg) | Sebastian Hausler (Hospital St Hedwig of the Order of St John)[409] | 2020-04-09 - 2020-04-30 | Local | High | 151 | Adults (18-64 years) | Female | Sequential | Pregnant or parturient women | 2% | EUROIMMUN - IgG - Anti-SARS-CoV-2 ELISA IgG,EUROIMMUN - IgA - Anti-SARS-CoV-2 ELISA IgA | 0.997, 0.983 | Complex test algorithm: Manual calculation |
| Germany (Gangelt) | Hendrik Streeck (University of Bonn)[410] | 2020-03-31 - 2020-04-06 | Local | Moderate | 919 | Multiple groups | All | Probability | Household and community samples | 13.6% | EUROIMMUN - IgG - Anti-SARS-CoV-2 ELISA IgG | 0.9, 1 | Validated by manufacturer |
| Greece (Athens) | Helena Maltezou (University of Athens)[411] | 2021-06-01 - 2021-06-22 | Local | High | 901 | Multiple groups | All | Convenience | Residual sera | 55.7% | Abbott Laboratories - IgG - SARS-CoV-2 IgG II Quant | 0.994, 0.996 | Validated by manufacturer |
| Greece (Deskati) | Ourania Kotsiou (University of Thessaly)[412] | 2021-06-06 - 2021-06-06 | Local | High | 69 | Multiple groups | All | Convenience | Household and community samples | 79.7% | Abbott Laboratories - IgG - SARS-CoV-2 IgG II Quant | 0.994, 0.996 | Validated by manufacturer |
| Greece (Athens) | Helena Maltezou (University of Athens)[411] | 2021-03-01 - 2021-03-22 | Local | High | 764 | Multiple groups | All | Convenience | Residual sera | 11.6% | Abbott Laboratories - IgG - SARS-CoV-2 IgG II Quant | 0.994, 0.996 | Validated by manufacturer |
| Greece (Deskati) | Ourania S. Kotsiou (University of Thessaly)[413] | 2021-01-29 - 2021-01-29 | Local | High | 388 | Multiple groups | All | Convenience | Household and community samples | 17.8% | Prognosis Biotech - IgG, IgM, IgA - Rapid Test 2019-nCoV Total Ig,Abbott Laboratories - IgG - Abbott Architect SARS-CoV-2 IgG | 0.999, 1 | Complex test algorithm: Manual calculation |
| Greece | Zacharoula Bogogiannidou (University of Thessaly)[414] | 2020-04-01 - 2020-04-30 | National | High | 4,511 | Multiple groups | All | Convenience | Residual sera | 0.4% | Abbott Laboratories - IgG - Abbott Architect SARS-CoV-2 IgG | 0.99, 1 | Validated by manufacturer |
| Iceland | Daniel Gudbjartsson (deCODE Genetics)[415] | 2020-04-27 - 2020-06-12 | National | High | 23,452 | Multiple groups | All | Convenience | Household and community samples | 0.3% | Roche Diagnostics - IgG, IgM, IgA - Elecsys® Anti‐SARS‐CoV‐2 (S),Beijing Wantai Biological - IgG, IgM, IgA - Wantai SARS-CoV-2 Total Ab ELISA | 0.955, 1 | Complex test algorithm: Manual calculation |
| Ireland | Laura Heavey (Health Protection Surveillance Centre)[416] | 2020-06-22 - 2020-07-16 | Local | Low | 913 | Multiple groups | All | Probability | Household and community samples | 3.1% | Abbott Laboratories - IgG - Abbott Architect SARS-CoV-2 IgG,Beijing Wantai Biological - IgG, IgM, IgA - Wantai SARS-CoV-2 Total Ab ELISA | 0.957, 1 | Complex test algorithm: Manual calculation |
| Ireland | Laura Heavey (Health Protection Surveillance Centre)[416] | 2020-06-22 - 2020-07-16 | Local | Low | 820 | Multiple groups | All | Probability | Household and community samples | 0.6% | Abbott Laboratories - IgG - Abbott Architect SARS-CoV-2 IgG,Beijing Wantai Biological - IgG, IgM, IgA - Wantai SARS-CoV-2 Total Ab ELISA | 0.957, 1 | Complex test algorithm: Manual calculation |
| Israel (Jerusalem) | Adin Breuer (Shaare Zedek Medical Center)[417] | 2020-10-18 - 2021-01-12 | Local | Moderate | 1,138 | Children and Youth (0-17 years) | All | Convenience | Residual sera | 10% | Abbott Laboratories - IgG - Abbott Architect SARS-CoV-2 IgG,DiaSorin - IgG - Liaison SARS-CoV-2 S1/S2 IgG | 0.964, 1 | Complex test algorithm: Manual calculation |
| Israel (Modiin) | Yael Gozlan (Israel Ministry of Health)[418] | 2020-09-01 - 2020-09-30 | Local | High | 283 | Multiple groups | All | Convenience | Household and community samples | 16.6% | Abbott Laboratories - IgG - Abbott Architect SARS-CoV-2 IgG,DiaSorin - IgG - Liaison SARS-CoV-2 S1/S2 IgG,NA - IgG - Author designed (ELISA) -Spike | N/A | Complex test algorithm: Manual review |
| Israel | Anat Ekka Zohar (Maccabi Healthcare Services)[419] | 2020-06-28 - 2020-07-27 | National | High | 15,459 | Multiple groups | All | Convenience | Residual sera | 1% | Abbott Laboratories - IgG - Abbott Architect SARS-CoV-2 IgG,DiaSorin - IgG - LIAISON® SARS-CoV-2 TrimericS IgG | 0.977, 1 | Complex test algorithm: Manual calculation |
| Italy (Foggia) | Francescopaolo Antonucci (University Hospital “Ospedali Riuniti” Foggia)[420] | 2021-03-01 - 2021-03-15 | Local | Moderate | 484 | Multiple groups | All | Convenience | Blood donors | 18.8% | Ortho Clinical Diagnostics Inc. - IgG - VITROS Immunodiagnostic Products Anti-SARS-CoV-2 IgG,Ortho Clinical Diagnostics Inc. - IgG, IgM, IgA - VITROS Immunodiagnostic Products Anti-SARS-CoV-2 Total | 1, 1 | Complex test algorithm: Manual calculation |
| Italy (Campania) | Immacolata Polvere (Università degli Studi del Sannio)[421] | 2020-09-01 - 2020-09-30 | Local | Moderate | 1,383 | Seniors (65+ years) | All | Convenience | Household and community samples | 4.7% | Beijing Lepu Medical Technology - IgG, IgM - Leccurate SARS-CoV-2 Antibody Rapid Test Kit (Colloidal Gold Immunochromatography) | 0.989, 0.976 | Validated by manufacturer |
| Italy (Gardena Valley ( Ortisei, Santa Cristina, and Selva, the main municipalities)) | Roberto Melotti (Hospital of Bolzano)[422] | 2020-05-26 - 2020-06-08 | Local | Low | 2,106 | Multiple groups | All | Probability | Household and community samples | 26.2% | Abbott Laboratories - IgG - Abbott Architect SARS-CoV-2 IgG | 0.99, 1 | Validated by manufacturer |
| Italy (Castiglione D’Adda) | Gabriele Pagani (University of Milan)[423] | 2020-05-18 - 2020-06-07 | Local | Moderate | 4,143 | Multiple groups | All | Probability | Household and community samples | 22.2% | PRIMA Lab S.A. - IgG, IgM - PRIMA COVID-19 IgG/IgM Rapid Test | 0.92, 0.98 | Validated by manufacturer |
| Italy (Ariano Irpino) | Pellegrino Cerino (The Zoo-Prophylactic Institute of Southern Italy)[424] | 2020-05-16 - 2020-05-19 | Local | Moderate | 13,218 | Multiple groups | All | Convenience | Household and community samples | 5.6% | Roche Diagnostics - IgG, IgM, IgA - Elecsys® Anti‐SARS‐CoV‐2 (N) | 1, 0.995 | Validated by manufacturer |
| Italy (Foggia) | Jose Ramon Fiore (University of Foggia)[425] | 2020-05-01 - 2020-05-31 | Local | Moderate | 904 | Adults (18-64 years) | All | Convenience | Blood donors | 1% | Snibe Co. Ltd (Shenzhen New Industries Biomedical Engineering Co. Ltd) - IgG, IgM - MAGLUMI 2019-nCoV IgM/IgG | 0.94, 0.987 | Validated by manufacturer |
| Italy (Vo) | Ilaria Dorigatti (University of Padova)[426] | 2020-05-01 - 2020-05-31 | Local | Moderate | 2,602 | Multiple groups | All | Probability | Household and community samples | 6.2% | Abbott Laboratories - IgG - Abbott Architect SARS-CoV-2 IgG,Roche Diagnostics - IgG, IgM, IgA - Elecsys® Anti‐SARS‐CoV‐2 (N),DiaSorin - IgG - Liaison SARS-CoV-2 S1/S2 IgG | N/A | Complex test algorithm: Manual review |
| Italy (Lodi) | Irene Cassaniti (IRCCS Policlinico)[427] | 2020-03-18 - 2020-06-24 | Local | High | 1,922 | Multiple groups | All | Convenience | Blood donors | 19.7% | DiaSorin - IgG - Liaison SARS-CoV-2 S1/S2 IgG | 0.974, 0.985 | Validated by manufacturer |
| Italy (Verona) | Massimo Guerriero (Instituto di Ricovero e Cura A Caratterre)[428] | 2020-04-25 - 2020-05-08 | Local | Moderate | 1,515 | Multiple groups | All | Probability | Household and community samples | 2.7% | Abbott Laboratories - IgG - Abbott Architect SARS-CoV-2 IgG | 0.99, 1 | Validated by manufacturer |
| Italy (Lodi) | Alessandro Baracco (A.S.S.T.Lodi)[429] | 2020-04-23 - 2020-05-05 | Local | Moderate | 1,792 | Multiple groups | All | Probability | Household and community samples | 29.8% | DiaSorin - IgG - Liaison SARS-CoV-2 S1/S2 IgG | 0.974, 0.985 | Validated by manufacturer |
| Italy (Prato) | Anna Cavaliere (Santo Stefano Hospital)[430] | 2020-04-04 - 2020-05-16 | Local | High | 134 | Adults (18-64 years) | Female | Sequential | Pregnant or parturient women | 4.5% | ACRO Biotech Inc - IgG, IgM - Acro Biotech COVID-19 IgM/IgG Rapid POC test | 0.977, 0.99 | Validated by manufacturer |
| Italy (Villa Caldari) | Francesa Cito (Istituto Zooprofilattico Sperimentale dell’Abruzzo e del Molise G. Caporale )[431] | 2020-04-18 - 2020-04-19 | Local | Moderate | 667 | Multiple groups | All | Probability | Household and community samples | 10.9% | Beijing Wantai Biological - IgG, IgM, IgA - Wantai SARS-CoV-2 Total Ab ELISA | 0.967, 0.975 | Validated by manufacturer |
| Italy (Milan) | Luca Valenti ( Fondazione IRCCS Ca’ Granda Ospedale Maggiore )[432] | 2020-03-30 - 2020-04-08 | Local | High | 60 | Adults (18-64 years) | All | Convenience | Blood donors | 6.7% | PRIMA Lab S.A. - IgG, IgM - PRIMA COVID-19 IgG/IgM Rapid Test | 0.92, 0.98 | Validated by manufacturer |
| Italy (Arezzo) | Alessandro Pancrazzi (Ospedale San Donato)[433] | 2020-03-17 - 2020-03-21 | Local | High | 516 | Adults (18-64 years) | All | Sequential | Residual sera | 13% | ACRO Biotech Inc - IgG, IgM - Acro Biotech COVID-19 IgM/IgG Rapid POC test | 0.977, 0.99 | Validated by manufacturer |
| Netherlands (Amsterdam) | Liza Coyer (University of Amsterdam)[434] | 2020-11-23 - 2021-06-04 | Local | High | 2,075 | Multiple groups | All | Convenience | Household and community samples | 23.6% | Beijing Wantai Biological - IgG, IgM, IgA - Wantai SARS-CoV-2 Total Ab ELISA | 0.967, 0.975 | Validated by manufacturer |
| Netherlands (Limburg) | Demi Pagen (South Limburg Public Health Service)[435] | 2020-10-28 - 2020-12-23 | Subnational | High | 10,001 | Multiple groups | All | Convenience | Household and community samples | 19.5% | Beijing Wantai Biological - IgG, IgM, IgA - Wantai SARS-CoV-2 Total Ab ELISA | 0.967, 0.975 | Validated by manufacturer |
| Netherlands (Rotterdam) | I.L.M. Rotee ( Franciscus Gasthuis & Vlietland)[436] | 2020-10-19 - 2020-12-19 | Local | High | 240 | Children and Youth (0-17 years) | All | Sequential | Residual sera | 13.8% | Beijing Wantai Biological - IgG, IgM, IgA - Wantai SARS-CoV-2 Total Ab ELISA | 0.967, 0.975 | Validated by manufacturer |
| Netherlands (Rotterdam) | I.L.M. Rotee (Franciscus Gasthuis & Vlietland)[436] | 2020-07-19 - 2020-09-19 | Local | High | 209 | Children and Youth (0-17 years) | All | Sequential | Residual sera | 4.1% | Beijing Wantai Biological - IgG, IgM, IgA - Wantai SARS-CoV-2 Total Ab ELISA | 0.967, 0.975 | Validated by manufacturer |
| Netherlands (Amsterdam) | Liza Coyer (University of Amsterdam)[437] | 2020-06-24 - 2020-10-09 | Local | Moderate | 2,483 | Multiple groups | All | Probability | Household and community samples | 9.1% | Beijing Wantai Biological - IgG, IgM, IgA - Wantai SARS-CoV-2 Total Ab ELISA | 0.967, 0.975 | Validated by manufacturer |
| Netherlands (Amsterdam) | Maya Keuning (University of Amsterdam)[438] | 2020-04-12 - 2020-10-02 | Local | High | 487 | Children and Youth (0-17 years) | All | Convenience | Residual sera | 3.3% | Beijing Wantai Biological - IgG, IgM, IgA - Wantai SARS-CoV-2 Total Ab ELISA | 0.967, 0.975 | Validated by manufacturer |
| Netherlands | Ed Slot (Amsterdam UMC)[439] | 2020-04-01 - 2020-04-15 | National | High | 7,361 | Multiple groups | All | Sequential | Blood donors | 3.4% | Beijing Wantai Biological - IgG, IgM, IgA - Wantai Rapid Test for Total Antibody to SARS-CoV-2 | 0.947, 0.989 | Validated by manufacturer |
| Norway (Lorenskog) | Anne Eskild (University of Oslo)[440] | 2020-12-01 - 2020-12-31 | Local | Moderate | 418 | Adults (18-64 years) | Female | Sequential | Pregnant or parturient women | 5% | Roche Diagnostics - IgG, IgM, IgA - Elecsys® Anti‐SARS‐CoV‐2 (N),DiaSorin - IgG - Liaison SARS-CoV-2 S1/S2 IgG | 1, 0.98 | Complex test algorithm: Manual calculation |
| Poland (Katowice, Gliwice and Sosnowiec) | Jan Zejda (Slaski Uniwersytet Medyczny)[441] | 2020-10-15 - 2020-11-15 | Local | Moderate | 1,167 | Multiple groups | All | Probability | Household and community samples | 11.4% | EUROIMMUN - IgG - Anti-SARS-CoV-2 ELISA IgG | 0.9, 1 | Validated by manufacturer |
| Romania | Tudor Olariu (Victor Babes University of Medicine and Pharmacy)[442] | 2021-07-02 - 2021-09-02 | Local | High | 2,395 | Adults (18-64 years) | All | Sequential | Blood donors | 41% | Roche Diagnostics - IgG, IgM, IgA - Elecsys® Anti‐SARS‐CoV‐2 (N) | 1, 0.995 | Validated by manufacturer |
| Romania (Timisoara) | Tudor Olariu (Victor Babes University of Medicine and Pharmacy Timisoara)[443] | 2021-03-10 - 2021-06-10 | Local | High | 379 | Children and Youth (0-17 years) | All | Sequential | Residual sera | 46.7% | Roche Diagnostics - IgG, IgM, IgA - Elecsys® Anti‐SARS‐CoV‐2 (N) | 1, 0.995 | Validated by manufacturer |
| Romania | Tudor Rares Olariu (Victor Babes University of Medicine and Pharmacy)[444] | 2020-07-08 - 2020-09-01 | Local | Moderate | 2,115 | Adults (18-64 years) | All | Sequential | Blood donors | 1.5% | Roche Diagnostics - IgG, IgM, IgA - Elecsys® Anti‐SARS‐CoV‐2 (N) | 1, 0.995 | Validated by manufacturer |
| Spain (Catalonia) | Marianna Karachaliou (Instituto de Salud Global Barcelona)[445] | 2021-05-26 - 2021-07-21 | Subnational | High | 1,076 | Multiple groups | All | Convenience | Blood donors | 92.6% | NA - - Author designed (Multiplex) | 0.9578, 1 | Validated by developers |
| Spain (Madrid) | Vicente Soriano (UNIR Health Sciences School and Medical Centre)[446] | 2020-09-15 - 2020-12-24 | Local | High | 601 | Multiple groups | All | Sequential | Residual sera | 7.7% | PCL Inc - IgG, IgM - PCL COVID19 IgG/IgM Rapid Gold | 0.967, 1 | Validated by manufacturer |
| Spain (Catalonia) | Marianna Karachaliou (Barcelona Institute For Global Health)[185] | 2020-06-30 - 2020-11-15 | Subnational | High | 260 | Children and Youth (0-17 years) | All | Convenience | Household and community samples | 11.5% | NA - - Author designed (Multiplex) | N/A | Complex test algorithm: Manual review |
| Spain (Catalonia) | Manolis Kogevinas (Barcelona Institute for Global Health)[447] | 2020-05-17 - 2020-11-15 | Subnational | High | 4,103 | Adults (18-64 years) | All | Convenience | Household and community samples | 18.1% | NA - - Author designed (Luminex) | 0.9578, 1 | Validated by developers |
| Spain (Madrid) | Maria Vinuela (University Hospital Gregorio Marañón)[448] | 2020-05-06 - 2020-05-21 | Local | High | 100 | Adults (18-64 years) | Female | Sequential | Pregnant or parturient women | 13% | Abbott Laboratories - IgG - Abbott Architect SARS-CoV-2 IgG | 0.99, 1 | Validated by manufacturer |
| Spain (Madrid) | Vicente Soriano (UNIR Health Sciences School and Medical Centre)[446] | 2020-03-15 - 2020-06-30 | Local | High | 968 | Multiple groups | All | Sequential | Residual sera | 12.6% | PCL Inc - IgG, IgM - PCL COVID19 IgG/IgM Rapid Gold | 0.967, 1 | Validated by manufacturer |
| Spain (Barcelona) | Francesca Crovetto (Universitat de Barcelona)[449] | 2020-03-15 - 2020-05-31 | Local | High | 2,225 | Adults (18-64 years) | Female | Sequential | Pregnant or parturient women | 14.1% | Vircell S.L. - IgG - COVID-19 ELISA IgG,Ortho Clinical Diagnostics Inc. - IgG, IgM, IgA - VITROS Immunodiagnostic Products Anti-SARS-CoV-2 Total,NA - - Author designed (Luminex) | N/A | Complex test algorithm: Manual review |
| Spain (Madrid) | R. Alenda (Centro de Transfusión de la Comunidad de Madrid)[450] | 2020-03-15 - 2020-03-22 | Local | High | 89 | Multiple groups | All | Sequential | Blood donors | 0% | Quotient - IgG, IgM - MosaiQ COVID-19 Antibody Microarray | 1, 0.998 | Validated by manufacturer |
| Sweden (Stockholm) | Ake Lundkvist (Uppsala University )[451] | 2020-06-17 - 2020-06-18 | Local | High | 123 | Adults (18-64 years) | All | Probability | Household and community samples | 4.1% | Zhejiang Orient Gene Biotech - IgG, IgM - COVID-19 IgG/IgM Rapid Test | 0.972, 1 | Validated by manufacturer |
| Sweden (Stockholm) | Ake Lundkvist (Uppsala University )[451] | 2020-06-17 - 2020-06-18 | Local | High | 90 | Adults (18-64 years) | All | Probability | Household and community samples | 30% | Zhejiang Orient Gene Biotech - IgG, IgM - COVID-19 IgG/IgM Rapid Test | 0.972, 1 | Validated by manufacturer |
| Switzerland (Fribourg) | Emna El May (Swiss School of Public Health)[452] | 2021-05-20 - 2021-08-13 | Subnational | High | 504 | Multiple groups | All | Probability | Household and community samples | 78% | Swiss Federal Institute of Technology - IgG, IgA - Sensitive Anti-SARS-CoV-2 Spike Trimer Immunoglobulin Serological (SenASTrIS) | 0.966, 0.997 | Validated by independent authors/third party/non-developers |
| Switzerland (Geneva) | Silvia Stringhini (Geneva University Hospital )[453] | 2021-06-01 - 2021-07-07 | Local | Moderate | 3,355 | Multiple groups | All | Probability | Household and community samples | 27% | Roche Diagnostics - IgG, IgM, IgA - Elecsys® Anti‐SARS‐CoV‐2 (N) | N/A | Complex test algorithm: Manual review |
| Switzerland (Geneva) | Silvia Stringhini (Geneva University Hospitals)[453] | 2020-11-23 - 2020-12-23 | Local | Low | 4,000 | Multiple groups | All | Probability | Household and community samples | 20.5% | Roche Diagnostics - IgG, IgM, IgA - Elecsys® Anti‐SARS‐CoV‐2 (S) | 0.988, 1 | Validated by manufacturer |
| Switzerland (Southern Switzerland) | Elena Giuliano (Università della Svizzera Italiana)[454] | 2020-07-15 - 2020-12-15 | Subnational | High | 301 | Adults (18-64 years) | All | Sequential | Household and community samples | 3.3% | EUROIMMUN - IgG - Anti-SARS-CoV-2 ELISA IgG | 0.9, 1 | Validated by manufacturer |
| Switzerland (Zurich) | Marc Emmenegger (University of Zurich)[455] | 2020-07-01 - 2020-07-15 | Local | High | 1,067 | Adults (18-64 years) | All | Sequential | Blood donors | 0.7% | NA - - Author designed (ELISA) - MULTIPLEXED | 1, 1 | Validated by developers |
| Switzerland (Zurich) | Agne Ulyte (University of Zurich)[456] | 2020-06-16 - 2020-07-09 | Local | Low | 857 | Adults (18-64 years) | All | Probability | Household and community samples | 3.6% | NA - - Author designed (Luminex) | 0.983, 0.984 | Validated by developers |
| Switzerland (Geneva) | Stephanie Baggio (Geneva University Hospitals)[457] | 2020-06-01 - 2020-06-30 | Local | Moderate | 1,167 | Multiple groups | All | Convenience | Residual sera | 5.9% | EUROIMMUN - IgG - Anti-SARS-CoV-2 ELISA IgG | 0.9, 1 | Validated by manufacturer |
| Switzerland (Zurich) | Marc Emmenegger (University of Zurich)[455] | 2020-05-15 - 2020-07-15 | Local | High | 12,831 | Adults (18-64 years) | All | Sequential | Residual sera | 0.9% | NA - - Author designed (ELISA) - MULTIPLEXED | 1, 1 | Validated by developers |
| Switzerland (Grisons) | Gregorio Milani (Università della Svizzera Italiana)[458] | 2020-05-15 - 2020-05-31 | Local | High | 423 | Multiple groups | All | Convenience | Household and community samples | 2.6% | EUROIMMUN - IgG - Anti-SARS-CoV-2 ELISA IgG | 0.9, 1 | Validated by manufacturer |
| Switzerland (Zurich) | Marc Emmenegger (University of Zurich)[455] | 2020-04-01 - 2020-04-30 | Local | High | 1,469 | Adults (18-64 years) | All | Sequential | Blood donors | 1.2% | NA - - Author designed (ELISA) - MULTIPLEXED | 1, 1 | Validated by developers |
| Switzerland (Zurich) | Marc Emmenegger (University of Zurich)[455] | 2020-04-01 - 2020-04-30 | Local | High | 4,275 | Adults (18-64 years) | All | Sequential | Residual sera | 1.4% | NA - - Author designed (ELISA) - MULTIPLEXED | 1, 1 | Validated by developers |
| Switzerland (Zurich) | Marc Emmenegger (University of Zurich)[455] | 2020-03-15 - 2020-03-31 | Local | High | 3,806 | Adults (18-64 years) | All | Sequential | Residual sera | 0.3% | NA - - Author designed (ELISA) - MULTIPLEXED | 1, 1 | Validated by developers |
| The United Kingdom | Public Health Scotland (Public Health Scotland)[94] | 2021-12-20 - 2022-01-23 | National | High | 2,914 | Adults (18-64 years) | Female | Sequential | Pregnant or parturient women | 91% | Roche Diagnostics - IgG, IgM, IgA - Elecsys® Anti‐SARS‐CoV‐2 (S) | 0.988, 1 | Validated by manufacturer |
| The United Kingdom | Public Health Scotland (Public Health Scotland)[95] | 2021-11-15 - 2021-12-19 | National | High | 3,060 | Adults (18-64 years) | Female | Sequential | Pregnant or parturient women | 87.2% | Roche Diagnostics - IgG, IgM, IgA - Elecsys® Anti‐SARS‐CoV‐2 (S) | 0.988, 1 | Validated by manufacturer |
| The United Kingdom | Public Health Scotland (Public Health Scotland)[96] | 2021-10-11 - 2021-11-14 | National | High | 2,890 | Adults (18-64 years) | Female | Sequential | Pregnant or parturient women | 86.2% | Roche Diagnostics - IgG, IgM, IgA - Elecsys® Anti‐SARS‐CoV‐2 (S) | 0.988, 1 | Validated by manufacturer |
| The United Kingdom | Daniel Major-Smith (University of Bristol)[459] | 2021-05-01 - 2021-06-28 | Local | High | 66 | Adults (18-64 years) | All | Convenience | Household and community samples | 36.4% | Roche Diagnostics - IgG, IgM, IgA - Elecsys® Anti‐SARS‐CoV‐2 (S) | 0.988, 1 | Validated by manufacturer |
| The United Kingdom | Daniel Major-Smith (University of Bristol)[459] | 2021-04-08 - 2021-06-28 | Local | High | 2,555 | Adults (18-64 years) | All | Convenience | Household and community samples | 96.4% | Roche Diagnostics - IgG, IgM, IgA - Elecsys® Anti‐SARS‐CoV‐2 (S) | 0.988, 1 | Validated by manufacturer |
| The United Kingdom | Sarah Beale (University College London)[460] | 2021-02-01 - 2021-06-08 | National | High | 3,775 | Multiple groups | All | Convenience | Household and community samples | 12.1% | Roche Diagnostics - IgG, IgM, IgA - Elecsys® Anti‐SARS‐CoV‐2 (N) | 1, 0.995 | Validated by manufacturer |
| The United Kingdom (London) | Katherine Gaskell (London School of Hygiene & Tropical Medicine)[461] | 2020-10-19 - 2020-12-07 | Local | Moderate | 1,242 | Multiple groups | All | Probability | Household and community samples | 64.3% | NA - - Not reported/ Unable to specify | 0.962, 0.974 | Validated by independent authors/third party/non-developers |
| The United Kingdom | Clarissa Oeser (UK Health Security Agency (UKHSA, formerly PHE))[97] | 2020-09-01 - 2020-10-31 | National | High | 119 | Children and Youth (0-17 years) | All | Convenience | Residual sera | 6.7% | Roche Diagnostics - IgG, IgM, IgA - Elecsys® Anti‐SARS‐CoV‐2 (S) | 0.988, 1 | Validated by manufacturer |
| The United Kingdom (Blackpool) | Jane Oakey (NHS Foundation Trust )[462] | 2020-08-01 - 2020-08-31 | Local | High | 225 | Multiple groups | All | Convenience | Residual sera | 5.3% | Abbott Laboratories - IgG - Abbott Architect SARS-CoV-2 IgG | 0.99, 1 | Validated by manufacturer |
| The United Kingdom (Scotland) | Hani Abo-Leyah (University of Dundee)[463] | 2020-05-28 - 2020-09-02 | Subnational | High | 231 | Multiple groups | All | Sequential | Residual sera | 4.8% | Siemens - IgG, IgM, IgA - Atellica® IM SARS-CoV-2 Total (COV2T) | 0.985, 0.998 | Validated by manufacturer |
| The United Kingdom (South East England) | Philippa M Wells (King’s College London)[464] | 2020-04-27 - 2020-06-02 | Subnational | High | 431 | Multiple groups | All | Convenience | Household and community samples | 12% | NA - - Author designed (ELISA) - MULTIPLEXED | 0.9, 1 | Validated by independent authors/third party/non-developers |
| The United Kingdom (Oxford) | Sheila F Lumley (University of Oxford)[465] | 2020-04-14 - 2020-06-15 | Local | Moderate | 1,000 | Adults (18-64 years) | Female | Sequential | Pregnant or parturient women | 5.3% | NA - IgG - Author designed (ELISA) -Spike | 0.991, 0.99 | Validated by independent authors/third party/non-developers |
| The United Kingdom (Dublin) | Kate Glennon (University College Dublin)[466] | 2020-05-04 - 2020-05-15 | Local | High | 598 | Adults (18-64 years) | Female | Convenience | Pregnant or parturient women | 1.7% | Abbott Laboratories - IgG - Abbott Architect SARS-CoV-2 IgG,Roche Diagnostics - IgG, IgM, IgA - Elecsys® Anti‐SARS‐CoV‐2 (N) | 0.99, 1 | Complex test algorithm: Manual calculation |
| The United Kingdom (London) | Gayatri Amirthalingam (Public Health England)[467] | 2020-05-01 - 2020-05-03 | Local | Moderate | 974 | Multiple groups | All | Convenience | Blood donors | 13% | EUROIMMUN - IgG - Anti-SARS-CoV-2 ELISA IgG | 0.9, 1 | Validated by manufacturer |
| The United Kingdom (Leicester) | Paul William Bird (University of Leicester)[468] | 2020-03-16 - 2020-06-03 | Local | Moderate | 1,779 | Multiple groups | All | Probability | Household and community samples | 11.3% | DiaSorin - IgG - Liaison SARS-CoV-2 S1/S2 IgG | 0.974, 0.985 | Validated by manufacturer |
| The United Kingdom (Greater Glasgow area) | Ellen C Hughes (University of Glasgow)[469] | 2020-03-16 - 2020-05-24 | Local | Low | 6,635 | Multiple groups | All | Probability | Residual sera | 7.8% | NA - IgG - Author designed (ELISA) -Spike | 0.9531, 0.972 | Validated by developers |
| The United Kingdom | Public Health England (Public Health England)[111] | 2020-02-01 - 2020-03-31 | National | High | 443 | Children and Youth (0-17 years) | All | Probability | Residual sera | 4.9% | Abbott Laboratories - IgG - Abbott Architect SARS-CoV-2 IgG | 0.99, 1 | Validated by manufacturer |

###### South-East Asia region

| Country (Location) | Author (Organization) | Sampling Dates (YMD) | Geographic scope | Overall risk of bias | Sample size | Age | Sex | Sampling method | Sample frame | Seroprevalence (95% CI) | Test Manufacturers - Isotypes - Names | Test Sens, Spec | Sens, Spec Source |
| --- | --- | --- | --- | --- | --- | --- | --- | --- | --- | --- | --- | --- | --- |
| Bangladesh - HRP (Sitakunda subdistrict) | Taufiqur Bhuiyan (icddr,b [International Centre for Diarrhoeal Disease Research, Bangladesh])[470] | 2021-03-27 - 2021-06-13 | Local | Low | 2,307 | Multiple groups | All | Probability | Household and community samples | 62.5% | Beijing Wantai Biological - IgG, IgM, IgA - Wantai SARS-CoV-2 Total Ab ELISA | 0.967, 0.975 | Validated by manufacturer |
| Bangladesh - HRP (Dhaka) | Zabed Ahmed (Bangladesh Reference Institute for Chemical Measurements )[471] | 2020-10-15 - 2021-02-15 | Local | High | 239 | Adults (18-64 years) | All | Convenience | Household and community samples | 42.2% | Erba Manneim - IgG - ErbaLisa® COVID-19 IgG semi quantitative kit | 0.991, 0.993 | Validated by manufacturer |
| Bangladesh - HRP (FDMN populations in Cox’s Bazar Rohiyanga camps) | Prof. Dr. Tahmina Shirin (IEDCR)[472] | 2020-12-01 - 2020-12-30 | Local | Moderate | 3,446 | Multiple groups | All | Probability | Household and community samples | 58% | Beijing Wantai Biological - IgG IgM IgA - Wantai SARS-CoV-2 Total Ab ELISA | 0.967, 0.975 | Validated by manufacturer |
| India (Vellore) | Divya Dayanand (Christian Medical College)[473] | 2021-07-01 - 2021-07-31 | Local | Low | 1,205 | Multiple groups | All | Probability | Household and community samples | 71.6% | InBios International, Inc. - IgG - anti-SARS-CoV-2 IgM and IgG | 0.919, 0.99 | Validated by manufacturer |
| India (Athagar) | Pujarini Dash (Indian Council of Medical Research–Regional Medical Research Centre)[474] | 2021-03-15 - 2021-04-15 | Local | Moderate | 887 | Multiple groups | All | Probability | Household and community samples | 60.3% | Roche Diagnostics - IgG, IgM, IgA - Elecsys® Anti‐SARS‐CoV‐2 (N) | 1, 0.995 | Validated by manufacturer |
| India (Badamba) | Pujarini Dash (Indian Council of Medical Research–Regional Medical Research Centre)[474] | 2021-03-15 - 2021-04-15 | Local | Moderate | 862 | Multiple groups | All | Probability | Household and community samples | 55.5% | Roche Diagnostics - IgG, IgM, IgA - Elecsys® Anti‐SARS‐CoV‐2 (N) | 1, 0.995 | Validated by manufacturer |
| India (Narsinghpur) | Pujarini Dash (Indian Council of Medical Research–Regional Medical Research Centre)[474] | 2021-03-15 - 2021-04-15 | Local | Moderate | 988 | Multiple groups | All | Probability | Household and community samples | 39.8% | Roche Diagnostics - IgG, IgM, IgA - Elecsys® Anti‐SARS‐CoV‐2 (N) | 1, 0.995 | Validated by manufacturer |
| India (Tigiria) | Pujarini Dash (Indian Council of Medical Research–Regional Medical Research Centre)[474] | 2021-03-15 - 2021-04-15 | Local | Moderate | 885 | Multiple groups | All | Probability | Household and community samples | 63.3% | Roche Diagnostics - IgG, IgM, IgA - Elecsys® Anti‐SARS‐CoV‐2 (N) | 1, 0.995 | Validated by manufacturer |
| India (Chennai) | Matt Hitchings (The Johns Hopkins Bloomberg School of Public Health)[475] | 2021-01-15 - 2021-05-15 | Local | Moderate | 4,629 | Multiple groups | All | Probability | Household and community samples | 62% | Abbott Laboratories - IgG - AdviseDX SARS-CoV-2 anti-IgG II assay | 0.981, 0.996 | Validated by manufacturer |
| India (Ahmedabad) | Om Prakash (Nathiba Hargovandas Lakhmichand (NHL) Municipal Medical College)[476] | 2021-02-22 - 2021-03-01 | Local | Moderate | 10,136 | Multiple groups | All | Stratified non-probability | Household and community samples | 27.9% | Zydus Diagnostics - IgG - Covid Kavach™ Anti-SARS CoV-2 IgG Antibody Detection ELISA | 0.987, 1 | Validated by manufacturer |
| India (Hyderabad) | Avula Laxmaiah (CSIR-CentreForCellularAndMolecularBiology)[477] | 2021-01-08 - 2021-01-24 | Local | Moderate | 9,363 | Multiple groups | All | Probability | Household and community samples | 54.2% | Roche Diagnostics - IgG, IgM, IgA - Elecsys® Anti‐SARS‐CoV‐2 (N) | 1, 0.995 | Validated by manufacturer |
| India (Vellore) | Divya Dayanand (Christian Medical College)[473] | 2021-01-01 - 2021-01-31 | Local | Low | 1,228 | Multiple groups | All | Probability | Household and community samples | 28.5% | InBios International, Inc. - IgG - anti-SARS-CoV-2 IgM and IgG | 0.919, 0.99 | Validated by manufacturer |
| India (South Andaman district) | Deepak Kumar (Andaman and Nicobar Islands Institute of Medical Sciences)[478] | 2020-12-15 - 2021-02-14 | Local | Moderate | 4,089 | Multiple groups | All | Probability | Household and community samples | 39.7% | Erba Manneim - IgG - ErbaLisa® COVID-19 IgG semi quantitative kit | 0.991, 0.993 | Validated by manufacturer |
| India (Jabalpur) | Sudarshan Ramaswamy (National Centre for Disease Control)[479] | 2020-12-11 - 2020-12-21 | Local | Low | 9,279 | Multiple groups | All | Probability | Household and community samples | 28.7% | Zydus Diagnostics - IgG - Covid Kavach™ Anti-SARS CoV-2 IgG Antibody Detection ELISA | 0.987, 1 | Validated by manufacturer |
| India (Cuttack) | Smita Mahapatra (Sriram Chandra Bhanja Medical College and Hospital)[480] | 2020-11-01 - 2021-01-31 | Local | High | 1,032 | Adults (18-64 years) | All | Convenience | Blood donors | 35.9% | Abbott Laboratories - IgG - Abbott Architect SARS-CoV-2 IgG | 0.99, 1 | Validated by manufacturer |
| India (New Delhi) | Alpesh Goyal (All India Institue of Medical Sciences)[481] | 2020-08-15 - 2021-02-15 | Local | High | 240 | Adults (18-64 years) | All | Convenience | Multiple general populations | 45.4% | DiaSorin - IgG - Liaison SARS-CoV-2 S1/S2 IgG | 0.974, 0.985 | Validated by manufacturer |
| India (Tamil Nadu) | Rita Isaac (University of Edinburgh)[482] | 2020-11-06 - 2020-11-20 | Local | Moderate | 200 | Multiple groups | All | Probability | Household and community samples | 22% | Siemens - IgG, IgM, IgA - Atellica® IM SARS-CoV-2 Total (COV2T),Roche Diagnostics - IgG, IgM, IgA - Elecsys® Anti‐SARS‐CoV‐2 (N) | 1, 0.993 | Complex test algorithm: Manual calculation |
| India (Ahmedabad) | Om Prakash (Municipal Medical College)[483] | 2020-10-15 - 2020-10-31 | Local | Moderate | 17,009 | Multiple groups | All | Convenience | Household and community samples | 24.2% | Zydus Diagnostics - IgG - Covid Kavach™ Anti-SARS CoV-2 IgG Antibody Detection ELISA | 0.987, 1 | Validated by manufacturer |
| India (Srinagar) | Taha Ayub (Government Medical College Srinagar)[484] | 2020-10-17 - 2020-10-20 | Local | Moderate | 2,418 | Multiple groups | All | Probability | Household and community samples | 40.6% | Abbott Laboratories - IgG - Abbott Architect SARS-CoV-2 IgG | 0.99, 1 | Validated by manufacturer |
| India (Pimpri-Chinchwad) | Amitav Banerjee (Dr. D. Y. Patil Medical College, Hospital and Research Centre)[485] | 2020-10-07 - 2020-10-17 | Local | Moderate | 5,000 | Multiple groups | All | Probability | Household and community samples | 34% | Abbott Laboratories - IgG - Abbott Architect SARS-CoV-2 IgG | 0.99, 1 | Validated by manufacturer |
| India (Chennai) | Muthusamy Kumar (National Institute of Epidemiology)[486] | 2020-10-08 - 2020-10-15 | Local | Low | 6,366 | Multiple groups | All | Probability | Household and community samples | 30.4% | Abbott Laboratories - IgG - Abbott Architect SARS-CoV-2 IgG | 0.99, 1 | Validated by manufacturer |
| India (Bhubaneswar) | Jaya Singh Kshatri (Indian Council of Medical Research)[487] | 2020-09-01 - 2020-09-30 | Local | Low | 1,403 | Multiple groups | All | Probability | Household and community samples | 49% | Roche Diagnostics - IgG, IgM, IgA - Elecsys® Anti‐SARS‐CoV‐2 (N) | 1, 0.995 | Validated by manufacturer |
| India (Ahmedabad) | Om Prakash (AMC MET Medical College)[488] | 2020-08-15 - 2020-08-31 | Local | Moderate | 10,310 | Multiple groups | All | Probability | Household and community samples | 23.2% | Zydus Diagnostics - IgG - Covid Kavach™ Anti-SARS CoV-2 IgG Antibody Detection ELISA | 0.987, 1 | Validated by manufacturer |
| India (Mumbai) | Ullas Kolthur Seetharam (NITI-Aayog)[489] | 2020-08-16 - 2020-08-29 | Local | Low | 2,176 | Multiple groups | All | Probability | Household and community samples | 17.5% | Abbott Laboratories - IgG - Abbott Architect SARS-CoV-2 IgG | 0.99, 1 | Validated by manufacturer |
| India (Mumbai) | Ullas Kolthur Seetharam (NITI-Aayog)[489] | 2020-08-16 - 2020-08-29 | Local | Low | 3,024 | Multiple groups | All | Probability | Persons living in slums | 44.9% | Abbott Laboratories - IgG - Abbott Architect SARS-CoV-2 IgG | 0.99, 1 | Validated by manufacturer |
| India (Indore) | Salil Sakalle (National Centre for Disease Control)[490] | 2020-08-11 - 2020-08-23 | Local | Low | 7,103 | Multiple groups | All | Probability | Household and community samples | 7.7% | Zydus Diagnostics - IgG - Covid Kavach™ Anti-SARS CoV-2 IgG Antibody Detection ELISA | 0.987, 1 | Validated by manufacturer |
| India (Berhampur) | Jaya Singh Kshatri (ICMR-Regional Medical Research Centre)[491] | 2020-08-01 - 2020-08-31 | Local | Low | 1,375 | Multiple groups | All | Probability | Household and community samples | 31.1% | Roche Diagnostics - IgG, IgM, IgA - Elecsys® Anti‐SARS‐CoV‐2 (N) | 1, 0.995 | Validated by manufacturer |
| India (Bhubaneswar) | Jaya Singh Kshatri (ICMR-Regional Medical Research Centre)[491] | 2020-08-01 - 2020-08-31 | Local | Low | 1,319 | Multiple groups | All | Probability | Household and community samples | 5.2% | Roche Diagnostics - IgG, IgM, IgA - Elecsys® Anti‐SARS‐CoV‐2 (N) | 1, 0.995 | Validated by manufacturer |
| India (Rourkela) | Jaya Singh Kshatri (ICMR-Regional Medical Research Centre)[491] | 2020-08-01 - 2020-08-31 | Local | Low | 1,452 | Multiple groups | All | Probability | Household and community samples | 24.6% | Roche Diagnostics - IgG, IgM, IgA - Elecsys® Anti‐SARS‐CoV‐2 (N) | 1, 0.995 | Validated by manufacturer |
| India (Malegaon) | Pallavi Saple (The Municipality of Malegaon )[492] | 2020-07-25 - 2020-08-20 | Local | High | 336 | Multiple groups | All | Stratified non-probability | Household and community samples | 40% | NA - - Not reported/ Unable to specify | 0.93, 1 | Validated by independent authors/third party/non-developers |
| India (Tamil Nadu) | Rita Isaac (University of Edinburgh)[482] | 2020-07-06 - 2020-08-20 | Local | Moderate | 501 | Multiple groups | All | Probability | Household and community samples | 2.2% | Siemens - IgG, IgM, IgA - Atellica® IM SARS-CoV-2 Total (COV2T),Roche Diagnostics - IgG, IgM, IgA - Elecsys® Anti‐SARS‐CoV‐2 (N) | 1, 0.993 | Complex test algorithm: Manual calculation |
| India (Chennai) | Sriram Selvaraju (National Institute of Epidemiology)[493] | 2020-07-01 - 2020-07-31 | Local | Low | 12,405 | Multiple groups | All | Probability | Household and community samples | 18.7% | Abbott Laboratories - IgG - Abbott Architect SARS-CoV-2 IgG | 0.99, 1 | Validated by manufacturer |
| India (Mumbai) | Anup Malani (Tata Institute for Fundamental Research)[494] | 2020-06-29 - 2020-07-19 | Local | Low | 2,702 | Multiple groups | All | Probability | Household and community samples | 16% | Abbott Laboratories - IgG - Abbott Architect SARS-CoV-2 IgG | 0.99, 1 | Validated by manufacturer |
| India (Mumbai) | Anup Malani (Tata Institute for Fundamental Research)[494] | 2020-06-29 - 2020-07-19 | Local | Low | 4,202 | Multiple groups | All | Probability | Persons living in slums | 54.1% | Abbott Laboratories - IgG - Abbott Architect SARS-CoV-2 IgG | 0.99, 1 | Validated by manufacturer |
| India (Ahmedabad) | Om Prakash ( Municipal Medical College )[495] | 2020-06-16 - 2020-07-07 | Local | Moderate | 29,891 | Multiple groups | All | Convenience | Residual sera | 17.6% | Zydus Diagnostics - IgG - Covid Kavach™ Anti-SARS CoV-2 IgG Antibody Detection ELISA | 0.987, 1 | Validated by manufacturer |
| India | Hem Chandra Pandey (All India Institute of Medical Sciences )[496] | 2020-04-15 - 2020-07-15 | Local | High | 1,191 | Adults (18-64 years) | All | Convenience | Blood donors | 9.5% | Abbott Laboratories - IgG - Abbott Architect SARS-CoV-2 IgG | 0.99, 1 | Validated by manufacturer |
| India (Karnataka) | Sindhulina Chandrasingh (Bangalore Baptist Hospital)[497] | 2020-05-15 - 2020-06-15 | Subnational | High | 210 | Multiple groups | All | Convenience | Blood donors | 0% | Roche Diagnostics - IgG, IgM, IgA - Elecsys® Anti‐SARS‐CoV‐2 (N) | 1, 0.995 | Validated by manufacturer |
| India (Mumbai) | Viswanath Billa (University of Pittsburgh Medical Center)[498] | 2020-03-24 - 2020-05-17 | Local | High | 110 | Multiple groups | All | Convenience | Residual sera | 37.3% | EUROIMMUN - IgG - Anti-SARS-CoV-2 ELISA IgG | 0.9, 1 | Validated by manufacturer |
| Maldives (Male’) | Raheema Abdul-Raheem (Maldives National University)[499] | 2020-10-15 - 2020-11-16 | Local | Moderate | 1,940 | Multiple groups | All | Probability | Household and community samples | 13.4% | Ortho Clinical Diagnostics Inc. - IgG, IgM, IgA - VITROS Immunodiagnostic Products Anti-SARS-CoV-2 Total | 1, 1 | Validated by manufacturer |

###### Western Pacific region

| Country (Location) | Author (Organization) | Sampling Dates (YMD) | Geographic scope | Overall risk of bias | Sample size | Age | Sex | Sampling method | Sample frame | Seroprevalence (95% CI) | Test Manufacturers - Isotypes - Names | Test Sens, Spec | Sens, Spec Source |
| --- | --- | --- | --- | --- | --- | --- | --- | --- | --- | --- | --- | --- | --- |
| Australia (Sydney, Melbourne, Brisbane, Adelaide, Perth) | Kristine Macartney, Nicholas Wood, Archana Koirala (Australian National Centre for Immunisation Research and Surveillance)[@] | 2020-11-01 - 2021-03-13 | Subnational | High | 1,678 | Multiple groups | All | Convenience | Residual sera | 0.4% (95% CI 0.2-0.9%) | Beijing Wantai Biological - IgG IgM IgA - Wantai SARS-CoV-2 Total Ab ELISA | 0.967, 0.975 | Validated by manufacturer |
| Australia (Metropolitan Melbourne) | Dorothy Machalek (University of New South Wales)[500] | 2020-11-23 - 2020-12-17 | Local | Low | 4,799 | Adults (18-64 years) | All | Probability | Blood donors | 1.6% | Beijing Wantai Biological - IgG, IgM, IgA - Wantai SARS-CoV-2 Total Ab ELISA | 0.967, 0.975 | Validated by manufacturer |
| Australia (Melbourne) | Dorothy Machalek (The Kirby Institue at the University of New South Wales and Australian National Centre for Immunisation Research and Surveillance)[501] | 2020-11-23 - 2020-12-17 | Local | High | 4,799 | Multiple groups | All | Convenience | Blood donors | 1.6% (95% CI 1.3-2%) | Beijing Wantai Biological - IgG IgM IgA - Wantai SARS-CoV-2 Total Ab ELISA | 0.967, 0.975 | Validated by manufacturer |
| Australia (Sydney) | Heather F Gidding (Children’s Hospital at Westmead)[502] | 2020-05-04 - 2020-05-23 | Local | Moderate | 560 | Adults (18-64 years) | Female | Probability | Pregnant or parturient women | 1% | NA - IgG, IgM - Author designed (IFA) - MULTIPLEXED | 0.907, 0.993 | Validated by independent authors/third party/non-developers |
| Australia (Sydney) | Heather F Gidding (Children’s Hospital at Westmead)[502] | 2020-04-20 - 2020-05-22 | Local | Moderate | 3,231 | Multiple groups | All | Probability | Residual sera | 0.6% | NA - - Author designed (IFA) - Unknown | 0.907, 0.993 | Validated by independent authors/third party/non-developers |
| China (Taipei) | Hsiang-Ling Ho (Taipei Veterans General Hospital)[503] | 2020-07-06 - 2020-07-08 | Local | Moderate | 4,988 | Multiple groups | All | Sequential | Residual sera | 0.1% | Academia Sinica - IgG, IgM - Academia Sinica ELISA assay,Roche Diagnostics - IgG, IgM, IgA - Elecsys® Anti‐SARS‐CoV‐2 (N) | 1, 1 | Complex test algorithm: Manual calculation |
| China (Wuhan) | Lu He (Renmin Hospital of Wuhan University)[504] | 2020-05-15 - 2020-05-29 | Local | Moderate | 4,454 | Multiple groups | All | Convenience | Household and community samples | 2.9% | Zhuhai Livzon Diagnostics Inc - IgG, IgM - 2019-nCoV IgG/IgM Antibody Detection Kit | 0.906, 0.992 | Validated by manufacturer |
| China (Shenzen) | Zhen Zhang (Shenzhen Center for Disease Control and Prevention)[505] | 2020-04-17 - 2020-04-23 | Local | High | 350 | Multiple groups | All | Convenience | Household and community samples | 0.3% | Beijing Wantai Biological - IgG, IgM, IgA - Wantai SARS-CoV-2 Total Ab ELISA | 0.967, 0.975 | Validated by manufacturer |
| China (Shenzen) | Zhen Zhang (Shenzhen Center for Disease Control and Prevention)[505] | 2020-04-17 - 2020-04-23 | Local | High | 50 | Multiple groups | All | Convenience | Household and community samples | 0% | Beijing Wantai Biological - IgG, IgM, IgA - Wantai SARS-CoV-2 Total Ab ELISA | 0.967, 0.975 | Validated by manufacturer |
| China (Guangzhou) | Ru Xu (Institute of Blood Transfusion)[506] | 2020-03-23 - 2020-04-02 | Local | Moderate | 2,199 | Adults (18-64 years) | All | Probability | Blood donors | 0.3% | Beijing Wantai Biological - IgG, IgM, IgA - Wantai SARS-CoV-2 Total Ab ELISA | 0.967, 0.975 | Validated by manufacturer |
| China (Liaoning) | Jin Zhang (Shengjing Hospital of China Medical University)[507] | 2020-01-21 - 2020-02-16 | Subnational | High | 225 | Multiple groups | All | Convenience | Residual sera | 0.4% | Shenzhen Yhlo Biotech Co. Ltd - IgG, IgM - iFlash-SARS-CoV-2 IgM/IgG | 1, 0.9956 | Validated by independent authors/third party/non-developers |
| Japan (Yokohama) | Kosuke Mori (Yokohama Municipal Citizen’s Hospital)[508] | 2021-02-01 - 2021-05-31 | Local | High | 1,070 | Multiple groups | All | Convenience | Residual sera | 3.3% | Abbott Laboratories - IgG - SARS-CoV-2 IgG II Quant | 0.994, 0.996 | Validated by manufacturer |
| Japan (Okinawa Prefecture) | Kenji Mizumoto (Okinawa Prefecture Epidemiological Statistics and Analysis Committee)[509] | 2021-02-01 - 2021-02-28 | Local | Moderate | 582 | Multiple groups | All | Convenience | Household and community samples | 1.4% | NA - IgG - Author designed (ELISA) -Spike | 0.925, 1 | Validated by independent authors/third party/non-developers |
| Japan (Okinawa Prefecture) | Kenji Mizumoto (Okinawa Prefecture Epidemiological Statistics and Analysis Committee)[509] | 2021-02-01 - 2021-02-28 | Local | Moderate | 123 | Multiple groups | All | Convenience | Household and community samples | 1.6% | NA - IgG - Author designed (ELISA) -Spike | 0.925, 1 | Validated by independent authors/third party/non-developers |
| Japan (Okinawa Prefecture) | Kenji Mizumoto (Okinawa Prefecture Epidemiological Statistics and Analysis Committee)[509] | 2020-10-01 - 2020-12-31 | Local | Moderate | 1,448 | Multiple groups | All | Convenience | Household and community samples | 0.6% | NA - IgG - Author designed (ELISA) -Spike | 0.925, 1 | Validated by independent authors/third party/non-developers |
| Japan (Okinawa Prefecture) | Kenji Mizumoto (Okinawa Prefecture Epidemiological Statistics and Analysis Committee)[509] | 2020-10-01 - 2020-12-31 | Local | Moderate | 144 | Multiple groups | All | Convenience | Household and community samples | 0% | NA - IgG - Author designed (ELISA) -Spike | 0.925, 1 | Validated by independent authors/third party/non-developers |
| Japan (Okinawa Prefecture) | Kenji Mizumoto (Okinawa Prefecture Epidemiological Statistics and Analysis Committee)[509] | 2020-07-01 - 2020-07-31 | Local | Moderate | 392 | Multiple groups | All | Convenience | Household and community samples | 0% | NA - IgG - Author designed (ELISA) -Spike | 0.925, 1 | Validated by independent authors/third party/non-developers |
| Japan (Yamagata) | Keita Morikane (Yamagata University)[510] | 2020-06-01 - 2020-06-04 | Local | High | 1,009 | Multiple groups | All | Sequential | Residual sera | 0.5% | Roche Diagnostics - IgG, IgM, IgA - Elecsys® Anti‐SARS‐CoV‐2 (N) | 1, 0.995 | Validated by manufacturer |
| Republic of Korea | Kwangmin Lee (Korea Disease Control and Prevention Agency)[511] | 2020-08-14 - 2020-10-31 | National | High | 1,379 | Multiple groups | All | Convenience | Residual sera | 0.2% | NA - - Author designed (Neutralization Assay) | 0.933, 0.971 | Validated by independent authors/third party/non-developers |
| Republic of Korea | Kwangmin Lee (Korea Disease Control and Prevention Agency)[511] | 2020-06-10 - 2020-08-13 | National | High | 1,440 | Multiple groups | All | Convenience | Residual sera | 0.1% | NA - - Author designed (Neutralization Assay) | 0.933, 0.971 | Validated by independent authors/third party/non-developers |
| Republic of Korea (Chungbuk Province) | Hye Won Jeong (Chungbuk National University)[512] | 2020-05-01 - 2020-07-17 | Local | Low | 3,981 | Multiple groups | All | Probability | Residual sera | 0.8% | NA - - Author designed (IFA) - Unknown | 0.995, 1 |  |
| Republic of Korea (Daegu) | Hye Won Jeong (Chungbuk National University)[512] | 2020-05-01 - 2020-07-17 | Local | Low | 3,268 | Multiple groups | All | Probability | Residual sera | 1.2% | NA - - Author designed (IFA) - Unknown | 0.995, 1 | Validated by developers |
| Republic of Korea | Kwangmin Lee (Korea Disease Control and Prevention Agency)[511] | 2020-04-21 - 2020-06-16 | National | High | 1,500 | Multiple groups | All | Convenience | Residual sera | 0% | NA - - Author designed (Neutralization Assay) | 0.933, 0.971 | Validated by independent authors/third party/non-developers |
| Singapore | Hannah E Clapham (National University of Singapore)[513] | 2020-11-15 - 2020-12-15 | National | High | 937 | Multiple groups | All | Convenience | Household and community samples | 0.2% | GenScript - IgG, IgM - cPass SARS-CoV-2 Surrogate Virus Neutralization Test ELISA Kit | 0.938, 0.994 | Validated by manufacturer |

#### S4.2 Risk of Bias breakdown for all studies

##### Table S8. Risk of bias breakdown for all studies.

###### Africa region

| Country | Author | Item 1: Appropriate sample frame | Item 2: Probability sampling method | Item 3: Adequate sample size | Item 4: Subjects & setting described | Item 5: Good coverage of sample | Item 6: Sens>=90%, Spec>=97% | Item 7: Same test for all subjects | Item 8: Appropriate statistical analysis | Item 9: Adequate response rate | Overall risk of bias |
| --- | --- | --- | --- | --- | --- | --- | --- | --- | --- | --- | --- |
| Angola | Cruz Sebastiao [214] | No | No | No | Yes | No | Yes | Yes | No | N/A | High |
| Burkina Faso | Dr Isidore Traore [215] | Yes | Yes | Yes | Yes | Yes | Yes | Yes | No | Yes | Moderate |
| Cameroon | Kene Nwosu [216] | Yes | Yes | Yes | Yes | Yes | No | Yes | Yes | Yes | Low |
| Cameroon | Tiffany Harris [1] | Yes | No | Yes | Yes | Yes | Yes | Yes | Yes | Yes | Moderate |
| Central African Republic | Alexandre Manirakiza [217] | Yes | Yes | Yes | Yes | Yes | Yes | Yes | No | Unclear | Moderate |
| Congo | Novy Bonguili [218] | No | No | Yes | Yes | No | Unclear | Yes | No | N/A | High |
| Democratic Republic of the Congo | Antoine Nkuba [219] | Yes | Yes | Yes | Yes | Yes | Unclear | Yes | Yes | Yes | Low |
| Democratic Republic of the Congo | Makiala Mandanda Sheila [125] | Yes | Yes | Yes | Yes | Unclear | Yes | Yes | No | Yes | Moderate |
| Ethiopia | Nega Assefa [126] | No | Yes | Yes | No | Unclear | Yes | Yes | No | Unclear | Moderate |
| Ethiopia | Esayas Gudina [220] | Yes | No | No | No | Unclear | Yes | Yes | No | Unclear | High |
| Ethiopia | Esayas Gudina [220] | Yes | No | No | No | Unclear | Yes | Yes | No | Unclear | High |
| Ethiopia | Esayas Gudina [220] | Yes | No | No | No | Unclear | Yes | Yes | No | Unclear | High |
| Ethiopia | Esayas Gudina [220] | Yes | No | No | No | Unclear | Yes | Yes | No | Unclear | High |
| Ethiopia | Esayas Gudina [220] | Yes | No | No | No | Unclear | Yes | Yes | No | Unclear | High |
| Ethiopia | Esayas Gudina [220] | Yes | No | No | No | Unclear | Yes | Yes | No | Unclear | High |
| Ethiopia | Esayas Gudina [220] | Yes | No | No | No | Unclear | Yes | Yes | No | Unclear | High |
| Ethiopia | Esayas Gudina [220] | Yes | No | No | No | Unclear | Yes | Yes | No | Unclear | High |
| Ethiopia | Esayas Gudina [220] | Yes | No | No | No | Unclear | Yes | Yes | No | Unclear | High |
| Ethiopia | Esayas Gudina [220] | Yes | No | No | No | Unclear | Yes | Yes | No | Unclear | High |
| Ethiopia | Esayas Gudina [220] | Yes | No | No | No | Unclear | Yes | Yes | No | Unclear | High |
| Ethiopia | Esayas Gudina [220] | Yes | No | No | No | Unclear | Yes | Yes | No | Unclear | High |
| Ethiopia | Enyew Birru Tadesse [2] | Yes | Yes | Yes | Yes | Yes | Yes | Yes | Yes | Unclear | Low |
| Ethiopia | Tamrat Shaweno [222] | Yes | Yes | Yes | Yes | Yes | Yes | Yes | No | Yes | Moderate |
| Ethiopia | Saro Abdella [221] | Yes | Yes | Yes | Yes | Yes | Yes | Yes | Yes | Unclear | Low |
| Ethiopia | Saro Abdella [221] | Yes | Yes | Yes | Yes | Yes | Yes | Yes | Yes | Unclear | Low |
| Ethiopia | John Kempen [223] | No | No | No | Yes | No | Yes | Yes | No | Unclear | High |
| Gabon | Amandine Mveang Nzoghe [224] | No | No | Yes | No | Unclear | Yes | Yes | No | Unclear | High |
| Gabon | Dr Paulin ESSONE NDONG, Pr Edgar Brice NGOUNGOU, Pr Ayola Akim ADEGNIKA [3] | Yes | Yes | Yes | Yes | Unclear | Yes | Yes | No | Unclear | Moderate |
| Ghana | Irene Owusu Donkor [4] | Yes | Yes | Yes | Yes | Yes | Yes | Yes | No | Yes | Moderate |
| Ghana | Irene Owusu Donkor [4] | Yes | Yes | Yes | Yes | Yes | Yes | Yes | No | Yes | Moderate |
| Ghana | Irene Owusu Donkor [4] | Yes | Yes | Yes | Yes | Yes | Yes | Yes | No | Yes | Moderate |
| Ghana | Irene Owusu Donkor [4] | Yes | Yes | Yes | Yes | Yes | Yes | Yes | No | Yes | Moderate |
| Kenya | R Lucinde [225] | No | No | No | No | Yes | Unclear | Yes | No | N/A | High |
| Kenya | R Lucinde [225] | No | No | No | No | Yes | Unclear | Yes | No | N/A | High |
| Kenya | R Lucinde [225] | No | No | No | No | Yes | Unclear | Yes | No | N/A | High |
| Kenya | R Lucinde [225] | No | No | No | No | Yes | Unclear | Yes | No | N/A | High |
| Kenya | R Lucinde [225] | No | No | No | No | Yes | Unclear | Yes | No | N/A | High |
| Kenya | R Lucinde [225] | No | No | No | No | Yes | Unclear | Yes | No | N/A | High |
| Kenya | R Lucinde [225] | No | No | No | No | Yes | Unclear | Yes | No | N/A | High |
| Kenya | R Lucinde [225] | No | No | No | No | Yes | Unclear | Yes | No | N/A | High |
| Kenya | R Lucinde [225] | No | No | No | No | Yes | Unclear | Yes | No | N/A | High |
| Kenya | Anthony Etyang [227] | Yes | Yes | Yes | Yes | Yes | Unclear | Yes | Yes | Unclear | Low |
| Kenya | Anthony Etyang [227] | Yes | Yes | Yes | Yes | Yes | Unclear | Yes | Yes | Unclear | Low |
| Kenya | Anthony Etyang [227] | Yes | Yes | Yes | Yes | Yes | Unclear | Yes | Yes | Unclear | Low |
| Kenya | James Nyagwange [226] | No | No | No | No | No | Yes | Yes | No | N/A | High |
| Kenya | James Nyagwange [226] | No | No | No | No | No | Yes | Yes | No | N/A | High |
| Kenya | Isaac Ngere [230] | Yes | Yes | Yes | Yes | Yes | Yes | Yes | Yes | Yes | Low |
| Kenya | Sophie Uyoga [5] | No | No | Yes | Yes | No | Unclear | Yes | Yes | N/A | Moderate |
| Kenya | Trevor Crowell [231] | No | No | No | Yes | Unclear | Yes | Yes | No | N/A | High |
| Kenya | Patrick Munywoki [228] | Yes | Yes | No | Yes | Yes | Yes | Yes | Yes | Yes | Moderate |
| Kenya | Ifedayo M. O. Adetifa [6] | Yes | No | Yes | Yes | No | Unclear | Yes | Yes | N/A | Moderate |
| Kenya | R Lucinde [229] | No | No | No | No | Unclear | Unclear | Yes | No | Unclear | High |
| Kenya | R Lucinde [229] | No | No | No | No | Unclear | Unclear | Yes | No | Unclear | High |
| Kenya | R Lucinde [229] | No | No | No | No | Unclear | Unclear | Yes | No | Unclear | High |
| Kenya | R Lucinde [229] | No | No | No | No | Unclear | Unclear | Yes | No | Unclear | High |
| Madagascar | Solohery Razafimahatratra [232] | No | No | No | Yes | Unclear | Yes | Yes | No | N/A | High |
| Madagascar | Solohery Razafimahatratra [232] | No | No | No | Yes | Unclear | Yes | Yes | No | N/A | High |
| Madagascar | Solohery Razafimahatratra [232] | No | No | No | Yes | Unclear | Yes | Yes | No | N/A | High |
| Madagascar | Solohery Razafimahatratra [232] | No | No | No | Yes | Unclear | Yes | Yes | No | N/A | High |
| Madagascar | Solohery Razafimahatratra [232] | No | No | No | Yes | Unclear | Yes | Yes | No | N/A | High |
| Madagascar | Solohery Razafimahatratra [232] | No | No | No | Yes | Unclear | Yes | Yes | No | N/A | High |
| Madagascar | Solohery Razafimahatratra [232] | No | No | No | Yes | Unclear | Yes | Yes | No | N/A | High |
| Madagascar | Solohery Razafimahatratra [232] | No | No | No | Yes | Unclear | Yes | Yes | No | N/A | High |
| Madagascar | Matthieu Schoenhals [233] | No | No | No | No | No | Yes | Yes | Yes | Unclear | High |
| Madagascar | Matthieu Schoenhals [233] | No | No | No | No | No | Yes | Yes | Yes | Unclear | High |
| Madagascar | Matthieu Schoenhals [233] | No | No | No | No | No | Yes | Yes | Yes | Unclear | High |
| Madagascar | Matthieu Schoenhals [233] | No | No | No | No | No | Yes | Yes | Yes | Unclear | High |
| Madagascar | Matthieu Schoenhals [233] | No | No | No | No | No | Yes | Yes | Yes | Unclear | High |
| Madagascar | Matthieu Schoenhals [233] | No | No | No | No | No | Yes | Yes | Yes | Unclear | High |
| Madagascar | Matthieu Schoenhals [233] | No | No | No | No | No | Yes | Yes | Yes | Unclear | High |
| Madagascar | Matthieu Schoenhals [233] | No | No | No | No | No | Yes | Yes | Yes | Unclear | High |
| Madagascar | Matthieu Schoenhals [233] | No | No | No | No | No | Yes | Yes | Yes | Unclear | High |
| Madagascar | Matthieu Schoenhals [233] | No | No | No | No | No | Yes | Yes | Yes | Unclear | High |
| Madagascar | Matthieu Schoenhals [233] | No | No | No | No | No | Yes | Yes | Yes | Unclear | High |
| Madagascar | Matthieu Schoenhals [233] | No | No | No | No | No | Yes | Yes | Yes | Unclear | High |
| Madagascar | Matthieu Schoenhals [233] | No | No | No | No | No | Yes | Yes | Yes | Unclear | High |
| Madagascar | Matthieu Schoenhals [233] | No | No | No | No | No | Yes | Yes | Yes | Unclear | High |
| Madagascar | Matthieu Schoenhals [233] | No | No | No | No | No | Yes | Yes | Yes | Unclear | High |
| Madagascar | Matthieu Schoenhals [233] | No | No | No | No | No | Yes | Yes | Yes | Unclear | High |
| Madagascar | Matthieu Schoenhals [233] | No | No | No | No | No | Yes | Yes | Yes | Unclear | High |
| Madagascar | Matthieu Schoenhals [233] | No | No | No | No | No | Yes | Yes | Yes | Unclear | High |
| Madagascar | Matthieu Schoenhals [233] | No | No | No | No | No | Yes | Yes | Yes | Unclear | High |
| Madagascar | Matthieu Schoenhals [233] | No | No | No | No | No | Yes | Yes | Yes | Unclear | High |
| Madagascar | Matthieu Schoenhals [233] | No | No | No | No | No | Yes | Yes | Yes | Unclear | High |
| Madagascar | Matthieu Schoenhals [233] | No | No | No | No | No | Yes | Yes | Yes | Unclear | High |
| Madagascar | Matthieu Schoenhals [233] | No | No | No | No | No | Yes | Yes | Yes | Unclear | High |
| Madagascar | Matthieu Schoenhals [233] | No | No | No | No | No | Yes | Yes | Yes | Unclear | High |
| Madagascar | Matthieu Schoenhals [233] | No | No | No | No | No | Yes | Yes | Yes | Unclear | High |
| Madagascar | Matthieu Schoenhals [233] | No | No | No | No | No | Yes | Yes | Yes | Unclear | High |
| Madagascar | Matthieu Schoenhals [233] | No | No | No | No | No | Yes | Yes | Yes | Unclear | High |
| Madagascar | Matthieu Schoenhals [233] | No | No | No | No | No | Yes | Yes | Yes | Unclear | High |
| Madagascar | Matthieu Schoenhals [233] | No | No | No | No | No | Yes | Yes | Yes | Unclear | High |
| Madagascar | Matthieu Schoenhals [233] | No | No | No | No | No | Yes | Yes | Yes | Unclear | High |
| Madagascar | Matthieu Schoenhals [233] | No | No | No | No | No | Yes | Yes | Yes | Unclear | High |
| Madagascar | Matthieu Schoenhals [233] | No | No | No | No | No | Yes | Yes | Yes | Unclear | High |
| Madagascar | Matthieu Schoenhals [233] | No | No | No | No | No | Yes | Yes | Yes | Unclear | High |
| Malawi | Kondwani Jambo [7] | No | Yes | Yes | Yes | No | Yes | Yes | Yes | N/A | Low |
| Malawi | Kondwani Jambo [7] | No | Yes | Yes | Yes | No | Yes | Yes | Yes | N/A | Low |
| Malawi | Kondwani Jambo [7] | No | Yes | Yes | Yes | No | Yes | Yes | Yes | N/A | Low |
| Malawi | Kondwani Jambo [7] | No | Yes | Yes | Yes | No | Yes | Yes | Yes | N/A | Low |
| Malawi | Kondwani Jambo [7] | No | Yes | Yes | Yes | No | Yes | Yes | Yes | N/A | Low |
| Malawi | Kondwani Jambo [7] | No | Yes | Yes | Yes | No | Yes | Yes | Yes | N/A | Low |
| Malawi | Kondwani Jambo [7] | No | Yes | Yes | Yes | No | Yes | Yes | Yes | N/A | Low |
| Malawi | Kondwani Jambo [7] | No | Yes | Yes | Yes | No | Yes | Yes | Yes | N/A | Low |
| Malawi | Kondwani Jambo [7] | No | Yes | Yes | Yes | No | Yes | Yes | Yes | N/A | Low |
| Malawi | Kondwani Jambo [7] | No | Yes | Yes | Yes | No | Yes | Yes | Yes | N/A | Low |
| Malawi | Kondwani Jambo [7] | No | Yes | Yes | Yes | No | Yes | Yes | Yes | N/A | Low |
| Malawi | Kondwani Jambo [7] | No | Yes | Yes | Yes | No | Yes | Yes | Yes | N/A | Low |
| Malawi | Kondwani Jambo [7] | No | Yes | Yes | Yes | No | Yes | Yes | Yes | N/A | Low |
| Malawi | Kondwani Jambo [7] | No | Yes | Yes | Yes | No | Yes | Yes | Yes | N/A | Low |
| Malawi | Kondwani Jambo [7] | No | Yes | Yes | Yes | No | Yes | Yes | Yes | N/A | Low |
| Malawi | Kondwani Jambo [7] | No | Yes | Yes | Yes | No | Yes | Yes | Yes | N/A | Low |
| Malawi | Kondwani Jambo [7] | No | Yes | Yes | Yes | No | Yes | Yes | Yes | N/A | Low |
| Malawi | Kondwani Jambo [7] | No | Yes | Yes | Yes | No | Yes | Yes | Yes | N/A | Low |
| Malawi | Kondwani Jambo [7] | No | Yes | Yes | Yes | No | Yes | Yes | Yes | N/A | Low |
| Malawi | Matthews Kagoli/Joe Theu [8] | Yes | Yes | Yes | Yes | Yes | Yes | Yes | Yes | Yes | Low |
| Mali | Issaka Sagara [234] | Yes | No | Yes | No | No | Unclear | Yes | Yes | Unclear | Moderate |
| Mali | Issaka Sagara [234] | Yes | No | Yes | Yes | No | Unclear | Yes | Yes | Unclear | Moderate |
| Mali | Issaka Sagara [234] | Yes | No | Yes | No | No | Unclear | Yes | Yes | Unclear | Moderate |
| Mali | Issaka Sagara [234] | Yes | No | Yes | Yes | No | Unclear | Yes | Yes | Unclear | Moderate |
| Mali | Issaka Sagara [234] | Yes | No | No | No | No | Unclear | Yes | Yes | Unclear | Moderate |
| Mali | Issaka Sagara [234] | Yes | No | No | Yes | No | Unclear | Yes | Yes | Unclear | Moderate |
| Mozambique | Paulo Arnaldo [239] | Yes | Yes | Yes | Yes | Yes | No | Yes | No | No | Moderate |
| Mozambique | Mussagy Mahomed [235] | Yes | Yes | Yes | Yes | Unclear | No | Yes | No | Unclear | Moderate |
| Mozambique | Mussagy Mahomed [235] | Yes | Yes | Yes | Yes | Unclear | No | Yes | No | Unclear | Moderate |
| Mozambique | Arlete Mahumane [236] | Yes | Yes | Yes | Yes | Yes | No | Yes | No | Unclear | Moderate |
| Mozambique | Jeronimo Langa [237] | Yes | Yes | Yes | Yes | Unclear | No | Yes | No | Unclear | Moderate |
| Mozambique | Mussagy Mahomed [238] | Yes | Yes | Yes | Yes | Unclear | No | Yes | No | Unclear | Moderate |
| Mozambique | Eduardo Samo Gudo [240] | Yes | Yes | Yes | No | Unclear | No | Yes | No | Unclear | Moderate |
| Nigeria | OgoChukwu Vincent Okpala [127] | Yes | Yes | Yes | Yes | No | No | Yes | No | N/A | Moderate |
| Nigeria | Ijeoma Ifeorah [241] | No | No | Yes | Yes | Unclear | Yes | Yes | No | N/A | High |
| Nigeria | Hussaini Majiya [129] | Yes | Yes | No | No | Unclear | Unclear | Yes | No | Unclear | Moderate |
| Nigeria | Kristen A. Stafford (UMB); Laura Steinhardt (CDC); Elsie Ilori (NCDC); Rosemary Audu (NIMR) [128] | Yes | Yes | Yes | Yes | Yes | Yes | Yes | Yes | Yes | Low |
| Nigeria | Kristen A. Stafford (UMB); Laura Steinhardt (CDC); Elsie Ilori (NCDC); Rosemary Audu (NIMR) [128] | Yes | Yes | Yes | Yes | Yes | Yes | Yes | Yes | Yes | Low |
| Nigeria | Kristen A. Stafford (UMB); Laura Steinhardt (CDC); Elsie Ilori (NCDC); Rosemary Audu (NIMR) [128] | Yes | Yes | Yes | Yes | Yes | Yes | Yes | Yes | Yes | Low |
| Nigeria | Kristen A. Stafford (UMB); Laura Steinhardt (CDC); Elsie Ilori (NCDC); Rosemary Audu (NIMR) [128] | Yes | Yes | Yes | Yes | Yes | Yes | Yes | Yes | Yes | Low |
| Nigeria | Olatunji Kolawole [9] | No | Yes | Yes | Yes | Yes | Yes | Yes | No | Yes | Moderate |
| Senegal | Cheikh Talla [10] | Yes | Yes | Yes | Yes | Unclear | Yes | Yes | Yes | Yes | Low |
| Sierra Leone | Mohamed Bailor Barrie [11] | Yes | Yes | Yes | Yes | Unclear | Yes | Yes | Yes | Unclear | Low |
| South Africa | Jackie Kleynhans [242] | Yes | Yes | No | No | Unclear | Yes | Yes | No | Unclear | Moderate |
| South Africa | Jackie Kleynhans [242] | Yes | Yes | No | No | Unclear | Yes | Yes | No | Unclear | Moderate |
| South Africa | Jackie Kleynhans [242] | Yes | Yes | No | No | Unclear | Yes | Yes | No | Unclear | Moderate |
| South Africa | Jackie Kleynhans [242] | Yes | Yes | No | No | Unclear | Yes | Yes | No | Unclear | Moderate |
| South Africa | Jackie Kleynhans [242] | Yes | Yes | No | No | Unclear | Yes | Yes | No | Unclear | Moderate |
| South Africa | Jackie Kleynhans [242] | Yes | Yes | No | No | Unclear | Yes | Yes | No | Unclear | Moderate |
| South Africa | Jackie Kleynhans [242] | Yes | Yes | No | No | Unclear | Yes | Yes | No | Unclear | Moderate |
| South Africa | Jackie Kleynhans [242] | Yes | Yes | No | No | Unclear | Yes | Yes | No | Unclear | Moderate |
| South Africa | Nicole Wolter [12] | Yes | Yes | Yes | Yes | Unclear | Yes | Yes | No | No | Moderate |
| South Africa | Russel Cable [243] | No | No | Yes | No | Unclear | Yes | Yes | No | Unclear | High |
| South Africa | Jaya George [130] | No | No | Yes | Yes | Yes | Yes | Yes | Yes | N/A | Moderate |
| South Africa | Marion Vermeulen [13] | No | No | Yes | No | Unclear | Yes | Yes | Yes | N/A | Moderate |
| South Africa | Marvin Hsiao [246] | No | No | Yes | Yes | No | Yes | Yes | No | N/A | High |
| South Africa | Jackie Kleynhans [245] | Yes | Yes | Yes | Yes | No | Yes | Yes | No | Unclear | Moderate |
| South Africa | Jackie Kleynhans [245] | Yes | Yes | Yes | Yes | No | Yes | Yes | No | Unclear | Moderate |
| South Africa | Jackie Kleynhans [245] | Yes | Yes | Yes | Yes | No | Yes | Yes | No | Unclear | Moderate |
| South Africa | Jackie Kleynhans [245] | Yes | Yes | Yes | Yes | No | Yes | Yes | No | Unclear | Moderate |
| South Africa | Jackie Kleynhans [245] | Yes | Yes | Yes | Yes | No | Yes | Yes | No | Unclear | Moderate |
| South Africa | Jackie Kleynhans [245] | Yes | Yes | Yes | Yes | No | Yes | Yes | No | Unclear | Moderate |
| South Africa | Jackie Kleynhans [245] | Yes | Yes | Yes | Yes | No | Yes | Yes | No | Unclear | Moderate |
| South Africa | Jackie Kleynhans [245] | Yes | Yes | Yes | Yes | No | Yes | Yes | No | Unclear | Moderate |
| South Africa | Jackie Kleynhans [245] | Yes | Yes | Yes | Yes | No | Yes | Yes | No | Unclear | Moderate |
| South Africa | Jackie Kleynhans [245] | Yes | Yes | Yes | Yes | No | Yes | Yes | No | Unclear | Moderate |
| South Africa | Lee Fairlie [244] | No | No | Yes | Yes | Unclear | Yes | Yes | No | No | High |
| South Sudan | Kirsten E Wiens [247] | Yes | Yes | Yes | No | Unclear | Unclear | Yes | Yes | Unclear | Low |
| Togo | Didier K Ekouevi [14] | Yes | Yes | Yes | Yes | No | Yes | Yes | Yes | Unclear | Low |
| Uganda | Dr. Henry Kyobe Bosa [15] | Yes | Yes | Yes | Yes | Yes | Yes | Yes | No | Unclear | Moderate |
| Zambia | Kwame Shanaube [248] | Yes | Yes | Yes | Yes | No | Yes | Yes | No | Unclear | Moderate |
| Zambia | Jonas Z Hines [16] | Yes | No | Yes | No | No | Yes | Yes | No | Unclear | Moderate |
| Zambia | Lloyd B Mulenga [17] | Yes | Yes | Yes | Yes | Yes | Yes | Yes | Yes | Yes | Low |
| Zimbabwe | Arun Fryatt [249] | No | Yes | Yes | Yes | Yes | Unclear | Yes | No | Yes | Moderate |
| Zimbabwe | Arun Fryatt [249] | No | Yes | Yes | Yes | Yes | Unclear | Yes | No | Yes | Moderate |

###### Americas region

| Country | Author | Item 1: Appropriate sample frame | Item 2: Probability sampling method | Item 3: Adequate sample size | Item 4: Subjects & setting described | Item 5: Good coverage of sample | Item 6: Sens>=90%, Spec>=97% | Item 7: Same test for all subjects | Item 8: Appropriate statistical analysis | Item 9: Adequate response rate | Overall risk of bias |
| --- | --- | --- | --- | --- | --- | --- | --- | --- | --- | --- | --- |
| Argentina | Silvana Figar [251] | Yes | Yes | Yes | Yes | Yes | Unclear | Yes | Yes | Yes | Low |
| Argentina | Laura Munoz [250] | Yes | No | No | Yes | Yes | Unclear | Yes | No | Yes | High |
| Brazil | Mariam Fofana [255] | Yes | Yes | Yes | Yes | Yes | Yes | Yes | No | Yes | Moderate |
| Brazil | Carolina Ferrugini [260] | No | No | No | No | No | Yes | Yes | No | Yes | High |
| Brazil | Nayara Vale [132] | No | Yes | Yes | Yes | Yes | Yes | Yes | No | N/A | Moderate |
| Brazil | Nayara Vale [132] | No | Yes | Yes | Yes | Yes | Yes | Yes | No | N/A | Moderate |
| Brazil | Nayara Vale [132] | No | Yes | Yes | Yes | Yes | Yes | Yes | No | N/A | Moderate |
| Brazil | Daniel Chaves [137] | No | Yes | Yes | Yes | Unclear | Yes | Yes | No | N/A | Moderate |
| Brazil | Keise Pereira [253] | Unclear | No | No | Yes | Unclear | Yes | Yes | No | Unclear | High |
| Brazil | Ana Silva [263] | Yes | Yes | Yes | Yes | Yes | No | Yes | No | Unclear | Moderate |
| Brazil | Aluisio Barros [131] | Yes | Yes | Yes | Yes | Yes | Unclear | Yes | No | Unclear | Moderate |
| Brazil | Aluisio Barros [131] | Yes | Yes | Yes | Yes | Yes | Unclear | Yes | No | Unclear | Moderate |
| Brazil | Ana Schneider [135] | Yes | Yes | Yes | Yes | No | Yes | Yes | No | N/A | Moderate |
| Brazil | José Olimpio Moura de Albuquerque [133] | Yes | Yes | Yes | No | Yes | Unclear | Yes | No | No | Moderate |
| Brazil | José Olimpio Moura de Albuquerque [133] | Yes | Yes | Yes | No | Yes | Unclear | Yes | No | No | Moderate |
| Brazil | José Olimpio Moura de Albuquerque [133] | Yes | Yes | Yes | No | Yes | Unclear | Yes | No | No | Moderate |
| Brazil | José Olimpio Moura de Albuquerque [133] | Yes | Yes | Yes | No | Yes | Unclear | Yes | No | No | Moderate |
| Brazil | José Olimpio Moura de Albuquerque [133] | Yes | Yes | Yes | Yes | Yes | Unclear | Yes | No | No | Moderate |
| Brazil | José Olimpio Moura de Albuquerque [133] | Yes | Yes | Yes | No | Yes | Unclear | Yes | No | No | Moderate |
| Brazil | José Olimpio Moura de Albuquerque [133] | Yes | Yes | Yes | No | Yes | Unclear | Yes | No | No | Moderate |
| Brazil | José Olimpio Moura de Albuquerque [133] | Yes | Yes | Yes | Yes | Yes | Unclear | Yes | No | No | Moderate |
| Brazil | José Olimpio Moura de Albuquerque [133] | Yes | Yes | Yes | No | Yes | Unclear | Yes | No | No | Moderate |
| Brazil | José Olimpio Moura de Albuquerque [133] | Yes | Yes | Yes | No | Yes | Unclear | Yes | No | No | Moderate |
| Brazil | José Olimpio Moura de Albuquerque [133] | Yes | Yes | Yes | Yes | Yes | Unclear | Yes | No | No | Moderate |
| Brazil | Alanna Calheiros Santos [265] | Unclear | No | No | No | Unclear | Yes | Yes | No | Unclear | High |
| Brazil | Ethel Leonor Noia Maciel [138] | Yes | Yes | Yes | Yes | Unclear | Unclear | Yes | No | Unclear | Moderate |
| Brazil | Hilton Pereira da Silva [256] | Yes | Yes | No | Yes | Unclear | Yes | Yes | No | Yes | Moderate |
| Brazil | Gemilson Soares Pontes [258] | No | No | No | Yes | Unclear | Yes | Yes | No | N/A | High |
| Brazil | Breno Bernardes-Souza [266] | Yes | Yes | Yes | Yes | Unclear | No | Yes | No | N/A | Moderate |
| Brazil | Breno Bernardes-Souza [266] | Yes | Yes | Yes | Yes | Unclear | No | Yes | No | N/A | Moderate |
| Brazil | Beatriz Tess [268] | Yes | No | No | Yes | No | Yes | Yes | No | Unclear | High |
| Brazil | Joao Luiz Miraglia [257] | Yes | Yes | No | Yes | Unclear | Yes | Yes | Yes | Yes | Moderate |
| Brazil | Edson Zangiacomi Martinez [267] | Yes | Yes | Yes | Yes | Yes | No | Yes | No | Unclear | Moderate |
| Brazil | Edson Zangiacomi Martinez [267] | Yes | Yes | Yes | No | Yes | No | Yes | No | Unclear | Moderate |
| Brazil | Pamella Lugon [261] | Yes | No | No | No | Unclear | Yes | Yes | No | Yes | High |
| Brazil | Pedro C. Hallal [136] | Yes | Yes | Yes | Yes | Yes | No | Yes | No | Yes | Moderate |
| Brazil | Fernando C Barros [18] | Yes | Yes | Yes | No | Yes | No | Yes | No | Unclear | Moderate |
| Brazil | Beatriz Tess [252] | Yes | Yes | Yes | No | Yes | Yes | Yes | No | Unclear | Moderate |
| Brazil | City of Sao Paulo [254] | Yes | Yes | Yes | No | Unclear | Yes | Yes | No | Unclear | Moderate |
| Brazil | Henrique Diegoli [264] | Yes | Yes | Yes | No | Unclear | Unclear | Yes | No | No | Moderate |
| Brazil | Lewis Buss [259] | No | No | Yes | No | Yes | Yes | Yes | Yes | N/A | Moderate |
| Brazil | Lewis Buss [259] | No | No | Yes | No | Yes | Yes | Yes | Yes | N/A | Moderate |
| Brazil | Lewis Buss [259] | No | No | Yes | No | Yes | Yes | Yes | Yes | N/A | Moderate |
| Brazil | Lewis Buss [259] | No | No | Yes | No | Yes | Yes | Yes | Yes | N/A | Moderate |
| Brazil | Lewis Buss [259] | No | No | Yes | No | Yes | Yes | Yes | Yes | N/A | Moderate |
| Brazil | Lewis Buss [259] | No | No | Yes | No | Yes | Yes | Yes | Yes | N/A | Moderate |
| Brazil | Lewis Buss [259] | No | No | Yes | No | Yes | Yes | Yes | Yes | N/A | Moderate |
| Brazil | Lewis Buss [259] | No | No | Yes | No | Yes | Yes | Yes | Yes | N/A | Moderate |
| Brazil | Lewis Buss [259] | No | No | Yes | No | Yes | Yes | Yes | Yes | N/A | Moderate |
| Brazil | Lewis Buss [259] | No | No | Yes | No | Yes | Yes | Yes | Yes | N/A | Moderate |
| Brazil | Lewis Buss [259] | No | No | Yes | No | Yes | Yes | Yes | Yes | N/A | Moderate |
| Brazil | Lewis Buss [259] | No | No | Yes | No | Yes | Yes | Yes | Yes | N/A | Moderate |
| Brazil | Lewis Buss [259] | No | No | Yes | No | Yes | Yes | Yes | Yes | N/A | Moderate |
| Brazil | Lewis Buss [259] | No | No | Yes | No | Yes | Yes | Yes | Yes | N/A | Moderate |
| Brazil | Lewis Buss [259] | No | No | Yes | No | Yes | Yes | Yes | Yes | N/A | Moderate |
| Brazil | Lewis Buss [259] | No | No | Yes | No | Yes | Yes | Yes | Yes | N/A | Moderate |
| Brazil | Lewis Buss [259] | No | No | Yes | No | Yes | Yes | Yes | Yes | N/A | Moderate |
| Brazil | Lewis Buss [259] | No | No | Yes | No | Yes | Yes | Yes | Yes | N/A | Moderate |
| Brazil | Ricardo Queiroz Gurgel [269] | No | No | Yes | Yes | Yes | Yes | Yes | No | Unclear | High |
| Brazil | Luanda Mara da Silva Oliveira [262] | No | No | Yes | Yes | No | Yes | Yes | No | Unclear | High |
| Brazil | Antonio da Silva [134] | Yes | Yes | Yes | Yes | Yes | Yes | Yes | Yes | Yes | Low |
| Canada | Canadian Blood Service [25] | No | No | Yes | Yes | No | Yes | Yes | No | N/A | Moderate |
| Canada | David Goldfarb [285] | No | No | Yes | No | Unclear | Yes | Yes | Yes | Unclear | High |
| Canada | Antoine Lewin [143] | No | No | Yes | Yes | Yes | Unclear | Yes | Yes | N/A | Moderate |
| Canada | Guadalein Tanunliong [290] | No | Yes | No | Yes | Yes | Unclear | Yes | No | N/A | High |
| Canada | Canadian Blood Services [26] | No | No | Yes | Yes | No | Yes | Yes | Yes | N/A | Moderate |
| Canada | Ashleigh Tuite [287] | No | No | Yes | Yes | No | Yes | Yes | No | N/A | High |
| Canada | Xuyang Tang [286] | No | No | Yes | Yes | Yes | Yes | Yes | No | No | High |
| Canada | Canadian Blood Services [27] | No | No | Yes | Yes | Yes | Yes | Yes | Yes | N/A | Moderate |
| Canada | Canadian Blood Services [28] | No | Yes | Yes | Yes | Yes | Yes | Yes | Yes | N/A | Low |
| Canada | Canadian Blood Services [29] | No | Yes | Yes | Yes | Yes | Yes | Yes | Yes | N/A | Low |
| Canada | Canadian Blood Services [30] | No | Yes | Yes | Yes | No | Yes | Yes | Yes | N/A | Low |
| Canada | Canadian Blood Services [31] | No | Yes | Yes | Yes | No | Yes | Yes | Yes | N/A | Low |
| Canada | Canadian Blood Services [32] | No | Yes | Yes | Yes | Yes | Yes | Yes | Yes | N/A | Low |
| Canada | Canadian Blood Services [33] | No | No | Yes | Yes | Yes | Yes | Yes | Yes | N/A | Moderate |
| Canada | Carmen Charlton [144] | No | No | Yes | Yes | Yes | Yes | Yes | No | N/A | Moderate |
| Canada | Carmen Charlton [144] | No | No | Yes | Yes | Yes | Yes | Yes | No | N/A | Moderate |
| Canada | Carmen Charlton [144] | No | No | Yes | Yes | Yes | Yes | Yes | No | N/A | Moderate |
| Canada | Carmen Charlton [144] | No | No | Yes | Yes | Yes | Yes | Yes | No | N/A | Moderate |
| Canada | Carmen Charlton [144] | No | No | Yes | Yes | Yes | Yes | Yes | No | N/A | Moderate |
| Canada | Carmen Charlton [144] | No | No | Yes | Yes | Yes | Yes | Yes | No | N/A | Moderate |
| Canada | Carmen Charlton [144] | No | No | Yes | Yes | Yes | Yes | Yes | No | N/A | Moderate |
| Canada | Carmen Charlton [144] | No | No | Yes | Yes | No | Yes | Yes | No | N/A | Moderate |
| Canada | Canadian Blood Services [34] | No | No | Yes | Yes | Yes | Yes | Yes | Yes | N/A | Moderate |
| Canada | Canadian Blood Services [35] | No | No | Yes | Yes | Yes | Yes | Yes | Yes | N/A | Moderate |
| Canada | Canadian Blood Services [36] | No | No | Yes | Yes | Yes | Yes | Yes | Yes | N/A | Moderate |
| Canada | Hema-Quebec [289] | No | No | Yes | No | No | Unclear | Yes | Yes | Unclear | High |
| Canada | Canadian Blood Services [37] | No | No | Yes | Yes | Yes | Yes | Yes | Yes | N/A | Moderate |
| Canada | Canadian Blood Services [38] | No | No | Yes | Yes | Yes | Yes | Yes | Yes | Yes | Moderate |
| Canada | Sahar Saeed [39] | Yes | No | Yes | Yes | Yes | Yes | Yes | Yes | Unclear | Moderate |
| Canada | Canadian Blood Services [38] | No | No | Yes | Yes | Yes | Yes | Yes | Yes | Yes | Moderate |
| Canada | Canadian Blood Services [38] | No | No | Yes | Yes | Yes | Yes | Yes | Yes | Yes | Moderate |
| Canada | Emilie Manny [288] | Yes | No | No | Yes | Unclear | Yes | Yes | No | Unclear | High |
| Canada | Danuta Skowronski [146] | No | Yes | Yes | Yes | Unclear | Yes | Yes | No | Yes | Moderate |
| Canada | Danuta Skowronski [146] | No | Yes | Yes | Yes | Unclear | Yes | Yes | No | Yes | Moderate |
| Canada | Shelly Bolotin, Vanessa Tran [145] | No | No | Yes | Yes | Unclear | Yes | Yes | Yes | N/A | Moderate |
| Canada | Shelly Bolotin, Vanessa Tran [145] | No | No | Yes | Yes | Unclear | Yes | Yes | Yes | N/A | Moderate |
| Canada | Shelly Bolotin, Vanessa Tran [145] | No | No | Yes | Yes | Unclear | Yes | Yes | Yes | N/A | Moderate |
| Canada | Shelly Bolotin, Vanessa Tran [145] | No | No | Yes | Yes | Unclear | Yes | Yes | Yes | N/A | Moderate |
| Canada | Shelly Bolotin, Vanessa Tran [145] | No | No | Yes | Yes | Unclear | Yes | Yes | Yes | N/A | Moderate |
| Canada | Shelly Bolotin, Vanessa Tran [145] | No | No | Yes | Yes | Unclear | Yes | Yes | Yes | N/A | Moderate |
| Canada | Shelly Bolotin, Vanessa Tran [145] | No | No | Yes | Yes | Unclear | Yes | Yes | Yes | N/A | Moderate |
| Chile | Pablo Vial [40] | Yes | Yes | Yes | Yes | Yes | Yes | Yes | No | Yes | Low |
| Chile | Ximena Aguilera [147] | Yes | Yes | Yes | Yes | Unclear | Yes | Yes | No | Unclear | Moderate |
| Colombia | Marcela Mercado-Reye [19] | Yes | Yes | Yes | Yes | Yes | Unclear | Yes | No | Yes | Moderate |
| Colombia | Hector Serrano-Coll [270] | Unclear | No | No | Yes | Yes | Yes | Yes | No | Unclear | High |
| Colombia | Gustavo Concha [139] | Yes | No | Yes | Yes | Unclear | Yes | Yes | No | Unclear | Moderate |
| Colombia | Evelin Garay [140] | Yes | No | Yes | Yes | Unclear | Yes | Yes | No | Unclear | Moderate |
| Colombia | Nelson Alvis Guzma [141] | Yes | Yes | Yes | No | Unclear | Yes | Yes | No | Unclear | Moderate |
| Colombia | Salim Mattar [271] | Yes | Yes | Yes | No | Unclear | Yes | Yes | Yes | Unclear | Low |
| Dominican Republic | Robert Paulino Ramirez [20] | No | Yes | Yes | Yes | Unclear | No | Yes | No | Unclear | Moderate |
| Ecuador | Oscar Del Brutto [272] | No | No | Yes | Yes | Yes | Unclear | Yes | No | Yes | High |
| Ecuador | David Acurio Paez [273] | Yes | Yes | Yes | Yes | Yes | No | Yes | No | Unclear | Moderate |
| French Guiana | Claude Flamand [274] | No | No | No | Yes | No | Yes | Yes | Yes | Unclear | High |
| Honduras | Mario Rene Mejia Nunez [21] | Yes | Yes | Yes | Yes | Yes | Unclear | Yes | No | Unclear | Moderate |
| Jamaica | TeAnne Chisolm [275] | Yes | No | No | No | Unclear | Yes | Yes | No | N/A | High |
| Mexico | Miguel Fernández-Rojas [22] | No | No | Yes | Yes | Yes | Unclear | Yes | Yes | N/A | Moderate |
| Mexico | Ana Basto-Abreu [24] | Yes | Yes | Yes | Yes | Yes | Yes | Yes | No | No | Moderate |
| Mexico | Victor Monteon [276] | Yes | No | No | Yes | No | Unclear | Yes | No | Yes | High |
| Mexico | Francisco Canto-Osorio [23] | Yes | Yes | Yes | No | Unclear | Yes | Yes | Yes | No | Low |
| Mexico | Francisco Canto-Osorio [23] | Yes | Yes | Yes | No | Unclear | Yes | Yes | Yes | No | Low |
| Mexico | Natalia Martinez-Acu a [279] | No | No | Yes | Yes | Yes | Yes | Yes | No | Unclear | High |
| Mexico | Jose Munoz-Medina [277] | No | No | Yes | No | Yes | Yes | Yes | No | N/A | High |
| Mexico | Jose Maria Remes-Troche [278] | Unclear | No | Yes | Yes | Unclear | Yes | Yes | No | Unclear | High |
| Panama | Alcibiades Villarreal [291] | No | No | No | Yes | No | Unclear | Yes | No | N/A | High |
| Peru | Yordi Tarazona-Castro [281] | Unclear | No | No | Yes | Yes | No | Yes | No | N/A | High |
| Peru | Charles Huamani [142] | No | Yes | Yes | Yes | Yes | Yes | Yes | Yes | Unclear | Moderate |
| Peru | Benoit Champigneulle [280] | Yes | No | No | Yes | Unclear | Unclear | Yes | No | Unclear | High |
| Peru | Mary F Reyes Vega [283] | Yes | Yes | Yes | Yes | Yes | No | Yes | Yes | Yes | Low |
| Peru | Carlos Alvarez-Antonio [282] | Yes | Yes | Yes | No | Yes | Yes | Yes | Yes | Yes | Low |
| Peru | Carlos Alvarez-Antonio [282] | Yes | Yes | Yes | No | Unclear | Yes | Yes | Yes | Unclear | Low |
| Peru | Enrique Guevara Rios [284] | Yes | No | Yes | No | Unclear | Unclear | Yes | No | Yes | Moderate |
| United States of America | Kristie Clarke [293] | No | No | Yes | No | Unclear | Yes | Yes | No | N/A | High |
| United States of America | Isobel Routledge [295] | No | No | Yes | Yes | No | Yes | Yes | No | N/A | High |
| United States of America | Kristie Clarke [292] | Unclear | No | Yes | No | Unclear | Yes | Yes | Yes | Unclear | High |
| United States of America | Victor M. Cardenas [153] | Yes | Yes | Yes | Yes | Unclear | Yes | Yes | No | No | Moderate |
| United States of America | Teresa Janevic [311] | Yes | No | Yes | No | Yes | Unclear | Yes | No | Unclear | Moderate |
| United States of America | Minesh Khatri [328] | Yes | No | No | No | Unclear | Yes | Yes | No | Unclear | High |
| United States of America | Ryan Kilpatrick [299] | No | No | Yes | Yes | No | Yes | Yes | No | N/A | High |
| United States of America | Alexia Couture [150] | No | No | No | No | No | Yes | Yes | Yes | N/A | High |
| United States of America | Alexia Couture [150] | No | No | Yes | No | No | Yes | Yes | Yes | N/A | Moderate |
| United States of America | Alexia Couture [150] | No | No | Yes | No | No | Yes | Yes | Yes | N/A | Moderate |
| United States of America | Alexia Couture [150] | No | No | No | No | No | Yes | Yes | Yes | N/A | High |
| United States of America | Alexia Couture [150] | No | No | No | No | No | Yes | Yes | Yes | N/A | High |
| United States of America | Alexia Couture [150] | No | No | Yes | No | No | Yes | Yes | Yes | N/A | Moderate |
| United States of America | Alexia Couture [150] | No | No | Yes | No | No | Yes | Yes | Yes | N/A | Moderate |
| United States of America | Alexia Couture [150] | No | No | Yes | No | No | Yes | Yes | Yes | N/A | Moderate |
| United States of America | Neeraj Sood [294] | Yes | Yes | Yes | Yes | Yes | Yes | Yes | Yes | No | Low |
| United States of America | Megha Mehrotra [149] | Yes | Yes | Yes | No | Yes | Yes | Yes | Yes | No | Low |
| United States of America | Megha Mehrotra [149] | Yes | Yes | Yes | Yes | Yes | Yes | Yes | Yes | No | Low |
| United States of America | Alice Pressman [154] | Yes | No | Yes | No | Yes | Yes | Yes | No | N/A | Moderate |
| United States of America | Cameron Adams [157] | Yes | No | Yes | No | Yes | Yes | Yes | Yes | Yes | Moderate |
| United States of America | Sylvia LaCourse [298] | Yes | No | Yes | No | No | Yes | Yes | No | Unclear | Moderate |
| United States of America | Marcus Wong [151] | Yes | No | Yes | No | Unclear | Yes | Yes | No | Yes | Moderate |
| United States of America | Marcus Wong [151] | Yes | No | Yes | No | Unclear | Yes | Yes | No | Yes | Moderate |
| United States of America | Marcus Wong [151] | Yes | No | Yes | No | Unclear | Yes | Yes | No | Yes | Moderate |
| United States of America | Nina Molenaar [319] | No | No | Yes | No | Yes | Unclear | Yes | No | Unclear | High |
| United States of America | Charles Gonik [317] | Yes | No | Yes | No | Unclear | Yes | Yes | No | Unclear | Moderate |
| United States of America | Kiersten Kugeler [158] | Yes | Yes | No | Yes | Yes | Yes | Yes | Yes | Unclear | Moderate |
| United States of America | Kiersten Kugeler [158] | Yes | No | Yes | Yes | Yes | Yes | Yes | Yes | Unclear | Moderate |
| United States of America | Rebecca Levorson [155] | Yes | No | Yes | Yes | Yes | Yes | Yes | No | Unclear | Moderate |
| United States of America | The University of Texas Health Science Center at Houston [148] | Yes | No | Yes | Yes | No | Yes | Yes | Yes | N/A | Moderate |
| United States of America | Roger Dodd [41] | Yes | No | Yes | Yes | Yes | Yes | Yes | No | N/A | Moderate |
| United States of America | Cara Staszewski [302] | Yes | No | No | No | No | Yes | Yes | No | N/A | High |
| United States of America | Cara Staszewski [302] | Yes | No | No | No | No | Yes | Yes | No | N/A | High |
| United States of America | McKaylee Robertson [321] | No | No | Yes | No | No | Yes | Yes | No | Yes | High |
| United States of America | Jefferson Jones [42] | No | Yes | Yes | Yes | Yes | Yes | Yes | No | N/A | Moderate |
| United States of America | Jefferson Jones [42] | No | Yes | Yes | Yes | Yes | Yes | Yes | No | N/A | Moderate |
| United States of America | Jefferson Jones [42] | No | Yes | Yes | Yes | Yes | Yes | Yes | No | N/A | Moderate |
| United States of America | Jefferson Jones [42] | No | Yes | Yes | Yes | Yes | Yes | Yes | No | N/A | Moderate |
| United States of America | Jefferson Jones [42] | No | Yes | Yes | Yes | Yes | Yes | Yes | No | N/A | Moderate |
| United States of America | Matthew Samore [163] | Yes | Yes | Yes | Yes | No | Yes | Yes | Yes | No | Low |
| United States of America | Daniel Nesbitt [333] | No | No | Yes | Yes | No | Yes | Yes | No | N/A | High |
| United States of America | Brittany K. Smith [297] | No | No | Yes | Yes | Yes | Unclear | Yes | No | N/A | High |
| United States of America | Brittany K. Smith [297] | No | No | Yes | Yes | Yes | Unclear | Yes | No | N/A | High |
| United States of America | Brittany K. Smith [297] | No | No | Yes | Yes | Yes | Unclear | Yes | No | N/A | High |
| United States of America | Roger Dodd [301] | No | No | Yes | Yes | Unclear | Yes | Yes | No | N/A | High |
| United States of America | Olga Morozova [324] | No | No | Yes | Yes | No | Yes | Yes | Yes | No | High |
| United States of America | Joshua Kennedy [152] | No | No | Yes | Yes | Yes | Yes | Yes | No | N/A | Moderate |
| United States of America | Joshua Kennedy [152] | No | No | Yes | Yes | Yes | Yes | Yes | No | N/A | Moderate |
| United States of America | Joshua Kennedy [152] | No | No | Yes | Yes | Yes | Yes | Yes | No | N/A | Moderate |
| United States of America | Patrick Sullivan [43] | Yes | Yes | Yes | Yes | Yes | Yes | Yes | Yes | No | Low |
| United States of America | Sumanth Gandra [307] | No | No | No | Yes | No | Yes | Yes | No | Unclear | High |
| United States of America | Isobel Routledge [330] | No | No | Yes | Yes | No | Yes | Yes | Yes | N/A | Moderate |
| United States of America | Glenn J. Rapsinski [323] | No | No | Yes | Yes | Unclear | Yes | Yes | No | N/A | High |
| United States of America | Glenn J. Rapsinski [323] | No | No | Yes | Yes | Unclear | Yes | Yes | No | N/A | High |
| United States of America | Lingqing Xu [296] | No | No | No | Yes | No | Yes | Yes | No | N/A | High |
| United States of America | Jannae Parrott [313] | Yes | Yes | Yes | Yes | Unclear | Yes | Yes | Yes | Unclear | Low |
| United States of America | Oliver Laeyendecker [336] | No | No | Yes | Yes | Yes | Yes | Yes | No | N/A | High |
| United States of America | Elaine Symanski [308] | Yes | Yes | Yes | Yes | Yes | Yes | Yes | No | Unclear | Low |
| United States of America | Daniel Geisler [326] | No | No | Yes | No | Unclear | Yes | Yes | No | N/A | High |
| United States of America | Robert Stout [306] | No | No | Yes | Yes | Yes | Yes | Yes | No | N/A | High |
| United States of America | Fabrizio Bonelli [305] | No | No | Yes | No | Unclear | Yes | Yes | No | N/A | High |
| United States of America | Fabrizio Bonelli [305] | No | No | Yes | No | Unclear | Yes | Yes | No | N/A | High |
| United States of America | Travis Lim [162] | No | Yes | Yes | Yes | Unclear | Unclear | Yes | Yes | Unclear | Low |
| United States of America | Travis Lim [162] | No | Yes | Yes | Yes | Unclear | Unclear | Yes | Yes | Unclear | Low |
| United States of America | Travis Lim [162] | No | Yes | Yes | Yes | Unclear | Unclear | Yes | Yes | Unclear | Low |
| United States of America | Travis Lim [162] | No | Yes | Yes | Yes | Unclear | Unclear | Yes | Yes | Unclear | Low |
| United States of America | Travis Lim [162] | No | Yes | Yes | Yes | Unclear | Unclear | Yes | Yes | Unclear | Low |
| United States of America | Travis Lim [162] | No | Yes | Yes | Yes | Unclear | Unclear | Yes | Yes | Unclear | Low |
| United States of America | Travis Lim [162] | No | Yes | Yes | Yes | Unclear | Unclear | Yes | Yes | Unclear | Low |
| United States of America | Travis Lim [162] | No | Yes | Yes | Yes | Unclear | Unclear | Yes | Yes | Unclear | Low |
| United States of America | Travis Lim [162] | No | Yes | Yes | Yes | Unclear | Unclear | Yes | Yes | Unclear | Low |
| United States of America | Callum Arnold [303] | Yes | No | No | Yes | Unclear | Unclear | Yes | No | No | High |
| United States of America | Travis Lim [162] | No | Yes | Yes | Yes | Unclear | Unclear | Yes | Yes | Unclear | Low |
| United States of America | Xiaofan Jia [310] | Yes | No | No | No | Yes | Unclear | Yes | No | Unclear | High |
| United States of America | Xiaofan Jia [310] | Yes | No | No | No | Yes | Unclear | Yes | No | Unclear | High |
| United States of America | Eran Bendavid [339] | No | No | Yes | Yes | No | Yes | Yes | Yes | Unclear | High |
| United States of America | Haley Steffen [318] | Yes | No | Yes | No | Unclear | Yes | Yes | No | N/A | Moderate |
| United States of America | Adrienne Sherman [312] | No | No | Yes | Yes | No | Yes | Yes | No | No | High |
| United States of America | Philip A Chan [165] | Yes | Yes | Yes | Yes | Yes | Yes | Yes | Yes | No | Low |
| United States of America | Elizabeth T Rogawski McQuade [159] | No | No | Yes | Yes | Yes | Yes | Yes | Yes | N/A | Moderate |
| United States of America | Tim Bruckner [314] | Yes | Yes | Yes | Yes | Yes | Unclear | Yes | Yes | Unclear | Low |
| United States of America | Brittany Smith [332] | No | No | No | Yes | Unclear | Unclear | Yes | No | Unclear | High |
| United States of America | Brittany Smith [332] | No | No | No | Yes | Unclear | Unclear | Yes | No | Unclear | High |
| United States of America | Burak Bahar [309] | No | No | No | Yes | Yes | Yes | Yes | No | N/A | High |
| United States of America | Robert Stout [300] | No | No | Yes | No | No | Yes | Yes | No | No | High |
| United States of America | Robert Stout [300] | No | No | Yes | No | No | Yes | Yes | No | No | High |
| United States of America | Jennifer Rathe [338] | Unclear | Yes | No | No | Unclear | Yes | Yes | No | Unclear | High |
| United States of America | Kristina Bajema [335] | No | No | Yes | Yes | Yes | Unclear | Yes | Yes | Unclear | High |
| United States of America | Jason Yamaki [316] | No | No | Yes | Yes | No | Yes | Yes | No | Unclear | High |
| United States of America | Daniel Stadlbauer [320] | No | No | Yes | Yes | No | Unclear | Yes | No | Unclear | High |
| United States of America | Daniel Stadlbauer [320] | No | No | Yes | Yes | Yes | Unclear | Yes | No | Unclear | High |
| United States of America | Shiwani Mahajan [160] | Yes | Yes | No | Yes | No | Yes | Yes | Yes | No | Moderate |
| United States of America | Shiwani Mahajan [160] | Yes | Yes | No | Yes | Yes | Yes | Yes | Yes | Yes | Moderate |
| United States of America | Shiwani Mahajan [160] | Yes | Yes | No | Yes | No | Yes | Yes | Yes | No | Moderate |
| United States of America | Amy K Feehan [315] | Yes | Yes | Yes | Yes | Unclear | Yes | Yes | Yes | Yes | Low |
| United States of America | Kathy Kamath [322] | No | No | Yes | Yes | Yes | Unclear | Yes | No | Unclear | High |
| United States of America | Amir Barzin [325] | No | No | Yes | No | Unclear | Yes | Yes | No | Unclear | High |
| United States of America | Shuchi Anand [44] | No | Yes | Yes | Yes | Yes | Unclear | Yes | Yes | Yes | Low |
| United States of America | Lylach Haizler Cohen [161] | Yes | No | Yes | No | Unclear | Yes | Yes | No | Unclear | Moderate |
| United States of America | Dianna Ng [341] | No | No | Yes | No | Unclear | Yes | Yes | No | Unclear | High |
| United States of America | Adam S Dingens [340] | No | No | Yes | Yes | Unclear | Yes | Yes | No | N/A | High |
| United States of America | Melissa Sutton [164] | Yes | Yes | Yes | No | Yes | Yes | Yes | No | Unclear | Moderate |
| United States of America | Amy K Feehan [329] | Yes | Yes | Yes | Yes | Unclear | Yes | Yes | Yes | Yes | Low |
| United States of America | Dustin Flannery [331] | Yes | No | Yes | No | Yes | Unclear | Yes | No | Yes | Moderate |
| United States of America | Holly Biggs [334] | Yes | Yes | Yes | No | Yes | Yes | Yes | Yes | Unclear | Low |
| United States of America | Josh Reifer [327] | Unclear | No | Yes | No | Unclear | Yes | Yes | No | Unclear | High |
| United States of America | Neeraj Sood [337] | Yes | Yes | Yes | Yes | No | Yes | Yes | Yes | No | Low |
| United States of America | Nir Menachemi [166] | Yes | Yes | Yes | Yes | No | Unclear | Yes | Yes | No | Low |
| United States of America | Megan Jehn [304] | Yes | Yes | No | Yes | Yes | Yes | Yes | Yes | No | Moderate |

###### Eastern Mediterranean region

| Country | Author | Item 1: Appropriate sample frame | Item 2: Probability sampling method | Item 3: Adequate sample size | Item 4: Subjects & setting described | Item 5: Good coverage of sample | Item 6: Sens>=90%, Spec>=97% | Item 7: Same test for all subjects | Item 8: Appropriate statistical analysis | Item 9: Adequate response rate | Overall risk of bias |
| --- | --- | --- | --- | --- | --- | --- | --- | --- | --- | --- | --- |
| Egypt | Sahar Abdelmaksoud [342] | No | Yes | No | No | No | Yes | Yes | No | N/A | High |
| Egypt | Samia Girgis [343] | No | No | Yes | Yes | Unclear | Yes | Yes | No | Unclear | High |
| Iran (Islamic Republic of) | Hamed Zandian [345] | No | Yes | Yes | No | No | Yes | Yes | No | No | Moderate |
| Iran (Islamic Republic of) | Kazem Khalagi [45] | Yes | Yes | Yes | Yes | Yes | Yes | Yes | Yes | No | Low |
| Iran (Islamic Republic of) | Seyed Abbas Mousavi [168] | Yes | Yes | Yes | Yes | Unclear | Yes | Yes | No | Unclear | Low |
| Iran (Islamic Republic of) | Maryam Shakiba [167] | Yes | Yes | No | Yes | Yes | Yes | Yes | Yes | No | Moderate |
| Iran (Islamic Republic of) | Hossein Poustchi [46] | Yes | Yes | Yes | Yes | Yes | Yes | Yes | Yes | Yes | Low |
| Iran (Islamic Republic of) | Zahra Mohammadi [344] | Unclear | Yes | No | No | No | Yes | Yes | No | Unclear | High |
| Iraq | Nawfal Hussein [346] | Yes | Yes | Yes | Yes | Yes | Yes | Yes | No | N/A | Moderate |
| Jordan | Maher Sughayer [347] | No | No | No | Yes | Unclear | Yes | Yes | No | Unclear | High |
| Jordan | Saverio Bellizzi [47] | Yes | Yes | Yes | No | Unclear | No | Yes | No | Yes | Moderate |
| Jordan | Saverio Bellizzi [47] | Yes | Yes | Yes | Yes | Unclear | No | Yes | No | Yes | Moderate |
| Jordan | Saverio Bellizzi [47] | Yes | Yes | Yes | Yes | Unclear | No | Yes | No | Yes | Moderate |
| Jordan | Maher Sughayer [348] | No | No | No | Yes | No | Yes | Yes | No | N/A | High |
| Jordan | Maher Sughayer [348] | No | No | No | No | No | Yes | Yes | No | N/A | High |
| Jordan | Maher Sughayer [348] | No | No | Yes | No | No | Yes | Yes | No | N/A | High |
| Lebanon | Abbas Hoballah [48] | Yes | Yes | Yes | Yes | Yes | Yes | Yes | Yes | Yes | Low |
| Lebanon | Dr Alissar Rady [@] | Yes | Yes | Yes | Yes | Yes | Yes | Yes | Yes | Yes | Low |
| Lebanon | Dr Alissar Rady [@] | No | Yes | Yes | Yes | Unclear | Yes | Yes | Yes | No | Moderate |
| occupied Palestinian territory - including east Jerusalem | Beesan Maraqa [169] | No | Yes | Yes | Yes | Yes | Yes | Yes | No | Unclear | Moderate |
| occupied Palestinian territory - including east Jerusalem | Nouar Qutob [349] | No | No | Yes | Yes | Yes | Yes | Yes | No | N/A | High |
| occupied Palestinian territory - including east Jerusalem | Nouar Qutob [349] | Yes | Yes | Yes | Yes | No | Yes | Yes | Yes | Yes | Low |
| occupied Palestinian territory, including east Jerusalem | Sharif E. Qaddomi [@] | Yes | Yes | Yes | Yes | Yes | Yes | Yes | No | Yes | Moderate |
| Oman | Seif Al-Abri [49] | Yes | Yes | Yes | Yes | Unclear | Yes | Yes | No | Yes | Moderate |
| Oman | Seif Al-Abri [49] | Yes | Yes | Yes | Yes | Unclear | Yes | Yes | No | Yes | Moderate |
| Oman | Seif Al-Abri [49] | Yes | Yes | Yes | Yes | Unclear | Yes | Yes | No | Yes | Moderate |
| Oman | Seif Al-Abri [49] | Yes | Yes | Yes | Yes | Unclear | Yes | Yes | No | Yes | Moderate |
| Pakistan | Ahsan Ahmad [50] | Yes | Yes | Yes | Yes | Yes | No | Yes | No | Unclear | Moderate |
| Pakistan | Mohsina Haq [51] | Yes | Yes | Yes | Yes | No | Yes | Yes | No | Unclear | Moderate |
| Pakistan | Muhammed Imran Nisar [350] | Yes | Yes | No | No | Yes | Yes | Yes | Yes | Unclear | Moderate |
| Pakistan | Muhammed Imran Nisar [350] | Yes | Yes | No | No | Yes | Yes | Yes | Yes | Unclear | Moderate |
| Pakistan | Muhammed Imran Nisar [350] | Yes | Yes | No | No | Yes | Yes | Yes | Yes | Unclear | Moderate |
| Pakistan | Muhammed Imran Nisar [350] | Yes | Yes | No | No | Yes | Yes | Yes | Yes | Unclear | Moderate |
| Pakistan | Muhammed Imran Nisar [350] | Yes | Yes | No | No | Yes | Yes | Yes | Yes | Unclear | Moderate |
| Pakistan | Muhammed Imran Nisar [350] | Yes | Yes | No | No | Yes | Yes | Yes | Yes | Unclear | Moderate |
| Pakistan | Sheeba Ansari [351] | No | No | Yes | Yes | No | Unclear | Yes | No | Unclear | High |
| Pakistan | Wajiha Javed [352] | Unclear | Yes | Yes | No | Unclear | Yes | Yes | No | Yes | High |
| Qatar | Peter Coyle [52] | Yes | No | Yes | Yes | Yes | Yes | Yes | No | N/A | Moderate |
| Qatar | Peter Coyle [53] | No | No | Yes | Yes | Yes | Yes | Yes | Yes | N/A | Moderate |
| Saudi Arabia | Sami Almudarra [354] | No | Yes | Yes | Yes | Unclear | Yes | Yes | No | Unclear | Moderate |
| Saudi Arabia | Mohammed Alosaimi [353] | No | No | No | Yes | No | Yes | Yes | No | Unclear | High |
| United Arab Emirates | May Raouf [355] | Yes | Yes | Yes | Yes | No | Yes | Yes | No | Unclear | Moderate |
| United Arab Emirates | Ahmed R Alsuwaidi [170] | Yes | Yes | Yes | Yes | Unclear | Yes | Yes | No | Unclear | Moderate |
| Yemen | Abdulla Bin-Ghouth [356] | Yes | Yes | Yes | Yes | Yes | Yes | Yes | No | Unclear | Moderate |

###### Europe region

| Country | Author | Item 1: Appropriate sample frame | Item 2: Probability sampling method | Item 3: Adequate sample size | Item 4: Subjects & setting described | Item 5: Good coverage of sample | Item 6: Sens>=90%, Spec>=97% | Item 7: Same test for all subjects | Item 8: Appropriate statistical analysis | Item 9: Adequate response rate | Overall risk of bias |
| --- | --- | --- | --- | --- | --- | --- | --- | --- | --- | --- | --- |
| Albania | Genc Sulcebe [357] | No | Yes | No | Yes | Unclear | Yes | Yes | No | Yes | High |
| Albania | Genc Sulcebe [357] | No | Yes | Yes | Yes | Unclear | Yes | Yes | No | Yes | Moderate |
| Andorra | Cristina Royo Cebrecos [59] | Yes | Yes | Yes | Yes | Yes | Yes | Yes | No | Yes | Moderate |
| Austria | Anita Siller [367] | No | No | Yes | Yes | Unclear | Yes | Yes | No | N/A | High |
| Austria | Heike Rebholz [369] | Yes | No | No | Yes | Unclear | Yes | Yes | No | Yes | High |
| Austria | Wegene Borena [371] | Yes | No | Yes | Yes | Unclear | Yes | Yes | No | Unclear | Moderate |
| Austria | Lisa Weidner [370] | No | No | Yes | Yes | Unclear | Yes | Yes | No | Unclear | High |
| Austria | M. Riesenhuber [368] | Unclear | No | Yes | No | Unclear | Yes | Yes | No | Unclear | High |
| Austria | Dennis Ladage [372] | Yes | No | Yes | No | Unclear | Yes | Yes | No | Unclear | Moderate |
| Austria | Jens Lehmann [373] | Yes | Yes | No | Yes | Unclear | Yes | Yes | No | No | High |
| Belgium | Sereina Herzog [60] | No | Yes | Yes | Yes | Yes | Yes | Yes | Yes | N/A | Low |
| Belgium | Sereina Herzog [60] | No | Yes | Yes | Yes | Yes | Yes | Yes | Yes | N/A | Low |
| Belgium | Sereina Herzog [60] | No | Yes | Yes | Yes | Yes | Yes | Yes | Yes | N/A | Low |
| Belgium | Sereina Herzog [60] | No | Yes | Yes | Yes | Yes | Yes | Yes | Yes | N/A | Low |
| Belgium | Sereina Herzog [60] | No | Yes | Yes | Yes | Yes | Yes | Yes | Yes | N/A | Low |
| Belgium | Sereina Herzog [60] | No | Yes | Yes | Yes | Yes | Yes | Yes | Yes | N/A | Low |
| Belgium | Sereina Herzog [60] | No | Yes | Yes | Yes | Yes | Yes | Yes | Yes | N/A | Low |
| Belgium | Laura Heireman [374] | No | No | No | No | Unclear | Yes | Yes | No | Unclear | High |
| Belgium | Peter van Dam [375] | No | No | No | No | Unclear | Yes | Yes | No | Unclear | High |
| Bosnia and Herzegovina | Biljana Mijovic [171] | Yes | Yes | Yes | Yes | Unclear | Yes | Yes | No | Yes | Moderate |
| Bosnia and Herzegovina | Adna Asic [358] | No | No | Yes | No | Unclear | Yes | Yes | No | Unclear | High |
| Bosnia and Herzegovina | Adna Asic [358] | No | No | Yes | No | Unclear | Yes | Yes | No | N/A | High |
| Bosnia and Herzegovina | Sanjin Musa [172] | No | Yes | Yes | Yes | No | Yes | Yes | No | N/A | Moderate |
| Bulgaria | Denitsa Tsaneva-Damyanova [359] | Yes | No | No | No | Unclear | Yes | Yes | No | Unclear | High |
| Croatia | Tatjana Vilibic-Cavlek [376] | Unclear | No | Yes | Yes | Yes | Yes | Yes | No | Unclear | High |
| Croatia | Tatjana Vilibic-Cavlek [376] | Unclear | No | Yes | Yes | Yes | Yes | Yes | No | Unclear | High |
| Croatia | Jasna Lenicek Krleza [377] | No | No | No | Yes | Unclear | Yes | Yes | No | Yes | High |
| Czechia | Pavel Piler [378] | Unclear | No | Yes | Yes | Yes | Yes | Yes | No | Unclear | High |
| Czechia | Pavel Piler [378] | Unclear | No | Yes | Yes | Yes | Yes | Yes | No | Unclear | High |
| Czechia | Pavel Piler [378] | Unclear | No | Yes | Yes | Yes | Yes | Yes | No | Unclear | High |
| Czechia | Marketa Bloomfield [379] | No | No | No | Yes | Unclear | Yes | Yes | No | Unclear | High |
| Denmark | Laura Espenhain [62] | Yes | Yes | Yes | No | Unclear | Yes | Yes | Yes | No | Low |
| Denmark | Laura Espenhain [62] | Yes | Yes | Yes | No | Unclear | Yes | Yes | Yes | No | Low |
| Denmark | Laura Espenhain [62] | Yes | Yes | Yes | No | Unclear | Yes | Yes | Yes | No | Low |
| Denmark | Laura Espenhain [62] | Yes | Yes | Yes | No | Unclear | Yes | Yes | Yes | No | Low |
| Denmark | Kamille Fogh [63] | Yes | Yes | Yes | Yes | Yes | Yes | Yes | No | No | Moderate |
| Denmark | Kamille Fogh [61] | Yes | Yes | Yes | Yes | Unclear | Yes | Yes | No | Unclear | Moderate |
| Denmark | Maren J H Rytter [174] | No | No | Yes | Yes | No | Yes | Yes | No | N/A | High |
| Denmark | Maren J H Rytter [174] | No | Yes | Yes | No | No | Yes | Yes | No | N/A | Moderate |
| Denmark | Pia Egerup [381] | Yes | No | Yes | No | Unclear | Unclear | Yes | No | Yes | Moderate |
| Denmark | Ole Birger Pedersen [64] | No | Yes | Yes | Yes | Yes | Yes | Yes | Yes | No | Low |
| Denmark | Ole Birger Pedersen [64] | No | No | Yes | No | Unclear | Yes | Yes | Yes | Unclear | High |
| Denmark | Senne Jespersen [380] | No | No | No | No | Unclear | Yes | Yes | No | N/A | High |
| Denmark | Senne Jespersen [380] | No | No | No | No | Unclear | Yes | Yes | No | N/A | High |
| Denmark | Kasper Iversen [175] | Yes | No | Yes | No | Yes | Yes | Yes | No | Yes | Moderate |
| Denmark | Christian Erikstrup [65] | No | Yes | Yes | Yes | Yes | Yes | Yes | No | Yes | Moderate |
| Estonia | Hiie Soeorg [382] | No | No | Yes | No | Unclear | Yes | Yes | No | N/A | High |
| Estonia | Piret Veerus [383] | Yes | No | No | No | Yes | Yes | Yes | No | Unclear | High |
| Faroe Islands | Maria Skaalum Petersen [176] | Yes | Yes | Yes | Yes | Yes | Yes | Yes | No | Yes | Moderate |
| Finland | Finnish institute for health and welfare [66] | Yes | Yes | No | No | Unclear | Unclear | Yes | No | Unclear | Moderate |
| Finland | Finnish institute for health and welfare [66] | Yes | Yes | No | No | Unclear | Unclear | Yes | No | Unclear | Moderate |
| Finland | Finnish institute for health and welfare [66] | Yes | Yes | No | No | Unclear | Unclear | Yes | No | Unclear | Moderate |
| Finland | Finnish institute for health and welfare [66] | Yes | Yes | No | No | Unclear | Unclear | Yes | No | Unclear | Moderate |
| Finland | Finnish institute for health and welfare [66] | Yes | Yes | No | No | Unclear | Unclear | Yes | No | Unclear | Moderate |
| Finland | Finnish institute for health and welfare [66] | Yes | Yes | No | No | Unclear | Unclear | Yes | No | Unclear | Moderate |
| Finland | Finnish institute for health and welfare [66] | Yes | Yes | No | No | Unclear | Unclear | Yes | No | Unclear | Moderate |
| Finland | Finnish institute for health and welfare [66] | Yes | Yes | No | No | Unclear | Unclear | Yes | No | Unclear | Moderate |
| Finland | Finnish institute for health and welfare [66] | Yes | Yes | No | No | Unclear | Unclear | Yes | No | Unclear | Moderate |
| Finland | Finnish institute for health and welfare [66] | Yes | Yes | No | No | Unclear | Unclear | Yes | No | Unclear | Moderate |
| Finland | Finnish institute for health and welfare [66] | Yes | Yes | No | No | Unclear | Unclear | Yes | No | Unclear | Moderate |
| Finland | Finnish institute for health and welfare [66] | Yes | Yes | No | No | Unclear | Unclear | Yes | No | Unclear | Moderate |
| Finland | Finnish institute for health and welfare [66] | Yes | Yes | No | No | Unclear | Unclear | Yes | No | Unclear | Moderate |
| Finland | Finnish institute for health and welfare [66] | Yes | Yes | No | No | Unclear | Unclear | Yes | No | Unclear | Moderate |
| Finland | Finnish institute for health and welfare [66] | Yes | Yes | No | No | Unclear | Unclear | Yes | No | Unclear | Moderate |
| Finland | Finnish institute for health and welfare [66] | Yes | Yes | No | No | Unclear | Unclear | Yes | No | Unclear | Moderate |
| Finland | Finnish institute for health and welfare [66] | Yes | Yes | No | No | Unclear | Unclear | Yes | No | Unclear | Moderate |
| Finland | Finnish institute for health and welfare [66] | Yes | Yes | No | No | Unclear | Unclear | Yes | No | Unclear | Moderate |
| Finland | Finnish institute for health and welfare [66] | Yes | Yes | No | No | Unclear | Unclear | Yes | No | Unclear | Moderate |
| Finland | Finnish institute for health and welfare [66] | Yes | Yes | No | No | Unclear | Unclear | Yes | No | Unclear | Moderate |
| Finland | Finnish institute for health and welfare [66] | Yes | Yes | No | No | Unclear | Unclear | Yes | No | Unclear | Moderate |
| Finland | Finnish institute for health and welfare [66] | Yes | Yes | No | No | Unclear | Unclear | Yes | No | Unclear | Moderate |
| Finland | Finnish institute for health and welfare [66] | Yes | Yes | No | No | Unclear | Unclear | Yes | No | Unclear | Moderate |
| Finland | Finnish institute for health and welfare [66] | Yes | Yes | No | No | Unclear | Unclear | Yes | No | Unclear | Moderate |
| Finland | Finnish institute for health and welfare [66] | Yes | Yes | No | No | Unclear | Unclear | Yes | No | Unclear | Moderate |
| Finland | Finnish institute for health and welfare [66] | Yes | Yes | No | No | Unclear | Unclear | Yes | No | Unclear | Moderate |
| Finland | Finnish institute for health and welfare [66] | Yes | Yes | No | No | Unclear | Unclear | Yes | No | Unclear | Moderate |
| Finland | Finnish institute for health and welfare [66] | Yes | Yes | No | No | Unclear | Unclear | Yes | No | Unclear | Moderate |
| Finland | Finnish institute for health and welfare [66] | Yes | Yes | No | No | Unclear | Unclear | Yes | No | Unclear | Moderate |
| Finland | Finnish institute for health and welfare [66] | Yes | Yes | No | No | Unclear | Unclear | Yes | No | Unclear | Moderate |
| Finland | Finnish institute for health and welfare [66] | Yes | Yes | No | No | Unclear | Unclear | Yes | No | Unclear | Moderate |
| Finland | Finnish institute for health and welfare [66] | Yes | Yes | No | No | Unclear | Unclear | Yes | No | Unclear | Moderate |
| Finland | Finnish institute for health and welfare [66] | Yes | Yes | No | No | Unclear | Unclear | Yes | No | Unclear | Moderate |
| Finland | Finnish institute for health and welfare [66] | Yes | Yes | No | No | Unclear | Unclear | Yes | No | Unclear | Moderate |
| Finland | Finnish institute for health and welfare [66] | Yes | Yes | No | No | Unclear | Unclear | Yes | No | Unclear | Moderate |
| Finland | Finnish institute for health and welfare [66] | Yes | Yes | No | No | Unclear | Unclear | Yes | No | Unclear | Moderate |
| Finland | Finnish institute for health and welfare [66] | Yes | Yes | No | No | Unclear | Unclear | Yes | No | Unclear | Moderate |
| Finland | Finnish institute for health and welfare [66] | Yes | Yes | No | No | Unclear | Unclear | Yes | No | Unclear | Moderate |
| Finland | Finnish institute for health and welfare [66] | Yes | Yes | No | No | Unclear | Unclear | Yes | No | Unclear | Moderate |
| Finland | Finnish institute for health and welfare [66] | Yes | Yes | No | No | Unclear | Unclear | Yes | No | Unclear | Moderate |
| Finland | Finnish institute for health and welfare [66] | Yes | Yes | No | No | Unclear | Unclear | Yes | No | Unclear | Moderate |
| Finland | Finnish institute for health and welfare [66] | Yes | Yes | No | No | Unclear | Unclear | Yes | No | Unclear | Moderate |
| Finland | Finnish institute for health and welfare [66] | Yes | Yes | No | No | Unclear | Unclear | Yes | No | Unclear | Moderate |
| Finland | Finnish institute for health and welfare [66] | Yes | Yes | No | No | Unclear | Unclear | Yes | No | Unclear | Moderate |
| Finland | Finnish institute for health and welfare [66] | Yes | Yes | No | No | Unclear | Unclear | Yes | No | Unclear | Moderate |
| Finland | Finnish institute for health and welfare [66] | Yes | Yes | No | No | Unclear | Unclear | Yes | No | Unclear | Moderate |
| Finland | Finnish institute for health and welfare [66] | Yes | Yes | No | No | Unclear | Unclear | Yes | No | Unclear | Moderate |
| Finland | Finnish institute for health and welfare [66] | Yes | Yes | No | No | Unclear | Unclear | Yes | No | Unclear | Moderate |
| Finland | Finnish institute for health and welfare [66] | Yes | Yes | No | No | Unclear | Unclear | Yes | No | Unclear | Moderate |
| Finland | Finnish institute for health and welfare [66] | Yes | Yes | No | No | Unclear | Unclear | Yes | No | Unclear | Moderate |
| Finland | Finnish institute for health and welfare [66] | Yes | Yes | No | No | Unclear | Unclear | Yes | No | Unclear | Moderate |
| Finland | Finnish institute for health and welfare [66] | Yes | Yes | No | No | Unclear | Unclear | Yes | No | Unclear | Moderate |
| Finland | Finnish institute for health and welfare [66] | Yes | Yes | No | No | Unclear | Unclear | Yes | No | Unclear | Moderate |
| Finland | Finnish institute for health and welfare [66] | Yes | Yes | No | No | Unclear | Unclear | Yes | No | Unclear | Moderate |
| Finland | Finnish institute for health and welfare [66] | Yes | Yes | No | No | Unclear | Unclear | Yes | No | Unclear | Moderate |
| Finland | Finnish institute for health and welfare [66] | Yes | Yes | No | No | Unclear | Unclear | Yes | No | Unclear | Moderate |
| Finland | Finnish institute for health and welfare [66] | Yes | Yes | No | No | Unclear | Unclear | Yes | No | Unclear | Moderate |
| Finland | Finnish institute for health and welfare [66] | Yes | Yes | No | No | Unclear | Unclear | Yes | No | Unclear | Moderate |
| Finland | Finnish institute for health and welfare [66] | Yes | Yes | No | No | Unclear | Unclear | Yes | No | Unclear | Moderate |
| Finland | Finnish institute for health and welfare [66] | Yes | Yes | No | No | Unclear | Unclear | Yes | No | Unclear | Moderate |
| Finland | Finnish institute for health and welfare [66] | Yes | Yes | No | No | Unclear | Unclear | Yes | No | Unclear | Moderate |
| Finland | Finnish institute for health and welfare [66] | Yes | Yes | No | No | Unclear | Unclear | Yes | No | Unclear | Moderate |
| Finland | Finnish institute for health and welfare [66] | Yes | Yes | No | No | Unclear | Yes | Yes | No | Unclear | High |
| Finland | Finnish institute for health and welfare [66] | Yes | Yes | No | No | Unclear | Yes | Yes | No | Unclear | High |
| Finland | Finnish institute for health and welfare [66] | Yes | Yes | No | No | Unclear | Yes | Yes | No | Unclear | High |
| Finland | Finnish institute for health and welfare [66] | Yes | Yes | No | No | Unclear | Yes | Yes | No | Unclear | High |
| Finland | Finnish institute for health and welfare [66] | Yes | Yes | No | No | Unclear | Yes | Yes | No | Unclear | High |
| Finland | Finnish institute for health and welfare [66] | Yes | Yes | No | No | Unclear | Yes | Yes | No | Unclear | High |
| Finland | Finnish institute for health and welfare [66] | Yes | Yes | No | No | Unclear | Yes | Yes | No | Unclear | High |
| Finland | Finnish institute for health and welfare [66] | Yes | Yes | No | No | Unclear | Yes | Yes | No | Unclear | High |
| Finland | Finnish institute for health and welfare [66] | Yes | Yes | No | No | Unclear | Yes | Yes | No | Unclear | High |
| Finland | Finnish institute for health and welfare [66] | Yes | Yes | No | No | Unclear | Yes | Yes | No | Unclear | High |
| Finland | Finnish institute for health and welfare [66] | Yes | Yes | No | No | Unclear | Yes | Yes | No | Unclear | High |
| Finland | Finnish institute for health and welfare [66] | Yes | Yes | No | No | Unclear | Yes | Yes | No | Unclear | High |
| Finland | Finnish institute for health and welfare [66] | Yes | Yes | No | No | Unclear | Yes | Yes | No | Unclear | High |
| Finland | Finnish institute for health and welfare [66] | Yes | Yes | No | No | Unclear | Yes | Yes | No | Unclear | High |
| Finland | Finnish institute for health and welfare [66] | Yes | Yes | No | No | Unclear | Yes | Yes | No | Unclear | High |
| Finland | Finnish institute for health and welfare [66] | Yes | Yes | No | No | Unclear | Yes | Yes | No | Unclear | High |
| Finland | Finnish institute for health and welfare [66] | Yes | Yes | No | No | Unclear | Yes | Yes | No | Unclear | High |
| Finland | Finnish institute for health and welfare [66] | Yes | Yes | No | No | Unclear | Yes | Yes | No | Unclear | High |
| Finland | Finnish institute for health and welfare [66] | Yes | Yes | No | No | Unclear | Yes | Yes | No | Unclear | High |
| Finland | Finnish institute for health and welfare [66] | Yes | Yes | No | No | Unclear | Yes | Yes | No | Unclear | High |
| Finland | Finnish institute for health and welfare [66] | Yes | Yes | No | No | Unclear | Yes | Yes | No | Unclear | High |
| Finland | Finnish institute for health and welfare [66] | Yes | Yes | No | No | Unclear | Yes | Yes | No | Unclear | High |
| Finland | Finnish institute for health and welfare [66] | Yes | Yes | Yes | No | Unclear | Yes | Yes | No | Unclear | Moderate |
| Finland | Finnish institute for health and welfare [66] | Yes | Yes | No | No | Unclear | Yes | Yes | No | Unclear | High |
| France | Anael Ayrolles [384] | No | No | No | Yes | Unclear | Yes | Yes | No | Unclear | High |
| France | Josiane Warszawski [67] | Yes | No | Yes | Yes | Yes | Yes | Yes | No | Yes | Moderate |
| France | Fabrice Carrat [385] | No | No | Yes | Yes | No | Yes | Yes | No | Unclear | High |
| France | Adeline Beaumont [387] | No | Yes | Yes | Yes | No | Yes | Yes | Yes | Unclear | Moderate |
| France | Lisandru Capai [177] | No | No | Yes | Yes | Yes | Yes | Yes | Yes | N/A | Moderate |
| France | Tom Woudenberg [388] | No | No | Yes | No | No | Yes | Yes | No | N/A | High |
| France | Anne Petit [386] | Yes | No | Yes | Yes | Unclear | Yes | Yes | No | No | Moderate |
| France | Stephane Le Vu [68] | No | Yes | Yes | Yes | Yes | Yes | Yes | Yes | N/A | Low |
| France | Stephane Le Vu [68] | No | Yes | Yes | Yes | Yes | Yes | Yes | Yes | N/A | Low |
| France | Stephane Le Vu [68] | No | Yes | Yes | Yes | Yes | Yes | Yes | Yes | N/A | Low |
| France | Vassilis Tsatsarisa [389] | Yes | No | No | No | Unclear | Yes | Yes | No | Unclear | High |
| France | Robert Cohen [392] | No | No | Yes | Yes | Unclear | Yes | Yes | No | Unclear | High |
| France | Lisandru Capai [391] | No | No | Yes | Yes | No | Yes | Yes | No | Unclear | High |
| France | Jeremie Mattern [390] | No | No | No | No | Unclear | Yes | Yes | No | Yes | High |
| Georgia | Khatuna Zakhashvili [173] | Yes | Yes | Yes | Yes | Yes | Yes | Yes | No | Yes | Moderate |
| Georgia | Khatuna Zakhashvili [173] | Yes | Yes | Yes | Yes | Yes | Yes | Yes | No | Yes | Moderate |
| Germany | Insa Backhaus [178] | Yes | Yes | Yes | Yes | Unclear | Yes | Yes | No | Unclear | Moderate |
| Germany | Felix Wachter [393] | No | No | Yes | Yes | No | Yes | Yes | No | Unclear | High |
| Germany | Felix Wachter [393] | No | No | Yes | Yes | No | Yes | Yes | No | Unclear | High |
| Germany | Felix Wachter [393] | No | No | Yes | Yes | No | Yes | Yes | No | Unclear | High |
| Germany | Bastian Fischer [394] | No | No | Yes | No | Unclear | Yes | Yes | No | Unclear | High |
| Germany | Gabor Dunay [403] | Unclear | No | Yes | Yes | Unclear | Yes | Yes | No | Yes | High |
| Germany | Enrico Richter [395] | Yes | Yes | No | No | Unclear | Yes | Yes | No | No | Moderate |
| Germany | Enrico Richter [395] | Yes | Yes | No | No | Unclear | Yes | Yes | No | No | Moderate |
| Germany | Enrico Richter [395] | Yes | Yes | No | No | Unclear | Yes | Yes | No | Yes | Moderate |
| Germany | Folke Brinkmann [398] | No | No | Yes | No | Yes | Yes | Yes | No | N/A | High |
| Germany | Felizitas Eichner [400] | Yes | No | Yes | Yes | Unclear | Yes | Yes | No | Yes | Moderate |
| Germany | Otto Laub [179] | Yes | Yes | Yes | Yes | Yes | Yes | Yes | No | N/A | Moderate |
| Germany | Katja Radon [396] | Yes | Yes | Yes | Yes | Yes | Yes | Yes | Yes | Yes | Low |
| Germany | Daniela Gornyk [70] | Yes | Yes | Yes | Yes | Yes | Yes | Yes | No | No | Moderate |
| Germany | N Ahmad Aziz [405] | No | No | Yes | No | Unclear | Yes | Yes | No | Yes | High |
| Germany | Ralf Wagner [402] | Yes | Yes | Yes | Yes | Yes | Yes | Yes | No | Yes | Moderate |
| Germany | Michael Pritsch [406] | Yes | Yes | Yes | No | Yes | Yes | Yes | Yes | Yes | Low |
| Germany | Sebastian Hausler [409] | No | No | No | No | Yes | Yes | Yes | No | N/A | High |
| Germany | Robert Koch Institut [397] | Yes | Yes | Yes | No | Unclear | Yes | Yes | Yes | Yes | Low |
| Germany | Markus Hippich [401] | No | No | Yes | Yes | Yes | Unclear | Yes | No | N/A | High |
| Germany | Robert Koch Institute [399] | Yes | Yes | Yes | No | Unclear | Yes | Yes | Yes | Yes | Low |
| Germany | Clemens Fuest [69] | Yes | No | Yes | No | Unclear | Yes | Yes | Yes | Unclear | Moderate |
| Germany | Claudia Santos Hovener [404] | Yes | Yes | Yes | Yes | Yes | Yes | Yes | No | Yes | Moderate |
| Germany | Hendrik Streeck [410] | Yes | Yes | Yes | Yes | No | Yes | Yes | No | Unclear | Moderate |
| Germany | Stefan Runkel [407] | No | No | Yes | Yes | Unclear | Yes | Yes | No | N/A | High |
| Germany | Janine Zollkau [408] | No | No | No | No | Unclear | Yes | Yes | No | Unclear | High |
| Germany | Bastian Fischer [180] | Yes | No | Yes | No | Unclear | Yes | Yes | No | Yes | Moderate |
| Greece | Michalis Koureas [71] | No | Yes | Yes | Yes | Yes | Yes | Yes | No | N/A | Moderate |
| Greece | Ourania Kotsiou [412] | Yes | No | No | Yes | No | Yes | Yes | No | No | High |
| Greece | Helena Maltezou [411] | No | No | Yes | Yes | Yes | Yes | Yes | No | Unclear | High |
| Greece | Helena Maltezou [411] | No | No | Yes | Yes | Yes | Yes | Yes | No | Unclear | High |
| Greece | Ourania S. Kotsiou [413] | Yes | No | No | Yes | Unclear | Yes | Yes | No | No | High |
| Greece | Zacharoula Bogogiannidou [414] | No | No | Yes | Yes | No | Yes | Yes | No | N/A | High |
| Hungary | Bela Merkely [73] | Yes | Yes | Yes | Yes | Yes | Yes | Yes | Yes | No | Low |
| Iceland | Daniel Gudbjartsson [415] | No | No | Yes | No | Yes | Yes | Yes | No | Unclear | High |
| Ireland | Laura Heavey [416] | Yes | Yes | Yes | Yes | Yes | Yes | Yes | No | No | Low |
| Ireland | Laura Heavey [416] | Yes | Yes | Yes | Yes | Yes | Yes | Yes | No | No | Low |
| Israel | Victoria Indenbaum [74] | Yes | No | Yes | Yes | Yes | Yes | Yes | No | Unclear | Moderate |
| Israel | Anat Ekka Zohar [419] | No | No | Yes | Yes | No | Yes | Yes | No | Unclear | High |
| Israel | Adin Breuer [417] | Yes | No | Yes | Yes | Yes | Yes | Yes | No | Unclear | Moderate |
| Israel | Yael Gozlan [418] | No | No | No | Yes | Yes | Yes | Yes | No | No | High |
| Israel | Shay Reicher [75] | Yes | No | Yes | Yes | Yes | Yes | Yes | No | Yes | Moderate |
| Italy | Francescopaolo Antonucci [420] | Yes | No | Yes | Yes | Yes | Yes | Yes | No | Unclear | Moderate |
| Italy | Federica Calo [181] | Yes | No | Yes | Yes | Yes | Yes | Yes | No | N/A | Moderate |
| Italy | Ilaria Dorigatti [426] | Yes | Yes | Yes | No | Unclear | Yes | Yes | No | Yes | Moderate |
| Italy | Roberto Melotti [422] | Yes | Yes | Yes | Yes | Yes | Yes | Yes | Yes | Yes | Low |
| Italy | Immacolata Polvere [421] | Yes | No | Yes | Yes | Yes | Yes | Yes | No | Unclear | Moderate |
| Italy | Irene Cassaniti [427] | No | No | Yes | No | Unclear | Yes | Yes | No | Unclear | High |
| Italy | Gabriele Pagani [423] | Yes | Yes | Yes | Yes | Yes | Yes | Yes | No | Yes | Moderate |
| Italy | Luca Valenti [432] | No | No | Yes | Yes | Unclear | Yes | Yes | No | Unclear | High |
| Italy | Pellegrino Cerino [424] | Yes | No | Yes | Yes | No | Yes | Yes | No | Yes | Moderate |
| Italy | Massimo Guerriero [428] | Yes | Yes | Yes | Yes | Yes | Yes | Yes | No | Yes | Moderate |
| Italy | Paola Stefanelli [182] | Yes | No | Yes | Yes | Yes | Yes | Yes | No | Yes | Moderate |
| Italy | Francesa Cito [431] | Yes | Yes | Yes | Yes | Unclear | Yes | Yes | No | Yes | Moderate |
| Italy | Jose Ramon Fiore [425] | Yes | No | Yes | Yes | Unclear | Yes | Yes | No | Unclear | Moderate |
| Italy | Instituto Nazionale di Statistica [76] | Yes | No | Yes | No | Unclear | Yes | Yes | No | Unclear | Moderate |
| Italy | Anna Cavaliere [430] | Yes | No | No | No | Unclear | Yes | Yes | No | Yes | High |
| Italy | Alessandro Baracco [429] | Yes | Yes | Yes | Yes | Yes | Yes | Yes | No | Unclear | Moderate |
| Italy | Alessandro Pancrazzi [433] | Yes | No | No | Yes | No | Yes | Yes | No | Unclear | High |
| Jersey | Statistics Jersey [77] | Yes | Yes | Yes | Yes | Yes | Yes | Yes | Yes | Yes | Low |
| Jersey | Statistics Jersey [184] | Yes | Yes | Yes | Yes | Unclear | Yes | Yes | Yes | No | Low |
| Jersey | Statistics Jersey [183] | Yes | Yes | Yes | Yes | Unclear | Yes | Yes | No | No | Low |
| Kazakhstan | Manar Smagul [361] | Yes | Yes | Yes | Yes | No | Yes | Yes | Yes | Yes | Low |
| Kazakhstan | Manar Smagul [361] | Yes | Yes | Yes | Yes | No | Yes | Yes | Yes | Yes | Low |
| Kazakhstan | Manar Smagul [361] | Yes | Yes | Yes | Yes | No | Yes | Yes | Yes | Yes | Low |
| Kazakhstan | Yuliya Semenova [360] | No | No | Yes | No | Unclear | Unclear | Yes | No | N/A | High |
| Kazakhstan | Manar Smangul [54] | Yes | Yes | Yes | Yes | Yes | Yes | Yes | No | Yes | Moderate |
| Kyrgyzstan | Anna Popova [55] | Yes | Yes | Yes | Yes | Yes | Unclear | Yes | No | Unclear | Moderate |
| Kyrgyzstan | Nurmatov Zuridin, Kuchuk Tatyana [56] | Yes | No | Yes | Yes | Yes | Yes | Yes | No | Yes | Moderate |
| Lithuania | Kastytis Smigelskas [78] | No | Yes | Yes | Yes | Unclear | Yes | Yes | No | Unclear | Moderate |
| Luxembourg | Chantal Snoeck [79] | Yes | Yes | Yes | Yes | Unclear | Yes | Yes | No | Unclear | Low |
| Netherlands | Liza Coyer [437] | No | Yes | Yes | Yes | Yes | Yes | Yes | No | No | Moderate |
| Netherlands | Demi Pagen [435] | No | No | Yes | Yes | No | Yes | Yes | No | N/A | High |
| Netherlands | Liza Coyer [434] | No | No | Yes | Yes | No | Yes | Yes | No | Unclear | High |
| Netherlands | I.L.M. Rotee [436] | No | No | No | Yes | Unclear | Yes | Yes | Yes | N/A | High |
| Netherlands | I.L.M. Rotee [436] | No | No | No | Yes | Unclear | Yes | Yes | Yes | N/A | High |
| Netherlands | Maya Keuning [438] | No | No | No | Yes | Unclear | Yes | Yes | No | Unclear | High |
| Netherlands | Ed Slot [439] | No | No | Yes | Yes | Yes | Yes | Yes | No | Yes | High |
| North Macedonia | Tatjana Makarovska Bojadjieva [362] | No | No | Yes | No | Unclear | Yes | Yes | No | N/A | High |
| Norway | Anne Eskild [440] | Yes | No | Yes | No | Unclear | Yes | Yes | No | Unclear | Moderate |
| Norway | Gro Tunheim [80] | Yes | No | Yes | Yes | Unclear | Unclear | Yes | Yes | N/A | Moderate |
| Poland | Jan Zejda [441] | Yes | Yes | Yes | Yes | No | Yes | Yes | No | No | Moderate |
| Portugal | ISN COVID-19 Group [81] | Yes | No | Yes | Yes | Unclear | Yes | Yes | No | Yes | Moderate |
| Portugal | Luisa Castro [83] | Yes | No | Yes | Yes | Yes | Unclear | Yes | Yes | Unclear | Moderate |
| Portugal | Luisa Canto e Castro [82] | Yes | Yes | Yes | Yes | Unclear | Yes | Yes | Yes | Yes | Low |
| Portugal | Irina Kislaya [84] | No | No | Yes | Yes | Yes | Yes | Yes | Yes | N/A | Moderate |
| Republic of Moldova | Alexei Ceban Stela Gheorghita, WHO CO [57] | No | Yes | Yes | Yes | No | Yes | Yes | No | Yes | Moderate |
| Romania | Tudor Olariu [443] | No | No | No | Yes | Yes | Yes | Yes | No | Unclear | High |
| Romania | Tudor Olariu [442] | No | No | Yes | Yes | Unclear | Yes | Yes | No | N/A | High |
| Romania | Tudor Rares Olariu [444] | Yes | No | Yes | Yes | Yes | Yes | Yes | No | Unclear | Moderate |
| Russian Federation | Vladimir Gorodin [363] | Unclear | No | Yes | Yes | Unclear | Yes | Yes | No | Unclear | High |
| Russian Federation | Anton Barchuk [364] | Yes | Yes | Yes | Yes | Yes | No | Yes | Yes | No | Low |
| Russian Federation | Anton Barchuk [364] | Yes | Yes | No | Yes | Yes | No | Yes | Yes | No | Moderate |
| Russian Federation | Anton Barchuk [365] | Yes | Yes | Yes | Yes | Unclear | Yes | Yes | Yes | Unclear | Low |
| Slovenia | Mario Poljak [85] | Yes | Yes | Yes | Yes | Unclear | Yes | Yes | No | No | Low |
| Slovenia | Mario Poljak [85] | Yes | Yes | Yes | Yes | Unclear | Yes | Yes | Yes | No | Low |
| Spain | Marianna Karachaliou [445] | Unclear | No | Yes | Yes | Yes | Unclear | Yes | No | No | High |
| Spain | Mayte Perez-Olmeda [86] | Yes | Yes | Yes | No | Unclear | Yes | Yes | No | Unclear | Moderate |
| Spain | Manolis Kogevinas [447] | No | No | Yes | Yes | No | Unclear | Yes | No | Yes | High |
| Spain | Marianna Karachaliou [185] | Yes | No | Yes | No | Unclear | Unclear | Yes | Yes | No | Moderate |
| Spain | Marianna Karachaliou [185] | Yes | No | No | No | No | Unclear | Yes | No | Unclear | High |
| Spain | R. Alenda [450] | No | No | Yes | No | Unclear | Yes | Yes | No | N/A | High |
| Spain | Paula Iruzubieta [186] | Yes | Yes | Yes | Yes | Yes | No | Yes | No | Yes | Moderate |
| Spain | Francesca Crovetto [449] | No | No | Yes | No | Unclear | Yes | Yes | No | N/A | High |
| Spain | Vicente Soriano [446] | No | No | Yes | No | Yes | Yes | Yes | No | Unclear | High |
| Spain | Vicente Soriano [446] | No | No | Yes | Yes | Yes | Yes | Yes | No | Unclear | High |
| Spain | Nicolas Prados [88] | Yes | No | Yes | No | No | Yes | Yes | No | Unclear | Moderate |
| Spain | Maria Vinuela [448] | Yes | No | No | No | Yes | Yes | Yes | No | Yes | High |
| Spain | Ministry of Health [87] | Yes | Yes | Yes | Yes | Yes | Yes | Yes | Yes | Yes | Low |
| Sweden | Swedish Public Health Agency [89] | Yes | No | Yes | No | Unclear | Unclear | Yes | Yes | N/A | Moderate |
| Sweden | Swedish Public Health Agency [90] | Yes | No | Yes | No | Yes | Unclear | Yes | Yes | N/A | Moderate |
| Sweden | Swedish Public Health Agency [91] | No | No | Yes | No | Unclear | Unclear | Yes | Yes | N/A | Moderate |
| Sweden | Ake Lundkvist [451] | Yes | Yes | No | Yes | Unclear | Yes | Yes | No | Unclear | High |
| Sweden | Ake Lundkvist [451] | Yes | Yes | No | Yes | Unclear | Yes | Yes | No | Unclear | High |
| Switzerland | Erin West [190] | Yes | Yes | No | Yes | Yes | Unclear | Yes | Yes | Unclear | Moderate |
| Switzerland | Ottavio Beretta [187] | Yes | Yes | Yes | No | Yes | Yes | Yes | No | No | Moderate |
| Switzerland | Ottavio Beretta [187] | Yes | Yes | Yes | No | Yes | Yes | Yes | No | No | Moderate |
| Switzerland | Ottavio Beretta [187] | Yes | Yes | Yes | No | Yes | Yes | Yes | No | Yes | Moderate |
| Switzerland | Ottavio Beretta [187] | Yes | Yes | Yes | Yes | Yes | Yes | Yes | No | Yes | Moderate |
| Switzerland | Emna El May [452] | Unclear | Yes | No | Yes | Unclear | Unclear | Yes | Yes | No | High |
| Switzerland | Aude Richard [191] | Yes | Yes | Yes | Yes | No | Yes | Yes | Yes | No | Low |
| Switzerland | Silvia Stringhini [453] | No | Yes | Yes | Yes | Yes | Yes | Yes | Yes | No | Moderate |
| Switzerland | Agne Ulyte [456] | Yes | Yes | Yes | No | Unclear | Unclear | Yes | Yes | Unclear | Low |
| Switzerland | Stephanie Baggio [457] | Yes | No | Yes | Yes | No | Yes | Yes | No | Yes | Moderate |
| Switzerland | Emna El May [188] | Yes | Yes | No | Yes | Yes | Unclear | Yes | Yes | No | Moderate |
| Switzerland | Gregorio Milani [458] | Unclear | No | No | Yes | Unclear | Yes | Yes | No | Yes | High |
| Switzerland | Elena Giuliano [454] | Yes | No | No | No | Yes | Yes | Yes | No | No | High |
| Switzerland | Silvia Stringhini [453] | Yes | Yes | Yes | Yes | Yes | Yes | Yes | Yes | N/A | Low |
| Switzerland | Daniela Anker [189] | Yes | Yes | No | Yes | Yes | Unclear | Yes | Yes | No | Moderate |
| Switzerland | Craig Fenwick [192] | Yes | Yes | No | No | No | Unclear | Yes | No | Unclear | Moderate |
| Switzerland | Marc Emmenegger [455] | No | No | Yes | No | Unclear | Unclear | Yes | No | Unclear | High |
| Switzerland | Marc Emmenegger [455] | No | No | Yes | No | Unclear | Unclear | Yes | No | Unclear | High |
| Switzerland | Marc Emmenegger [455] | No | No | Yes | No | Unclear | Unclear | Yes | No | Unclear | High |
| Switzerland | Marc Emmenegger [455] | No | No | Yes | No | Unclear | Unclear | Yes | No | Unclear | High |
| Switzerland | Marc Emmenegger [455] | No | No | Yes | No | Unclear | Unclear | Yes | No | Unclear | High |
| Switzerland | Silvia Stringhini [193] | Yes | Yes | Yes | Yes | Yes | Yes | Yes | No | No | Moderate |
| Turkey | Ozge Yucel Celik [366] | Yes | No | No | No | Unclear | Yes | Yes | No | Unclear | High |
| Turkey | Ozge Yucel Celik [366] | Yes | No | No | No | Unclear | Yes | Yes | No | Unclear | High |
| United Kingdom of Great Britain and Northern Ireland | Public Health Scotland [92] | No | Yes | Yes | Yes | Yes | Yes | Yes | Yes | N/A | Low |
| United Kingdom of Great Britain and Northern Ireland | Public Health Scotland [92] | No | No | Yes | No | Yes | Yes | Yes | Yes | N/A | Moderate |
| United Kingdom of Great Britain and Northern Ireland | Public Health Scotland [92] | No | No | Yes | Yes | Yes | Yes | Yes | Yes | N/A | Moderate |
| United Kingdom of Great Britain and Northern Ireland | Sarah Beale [460] | No | No | Yes | Yes | Unclear | Yes | Yes | No | N/A | High |
| United Kingdom of Great Britain and Northern Ireland | Heather Whitaker [108] | Yes | No | Yes | No | Unclear | Yes | Yes | No | N/A | Moderate |
| United Kingdom of Great Britain and Northern Ireland | Heather Whitaker [108] | Yes | No | Yes | No | Unclear | Yes | Yes | No | N/A | Moderate |
| United Kingdom of Great Britain and Northern Ireland | Heather Whitaker [108] | Yes | No | Yes | No | Unclear | Yes | Yes | No | N/A | Moderate |
| United Kingdom of Great Britain and Northern Ireland | Heather Whitaker [108] | Yes | No | Yes | No | Unclear | Yes | Yes | No | N/A | Moderate |
| United Kingdom of Great Britain and Northern Ireland | Public Health Scotland [93] | No | Yes | Yes | Yes | Yes | Yes | Yes | Yes | N/A | Low |
| United Kingdom of Great Britain and Northern Ireland | Public Health Scotland [93] | No | No | Yes | No | Yes | Yes | Yes | Yes | N/A | Moderate |
| United Kingdom of Great Britain and Northern Ireland | Public Health Scotland [93] | No | No | Yes | Yes | Yes | Yes | Yes | Yes | N/A | Moderate |
| United Kingdom of Great Britain and Northern Ireland | Public Health Scotland [94] | No | Yes | Yes | Yes | Yes | Yes | Yes | Yes | N/A | Low |
| United Kingdom of Great Britain and Northern Ireland | Public Health Scotland [94] | No | No | Yes | No | Yes | Yes | Yes | No | N/A | High |
| United Kingdom of Great Britain and Northern Ireland | Public Health Scotland [94] | No | No | Yes | Yes | Yes | Yes | Yes | Yes | N/A | Moderate |
| United Kingdom of Great Britain and Northern Ireland | Public Health Scotland [95] | No | Yes | Yes | Yes | Yes | Yes | Yes | Yes | N/A | Low |
| United Kingdom of Great Britain and Northern Ireland | Public Health Scotland [95] | No | No | Yes | No | Yes | Yes | Yes | No | N/A | High |
| United Kingdom of Great Britain and Northern Ireland | Public Health Scotland [95] | No | No | Yes | Yes | Yes | Yes | Yes | Yes | N/A | Moderate |
| United Kingdom of Great Britain and Northern Ireland | Public Health Scotland [96] | No | Yes | Yes | Yes | Yes | Yes | Yes | Yes | N/A | Low |
| United Kingdom of Great Britain and Northern Ireland | Public Health Scotland [96] | No | No | Yes | No | Yes | Yes | Yes | No | N/A | High |
| United Kingdom of Great Britain and Northern Ireland | Public Health Scotland [96] | No | No | Yes | Yes | Yes | Yes | Yes | Yes | N/A | Moderate |
| United Kingdom of Great Britain and Northern Ireland | Clarissa Oeser [97] | No | No | Yes | No | Unclear | Yes | Yes | Yes | N/A | Moderate |
| United Kingdom of Great Britain and Northern Ireland | Clarissa Oeser [97] | No | No | Yes | No | Unclear | Yes | Yes | Yes | N/A | Moderate |
| United Kingdom of Great Britain and Northern Ireland | Clarissa Oeser [97] | No | No | Yes | No | Unclear | Yes | Yes | Yes | N/A | Moderate |
| United Kingdom of Great Britain and Northern Ireland | Clarissa Oeser [97] | No | No | Yes | No | Unclear | Yes | Yes | Yes | N/A | Moderate |
| United Kingdom of Great Britain and Northern Ireland | Clarissa Oeser [97] | No | No | Yes | No | Unclear | Yes | Yes | Yes | N/A | Moderate |
| United Kingdom of Great Britain and Northern Ireland | Clarissa Oeser [97] | No | No | No | No | Unclear | Yes | Yes | Yes | N/A | High |
| United Kingdom of Great Britain and Northern Ireland | Clarissa Oeser [97] | No | No | Yes | No | Unclear | Yes | Yes | Yes | N/A | Moderate |
| United Kingdom of Great Britain and Northern Ireland | Public Health Scotland [98] | No | Yes | Yes | Yes | Yes | Yes | Yes | Yes | N/A | Low |
| United Kingdom of Great Britain and Northern Ireland | Public Health Scotland [98] | No | No | Yes | No | Yes | Yes | Yes | Yes | N/A | Moderate |
| United Kingdom of Great Britain and Northern Ireland | Public Health Scotland [98] | No | No | Yes | Yes | Yes | Yes | Yes | Yes | N/A | Moderate |
| United Kingdom of Great Britain and Northern Ireland | Daniel Major-Smith [459] | No | No | Yes | No | No | Yes | Yes | No | Yes | High |
| United Kingdom of Great Britain and Northern Ireland | Daniel Major-Smith [459] | No | No | Yes | No | No | Yes | Yes | No | Yes | High |
| United Kingdom of Great Britain and Northern Ireland | Kate Glennon [466] | Yes | No | No | No | Unclear | Yes | Yes | No | Unclear | High |
| United Kingdom of Great Britain and Northern Ireland | Public Health Scotland [99] | Yes | Yes | Yes | Yes | Unclear | Yes | Yes | Yes | N/A | Low |
| United Kingdom of Great Britain and Northern Ireland | Public Health Scotland [99] | Yes | No | Yes | No | Yes | Yes | Yes | Yes | N/A | Moderate |
| United Kingdom of Great Britain and Northern Ireland | Public Health Scotland [99] | No | No | Yes | Yes | Unclear | Yes | Yes | Yes | N/A | Moderate |
| United Kingdom of Great Britain and Northern Ireland | Public Health Scotland [100] | No | Yes | Yes | Yes | Yes | Yes | Yes | Yes | N/A | Low |
| United Kingdom of Great Britain and Northern Ireland | Public Health Scotland [100] | Yes | No | Yes | No | Yes | Yes | Yes | Yes | N/A | Moderate |
| United Kingdom of Great Britain and Northern Ireland | Public Health Scotland [100] | No | No | Yes | Yes | Yes | Yes | Yes | Yes | N/A | Moderate |
| United Kingdom of Great Britain and Northern Ireland | Katherine Gaskell [461] | No | Yes | Yes | Yes | Yes | Unclear | Yes | No | Yes | Moderate |
| United Kingdom of Great Britain and Northern Ireland | Paul William Bird [468] | Yes | Yes | Yes | Yes | Unclear | Yes | Yes | No | Unclear | Moderate |
| United Kingdom of Great Britain and Northern Ireland | Public Health Scotland [101] | No | Yes | Yes | Yes | Yes | Yes | Yes | Yes | N/A | Low |
| United Kingdom of Great Britain and Northern Ireland | Public Health Scotland [101] | No | No | Yes | No | Yes | Yes | Yes | Yes | N/A | Moderate |
| United Kingdom of Great Britain and Northern Ireland | Public Health Scotland [101] | No | No | Yes | Yes | Yes | Yes | Yes | Yes | N/A | Moderate |
| United Kingdom of Great Britain and Northern Ireland | Hani Abo-Leyah [463] | No | No | No | No | Yes | Yes | Yes | No | N/A | High |
| United Kingdom of Great Britain and Northern Ireland | Gayatri Amirthalingam [467] | No | No | Yes | Yes | Unclear | Yes | Yes | Yes | N/A | Moderate |
| United Kingdom of Great Britain and Northern Ireland | Robert W Aldridge [103] | Yes | No | Yes | Yes | Yes | Yes | Yes | No | Unclear | Moderate |
| United Kingdom of Great Britain and Northern Ireland | Public Health Scotland [102] | No | Yes | Yes | Yes | Yes | Yes | Yes | Yes | N/A | Low |
| United Kingdom of Great Britain and Northern Ireland | Public Health Scotland [102] | No | No | Yes | No | Yes | Yes | Yes | Yes | N/A | Moderate |
| United Kingdom of Great Britain and Northern Ireland | Public Health Scotland [102] | No | No | Yes | Yes | Yes | Yes | Yes | Yes | N/A | Moderate |
| United Kingdom of Great Britain and Northern Ireland | Heather J Whitaker [104] | Yes | No | Yes | No | Unclear | Yes | Yes | Yes | Yes | Moderate |
| United Kingdom of Great Britain and Northern Ireland | Jane Oakey [462] | No | No | No | Yes | Yes | Yes | Yes | No | N/A | High |
| United Kingdom of Great Britain and Northern Ireland | Public Health Scotland [105] | Yes | No | Yes | No | Yes | Yes | Yes | Yes | N/A | Moderate |
| United Kingdom of Great Britain and Northern Ireland | Public Health Scotland [105] | No | No | Yes | Yes | Yes | Yes | Yes | Yes | N/A | Moderate |
| United Kingdom of Great Britain and Northern Ireland | Office for National Statistics [194] | Yes | Yes | Yes | Yes | Unclear | Unclear | Yes | Yes | Unclear | Low |
| United Kingdom of Great Britain and Northern Ireland | Office for National Statistics [194] | Yes | Yes | Yes | Yes | Unclear | Unclear | Yes | Yes | Unclear | Low |
| United Kingdom of Great Britain and Northern Ireland | Office for National Statistics [194] | Yes | Yes | Yes | Yes | Unclear | Unclear | Yes | Yes | Unclear | Low |
| United Kingdom of Great Britain and Northern Ireland | Office for National Statistics [194] | Yes | Yes | Yes | Yes | Unclear | Unclear | Yes | Yes | Unclear | Low |
| United Kingdom of Great Britain and Northern Ireland | Public Health Scotland [106] | Yes | No | Yes | No | Yes | Yes | Yes | Yes | N/A | Moderate |
| United Kingdom of Great Britain and Northern Ireland | Public Health Scotland [106] | No | No | Yes | Yes | Yes | Yes | Yes | Yes | N/A | Moderate |
| United Kingdom of Great Britain and Northern Ireland | Public Health Scotland [107] | Yes | No | Yes | No | Yes | Yes | Yes | Yes | N/A | Moderate |
| United Kingdom of Great Britain and Northern Ireland | Public Health Scotland [107] | No | No | Yes | Yes | Yes | Yes | Yes | Yes | N/A | Moderate |
| United Kingdom of Great Britain and Northern Ireland | Office for National Statistics [195] | Yes | Yes | Yes | No | Unclear | Unclear | Yes | No | Unclear | Moderate |
| United Kingdom of Great Britain and Northern Ireland | Office for National Statistics [195] | Yes | Yes | Yes | No | Unclear | Unclear | Yes | No | Unclear | Moderate |
| United Kingdom of Great Britain and Northern Ireland | Office for National Statistics [195] | Yes | Yes | No | No | Unclear | Unclear | Yes | No | Unclear | Moderate |
| United Kingdom of Great Britain and Northern Ireland | Office for National Statistics [195] | Yes | Yes | Yes | No | Unclear | Unclear | Yes | No | Unclear | Moderate |
| United Kingdom of Great Britain and Northern Ireland | Shamez Ladhani [109] | Yes | Yes | Yes | No | Yes | Yes | Yes | No | Unclear | Moderate |
| United Kingdom of Great Britain and Northern Ireland | Office for National Statistics [195] | Yes | Yes | Yes | No | Unclear | Unclear | Yes | No | Unclear | Moderate |
| United Kingdom of Great Britain and Northern Ireland | Office for National Statistics [195] | Yes | Yes | Yes | No | Unclear | Unclear | Yes | No | Unclear | Moderate |
| United Kingdom of Great Britain and Northern Ireland | Office for National Statistics [195] | Yes | Yes | No | No | Unclear | Unclear | Yes | No | Unclear | Moderate |
| United Kingdom of Great Britain and Northern Ireland | Office for National Statistics [195] | Yes | Yes | Yes | No | Unclear | Unclear | Yes | No | Unclear | Moderate |
| United Kingdom of Great Britain and Northern Ireland | Ellen C Hughes [469] | Yes | Yes | Yes | Yes | Yes | Unclear | Yes | Yes | N/A | Low |
| United Kingdom of Great Britain and Northern Ireland | Public Health Scotland [110] | No | No | Yes | Yes | Yes | Yes | Yes | Yes | N/A | Moderate |
| United Kingdom of Great Britain and Northern Ireland | Office for National Statistics [195] | Yes | Yes | Yes | No | Unclear | Unclear | Yes | No | Unclear | Moderate |
| United Kingdom of Great Britain and Northern Ireland | Office for National Statistics [195] | Yes | Yes | No | No | Unclear | Unclear | Yes | No | Unclear | Moderate |
| United Kingdom of Great Britain and Northern Ireland | Office for National Statistics [195] | Yes | Yes | No | No | Unclear | Unclear | Yes | No | Unclear | Moderate |
| United Kingdom of Great Britain and Northern Ireland | Office for National Statistics [195] | Yes | Yes | No | No | Unclear | Unclear | Yes | No | Unclear | Moderate |
| United Kingdom of Great Britain and Northern Ireland | Office for National Statistics [195] | Yes | Yes | No | No | Unclear | Unclear | Yes | No | Unclear | Moderate |
| United Kingdom of Great Britain and Northern Ireland | Office for National Statistics [195] | Yes | Yes | No | No | Unclear | Unclear | Yes | No | Unclear | Moderate |
| United Kingdom of Great Britain and Northern Ireland | Office for National Statistics [195] | Yes | Yes | No | No | Unclear | Unclear | Yes | No | Unclear | Moderate |
| United Kingdom of Great Britain and Northern Ireland | Office for National Statistics [195] | Yes | Yes | Yes | No | Unclear | Unclear | Yes | No | Unclear | Moderate |
| United Kingdom of Great Britain and Northern Ireland | Craig P Thompson [196] | No | Yes | Yes | Yes | Yes | Unclear | Yes | No | N/A | Moderate |
| United Kingdom of Great Britain and Northern Ireland | Sheila F Lumley [465] | Yes | No | Yes | No | Unclear | Unclear | Yes | No | Yes | Moderate |
| United Kingdom of Great Britain and Northern Ireland | Philippa M Wells [464] | No | No | No | Yes | Unclear | Unclear | Yes | No | No | High |
| United Kingdom of Great Britain and Northern Ireland | Public Health England [111] | No | Yes | Yes | No | Yes | Yes | Yes | Yes | Unclear | Low |
| United Kingdom of Great Britain and Northern Ireland | Public Health England [111] | No | Yes | Yes | No | Yes | Yes | Yes | Yes | Unclear | Low |
| United Kingdom of Great Britain and Northern Ireland | Public Health England [111] | No | Yes | Yes | No | Yes | Yes | Yes | Yes | Unclear | Low |
| United Kingdom of Great Britain and Northern Ireland | Office for National Statistics [195] | Yes | Yes | Yes | No | Unclear | Unclear | Yes | No | Unclear | Moderate |
| United Kingdom of Great Britain and Northern Ireland | Office for National Statistics [195] | Yes | Yes | No | No | Unclear | Unclear | Yes | No | Unclear | Moderate |
| United Kingdom of Great Britain and Northern Ireland | Public Health England [111] | Yes | Yes | No | No | Yes | Yes | Yes | Yes | Yes | Moderate |
| United Kingdom of Great Britain and Northern Ireland | Public Health England [111] | No | Yes | Yes | No | Yes | Yes | Yes | No | Unclear | Moderate |
| United Kingdom of Great Britain and Northern Ireland | Public Health England [111] | No | Yes | Yes | No | Yes | Yes | Yes | Yes | Unclear | Low |
| United Kingdom of Great Britain and Northern Ireland | Public Health England [111] | No | Yes | Yes | No | Yes | Yes | Yes | Yes | Unclear | Low |
| United Kingdom of Great Britain and Northern Ireland | Public Health England [111] | No | Yes | Yes | No | Yes | Yes | Yes | Yes | Unclear | Low |
| United Kingdom of Great Britain and Northern Ireland | Public Health England [111] | No | Yes | Yes | No | Yes | Yes | Yes | Yes | Unclear | Low |
| United Kingdom of Great Britain and Northern Ireland | Public Health England [111] | No | Yes | Yes | No | Yes | Yes | Yes | Yes | Unclear | Low |
| United Kingdom of Great Britain and Northern Ireland | Public Health England [111] | No | Yes | Yes | No | Yes | Yes | Yes | Yes | Unclear | Low |
| United Kingdom of Great Britain and Northern Ireland | Public Health England [111] | No | Yes | Yes | No | Yes | Yes | Yes | Yes | Unclear | Low |
| United Kingdom of Great Britain and Northern Ireland | Public Health England [111] | No | Yes | Yes | No | Yes | Yes | Yes | Yes | Unclear | Low |
| United Kingdom of Great Britain and Northern Ireland | Office for National Statistics [195] | Yes | Yes | Yes | No | Unclear | Unclear | Yes | No | Unclear | Moderate |
| United Kingdom of Great Britain and Northern Ireland | Office for National Statistics [195] | Yes | Yes | Yes | No | Unclear | Unclear | Yes | No | Unclear | Moderate |
| United Kingdom of Great Britain and Northern Ireland | Public Health England [111] | Yes | Yes | No | No | Yes | Yes | Yes | No | Unclear | Moderate |
| United Kingdom of Great Britain and Northern Ireland | Public Health England [111] | Yes | Yes | No | No | Yes | Yes | Yes | No | Unclear | Moderate |
| United Kingdom of Great Britain and Northern Ireland | Public Health England [111] | No | Yes | Yes | No | Yes | Yes | Yes | No | Unclear | Moderate |
| United Kingdom of Great Britain and Northern Ireland | Public Health England [111] | No | Yes | No | No | Yes | Yes | Yes | No | Unclear | High |
| United Kingdom of Great Britain and Northern Ireland | Public Health England [111] | No | Yes | Yes | No | Yes | Yes | Yes | Yes | Unclear | Low |
| United Kingdom of Great Britain and Northern Ireland | Public Health England [111] | No | Yes | Yes | No | Yes | Yes | Yes | Yes | Unclear | Low |
| United Kingdom of Great Britain and Northern Ireland | Office for National Statistics [195] | Yes | Yes | Yes | No | Unclear | Unclear | Yes | No | Unclear | Moderate |
| United Kingdom of Great Britain and Northern Ireland | Public Health England [111] | No | Yes | Yes | No | Yes | Yes | Yes | Yes | Unclear | Low |
| United Kingdom of Great Britain and Northern Ireland | Public Health England [111] | No | Yes | Yes | No | Yes | Yes | Yes | Yes | Unclear | Low |
| United Kingdom of Great Britain and Northern Ireland | Office for National Statistics [195] | Yes | Yes | Yes | No | Unclear | Unclear | Yes | Yes | Unclear | Low |
| Uzbekistan | R A Rakhimov [58] | Yes | No | Yes | No | Unclear | Yes | Yes | No | Unclear | Moderate |

###### South-East Asia region

| Country | Author | Item 1: Appropriate sample frame | Item 2: Probability sampling method | Item 3: Adequate sample size | Item 4: Subjects & setting described | Item 5: Good coverage of sample | Item 6: Sens>=90%, Spec>=97% | Item 7: Same test for all subjects | Item 8: Appropriate statistical analysis | Item 9: Adequate response rate | Overall risk of bias |
| --- | --- | --- | --- | --- | --- | --- | --- | --- | --- | --- | --- |
| Bangladesh | Zabed Ahmed [471] | Yes | No | No | Yes | Yes | Yes | Yes | No | Unclear | High |
| Bangladesh | Taufiqur Bhuiyan [470] | Yes | Yes | Yes | Yes | Yes | Yes | Yes | Yes | Unclear | Low |
| Bangladesh | Prof. Dr. Tahmina Shirin [472] | No | Yes | Yes | Yes | Unclear | Yes | Yes | Yes | No | Moderate |
| India | Alpesh Goyal [481] | Unclear | No | No | Yes | No | Yes | Yes | No | Unclear | High |
| India | Pujarini Dash [474] | Yes | Yes | No | No | Yes | Yes | Yes | Yes | Yes | Moderate |
| India | Pujarini Dash [474] | Yes | Yes | No | No | Yes | Yes | Yes | Yes | Yes | Moderate |
| India | Pujarini Dash [474] | Yes | Yes | No | No | Yes | Yes | Yes | Yes | Yes | Moderate |
| India | Pujarini Dash [474] | Yes | Yes | No | No | Yes | Yes | Yes | Yes | Yes | Moderate |
| India | Om Prakash [476] | Yes | No | Yes | Yes | Unclear | Yes | Yes | No | Yes | Moderate |
| India | Matt Hitchings [475] | Yes | Yes | Yes | Yes | Unclear | Yes | Yes | No | Unclear | Moderate |
| India | Deepak Kumar [478] | Yes | Yes | Yes | Yes | Yes | Yes | Yes | No | Yes | Moderate |
| India | Pragya Sharma [197] | Yes | Yes | Yes | Yes | Yes | Yes | Yes | Yes | Unclear | Low |
| India | Divya Dayanand [473] | Yes | Yes | Yes | Yes | Yes | Yes | Yes | Yes | Yes | Low |
| India | Divya Dayanand [473] | Yes | Yes | Yes | Yes | Yes | Yes | Yes | Yes | Yes | Low |
| India | Sindhulina Chandrasingh [497] | No | No | No | No | Unclear | Yes | Yes | No | N/A | High |
| India | Vasanthakumar Namasivayam [204] | Yes | Yes | Yes | Yes | Yes | Yes | Yes | Yes | Unclear | Low |
| India | Smita Mahapatra [480] | No | No | Yes | Yes | No | Yes | Yes | No | N/A | High |
| India | Government of Kerala [198] | Yes | Yes | Yes | Yes | Unclear | Yes | Yes | No | Unclear | Moderate |
| India | Government of Kerala [198] | Yes | Yes | Yes | Yes | Unclear | Yes | Yes | No | Unclear | Moderate |
| India | Government of Kerala [198] | Yes | Yes | Yes | Yes | Unclear | Yes | Yes | No | Unclear | Moderate |
| India | Government of Kerala [198] | Yes | Yes | Yes | Yes | Unclear | Yes | Yes | No | Unclear | Moderate |
| India | Government of Kerala [198] | Yes | Yes | Yes | Yes | Unclear | Yes | Yes | No | Unclear | Moderate |
| India | Government of Kerala [198] | Yes | No | Yes | No | Unclear | Yes | Yes | No | Unclear | Moderate |
| India | Muhammad Khan [201] | Yes | Yes | Yes | Yes | Yes | Yes | Yes | Yes | Yes | Low |
| India | Amitav Banerjee [485] | Yes | Yes | Yes | No | Unclear | Yes | Yes | No | Yes | Moderate |
| India | Viswanath Billa [498] | Yes | No | No | No | Unclear | Yes | Yes | No | Unclear | High |
| India | Muthusamy Kumar [486] | Yes | Yes | Yes | Yes | Yes | Yes | Yes | Yes | Yes | Low |
| India | Nandini Sharma [200] | Yes | Yes | Yes | Yes | Yes | Yes | Yes | Yes | Yes | Low |
| India | Nandini Sharma [202] | Yes | Yes | Yes | Yes | Yes | Yes | Yes | Yes | N/A | Low |
| India | Nandini Sharma [202] | Yes | Yes | Yes | Yes | Yes | Yes | Yes | Yes | N/A | Low |
| India | Nandini Sharma [202] | Yes | Yes | Yes | Yes | Yes | Unclear | Yes | Yes | N/A | Low |
| India | Avula Laxmaiah [477] | Yes | Yes | Yes | Yes | Unclear | Yes | Yes | No | Unclear | Moderate |
| India | Om Prakash [488] | Yes | Yes | Yes | Yes | Unclear | Yes | Yes | No | Unclear | Moderate |
| India | Manoj V Murhekar [112] | Yes | Yes | Yes | Yes | Yes | Yes | Yes | Yes | Yes | Low |
| India | Jaya Singh Kshatri [491] | Yes | Yes | Yes | Yes | Unclear | Yes | Yes | Yes | Yes | Low |
| India | Jaya Singh Kshatri [491] | Yes | Yes | Yes | Yes | Unclear | Yes | Yes | Yes | Yes | Low |
| India | Jaya Singh Kshatri [491] | Yes | Yes | Yes | Yes | Unclear | Yes | Yes | Yes | Yes | Low |
| India | Om Prakash [483] | Yes | No | Yes | Yes | Yes | Yes | Yes | No | Unclear | Moderate |
| India | Jaya Singh Kshatri [487] | Yes | Yes | Yes | Yes | Yes | Yes | Yes | Yes | Yes | Low |
| India | Pallavi Saple [492] | Yes | No | No | Yes | Unclear | Unclear | Yes | No | Unclear | High |
| India | Salil Sakalle [490] | Yes | Yes | Yes | Yes | Unclear | Yes | Yes | Yes | Unclear | Low |
| India | Taha Ayub [484] | Yes | Yes | Yes | Yes | Yes | Yes | Yes | No | Yes | Moderate |
| India | Rita Isaac [482] | Yes | Yes | No | Yes | Yes | Yes | Yes | No | N/A | Moderate |
| India | Rita Isaac [482] | Yes | Yes | No | Yes | Yes | Yes | Yes | No | N/A | Moderate |
| India | Hem Chandra Pandey [496] | No | No | Yes | Yes | No | Yes | Yes | No | N/A | High |
| India | Sudarshan Ramaswamy [479] | Yes | Yes | Yes | Yes | Yes | Yes | Yes | Yes | N/A | Low |
| India | Manoj V Murhekar [112] | Yes | Yes | Yes | Yes | Yes | Yes | Yes | Yes | Yes | Low |
| India | Sriram Selvaraju [493] | Yes | Yes | Yes | Yes | Yes | Yes | Yes | Yes | Yes | Low |
| India | Sitanshu Sekhar Kar [203] | Yes | Yes | Yes | Yes | Yes | Yes | Yes | No | Yes | Moderate |
| India | Sitanshu Sekhar Kar [203] | Yes | Yes | Yes | Yes | Yes | Yes | Yes | No | Yes | Moderate |
| India | Sitanshu Sekhar Kar [203] | Yes | Yes | Yes | Yes | Yes | Yes | Yes | No | Yes | Moderate |
| India | Om Prakash [495] | Yes | No | Yes | Yes | Yes | Yes | Yes | No | Unclear | Moderate |
| India | Anup Malani [494] | Yes | Yes | Yes | Yes | Yes | Yes | Yes | Yes | Unclear | Low |
| India | Anup Malani [494] | Yes | Yes | Yes | Yes | Yes | Yes | Yes | Yes | Unclear | Low |
| India | S Muhammad Salim Khan [206] | No | No | Yes | Yes | Yes | Yes | Yes | Yes | Yes | Moderate |
| India | Parthasarathi Satpati [205] | Yes | Yes | No | Yes | Yes | Yes | Yes | No | Unclear | Moderate |
| India | Ullas Kolthur Seetharam [489] | Yes | Yes | Yes | Yes | Yes | Yes | Yes | Yes | Yes | Low |
| India | Ullas Kolthur Seetharam [489] | Yes | Yes | Yes | Yes | Yes | Yes | Yes | Yes | Yes | Low |
| India | Ministry of Health and Family Welfare [207] | Yes | Yes | Yes | No | No | Yes | Yes | No | Unclear | Moderate |
| India | Dr Puneet Misra [199] | Yes | Yes | Yes | Yes | Unclear | Yes | Yes | No | No | Moderate |
| Indonesia | Ni Luh Ayu Megasari [208] | Yes | No | Yes | Yes | Unclear | No | Yes | No | Unclear | Moderate |
| Indonesia | Dr. dr. Tri Yunis Miko Wahyono, M.Sc [113] | Yes | Yes | Yes | Yes | Yes | Yes | Yes | No | Yes | Moderate |
| Maldives | Raheema Abdul-Raheem [499] | Yes | Yes | Yes | Yes | Unclear | Yes | Yes | No | Yes | Moderate |
| Nepal | Government of Nepal [114] | Yes | Yes | Yes | Yes | Yes | Yes | Yes | Yes | Yes | Low |
| Thailand | Sutthichai Nakphook, MD [115] | Yes | Yes | Yes | Yes | Yes | Yes | Yes | No | Yes | Moderate |

###### Western Pacific region

| Country | Author | Item 1: Appropriate sample frame | Item 2: Probability sampling method | Item 3: Adequate sample size | Item 4: Subjects & setting described | Item 5: Good coverage of sample | Item 6: Sens>=90%, Spec>=97% | Item 7: Same test for all subjects | Item 8: Appropriate statistical analysis | Item 9: Adequate response rate | Overall risk of bias |
| --- | --- | --- | --- | --- | --- | --- | --- | --- | --- | --- | --- |
| Australia | Dorothy Machalek [500] | No | Yes | Yes | Yes | Yes | Yes | Yes | Yes | N/A | Low |
| Australia | Kaitlyn Vette [116] | Yes | No | Yes | No | Yes | Yes | Yes | No | Unclear | Moderate |
| Australia | Heather F Gidding [502] | No | Yes | Yes | Yes | Yes | Unclear | Yes | No | N/A | Moderate |
| Australia | Heather F Gidding [502] | Yes | Yes | No | No | Yes | Unclear | Yes | No | N/A | Moderate |
| Australia | Nicholas Coatsworth [117] | Yes | No | Yes | No | Yes | Unclear | Yes | No | Yes | Moderate |
| Australia | Dorothy Machalek [501] | No | No | Yes | Yes | Unclear | Yes | Yes | No | N/A | High |
| Australia | Kristine Macartney, Nicholas Wood, Archana Koirala [@] | No | No | Yes | Yes | Unclear | Yes | Yes | No | N/A | High |
| China | Lin-Lei Chen [209] | Yes | No | Yes | No | No | Yes | Yes | No | N/A | Moderate |
| China | Zhenyu He [210] | Yes | Yes | Yes | Yes | Yes | Yes | Yes | Yes | Yes | Low |
| China | Zhongjie Li [118] | Yes | Yes | Yes | Yes | Yes | Yes | Yes | No | Yes | Moderate |
| China | Zhen Zhang [505] | No | No | No | No | Unclear | Yes | Yes | No | Unclear | High |
| China | Zhen Zhang [505] | No | No | No | No | Unclear | Yes | Yes | No | Unclear | High |
| China | Lu He [504] | Yes | No | Yes | Yes | Unclear | Yes | Yes | No | Unclear | Moderate |
| China | Hsiang-Ling Ho [503] | No | No | Yes | Yes | Unclear | Yes | Yes | No | N/A | Moderate |
| China | Ru Xu [506] | No | Yes | Yes | Yes | Yes | Yes | Yes | No | Unclear | Moderate |
| China | Jin Zhang [507] | Yes | No | No | Yes | Yes | Unclear | Yes | No | Unclear | High |
| Japan | Kosuke Mori [508] | No | No | Yes | No | No | Yes | Yes | No | N/A | High |
| Japan | Atsushi Goto [211] | Yes | Yes | Yes | Yes | Yes | Yes | Yes | Yes | Unclear | Low |
| Japan | Kenji Mizumoto [509] | Yes | No | Yes | Yes | Unclear | Unclear | Yes | No | Unclear | Moderate |
| Japan | Kenji Mizumoto [509] | Yes | No | Yes | Yes | Unclear | Unclear | Yes | No | Unclear | Moderate |
| Japan | Kenji Mizumoto [509] | Yes | No | Yes | Yes | Unclear | Unclear | Yes | No | Unclear | Moderate |
| Japan | Kenji Mizumoto [509] | Yes | No | Yes | Yes | Unclear | Unclear | Yes | No | Unclear | Moderate |
| Japan | Kenji Mizumoto [509] | Yes | No | Yes | Yes | Unclear | Unclear | Yes | No | No | Moderate |
| Japan | Prefecture of Hiroshima [212] | Yes | Yes | Yes | No | Unclear | Yes | Yes | No | No | Moderate |
| Japan | Keita Morikane [510] | No | No | Yes | No | Unclear | Yes | Yes | No | N/A | High |
| Japan | Takashi Yoshiyama [119] | No | Yes | Yes | Yes | Unclear | Yes | Yes | No | No | Moderate |
| Malaysia | I-Ching Sam [213] | No | No | Yes | No | Unclear | Yes | Yes | Yes | N/A | Moderate |
| Malaysia | Chong Zhuo Lin [120] | Yes | Yes | Yes | Yes | Yes | Yes | Yes | No | Unclear | Moderate |
| Mongolia | Battogtokh Chimeddorj [121] | Yes | No | Yes | Yes | Yes | Yes | Yes | No | Unclear | Moderate |
| Mongolia | Battogtokh Chimeddorj [121] | Yes | No | Yes | Yes | Yes | Yes | Yes | No | Unclear | Moderate |
| Papua New Guinea | Moses Laman [122] | Yes | No | Yes | Yes | Unclear | Yes | Yes | No | Unclear | Moderate |
| Republic of Korea | Ah-Ra Kim [123] | Yes | No | Yes | Yes | Unclear | Yes | Yes | No | Unclear | Moderate |
| Republic of Korea | Kwangmin Lee [511] | No | No | Yes | No | Unclear | Unclear | Yes | No | N/A | High |
| Republic of Korea | Kwangmin Lee [511] | No | No | Yes | No | Unclear | Unclear | Yes | No | N/A | High |
| Republic of Korea | Kwangmin Lee [511] | No | No | Yes | No | Unclear | Unclear | Yes | No | N/A | High |
| Republic of Korea | Hye Won Jeong [512] | Yes | Yes | Yes | No | Unclear | Unclear | Yes | Yes | Unclear | Low |
| Republic of Korea | Hye Won Jeong [512] | Yes | Yes | Yes | No | Unclear | Unclear | Yes | Yes | Unclear | Low |
| Republic of Korea | Eun Hee Nah [124] | Yes | Yes | Yes | Yes | Unclear | Yes | Yes | No | Unclear | Moderate |
| Singapore | Hannah E Clapham [513] | Unclear | No | Yes | Yes | Yes | Yes | Yes | No | Unclear | High |

#### S4.3 Additional figures and results

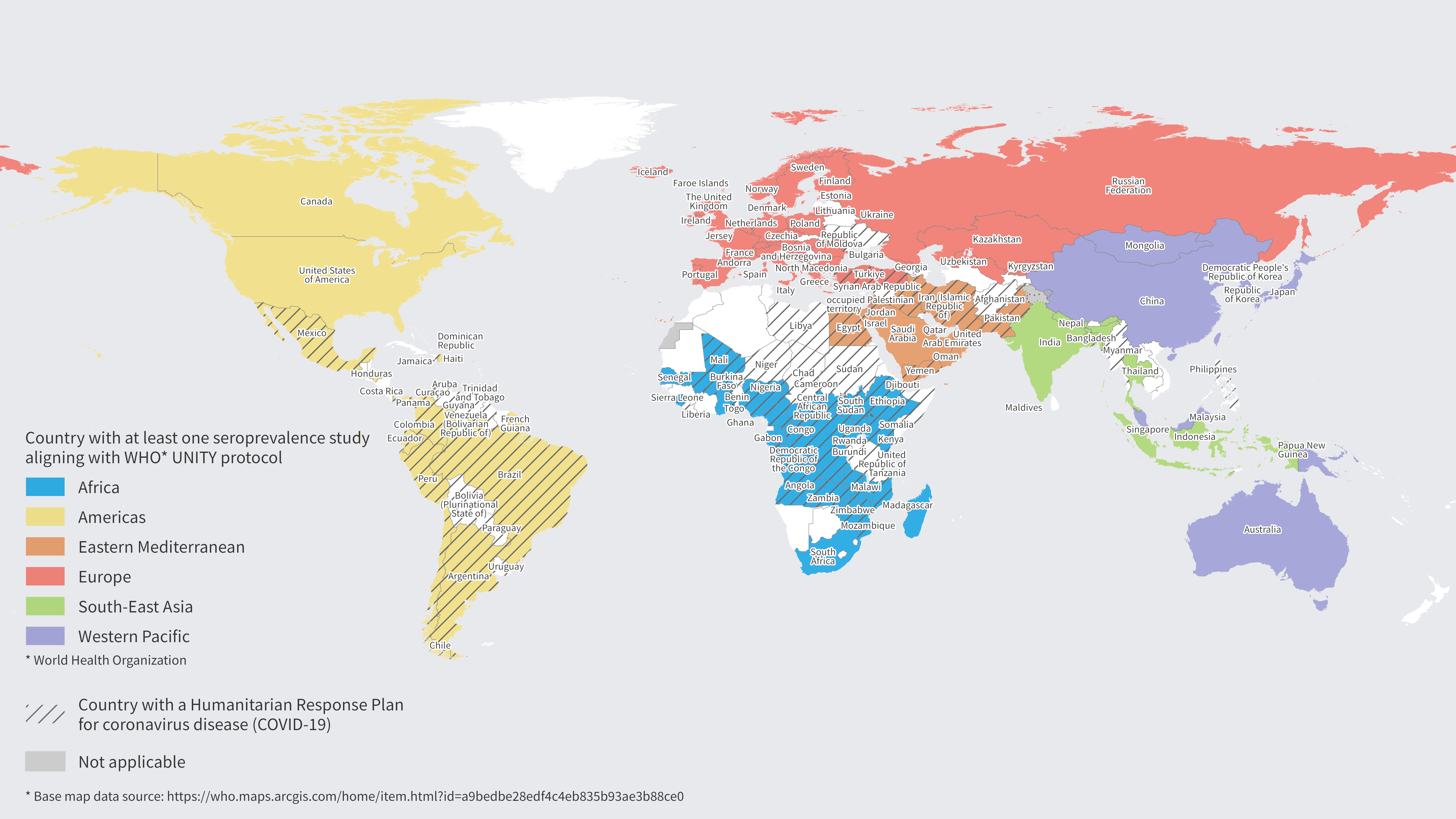

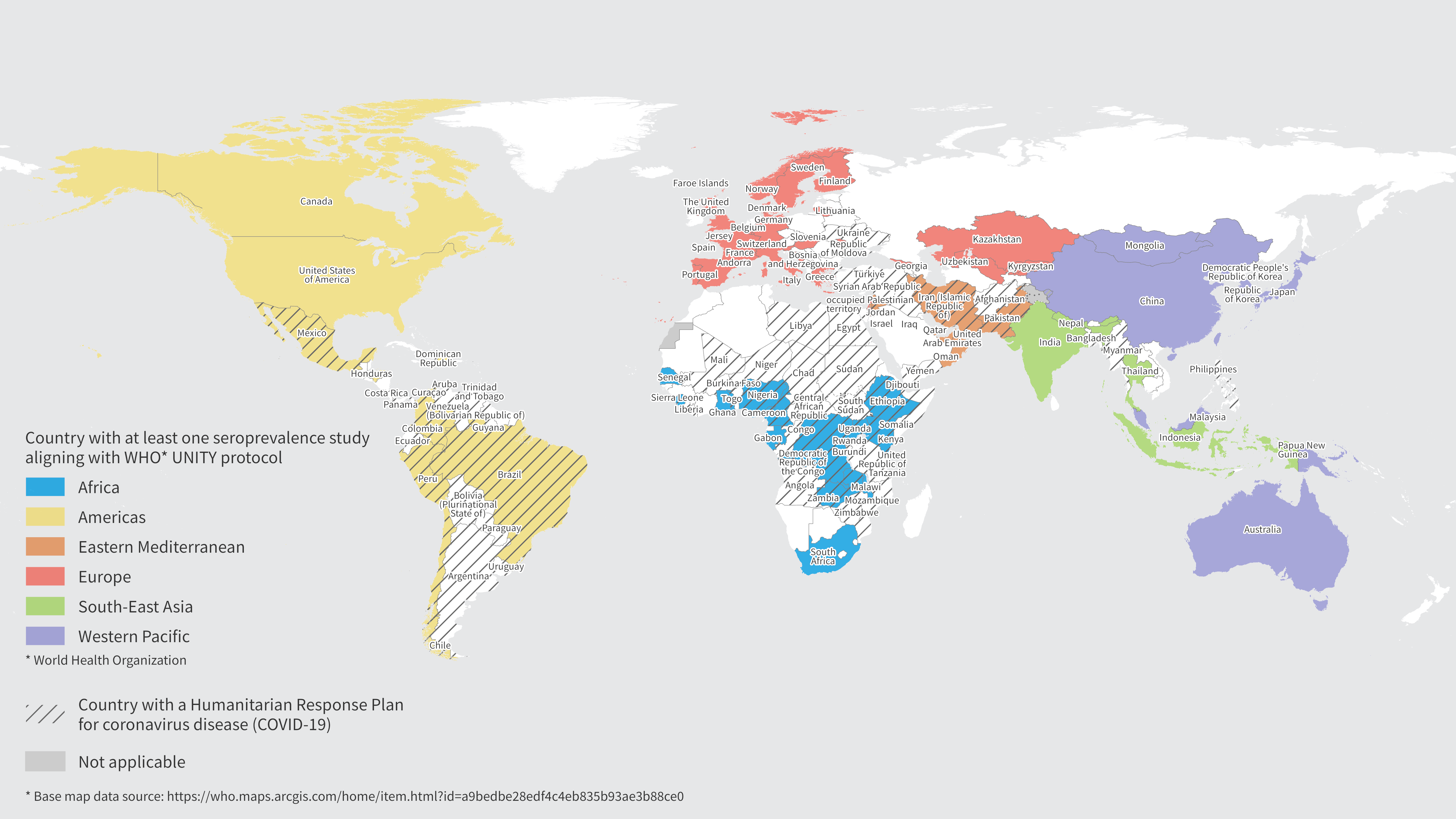

##### Figure S1: WHO Member States with seroprevalence data identified.

Top panel: MS included in descriptive analysis (dataset 0: all studies). Bottom panel: MS included in meta-analysis (sub-dataset 1: national or sub-national studies rated low and moderate risk of bias). Base layer of the map used are from ArcGIS, ESR: <https://who.maps.arcgis.com/home/item.html?id=a9bedbe28edf4c4eb835b93ae3b88ce0>

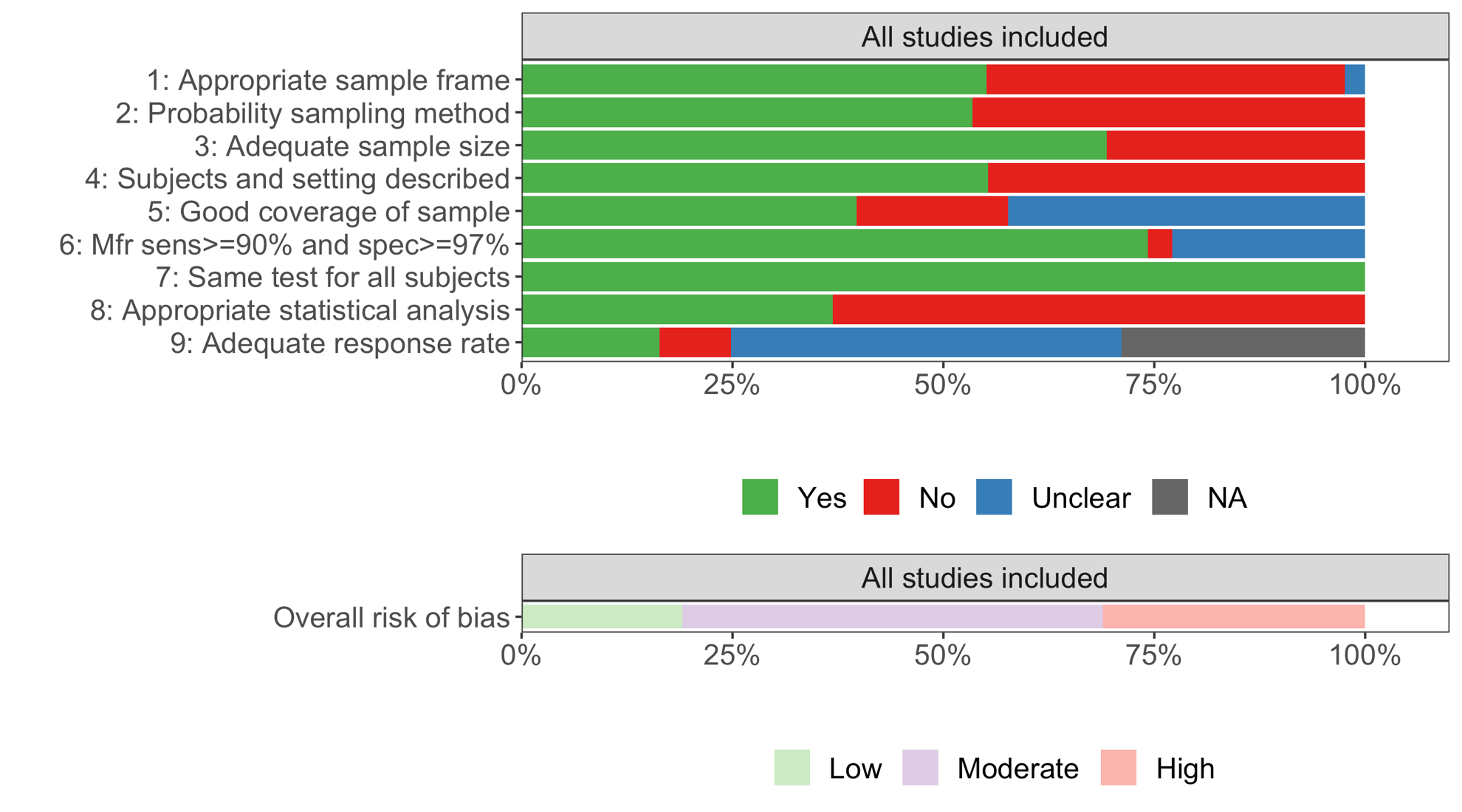

##### Figure S2: Risk of Bias Assessment for Included Studies.

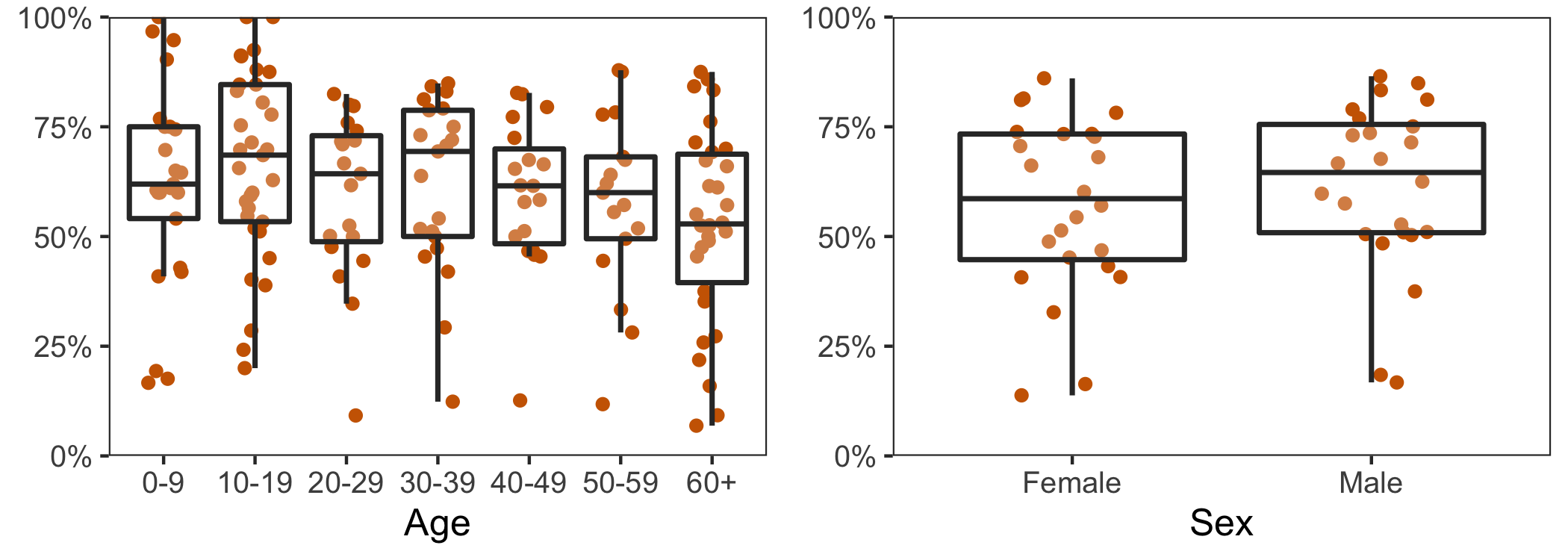

##### Figure S3: Asymptomatic Prevalence in SEROPREV Protocol Studies by Age and Sex.

The inner horizontal line and edges of the box indicate the median and interquartile range of the reported percentage of seropositives with no symptoms, while the whiskers extend from the box to the largest value no further than 1.5 times the interquartile range and indicate the spread of the data. Each point represents an individual SEROPREV protocol seroprevalence study.

##### Table S9. Estimated seroprevalence over time by region and globally (uncorrected for test characteristics)

Infection = infection-induced. NA = no data. See S3.2 for analysis details.

| WHO region | Population (millions) | Month | Countries | Overall N | Overall seroprev [95% CI] | Infection N | Infection seroprev [95% CI] |
| --- | --- | --- | --- | --- | --- | --- | --- |
| Africa | 1118.8 | Feb 2020 | Malawi | 2 | 0.00% [0.12%-2.07%] | 2 | 0.00% [0.12%-2.07%] |
| Africa | 1118.8 | Mar 2020 | Malawi | 3 | 0.00% [0.16%-1.58%] | 3 | 0.00% [0.16%-1.58%] |
| Africa | 1118.8 | Apr 2020 | Malawi | 3 | 0.67% [0.25%-1.80%] | 3 | 0.67% [0.25%-1.80%] |
| Africa | 1118.8 | May 2020 | Kenya, Malawi | 4 | 4.08% [3.23%-5.15%] | 4 | 4.08% [3.23%-5.15%] |
| Africa | 1118.8 | May 2020 | Ethiopia, Kenya, Malawi, Zambia | 7 | 2.93% [2.53%-3.40%] | 7 | 2.93% [2.53%-3.40%] |
| Africa | 1118.8 | Jun 2020 | Ethiopia, Kenya, Malawi, Zambia | 7 | 3.48% [2.88%-4.20%] | 7 | 3.48% [2.88%-4.20%] |
| Africa | 1118.8 | Jul 2020 | Ethiopia, Malawi, South Africa, Zambia | 7 | 10.14% [9.00%-11.42%] | 7 | 10.14% [9.00%-11.42%] |
| Africa | 1118.8 | Aug 2020 | Ethiopia, Malawi, Nigeria, South Africa, Zambia | 12 | 15.03% [12.19%-18.38%] | 12 | 15.03% [12.19%-18.38%] |
| Africa | 1118.8 | Sep 2020 | Ethiopia, Kenya, Malawi, Nigeria, South Africa | 12 | 14.95% [12.34%-17.99%] | 12 | 14.95% [12.34%-17.99%] |
| Africa | 1118.8 | Oct 2020 | Cameroon, Ethiopia, Kenya, Malawi, Nigeria, Senegal, South Africa | 15 | 14.95% [12.55%-17.71%] | 15 | 14.95% [12.55%-17.71%] |
| Africa | 1118.8 | Nov 2020 | Cameroon, Malawi, Nigeria, Senegal, South Africa | 14 | 19.19% [15.77%-23.14%] | 13 | 19.23% [15.05%-24.25%] |
| Africa | 1118.8 | Dec 2020 | Cameroon, Malawi, Nigeria, Senegal, South Africa | 10 | 19.21% [16.73%-21.95%] | NA | NA |
| Africa | 1118.8 | Dec 2020 | Cameroon, Malawi, Senegal, South Africa | NA | NA | 9 | 21.92% [15.53%-30.01%] |
| Africa | 1118.8 | Jan 2021 | Ghana, Kenya, Malawi, Nigeria, South Africa | 9 | 26.64% [24.60%-28.79%] | NA | NA |
| Africa | 1118.8 | Jan 2021 | Ghana, Kenya, Malawi, South Africa | NA | NA | 8 | 38.06% [33.21%-43.16%] |
| Africa | 1118.8 | Feb 2021 | Democratic Republic of the Congo, Ghana, Kenya, Malawi, Sierra Leone, South Africa, Uganda | 12 | 46.18% [44.81%-47.57%] | 12 | 46.18% [44.81%-47.57%] |
| Africa | 1118.8 | Mar 2021 | Democratic Republic of the Congo, Ghana, Kenya, Malawi, Sierra Leone, South Africa, Uganda | 12 | 47.03% [45.95%-48.11%] | 12 | 47.03% [45.95%-48.11%] |
| Africa | 1118.8 | Apr 2021 | Democratic Republic of the Congo, Malawi, Sierra Leone, South Africa, Uganda | 9 | 50.27% [49.11%-51.42%] | 9 | 50.27% [49.11%-51.42%] |
| Africa | 1118.8 | May 2021 | Ghana, Malawi, South Africa, Togo, Uganda | 7 | 44.54% [43.42%-45.65%] | 7 | 44.54% [43.42%-45.65%] |
| Africa | 1118.8 | May 2021 | Ghana, Malawi, Togo | 6 | 65.30% [63.72%-66.85%] | 6 | 65.30% [63.72%-66.85%] |
| Africa | 1118.8 | Jun 2021 | Ghana, Malawi, Nigeria, Togo | 7 | 75.91% [74.86%-76.93%] | 7 | 75.91% [74.86%-76.93%] |
| Africa | 1118.8 | Jul 2021 | Ghana, Malawi, Nigeria | 4 | 76.48% [75.38%-77.56%] | 4 | 76.48% [75.38%-77.56%] |
| Africa | 1118.8 | Aug 2021 | Ghana, Malawi, Nigeria | 3 | 76.80% [75.70%-77.86%] | 3 | 76.80% [75.70%-77.86%] |
| Africa | 1118.8 | Nov 2021 | Gabon, Ghana | 2 | 86.67% [84.56%-88.53%] | NA | NA |
| Africa | 1118.8 | Nov 2021 | Gabon | NA | NA | 1 | 85.57% [84.37%-86.69%] |
| Africa | 1118.8 | Dec 2021 | Gabon, Ghana | 2 | 86.67% [84.56%-88.53%] | NA | NA |
| Americas: High income | 401.1 | Mar 2020 | Canada | 2 | 0.00% [0.36%-1.18%] | 2 | 0.00% [0.36%-1.18%] |
| Americas: High income | 401.1 | Apr 2020 | Canada, United States of America | 6 | 2.38% [1.18%-4.73%] | 4 | 4.84% [4.22%-5.55%] |
| Americas: High income | 401.1 | May 2020 | Canada, United States of America | 19 | 2.74% [1.98%-3.79%] | 17 | 3.00% [2.13%-4.21%] |
| Americas: High income | 401.1 | May 2020 | Canada, United States of America | 27 | 3.77% [2.68%-5.28%] | 23 | 3.97% [2.79%-5.61%] |
| Americas: High income | 401.1 | Jun 2020 | Canada, United States of America | 29 | 3.65% [2.52%-5.25%] | 27 | 3.48% [2.38%-5.06%] |
| Americas: High income | 401.1 | Jul 2020 | Canada, United States of America | 21 | 4.59% [2.83%-7.35%] | 19 | 4.43% [2.64%-7.35%] |
| Americas: High income | 401.1 | Aug 2020 | Canada, United States of America | 17 | 3.80% [2.36%-6.06%] | 17 | 3.80% [2.36%-6.06%] |
| Americas: High income | 401.1 | Sep 2020 | Canada, Chile, United States of America | 15 | 3.93% [2.42%-6.31%] | 15 | 3.93% [2.42%-6.31%] |
| Americas: High income | 401.1 | Oct 2020 | Canada, Chile, United States of America | 17 | 5.02% [3.17%-7.85%] | 17 | 5.02% [3.17%-7.85%] |
| Americas: High income | 401.1 | Nov 2020 | Canada, Chile, United States of America | 14 | 4.99% [2.52%-9.65%] | 14 | 4.99% [2.52%-9.65%] |
| Americas: High income | 401.1 | Dec 2020 | Canada, United States of America | 12 | 6.14% [2.69%-13.37%] | 14 | 5.86% [2.67%-12.37%] |
| Americas: High income | 401.1 | Jan 2021 | Canada, United States of America | 7 | 24.14% [17.11%-32.90%] | 9 | 15.67% [14.07%-17.42%] |
| Americas: High income | 401.1 | Feb 2021 | Canada, United States of America | 9 | 34.78% [24.44%-46.77%] | 11 | 9.95% [4.04%-22.47%] |
| Americas: High income | 401.1 | Mar 2021 | Canada, United States of America | 8 | 45.75% [33.35%-58.70%] | 8 | 10.87% [4.27%-24.99%] |
| Americas: High income | 401.1 | Apr 2021 | Canada, United States of America | 8 | 57.24% [40.74%-72.27%] | 8 | 10.94% [4.25%-25.37%] |
| Americas: High income | 401.1 | May 2021 | Canada, United States of America | 5 | 71.50% [62.39%-79.14%] | 6 | 15.65% [12.42%-19.54%] |
| Americas: High income | 401.1 | May 2021 | Canada, United States of America | 6 | 80.03% [67.54%-88.53%] | 6 | 14.69% [11.13%-19.13%] |
| Americas: High income | 401.1 | Jun 2021 | Canada, United States of America | 6 | 82.27% [70.85%-89.85%] | 6 | 14.72% [11.16%-19.17%] |
| Americas: High income | 401.1 | Jul 2021 | Canada, United States of America | 4 | 90.38% [89.96%-90.78%] | 4 | 16.38% [16.29%-16.47%] |
| Americas: High income | 401.1 | Aug 2021 | Canada, United States of America | 4 | 77.49% [77.02%-77.95%] | 4 | 21.18% [20.91%-21.44%] |
| Americas: High income | 401.1 | Sep 2021 | Canada, Chile, United States of America | 5 | 78.56% [78.11%-79.00%] | 5 | 23.70% [23.41%-23.99%] |
| Americas: High income | 401.1 | Oct 2021 | Canada, Chile, United States of America | 5 | 78.63% [78.16%-79.10%] | 5 | 23.72% [23.42%-24.02%] |
| Americas: High income | 401.1 | Nov 2021 | Canada, Chile | 4 | 97.17% [96.80%-97.50%] | 5 | 27.93% [25.69%-30.29%] |
| Americas: High income | 401.1 | Dec 2021 | Canada | 3 | 97.49% [97.23%-97.72%] | 4 | 7.15% [5.12%-9.90%] |
| Americas: High income | 401.1 | Jan 2022 | Canada | 2 | 97.53% [97.22%-97.81%] | 3 | 8.01% [5.64%-11.27%] |
| Americas: High income | 401.1 | Feb 2022 | Canada | 1 | 97.72% [97.55%-97.88%] | 1 | 12.12% [11.76%-12.49%] |
| Americas: High income | 401.1 | Mar 2022 | Canada | 1 | 99.80% [99.71%-99.86%] | 2 | 33.73% [31.58%-35.96%] |
| Americas: Low and middle income | 620.5 | Apr 2020 | Brazil, Dominican Republic | 5 | 0.38% [0.22%-0.65%] | 4 | 1.76% [1.55%-2.01%] |
| Americas: Low and middle income | 620.5 | May 2020 | Brazil, Colombia, Dominican Republic, Honduras | 11 | 8.17% [2.78%-21.69%] | NA | NA |
| Americas: Low and middle income | 620.5 | May 2020 | Brazil, Colombia, Dominican Republic | NA | NA | 8 | 10.26% [6.05%-16.85%] |
| Americas: Low and middle income | 620.5 | Jun 2020 | Brazil, Colombia, Dominican Republic, Honduras | 13 | 8.87% [3.86%-19.08%] | NA | NA |
| Americas: Low and middle income | 620.5 | Jun 2020 | Brazil, Colombia, Dominican Republic | NA | NA | 9 | 11.64% [7.11%-18.50%] |
| Americas: Low and middle income | 620.5 | Jul 2020 | Brazil, Colombia, Honduras | 15 | 10.49% [6.08%-17.50%] | NA | NA |
| Americas: Low and middle income | 620.5 | Jul 2020 | Brazil, Colombia | NA | NA | 11 | 13.02% [9.29%-17.96%] |
| Americas: Low and middle income | 620.5 | Aug 2020 | Brazil, Peru | 13 | 8.27% [4.80%-13.88%] | 10 | 10.94% [7.48%-15.72%] |
| Americas: Low and middle income | 620.5 | Aug 2020 | Brazil, Colombia, Mexico, Peru | 16 | 18.98% [14.55%-24.37%] | 14 | 20.03% [15.91%-24.89%] |
| Americas: Low and middle income | 620.5 | Sep 2020 | Brazil, Colombia, Mexico, Peru | 12 | 20.95% [14.57%-29.18%] | 12 | 20.95% [14.57%-29.18%] |
| Americas: Low and middle income | 620.5 | Oct 2020 | Brazil, Colombia, Mexico, Peru | 9 | 23.15% [15.05%-33.87%] | 9 | 23.15% [15.05%-33.87%] |
| Americas: Low and middle income | 620.5 | Nov 2020 | Brazil, Colombia | 4 | 25.19% [16.84%-35.90%] | 4 | 25.19% [16.84%-35.90%] |
| Americas: Low and middle income | 620.5 | Dec 2020 | Brazil | 4 | 13.28% [11.10%-15.82%] | 4 | 13.18% [10.91%-15.85%] |
| Americas: Low and middle income | 620.5 | Jan 2021 | Brazil | 5 | 15.96% [11.16%-22.30%] | 5 | 15.79% [11.07%-22.02%] |
| Americas: Low and middle income | 620.5 | Feb 2021 | Brazil | 5 | 15.96% [11.16%-22.30%] | 5 | 15.79% [11.07%-22.02%] |
| Americas: Low and middle income | 620.5 | Mar 2021 | Brazil, Colombia | 5 | 19.53% [13.96%-26.62%] | 5 | 17.33% [11.95%-24.46%] |
| Americas: Low and middle income | 620.5 | Apr 2021 | Brazil, Colombia | 2 | 19.99% [18.83%-21.21%] | 2 | 13.98% [12.97%-15.06%] |
| Eastern Mediterranean | 730.8 | Apr 2020 | Iran (Islamic Republic of) | 3 | 16.92% [13.04%-21.66%] | 3 | 16.92% [13.04%-21.66%] |
| Eastern Mediterranean | 730.8 | May 2020 | Iran (Islamic Republic of) | 3 | 16.92% [13.04%-21.66%] | 3 | 16.92% [13.04%-21.66%] |
| Eastern Mediterranean | 730.8 | Jun 2020 | Iran (Islamic Republic of), Oman, Pakistan, Qatar | 4 | 33.61% [32.82%-34.40%] | NA | NA |
| Eastern Mediterranean | 730.8 | Jun 2020 | Iran (Islamic Republic of), Oman, Pakistan | NA | NA | 3 | 33.79% [33.00%-34.60%] |
| Eastern Mediterranean | 730.8 | Jul 2020 | Jordan, Oman, Pakistan, Qatar, United Arab Emirates | 6 | 38.38% [37.50%-39.27%] | NA | NA |
| Eastern Mediterranean | 730.8 | Jul 2020 | Jordan, Oman, Pakistan, United Arab Emirates | NA | NA | 5 | 38.68% [37.78%-39.58%] |
| Eastern Mediterranean | 730.8 | Aug 2020 | Iran (Islamic Republic of), Jordan, Oman, Pakistan, Qatar, United Arab Emirates | 8 | 31.71% [31.02%-32.41%] | NA | NA |
| Eastern Mediterranean | 730.8 | Aug 2020 | Iran (Islamic Republic of), Jordan, Oman, Pakistan, United Arab Emirates | NA | NA | 7 | 31.87% [31.17%-32.58%] |
| Eastern Mediterranean | 730.8 | Aug 2020 | Iran (Islamic Republic of), Jordan, Oman, Pakistan, Qatar, United Arab Emirates | 8 | 32.30% [30.67%-33.98%] | 8 | 32.30% [30.67%-33.98%] |
| Eastern Mediterranean | 730.8 | Sep 2020 | Iran (Islamic Republic of), Jordan, Oman, Pakistan, Qatar | 8 | 8.80% [7.98%-9.69%] | 8 | 8.80% [7.98%-9.69%] |
| Eastern Mediterranean | 730.8 | Oct 2020 | Iran (Islamic Republic of), Jordan, occupied Palestinian territory, including east Jerusalem, Oman, Pakistan, Qatar | 7 | 9.27% [8.64%-9.93%] | 7 | 9.27% [8.64%-9.93%] |
| Eastern Mediterranean | 730.8 | Nov 2020 | Jordan, Lebanon, occupied Palestinian territory, including east Jerusalem, Oman, Pakistan, Qatar | 8 | 9.20% [8.30%-10.18%] | 8 | 9.20% [8.30%-10.18%] |
| Eastern Mediterranean | 730.8 | Dec 2020 | Jordan, Lebanon, occupied Palestinian territory, including east Jerusalem, Oman | 5 | 26.80% [24.79%-28.92%] | 5 | 26.80% [24.79%-28.92%] |
| Eastern Mediterranean | 730.8 | Jan 2021 | Jordan, Lebanon, occupied Palestinian territory, including east Jerusalem | 5 | 34.83% [29.64%-40.40%] | 5 | 34.83% [29.64%-40.40%] |
| Eastern Mediterranean | 730.8 | Feb 2021 | Lebanon | 2 | 42.71% [37.57%-48.02%] | 2 | 42.71% [37.57%-48.02%] |
| Europe: High income | 529.3 | Mar 2020 | Belgium, Denmark, Finland, France, Germany, Italy, Luxembourg, Slovenia, Switzerland, The United Kingdom | 26 | 2.12% [1.41%-3.17%] | NA | NA |
| Europe: High income | 529.3 | Mar 2020 | Belgium, Finland, France, Germany, Italy, Slovenia, Switzerland, The United Kingdom | NA | NA | 23 | 2.11% [1.40%-3.19%] |
| Europe: High income | 529.3 | Apr 2020 | Belgium, Denmark, Finland, France, Germany, Hungary, Italy, Luxembourg, Slovenia, Spain, Switzerland, The United Kingdom | 43 | 2.87% [1.89%-4.31%] | NA | NA |
| Europe: High income | 529.3 | Apr 2020 | Belgium, Finland, France, Germany, Hungary, Italy, Slovenia, Switzerland, The United Kingdom | NA | NA | 38 | 2.84% [1.76%-4.57%] |
| Europe: High income | 529.3 | May 2020 | Belgium, Denmark, Finland, France, Germany, Hungary, Italy, Luxembourg, Portugal, Slovenia, Spain, Switzerland, The United Kingdom | 59 | 3.16% [2.42%-4.11%] | NA | NA |
| Europe: High income | 529.3 | May 2020 | Belgium, Finland, France, Germany, Hungary, Italy, Slovenia, Switzerland, The United Kingdom | NA | NA | 49 | 3.31% [2.34%-4.66%] |
| Europe: High income | 529.3 | May 2020 | Belgium, Denmark, Finland, France, Germany, Hungary, Italy, Luxembourg, Portugal, Slovenia, Spain, Switzerland, The United Kingdom | 52 | 3.40% [2.62%-4.42%] | NA | NA |
| Europe: High income | 529.3 | May 2020 | Belgium, Finland, France, Germany, Hungary, Italy, Slovenia, Switzerland, The United Kingdom | NA | NA | 42 | 3.61% [2.56%-5.06%] |
| Europe: High income | 529.3 | Jun 2020 | Belgium, Denmark, France, Israel, Italy, Portugal, Spain, Switzerland, The United Kingdom | 35 | 4.34% [3.43%-5.49%] | NA | NA |
| Europe: High income | 529.3 | Jun 2020 | Belgium, France, Israel, Italy, Switzerland, The United Kingdom | NA | NA | 29 | 5.06% [3.87%-6.60%] |
| Europe: High income | 529.3 | Jul 2020 | Belgium, Denmark, Israel, Italy, Lithuania, Portugal, Spain, Switzerland, The United Kingdom | 32 | 5.05% [4.24%-5.99%] | NA | NA |
| Europe: High income | 529.3 | Jul 2020 | Belgium, Israel, Spain, Switzerland, The United Kingdom | NA | NA | 26 | 8.49% [7.99%-9.01%] |
| Europe: High income | 529.3 | Aug 2020 | Belgium, Denmark, Finland, Germany, Greece, Israel, Lithuania, Portugal, Spain, Switzerland, The United Kingdom | 30 | 5.56% [5.21%-5.93%] | NA | NA |
| Europe: High income | 529.3 | Aug 2020 | Belgium, Denmark, Finland, Germany, Greece, Israel, Portugal, Spain, Switzerland, The United Kingdom | NA | NA | 28 | 5.59% [5.25%-5.95%] |
| Europe: High income | 529.3 | Sep 2020 | Belgium, Denmark, Finland, Germany, Greece, Lithuania, Portugal, Slovenia, Spain, Switzerland, The United Kingdom | 33 | 5.60% [4.86%-6.44%] | NA | NA |
| Europe: High income | 529.3 | Sep 2020 | Belgium, Denmark, Finland, Germany, Greece, Portugal, Slovenia, Spain, Switzerland, The United Kingdom | NA | NA | 30 | 5.62% [4.88%-6.47%] |
| Europe: High income | 529.3 | Oct 2020 | Belgium, Denmark, Finland, France, Germany, Greece, Portugal, Slovenia, Spain, Sweden, Switzerland, The United Kingdom | 44 | 5.38% [4.50%-6.42%] | 43 | 5.40% [4.51%-6.44%] |
| Europe: High income | 529.3 | Nov 2020 | Belgium, Denmark, Finland, France, Germany, Greece, Slovenia, Spain, Sweden, Switzerland, The United Kingdom | 46 | 5.39% [4.38%-6.63%] | 45 | 5.39% [4.38%-6.62%] |
| Europe: High income | 529.3 | Dec 2020 | Denmark, Finland, France, Germany, Greece, Norway, Spain, Sweden, Switzerland, The United Kingdom | 40 | 5.68% [4.59%-7.00%] | NA | NA |
| Europe: High income | 529.3 | Dec 2020 | Finland, France, Germany, Greece, Norway, Spain, Sweden, Switzerland, The United Kingdom | NA | NA | 39 | 5.71% [4.60%-7.06%] |
| Europe: High income | 529.3 | Jan 2021 | Denmark, Finland, France, Greece, Norway, Switzerland, The United Kingdom | 34 | 9.61% [8.32%-11.09%] | NA | NA |
| Europe: High income | 529.3 | Jan 2021 | Finland, France, Greece, Norway, Switzerland, The United Kingdom | NA | NA | 29 | 8.34% [7.58%-9.18%] |
| Europe: High income | 529.3 | Feb 2021 | Denmark, Finland, France, Norway, Portugal, Switzerland, The United Kingdom | 26 | 13.38% [11.47%-15.55%] | NA | NA |
| Europe: High income | 529.3 | Feb 2021 | Finland, France, Norway, Portugal, Switzerland, The United Kingdom | NA | NA | 19 | 10.17% [9.28%-11.14%] |
| Europe: High income | 529.3 | Mar 2021 | Finland, France, Portugal, The United Kingdom | 23 | 20.69% [18.36%-23.23%] | 11 | 9.85% [8.62%-11.24%] |
| Europe: High income | 529.3 | Apr 2021 | Finland, Portugal, The United Kingdom | 22 | 37.52% [31.90%-43.49%] | 8 | 10.84% [9.26%-12.65%] |
| Europe: High income | 529.3 | May 2021 | Finland, Sweden, The United Kingdom | 22 | 50.32% [42.99%-57.64%] | NA | NA |
| Europe: High income | 529.3 | May 2021 | Switzerland, The United Kingdom | NA | NA | 4 | 12.16% [11.45%-12.91%] |
| Europe: High income | 529.3 | May 2021 | Finland, Sweden, The United Kingdom | 22 | 62.13% [48.67%-73.95%] | NA | NA |
| Europe: High income | 529.3 | May 2021 | Switzerland, The United Kingdom | NA | NA | 4 | 12.52% [11.32%-13.83%] |
| Europe: High income | 529.3 | Jun 2021 | Finland, Sweden, The United Kingdom | 18 | 74.57% [58.30%-86.01%] | NA | NA |
| Europe: High income | 529.3 | Jun 2021 | Switzerland, The United Kingdom | NA | NA | 3 | 13.12% [12.08%-14.22%] |
| Europe: High income | 529.3 | Jul 2021 | Finland, The United Kingdom | 14 | 85.20% [61.25%-95.45%] | NA | NA |
| Europe: High income | 529.3 | Jul 2021 | The United Kingdom | NA | NA | 1 | 12.71% [11.54%-13.98%] |
| Europe: High income | 529.3 | Aug 2021 | Finland, Sweden, The United Kingdom | 15 | 85.78% [73.00%-93.09%] | NA | NA |
| Europe: High income | 529.3 | Aug 2021 | The United Kingdom | NA | NA | 2 | 15.22% [11.79%-19.43%] |
| Europe: High income | 529.3 | Sep 2021 | Finland, Portugal, Sweden, The United Kingdom | 14 | 86.84% [77.71%-92.59%] | NA | NA |
| Europe: High income | 529.3 | Sep 2021 | The United Kingdom | NA | NA | 1 | 18.09% [16.74%-19.54%] |
| Europe: High income | 529.3 | Oct 2021 | Finland, Portugal, Sweden, The United Kingdom | 17 | 88.09% [81.52%-92.53%] | NA | NA |
| Europe: High income | 529.3 | Oct 2021 | The United Kingdom | NA | NA | 1 | 18.09% [16.74%-19.54%] |
| Europe: High income | 529.3 | Nov 2021 | Finland, Portugal, The United Kingdom | 14 | 90.19% [82.71%-94.64%] | NA | NA |
| Europe: High income | 529.3 | Dec 2021 | Finland, The United Kingdom | 12 | 94.29% [86.49%-97.71%] | NA | NA |
| Europe: High income | 529.3 | Jan 2022 | Finland, The United Kingdom | 11 | 94.06% [88.93%-96.90%] | NA | NA |
| Europe: High income | 529.3 | Jan 2022 | The United Kingdom | NA | NA | 1 | 42.96% [41.22%-44.71%] |
| Europe: High income | 529.3 | Feb 2022 | The United Kingdom | 8 | 95.94% [92.57%-97.82%] | 2 | 47.88% [40.95%-54.89%] |
| Europe: High income | 529.3 | Mar 2022 | The United Kingdom | 6 | 95.88% [92.30%-97.83%] | 2 | 47.88% [40.95%-54.89%] |
| Europe: Low and middle income | 397.4 | Jul 2020 | Georgia, Kyrgyzstan, Uzbekistan | 3 | 22.44% [21.15%-23.79%] | NA | NA |
| Europe: Low and middle income | 397.4 | Jul 2020 | Georgia, Kyrgyzstan | NA | NA | 2 | 20.35% [15.74%-25.89%] |
| Europe: Low and middle income | 397.4 | Aug 2020 | Georgia, Kyrgyzstan, Uzbekistan | 3 | 22.44% [21.15%-23.79%] | NA | NA |
| Europe: Low and middle income | 397.4 | Aug 2020 | Georgia, Kyrgyzstan | NA | NA | 2 | 20.35% [15.74%-25.89%] |
| Europe: Low and middle income | 397.4 | Sep 2020 | Georgia, Uzbekistan | 2 | 20.69% [19.28%-22.18%] | NA | NA |
| Europe: Low and middle income | 397.4 | Sep 2020 | Georgia | NA | NA | 1 | 0.49% [0.22%-1.11%] |
| Europe: Low and middle income | 397.4 | Oct 2020 | Bosnia and Herzegovina, Uzbekistan | 2 | 22.74% [22.39%-23.11%] | NA | NA |
| Europe: Low and middle income | 397.4 | Oct 2020 | Bosnia and Herzegovina | NA | NA | 1 | 19.11% [16.77%-21.70%] |
| Europe: Low and middle income | 397.4 | Nov 2020 | Bosnia and Herzegovina, Georgia, Kazakhstan, Republic of Moldova | 5 | 51.03% [48.82%-53.23%] | 5 | 51.03% [48.82%-53.23%] |
| Europe: Low and middle income | 397.4 | Dec 2020 | Bosnia and Herzegovina, Georgia, Kazakhstan, Republic of Moldova | 5 | 51.03% [48.82%-53.23%] | 5 | 51.03% [48.82%-53.23%] |
| Europe: Low and middle income | 397.4 | Jan 2021 | Bosnia and Herzegovina, Georgia, Kazakhstan, Republic of Moldova | 4 | 52.30% [51.40%-53.20%] | 4 | 52.30% [51.40%-53.20%] |
| Europe: Low and middle income | 397.4 | Jan 2021 | Bosnia and Herzegovina | 1 | 40.27% [38.01%-42.57%] | 1 | 40.27% [38.01%-42.57%] |
| Europe: Low and middle income | 397.4 | May 2021 | Kyrgyzstan | 1 | 48.70% [47.67%-49.72%] | 1 | 47.93% [46.90%-48.95%] |
| Europe: Low and middle income | 397.4 | Jun 2021 | Kyrgyzstan | 1 | 48.70% [47.67%-49.72%] | 1 | 47.93% [46.90%-48.95%] |
| South-East Asia | 2021.4 | Jun 2020 | India | 5 | 9.22% [3.84%-20.50%] | 5 | 9.22% [3.84%-20.50%] |
| South-East Asia | 2021.4 | Jul 2020 | India | 8 | 11.44% [6.14%-20.33%] | 8 | 11.44% [6.14%-20.33%] |
| South-East Asia | 2021.4 | Aug 2020 | India, Indonesia | 8 | 12.83% [7.79%-20.41%] | 8 | 12.83% [7.79%-20.41%] |
| South-East Asia | 2021.4 | Sep 2020 | India, Indonesia, Nepal | 10 | 17.84% [11.94%-25.80%] | 10 | 17.84% [11.94%-25.80%] |
| South-East Asia | 2021.4 | Oct 2020 | India, Indonesia, Nepal | 7 | 24.59% [20.42%-29.30%] | 7 | 24.59% [20.42%-29.30%] |
| South-East Asia | 2021.4 | Nov 2020 | India, Nepal | 5 | 29.60% [24.56%-35.20%] | 5 | 29.60% [24.56%-35.20%] |
| South-East Asia | 2021.4 | Dec 2020 | India, Indonesia, Thailand | 4 | 32.57% [20.19%-47.99%] | 4 | 32.57% [20.19%-47.99%] |
| South-East Asia | 2021.4 | Jan 2021 | India, Indonesia, Thailand | 4 | 32.57% [20.19%-47.99%] | 4 | 32.57% [20.19%-47.99%] |
| South-East Asia | 2021.4 | Feb 2021 | India, Indonesia, Thailand | 4 | 32.57% [20.19%-47.99%] | 4 | 32.57% [20.19%-47.99%] |
| South-East Asia | 2021.4 | Apr 2021 | India | 1 | 64.40% [63.05%-65.72%] | 1 | 61.10% [59.73%-62.45%] |
| South-East Asia | 2021.4 | May 2021 | India | 1 | 64.40% [63.05%-65.72%] | 1 | 61.10% [59.73%-62.45%] |
| South-East Asia | 2021.4 | May 2021 | India | 1 | 64.40% [63.05%-65.72%] | 1 | 61.10% [59.73%-62.45%] |
| South-East Asia | 2021.4 | Jul 2021 | India | 1 | 65.39% [63.37%-67.36%] | 2 | 29.53% [12.41%-55.34%] |
| South-East Asia | 2021.4 | Aug 2021 | India | 6 | 82.21% [75.89%-87.15%] | 11 | 54.76% [38.02%-70.50%] |
| South-East Asia | 2021.4 | Sep 2021 | India | 6 | 82.21% [75.89%-87.15%] | 11 | 54.76% [38.02%-70.50%] |
| Western Pacific | 1932.3 | Apr 2020 | China | 2 | 2.95% [0.85%-9.71%] | 2 | 2.95% [0.85%-9.71%] |
| Western Pacific | 1932.3 | May 2020 | Australia, China, Japan, Papua New Guinea | 5 | 2.68% [0.88%-7.91%] | 5 | 2.68% [0.88%-7.91%] |
| Western Pacific | 1932.3 | Jun 2020 | Australia, Japan, Papua New Guinea, Republic of Korea | 5 | 0.22% [0.12%-0.43%] | 4 | 0.21% [0.11%-0.40%] |
| Western Pacific | 1932.3 | Jul 2020 | Australia, Japan, Papua New Guinea, Republic of Korea | 5 | 0.22% [0.12%-0.43%] | 5 | 0.24% [0.15%-0.38%] |
| Western Pacific | 1932.3 | Aug 2020 | Australia, Malaysia, Republic of Korea | 4 | 0.28% [0.11%-0.74%] | 4 | 0.32% [0.22%-0.45%] |
| Western Pacific | 1932.3 | Aug 2020 | Malaysia, Republic of Korea | 3 | 0.32% [0.17%-0.60%] | 4 | 0.36% [0.26%-0.50%] |
| Western Pacific | 1932.3 | Sep 2020 | Japan, Malaysia, Mongolia, Republic of Korea | 5 | 0.35% [0.21%-0.57%] | NA | NA |
| Western Pacific | 1932.3 | Sep 2020 | Malaysia, Mongolia, Republic of Korea | NA | NA | 5 | 0.45% [0.36%-0.58%] |
| Western Pacific | 1932.3 | Oct 2020 | Japan, Mongolia, Republic of Korea | 5 | 0.29% [0.16%-0.52%] | NA | NA |
| Western Pacific | 1932.3 | Oct 2020 | Mongolia, Republic of Korea | NA | NA | 4 | 0.38% [0.25%-0.58%] |
| Western Pacific | 1932.3 | Nov 2020 | Japan, Mongolia, Republic of Korea | 4 | 0.30% [0.17%-0.53%] | NA | NA |
| Western Pacific | 1932.3 | Nov 2020 | Mongolia, Republic of Korea | NA | NA | 3 | 0.41% [0.26%-0.64%] |
| Western Pacific | 1932.3 | Dec 2020 | Mongolia, Republic of Korea | 2 | 0.19% [0.03%-1.23%] | 2 | 0.19% [0.03%-1.23%] |
| Western Pacific | 1932.3 | Jan 2021 | Malaysia, Mongolia | 2 | 3.90% [2.76%-5.49%] | 2 | 3.90% [2.76%-5.49%] |
| Western Pacific | 1932.3 | Feb 2021 | China, Malaysia, Mongolia | 3 | 1.72% [0.72%-4.05%] | 3 | 1.72% [0.72%-4.05%] |
| Western Pacific | 1932.3 | Mar 2021 | China, Malaysia, Mongolia | 3 | 1.72% [0.72%-4.05%] | 3 | 1.72% [0.72%-4.05%] |
| Western Pacific | 1932.3 | Apr 2021 | China | 1 | 1.67% [0.68%-4.01%] | 1 | 1.67% [0.68%-4.01%] |
| Western Pacific | 1932.3 | May 2021 | China | 1 | 1.67% [0.68%-4.01%] | 1 | 1.67% [0.68%-4.01%] |
| Western Pacific | 1932.3 | Aug 2021 | China | 1 | 34.33% [29.08%-40.00%] | NA | NA |
| Western Pacific | 1932.3 | Sep 2021 | China | 1 | 34.33% [29.08%-40.00%] | NA | NA |
| Western Pacific | 1932.3 | Oct 2021 | China | 2 | 32.33% [28.64%-36.26%] | 2 | 4.32% [1.88%-9.62%] |
| Western Pacific | 1932.3 | Nov 2021 | China | 1 | 30.33% [25.30%-35.89%] | 2 | 4.32% [1.88%-9.62%] |
| Western Pacific | 1932.3 | Dec 2021 | China | 1 | 30.33% [25.30%-35.89%] | NA | NA |
| Western Pacific | 1932.3 | Jan 2022 | Japan | 1 | 94.21% [92.75%-95.38%] | NA | NA |
| Western Pacific | 1932.3 | Feb 2022 | Japan | 1 | 94.21% [92.75%-95.38%] | NA | NA |

| Global | Month | Effect size and 95% CI | With 95% prediction interval |
| --- | --- | --- | --- |
| Global (combined seroprevalence) | Feb 2020 | 0.51% [0.12%-2.07%] | 0.51% [0.51%-0.51%] |
| Global (combined seroprevalence) | Mar 2020 | 0.95% [0.50%-1.80%] | 0.95% [0.51%-1.77%] |
| Global (combined seroprevalence) | Apr 2020 | 4.04% [2.45%-6.57%] | 4.04% [3.18%-5.12%] |
| Global (combined seroprevalence) | May 2020 | 4.95% [3.65%-6.69%] | 4.95% [3.91%-6.26%] |
| Global (combined seroprevalence) | Jun 2020 | 7.72% [5.73%-10.33%] | 7.72% [4.18%-13.83%] |
| Global (combined seroprevalence) | Jul 2020 | 10.69% [8.55%-13.30%] | 10.69% [6.40%-17.34%] |
| Global (combined seroprevalence) | Aug 2020 | 10.96% [8.91%-13.43%] | 10.96% [7.66%-15.45%] |
| Global (combined seroprevalence) | Sep 2020 | 11.05% [9.37%-12.99%] | 11.05% [6.63%-17.85%] |
| Global (combined seroprevalence) | Oct 2020 | 13.16% [11.35%-15.22%] | 13.16% [7.98%-20.95%] |
| Global (combined seroprevalence) | Nov 2020 | 16.69% [14.40%-19.26%] | 16.69% [10.28%-25.94%] |
| Global (combined seroprevalence) | Dec 2020 | 18.22% [11.89%-26.90%] | 18.22% [11.24%-28.18%] |
| Global (combined seroprevalence) | Jan 2021 | 23.33% [20.14%-26.85%] | 23.33% [15.03%-34.36%] |
| Global (combined seroprevalence) | Feb 2021 | 25.67% [20.68%-31.39%] | 25.67% [16.46%-37.72%] |
| Global (combined seroprevalence) | Mar 2021 | 27.15% [21.96%-33.04%] | 27.15% [17.52%-39.52%] |
| Global (combined seroprevalence) | Apr 2021 | 37.68% [32.45%-43.20%] | 37.68% [25.59%-51.52%] |
| Global (combined seroprevalence) | May 2021 | 48.19% [44.37%-52.03%] | 48.19% [35.65%-60.96%] |
| Global (combined seroprevalence) | Jun 2021 | 45.64% [39.90%-51.49%] | 45.64% [32.37%-59.55%] |
| Global (combined seroprevalence) | Jul 2021 | 47.12% [41.12%-53.21%] | 47.12% [33.69%-60.99%] |
| Global (combined seroprevalence) | Aug 2021 | 45.74% [41.23%-50.31%] | 45.74% [39.94%-51.65%] |
| Global (combined seroprevalence) | Sep 2021 | 59.20% [56.10%-62.22%] | 59.20% [51.23%-66.71%] |

| Global | Month | Effect size and 95% CI | With 95% prediction interval |
| --- | --- | --- | --- |
| Global (Infection-induced seroprevalence) | Feb 2020 | 0.51% [0.12%-2.07%] | 0.51% [0.51%-0.51%] |
| Global (Infection-induced seroprevalence) | Mar 2020 | 0.95% [0.50%-1.80%] | 0.95% [0.51%-1.77%] |
| Global (Infection-induced seroprevalence) | Apr 2020 | 4.38% [2.68%-7.07%] | 4.38% [3.53%-5.43%] |
| Global (Infection-induced seroprevalence) | May 2020 | 5.18% [3.85%-6.92%] | 5.18% [4.10%-6.52%] |
| Global (Infection-induced seroprevalence) | Jun 2020 | 8.01% [5.98%-10.66%] | 8.01% [4.34%-14.31%] |
| Global (Infection-induced seroprevalence) | Jul 2020 | 11.05% [9.10%-13.36%] | 11.05% [6.62%-17.87%] |
| Global (Infection-induced seroprevalence) | Aug 2020 | 11.12% [9.83%-12.57%] | 11.12% [7.78%-15.66%] |
| Global (Infection-induced seroprevalence) | Sep 2020 | 10.04% [8.73%-11.53%] | 10.04% [6.00%-16.33%] |
| Global (Infection-induced seroprevalence) | Oct 2020 | 13.00% [11.54%-14.62%] | 13.00% [7.87%-20.72%] |
| Global (Infection-induced seroprevalence) | Nov 2020 | 16.72% [14.73%-18.92%] | 16.72% [10.30%-25.99%] |
| Global (Infection-induced seroprevalence) | Dec 2020 | 18.60% [12.12%-27.45%] | 18.60% [11.50%-28.66%] |
| Global (Infection-induced seroprevalence) | Jan 2021 | 24.38% [21.09%-28.01%] | 24.38% [15.80%-35.65%] |
| Global (Infection-induced seroprevalence) | Feb 2021 | 24.15% [19.33%-29.73%] | 24.15% [15.28%-36.00%] |
| Global (Infection-induced seroprevalence) | Mar 2021 | 24.42% [19.55%-30.05%] | 24.42% [15.45%-36.36%] |
| Global (Infection-induced seroprevalence) | Apr 2021 | 32.12% [27.27%-37.39%] | 32.12% [21.11%-45.56%] |
| Global (Infection-induced seroprevalence) | May 2021 | 38.68% [35.19%-42.29%] | 38.68% [27.28%-51.47%] |
| Global (Infection-induced seroprevalence) | Jun 2021 | 36.56% [31.47%-41.98%] | 36.56% [24.79%-50.20%] |
| Global (Infection-induced seroprevalence) | Jul 2021 | 28.47% [21.70%-36.38%] | 28.47% [18.55%-41.04%] |
| Global (Infection-induced seroprevalence) | Aug 2021 | 28.86% [23.88%-34.41%] | 28.86% [20.48%-38.97%] |
| Global (Infection-induced seroprevalence) | Sep 2021 | 35.85% [29.54%-42.69%] | 35.85% [22.78%-51.42%] |

##### Table S10. Estimated seroprevalence over time by region and globally (corrected for test characteristics)

Infection = infection-induced. NA = no data. See S3.2 for analysis details.

| WHO region | Population (millions) | Month | Countries | Overall N | Overall seroprev [95% CI] |
| --- | --- | --- | --- | --- | --- |
| Africa | 1118.8 | Jan 2020 | Malawi | 2 | 0.00% [0.12%-1.93%] |
| Africa | 1118.8 | Feb 2020 | Malawi | 3 | 0.00% [0.15%-1.52%] |
| Africa | 1118.8 | Mar 2020 | Malawi | 3 | 0.00% [0.16%-1.58%] |
| Africa | 1118.8 | Apr 2020 | Malawi | 3 | 0.67% [0.25%-1.80%] |
| Africa | 1118.8 | May 2020 | Kenya, Malawi | 4 | 2.99% [2.30%-3.88%] |
| Africa | 1118.8 | May 2020 | Ethiopia, Kenya, Malawi, Zambia | 7 | 2.10% [1.78%-2.49%] |
| Africa | 1118.8 | Jun 2020 | Ethiopia, Kenya, Malawi, Zambia | 7 | 2.64% [2.09%-3.32%] |
| Africa | 1118.8 | Jul 2020 | Ethiopia, Kenya, Malawi, South Africa, Zambia | 9 | 9.57% [8.53%-10.72%] |
| Africa | 1118.8 | Aug 2020 | Ethiopia, Kenya, Malawi, Nigeria, South Africa, Zambia | 14 | 14.05% [11.44%-17.15%] |
| Africa | 1118.8 | Sep 2020 | Ethiopia, Kenya, Malawi, Nigeria, South Africa | 12 | 14.73% [11.94%-18.03%] |
| Africa | 1118.8 | Oct 2020 | Cameroon, Ethiopia, Kenya, Malawi, Nigeria, Senegal, South Africa | 15 | 14.75% [12.18%-17.75%] |
| Africa | 1118.8 | Nov 2020 | Cameroon, Malawi, Nigeria, Senegal, South Africa | 14 | 18.96% [15.09%-23.53%] |
| Africa | 1118.8 | Dec 2020 | Cameroon, Malawi, Nigeria, Senegal, South Africa | 10 | 16.18% [13.96%-18.68%] |
| Africa | 1118.8 | Jan 2021 | Ghana, Kenya, Malawi, Nigeria, South Africa | 9 | 24.60% [22.51%-26.82%] |
| Africa | 1118.8 | Feb 2021 | Democratic Republic of the Congo, Ghana, Kenya, Malawi, Sierra Leone, South Africa, Uganda | 12 | 46.47% [44.96%-48.00%] |
| Africa | 1118.8 | Mar 2021 | Democratic Republic of the Congo, Ghana, Kenya, Malawi, Sierra Leone, South Africa, Uganda | 12 | 47.32% [46.12%-48.52%] |
| Africa | 1118.8 | Apr 2021 | Democratic Republic of the Congo, Malawi, Sierra Leone, South Africa, Uganda | 9 | 49.75% [48.37%-51.14%] |
| Africa | 1118.8 | May 2021 | Ghana, Malawi, South Africa, Togo, Uganda | 7 | 43.37% [42.23%-44.52%] |
| Africa | 1118.8 | May 2021 | Ghana, Malawi, Togo | 6 | 66.19% [64.59%-67.75%] |
| Africa | 1118.8 | Jun 2021 | Ghana, Malawi, Nigeria, Togo | 7 | 77.13% [76.10%-78.14%] |
| Africa | 1118.8 | Jul 2021 | Ghana, Malawi, Nigeria | 4 | 77.73% [76.64%-78.79%] |
| Africa | 1118.8 | Aug 2021 | Ghana, Malawi, Nigeria | 3 | 78.05% [76.97%-79.09%] |
| Africa | 1118.8 | Nov 2021 | Gabon, Ghana | 2 | 88.32% [86.31%-90.07%] |
| Africa | 1118.8 | Dec 2021 | Gabon, Ghana | 2 | 88.32% [86.31%-90.07%] |
| Americas: High income | 401.1 | Mar 2020 | Canada | 2 | 0.00% [0.16%-0.80%] |
| Americas: High income | 401.1 | Apr 2020 | Canada, United States of America | 6 | 1.57% [0.54%-4.48%] |
| Americas: High income | 401.1 | May 2020 | Canada, United States of America | 19 | 1.75% [1.16%-2.63%] |
| Americas: High income | 401.1 | May 2020 | Canada, United States of America | 27 | 2.74% [1.77%-4.21%] |
| Americas: High income | 401.1 | Jun 2020 | Canada, United States of America | 29 | 2.53% [1.58%-4.04%] |
| Americas: High income | 401.1 | Jul 2020 | Canada, United States of America | 21 | 3.53% [1.89%-6.51%] |
| Americas: High income | 401.1 | Aug 2020 | Canada, United States of America | 17 | 2.74% [1.45%-5.12%] |
| Americas: High income | 401.1 | Sep 2020 | Canada, Chile, United States of America | 15 | 2.96% [1.56%-5.54%] |
| Americas: High income | 401.1 | Oct 2020 | Canada, Chile, United States of America | 17 | 3.95% [2.12%-7.25%] |
| Americas: High income | 401.1 | Nov 2020 | Canada, Chile, United States of America | 14 | 3.74% [1.46%-9.25%] |
| Americas: High income | 401.1 | Dec 2020 | Canada, United States of America | 12 | 4.83% [1.63%-13.42%] |
| Americas: High income | 401.1 | Jan 2021 | Canada, United States of America | 7 | 23.28% [16.07%-32.48%] |
| Americas: High income | 401.1 | Feb 2021 | Canada, United States of America | 9 | 34.30% [23.68%-46.78%] |
| Americas: High income | 401.1 | Mar 2021 | Canada, United States of America | 8 | 45.72% [33.05%-58.96%] |
| Americas: High income | 401.1 | Apr 2021 | Canada, United States of America | 8 | 57.67% [40.61%-73.08%] |
| Americas: High income | 401.1 | May 2021 | Canada, United States of America | 5 | 72.22% [62.46%-80.25%] |
| Americas: High income | 401.1 | May 2021 | Canada, United States of America | 6 | 81.67% [68.11%-90.28%] |
| Americas: High income | 401.1 | Jun 2021 | Canada, United States of America | 6 | 83.89% [71.58%-91.50%] |
| Americas: High income | 401.1 | Jul 2021 | Canada, United States of America | 4 | 92.09% [91.64%-92.52%] |
| Americas: High income | 401.1 | Aug 2021 | Canada, United States of America | 4 | 80.97% [80.46%-81.47%] |
| Americas: High income | 401.1 | Sep 2021 | Canada, Chile, United States of America | 5 | 81.94% [81.28%-82.58%] |
| Americas: High income | 401.1 | Oct 2021 | Canada, Chile, United States of America | 5 | 81.98% [81.29%-82.65%] |
| Americas: High income | 401.1 | Nov 2021 | Canada, Chile | 4 | 99.16% [98.91%-99.35%] |
| Americas: High income | 401.1 | Dec 2021 | Canada | 3 | 99.11% [98.98%-99.21%] |
| Americas: High income | 401.1 | Jan 2022 | Canada | 2 | 99.12% [98.97%-99.25%] |
| Americas: High income | 401.1 | Feb 2022 | Canada | 1 | 99.20% [99.10%-99.29%] |
| Americas: High income | 401.1 | Mar 2022 | Canada | 1 | 99.80% [99.71%-99.86%] |
| Americas: High income | 401.1 | Apr 2022 | Canada | 1 | 99.80% [99.71%-99.86%] |
| Americas: Low and middle income | 620.5 | Apr 2020 | Brazil, Dominican Republic | 5 | 0.45% [0.23%-0.88%] |
| Americas: Low and middle income | 620.5 | May 2020 | Brazil, Colombia, Dominican Republic, Honduras | 11 | 8.04% [2.30%-24.55%] |
| Americas: Low and middle income | 620.5 | Jun 2020 | Brazil, Colombia, Dominican Republic, Honduras | 13 | 8.41% [2.84%-22.36%] |
| Americas: Low and middle income | 620.5 | Jul 2020 | Brazil, Colombia, Honduras | 15 | 9.51% [4.39%-19.40%] |
| Americas: Low and middle income | 620.5 | Aug 2020 | Brazil, Peru | 13 | 7.22% [3.19%-15.54%] |
| Americas: Low and middle income | 620.5 | Aug 2020 | Brazil, Colombia, Mexico, Peru | 16 | 18.57% [12.70%-26.35%] |
| Americas: Low and middle income | 620.5 | Sep 2020 | Brazil, Colombia, Mexico, Peru | 12 | 20.99% [13.15%-31.78%] |
| Americas: Low and middle income | 620.5 | Oct 2020 | Brazil, Colombia, Mexico, Peru | 9 | 23.98% [15.53%-35.11%] |
| Americas: Low and middle income | 620.5 | Nov 2020 | Brazil, Colombia | 4 | 27.18% [18.41%-38.16%] |
| Americas: Low and middle income | 620.5 | Dec 2020 | Brazil | 4 | 13.23% [9.57%-18.01%] |
| Americas: Low and middle income | 620.5 | Jan 2021 | Brazil | 5 | 16.03% [10.42%-23.85%] |
| Americas: Low and middle income | 620.5 | Feb 2021 | Brazil | 5 | 16.03% [10.42%-23.85%] |
| Americas: Low and middle income | 620.5 | Mar 2021 | Brazil, Colombia | 5 | 18.89% [12.40%-27.68%] |
| Americas: Low and middle income | 620.5 | Apr 2021 | Brazil, Colombia | 2 | 19.03% [17.88%-20.22%] |
| Eastern Mediterranean | 730.8 | Apr 2020 | Iran (Islamic Republic of) | 3 | 18.66% [12.08%-27.70%] |
| Eastern Mediterranean | 730.8 | May 2020 | Iran (Islamic Republic of) | 3 | 18.66% [12.08%-27.70%] |
| Eastern Mediterranean | 730.8 | Jun 2020 | Iran (Islamic Republic of), Oman, Pakistan, Qatar | 4 | 35.45% [34.67%-36.23%] |
| Eastern Mediterranean | 730.8 | Jul 2020 | Jordan, Oman, Pakistan, Qatar, United Arab Emirates | 6 | 39.89% [38.77%-41.02%] |
| Eastern Mediterranean | 730.8 | Aug 2020 | Iran (Islamic Republic of), Jordan, Oman, Pakistan, Qatar, United Arab Emirates | 8 | 32.98% [32.13%-33.85%] |
| Eastern Mediterranean | 730.8 | Aug 2020 | Iran (Islamic Republic of), Jordan, Oman, Pakistan, Qatar, United Arab Emirates | 8 | 33.60% [31.63%-35.63%] |
| Eastern Mediterranean | 730.8 | Sep 2020 | Iran (Islamic Republic of), Jordan, Oman, Pakistan, Qatar | 8 | 8.91% [8.00%-9.91%] |
| Eastern Mediterranean | 730.8 | Oct 2020 | Iran (Islamic Republic of), Jordan, occupied Palestinian territory, including east Jerusalem, Oman, Pakistan, Qatar | 7 | 9.38% [8.75%-10.05%] |
| Eastern Mediterranean | 730.8 | Nov 2020 | Jordan, Lebanon, occupied Palestinian territory, including east Jerusalem, Oman, Pakistan, Qatar | 8 | 9.24% [8.29%-10.27%] |
| Eastern Mediterranean | 730.8 | Dec 2020 | Jordan, Lebanon, occupied Palestinian territory, including east Jerusalem, Oman | 5 | 28.15% [26.07%-30.33%] |
| Eastern Mediterranean | 730.8 | Jan 2021 | Jordan, Lebanon, occupied Palestinian territory, including east Jerusalem | 5 | 36.72% [31.75%-41.99%] |
| Eastern Mediterranean | 730.8 | Feb 2021 | Lebanon | 2 | 42.66% [37.27%-48.24%] |
| Europe: High income | 529.3 | Mar 2020 | Belgium, Denmark, Finland, France, Germany, Italy, Luxembourg, Slovenia, Switzerland, The United Kingdom | 25 | 1.37% [0.83%-2.26%] |
| Europe: High income | 529.3 | Apr 2020 | Belgium, Denmark, Finland, France, Germany, Hungary, Italy, Luxembourg, Slovenia, Spain, Switzerland, The United Kingdom | 42 | 1.80% [1.08%-2.96%] |
| Europe: High income | 529.3 | May 2020 | Belgium, Denmark, Finland, France, Germany, Hungary, Italy, Luxembourg, Portugal, Slovenia, Spain, Switzerland, The United Kingdom | 58 | 2.13% [1.56%-2.90%] |
| Europe: High income | 529.3 | May 2020 | Belgium, Denmark, Finland, France, Germany, Hungary, Italy, Luxembourg, Portugal, Slovenia, Spain, Switzerland, The United Kingdom | 51 | 2.29% [1.68%-3.10%] |
| Europe: High income | 529.3 | Jun 2020 | Belgium, Denmark, France, Israel, Italy, Portugal, Spain, Switzerland, The United Kingdom | 35 | 2.94% [2.41%-3.58%] |
| Europe: High income | 529.3 | Jul 2020 | Belgium, Denmark, Israel, Italy, Lithuania, Portugal, Spain, Switzerland, The United Kingdom | 32 | 3.75% [2.97%-4.73%] |
| Europe: High income | 529.3 | Aug 2020 | Belgium, Denmark, Finland, Germany, Greece, Israel, Lithuania, Portugal, Spain, Switzerland, The United Kingdom | 31 | 4.41% [4.07%-4.78%] |
| Europe: High income | 529.3 | Sep 2020 | Belgium, Denmark, Finland, Germany, Greece, Lithuania, Portugal, Slovenia, Spain, Switzerland, The United Kingdom | 35 | 4.53% [4.20%-4.88%] |
| Europe: High income | 529.3 | Oct 2020 | Belgium, Denmark, Finland, France, Germany, Greece, Portugal, Slovenia, Spain, Sweden, Switzerland, The United Kingdom | 46 | 4.05% [3.20%-5.12%] |
| Europe: High income | 529.3 | Nov 2020 | Belgium, Denmark, Finland, France, Germany, Greece, Slovenia, Spain, Sweden, Switzerland, The United Kingdom | 50 | 4.15% [3.19%-5.39%] |
| Europe: High income | 529.3 | Dec 2020 | Denmark, Finland, France, Germany, Greece, Norway, Spain, Sweden, Switzerland, The United Kingdom | 46 | 4.48% [3.43%-5.82%] |
| Europe: High income | 529.3 | Jan 2021 | Denmark, Finland, France, Greece, Norway, Switzerland, The United Kingdom | 40 | 7.83% [6.55%-9.33%] |
| Europe: High income | 529.3 | Feb 2021 | Denmark, Finland, France, Norway, Portugal, Switzerland, The United Kingdom | 31 | 11.93% [10.04%-14.13%] |
| Europe: High income | 529.3 | Mar 2021 | Finland, France, Portugal, The United Kingdom | 24 | 19.82% [17.31%-22.60%] |
| Europe: High income | 529.3 | Apr 2021 | Finland, Portugal, The United Kingdom | 22 | 38.76% [32.43%-45.48%] |
| Europe: High income | 529.3 | May 2021 | Finland, Sweden, The United Kingdom | 22 | 53.23% [44.19%-62.07%] |
| Europe: High income | 529.3 | May 2021 | Finland, Sweden, The United Kingdom | 22 | 67.58% [49.80%-81.41%] |
| Europe: High income | 529.3 | Jun 2021 | Finland, Sweden, The United Kingdom | 18 | 80.75% [59.31%-92.35%] |
| Europe: High income | 529.3 | Jul 2021 | Finland, The United Kingdom | 14 | 91.32% [61.54%-98.57%] |
| Europe: High income | 529.3 | Aug 2021 | Finland, Sweden, The United Kingdom | 15 | 91.36% [76.48%-97.18%] |
| Europe: High income | 529.3 | Sep 2021 | Finland, Portugal, Sweden, The United Kingdom | 14 | 91.56% [80.88%-96.53%] |
| Europe: High income | 529.3 | Oct 2021 | Finland, Portugal, Sweden, The United Kingdom | 17 | 92.49% [85.36%-96.30%] |
| Europe: High income | 529.3 | Nov 2021 | Finland, Portugal, The United Kingdom | 14 | 94.41% [86.93%-97.72%] |
| Europe: High income | 529.3 | Dec 2021 | Finland, The United Kingdom | 12 | 97.43% [91.85%-99.22%] |
| Europe: High income | 529.3 | Jan 2022 | Finland, The United Kingdom | 11 | 96.29% [91.74%-98.38%] |
| Europe: High income | 529.3 | Feb 2022 | The United Kingdom | 8 | 98.57% [96.92%-99.34%] |
| Europe: High income | 529.3 | Mar 2022 | The United Kingdom | 6 | 98.53% [96.77%-99.33%] |
| Europe: Low and middle income | 397.4 | Jul 2020 | Georgia, Kyrgyzstan, Uzbekistan | 3 | 24.03% [22.15%-26.01%] |
| Europe: Low and middle income | 397.4 | Aug 2020 | Georgia, Kyrgyzstan, Uzbekistan | 3 | 24.03% [22.15%-26.01%] |
| Europe: Low and middle income | 397.4 | Sep 2020 | Georgia, Uzbekistan | 2 | 22.66% [20.57%-24.91%] |
| Europe: Low and middle income | 397.4 | Oct 2020 | Bosnia and Herzegovina, Uzbekistan | 2 | 24.87% [24.50%-25.24%] |
| Europe: Low and middle income | 397.4 | Nov 2020 | Bosnia and Herzegovina, Georgia, Kazakhstan, Republic of Moldova | 5 | 52.33% [50.26%-54.40%] |
| Europe: Low and middle income | 397.4 | Dec 2020 | Bosnia and Herzegovina, Georgia, Kazakhstan, Republic of Moldova | 5 | 52.33% [50.26%-54.40%] |
| Europe: Low and middle income | 397.4 | Jan 2021 | Bosnia and Herzegovina, Georgia, Kazakhstan, Republic of Moldova | 4 | 53.52% [52.62%-54.42%] |
| Europe: Low and middle income | 397.4 | Jan 2021 | Bosnia and Herzegovina | 1 | 40.05% [37.80%-42.35%] |
| Europe: Low and middle income | 397.4 | May 2021 | Kyrgyzstan | 1 | 48.70% [47.67%-49.72%] |
| Europe: Low and middle income | 397.4 | Jun 2021 | Kyrgyzstan | 1 | 48.70% [47.67%-49.72%] |
| South-East Asia | 2021.4 | Jun 2020 | India | 5 | 8.73% [3.43%-20.47%] |
| South-East Asia | 2021.4 | Jul 2020 | India | 8 | 11.06% [5.72%-20.33%] |
| South-East Asia | 2021.4 | Aug 2020 | India, Indonesia | 8 | 12.63% [7.53%-20.44%] |
| South-East Asia | 2021.4 | Sep 2020 | India, Indonesia, Nepal | 10 | 17.88% [11.59%-26.55%] |
| South-East Asia | 2021.4 | Oct 2020 | India, Indonesia, Nepal | 7 | 25.22% [20.48%-30.64%] |
| South-East Asia | 2021.4 | Nov 2020 | India, Nepal | 5 | 30.47% [24.41%-37.30%] |
| South-East Asia | 2021.4 | Dec 2020 | India, Indonesia, Thailand | 4 | 35.20% [19.16%-55.47%] |
| South-East Asia | 2021.4 | Jan 2021 | India, Indonesia, Thailand | 4 | 35.20% [19.16%-55.47%] |
| South-East Asia | 2021.4 | Feb 2021 | India, Indonesia, Thailand | 4 | 35.20% [19.16%-55.47%] |
| South-East Asia | 2021.4 | Apr 2021 | India | 1 | 65.10% [63.76%-66.42%] |
| South-East Asia | 2021.4 | May 2021 | India | 1 | 65.10% [63.76%-66.42%] |
| South-East Asia | 2021.4 | May 2021 | India | 1 | 65.10% [63.76%-66.42%] |
| South-East Asia | 2021.4 | Jul 2021 | India | 1 | 68.65% [66.67%-70.56%] |
| South-East Asia | 2021.4 | Aug 2021 | India | 6 | 88.66% [79.95%-93.88%] |
| South-East Asia | 2021.4 | Sep 2021 | India | 6 | 88.66% [79.95%-93.88%] |
| Western Pacific | 1932.3 | Apr 2020 | China | 2 | 2.85% [0.87%-8.99%] |
| Western Pacific | 1932.3 | May 2020 | Australia, China, Japan, Papua New Guinea | 5 | 2.59% [0.88%-7.34%] |
| Western Pacific | 1932.3 | Jun 2020 | Australia, Japan, Papua New Guinea, Republic of Korea | 5 | 0.20% [0.11%-0.38%] |
| Western Pacific | 1932.3 | Jul 2020 | Australia, Japan, Papua New Guinea, Republic of Korea | 5 | 0.20% [0.11%-0.38%] |
| Western Pacific | 1932.3 | Aug 2020 | Australia, Malaysia, Republic of Korea | 4 | 0.12% [0.05%-0.33%] |
| Western Pacific | 1932.3 | Aug 2020 | Malaysia, Republic of Korea | 3 | 0.17% [0.09%-0.33%] |
| Western Pacific | 1932.3 | Sep 2020 | Japan, Malaysia, Mongolia, Republic of Korea | 5 | 0.17% [0.08%-0.34%] |
| Western Pacific | 1932.3 | Oct 2020 | Japan, Mongolia, Republic of Korea | 5 | 0.16% [0.07%-0.35%] |
| Western Pacific | 1932.3 | Nov 2020 | Japan, Mongolia, Republic of Korea | 4 | 0.16% [0.07%-0.35%] |
| Western Pacific | 1932.3 | Dec 2020 | Mongolia, Republic of Korea | 2 | 0.14% [0.02%-0.91%] |
| Western Pacific | 1932.3 | Jan 2021 | Malaysia, Mongolia | 2 | 2.29% [1.46%-3.58%] |
| Western Pacific | 1932.3 | Feb 2021 | China, Malaysia, Mongolia | 3 | 1.68% [0.70%-3.96%] |
| Western Pacific | 1932.3 | Mar 2021 | China, Malaysia, Mongolia | 3 | 1.68% [0.70%-3.96%] |
| Western Pacific | 1932.3 | Apr 2021 | China | 1 | 1.67% [0.68%-4.01%] |
| Western Pacific | 1932.3 | May 2021 | China | 1 | 1.67% [0.68%-4.01%] |
| Western Pacific | 1932.3 | Aug 2021 | China | 1 | 34.33% [29.08%-40.00%] |
| Western Pacific | 1932.3 | Sep 2021 | China | 1 | 34.33% [29.08%-40.00%] |
| Western Pacific | 1932.3 | Oct 2021 | China | 2 | 32.33% [28.64%-36.26%] |
| Western Pacific | 1932.3 | Nov 2021 | China | 1 | 30.33% [25.30%-35.89%] |
| Western Pacific | 1932.3 | Dec 2021 | China | 1 | 30.33% [25.30%-35.89%] |
| Western Pacific | 1932.3 | Jan 2022 | Japan | 1 | 94.21% [92.75%-95.38%] |
| Western Pacific | 1932.3 | Feb 2022 | Japan | 1 | 94.21% [92.75%-95.38%] |

| Global | Month | Effect size and 95% CI | With 95% prediction interval |
| --- | --- | --- | --- |
| Global (combined seroprevalence) | Jan 2020 | 0.48% [0.12%-1.93%] | 0.48% [0.48%-0.48%] |
| Global (combined seroprevalence) | Feb 2020 | 0.48% [0.15%-1.52%] | 0.48% [0.15%-1.52%] |
| Global (combined seroprevalence) | Mar 2020 | 0.70% [0.36%-1.35%] | 0.70% [0.37%-1.30%] |
| Global (combined seroprevalence) | Apr 2020 | 4.08% [2.50%-6.60%] | 4.08% [3.06%-5.42%] |
| Global (combined seroprevalence) | May 2020 | 4.53% [3.34%-6.13%] | 4.53% [3.49%-5.88%] |
| Global (combined seroprevalence) | Jun 2020 | 7.44% [5.42%-10.13%] | 7.44% [3.85%-13.89%] |
| Global (combined seroprevalence) | Jul 2020 | 10.51% [8.31%-13.21%] | 10.51% [6.06%-17.61%] |
| Global (combined seroprevalence) | Aug 2020 | 10.83% [8.72%-13.37%] | 10.83% [7.36%-15.66%] |
| Global (combined seroprevalence) | Sep 2020 | 10.97% [8.92%-13.43%] | 10.97% [6.36%-18.29%] |
| Global (combined seroprevalence) | Oct 2020 | 13.31% [10.90%-16.14%] | 13.31% [7.79%-21.80%] |
| Global (combined seroprevalence) | Nov 2020 | 16.93% [13.88%-20.48%] | 16.93% [10.09%-27.01%] |
| Global (combined seroprevalence) | Dec 2020 | 18.50% [11.88%-27.66%] | 18.50% [11.00%-29.44%] |
| Global (combined seroprevalence) | Jan 2021 | 23.38% [19.38%-27.93%] | 23.38% [14.56%-35.34%] |
| Global (combined seroprevalence) | Feb 2021 | 26.26% [20.66%-32.75%] | 26.26% [16.39%-39.28%] |
| Global (combined seroprevalence) | Mar 2021 | 27.74% [21.92%-34.42%] | 27.74% [17.44%-41.09%] |
| Global (combined seroprevalence) | Apr 2021 | 37.80% [32.56%-43.34%] | 37.80% [25.02%-52.54%] |
| Global (combined seroprevalence) | May 2021 | 48.80% [44.91%-52.71%] | 48.80% [35.52%-62.26%] |
| Global (combined seroprevalence) | Jun 2021 | 46.42% [40.51%-52.43%] | 46.42% [32.27%-61.18%] |
| Global (combined seroprevalence) | Jul 2021 | 48.58% [42.11%-55.09%] | 48.58% [34.18%-63.21%] |
| Global (combined seroprevalence) | Aug 2021 | 47.33% [42.05%-52.67%] | 47.33% [38.28%-56.55%] |
| Global (combined seroprevalence) | Sep 2021 | 61.48% [56.69%-66.05%] | 61.48% [48.94%-72.66%] |

##### Table S11: Meta-regression results and model comparison.

|  | **Model 1 (full model)** | | | **Model 2** | | | **Model 3** | | | **Model 4** | | |
| --- | --- | --- | --- | --- | --- | --- | --- | --- | --- | --- | --- | --- |
| **Characteristic** | **PR** | **95% CI** | **p-value** | **PR** | **95% CI** | **p-value** | **PR** | **95% CI** | **p-value** | **PR** | **95% CI** | **p-value** |
| **WHO region** |  |  |  |  |  |  |  |  |  |  |  |  |
| Americas | — | — |  | — | — |  | — | — |  | — | — |  |
| Europe | 1.35 | 1.02, 1.78 | 0.038 | 1.32 | 1.01, 1.72 | 0.042 | 1.22 | 0.93, 1.59 | 0.14 | 1.40 | 1.06, 1.86 | 0.017 |
| Western Pacific | 0.17 | 0.10, 0.29 | <0.001 | 0.17 | 0.10, 0.29 | <0.001 | 0.15 | 0.09, 0.26 | <0.001 | 0.17 | 0.10, 0.30 | <0.001 |
| Africa | 1.28 | 0.71, 2.29 | 0.4 | 1.36 | 0.76, 2.42 | 0.3 | 1.18 | 0.66, 2.11 | 0.6 | 1.31 | 0.72, 2.36 | 0.4 |
| South-East Asia | 1.41 | 0.79, 2.53 | 0.2 | 1.42 | 0.80, 2.54 | 0.2 | 1.41 | 0.79, 2.54 | 0.2 | 1.36 | 0.76, 2.45 | 0.3 |
| Eastern Mediterranean | 1.69 | 1.08, 2.64 | 0.023 | 1.66 | 1.07, 2.59 | 0.025 | 1.45 | 0.94, 2.23 | 0.090 | 1.74 | 1.11, 2.74 | 0.016 |
| **Income class** |  |  |  |  |  |  |  |  |  |  |  |  |
| High income country | — | — |  | — | — |  | — | — |  | — | — |  |
| Low income country | 7.33 | 3.49, 15.4 | <0.001 | 6.55 | 3.19, 13.5 | <0.001 | 7.19 | 3.41, 15.2 | <0.001 | 5.54 | 2.73, 11.3 | <0.001 |
| Lower middle income country | 7.75 | 4.69, 12.8 | <0.001 | 7.25 | 4.42, 11.9 | <0.001 | 7.76 | 4.69, 12.8 | <0.001 | 6.55 | 4.03, 10.6 | <0.001 |
| Upper middle income country | 3.97 | 2.88, 5.49 | <0.001 | 3.86 | 2.84, 5.26 | <0.001 | 4.02 | 2.91, 5.56 | <0.001 | 3.62 | 2.64, 4.96 | <0.001 |
| **Geographic scope** |  |  |  |  |  |  |  |  |  |  |  |  |
| National | — | — |  | — | — |  |  |  |  | — | — |  |
| Subnational | 1.27 | 1.02, 1.59 | 0.033 | 1.27 | 1.02, 1.59 | 0.032 |  |  |  | 1.34 | 1.08, 1.67 | 0.009 |
| **Cases per 100 pop.** | 1.39 | 1.30, 1.49 | <0.001 | 1.38 | 1.29, 1.48 | <0.001 | 1.40 | 1.30, 1.50 | <0.001 | 1.35 | 1.27, 1.44 | <0.001 |
| **Transmission phase** |  |  |  |  |  |  |  |  |  |  |  |  |
| Before peak of 1st wave | — | — |  | — | — |  | — | — |  | — | — |  |
| After peak of 1st wave | 1.41 | 1.26, 1.58 | <0.001 | 1.41 | 1.26, 1.58 | <0.001 | 1.41 | 1.26, 1.58 | <0.001 | 1.38 | 1.24, 1.54 | <0.001 |
| After peak of 2nd wave | 3.40 | 2.85, 4.05 | <0.001 | 3.41 | 2.86, 4.07 | <0.001 | 3.39 | 2.84, 4.04 | <0.001 | 3.34 | 2.80, 3.99 | <0.001 |
| **Sample frame** |  |  |  |  |  |  |  |  |  |  |  |  |
| Household and community samples | — | — |  |  |  |  | — | — |  | — | — |  |
| Blood donors | 1.04 | 0.77, 1.40 | 0.8 |  |  |  | 0.99 | 0.74, 1.33 | >0.9 | 1.08 | 0.80, 1.46 | 0.6 |
| Residual sera | 1.08 | 0.83, 1.41 | 0.6 |  |  |  | 1.05 | 0.81, 1.37 | 0.7 | 1.10 | 0.85, 1.43 | 0.5 |
| Pregnant or parturient women | 0.79 | 0.38, 1.65 | 0.5 |  |  |  | 0.76 | 0.36, 1.60 | 0.5 | 0.90 | 0.43, 1.87 | 0.8 |
| Multiple general populations | 2.13 | 0.78, 5.86 | 0.14 |  |  |  | 2.19 | 0.80, 6.05 | 0.13 | 2.05 | 0.74, 5.66 | 0.2 |
| **PHSM index (0-10)** | 0.89 | 0.80, 0.98 | 0.022 | 0.89 | 0.81, 0.99 | 0.027 | 0.87 | 0.79, 0.96 | 0.006 |  |  |  |
| **Age** |  |  |  |  |  |  |  |  |  |  |  |  |
| Adults (18-64 years) | — | — |  | — | — |  | — | — |  | — | — |  |
| Multiple groups | 0.84 | 0.77, 0.91 | <0.001 | 0.84 | 0.78, 0.92 | <0.001 | 0.84 | 0.77, 0.91 | <0.001 | 0.85 | 0.78, 0.92 | <0.001 |
| Children and Youth (0-17 years) | 0.58 | 0.47, 0.71 | <0.001 | 0.59 | 0.48, 0.72 | <0.001 | 0.57 | 0.47, 0.70 | <0.001 | 0.59 | 0.48, 0.72 | <0.001 |
| Seniors (65+ years) | 0.79 | 0.29, 2.16 | 0.6 | 0.82 | 0.30, 2.25 | 0.7 | 0.72 | 0.26, 1.97 | 0.5 | 0.85 | 0.31, 2.35 | 0.8 |
| AIC | 3,854 |  |  | 3,849 |  |  | 3,856 |  |  | 3,857 |  |  |
| BIC | 3,941 |  |  | 3,920 |  |  | 3,940 |  |  | 3,940 |  |  |
| PR = Prevalence Ratio, CI = Confidence Interval | | | | | | | | | | | | |

|  | **Model 5** | | | **Model 6** | | | **Model 7** | | | **Model 8** | | | **Model 9** | | |
| --- | --- | --- | --- | --- | --- | --- | --- | --- | --- | --- | --- | --- | --- | --- | --- |
| **Characteristic** | **PR** | **95% CI** | **p-value** | **PR** | **95% CI** | **p-value** | **PR** | **95% CI** | **p-value** | **PR** | **95% CI** | **p-value** | **PR** | **95% CI** | **p-value** |
| **WHO region** |  |  |  |  |  |  |  |  |  |  |  |  |  |  |  |
| Americas | — | — |  |  |  |  | — | — |  | — | — |  | — | — |  |
| Europe | 1.39 | 1.06, 1.84 | 0.017 |  |  |  | 1.29 | 0.97, 1.72 | 0.085 | 0.91 | 0.67, 1.23 | 0.5 | 1.69 | 1.27, 2.26 | <0.001 |
| Western Pacific | 0.17 | 0.10, 0.30 | <0.001 |  |  |  | 0.13 | 0.07, 0.22 | <0.001 | 0.24 | 0.13, 0.45 | <0.001 | 0.23 | 0.13, 0.41 | <0.001 |
| Africa | 1.35 | 0.76, 2.40 | 0.3 |  |  |  | 0.98 | 0.54, 1.80 | >0.9 | 4.26 | 2.73, 6.64 | <0.001 | 2.10 | 1.15, 3.86 | 0.016 |
| South-East Asia | 1.43 | 0.81, 2.54 | 0.2 |  |  |  | 1.14 | 0.62, 2.08 | 0.7 | 4.21 | 2.46, 7.20 | <0.001 | 1.47 | 0.80, 2.69 | 0.2 |
| Eastern Mediterranean | 1.76 | 1.13, 2.74 | 0.012 |  |  |  | 1.86 | 1.16, 2.96 | 0.009 | 2.45 | 1.47, 4.06 | <0.001 | 1.86 | 1.16, 2.96 | 0.010 |
| **Income class** |  |  |  |  |  |  |  |  |  |  |  |  |  |  |  |
| High income country | — | — |  | — | — |  | — | — |  |  |  |  | — | — |  |
| Low income country | 6.34 | 3.06, 13.2 | <0.001 | 8.01 | 4.63, 13.9 | <0.001 | 4.12 | 1.93, 8.82 | <0.001 |  |  |  | 7.74 | 3.58, 16.8 | <0.001 |
| Lower middle income country | 7.25 | 4.42, 11.9 | <0.001 | 8.19 | 5.62, 11.9 | <0.001 | 5.75 | 3.43, 9.63 | <0.001 |  |  |  | 7.67 | 4.55, 12.9 | <0.001 |
| Upper middle income country | 3.81 | 2.77, 5.23 | <0.001 | 3.21 | 2.34, 4.39 | <0.001 | 3.87 | 2.77, 5.40 | <0.001 |  |  |  | 3.59 | 2.57, 5.02 | <0.001 |
| **Geographic scope** |  |  |  |  |  |  |  |  |  |  |  |  |  |  |  |
| National | — | — |  | — | — |  | — | — |  | — | — |  | — | — |  |
| Subnational | 1.32 | 1.06, 1.65 | 0.012 | 1.26 | 1.01, 1.58 | 0.041 | 1.39 | 1.11, 1.75 | 0.005 | 1.32 | 1.02, 1.70 | 0.033 | 1.23 | 0.98, 1.55 | 0.080 |
| Cases per 100 pop. | 1.37 | 1.28, 1.47 | <0.001 | 1.43 | 1.34, 1.54 | <0.001 |  |  |  | 1.37 | 1.27, 1.47 | <0.001 | 1.82 | 1.71, 1.93 | <0.001 |
| **Transmission phase** |  |  |  |  |  |  |  |  |  |  |  |  |  |  |  |
| Before peak of 1st wave | — | — |  | — | — |  | — | — |  | — | — |  |  |  |  |
| After peak of 1st wave | 1.40 | 1.26, 1.57 | <0.001 | 1.45 | 1.30, 1.62 | <0.001 | 1.56 | 1.39, 1.74 | <0.001 | 1.39 | 1.24, 1.56 | <0.001 |  |  |  |
| After peak of 2nd wave | 3.38 | 2.84, 4.03 | <0.001 | 3.46 | 2.90, 4.12 | <0.001 | 5.04 | 4.31, 5.89 | <0.001 | 3.36 | 2.80, 4.02 | <0.001 |  |  |  |
| **PHSM index (0-10)** | 0.92 | 0.83, 1.01 | 0.082 | 0.88 | 0.80, 0.98 | 0.024 | 1.06 | 0.96, 1.17 | 0.2 | 1.03 | 0.93, 1.14 | 0.6 | 0.92 | 0.83, 1.02 | 0.10 |
| **Sample frame** |  |  |  |  |  |  |  |  |  |  |  |  |  |  |  |
| Household and community samples | — | — |  | — | — |  | — | — |  | — | — |  | — | — |  |
| Blood donors | 1.11 | 0.83, 1.49 | 0.5 | 1.04 | 0.75, 1.43 | 0.8 | 1.13 | 0.83, 1.54 | 0.4 | 0.91 | 0.66, 1.26 | 0.6 | 1.09 | 0.80, 1.48 | 0.6 |
| Residual sera | 1.06 | 0.82, 1.37 | 0.6 | 0.92 | 0.70, 1.21 | 0.6 | 1.07 | 0.81, 1.40 | 0.6 | 0.78 | 0.59, 1.04 | 0.085 | 1.13 | 0.86, 1.48 | 0.4 |
| Pregnant or parturient women | 0.97 | 0.47, 1.99 | >0.9 | 0.53 | 0.24, 1.19 | 0.12 | 1.30 | 0.61, 2.78 | 0.5 | 0.57 | 0.25, 1.32 | 0.2 | 0.69 | 0.32, 1.48 | 0.3 |
| Multiple general populations | 1.63 | 0.61, 4.37 | 0.3 | 2.21 | 0.73, 6.63 | 0.2 | 2.19 | 0.77, 6.24 | 0.14 | 1.27 | 0.40, 3.95 | 0.7 | 2.24 | 0.78, 6.39 | 0.13 |
| **Age** |  |  |  |  |  |  |  |  |  |  |  |  |  |  |  |
| Adults (18-64 years) |  |  |  | — | — |  | — | — |  | — | — |  | — | — |  |
| Multiple groups |  |  |  | 0.83 | 0.77, 0.91 | <0.001 | 0.87 | 0.80, 0.94 | <0.001 | 0.85 | 0.78, 0.93 | <0.001 | 0.84 | 0.77, 0.91 | <0.001 |
| Children and Youth (0-17 years) |  |  |  | 0.57 | 0.46, 0.70 | <0.001 | 0.59 | 0.48, 0.72 | <0.001 | 0.59 | 0.48, 0.72 | <0.001 | 0.58 | 0.47, 0.71 | <0.001 |
| Seniors (65+ years) |  |  |  | 0.95 | 0.31, 2.90 | >0.9 | 0.79 | 0.28, 2.24 | 0.7 | 0.95 | 0.30, 3.02 | >0.9 | 0.82 | 0.29, 2.36 | 0.7 |
| AIC | 3,881 |  |  | 3,903 |  |  | 3,939 |  |  | 3,931 |  |  | 4,059 |  |  |
| BIC | 3,956 |  |  | 3,970 |  |  | 4,023 |  |  | 4,007 |  |  | 4,139 |  |  |
| PR = Prevalence Ratio, CI = Confidence Interval | | | | | | | | | | | | | | | |

### S5 References for all included studies

1.	Sachathep K, Kainne E, Cecile BA, Harris T, Pottinger Y, Longwe H, et al. Seroprevalence of SARS-CoV-2 in 10 Regional Capitals of Cameroon. Zenodo; 2021. doi:[10.5281/ZENODO.5680032](https://doi.org/10.5281/ZENODO.5680032)

2.	Tadesse EB, Endris AA, Solomon H, Alayu M, Kebede A, Eshetu K, et al. Seroprevalence and risk factors for SARS-CoV-2 Infection in selected urban areas in Ethiopia: A cross-sectional evaluation during July 2020. International Journal of Infectious Diseases. 2021;111: 179–185. doi:[10.1016/j.ijid.2021.08.028](https://doi.org/10.1016/j.ijid.2021.08.028)

3.	Adegnika AA, Ngoungou EB, Abuaku B, Essone Ndong P, Djoba Siawaya JF, Lekana-Douki, Jean Bernard, et al. Seroepidemiological survey of coronavirus 2019 (COVID-19) infection in the general population in Gabon. Zenodo; 2022. doi:[10.5281/zenodo.6354737](https://doi.org/10.5281/zenodo.6354737)

4.	Donkor IO, Akorli J, Abuaku B, Opoku M, Andoh NE, Mensah SK, et al. SARS CoV-2 Seroprevalence in Rural and Urban Populations In Ghana. Zenodo; 2021. doi:[10.5281/ZENODO.5771207](https://doi.org/10.5281/ZENODO.5771207)

5.	Uyoga S, Adetifa IMO, Otiende M, Yegon C, Agweyu A, Warimwe GM, et al. Prevalence of SARS-CoV-2 Antibodies From a National Serosurveillance of Kenyan Blood Donors, January-March 2021. JAMA. 2021;326: 1436–1438. doi:[10.1001/jama.2021.15265](https://doi.org/10.1001/jama.2021.15265)

6.	Adetifa IMO, Uyoga S, Gitonga JN, Mugo D, Otiende M, Nyagwange J, et al. Temporal trends of SARS-CoV-2 seroprevalence during the first wave of the COVID-19 epidemic in Kenya. Nature Communications. 2021;12: 3966. doi:[10.1038/s41467-021-24062-3](https://doi.org/10.1038/s41467-021-24062-3)

7.	Mandolo J, Msefula J, Henrion M, Brown C, Moyo B, Samon A, et al. Data from: SARS-CoV-2 exposure in Malawian blood donors: An analysis of seroprevalence and variant dynamics between January 2020 and July 2021. Zenodo; 2021. doi:[10.5281/ZENODO.5644207](https://doi.org/10.5281/ZENODO.5644207)

8.	Kagoli MM, Banda EC, Mwale A, Divala T, Mankhamba R, Chiwaula M, et al. Community and Facility-based COVID-19 Surveillance in Selected Districts of Malawi. Zenodo; 2021. doi:[10.5281/ZENODO.5711715](https://doi.org/10.5281/ZENODO.5711715)

9.	Kolawole O, Tomori O, Agbonlahor D, Ekanem E, Bakare R, Nasidi A, et al. Population-based Age-stratified Seroprevalence Investigation on SARS-CoV-2 Virus in Selected States of High and Low Burden of Disease in Nigeria. Zenodo; 2021. doi:[10.5281/ZENODO.5644223](https://doi.org/10.5281/ZENODO.5644223)

10.	Talla C, Loucoubar C, Roka JL, Barry A, Ndiaye S, Diarra M, et al. Seroprevalence of Anti-SARS-CoV-2 Antibodies in Senegal: A National Population-Based Cross-Sectional Survey, between October and November 2020. SSRN. 2021. doi:[10.2139/ssrn.3925475](https://doi.org/10.2139/ssrn.3925475)

11.	Barrie MB, Lakoh S, Kelly JD, Kanu JS, Squire J, Koroma Z, et al. SARS-CoV-2 antibody prevalence in Sierra Leone, March 2021: A cross-sectional, nationally representative, age-stratified serosurvey. medRxiv. 2021. doi:[10.1101/2021.06.27.21259271](https://doi.org/10.1101/2021.06.27.21259271)

12.	Wolter N, Tempia S, von Gottberg A, Bhiman JN, Walaza S, Kleynhans J, et al. Seroprevalence of SARS-CoV-2 after the second wave in South Africa in HIV-infected and uninfected persons: A cross-sectional household survey. Clinical Infectious Diseases. 2022. doi:[10.1093/cid/ciac198](https://doi.org/10.1093/cid/ciac198)

13.	Vermeulen M. Prevalence of anti-SARS-CoV-2 antibodies among blood donors in South Africa during the period January-May 2021 | Research Square. https://www.researchsquare.com/article/rs-690372/v1; 2021.

14.	Ekouevi DK, Gbeasor-Komlanvi, Fifonsi, Malou-Adom, Vicky, Sadio A, Konu YR, Tchankoni M, et al. Seroprevalence of SARS-CoV-2 infection in the general population in Togo in 2021. Zenodo; 2022. doi:[10.5281/ZENODO.6519820](https://doi.org/10.5281/ZENODO.6519820)

15.	Bosa, Henry Kyobe, Ario, Alex R., Wamala, Robert, Makumbi, Issa, Ssennono, Vincent, Wayengera, Misaki, et al. Population-based, Age- and Gender- Stratified Sero-Survey Study for SARS-CoV-2 in Uganda. Zenodo; 2022. doi:[10.5281/ZENODO.5961443](https://doi.org/10.5281/ZENODO.5961443)

16.	Hines JZ, Fwoloshi S, Kampamba D, Barradas DT, Banda D, Zulu JE, et al. SARS-CoV-2 Prevalence among Outpatients during Community Transmission, Zambia, July 2020. Emerging Infectious Diseases. 2021;27: 2166–2168. doi:[10.3201/eid2708.210502](https://doi.org/10.3201/eid2708.210502)

17.	Mulenga LB, Hines JZ, Fwoloshi S, Chirwa L, Siwingwa M, Yingst S, et al. Prevalence of SARS-CoV-2 in six districts in Zambia in July, 2020: A cross-sectional cluster sample survey. The Lancet Global Health. 2021;0. doi:[10.1016/S2214-109X(21)00053-X](https://doi.org/10.1016/S2214-109X(21)00053-X)

18.	Barros FC, Hartwig FP, Barros AJD, Menezes AMB, Horta BL, Struchiner CJ, et al. COVID-19 and social distancing among children and adolescents in Brazil. Revista De Saude Publica. 2021;55: 42. doi:[10.11606/s1518-8787.2021055003832](https://doi.org/10.11606/s1518-8787.2021055003832)

19.	Mercado-Reyes M, Malagón-Rojas J, Rodríguez-Barraquer I, Zapata-Bedoya S, Wiesner M, Cucunubá Z, et al. Seroprevalence of anti-SARS-CoV-2 antibodies in Colombia, 2020: A population-based study. The Lancet Regional Health - Americas. 2022;9: 100195. doi:[10.1016/j.lana.2022.100195](https://doi.org/10.1016/j.lana.2022.100195)

20.	Paulino-Ramirez R, Báez AA, Degaudenzi AV, Tapia L. Seroprevalence of Specific Antibodies against SARS-CoV-2 from Hotspot Communities in the Dominican Republic. The American Journal of Tropical Medicine and Hygiene. 2020;103: 2343–2346. doi:[10.4269/ajtmh.20-0907](https://doi.org/10.4269/ajtmh.20-0907)

21.	Rene Mejía Nùñez M, Lara J Ángel, Araujo Pleitez S, Ariel Ramírez G. Vista de Vigilancia seroepidemiológica de la circulación del SARS-CoV-2 en 41 municipios de Honduras sin reporte de casos activos, COVID-19, del 16- 23 junio 2020. Población y Salud en Mesoamérica. Revista Población y Salud en Mesoamérica. 2021;18. doi:[10.15517/psm.v18i2.43261](https://doi.org/10.15517/psm.v18i2.43261)

22.	Fernández-Rojas MA, Luna-Ruiz Esparza MA, Campos-Romero A, Calva-Espinosa DY, Moreno-Camacho JL, Mendlovic F, et al. Seroconversion dynamic and SARS-CoV-2 seropositivity in unvaccinated population during the first and second outbreaks in Mexico. Scientific Reports. 2022;12: 5241. doi:[10.1038/s41598-022-09395-3](https://doi.org/10.1038/s41598-022-09395-3)

23.	Canto-Osorio F, Stern D, Pérez-Ferrer C, Arias-de la Garza E, Danon L, Barrientos-Gutiérrez T, et al. Seroprevalencia de SARS-CoV-2 en niños y adolescentes mexicanos en edad escolar. Salud Pública de México. 2021;63: 803–806. doi:[10.21149/12847](https://doi.org/10.21149/12847)

24.	Basto-Abreu A, Carnalla M, Torres-Ibarra L, Romero-Martínez M, Martínez-Barnetche J, López-Martínez I, et al. Nationally representative SARS-CoV-2 antibody prevalence estimates after the first epidemic wave in Mexico. Nature Communications. 2022;13: 589. doi:[10.1038/s41467-022-28232-9](https://doi.org/10.1038/s41467-022-28232-9)

25.	Canadian Blood Services. COVID-19 Seroprevalence Brief Report Report #21A: April 1-15, 2022, Survey. 2022 May.

26.	Canadian Blood Services. COVID-19 Seroprevalence Report March 11th, 2022 Report #18: January 2022 Survey The advance of Omicron. 2022 Mar.

27.	Canadian Blood Services. COVID-19 Seroprevalence Report January 3rd, 2022 Report #16 - December 2021 Survey The advance of Omicron. 2022 Feb.

28.	Canadian Blood Services. COVID-19 Seroprevalence Report January 3rd, 2022 Report #16 - November 2021 Survey Tracking seroprevalence in the vaccine era. 2022 Jan.

29.	Canadian Blood Services. COVID-19 Seroprevalence Report January 3rd, 2022 Report #15 - October 2021 Survey Tracking seroprevalence in the vaccine era. 2022 Jan.

30.	Canadian Blood Services. COVID-19 Seroprevalence Report November 30th, 2021 Report #14 - September 2021 Survey Tracking seroprevalence in the vaccine era. 2021 Nov.

31.	Canadian Blood Services. COVID-19 Seroprevalence Report October 13, 2021 Report #13 - August 2021 Survey Tracking seroprevalence in the vaccine era. 2021 Oct.

32.	Canadian Blood Services. COVID-19 Seroprevalence Report October 13, 2021 Report #12 - July 2021 Survey Tracking seroprevalence in the vaccine era. 2021 Oct.

33.	Canadian Blood Services. COVID-19 Seroprevalence Report August 30, 2021 Report #11 - June 2021 Survey Tracking seroprevalence in the vaccine era. 2021 Aug.

34.	Canadian Blood Services. COVID-19 Seroprevalence Report July 27, 2021 Report #10 - May 2021 Survey Tracking seroprevalence in the vaccine era. 2021 Jul.

35.	Canadian Blood Services. COVID-19 Seroprevalence Report June 30, 2021 Report #9 - April 2021 Survey Tracking seroprevalence in the vaccine era. 2021 Jun.

36.	Canadian Blood Services. COVID-19 Seroprevalence Report June 9, 2021 Report #8 - March 2021 Survey Tracking seroprevalence in the vaccine era. 2021 Jun.

37.	Canadian Blood Services. COVID-19 Seroprevalence Report May 10, 2021 Report #7 - January 2021 Survey Monitoring immunity in the vaccine era. 2021 May.

38.	Canadian Blood Services. COVID-19 Seroprevalence Report 4 January 12, 2021. 2021 Jan.

39.	Saeed S, Drews SJ, Pambrun C, Yi Q-L, Osmond L, O’Brien SF. SARS-CoV-2 seroprevalence among blood donors after the first COVID-19 wave in Canada. Transfusion. 2021;61: 862–872. doi:[10.1111/trf.16296](https://doi.org/10.1111/trf.16296)

40.	Vial PA, González C, Icaza G, Ramirez-Santana M, Quezada-Gaete R, Nuñez-Franz L, et al. Seroprevalence, Spatial Distribution, and Social Determinants of SARS-CoV-2 in Three Urban Centers of Chile. SSRN Electronic Journal. 2021. doi:[10.2139/ssrn.3933650](https://doi.org/10.2139/ssrn.3933650)

41.	Dodd RY, Notari EP, Brodsky JP, Foster GA, Xu M, Saá P, et al. Patterns of antibody response to SARS-CoV-2 among 1.6 million blood donors: Impact of vaccination, United States December 2020 June 2021. The Journal of Infectious Diseases. 2022;225: 5–9. doi:[10.1093/infdis/jiab514](https://doi.org/10.1093/infdis/jiab514)

42.	Jones JM, Stone M, Sulaeman H, Fink RV, Dave H, Levy ME, et al. Estimated US Infection- and Vaccine-Induced SARS-CoV-2 Seroprevalence Based on Blood Donations, July 2020-May 2021. JAMA. 2021;326: 1400–1409. doi:[10.1001/jama.2021.15161](https://doi.org/10.1001/jama.2021.15161)

43.	Sullivan PS, Siegler AJ, Shioda K, Hall EW, Bradley H, Sanchez T, et al. Severe Acute Respiratory Syndrome Coronavirus 2 Cumulative Incidence, United States, August 2020 2020. Clinical Infectious Diseases. 2022;74: 1141–1150. doi:[10.1093/cid/ciab626](https://doi.org/10.1093/cid/ciab626)

44.	Anand S, Montez-Rath M, Han J, Bozeman J, Kerschmann R, Beyer P, et al. Prevalence of SARS-CoV-2 antibodies in a large nationwide sample of patients on dialysis in the USA: A cross-sectional study. The Lancet. 2020;396: 1335–1344. doi:[10.1016/S0140-6736(20)32009-2](https://doi.org/10.1016/S0140-6736(20)32009-2)

45.	Khalagi K, Gharibzadeh S, Khalili D, Mansournia MA, Samiee SM, Aghamohamadi S, et al. Prevalence of COVID-19 in Iran: Results of the first survey of the Iranian COVID-19 Serological Surveillance programme. Clinical Microbiology and Infection. 2021;27: 1666–1671. doi:[10.1016/j.cmi.2021.06.002](https://doi.org/10.1016/j.cmi.2021.06.002)

46.	Poustchi H, Darvishian M, Mohammadi Z, Shayanrad A, Delavari A, Bahadorimonfared A, et al. SARS-CoV-2 antibody seroprevalence in the general population and high-risk occupational groups across 18 cities in Iran: A population-based cross-sectional study. The Lancet Infectious Diseases. 2021;21: 473–481. doi:[10.1016/S1473-3099(20)30858-6](https://doi.org/10.1016/S1473-3099(20)30858-6)

47.	Bellizzi S, Alsawalha L, Sheikh Ali S, Sharkas G, Muthu N, Ghazo M, et al. A three-phase population based sero-epidemiological study: Assessing the trend in prevalence of SARS-CoV-2 during COVID-19 pandemic in Jordan. One Health. 2021;13: 100292. doi:[10.1016/j.onehlt.2021.100292](https://doi.org/10.1016/j.onehlt.2021.100292)

48.	Hoballah A, El Haidari R, Siblany G, Abdel Sater F, Mansour S, Hassan H, et al. SARS-CoV-2 antibody seroprevalence in Lebanon: Findings from the first nationwide serosurvey. BMC Infectious Diseases. 2022;22: 42. doi:[10.1186/s12879-022-07031-z](https://doi.org/10.1186/s12879-022-07031-z)

49.	Al-Abri SS, Al-Wahaibi A, Al-Kindi H, Kurup PJ, Al-Maqbali A, Al-Mayahi Z, et al. SARS-COV-2 antibody seroprevalence in the general population of Oman: Results from four successive nationwide seroepidemiological surveys. International Journal of Infectious Diseases. 2021;112: 269–277. doi:[10.1016/j.ijid.2021.09.062](https://doi.org/10.1016/j.ijid.2021.09.062)

50.	Ahmad AM, Shahzad K, Masood M, Umar M, Abbasi F, Hafeez A. COVID-19 seroprevalence in Pakistan: A cross-sectional study. BMJ Open. 2022;12: e055381. doi:[10.1136/bmjopen-2021-055381](https://doi.org/10.1136/bmjopen-2021-055381)

51.	Haq M, Rehman A, Ahmad J, Zafar U, Ahmed S, Khan MA, et al. SARS-CoV-2: Big seroprevalence data from Pakistanis herd immunity at hand? Infection. 2021;49: 983–988. doi:[10.1007/s15010-021-01629-2](https://doi.org/10.1007/s15010-021-01629-2)

52.	Coyle PV, Kahlout RAE, Dargham SR, Chemaitelly H, Kacem MABH, Al-Mawlawi NHA, et al. Assessing the performance of a serological point-of-care test in measuring detectable antibodies against SARS-CoV-2. PLoS One. 2022;17: e0262897. doi:[10.1371/journal.pone.0262897](https://doi.org/10.1371/journal.pone.0262897)

53.	Coyle PV, Chemaitelly H, Ben Hadj Kacem MA, Abdulla Al Molawi NH, El Kahlout RA, Gilliani I, et al. SARS-CoV-2 seroprevalence in the urban population of Qatar: An analysis of antibody testing on a sample of 112,941 individuals. iScience. 2021;24: 102646. doi:[10.1016/j.isci.2021.102646](https://doi.org/10.1016/j.isci.2021.102646)

54.	Zhussupov B, Kaidar E, Suleimenova Z, Aleksandrova N, Smagul M, Nusupbayeva G, et al. Population-based age-stratified seroepidemiological investigation protocol for COVID-19 virus infection, Republic of Kazakhstan. Zenodo; 2021. doi:[10.5281/ZENODO.5646794](https://doi.org/10.5281/ZENODO.5646794)

55.	Popova AY, Kasymov OT, Smolenski VY, Egorova SA, Nurmatov ZS, Milichkina AM, et al. SARS-CoV-2 Herd Immunity of the Kyrgyz Population in 2021. Research Square. 2021. doi:[10.21203/rs.3.rs-1138128/v1](https://doi.org/10.21203/rs.3.rs-1138128/v1)

56.	Kuchuk T, Nurmatov Z, Kasymbekova K. Population-based age-stratified seroepidemiological investigation protocol for coronavirus 2019 (COVID-19) infection in Kyrgyz Republic. Zenodo; 2021. doi:[10.5281/ZENODO.5645779](https://doi.org/10.5281/ZENODO.5645779)

57.	Ceban A, Gheorghita S. Serological surveillance of COVID-19 in the general population of Moldova. Zenodo; 2021. doi:[10.5281/ZENODO.5645781](https://doi.org/10.5281/ZENODO.5645781)

58.	R. A. R, N. S. I, A. S. K, M. A. A, S. SS, L. M. L, et al. FORMATION OF HERD IMMUNITY TO SARS-CoV-2 IN THE REGIONS OF UZBEKISTAN. European Journal of Molecular & Clinical Medicine. 2021;8: 574–581.

59.	Royo-Cebrecos C, Vilanova D, López J, Arroyo V, Pons M, Francisco G, et al. Mass SARS-CoV-2 serological screening, a population-based study in the Principality of Andorra. The Lancet Regional Health - Europe. 2021;5: 100119. doi:[10.1016/j.lanepe.2021.100119](https://doi.org/10.1016/j.lanepe.2021.100119)

60.	Herzog SA, Bie JD, Abrams S, Wouters I, Ekinci E, Patteet L, et al. Seroprevalence of IgG antibodies against SARS-CoV-2 a serial prospective cross-sectional nationwide study of residual samples, Belgium, March to October 2020. Eurosurveillance. 2022;27: 2100419. doi:[10.2807/1560-7917.ES.2022.27.9.2100419](https://doi.org/10.2807/1560-7917.ES.2022.27.9.2100419)

61.	Fogh K, Eriksen AR, Hasselbalch RB, Kristensen ES, Bundgaard H, Nielsen SD, et al. Seroprevalence of SARS-CoV-2 antibodies in social housing areas in Denmark. Epidemiology; 2021 May. doi:[10.1101/2021.05.07.21256725](https://doi.org/10.1101/2021.05.07.21256725)

62.	Espenhain L, Tribler S, Sværke Jørgensen C, Holm Hansen C, Wolff Sönksen U, Ethelberg S. Prevalence of SARS-CoV-2 antibodies in Denmark: Nationwide, population-based seroepidemiological study. European Journal of Epidemiology. 2021;36: 715–725. doi:[10.1007/s10654-021-00796-8](https://doi.org/10.1007/s10654-021-00796-8)

63.	Fogh K, Strange JE, Scharff BF, Eriksen AR, Hasselbalch RB, Bundgaard H, et al. Testing Denmark: A Danish nationwide surveillance study of COVID-19. medRxiv. 2021. doi:[10.1101/2021.08.10.21261777](https://doi.org/10.1101/2021.08.10.21261777)

64.	Pedersen OB, Nissen J, Dinh KM, Schwinn M, Kaspersen KA, Boldsen JK, et al. Severe Acute Respiratory Syndrome Coronavirus 2 (SARS-CoV-2) Infection Fatality Rate Among Elderly Danes: A Cross-sectional Study on Retired Blood Donors. Clinical Infectious Diseases. 2021;73: e2962–e2969. doi:[10.1093/cid/ciaa1627](https://doi.org/10.1093/cid/ciaa1627)

65.	Erikstrup C, Hother CE, Pedersen OBV, Mølbak K, Skov RL, Holm DK, et al. Estimation of SARS-CoV-2 infection fatality rate by real-time antibody screening of blood donors. medRxiv. 2020. doi:[10.1101/2020.04.24.20075291](https://doi.org/10.1101/2020.04.24.20075291)

66.	Melin M, Palmu A. Report of THL serological population study of the coronavirus epidemic. thl. https://www.thl.fi/roko/cov-vaestoserologia/sero_report_weekly_en.html; 2021.

67.	Warszawski J, Beaumont A-L, Seng R, de Lamballerie X, Rahib D, Lydié N, et al. Prevalence of SARS-Cov-2 antibodies and living conditions: The French national random population-based EPICOV cohort. BMC Infectious Diseases. 2022;22: 41. doi:[10.1186/s12879-021-06973-0](https://doi.org/10.1186/s12879-021-06973-0)

68.	Le Vu S, Jones G, Anna F, Rose T, Richard J-B, Bernard-Stoecklin S, et al. Prevalence of SARS-CoV-2 antibodies in France: Results from nationwide serological surveillance. Nature Communications. 2021;12: 3025. doi:[10.1038/s41467-021-23233-6](https://doi.org/10.1038/s41467-021-23233-6)

69.	Fuest C, Neumeier F, Peichl A, Charité - Universitätsmedizin Berlin, Forsa Gesellschaft für Sozialforschung und statistische Analysen mbH, GmbH PHS. Die Deutschen und Corona. ifo Institut; 2020 Nov.

70.	Gornyk D, Harries M, Glöckner S, Strengert M, Kerrinnes T, Bojara G, et al. SARS-CoV-2 Seroprevalence in Germany - A Population Based Sequential Study in Five Regions. SSRN. 2021.

71.	Koureas M, Bogogiannidou Z, Vontas A, Kyritsi MA, Mouchtouri VA, Dadouli K, et al. SARS-CoV-2 Sero-Surveillance in Greece: Evolution over Time and Epidemiological Attributes during the Pre-Vaccination Pandemic Era. Diagnostics. 2022;12: 295. doi:[10.3390/diagnostics12020295](https://doi.org/10.3390/diagnostics12020295)

72.	Ralli M, Arcangeli A, Soave PM, Voglino MC, De-Giorgio F. SARS-CoV-2 seroprevalence in the Vatican City State. European Journal of Internal Medicine. 2021;0. doi:[10.1016/j.ejim.2021.01.029](https://doi.org/10.1016/j.ejim.2021.01.029)

73.	Merkely B, Szabó AJ, Kosztin A, Berényi E, Sebestyén A, Lengyel C, et al. Novel coronavirus epidemic in the Hungarian population, a cross-sectional nationwide survey to support the exit policy in Hungary. GeroScience. 2020;42: 1063–1074. doi:[10.1007/s11357-020-00226-9](https://doi.org/10.1007/s11357-020-00226-9)

74.	Indenbaum V, Lustig Y, Mendelson E, Hershkovitz Y, Glatman-Freedman A, Keinan-Boker L, et al. Under-diagnosis of SARS-CoV-2 infections among children aged 0 years, a nationwide seroprevalence study, Israel, January 2020 to March 2021. Eurosurveillance. 2021;26: 2101040. doi:[10.2807/1560-7917.ES.2021.26.48.2101040](https://doi.org/10.2807/1560-7917.ES.2021.26.48.2101040)

75.	Reicher S, Ratzon R, Ben-Sahar S, Hermoni-Alon S, Mossinson D, Shenhar Y, et al. Nationwide seroprevalence of antibodies against SARS-CoV-2 in Israel. European Journal of Epidemiology. 2021;36: 727–734. doi:[10.1007/s10654-021-00749-1](https://doi.org/10.1007/s10654-021-00749-1)

76.	Instituto Nazionale di Statistica. PRIMI RISULTATI DELL’INDAGINE DI SIEROPREVALENZA SUL SARS-CoV-2. Instituto Nazionale di Statistica; 2020 Aug.

77.	Statistics Jersey. SARS-CoV-2 - Prevalence of antibodies in Jersey Community survey round 3. Statistics Jersey; 2020 Jul.

78.	Šmigelskas K, Petrikonis K, Kasiulevičius V, Kalėdienė R, Jakaitienė A, Kaselienė S, et al. SARS-CoV-2 Seroprevalence in Lithuania: Results of National Population Survey. Acta medica Lituanica. 2021;28: 48–58. doi:[10.15388/Amed.2020.28.1.2](https://doi.org/10.15388/Amed.2020.28.1.2)

79.	Snoeck C. Prevalence of SARS-CoV-2 infection in the Luxembourgish population: The CON-VINCE study. Pre-print.

80.	Tunheim G, Rø GØI, Tran T, Kran A-MB, Andersen JT, Vaage EB, et al. Trends in seroprevalence of SARS-CoV-2 and infection fatality rate in the Norwegian population through the first year of the COVID-19 pandemic. Influenza and Other Respiratory Viruses. 2022;16: 204–212. doi:[10.1111/irv.12932](https://doi.org/10.1111/irv.12932)

81.	ISN COVID-19 Group. National Serological Survey COVID-19 (3rd phase). National Institute of Health; 2021.

82.	Castro LC e, Gomes A, Serrano M, Pereira AH, Ribeiro R, Napoleão P, et al. Longitudinal SARS-CoV-2 seroprevalence in Portugal and antibody maintenance 12 months after the start of the COVID-19 pandemic. Research Square. 2021. doi:[10.21203/rs.3.rs-603060/v1](https://doi.org/10.21203/rs.3.rs-603060/v1)

83.	Castro LC e, Pereira AHG, Ribeiro R, Alves C, Veloso L, Vicente V, et al. Early Release - Prevalence of SARS-CoV-2 Antibodies after First 6 Months of COVID-19 Pandemic, Portugal - Volume 27, Number 11 2021 - Emerging Infectious Diseases journal - CDC. 2021. doi:[10.3201/eid2711.210636](https://doi.org/10.3201/eid2711.210636)

84.	Kislaya I, Gonçalves P, Barreto M, Sousa R de, Garcia AC, Matos R, et al. Seroprevalence of SARS-CoV-2 Infection in Portugal in May-July 2020: Results of the First National Serological Survey (ISNCOVID-19). Acta Médica Portuguesa. 2021;34: 87–94. doi:[10.20344/amp.15122](https://doi.org/10.20344/amp.15122)

85.	Poljak M, Oštrbenk Valenčak A, Štrumbelj E, Maver Vodičar P, Vehovar V, Resman Rus K, et al. Seroprevalence of severe acute respiratory syndrome coronavirus 2 in Slovenia: Results of two rounds of a nationwide population study on a probability-based sample, challenges and lessons learned. Clinical Microbiology and Infection. 2021;27: 1039.e1–1039.e7. doi:[10.1016/j.cmi.2021.03.009](https://doi.org/10.1016/j.cmi.2021.03.009)

86.	Pérez-Olmeda M, Saugar JM, Fernández-García A, Pérez-Gómez B, Pollán M, Avellón A, et al. Evolution of antibodies against SARS-CoV-2 over seven months: Experience of the nationwide seroprevalence ENE-COVID study in Spain. Journal of Clinical Virology. 2022;149: 105130. doi:[10.1016/j.jcv.2022.105130](https://doi.org/10.1016/j.jcv.2022.105130)

87.	Ministerio de Salud. ESTUDIO ENE-COVID: INFORME FINAL ESTUDIO NACIONAL DE SERO-EPIDEMIOLOGÍA DE LA INFECCIÓN POR SARS-COV-2 EN ESPAÑA. España: Ministerio de Salud; 2020 Jun.

88.	Prados N, González-Ravina C, Vergara V, Herrero J, Cruz M, Requena A. Risk Factor Analysis For SARS-CoV-2 Seropositivity Within Assisted Reproductive. Fertility and Sterility. 2020;114: e547. doi:[10.1016/j.fertnstert.2020.09.077](https://doi.org/10.1016/j.fertnstert.2020.09.077)

89.	Agency SPH. Påvisning av antikroppar mot SARS-CoV-2 i blodprov från öppenvården.

90.	Agency SPH. Påvisning av antikroppar mot SARS-CoV-2 i blodprov från öppenvården. 2021-07-19.

91.	Agency SPH. Påvisning av antikroppar efter genomgången covid-19 hos blodgivare (Delrapport 2). Swedish Public Health Agency; 2021 Mar.

92.	Scotland PH. Enhanced Surveillance of COVID-19 in Scotland - Population-based seroprevalence surveillance 4 May 2022. Public Health Scotland; 2022 May. doi:[10.52487/83780](https://doi.org/10.52487/83780)

93.	Scotland PH. Enhanced Surveillance of COVID-19 in Scotland - Population-based seroprevalence surveillance 30 March 2022. Public Health Scotland; 2022 Mar.

94.	Scotland PH. Enhanced Surveillance of COVID-19 in Scotland - Population-based seroprevalence surveillance 23 February 2022. Public Health Scotland; 2022 Feb.

95.	Scotland PH. Enhanced Surveillance of COVID-19 in Scotland - Population-based seroprevalence surveillance 19 January 2022. Public Health Scotland; 2022 Jan.

96.	Scotland PH. Enhanced Surveillance of COVID-19 in Scotland - Population-based seroprevalence surveillance 8 December 2021. Public Health Scotland; 2021 Dec.

97.	Oeser C, Whitaker H, Linley E, Borrow R, Tonge S, Brown CS, et al. Large increases in SARS-CoV-2 seropositivity in children in England: Effects of the delta wave and vaccination. Journal of Infection. 2022;84: 418–467. doi:[10.1016/j.jinf.2021.11.019](https://doi.org/10.1016/j.jinf.2021.11.019)

98.	Scotland PH. Enhanced Surveillance of COVID-19 in Scotland - Population-based seroprevalence surveillance 3 November 2021. Public Health Scotland; 2021 Nov.

99.	Scotland PH. Enhanced Surveillance of COVID-19 in Scotland - Population-based seroprevalence surveillance 25 August 2021. Public Health Scotland; 2021 Aug.

100.	Scotland PH. Enhanced Surveillance of COVID-19 in Scotland - Population-based seroprevalence surveillance 21 July 2021. Public Health Scotland; 2021 Jul.

101.	Public health Scotland. Enhanced Surveillance of COVID-19 in Scotland 16 June 2021. Public Health Scotland; 2021 Jun.

102.	Public health Scotland. Enhanced Surveillance of COVID-19 in Scotland 12 May 2021. Public Health Scotland; 2021 May.

103.	Aldridge RW, Pineo H, Fragaszy E, Eyre M, Kovar J, Nguyen V, et al. Household overcrowding and risk of SARS-CoV-2: Analysis of the Virus Watch prospective community cohort study in England and Wales. medRxiv. 2021. doi:[10.1101/2021.05.10.21256912](https://doi.org/10.1101/2021.05.10.21256912)

104.	Whitaker HJ, Elgohari S, Rowe C, Otter AD, Brooks T, Linley E, et al. Impact of COVID-19 vaccination program on seroprevalence in blood donors in England, 2021. Journal of Infection. 2021;0. doi:[10.1016/j.jinf.2021.04.037](https://doi.org/10.1016/j.jinf.2021.04.037)

105.	Public health Scotland. Enhanced Surveillance of COVID-19 in Scotland 7 April 2021. Public Health Scotland; 2021 Apr.

106.	Public health Scotland. Enhanced Surveillance of COVID-19 in Scotland 3 March 2021. Public Health Scotland; 2021 Mar.

107.	Public health Scotland. Enhanced Surveillance of COVID-19 in Scotland 27 January 2021. Public Health Scotland; 2021 Jan.

108.	Whitaker H, Tsang RS, Button E, Andrews N, Byford R, Borrow R, et al. Sociodemographic disparities in COVID-19 seroprevalence across England in the Oxford RCGP primary care sentinel network. Journal of Infection. doi:[10.1016/j.jinf.2022.04.016](https://doi.org/10.1016/j.jinf.2022.04.016)

109.	Ladhani S. Prospective Active National Surveillance of Preschools and Primary Schools for SARS-CoV-2 Infection and Transmission in England, June 2020. Rochester, NY: Social Science Research Network; 2021 Jan. Report No.: ID 3764198.

110.	Public health Scotland. Enhanced Surveillance of COVID-19 in Scotland 23 December 2020. Scotland; 2020 Dec.

111.	England PH. National COVID-19 surveillance report: 28 May 2020 (week 22). Public Health England; 2020 May.

112.	Murhekar MV, Bhatnagar T, Thangaraj JWV, Saravanakumar V, Kumar MS, Selvaraju S, et al. SARS-CoV-2 seroprevalence among the general population and healthcare workers in India, December 2020-January 2021. International journal of infectious diseases: IJID: official publication of the International Society for Infectious Diseases. 2021;108: 145–155. doi:[10.1016/j.ijid.2021.05.040](https://doi.org/10.1016/j.ijid.2021.05.040)

113.	Wahyono TYM, Mahkota R, Ansariadi H, Syahrul F, Hidajah AC, Dwinata I, et al. Population-based Age-Stratified Seroepidemiological Investigation for Coronavirus 2019 (COVID-19) Infection in Indonesia. Zenodo; 2021. doi:[10.5281/ZENODO.5665348](https://doi.org/10.5281/ZENODO.5665348)

114.	Government of Nepal. Enhanced Surveillance on Sero-Prevalence of SARS-CoV-2 In General Population. Government of Nepal; 2021 Apr.

115.	Nakphook S, Iamsirithavorn S, Puthavathana P Pilaipan, Lerdsamran H, Prasertsophon, Chanma T, et al. Age-stratified sero-surveillance for SARS-CoV-2 infections in four cities in Thailand. Zenodo; 2021. doi:[10.5281/ZENODO.5703307](https://doi.org/10.5281/ZENODO.5703307)

116.	Vette KM, Machalek DA, Gidding HF, Nicholson S, O’Sullivan MVN, Carlin JB, et al. Seroprevalence of Severe Acute Respiratory Syndrome Coronavirus 2-Specific Antibodies in Australia After the First Epidemic Wave in 2020: A National Survey. Open Forum Infectious Diseases. 2022;9: ofac002. doi:[10.1093/ofid/ofac002](https://doi.org/10.1093/ofid/ofac002)

117.	Coatsworth N, Myles PS, Mann GJ, Cockburn IA, Forbes AB, Gardiner EE, et al. Prevalence of asymptomatic SARS-CoV-2 infection in elective surgical patients in Australia: A prospective surveillance study. ANZ Journal of Surgery. 2021;91: 27–32. doi:[10.1111/ans.16564](https://doi.org/10.1111/ans.16564)

118.	Li Z, Guan X, Mao N, Luo H, Qin Y, He N, et al. Antibody seroprevalence in the epicenter Wuhan, Hubei, and six selected provinces after containment of the first epidemic wave of COVID-19 in China. The Lancet Regional Health Western Pacific. 2021;8: 100094. doi:[10.1016/j.lanwpc.2021.100094](https://doi.org/10.1016/j.lanwpc.2021.100094)

119.	Yoshiyama T, Saito Y, Masuda K, Nakanishi Y, Kido Y, Uchimura K, et al. Prevalence of SARS-CoV-2, Japan, June 2020. Emerging Infectious Diseases. 2021;27: 628–631. doi:[10.3201/eid2702.204088](https://doi.org/10.3201/eid2702.204088)

120.	Zhuo Lin C, Hatta Bin Abdul Mutalip M, Ani binti Ahmad N. POPULATION-BASED SEROPREVALENCE SURVEY OF COVID-19 VIRUS INFECTION IN MALAYSIA. Zenodo; 2021. doi:[10.5281/ZENODO.5645887](https://doi.org/10.5281/ZENODO.5645887)

121.	Chimeddorj B, Nyamadawa P, Davaalkham D, Le L-V, Ya A, Khurelbaatar N, et al. Novel Coronavirus (COVID-19) Population-Based Age-Stratified Sero-Epidemiological Study in Mongolia (A Prospective, Nationwide Study). Zenodo; 2021. doi:[10.5281/ZENODO.5646781](https://doi.org/10.5281/ZENODO.5646781)

122.	Lavu E, Mapira P, Ila R, Vellemu E, Maha A, Menda L, et al. Seroprevalence of COVID-19 in Papua New Guinea. Zenodo; 2021. doi:[10.5281/ZENODO.5703345](https://doi.org/10.5281/ZENODO.5703345)

123.	Kim A-R, Minn D, Kim S, Do H, Kim BB, Choi YS, et al. Seroprevalence study of SARS-CoV-2 antibodies in community based on participants in the 2020 Korea National Health and Nutrition Examination Survey (KNHANES). Epidemiology and Health. 2022; e2022028. doi:[10.4178/epih.e2022028](https://doi.org/10.4178/epih.e2022028)

124.	Nah E-H, Cho S, Park H, Hwang I, Cho H-I. Nationwide seroprevalence of antibodies to SARS-CoV-2 in asymptomatic population in South Korea: A cross-sectional study. BMJ Open. 2021;11: e049837. doi:[10.1136/bmjopen-2021-049837](https://doi.org/10.1136/bmjopen-2021-049837)

125.	Makiala-Mandanda, Sheila, Folefack, Gervais Léon, Ahuka, Steve, Nkwembe, Edith, Nsio, Justus, Otshudiema, John, et al. Seroprevalence survey on infection with the SARS-CoV-2 virus after the second wave in Kinshasa, Democratic Republic of the Congo. 2021. Zenodo; 2021. doi:[10.5281/ZENODO.5644148](https://doi.org/10.5281/ZENODO.5644148)

126.	Assefa N, Regassa LD, Teklemariam Z, Oundo J, Madrid L, Dessie Y, et al. Seroprevalence of anti-SARS-CoV-2 antibodies in women attending antenatal care in eastern Ethiopia: A facility-based surveillance. BMJ Open. 2021;11: e055834. doi:[10.1136/bmjopen-2021-055834](https://doi.org/10.1136/bmjopen-2021-055834)

127.	Okpala OV, Dim CC, Ugwu CI, Onyemaechi S, Uchebo O, Chukwulobelu U, et al. Population seroprevalence of SARS-CoV-2 antibodies in Anambra state, South-East, Nigeria. International Journal of Infectious Diseases. 2021;110: 171–178. doi:[10.1016/j.ijid.2021.07.040](https://doi.org/10.1016/j.ijid.2021.07.040)

128.	Ilori E, Stafford K, Steinhardt L, Ihekweazu C, Nwachukwu W, Igumbor E, et al. Nigeria COVID-19 Household Seroprevalence Survey. Zenodo; 2021. doi:[10.5281/ZENODO.5644231](https://doi.org/10.5281/ZENODO.5644231)

129.	Majiya H, Aliyu-Paiko M, Balogu VT, Musa DA, Salihu IM, Kawu AA, et al. Seroprevalence of COVID-19 in Niger State. medRxiv. 2020. doi:[10.1101/2020.08.04.20168112](https://doi.org/10.1101/2020.08.04.20168112)

130.	George JA, Khoza S, Mayne E, Dlamini S, Kone N, Jassat W, et al. Sentinel seroprevalence of SARS-CoV-2 in Gauteng Province, South Africa, August - October 2020. South African Medical Journal. 2021;111: 1078–1083. doi:[10.7196/SAMJ.2021.v111i11.15669](https://doi.org/10.7196/SAMJ.2021.v111i11.15669)

131.	Barros AJD, Victora CG, Menezes AMB, Horta BL, Barros FC, Hartwig FP, et al. Population-level seropositivity trend for SARS-Cov-2 in Rio Grande do Sul, Brazil. Revista de Saúde Pública. 2021;55: 78. doi:[10.11606/s1518-8787.2021055004075](https://doi.org/10.11606/s1518-8787.2021055004075)

132.	Moya Rios do Vale N, Roche Moreira Latini F, Prisco Arnoni C, Martins Parreira R, Batista Castelo Girão MJ, Pereira Cortez AJ, et al. Increasing rate of anti-SARS-CoV-2 antibodies between the first and second waves of COVID-19 in São Paulo, Brazil: A cross-sectional blood donors-based study. Clinics. 2022;77: 100016. doi:[10.1016/j.clinsp.2022.100016](https://doi.org/10.1016/j.clinsp.2022.100016)

133.	Albuquerque JOM de, Kamioka GA, Madalosso G, Costa SA, Ferreira PB, Pino FA, et al. Prevalence evolution of SARS-CoV-2 infection in the city of São Paulo, 2020. Revista de Saúde Pública. 2021;55: 62–62. doi:[10.11606/s1518-8787.2021055003970](https://doi.org/10.11606/s1518-8787.2021055003970)

134.	da Silva AAM. Prevalência da infecção pelo vírus SARS-Cov-2 no Maranhão, Brasil - Fase 2. Universidade Federal do Maranhão; 2020 Nov.

135.	Schneider APH, Gaedke MÂ, Koepp J, Reuter ÉM, Darsie C, Possuelo LG, et al. [Social distancing as protection factor against COVID-19 in a non-metropolitan area in the State of Rio Grande do Sul, BrazilLas medidas de distanciamiento social como factor de protección contra la COVID-19 en el interior de Rio Grande do Sul, Brasil]. Revista Panamericana De Salud Publica = Pan American Journal of Public Health. 2021;45: e145. doi:[10.26633/RPSP.2021.145](https://doi.org/10.26633/RPSP.2021.145)

136.	Hallal PC, Silveira MF, Menezes AMB, Horta BL, Barros AJD, Pellanda LC, et al. Slow Spread of SARS-CoV-2 in Southern Brazil Over a 6-Month Period: Report on 8 Sequential Statewide Serological Surveys Including 35611 Participants. American Journal of Public Health. 2021;111: 1542–1550. doi:[10.2105/AJPH.2021.306351](https://doi.org/10.2105/AJPH.2021.306351)

137.	Goncalves Chaves D, Hiroshi Caldeira Takahashi R, Campelo F, Fernandes da Silva Malta MC, Rocha de Oliveira I, Figueiredo Barbosa-Stancioli E, et al. SARS-CoV-2 IgG Seroprevalence among Blood Donors as a Monitor of the COVID-19 Epidemic, Brazil. Emerging Infectious Diseases. 2022;28: 734–742. doi:[10.3201/eid2804.211961](https://doi.org/10.3201/eid2804.211961)

138.	Maciel ELN, Gomes CC, Almada GL, Medeiros Junior NF de, Cardoso OA, Jabor PM, et al. COVID-19 em crianças, adolescentes e jovens: Estudo transversal no Espírito Santo, 2020. Epidemiologia e Serviços de Saúde. 2021;30: e20201029. doi:[10.1590/s1679-49742021000400001](https://doi.org/10.1590/s1679-49742021000400001)

139.	Concha G, Frickmann H, Oey A, Strengert M, Kreienbrock L, Kann S. Direct and Indirect Proof of SARS-CoV-2 Infections in Indigenous Wiwa Communities in North-Eastern Colombia. Vaccines. 2021;9: 1120. doi:[10.3390/vaccines9101120](https://doi.org/10.3390/vaccines9101120)

140.	Garay E, Serrano-Coll H, Rivero R, Gastelbondo B, Faccini-Martínez Á, Berrocal J, et al. SARS-CoV-2 in eight municipalities of the Colombian tropics: High immunity, clinical and sociodemographic outcomes. Transactions of The Royal Society of Tropical Medicine and Hygiene. 2022;116: 139–147. doi:[10.1093/trstmh/trab094](https://doi.org/10.1093/trstmh/trab094)

141.	Guzman NA, De la Hoz Restrepo F, Serrano-Coll H, Gastelbondo B, Mattar S. Using serological studies to assess COVID-19 infection fatality rate in developing countries. A case study from one Colombian department. International Journal of Infectious Diseases. 2021;110: 4–5. doi:[10.1016/j.ijid.2021.06.018](https://doi.org/10.1016/j.ijid.2021.06.018)

142.	Huamaní C, Velásquez L, Montes S, Mayanga-Herrera A, Bernabé-Ortiz A. SARS-CoV-2 seroprevalence in a high-altitude setting in Peru: Adult population-based cross-sectional study. PeerJ. 2021;9: e12149. doi:[10.7717/peerj.12149](https://doi.org/10.7717/peerj.12149)

143.	Lewin A, De Serres G, Grégoire Y, Perreault J, Drouin M, Fournier M-J, et al. Seroprevalence of SARS-CoV-2 antibodies among blood donors in Québec: An update from a serial cross-sectional study. Canadian Journal of Public Health. 2022;113: 385–393. doi:[10.17269/s41997-022-00622-y](https://doi.org/10.17269/s41997-022-00622-y)

144.	Charlton CL, Nguyen LT, Bailey A, Fenton J, Plitt SS, Marohn C, et al. Pre-Vaccine Positivity of SARS-CoV-2 Antibodies in Alberta, Canada during the First Two Waves of the COVID-19 Pandemic. Bard JD, editor. Microbiology Spectrum. 2021;9: e00291–21. doi:[10.1128/Spectrum.00291-21](https://doi.org/10.1128/Spectrum.00291-21)

145.	Bolotin S, Tran V. COVID-19 Seroprevalence in Ontario; March 27 - October 30, 2020. Zenodo; 2021. doi:[10.5281/ZENODO.5703273](https://doi.org/10.5281/ZENODO.5703273)

146.	Skowronski DM, Sekirov I, Sabaiduc S, Zou M, Morshed M, Lawrence D, et al. Low SARS-CoV-2 sero-prevalence based on anonymized residual sero-survey before and after first wave measures in British Columbia, Canada, March-May 2020. medRxiv. 2020. doi:[10.1101/2020.07.13.20153148](https://doi.org/10.1101/2020.07.13.20153148)

147.	Vial, Pablo, González, Claudia, Icaza, Gloria, Ramirez-Santana, Muriel, Quezada-Gaete, Rubén, Núñez-Franz, Loreto, et al. Serial seroprevalence study: Follow-up of immunity to SARS-Cov-2 infection and monitoring of effective vaccination coverage, in three Chilean cities. Zenodo; 2022. doi:[10.5281/ZENODO.5998971](https://doi.org/10.5281/ZENODO.5998971)

148.	University of Texas Health Science Center, Texas Health and Human Services. Texas Coronavirus Antibody Response Survey - October 2021. Texas; 2021 Oct p. 20.

149.	Mehrotra ML, Lim E, Lamba K, Kamali A, Lai KW, Meza E, et al. CalScope: Monitoring SARS-CoV-2 Seroprevalence from Vaccination and Prior Infection in Adults and Children in California May 2021 July 2021. Epidemiology; 2021 Dec. doi:[10.1101/2021.12.09.21267565](https://doi.org/10.1101/2021.12.09.21267565)

150.	Couture A, Lyons BC, Mehrotra ML, Sosa L, Ezike N, Ahmed FS, et al. Severe Acute Respiratory Syndrome Coronavirus 2 Seroprevalence and Reported Coronavirus Disease 2019 Cases in US Children, August 2020 2021. Open Forum Infectious Diseases. 2022;9: ofac044. doi:[10.1093/ofid/ofac044](https://doi.org/10.1093/ofid/ofac044)

151.	Wong MP, Meas MA, Adams C, Hernandez S, Green V, Montoya M, et al. Development and Implementation of Dried Blood Spot-based COVID-19 Serological Assays for Epidemiologic Studies. Cold Spring Harbor Laboratory Press; 2021. p. 2021.11.25.21266786. doi:[10.1101/2021.11.25.21266786](https://doi.org/10.1101/2021.11.25.21266786)

152.	Kennedy JL, Forrest JC, Young SG, Amick B, Williams M, James L, et al. Temporal Variations in Seroprevalence of SARS-CoV-2 Infections by Race and Ethnicity in Arkansas. medRxiv. 2021. doi:[10.1101/2021.07.15.21260213](https://doi.org/10.1101/2021.07.15.21260213)

153.	Cardenas VM, Kennedy JL, Williams M, Nembhard WN, Zohoori N, Du R, et al. State-wide random seroprevalence survey of SARS-CoV-2 past infection in a southern US State, 2020. PLoS One. 2022;17: 1–12. doi:[10.1371/journal.pone.0267322](https://doi.org/10.1371/journal.pone.0267322)

154.	Pressman A, Lockhart SH, Wilcox J, Smits K, Etzell J, Albeiroti S, et al. COVID-19 in pregnancy by race and ethnicity: Implications for development of a vaccination strategy. Women’s Health. 2021;17: 174550652110633. doi:[10.1177/17455065211063300](https://doi.org/10.1177/17455065211063300)

155.	Levorson RE, Christian E, Hunter B, Sayal J, Sun J, Bruce SA, et al. A cross-sectional investigation of SARS-CoV-2 seroprevalence and associated risk factors in children and adolescents in the United States. PLoS One. 2021;16: e0259823. doi:[10.1371/journal.pone.0259823](https://doi.org/10.1371/journal.pone.0259823)

156.	Lopez CA, Cunningham CH, Pugh S, Brandt K, Vanna UP, Delacruz MJ, et al. Disparities in SARS-CoV-2 seroprevalence among individuals presenting for care in central North Carolina over a six-month period. medRxiv. 2021. doi:[10.1101/2021.03.25.21254320](https://doi.org/10.1101/2021.03.25.21254320)

157.	Adams C, Horton M, Solomon O, Wong M, Wu SL, Fuller S, et al. Impact of individual-level characteristics and transmission mitigation behaviors on SARS-CoV-2 infection and seroprevalence in a large Northern California Bay Area cohort. Epidemiology; 2021 Dec. doi:[10.1101/2021.12.02.21266871](https://doi.org/10.1101/2021.12.02.21266871)

158.	Kugeler KJ, Podewils LJ, Alden NB, Burket TL, Kawasaki B, Biggerstaff BJ, et al. Assessment of SARS-CoV-2 Seroprevalence by Community Survey and Residual Specimens, Denver, Colorado, July 2020. Public Health Reports. 2022;137: 128–136. doi:[10.1177/00333549211055137](https://doi.org/10.1177/00333549211055137)

159.	Rogawski McQuade ET, Guertin KA, Becker L, Operario D, Gratz J, Guan D, et al. Assessment of Seroprevalence of SARS-CoV-2 and Risk Factors Associated With COVID-19 Infection Among Outpatients in Virginia. JAMA Network Open. 2021;4: e2035234. doi:[10.1001/jamanetworkopen.2020.35234](https://doi.org/10.1001/jamanetworkopen.2020.35234)

160.	Mahajan S, Srinivasan R, Redlich CA, Huston SK, Anastasio KM, Cashman L, et al. Seroprevalence of SARS-CoV-2-Specific IgG Antibodies Among Adults Living in Connecticut: Post-Infection Prevalence (PIP) Study. The American Journal of Medicine. 2021;134: 526–534.e11. doi:[10.1016/j.amjmed.2020.09.024](https://doi.org/10.1016/j.amjmed.2020.09.024)

161.	Haizler-Cohen L, Davidov A, Blitz MJ, Fruhman G. Severe acute respiratory syndrome coronavirus 2 antibodies in pregnant women admitted to labor and delivery units. American Journal of Obstetrics & Gynecology. 2020;0. doi:[10.1016/j.ajog.2020.09.022](https://doi.org/10.1016/j.ajog.2020.09.022)

162.	Lim T, Delorey M, Bestul N, Johannsen M, Reed C, Hall AJ, et al. Changes in SARS CoV-2 Seroprevalence Over Time in Ten Sites in the United States, March August, 2020. Clinical Infectious Diseases. 2021. doi:[10.1093/cid/ciab185](https://doi.org/10.1093/cid/ciab185)

163.	Samore MH, Looney A, Orleans B, Greene T, Seegert N, Delgado JC, et al. Probability-Based Estimates of Severe Acute Respiratory Syndrome Coronavirus 2 Seroprevalence and Detection Fraction, Utah, USA. Emerging Infectious Diseases. 2021;27: 2786–2794. doi:[10.3201/eid2711.204435](https://doi.org/10.3201/eid2711.204435)

164.	Sutton M, Cieslak P, Linder M. Notes from the Field: Seroprevalence Estimates of SARS-CoV-2 Infection in Convenience Sample Oregon, May 11 15, 2020. MMWR Morbidity and Mortality Weekly Report. 2020;69: 1100–1101. doi:[10.15585/mmwr.mm6932a4](https://doi.org/10.15585/mmwr.mm6932a4)

165.	Chan PA, King E, Xu Y, Goedel W, Lasher L, Vargas M, et al. Seroprevalence of SARS-CoV-2 Antibodies in Rhode Island From a Statewide Random Sample. American Journal of Public Health. 2021;111: 700–703. doi:[10.2105/AJPH.2020.306115](https://doi.org/10.2105/AJPH.2020.306115)

166.	Menachemi N, Yiannoutsos CT, Dixon BE, Duszynski TJ, Fadel WF, Wools-Kaloustian KK, et al. Population Point Prevalence of SARS-CoV-2 Infection Based on a Statewide Random Sample Indiana, April 25, 2020. MMWR Morbidity and Mortality Weekly Report. 2020;69: 960–964. doi:[10.15585/mmwr.mm6929e1](https://doi.org/10.15585/mmwr.mm6929e1)

167.	Shakiba M, Nazemipour M, Salari A, Mehrabian F, Nazari SS, Rezvani SM, et al. Seroprevalence of SARS-CoV-2 in Guilan Province, Iran, April 2020. Emerging Infectious Diseases. 2021;27: 636–638. doi:[10.3201/eid2702.201960](https://doi.org/10.3201/eid2702.201960)

168.	Mousavi SA, Rouhanizadeh H, Saeedi M, Moosazadeh M, Ajami A, Fendereski A, et al. Seroprevalence of SARS-CoV-2 in Mazandaran province, Iran. Asian Pacific Journal of Tropical Medicine. 2021;14: 10–16. doi:[10.4103/1995-7645.304296](https://doi.org/10.4103/1995-7645.304296)

169.	Maraqa B, Basha W, Khayyat R, Abdul-Hadi A-R, Jabareen J, Al-Shakhra K, et al. Prevalence of SARS-CoV-2 antibodies in the Palestinian population: A primary health center-based cross-sectional study. PLoS One. 2021;16: e0258255. doi:[10.1371/journal.pone.0258255](https://doi.org/10.1371/journal.pone.0258255)

170.	Alsuwaidi AR, Al Hosani FI, Al Memari S, Narchi H, Abdel Wareth L, Kamal H, et al. Seroprevalence of COVID-19 infection in the Emirate of Abu Dhabi, United Arab Emirates: A population-based cross-sectional study. International Journal of Epidemiology. 2021. doi:[10.1093/ije/dyab077](https://doi.org/10.1093/ije/dyab077)

171.	Mijović B, Mašić S, Petković M, Knežević D, Aćimović J, Djaković-Dević J, et al. Seroprevalence of SARS-CoV-2 antibodies and knowledge, attitude and practice toward COVID-19 in the Republic of Srpska-Bosnia & Herzegovina: A population-based study. Borrow R, editor. PLoS One. 2022;17: e0262738. doi:[10.1371/journal.pone.0262738](https://doi.org/10.1371/journal.pone.0262738)

172.	Musa S. Population-based age-stratified seroepidemiological investigation protocol for coronavirus 2019 (COVID-19) infection in the Federation of Bosnia and Herzegovina. Zenodo; 2021. doi:[10.5281/ZENODO.5645820](https://doi.org/10.5281/ZENODO.5645820)

173.	Mouravi-Tarkhan O, Chitadze N. Sero-epidemiological Cross-sectional investigation on COVID-19 virus infection in Georgia. Zenodo; 2021. doi:[10.5281/ZENODO.5703254](https://doi.org/10.5281/ZENODO.5703254)

174.	Rytter MJH, Nygaard U, Mandic IN, Glenthøj JP, Schmidt LS, Cortes D, et al. Prevalence of SARS-CoV-2-Antibodies in Danish Children and Adults. Pediatric Infectious Disease Journal. 2021;40: e157–e159. doi:[10.1097/INF.0000000000003048](https://doi.org/10.1097/INF.0000000000003048)

175.	Iversen K, Bundgaard H, Hasselbalch RB, Kristensen JH, Nielsen PB, Pries-Heje M, et al. Risk of COVID-19 in health-care workers in Denmark: An observational cohort study. The Lancet Infectious diseases. 2020;20: 1401–1408. doi:[10.1016/S1473-3099(20)30589-2](https://doi.org/10.1016/S1473-3099(20)30589-2)

176.	Petersen MS, Strøm M, Christiansen DH, Fjallsbak JP, Eliasen EH, Johansen M, et al. Seroprevalence of SARS-CoV-2, Faroe Islands. Emerging Infectious Diseases. 2020;26: 2760–2762. doi:[10.3201/eid2611.202736](https://doi.org/10.3201/eid2611.202736)

177.	Capai L, Masse S, Fourié T, Decarreaux D, Canarelli J, Simeoni M-H, et al. Impact of the Second Epidemic Wave of SARS-CoV-2: Increased Exposure of Young People. Frontiers in Public Health. 2021;9: 1078. doi:[10.3389/fpubh.2021.715192](https://doi.org/10.3389/fpubh.2021.715192)

178.	Backhaus I, Hermsen D, Timm J, Boege F, Lübke N, Degode T, et al. SARS-CoV-2 seroprevalence and determinants of infection in young adults: A population-based seroepidemiological study. Public Health. 2022;207: 54–61. doi:[10.1016/j.puhe.2022.03.009](https://doi.org/10.1016/j.puhe.2022.03.009)

179.	Laub O, Leipold G, Toncheva AA, Peterhoff D, Einhauser S, Neckermann P, et al. Symptoms, SARS-CoV-2 Antibodies, and Neutralization Capacity in a Cross Sectional-Population of German Children. Frontiers in Pediatrics. 2021;9: 678937. doi:[10.3389/fped.2021.678937](https://doi.org/10.3389/fped.2021.678937)

180.	Fischer B, Knabbe C, Vollmer T. SARS-CoV-2 IgG seroprevalence in blood donors located in three different federal states, Germany, March to June 2020. Eurosurveillance. 2020;25. doi:[10.2807/1560-7917.ES.2020.25.28.2001285](https://doi.org/10.2807/1560-7917.ES.2020.25.28.2001285)

181.	Calò F, Di Fraia A, Russo A, Di Biase A, Misso S, Coppola N. Blood donor serological screening for SARS-CoV-2 as a tool to estimate the prevalence of asymptomatic infection in a low-intermediate endemic area of southern Italy after the first wave of the pandemic. Blood Transfusion. 2021. doi:[10.2450/2021.0196-21](https://doi.org/10.2450/2021.0196-21)

182.	Stefanelli P, Bella A, Fedele G, Pancheri S, Leone P, Vacca P, et al. Prevalence of SARS-CoV-2 IgG antibodies in an area of northeastern Italy with a high incidence of COVID-19 cases: A population-based study. Clinical Microbiology and Infection. 2021;27: 633.e1–633.e7. doi:[10.1016/j.cmi.2020.11.013](https://doi.org/10.1016/j.cmi.2020.11.013)

183.	Statistics Jersey. SARS-CoV-2 - Prevalence of antibodies in Jersey - Preliminary Analysis. Statistics Jersey; 2020 May.

184.	Statistics Jersey. SARS-CoV-2 - Prevalence of antibodies in Jersey - Community survey round 2. Statistics Jersey; 2020 Jun.

185.	Karachaliou M, Moncunill G, Espinosa A, Castaño-Vinyals G, Jiménez A, Vidal M, et al. Infection induced SARS-CoV-2 seroprevalence and heterogeneity of antibody responses in a general population cohort study in Catalonia Spain. Scientific Reports. 2021;11: 21571. doi:[10.1038/s41598-021-00807-4](https://doi.org/10.1038/s41598-021-00807-4)

186.	Iruzubieta P, Fernández-Lanas T, Rasines L, Cayon L, Álvarez-Cancelo A, Santos-Laso A, et al. Feasibility of large-scale population testing for SARS-CoV-2 detection by self-testing at home. Scientific Reports. 2021;11: 9819. doi:[10.1038/s41598-021-89236-x](https://doi.org/10.1038/s41598-021-89236-x)

187.	Beretta O, Pagani SC, Lazzaro M, Merlani G, Gallacchi MB. Seroprevalence of the SARS-CoV-2 virus in the population of the southern Switzerland (Canton Ticino) cohort study, results at 12 months. Swiss Medical Weekly. 2021;151: w30116. doi:[10.4414/smw.2021.w30116](https://doi.org/10.4414/smw.2021.w30116)

188.	El May E, Anker D, Epure A, Wagner C, Magnin J-L, Schmid A, et al. Corona Immunitas Fribourg - Immunité de la population Episode 2/2021. Corona Immunitas; 2021 Apr.

189.	Anker D, Chiorelo A, Epure A, Schmid A, Carmeli C, Rodondi P-Y, et al. Corona Immunitas Fribourg - Immunté de la population Episode 1/2020. Corona Immunitas; 2020 Nov.

190.	West EA, Kotoun OJ, Schori LJ, Kopp J, Kaufmann M, Rasi M, et al. Seroprevalence of SARS-CoV-2 antibodies, associated factors, experiences and attitudes of nursing home and home healthcare employees in Switzerland. BMC Infectious Diseases. 2022;22: 259. doi:[10.1186/s12879-022-07222-8](https://doi.org/10.1186/s12879-022-07222-8)

191.	Richard A, Wisniak A, Perez-Saez J, Garrison-Desany H, Petrovic D, Piumatti G, et al. Seroprevalence of anti-SARS-CoV-2 IgG antibodies, risk factors for infection and associated symptoms in Geneva, Switzerland: A population-based study. Scandinavian Journal of Public Health. 2022;50: 124–135. doi:[10.1177/14034948211048050](https://doi.org/10.1177/14034948211048050)

192.	Fenwick C, Croxatto A, Coste AT, Pojer F, André C, Pellaton C, et al. Changes in SARS-CoV-2 Spike versus Nucleoprotein Antibody Responses Impact the Estimates of Infections in Population-Based Seroprevalence Studies. Subbarao K, editor. Journal of Virology. 2020;95: e01828–20, /jvi/95/3/JVI.01828–20.atom. doi:[10.1128/JVI.01828-20](https://doi.org/10.1128/JVI.01828-20)

193.	Stringhini S, Wisniak A, Piumatti G, Azman AS, Lauer SA, Baysson H, et al. Seroprevalence of anti-SARS-CoV-2 IgG antibodies in Geneva, Switzerland (SEROCoV-POP): A population-based study. The Lancet. 2020;0. doi:[10.1016/S0140-6736(20)31304-0](https://doi.org/10.1016/S0140-6736(20)31304-0)

194.	Coronavirus (COVID-19) antibody and vaccination data for the UK. Office for National Statistics; 2021 May.

195.	Steel K, Davies B. Coronavirus (COVID-19) Infection Survey: Characteristics of people testing positive for COVID-19 in England and antibody data for the UK - Office for National Statistics. Office for National Statistics; 2020 Nov p. 12.

196.	Thompson CP, Grayson NE, Paton RS, Bolton JS, Lourenço J, Penman BS, et al. Detection of neutralising antibodies to SARS-CoV-2 to determine population exposure in Scottish blood donors between March and May 2020. Eurosurveillance. 2020;25: 2000685. doi:[10.2807/1560-7917.ES.2020.25.42.2000685](https://doi.org/10.2807/1560-7917.ES.2020.25.42.2000685)

197.	Sharma P, Basu S, Mishra S, Gupta E, Aggarwal R, Kale P, et al. SARS-CoV-2 seroprevalence in Delhi, India - September-October 2021 a population based seroepidemiological study. medRxiv. 2021. doi:[10.1101/2021.12.28.21268451](https://doi.org/10.1101/2021.12.28.21268451)

198.	Department of Health & Family Welfare, Government of Kerala. COVID 19 Sero Surveillance Report August - September 2021. Government of Kerala; 2021 p. 84.

199.	Misra P, Kant S, Rai S, Chaturvedi PK, Medigeshi, Yadav K, et al. A Multi-Centric Novel Coronavirus (COVID-19) Population-Based Age-Stratified Sero-Epidemiological Study (In Urban, Rural and Tribal Communities at Selected Centers in India: A Prospective Study). Zenodo; 2021. doi:[10.5281/ZENODO.5703289](https://doi.org/10.5281/ZENODO.5703289)

200.	Sharma N, Sharma P, Basu S, Bakshi R, Gupta E, Agarwal R, et al. Second wave of the Covid-19 pandemic in Delhi, India: High seroprevalence not a deterrent? medRxiv. 2021. doi:[10.1101/2021.09.09.21263331](https://doi.org/10.1101/2021.09.09.21263331)

201.	Khan SMS, Qurieshi MA, Haq I, Majid S, Ahmad J, Ayub T, et al. Seroprevalence of SARS-CoV-2-specific IgG antibodies in Kashmir, India, 7 months after the first reported local COVID-19 case: Results of a population-based seroprevalence survey from October to November 2020. BMJ Open. 2021;11: e053791. doi:[10.1136/bmjopen-2021-053791](https://doi.org/10.1136/bmjopen-2021-053791)

202.	Sharma N, Sharma P, Basu S, Saxena S, Chawla R, Dushyant K, et al. The seroprevalence of severe acute respiratory syndrome coronavirus 2 in Delhi, India: A repeated population-based seroepidemiological study. Transactions of The Royal Society of Tropical Medicine and Hygiene. 2021; trab109. doi:[10.1093/trstmh/trab109](https://doi.org/10.1093/trstmh/trab109)

203.	Kar SS, Sarkar S, Murali S, Dhodapkar R, Joseph NM, Aggarwal R. Prevalence and Time Trend of SARS-CoV-2 Infection in Puducherry, India, August 2020. Emerging Infectious Diseases. 2021;27: 666–669. doi:[10.3201/eid2702.204480](https://doi.org/10.3201/eid2702.204480)

204.	Namasivayam V, Jain A, Agrawal V, Prakash R, Dehury B, Becker M, et al. Understanding the Prevalence and Geographic Heterogeneity of SARS-CoV-2 Infection: Findings of the First Serosurvey in Uttar Pradesh, India. Journal of Epidemiology and Global Health. 2021;11: 364–376. doi:[10.1007/s44197-021-00012-6](https://doi.org/10.1007/s44197-021-00012-6)

205.	Satpati P, Sarangi S, Gantait K, Endow S, Mandal N, Panchanan K, et al. Sero-surveillance (IgG) of SARS-CoV-2 among Asymptomatic General population of Paschim Medinipur, West Bengal, India. 2020. doi:[10.1101/2020.09.12.20193219](https://doi.org/10.1101/2020.09.12.20193219)

206.	Khan SMS, Qurieshi MA, Haq I, Majid S, Bhat AA, Nabi S, et al. Seroprevalence of SARS-CoV-2 specific IgG antibodies in District Srinagar, northern India A cross-sectional study. PLoS One. 2020;15: e0239303. doi:[10.1371/journal.pone.0239303](https://doi.org/10.1371/journal.pone.0239303)

207.	Delhi P. Sero-prevalence study conducted by National Center for Disease Control NCDC, MoHFW, in Delhi, June 2020. Indian Ministry of Health and Family Welfare; 2020 Jul.

208.	Megasari NLA, Utsumi T, Yamani LN, Juniastuti, Gunawan E, Furukawa K, et al. Seroepidemiological study of SARS-CoV-2 infection in East Java, Indonesia. PLoS One. 2021;16: e0251234. doi:[10.1371/journal.pone.0251234](https://doi.org/10.1371/journal.pone.0251234)

209.	Chen L-L, Abdullah SMU, Chan W-M, Chan BP-C, Ip JD, Chu AWH, et al. Contribution of low population immunity to the severe Omicron BA.2 outbreak in Hong Kong. In Review; 2022 Apr. doi:[10.21203/rs.3.rs-1512533/v1](https://doi.org/10.21203/rs.3.rs-1512533/v1)

210.	He Z, Ren L, Yang J, Guo L, Feng L, Ma C, et al. Seroprevalence and humoral immune durability of anti-SARS-CoV-2 antibodies in Wuhan, China: A longitudinal, population-level, cross-sectional study. The Lancet. 2021;397: 1075–1084. doi:[10.1016/S0140-6736(21)00238-5](https://doi.org/10.1016/S0140-6736(21)00238-5)

211.	Goto A, Miyakawa K, Nakayama I, Yagome S, Xu J, Kaneko M, et al. Analysis of humoral immunity against emerging SARS-CoV-2 variants: A population-based prevalence study in Yokohama, Japan. medRxiv; 2022. p. 2022.03.26.22272766. doi:[10.1101/2022.03.26.22272766](https://doi.org/10.1101/2022.03.26.22272766)

212.	Prefecture of Hiroshima. About the result of the 2nd new coronavirus antibody prevalence survey. Japan: Prefecture of Hiroshima; 2021 Feb.

213.	Sam I-C, Chong YM, Abdullah A, Fu JYL, Hasan MS, Jamaluddin FH, et al. Changing predominant SARS-CoV-2 lineages drives successive COVID-19 waves in Malaysia, February 2020 to March 2021. Journal of Medical Virology. 2021;94: 1146–1153. doi:[10.1002/jmv.27441](https://doi.org/10.1002/jmv.27441)

214.	Sebastião CS, Galangue M, Gaston C, Van-Dunen R, Quivinja J, Lunbungululo E, et al. Seroprevalence of anti-SARS-CoV-2 antibodies and risk factors among healthy blood donors in Luanda, Angola. BMC Infectious Diseases. 2021;21: 1131. doi:[10.1186/s12879-021-06814-0](https://doi.org/10.1186/s12879-021-06814-0)

215.	Dabakuyo Yonli S, Traoré I, Tinto H, Ouédraogo S. ÉMuL-COVID-19: A Multidisciplinary study against COVID-19 in Burkina Faso. Zenodo; 2021. doi:[10.5281/ZENODO.5644217](https://doi.org/10.5281/ZENODO.5644217)

216.	Nwosu K, Fokam J, Wanda F, Mama L, Orel E, Ray N, et al. SARS-CoV-2 antibody seroprevalence and associated risk factors in an urban district in Cameroon. Nature Communications. 2021;12: 5851. doi:[10.1038/s41467-021-25946-0](https://doi.org/10.1038/s41467-021-25946-0)

217.	Manirakiza A, Malaka C, Yambiyo BM, Marie-Astrid Vernet, Vernet G, Nakoune E. Seroprevalence of anti-SARS-CoV-2 antibodies among communities in Bangui, Central African Republic. Zenodo; 2021. doi:[10.5281/ZENODO.5794867](https://doi.org/10.5281/ZENODO.5794867)

218.	Bonguili NCB, Fritz M, Lenguiya H, Mayengue PI, Koukouikila-Koussounda F, Dossou-Yovo LR, et al. Early Release - Early Circulation of SARS-CoV-2, Congo, 2020 - Volume 28, Number 4 2022 - Emerging Infectious Diseases journal - CDC. 2022. doi:[10.3201/eid2804.212476](https://doi.org/10.3201/eid2804.212476)

219.	Nkuba AN, Makiala SM, Guichet E, Tshiminyi PM, Bazitama YM, Yambayamba MK, et al. High Prevalence of Anti 2 (Anti) Antibodies After the First Wave of Coronavirus Disease 2019 (COVID-19) in Kinshasa, Democratic Republic of the Congo: Results of a Cross-sectional Household-Based Survey. Clinical Infectious Diseases. 2022;74: 882–890. doi:[10.1093/cid/ciab515](https://doi.org/10.1093/cid/ciab515)

220.	Gudina EK, Ali S, Girma E, Gize A, Tegene B, Hundie GB, et al. Seroepidemiology and model-based prediction of SARS-CoV-2 in Ethiopia: Longitudinal cohort study among front-line hospital workers and communities. The Lancet Global Health. 2021;9: e1517–e1527. doi:[10.1016/S2214-109X(21)00386-7](https://doi.org/10.1016/S2214-109X(21)00386-7)

221.	Abdella S, Riou S, Tessema M, Assefa A, Seifu A, Blachman A, et al. Prevalence of SARS-CoV-2 in urban and rural Ethiopia: Randomized household serosurveys reveal level of spread during the first wave of the pandemic. eClinicalMedicine. 2021;35: 100880. doi:[10.1016/j.eclinm.2021.100880](https://doi.org/10.1016/j.eclinm.2021.100880)

222.	Shaweno T, Abdulhamid I, Bezabih L, Teshome D, Derese B, Tafesse H, et al. Sero-prevalence of SARS-CoV-2 Antibody Among Adults in the General Population in Diredawa, Ethiopia. Research Square. 2021. doi:[10.21203/rs.3.rs-266413/v1](https://doi.org/10.21203/rs.3.rs-266413/v1)

223.	Kempen JH, Abashawl A, Suga HK, Nigussie Difabachew M, Kempen CJ, Tesfaye Debele M, et al. SARS-CoV-2 Serosurvey in Addis Ababa, Ethiopia. The American Journal of Tropical Medicine and Hygiene. 2020;103: 2022–2023. doi:[10.4269/ajtmh.20-0816](https://doi.org/10.4269/ajtmh.20-0816)

224.	Mveang Nzoghe A, Leboueny M, Kuissi Kamgaing E, Maloupazoa Siawaya AC, Bongho EC, Mvoundza Ndjindji O, et al. Circulating anti-SARS-CoV-2 nucleocapsid (N)-protein antibodies and anti-SARS-CoV-2 spike (S)-protein antibodies in an African setting: Herd immunity, not there yet! BMC Research Notes. 2021;14: 152. doi:[10.1186/s13104-021-05570-3](https://doi.org/10.1186/s13104-021-05570-3)

225.	Lucinde R, Mugo D, Bottomley C, Karani A, Gardiner E, Aziza R, et al. Sero-surveillance for IgG to SARS-CoV-2 at antenatal care clinics in three Kenyan referral hospitals: Repeated cross-sectional surveys 2020-21. medRxiv; 2022. p. 2022.03.03.22271860. doi:[10.1101/2022.03.03.22271860](https://doi.org/10.1101/2022.03.03.22271860)

226.	Nyagwange J, Kutima B, Mwai K, Karanja HK, Gitonga JN, Mugo D, et al. Comparative performance of WANTAI ELISA for total immunoglobulin to receptor binding protein and an ELISA for IgG to spike protein in detecting SARS-CoV-2 antibodies in Kenyan populations. Journal of Clinical Virology. 2022;146: 105061. doi:[10.1016/j.jcv.2021.105061](https://doi.org/10.1016/j.jcv.2021.105061)

227.	Etyang AO, Adetifa I, Omore R, Misore T, Ziraba AK, Ng’oda MA, et al. SARS-CoV-2 Seroprevalence in three Kenyan Health and Demographic Surveillance Sites, December 2020-May 2021. medRxiv. 2022. doi:[10.1101/2022.02.07.22270012](https://doi.org/10.1101/2022.02.07.22270012)

228.	Munywoki PK, Nasimiyu C, Alando MD, Otieno N, Ombok C, Njoroge R, et al. Seroprevalence and risk factors of SARS-CoV-2 infection in an urban informal settlement in Nairobi, Kenya, December 2020. F1000Research. 2022;10: 853. doi:[10.12688/f1000research.72914.2](https://doi.org/10.12688/f1000research.72914.2)

229.	Lucinde R, Mugo D, Bottomley C, Aziza R, Gitonga J, Karanja H, et al. Sero-surveillance for IgG to SARS-CoV-2 at antenatal care clinics in two Kenyan referral hospitals. medRxiv. 2021; 2021.02.05.21250735. doi:[10.1101/2021.02.05.21250735](https://doi.org/10.1101/2021.02.05.21250735)

230.	Ngere I, Dawa J, Hunsperger E, Otieno N, Masika M, Amoth P, et al. High seroprevalence of SARS-CoV-2 but low infection fatality ratio eight months after introduction in Nairobi, Kenya. International Journal of Infectious Diseases. 2021;112: 25–34. doi:[10.1016/j.ijid.2021.08.062](https://doi.org/10.1016/j.ijid.2021.08.062)

231.	Crowell TA, Daud II, Maswai J, Owuoth J, Sing’oei V, Imbach M, et al. SARS-CoV-2 antibody prevalence in people with and without HIV in rural western Kenya, January to March 2020. AIDS. 2021;35: 2401–2404. doi:[10.1097/QAD.0000000000003054](https://doi.org/10.1097/QAD.0000000000003054)

232.	Razafimahatratra SL, Ndiaye MDB, Rasoloharimanana LT, Dussart P, Sahondranirina PH, Randriamanantany ZA, et al. Seroprevalence of ancestral and Beta SARS-CoV-2 antibodies in Malagasy blood donors. The Lancet Global Health. 2021;9: e1363–e1364. doi:[10.1016/S2214-109X(21)00361-2](https://doi.org/10.1016/S2214-109X(21)00361-2)

233.	Schoenhals M, Rabenindrina N, Rakotondramanga JM, Dussart P, Randremanana R, Heraud J-M, et al. SARS-CoV-2 antibody seroprevalence follow-up in Malagasy blood donors during the 2020 COVID-19 Epidemic. eBioMedicine. 2021;68: 103419. doi:[10.1016/j.ebiom.2021.103419](https://doi.org/10.1016/j.ebiom.2021.103419)

234.	Sagara I, Woodford J, Kone M, Assadou MH, Katile A, Attaher O, et al. Rapidly Increasing Severe Acute Respiratory Syndrome Coronavirus 2 Seroprevalence and Limited Clinical Disease in 3 Malian Communities: A Prospective Cohort Study. Clinical Infectious Diseases. 2022;74: 1030–1038. doi:[10.1093/cid/ciab589](https://doi.org/10.1093/cid/ciab589)

235.	Mussagy Mahomed. Inquérito Sero-epidemiológico de SARS-CoV-2 na Cidade de Maxixe e Vila de Massinga (InCOVID 2020). República de Moçambique Ministério da Saúde; 2020 Dec.

236.	Arlete Mahumane. Inquérito Sero-epidemiológico de SARS-CoV-2 na Cidade de Chimoio (InCOVID 2020). República de Moçambique Ministério da Saúde; 2020 Nov.

237.	Jerónimo Langa. Inquérito Sero-epidemiológico de SARS-CoV-2 na Cidade de Lichinga (InCOVID 2020). República de Moçambique Ministério da Saúde; 2020 Oct.

238.	Mussagy Mahomed. Inquérito Sero-epidemiológico de SARS-CoV-2 na Cidade de Tete (InCOVID 2020). República de Moçambique Ministério da Saúde; 2020 Sep.

239.	Paulo Arnaldo. Inquérito Sero-epidemiológico de SARS-CoV-2 na Cidade de Pemba (InCOVID 2020). República de Moçambique Ministério da Saúde; 2020 Jul.

240.	Eduardo Samo Gudo. Inquérito Sero-epidemiológico de SARS-CoV-2 na Cidade de Nampula. República de Moçambique Ministério da Saúde; 2020 Jul p. 19.

241.	Ifeorah I, Nna E, Okeke U, Ivo E, Ayemoba O, Yunusa T, et al. Sero-pravelence of SARS CoV-2 IgM and IgG Antibodies Amongst Blood Donors in Nigeria. Research Square. 2021. doi:[10.21203/rs.3.rs-151037/v1](https://doi.org/10.21203/rs.3.rs-151037/v1)

242.	Kleynhans J, Tempia S, Wolter N, von Gottberg A, Bhiman JN, Buys A, et al. SARS-CoV-2 Seroprevalence after Third Wave of Infections, South Africa. Emerging Infectious Diseases. 2022;28. doi:[10.3201/eid2805.220278](https://doi.org/10.3201/eid2805.220278)

243.	Cable R, Coleman C, Glatt T, Grebe E, Mhlanga L, Nyano C, et al. Estimates of prevalence of anti-SARS-CoV-2 antibodies among blood donors in eight provinces of South Africa in November 2021. 2022. doi:[10.21203/rs.3.rs-1359658/v1](https://doi.org/10.21203/rs.3.rs-1359658/v1)

244.	Sawry S, le Roux JG, Mbatha P, Wolter N, Bhiman J, Kekana M, et al. High prevalence of SARS-CoV-2 antibodies in pregnant women in the inner city of Johannesburg, Gauteng Province, South Africa. Zenodo; 2021. doi:[10.5281/ZENODO.5643228](https://doi.org/10.5281/ZENODO.5643228)

245.	Cohen C, Kleynhans J, von Gottberg, Anne, McMorrow M, Wolter N, Bhiman J, et al. SARS-CoV-2 Seroprevalence in a Rural and Urban Household Cohort, South Africa, July 2020 2021. Zenodo; 2022. doi:[10.5281/ZENODO.5898414](https://doi.org/10.5281/ZENODO.5898414)

246.	Hsiao M, Davies M-A, Kalk E, Hardie D, Naidoo M, Center C, et al. SARS-CoV-2 seroprevalence in the Cape Town Metropolitan sub-districts after the peak of infections. NICD COVID-19 Special Public Health Surveillance Bulletin. 2020;18: 1–9.

247.	Wiens KE, Mawien PN, Rumunu J, Slater D, Jones FK, Moheed S, et al. Seroprevalence of Severe Acute Respiratory Syndrome Coronavirus 2 IgG in Juba, South Sudan, 2020. Emerging Infectious Diseases. 2021;27: 1598–1606. doi:[10.3201/eid2706.210568](https://doi.org/10.3201/eid2706.210568)

248.	Shanaube K, Schaap A, Klinkenberg E, Floyd S, Bwalya J, M C, et al. SARS-CoV-2 seroprevalence and associated risk factors in peri-urban Zambia: A population-based study. International Journal of Infectious Diseases. 2022; S1201971222001588. doi:[10.1016/j.ijid.2022.03.021](https://doi.org/10.1016/j.ijid.2022.03.021)

249.	Fryatt A, Simms V, Bandason T, Redzo N, Olaru ID, Ndhlovu CE, et al. Community SARS-CoV-2 seroprevalence before and after the second wave of SARS-CoV-2 infection in Harare, Zimbabwe. EClinicalMedicine. 2021;41: 101172. doi:[10.1016/j.eclinm.2021.101172](https://doi.org/10.1016/j.eclinm.2021.101172)

250.	Muñoz L, Pífano M, Bolzán A, Varela T, Comes Y, Specogna M, et al. Surveillance and Seroprevalence: Evaluation of IgG antibodies for SARS-Cov2 by ELISA in the popular neighborhood Villa Azul, Quilmes, province of Buenos Aires, Argentina. 2020 Aug. doi:[10.1590/SciELOPreprints.1147](https://doi.org/10.1590/SciELOPreprints.1147)

251.	Figar S, Pagotto V, Luna L, Salto J, Manslau MW, Mistchenko AS, et al. SEVERE ACUTE RESPIRATORY SYNDROME CORONAVIRUS 2 SEROEPIDEMIOLOGY STUDY IN ARGENTINIAN SLUM. Medicina Buenos Aires. 2021;81: 2020.07.14.20153858.

252.	Tess BH, Alves MCGP, Reinach FC, Granato CFH, Rizzatti E, Pintao MC, et al. SoroEpi MSP Inquéritos soroepidemiológicos seriados para monitorar a prevalência da infecção por SARS-CoV-2 no Município de São Paulo, SP, Brasil: Fase 6. SoroEpi MSP; 2020 May.

253.	Pereira KAS. PREVALÊNCIA DO CORONAVÍRUS 2 DA SÍNDROME RESPIRATÓRIA AGUDA GRAVE (SARS-COV-2) EM COMUNIDADES QUILOMBOLAS DO MUNÍCIPIO DE CAMETÁ, PARÁ. The Brazilian Journal of Infectious Diseases. 2022; 102060–102060.

254.	Cidade de São Paulo. Inquérito sorológico adultos ($>$18)para sars-cov-2 - resultado da fase 4 de 2021. Cidade de São Paulo; 2021 Mar.

255.	Fofana MO, Nery N, Ticona JPA, Belitardo EMMA, Victoriano R, Anjos RO, et al. Structural factors contributing to SARS-CoV-2 infection risk in the urban slum setting. medRxiv. 2022. doi:[10.1101/2022.02.13.22270856](https://doi.org/10.1101/2022.02.13.22270856)

256.	da Silva HP, Abreu IN, Lima CNC, de Lima ACR, do Nascimento Barbosa A, de Oliveira LR, et al. Migration in times of pandemic: SARS-CoV-2 infection among the Warao indigenous refugees in Belém, Pará, Amazonia, Brazil. BMC Public Health. 2021;21: 1659. doi:[10.1186/s12889-021-11696-7](https://doi.org/10.1186/s12889-021-11696-7)

257.	Miraglia JL, Nascimento Monteiro C, Giannecchini Romagnolo A, Xavier Gomes R, Pitangueiras Mangueira C, Aparecida Rosseto-Welter E, et al. A seroprevalence survey of anti-SARS-CoV-2 antibodies among individuals 18 years of age or older living in a vulnerable region of the city of São Paulo, Brazil. PLoS One. 2021;16: 1–9. doi:[10.1371/journal.pone.0255412](https://doi.org/10.1371/journal.pone.0255412)

258.	Pontes GS, de Melo Silva J, Pinheiro-Silva R, Barbosa AN, Santos LC, de Pádua Quirino Ramalho A, et al. Increased vulnerability to SARS-CoV-2 infection among indigenous people living in the urban area of Manaus. Scientific Reports. 2021;11: 17534. doi:[10.1038/s41598-021-96843-1](https://doi.org/10.1038/s41598-021-96843-1)

259.	Buss LF, Prete CA, Abrahim CMM, Mendrone A, Salomon T, Almeida-Neto C de, et al. Three-quarters attack rate of SARS-CoV-2 in the Brazilian Amazon during a largely unmitigated epidemic. Science. 2021;371: 288–292. doi:[10.1126/science.abe9728](https://doi.org/10.1126/science.abe9728)

260.	Ferrugini CLP, Boldrini NAT, Costa FLS, Salgueiro MA de OB, Coelho PD de P, Miranda AE. SARS-CoV-2 infection in pregnant women assisted in a high-risk maternity hospital in Brazil: Clinical aspects and obstetric outcomes. Kalk EK, editor. PLoS One. 2022;17: e0264901. doi:[10.1371/journal.pone.0264901](https://doi.org/10.1371/journal.pone.0264901)

261.	Lugon P, Fuller T, Damasceno L, Calvet G, Resende PC, Matos AR, et al. SARS-CoV-2 Infection Dynamics in Children and Household Contacts in a Slum in Rio de Janeiro. Pediatrics. 2021;148: e2021050182. doi:[10.1542/peds.2021-050182](https://doi.org/10.1542/peds.2021-050182)

262.	Oliveira LM da S, Tiyo BT, Silva LT da, Fonseca LAM, Rocha RC, Santos VA dos, et al. Prevalence of anti-SARS-CoV-2 antibodies in outpatients of a large public university hospital in Sao Paulo, Brazil. Revista do Instituto de Medicina Tropical de São Paulo. 2020;62. doi:[10.1590/s1678-9946202062091](https://doi.org/10.1590/s1678-9946202062091)

263.	Silva AVFG, Menezes D, Moreira FRR, Torres OA, Fonseca PLC, Moreira RG, et al. Seroprevalence, Prevalence, and Genomic Surveillance: Monitoring the Initial Phases of the SARS-CoV-2 Pandemic in Betim, Brazil. Frontiers in Microbiology. 2022;13.

264.	Diegoli H, Conzatti VS, Mazin SC, Safanelli J, Chiaradia Dellatorre LD, Bett K, et al. Population-Based Study of anti-SARS-CoV-2, Social Distancing and Government Responses in Joinville, Brazil. Public and Global Health; 2021 Feb. doi:[10.1101/2021.02.08.21251009](https://doi.org/10.1101/2021.02.08.21251009)

265.	Calheiros Santos A, Salete de Paula V, Custódio Miguel J, Ferreira da Silva E, Machado Pinto LC, Lewis-Ximenez LL, et al. P-87 HIGH PREVALENCE OF SARS-COV-2 ANTIBODIES IN PREGNANT WOMEN INFECTED WITH VIRAL HEPATITIS IN BRAZIL. Annals of Hepatology. 2021;24: 100450. doi:[10.1016/j.aohep.2021.100450](https://doi.org/10.1016/j.aohep.2021.100450)

266.	Bernardes-Souza B, Júnior SRC, Santos CA, Neto RMDN, Bottega FDC, Godoy DC, et al. Logistics workers are a key factor for SARS-CoV-2 spread in Brazilian small towns: Case-control study (Preprint). JMIR Public Health and Surveillance. 2021. doi:[10.2196/30406](https://doi.org/10.2196/30406)

267.	Martinez EZ, Passos ADC, Fabbro ALD, Silva AS da, Escarso AC, Pazin-Filho A, et al. Prevalence of virological and serological markers of SARS-CoV-2 infection in the population of Ribeirão Preto, Southeast Brazil: An epidemiological survey. Revista da Sociedade Brasileira de Medicina Tropical. 2021;54: e0210–2021. doi:[10.1590/0037-8682-0210-2021](https://doi.org/10.1590/0037-8682-0210-2021)

268.	Tess BH, Granato CFH, Alves MCGP, Pintão MCT, Nunes MC, Rizzatti EG, et al. Assessment of initial SARS-CoV-2 seroprevalence in the most affected districts in the municipality of São Paulo, Brazil. The Brazilian Journal of Infectious Diseases. 2021;25: 101604. doi:[10.1016/j.bjid.2021.101604](https://doi.org/10.1016/j.bjid.2021.101604)

269.	Gurgel RQ, Sá LC de, Souza DRV, Martins AF, Matos ILS, Lima AGA, et al. SARS-CoV-2 has been circulating in northeastern Brazil since February 2020: Evidence for antibody detection in asymptomatic patients. Journal of Infection. 2020;0. doi:[10.1016/j.jinf.2020.11.037](https://doi.org/10.1016/j.jinf.2020.11.037)

270.	Serrano-Coll H, Miller H, Rodríguez-Van Der Hamen C, Gastelbondo B, Novoa W, Oviedo M, et al. High Prevalence of SARS-CoV-2 in an Indigenous Community of the Colombian Amazon Region. Tropical Medicine and Infectious Disease. 2021;6: 191. doi:[10.3390/tropicalmed6040191](https://doi.org/10.3390/tropicalmed6040191)

271.	Mattar S, Alvis-Guzman N, Garay E, Rivero R, García A, Botero Y, et al. Severe Acute Respiratory Syndrome Coronavirus 2 Seroprevalence Among Adults in a Tropical City of the Caribbean Area, Colombia: Are We Much Closer to Herd Immunity Than Developed Countries? Open Forum Infectious Diseases. 2020;7. doi:[10.1093/ofid/ofaa550](https://doi.org/10.1093/ofid/ofaa550)

272.	Del Brutto OH, Costa AF, Recalde BY, Mera RM. Factors Associated With a Persistent Seronegative Status 1 Year After a SARS-CoV-2 Massive Infection Outbreak in Community Dwellers Living in Rural Ecuador: A Prospective Population-based Study. Journal of Primary Care & Community Health. 2021;12. doi:[10.1177/21501327211054989](https://doi.org/10.1177/21501327211054989)

273.	Acurio-Páez D, Vega B, Orellana D, Charry R, Gómez A, Obimpeh M, et al. Seroprevalence of SARS-CoV-2 Infection and Adherence to Preventive Measures in Cuenca, Ecuador, October 2020, a Cross-Sectional Study. International Journal of Environmental Research and Public Health. 2021;18: 4657. doi:[10.3390/ijerph18094657](https://doi.org/10.3390/ijerph18094657)

274.	Flamand C, Alves Sarmento C, Enfissi A, Bailly S, Beillard E, Gaillet M, et al. Seroprevalence of anti-SARS-CoV-2 IgG at the first epidemic peak in French Guiana, July 2020. Bowman N, editor. PLoS Neglected Tropical Diseases. 2021;15: e0009945. doi:[10.1371/journal.pntd.0009945](https://doi.org/10.1371/journal.pntd.0009945)

275.	Chisolm T, Walker JP, Leys Y, Butterfield TR, Medley C, Thompson TK, et al. SARS-CoV-2 Seroprevalence Among Antenatal Clinic Attendees in Kingston, Jamaica, September-November 2020. 2021. doi:[10.1101/2021.02.08.21251367](https://doi.org/10.1101/2021.02.08.21251367)

276.	Monteón V, Pérez FL, Hernández VP, Pacheco AO, Guzman PF, Torres GIG. Seroprevalence of SARS-CoV-2 antibodies in blood donors during the third wave of infection in Campeche Mexico. Transfusion and Apheresis Science. 2022; 103374. doi:[10.1016/j.transci.2022.103374](https://doi.org/10.1016/j.transci.2022.103374)

277.	Muñoz-Medina JE, Grajales-Muñiz C, Salas-Lais AG, Fernandes-Matano L, López-Macías C, Monroy-Muñoz IE, et al. SARS-CoV-2 IgG Antibodies Seroprevalence and Sera Neutralizing Activity in MEXICO: A National Cross-Sectional Study during 2020. Microorganisms. 2021;9: 850. doi:[10.3390/microorganisms9040850](https://doi.org/10.3390/microorganisms9040850)

278.	Remes-Troche JM, Ramos-De-la-Medina A, Manríquez-Reyes M, Martínez-Pérez Maldonado L, Solis-Gonzalez MA, Hernández Flores KG, et al. Prevalence of SARS-CoV-2 IgG antibodies in a population from Veracruz (Southeastern Mexico). medRxiv. 2020. doi:[10.1101/2020.10.19.20215558](https://doi.org/10.1101/2020.10.19.20215558)

279.	Martinez-Acuña N, Avalos-Nolazco DM, Rodriguez-Rodriguez DR, Martinez-Liu CG, Galan-Huerta KA, Padilla-Rivas GR, et al. Seroprevalence of Anti-SARS-CoV-2 Antibodies in Blood Donors from Nuevo Leon State, Mexico, during 2020: A Retrospective Cross-Sectional Evaluation. Viruses. 2021;13: 1225. doi:[10.3390/v13071225](https://doi.org/10.3390/v13071225)

280.	Champigneulle B, Hancco I, Renan R, Doutreleau S, Stauffer E, Pichon A, et al. High-Altitude Environment and COVID-19: SARS-CoV-2 Seropositivity in the Highest City in the World. High Altitude Medicine & Biology. 2021; ham.2021.0020. doi:[10.1089/ham.2021.0020](https://doi.org/10.1089/ham.2021.0020)

281.	Tarazona-Castro Y, Troyes-Rivera L, Martins-Luna J, Cabellos-Altamirano F, Aguilar-Luis MA, Carrillo-Ng H, et al. Detection of SARS-CoV-2 antibodies in febrile patients from an endemic region of dengue and chikungunya in Peru. PLoS One. 2022;17: e0265820. doi:[10.1371/journal.pone.0265820](https://doi.org/10.1371/journal.pone.0265820)

282.	Álvarez-Antonio C, Meza-Sánchez G, Calampa C, Casanova W, Carey C, Alava F, et al. Seroprevalence of anti-SARS-CoV-2 antibodies in Iquitos, Loreto, Peru. medRxiv. 2021; 2021.01.17.21249913. doi:[10.1101/2021.01.17.21249913](https://doi.org/10.1101/2021.01.17.21249913)

283.	Reyes-Vega MF, Soto-Cabezas MG, Cárdenas F, Martel KS, Valle A, Valverde J, et al. SARS-CoV-2 prevalence associated to low socioeconomic status and overcrowding in an LMIC megacity: A population-based seroepidemiological survey in Lima, Peru. eClinicalMedicine. 2021;34: 100801. doi:[10.1016/j.eclinm.2021.100801](https://doi.org/10.1016/j.eclinm.2021.100801)

284.	Guevara-Ríos E, Espinola-Sánchez M, Carranza-Asmat C, Ayala-Peralta F, Álvarez-Carrasco R, Luna-Figueroa A, et al. SARS-COVID-19 antibodies in pregnant women at a level III hospital in PeruSARS-COVID-19 antibodies in pregnant women at a level III hospital in Peru. Revista Peruana de Ginecología y Obstetricia. 2020;66. doi:[10.31403/rpgo.v66i2259](https://doi.org/10.31403/rpgo.v66i2259)

285.	Goldfarb DM, Mâsse LC, Watts AW, Hutchison SM, Muttucomaroe L, Bosman ES, et al. SARS-CoV-2 seroprevalence among Vancouver public school staff in British Columbia, Canada: A cross-sectional study. BMJ Open. 2022;12: e057846. doi:[10.1136/bmjopen-2021-057846](https://doi.org/10.1136/bmjopen-2021-057846)

286.	Tang X, Sharma A, Pasic M, Brown P, Colwill K, Gelband H, et al. Assessment of SARS-CoV-2 Seropositivity During the First and Second Viral Waves in 2020 and 2021 Among Canadian Adults. JAMA Network Open. 2022;5: e2146798. doi:[10.1001/jamanetworkopen.2021.46798](https://doi.org/10.1001/jamanetworkopen.2021.46798)

287.	Tuite AR, Fisman D, Abe KT, Rathod B, Pasculescu A, Colwill K, et al. Estimating SARS-CoV-2 Seroprevalence in Canadian Blood Donors, April 2020 to March 2021: Improving Accuracy with Multiple Assays. Pride DT, editor. Microbiology Spectrum. 2022;10: e02563–21. doi:[10.1128/spectrum.02563-21](https://doi.org/10.1128/spectrum.02563-21)

288.	Manny E, Carroll A, Charlton C, Robinson J, Subbarao P, Azad MB, et al. Increased Mask Use and Fewer Gatherings Associated with Lower SARS-CoV-2 Seropositivity Among Young School-Age Children. SSRN. 2020. doi:[10.2139/ssrn.3728570](https://doi.org/10.2139/ssrn.3728570)

289.	Lewin A, Therrien R, De Serres G, Grégoire Y, Perreault J, Drouin M, et al. SARS-CoV-2 seroprevalence among blood donors in Québec, and analysis of symptoms associated with seropositivity: A nested case-control study. Canadian Journal of Public Health. 2021;112: 576–586. doi:[10.17269/s41997-021-00531-6](https://doi.org/10.17269/s41997-021-00531-6)

290.	Tanunliong G, Liu AC, Kaweski S, Irvine M, Reyes RC, Purych D, et al. Age-Associated Seroprevalence of Coronavirus Antibodies: Population-Based Serosurveys in 2013 and 2020, British Columbia, Canada. Frontiers in Immunology. 2022;13: 836449. doi:[10.3389/fimmu.2022.836449](https://doi.org/10.3389/fimmu.2022.836449)

291.	Villarreal A, Rangel G, Zhang X, Wong D, Britton G, Fernandez PL, et al. Performance of a Point of Care Test for Detecting IgM and IgG Antibodies Against SARS-CoV-2 and Seroprevalence in Blood Donors and Health Care Workers in Panama. Frontiers in Medicine. 2021;8: 616106. doi:[10.3389/fmed.2021.616106](https://doi.org/10.3389/fmed.2021.616106)

292.	Clarke KEN, Jones JM, Deng Y, Nycz E, Lee A, Iachan R, et al. Seroprevalence of Infection-Induced SARS-CoV-2 Antibodies United States, September 2021 2022. MMWR Morbidity and Mortality Weekly Report. 2022;71: 606–608. doi:[10.15585/mmwr.mm7117e3](https://doi.org/10.15585/mmwr.mm7117e3)

293.	Clarke KEN, Kim Y, Jones J, Lee A, Deng Y, Nycz E, et al. Pediatric Infection-Induced SARS-CoV-2 Seroprevalence Estimation Using Commercial Laboratory Specimens: How Representative Is It of the General U.S. Pediatric Population? SSRN. 2022. doi:[10.2139/ssrn.4092074](https://doi.org/10.2139/ssrn.4092074)

294.	Sood N, Pernet O, Lam CN, Klipp A, Kotha R, Kovacs A, et al. Seroprevalence of Antibodies Specific to Receptor Binding Domain of SARS-CoV-2 and Vaccination Coverage Among Adults in Los Angeles County, April 2021: The LA Pandemic Surveillance Cohort Study. JAMA Network Open. 2022;5: e2144258. doi:[10.1001/jamanetworkopen.2021.44258](https://doi.org/10.1001/jamanetworkopen.2021.44258)

295.	Routledge I, Takahashi S, Epstein A, Hakim J, Janson O, Turcios K, et al. Using sero-epidemiology to monitor disparities in vaccination and infection with SARS-CoV-2. Nature Communications. 2022;13: 2451. doi:[10.1038/s41467-022-30051-x](https://doi.org/10.1038/s41467-022-30051-x)

296.	Xu L, Doyle J, Barbeau DJ, Le Sage V, Wells A, Duprex WP, et al. A Cross-Sectional Study of SARS-CoV-2 Seroprevalence between Fall 2020 and February 2021 in Allegheny County, Western Pennsylvania, USA. Pathogens. 2021;10: 710. doi:[10.3390/pathogens10060710](https://doi.org/10.3390/pathogens10060710)

297.	Smith BK, Janowski AB, Fremont AC, Adams LJ, Dai Y-N, Farnsworth CW, et al. Progression of SARS-CoV-2 Seroprevalence in St. Louis, Missouri, through January 2021. mSphere. 2021;6: e00450–21. doi:[10.1128/mSphere.00450-21](https://doi.org/10.1128/mSphere.00450-21)

298.	LaCourse S. SARS-CoV-2 Seroprevalence and Antibody Response Among Pregnant People in Seattle, WA. https://academic.oup.com/HTTPHandlers/Sigma/LoginHandler.ashx?error=login_required&state=86d424ea-437a-410f-aa59-b0e6f438970eredirecturl%3Dhttpszazjzjacademiczwoupzwcomzjofidzjarticlezj8zjSupplementzx1zjS345zj6450201;

299.	Kilpatrick RD, Sánchez-Soliño O, Alami NN, Johnson C, Fang Y, Wegrzyn LR, et al. EpidemiologiCal POpulatioN STudy of SARS-CoV-2 in Lake CounTy, Illinois (CONTACT): Methodology and Baseline Characteristics of a Community-Based Surveillance Study. Infectious Diseases and Therapy. 2022;11: 899–911. doi:[10.1007/s40121-022-00593-0](https://doi.org/10.1007/s40121-022-00593-0)

300.	Stout RL, Rigatti SJ. The Silent Pandemic COVID-19 in the Asymptomatic Population. medRxiv. 2021. doi:[doi.org/10.1101/2020.11.10.20215145](https://doi.org/doi.org/10.1101/2020.11.10.20215145)

301.	Dodd RY, Spencer BR, Xu M, Foster GA, Saá P, Brodsky JP, et al. Characteristics of US Blood Donors Testing Reactive for Antibodies to SARS-CoV-2 Prior to the Availability of Authorized Vaccines. Transfusion Medicine Reviews. 2021;35: 1–7. doi:[10.1016/j.tmrv.2021.07.001](https://doi.org/10.1016/j.tmrv.2021.07.001)

302.	Staszewski C, Rochelson B, Krantz DA, Gerber R, Juhel H, Reddy S, et al. Persistence of infection-induced SARS-CoV-2 seropositivity throughout gestation. American Journal of Obstetrics and Gynecology. 2021; S0002-9378(21)01087-5. doi:[10.1016/j.ajog.2021.09.037](https://doi.org/10.1016/j.ajog.2021.09.037)

303.	Arnold CRK, Srinivasan S, Herzog CM, Gontu A, Bharti N, Small M, et al. SARS-CoV-2 Seroprevalence in a University Community: A Longitudinal Study of the Impact of Student Return to Campus on Infection Risk Among Community Members. medRxiv. 2021. doi:[10.1101/2021.02.17.21251942](https://doi.org/10.1101/2021.02.17.21251942)

304.	Jehn M, Pandit U, Sabin S, Tompkins C, White J, Kaleta E, et al. Accuracy of Case-Based Seroprevalence of SARS-CoV-2 Antibodies in Maricopa County, Arizona. American Journal of Public Health. 2022;112: 38–42. doi:[10.2105/AJPH.2021.306568](https://doi.org/10.2105/AJPH.2021.306568)

305.	Bonelli F, Blocki FA, Bunnell T, Chu E, O ADL, Grenache DG, et al. Evaluation of the automated LIAISON SARS-CoV-2 TrimericS IgG assay for the detection of circulating antibodies. Clinical Chemistry and Laboratory Medicine (CCLM). 2021. doi:[10.1515/cclm-2021-0023](https://doi.org/10.1515/cclm-2021-0023)

306.	Stout RL, Rigatti SJ. Seroprevalence of SARS-CoV-2 Antibodies in the US Adult Asymptomatic Population as of September 30, 2020. JAMA Network Open. 2021;4: e211552. doi:[10.1001/jamanetworkopen.2021.1552](https://doi.org/10.1001/jamanetworkopen.2021.1552)

307.	Gandra S, Li T, Reske KA, Dang NL, Farnsworth CW, Hock KG, et al. SARS-CoV-2 Infection Risk Factors among Maintenance Hemodialysis Patients and Health Care Personnel In Outpatient Hemodialysis Centers. Kidney360. 2021;2: 996–1001. doi:[10.34067/KID.0001282021](https://doi.org/10.34067/KID.0001282021)

308.	Symanski E, Ensor KB, Piedra PA, Sheth K, Caton K, Williams SL, et al. Population-Based Estimates of SARS-CoV-2 Seroprevalence in Houston, TX as of September 2020. The Journal of Infectious Diseases. 2021;224: 1649–1657. doi:[10.1093/infdis/jiab203](https://doi.org/10.1093/infdis/jiab203)

309.	Bahar B, Simpson JN, Biddle C, Campbell A, Dome JS, DeBiasi RL, et al. Estimated SARS-CoV-2 Seroprevalence in Healthy Children and Those with Chronic Illnesses in The Washington Metropolitan Area as of October 2020. medRxiv. 2021; 2021.01.30.21250830. doi:[10.1101/2021.01.30.21250830](https://doi.org/10.1101/2021.01.30.21250830)

310.	Jia X, Gesualdo P, Geno Rasmussen C, Alkanani AA, He L, Dong F, et al. Prevalence of SARS-CoV-2 Antibodies in Children and Adults with Type 1 Diabetes. Diabetes Technology & Therapeutics. 2021;23: 517–521. doi:[10.1089/dia.2020.0609](https://doi.org/10.1089/dia.2020.0609)

311.	Janevic T, Lieb W, Ibroci E, Lynch J, Lieber M, Molenaar NM, et al. The influence of structural racism, pandemic stress, and SARS-CoV-2 infection during pregnancy with adverse birth outcomes. American Journal of Obstetrics & Gynecology MFM. 2022;0. doi:[10.1016/j.ajogmf.2022.100649](https://doi.org/10.1016/j.ajogmf.2022.100649)

312.	Sherman A, Reuben J, David N, Quasie-Woode DP, Gunn JKL, Nielsen CF, et al. SARS-CoV-2 Seroprevalence Survey Among District Residents Presenting for Serologic Testing at Three Community-Based Test Sites Washington, DC, July, 2020. medRxiv. 2021. doi:[10.1101/2021.02.15.21251764](https://doi.org/10.1101/2021.02.15.21251764)

313.	Parrott JC, Maleki AN, Vassor VE, Osahan S, Hsin Y, Sanderson M, et al. Prevalence of SARS-CoV-2 Antibodies in New York City Adults, June 2020: A Population-Based Survey. The Journal of Infectious Diseases. 2021;224: 188–195. doi:[10.1093/infdis/jiab296](https://doi.org/10.1093/infdis/jiab296)

314.	Bruckner TA, Parker DM, Bartell SM, Vieira VM, Khan S, Noymer A, et al. Estimated Seroprevalence of SARS-CoV-2 Antibodies Among Adults in Orange County, California. medRxiv. 2020. doi:[10.1101/2020.10.07.20208660](https://doi.org/10.1101/2020.10.07.20208660)

315.	Feehan AK, Velasco C, Fort D, Burton JH, Price-Haywood EG, Katzmarzyk PT, et al. Racial and Workplace Disparities in Seroprevalence of SARS-CoV-2, Baton Rouge, Louisiana, USA. Emerging Infectious Diseases. 2021;27: 314–317. doi:[10.3201/eid2701.203808](https://doi.org/10.3201/eid2701.203808)

316.	Yamaki J, Peled H, Mathews S, Park D, Firoozi M, Smith K, et al. Seroprevalence of Novel Coronavirus SARS-CoV-2 at a Community Hospital Emergency Department and Outpatient Laboratory in Northern Orange County, California. Journal of Racial and Ethnic Health Disparities. 2020;8: 1551–1555. doi:[10.1007/s40615-020-00918-0](https://doi.org/10.1007/s40615-020-00918-0)

317.	Gonik CO, Alonso AM, Gonik B. SARS-CoV-2 Seroprevalence in Florida Department of Health in Palm Beach County Obstetric Clinics: A Cross-Sectional Study during the First Pandemic Surge. American Journal of Perinatology. 2021. doi:[10.1055/s-0041-1739353](https://doi.org/10.1055/s-0041-1739353)

318.	Steffen HA, Swartz SR, Jackson JB, Kenne KA, Eyck PPT, Merryman AS, et al. SARS-CoV-2 Infection during Pregnancy in a Rural Midwest All-delivery Cohort and Associated Maternal and Neonatal Outcomes. American Journal of Perinatology. 2021. doi:[10.1055/s-0041-1723938](https://doi.org/10.1055/s-0041-1723938)

319.	Molenaar NM, Rommel A-S, de Witte L, Dolan SM, Lieb W, Ibroci E, et al. SARS-CoV-2 during pregnancy and associated outcomes: Results from an ongoing prospective cohort. Paediatric and Perinatal Epidemiology. 2021;n/a. doi:[10.1111/ppe.12812](https://doi.org/10.1111/ppe.12812)

320.	Stadlbauer D, Tan J, Jiang K, Hernandez MM, Fabre S, Amanat F, et al. Repeated cross-sectional sero-monitoring of SARS-CoV-2 in New York City. Nature. 2020. doi:[10.1038/s41586-020-2912-6](https://doi.org/10.1038/s41586-020-2912-6)

321.	Robertson MM, Kulkarni SG, Rane M, Kochhar S, Berry A, Chang M, et al. Cohort profile: A national, community-based prospective cohort study of SARS-CoV-2 pandemic outcomes in the USAthe CHASING COVID Cohort study. BMJ Open. 2021;11: e048778. doi:[10.1136/bmjopen-2021-048778](https://doi.org/10.1136/bmjopen-2021-048778)

322.	Kamath K, Baum-Jones E, Jordan G, Haynes W, Waitz R, Shon J, et al. Prevalence of antibodies to SARS-CoV-2 in healthy blood donors in New York. medRxiv. 2020; 2020.10.19.20215368. doi:[10.1101/2020.10.19.20215368](https://doi.org/10.1101/2020.10.19.20215368)

323.	Rapsinski GJ, Freeman MC, Haidar G, Belle SH, Hasskamp JH, Wheeler SE. Pediatric SARS-CoV-2 seroprevalence during mitigation procedures in Southwestern Pennsylvania. Journal of Clinical Virology Plus. 2021;1: 100026. doi:[10.1016/j.jcvp.2021.100026](https://doi.org/10.1016/j.jcvp.2021.100026)

324.	Morozova O, Clouston SAP, Valentine J, Newman A, Carr M, Luft BJ. COVID-19 cumulative incidence, asymptomatic infections, and fatality in Long Island, NY, January 2020: A cohort of World Trade Center responders. Yon DK, editor. PLoS One. 2021;16: e0254713. doi:[10.1371/journal.pone.0254713](https://doi.org/10.1371/journal.pone.0254713)

325.	Barzin A, Schmitz JL, Rosin S, Sirpal R, Almond M, Robinette C, et al. SARS-CoV-2 Seroprevalence among a Southern U.S. Population Indicates Limited Asymptomatic Spread under Physical Distancing Measures. Schultz-Cherry S, editor. mBio. 2020;11: e02426–20. doi:[10.1128/mBio.02426-20](https://doi.org/10.1128/mBio.02426-20)

326.	Geisler D, Freeman MC, Rapsinski GJ, Wheeler SE. Unexpected False-Positive Rates in Pediatric SARS-CoV-2 Serology Using the EUROIMMUN Anti-SARS-CoV-2 ELISA IgG Assay. American Journal of Clinical Pathology. 2021;155: 773–775. doi:[10.1093/ajcp/aqab033](https://doi.org/10.1093/ajcp/aqab033)

327.	Reifer J, Hayum N, Heszkel B, Klagsbald I, Streva VA. SARS-CoV-2 IgG antibody responses in New York City. Diagnostic Microbiology and Infectious Disease. 2020;98: 115128. doi:[10.1016/j.diagmicrobio.2020.115128](https://doi.org/10.1016/j.diagmicrobio.2020.115128)

328.	Khatri M, Islam S, Dutka P, Carson J, Drakakis J, Imbriano L, et al. COVID-19 Antibodies and Outcomes among Outpatient Maintenance Hemodialysis Patients. Kidney360. 2021;2: 263–269. doi:[10.34067/KID.0006292020](https://doi.org/10.34067/KID.0006292020)

329.	Feehan AK, Fort D, Garcia-Diaz J, Price-Haywood EG, Velasco C, Sapp E, et al. Seroprevalence of SARS-CoV-2 and Infection Fatality Ratio, Orleans and Jefferson Parishes, Louisiana, USA, May 2020. Emerging Infectious Diseases. 2020;26: 2765–2768. doi:[10.3201/eid2611.203029](https://doi.org/10.3201/eid2611.203029)

330.	Routledge I, Epstein A, Takahashi S, Janson O, Hakim J, Duarte E, et al. Citywide serosurveillance of the initial SARS-CoV-2 outbreak in San Francisco using electronic health records. Nature Communications. 2021;12: 3566. doi:[10.1038/s41467-021-23651-6](https://doi.org/10.1038/s41467-021-23651-6)

331.	Flannery DD, Gouma S, Dhudasia MB, Mukhopadhyay S, Pfeifer MR, Woodford EC, et al. SARS-CoV-2 Seroprevalence Among Parturient Women. medRxiv. 2020. doi:[10.21203/rs.3.rs-27402/v1](https://doi.org/10.21203/rs.3.rs-27402/v1)

332.	Smith BK, Janowski AB, Danis JE, Harvey IB, Zhao H, Dai Y-N, et al. Seroprevalence of SARS-CoV-2 Antibodies in Children and Adults in St. Louis, Missouri, USA. mSphere. 2021;6: e01207–20. doi:[10.1128/mSphere.01207-20](https://doi.org/10.1128/mSphere.01207-20)

333.	Nesbitt DJ, Jin DP, Hogan JW, Yang J, Chen H, Chan PA, et al. Low Seroprevalence of SARS-CoV-2 in Rhode Island blood donors during may 2020 as determined using multiple serological assay formats. BMC Infectious Diseases. 2021;21: 871. doi:[10.1186/s12879-021-06438-4](https://doi.org/10.1186/s12879-021-06438-4)

334.	Biggs HM, Harris JB, Breakwell L, Dahlgren FS, Abedi GR, Szablewski CM, et al. Estimated Community Seroprevalence of SARS-CoV-2 Antibodies Two Georgia Counties, April 28 3, 2020. MMWR Morbidity and Mortality Weekly Report. 2020;69: 965–970. doi:[10.15585/mmwr.mm6929e2](https://doi.org/10.15585/mmwr.mm6929e2)

335.	Bajema KL, Dahlgren FS, Lim TW, Bestul N, Biggs HM, Tate JE, et al. Comparison of Estimated SARS-CoV-2 Seroprevalence through Commercial Laboratory Residual Sera Testing and a Community Survey. Clinical Infectious Diseases. 2020. doi:[10.1093/cid/ciaa1804](https://doi.org/10.1093/cid/ciaa1804)

336.	Laeyendecker O, Hsieh Y-H, Rothman RE, Dashler G, Kickler T, Fernandez RE, et al. Demographic and clinical correlates of acute and convalescent SARS-CoV-2 infection among patients of a U.S. Emergency department. The American Journal of Emergency Medicine. 2021;48: 261–268. doi:[10.1016/j.ajem.2021.04.081](https://doi.org/10.1016/j.ajem.2021.04.081)

337.	Sood N, Simon P, Ebner P, Eichner D, Reynolds J, Bendavid E, et al. Seroprevalence of SARS-CoV-2 in Los Angeles County, California, on April 10-11, 2020. The Journal of the American Medical Association. 2020;323: 2425–2427. doi:[10.1001/jama.2020.8279](https://doi.org/10.1001/jama.2020.8279)

338.	Rathe JA, Hemann EA, Eggenberger J, Li Z, Knoll ML, Stokes C, et al. SARS-CoV-2 Serologic Assays in Control and Unknown Populations Demonstrate the Necessity of Virus Neutralization Testing. The Journal of Infectious Diseases. 2021;223: 1120–1131. doi:[10.1093/infdis/jiaa797](https://doi.org/10.1093/infdis/jiaa797)

339.	Bendavid E, Mulaney B, Sood N, Shah S, Ling E, Bromley-Dulfano R, et al. COVID-19 Antibody Seroprevalence in Santa Clara County, California. medRxiv. 2020. doi:[10.1101/2020.04.14.20062463](https://doi.org/10.1101/2020.04.14.20062463)

340.	Dingens AS, Crawford KH, Adler A, Steele SL, Lacombe K, Eguia R, et al. Serological identification of SARS-CoV-2 infections among children visiting a hospital during the initial Seattle outbreak. medRxiv. 2020. doi:[10.1101/2020.05.26.20114124](https://doi.org/10.1101/2020.05.26.20114124)

341.	Ng DL, Goldgof GM, Shy BR, Levine AG, Balcerek J, Bapat SP, et al. SARS-CoV-2 seroprevalence and neutralizing activity in donor and patient blood from the San Francisco Bay Area. medRxiv. 2020. doi:[10.1101/2020.05.19.20107482](https://doi.org/10.1101/2020.05.19.20107482)

342.	Abdelmaksoud SS, Youssef MK, Aboulmagd MM, Abdelmaksoud MF. Seropositive coronavirus disease 2019 detection among apparently healthy voluntary blood donors: A preliminary step to catch asymptomatic cases as a potential viral spread route. Egyptian Journal of Haematology. 2021;46: 65–69. doi:[10.4103/ejh.ejh_59_20](https://doi.org/10.4103/ejh.ejh_59_20)

343.	Girgis SA, Hafez HM, Elarab HE, Sherif B, Sabry MH, Afifi I, et al. SARS-CoV-2 PCR positivity rate and seroprevalence of related antibodies among a sample of patients in Cairo: Pre-wave 2 results of a screening program in a university hospital. Blackard JT, editor. PLoS One. 2021;16: e0254581. doi:[10.1371/journal.pone.0254581](https://doi.org/10.1371/journal.pone.0254581)

344.	Mohammadi Z, Eghtesad S, Hashemi-Shahri SM, Tabatabaei SM, Sharafkhah M, Poustchi H. The Seroprevalence of COVID-19 in Intravenous Drug Users in Comparison to Non-drug Users. Middle East Journal of Digestive Diseases (MEJDD). 2021;13: 67–70. doi:[10.34172/mejdd.2021.206](https://doi.org/10.34172/mejdd.2021.206)

345.	Zandian H, Moghadam TZ, Habibzadeh S, Pourfarzi F. Seroprevalence of SARS-CoV-2 Antibodies and its Risk Factors in the North-West of Iran: A Population-Based Cross-Sectional Study. The Open Public Health Journal. 2022;15: e187494452201140. doi:[10.2174/18749445-v15-e2201140](https://doi.org/10.2174/18749445-v15-e2201140)

346.	Hussein NR, Balatay AA, Naqid IA, Jamal SA, Rasheed NA, Ahmed AN, et al. COVID-19 antibody seroprevalence in Duhok, Kurdistan Region, Iraq: A population-based study. medRxiv. 2021; 2021.03.23.21254169. doi:[10.1101/2021.03.23.21254169](https://doi.org/10.1101/2021.03.23.21254169)

347.	Sughayer MA, Mansour A, Nuirat AA, Souan L, Abdel-Razeq R, Siag M. A second dramatic rise in seroprevalence rates of SARS-CoV-2 antibodies among adult healthy blood donors in Jordan; have we achieved herd immunity? medRxiv. 2021. doi:[10.1101/2021.08.15.21261584](https://doi.org/10.1101/2021.08.15.21261584)

348.	Sughayer MA, Mansour A, Al Nuirat A, Souan L, Ghanem M, Siag M. Dramatic rise in seroprevalence rates of SARS-CoV-2 antibodies among healthy blood donors: The evolution of a pandemic. International Journal of Infectious Diseases. 2021;107: 116–120. doi:[10.1016/j.ijid.2021.04.059](https://doi.org/10.1016/j.ijid.2021.04.059)

349.	Qutob N, Awartani F, Salah Z, Asia M, Khader IA, Herzallah K, et al. Seroprevalence of SARS-CoV-2 in the West Bank region of Palestine: A cross-sectional seroepidemiological study. BMJ Open. 2021;11: e044552. doi:[10.1136/bmjopen-2020-044552](https://doi.org/10.1136/bmjopen-2020-044552)

350.	Nisar MI, Ansari N, Khalid F, Amin M, Shahbaz H, Hotwani A, et al. Serial population-based serosurveys for COVID-19 in two neighbourhoods of Karachi, Pakistan. International Journal of Infectious Diseases. 2021;106: 176–182. doi:[10.1016/j.ijid.2021.03.040](https://doi.org/10.1016/j.ijid.2021.03.040)

351.	Ansari S, Memon M, Kumar R, Memon S, Memon MK. Do COVID-19 Antibodies Provide Long-Term Protection? Cureus. 2021;13: e12441. doi:[10.7759/cureus.12441](https://doi.org/10.7759/cureus.12441)

352.	Javed W, Baqar JB, Abidi SHB, Farooq W. Sero-prevalence Findings from Metropoles in Pakistan: Implications for Assessing COVID-19 Prevalence and Case-fatality within a Dense, Urban Working Population. medRxiv. 2020; 2020.08.13.20173914. doi:[10.1101/2020.08.13.20173914](https://doi.org/10.1101/2020.08.13.20173914)

353.	Alosaimi MF, Alhetheel A, Aleisa KA, Altwerki AA, Alenezy NM, Almutairi EM, et al. Risk factors and predictors that influence SARS-Cov-2 IgG positivity: A cross-sectional study of blood donors in Riyadh, Saudi Arabia. Saudi Medical Journal. 2021;42: 853–861. doi:[10.15537/smj.2021.42.8.20210238](https://doi.org/10.15537/smj.2021.42.8.20210238)

354.	Almudarra S, Kamel S, Saleh E, Alaswad R, Alruwaily A, Almowald S, et al. High seroprevalence of SARS-CoV-2 among high-density communities in Saudi Arabia. Infection. 2021. doi:[10.1007/s15010-021-01726-2](https://doi.org/10.1007/s15010-021-01726-2)

355.	Raouf M, Rabeh M, Kaur S, Sharma R, Thottumkal N, Mohammed R. Seroprevalence of IgG Anti-SARS-CoV-2 among Voluntary Blood Donors in Dubai: Demographic and Risk Factors. Dubai Medical Journal. 2021;4: 204–211. doi:[10.1159/000517456](https://doi.org/10.1159/000517456)

356.	Bin-Ghouth AS, Al-Shoteri S, Mahmoud N, Musani A, Baoom NM, Al-Waleedi AA, et al. SARS-CoV-2 seroprevalence in Aden, Yemen: A population-based study. International Journal of Infectious Diseases. 2022;115: 239–244. doi:[10.1016/j.ijid.2021.12.330](https://doi.org/10.1016/j.ijid.2021.12.330)

357.	Sulcebe G, Ylli A, Cenko F, Kurti-Prifti M. Rapid increase of SARS-CoV-2 seroprevalence during the 2020 pandemic year in the population of the city of Tirana, Albania. medRxiv. 2021. doi:[10.1101/2021.02.18.21251776](https://doi.org/10.1101/2021.02.18.21251776)

358.	Ašić A, Prguda-Mujić J, Salihefendić L, Bešić L, Ler D, Čeko I, et al. Serological testing for SARS-CoV-2 in Bosnia and Herzegovina: First report. Archives of Medical Science. 2021;17: 823–826. doi:[10.5114/aoms/134143](https://doi.org/10.5114/aoms/134143)

359.	Tsaneva-Damyanova D. SARS-CoV-2: Seroepidemiological pattern in northeastern Bulgaria. Biotechnology & Biotechnological Equipment. 2020;34: 441–446. doi:[10.1080/13102818.2020.1772105](https://doi.org/10.1080/13102818.2020.1772105)

360.	Semenova Y, Kalmatayeva Z, Oshibayeva A, Mamyrbekova S, Kudirbekova A, Nurbakyt A, et al. Seropositivity of SARS-CoV-2 in the Population of Kazakhstan: A Nationwide Laboratory-Based Surveillance. International Journal of Environmental Research and Public Health. 2022;19: 2263. doi:[10.3390/ijerph19042263](https://doi.org/10.3390/ijerph19042263)

361.	Smagul M, Esmagambetova A, Nusupbaeva G, Kirpicheva U, Kasabekova L, Nukenova G, et al. Sero-prevalence of SARS-CoV-2 in certain cities of Kazakhstan. Health Science Reports. 2022;5. doi:[10.1002/hsr2.562](https://doi.org/10.1002/hsr2.562)

362.	T. Makarovska Bojadjieva, S. Useini, J. Nikolova, E. Ristovska, E. Velkova, E. Petkovic, et al. Seroprevalence of SARS-CoV-2 among blood donors. Vox Sanguinis. 2021;116: 5–188. doi:[10.1111/vox.13117](https://doi.org/10.1111/vox.13117)

363.	Kuban State Medical University, Krasnodar, Russian Federation, Gorodin VN, Redko AN, Kuban State Medical University, Krasnodar, Russian Federation, Skoblikov NE, Kuban State Medical University, Krasnodar, Russian Federation, et al. Dynamics of seroprevalence of anti-SARS-CoV-2 antibodies in non-vaccinated people. Infekcionnye bolezni. 2021;19: 98–102. doi:[10.20953/1729-9225-2021-4-98-102](https://doi.org/10.20953/1729-9225-2021-4-98-102)

364.	Barchuk A, Skougarevskiy D, Kouprianov A, Shirokov D, Dudkina O, Tursun-zade R, et al. COVID-19 pandemic in Saint Petersburg, Russia: Combining surveillance and population-based serological study data in May, 2020, 2021. medRxiv. 2021. doi:[10.1101/2021.07.31.21261428](https://doi.org/10.1101/2021.07.31.21261428)

365.	Barchuk A, Skougarevskiy D, Titaev K, Shirokov D, Raskina Y, Novkunkskaya A, et al. Seroprevalence of SARS-CoV-2 antibodies in Saint Petersburg, Russia: A population-based study. Scientific Reports. 2021;11: 12930. doi:[10.1038/s41598-021-92206-y](https://doi.org/10.1038/s41598-021-92206-y)

366.	Celik OY, Çelen Ş, Üstün YE. Did the SARS-CoV-2 effect pregnancy complications? Ginekologia Polska. 2021;0. doi:[10.5603/GP.a2021.0134](https://doi.org/10.5603/GP.a2021.0134)

367.	Siller A, Seekircher L, Wachter GA, Astl M, Tschiderer L, Pfeifer B, et al. Seroprevalence, Waning and Correlates of Anti-SARS-CoV-2 IgG Antibodies in Tyrol, Austria: Large-Scale Study of 35,193 Blood Donors Conducted between June 2020 and September 2021. Viruses. 2022;14: 568. doi:[10.3390/v14030568](https://doi.org/10.3390/v14030568)

368.	Riesenhuber M, Nitsche C, Binder CJ, Schernhammer ES, Stamm T, Anwari E, et al. Seroprevalence of SARS-CoV-2 antibodies in healthcare workers and an unselected all comer patient population. Wiener klinische Wochenschrift. 2021. pp. 33–154. doi:[10.1007/s00508-021-01884-1](https://doi.org/10.1007/s00508-021-01884-1)

369.	Rebholz H, Braun RJ, Saha T, Harzer O, Schneider M, Ladage D. Longitudinal monitoring of SARS-CoV-2-specific immune responses. Cold Spring Harbor Laboratory Press; 2021. p. 2021.08.14.21262042. doi:[10.1101/2021.08.14.21262042](https://doi.org/10.1101/2021.08.14.21262042)

370.	Weidner L, Nunhofer V, Jungbauer C, Hoeggerl AD, Grüner L, Grabmer C, et al. Seroprevalence of anti-SARS-CoV-2 total antibody is higher in younger Austrian blood donors. Infection. 2021;49: 1187–1194. doi:[10.1007/s15010-021-01639-0](https://doi.org/10.1007/s15010-021-01639-0)

371.	Borena W, Bánki Z, Bates K, Winner H, Riepler L, Rössler A, et al. Persistence of immunity to SARS-CoV-2 over time in the ski resort Ischgl. eBioMedicine. 2021;70: 103534. doi:[10.1016/j.ebiom.2021.103534](https://doi.org/10.1016/j.ebiom.2021.103534)

372.	Ladage D, Höglinger Y, Ladage D, Adler C, Yalcin I, Harzer O, et al. SARS-CoV-2-Specific Antibody Prevalence and Symptoms in a Local Austrian Population. Frontiers in Medicine. 2021;8: 632942. doi:[10.3389/fmed.2021.632942](https://doi.org/10.3389/fmed.2021.632942)

373.	Lehmann J, Giesinger JM, Rumpold G, Borena W, Knabl L, Falkensammer B, et al. Estimating seroprevalence of SARS-CoV-2 antibodies using three self-reported symptoms: Development of a prediction model based on data from Ischgl, Austria. Epidemiology and Infection. 2021;149: e52. doi:[10.1017/S0950268821000418](https://doi.org/10.1017/S0950268821000418)

374.	Heireman L, Baetens M, Menten B, Dehaene I, Padalko E. Comparison of the positivity rate of anti-spike and anti-nucleocapsid SARS-CoV-2 IgG in asymptomatic pregnant women. Journal of Obstetrics and Gynaecology. 2021; 1–2. doi:[10.1080/01443615.2021.1997960](https://doi.org/10.1080/01443615.2021.1997960)

375.	van Dam P, Huizing M, Roelant E, Hotterbeekx A, De Winter FHR, Kumar-Singh S, et al. Immunoglobin G/total antibody testing for SARS-CoV-2: A prospective cohort study of ambulatory patients and health care workers in two Belgian oncology units comparing three commercial tests. European Journal of Cancer (Oxford, England: 1990). 2021;148: 328–339. doi:[10.1016/j.ejca.2021.02.024](https://doi.org/10.1016/j.ejca.2021.02.024)

376.	Vilibic-Cavlek T, Stevanovic V, Ilic M, Barbic L, Capak K, Tabain I, et al. SARS-CoV-2 Seroprevalence and Neutralizing Antibody Response after the First and Second COVID-19 Pandemic Wave in Croatia. Pathogens. 2021;10: 774. doi:[10.3390/pathogens10060774](https://doi.org/10.3390/pathogens10060774)

377.	Lenicek Krleza J, Zrinski Topic R, Stevanovic V, Lukic-Grlic A, Tabain I, Misak Z, et al. Seroprevalence of SARS-CoV-2 infection among children in Children’s Hospital Zagreb during the initial and second wave of COVID-19 pandemic in Croatia. Biochemia Medica. 2021;31: 020706. doi:[10.11613/BM.2021.020706](https://doi.org/10.11613/BM.2021.020706)

378.	Piler P, Thon V, Andrýsková L, Doležel K, Kostka D, Pavlík T, et al. Nationwide increases in anti-SARS-CoV-2 IgG antibodies between October 2020 and March 2021 in the unvaccinated Czech population. Communications Medicine. 2022;2: 1–7. doi:[10.1038/s43856-022-00080-0](https://doi.org/10.1038/s43856-022-00080-0)

379.	Bloomfield M, Pospisilova I, Cabelova T, Sediva A, Ibrahimova M, Borecka K, et al. Searching for COVID-19 Antibodies in Czech Children in the Haystack. Frontiers in Pediatrics. 2020;8. doi:[10.3389/fped.2020.597736](https://doi.org/10.3389/fped.2020.597736)

380.	Jespersen S, Mikkelsen S, Greve T, Kaspersen KA, Tolstrup M, Boldsen JK, et al. Severe Acute Respiratory Syndrome Coronavirus 2 Seroprevalence Survey Among 17 971 Healthcare and Administrative Personnel at Hospitals, Prehospital Services, and Specialist Practitioners in the Central Denmark Region. Clinical Infectious Diseases. 2021;73: e2853–e2860. doi:[10.1093/cid/ciaa1471](https://doi.org/10.1093/cid/ciaa1471)

381.	Egerup P, Fich Olsen L, Christiansen A-MH, Westergaard D, Severinsen ER, Hviid KVR, et al. Severe Acute Respiratory Syndrome Coronavirus 2 (SARS-CoV-2) Antibodies at Delivery in Women, Partners, and Newborns. Obstetrics & Gynecology. 2021;137: 49–55. doi:[10.1097/AOG.0000000000004199](https://doi.org/10.1097/AOG.0000000000004199)

382.	Soeorg H, Jõgi P, Naaber P, Ottas A, Toompere K, Lutsar I. Seroprevalence and levels of IgG antibodies after COVID-19 infection or vaccination. Infectious Diseases. 2022;54: 63–71. doi:[10.1080/23744235.2021.1974540](https://doi.org/10.1080/23744235.2021.1974540)

383.	Veerus P, Salumets A, Naaber P, Krjutškov K, Tilk K, Laanpere M, et al. Seroprevalence of SARS-CoV-2 antibodies among pregnant women in Estonia: A call for epidemiological studies. Acta Obstetricia Et Gynecologica Scandinavica. 2020;99: 1736–1737. doi:[10.1111/aogs.13995](https://doi.org/10.1111/aogs.13995)

384.	Ayrolles A, Ellul P, Trebossen V, Houhou-Fidouh N, Bonacorsi S, Descamps D, et al. Is SARS-CoV-2 seroconversion a risk factor for severe and acute psychiatric symptoms in children? Neuropsychopharmacology Reports. 2022;n/a. doi:[10.1002/npr2.12236](https://doi.org/10.1002/npr2.12236)

385.	Carrat F, Lapidus N, Ninove L, Blanché H, Rahib D, Saba Villarroel PM, et al. Age, COVID-19-like symptoms and SARS-CoV-2 seropositivity profiles after the first wave of the pandemic in France. Infection. 2022;50: 257–262. doi:[10.1007/s15010-021-01731-5](https://doi.org/10.1007/s15010-021-01731-5)

386.	Gégout Petit A, Jeulin H, Legrand K, Jay N, Bochnakian A, Vallois P, et al. Seroprevalence of SARS-CoV-2, symptom profiles and sero-neutralization in a suburban area, France. Viruses. 2021;13: 1076. doi:[10.3390/v13061076](https://doi.org/10.3390/v13061076)

387.	Beaumont A, Durand C, Ledrans M, Schwoebel V, Noel H, Strat YL, et al. Seroprevalence of anti-SARS-CoV-2 antibodies after the first wave of the COVID-19 pandemic in a vulnerable population in France: A cross-sectional study. BMJ Open. 2021;11: e053201. doi:[10.1136/bmjopen-2021-053201](https://doi.org/10.1136/bmjopen-2021-053201)

388.	Woudenberg T, Pelleau S, Anna F, Attia M, Donnadieu F, Gravet A, et al. Humoral immunity to SARS-CoV-2 and seasonal coronaviruses in children and adults in north-eastern France. EBioMedicine. 2021;70: 103495. doi:[10.1016/j.ebiom.2021.103495](https://doi.org/10.1016/j.ebiom.2021.103495)

389.	Tsatsaris V, Mariaggi A-A, Launay O, Couffignal C, Rousseau J, Ancel PY, et al. SARS-COV-2 IgG antibody response in pregnant women at delivery. Journal of Gynecology Obstetrics and Human Reproduction. 2020;50: 102041. doi:[10.1016/j.jogoh.2020.102041](https://doi.org/10.1016/j.jogoh.2020.102041)

390.	Mattern J, Vauloup-Fellous C, Zakaria H, Benachi A, Carrara J, Letourneau A, et al. Post lockdown COVID-19 seroprevalence and circulation at the time of delivery, France. PLoS One. 2020;15: e0240782. doi:[10.1371/journal.pone.0240782](https://doi.org/10.1371/journal.pone.0240782)

391.	Capai L, Ayhan N, Masse S, Canarelli J, Priet S, Simeoni M-H, et al. Seroprevalence of SARS-CoV-2 IgG Antibodies in Corsica (France), April and June 2020. Journal of Clinical Medicine. 2020;9: 3569. doi:[10.3390/jcm9113569](https://doi.org/10.3390/jcm9113569)

392.	Cohen R, Jung C, Ouldali N, Sellam A, Batard C, Cahn-Sellem F, et al. Assessment of SARS-CoV-2 infection by Reverse transcription-PCR and serology in the Paris area: A cross-sectional study. BMJ Paediatrics Open. 2020;4: e000887. doi:[10.1136/bmjpo-2020-000887](https://doi.org/10.1136/bmjpo-2020-000887)

393.	Wachter F, Regensburger AP, Peter AS, Knieling F, Wagner AL, Simon D, et al. Continuous monitoring of SARS-CoV-2 seroprevalence in children using residual blood samples from routine clinical chemistry. Clinical Chemistry and Laboratory Medicine (CCLM). 2022. doi:[10.1515/cclm-2022-0037](https://doi.org/10.1515/cclm-2022-0037)

394.	Fischer B, Vollmer T, Knabbe C. SARS-CoV-2 IgG seroprevalence in blood donors located in three different federal states, Germany, July 2020 to June 2021 a follow-up. medRxiv. 2022; 2022.02.11.22270833. doi:[10.1101/2022.02.11.22270833](https://doi.org/10.1101/2022.02.11.22270833)

395.	Richter E, Liebl D, Schulte B, Lehmann N, Fuhrmann C, Jöckel K-H, et al. Analysis of Fatality Impact and Seroprevalence Surveys in a Community Sustaining a SARS-CoV-2 Superspreading Event. medRxiv; 2022. p. 2022.01.26.22269805. doi:[10.1101/2022.01.26.22269805](https://doi.org/10.1101/2022.01.26.22269805)

396.	Radon K, Bakuli A, Pütz P, Gleut RL, Noller JMG, Olbrich L, et al. From first to second wave: Follow-up of the prospective Covid-19 cohort (KoCo19) in Munich (Germany). medRxiv. 2021; 2021.04.27.21256133. doi:[10.1101/2021.04.27.21256133](https://doi.org/10.1101/2021.04.27.21256133)

397.	Robert Koch Institut. RKI - CORONA MONITORING local - Corona monitoring local. Robert Koch Institut. 2021.

398.	Brinkmann F, Diebner HH, Matenar C, Schlegtendal A, Spiecker J, Eitner L, et al. Longitudinal Rise in Seroprevalence of SARS-CoV-2 Infections in Children in Western Germany in Epidemiology? Infectious Disease Reports. 2021;13: 957–964. doi:[10.3390/idr13040088](https://doi.org/10.3390/idr13040088)

399.	Robert Koch Institut. RKI - CORONA MONITORING local - Corona monitoring local. Robert Koch Institut. 2020.

400.	Eichner FA, Gelbrich G, Weißbrich B, Dölken L, Kurzai O, Deckert J, et al. Seroprävalenz von COVID-19 und psychosoziale Auswirkungen in der Allgemeinbevölkerung: Ergebnisse des STAAB-COVID-One Programms. Das Gesundheitswesen. 2021;83: 965–975. doi:[10.1055/a-1630-7601](https://doi.org/10.1055/a-1630-7601)

401.	Hippich M, Holthaus L, Assfalg R, Zapardiel-Gonzalo J, Kapfelsperger H, Heigermoser M, et al. A Public Health Antibody Screening Indicates a 6-Fold Higher SARS-CoV-2 Exposure Rate than Reported Cases in Children. Med. 2021;2: 149–163.e4. doi:[10.1016/j.medj.2020.10.003](https://doi.org/10.1016/j.medj.2020.10.003)

402.	Wagner R, Peterhoff D, Beileke S, Guenther F, Berr M, Einhauser S, et al. Estimates and determinants of SARS-CoV-2 seroprevalence and infection fatality ratio using latent class analysis: The population-based Tirschenreuth study in the hardest-hit German county in spring 2020. medRxiv. 2021. doi:[10.1101/2021.03.29.21254343](https://doi.org/10.1101/2021.03.29.21254343)

403.	Dunay GA, Barroso M, Woidy M, Danecka MK, Engels G, Hermann K, et al. Long-term antibody response to SARS-CoV-2 in children. Infectious Diseases (except HIV/AIDS); 2022 Feb. doi:[10.1101/2022.02.11.22270611](https://doi.org/10.1101/2022.02.11.22270611)

404.	Santos-Hövener C, Neuhauser HK, Rosario AS, Busch M, Schlaud M, Hoffmann R, et al. Serology- and PCR-based cumulative incidence of SARS-CoV-2 infection in adults in a successfully contained early hotspot (CoMoLo study), Germany, May to June 2020. Eurosurveillance. 2020;25: 2001752. doi:[10.2807/1560-7917.ES.2020.25.47.2001752](https://doi.org/10.2807/1560-7917.ES.2020.25.47.2001752)

405.	Aziz NA, Corman VM, Echterhoff AKC, Müller MA, Richter A, Schmandke A, et al. Seroprevalence and correlates of SARS-CoV-2 neutralizing antibodies from a population-based study in Bonn, Germany. Nature Communications. 2021;12: 2117. doi:[10.1038/s41467-021-22351-5](https://doi.org/10.1038/s41467-021-22351-5)

406.	Pritsch M, Radon K, Bakuli A, Le Gleut R, Olbrich L, Guggenbuehl Noller JM, et al. Prevalence and Risk Factors of Infection in the Representative COVID-19 Cohort Munich. Rochester, NY: Social Science Research Network; 2021 Jan. Report No.: ID 3745128. doi:[10.2139/ssrn.3745128](https://doi.org/10.2139/ssrn.3745128)

407.	Runkel S, Kowalzik F, Gehring S, Winter J, Grandt CL, Marron M, et al. Prevalence of Severe Acute Respiratory Syndrome Coronavirus-2-specific Antibodies in German Blood Donors during the COVID-19 Pandemic. Clinical laboratory. 2020;66. doi:[10.7754/Clin.Lab.2020.200915](https://doi.org/10.7754/Clin.Lab.2020.200915)

408.	Zöllkau J, Baier M, Scherag A, Schleußner E, Groten T. Periodenprävalenz von SARS-CoV-2 in einer unselektierten Stichprobe schwangerer Frauen in Jena, Thüringen. Zeitschrift für Geburtshilfe und Neonatologie. 2020;224: 194–198. doi:[10.1055/a-1206-1033](https://doi.org/10.1055/a-1206-1033)

409.	Häusler S, Weigl M, Ambrosch A, Gruber R, Seelbach-Göbel B, Malfertheiner SF. Peripartal anti-SARS-CoV-2-IgA/IgG in asymptomatic pregnant women during regional SARS-CoV-2-outbreak. Journal of Perinatal Medicine. 2021. doi:[10.1515/jpm-2021-0001](https://doi.org/10.1515/jpm-2021-0001)

410.	Streeck H, Schulte B, Kümmerer BM, Richter E, Höller T, Fuhrmann C, et al. Infection fatality rate of SARS-CoV2 in a super-spreading event in Germany. Nature Communications. 2020;11: 5829. doi:[10.1038/s41467-020-19509-y](https://doi.org/10.1038/s41467-020-19509-y)

411.	Maltezou HC, Krumbholz B, Mavrouli M, Tseroni M, Gamaletsou MN, Botsa E, et al. A study of the evolution of the third COVID-19 pandemic wave in the Athens metropolitan area, Greece, through two cross-sectional seroepidemiological surveys: March, June 2021. Journal of Medical Virology. 2021;n/a. doi:[10.1002/jmv.27465](https://doi.org/10.1002/jmv.27465)

412.	Kotsiou OS, Papagiannis D, Fradelos EC, Siachpazidou DI, Perlepe G, Miziou A, et al. Defining Antibody Seroprevalence and Duration of Humoral Responses to SARS-CoV-2 Infection and/or Vaccination in a Greek Community. International Journal of Environmental Research and Public Health. 2022;19: 407. doi:[10.3390/ijerph19010407](https://doi.org/10.3390/ijerph19010407)

413.	Kotsiou OS, Papagiannis D, Fradelos EC, Perlepe G, Miziou A, Siachpazidou DS, et al. Understanding COVID-19 Epidemiology and Implications for Control: The Experience from a Greek Semi-Closed Community. Journal of Clinical Medicine. 2021;10: 2765. doi:[10.3390/jcm10132765](https://doi.org/10.3390/jcm10132765)

414.	Bogogiannidou Z, Vontas A, Dadouli K, Kyritsi MA, Soteriades S, Nikoulis DJ, et al. Repeated leftover serosurvey of SARS-CoV-2 IgG antibodies, Greece, March and April 2020. Eurosurveillance. 2020;25: 2001369. doi:[10.2807/1560-7917.ES.2020.25.31.2001369](https://doi.org/10.2807/1560-7917.ES.2020.25.31.2001369)

415.	Gudbjartsson DF, Norddahl GL, Melsted P, Gunnarsdottir K, Holm H, Eythorsson E, et al. Humoral Immune Response to SARS-CoV-2 in Iceland. New England Journal of Medicine. 2020;383: 1724–1734. doi:[10.1056/NEJMoa2026116](https://doi.org/10.1056/NEJMoa2026116)

416.	Heavey L, Garvey P, Colgan AM, Thornton L, Connell J, Roux T, et al. The Study to Investigate COVID-19 Infection in People Living in Ireland (SCOPI): A seroprevalence study, June to July 2020. Eurosurveillance. 2021;26: 2001741. doi:[10.2807/1560-7917.ES.2021.26.48.2001741](https://doi.org/10.2807/1560-7917.ES.2021.26.48.2001741)

417.	Breuer A, Raphael A, Stern H, Odeh M, Fiszlinski J, Algur N, et al. SARS-CoV-2 antibodies started to decline just four months after COVID-19 infection in a paediatric population. Acta Paediatrica. 2021;110: 3054–3062. doi:[10.1111/apa.16031](https://doi.org/10.1111/apa.16031)

418.	Gozlan Y, Reingold S, Koren R, Halpern O, Regev-Yochay G, Cohen C, et al. SARS-CoV-2 outbreak in a synagogue community: Longevity and strength of anti-SARS-CoV-2 IgG responses. Epidemiology and Infection. 2021; 1–14. doi:[10.1017/S0950268821001369](https://doi.org/10.1017/S0950268821001369)

419.	Ekka Zohar A, Kertes J, Cohen-Lunger E, Novikov I, Shamir Stein N, Hermoni Alon S, et al. COVID-19 Seropositive Rates between the Waves, Israel. The Israel Medical Association journal: IMAJ. 2021;23: 611–614.

420.	Antonucci F, Fiore JR, De Feo L, Granato T, Di Stefano M, Faleo G, et al. Increased SARS-CoV-2 seroprevalence in healthy blood donors after the second pandemic wave in South-Eastern Italy: Evidence for asymptomatic young donors as potential virus spreaders. Infectious Diseases. 2022;54: 241–246. doi:[10.1080/23744235.2021.2003856](https://doi.org/10.1080/23744235.2021.2003856)

421.	Polvere I, Parrella A, Casamassa G, D’Andrea S, Tizzano A, Cardinale G, et al. Seroprevalence of Anti-SARS-CoV-2 IgG and IgM among Adults over 65 Years Old in the South of Italy. Diagnostics. 2021;11: 483. doi:[10.3390/diagnostics11030483](https://doi.org/10.3390/diagnostics11030483)

422.	Melotti R, Scaggiante F, Falciani M, Weichenberger CX, Foco L, Lombardo S, et al. Prevalence and determinants of serum antibodies to SARS-CoV-2 in the general population of the Gardena Valley. medRxiv. 2021; 2021.03.19.21253883. doi:[10.1101/2021.03.19.21253883](https://doi.org/10.1101/2021.03.19.21253883)

423.	Pagani G, Giacomelli A, Conti F, Bernacchia D, Rondanin R, Prina A, et al. Prevalence of SARS-CoV-2 in an area of unrestricted viral circulation: Mass seroepidemiological screening in Castiglione d’Adda, Italy. Hozbor DF, editor. PLoS One. 2021;16: e0246513. doi:[10.1371/journal.pone.0246513](https://doi.org/10.1371/journal.pone.0246513)

424.	Cerino P, Coppola A, Volzone P, Pizzolante A, Pierri B, Atripaldi L, et al. Seroprevalence of SARS-CoV-2-specific antibodies in the town of Ariano Irpino (Avellino, Campania, Italy): A population-based study. Future Science OA. 2021;7: FSO673. doi:[10.2144/fsoa-2020-0203](https://doi.org/10.2144/fsoa-2020-0203)

425.	Fiore JR, Centra M, De Carlo A, Granato T, Rosa A, Sarno M, et al. Results from a survey in healthy blood donors in South Eastern Italy indicate that we are far away from herd immunity to SARS-CoV-2. Journal of Medical Virology. 2021;93: 1739–1742. doi:[10.1002/jmv.26425](https://doi.org/10.1002/jmv.26425)

426.	Dorigatti I, Lavezzo E, Manuto L, Ciavarella C, Pacenti M, Boldrin C, et al. SARS-CoV-2 antibody dynamics and transmission from community-wide serological testing in the Italian municipality of Vo’. Nature Communications. 2021;12: 4383. doi:[10.1038/s41467-021-24622-7](https://doi.org/10.1038/s41467-021-24622-7)

427.	Cassaniti I, Percivalle E, Sarasini A, Cambiè G, Nepita EV, Maserati R, et al. Seroprevalence of SARS-CoV-2 in blood donors from the Lodi Red Zone and adjacent Lodi metropolitan and suburban area. Clinical Microbiology and Infection. 2021;0. doi:[10.1016/j.cmi.2021.01.030](https://doi.org/10.1016/j.cmi.2021.01.030)

428.	Guerriero M, Bisoffi Z, Poli A, Micheletto C, Conti A, Pomari C. Prevalence of SARS-CoV-2, Verona, Italy, April 2020 - Volume 27, Number 1 2021 - Emerging Infectious Diseases journal - CDC. Emerging Infectious Diseases. 2020. doi:[10.3201/eid2701.202740](https://doi.org/10.3201/eid2701.202740)

429.	Baracco A, Perotti G, Filippin A, Anesi A, Beccarini V, Raimondi L, et al. SARS-CoV-2 Antibody Prevalence in Health Care Workers of Lodi Hospital, the COVID-19 Italian Epicentre. Rochester, NY: Social Science Research Network; 2020 Jul. Report No.: ID 3650227.

430.	Cavaliere AF, Carabaneanu AI, Perelli F, Matarrese D, Brunelli T, Casprini P, et al. Universal screening for SARS-CoV-2 in pregnant women admitted for delivery: How to manage antibody testing? The Journal of Maternal-Fetal & Neonatal Medicine. 2020; 1–2. doi:[10.1080/14767058.2020.1793317](https://doi.org/10.1080/14767058.2020.1793317)

431.	Cito F, Amato L, Di Giuseppe A, Danzetta ML, Iannetti S, Petrini A, et al. A COVID-19 Hotspot Area: Activities and Epidemiological Findings. Microorganisms. 2020;8: 1711. doi:[10.3390/microorganisms8111711](https://doi.org/10.3390/microorganisms8111711)

432.	Valenti L, Bergna A, Pelusi S, Facciotti F, Lai A, Tarkowski M, et al. SARS-CoV-2 seroprevalence trends in healthy blood donors during the COVID-19 outbreak in Milan. Blood Transfusion. 2021;19: 181–189. doi:[10.2450/2021.0324-20](https://doi.org/10.2450/2021.0324-20)

433.	Pancrazzi A, Magliocca P, Lorubbio M, Vaggelli G, Galano A, Mafucci M, et al. Comparison of serologic and molecular SARS-CoV 2 results in a large cohort in Southern Tuscany demonstrates a role for serologic testing to increase diagnostic sensitivity. Clinical Biochemistry. 2020;84: 87–92. doi:[10.1016/j.clinbiochem.2020.07.002](https://doi.org/10.1016/j.clinbiochem.2020.07.002)

434.	Coyer L, Boyd A, Schinkel J, Agyemang C, Galenkamp H, Koopman ADM, et al. Differences in SARS-CoV-2 infections during the first and second wave of SARS-CoV-2 between six ethnic groups in Amsterdam, the Netherlands: A population-based longitudinal serological study. The Lancet Regional Health - Europe. 2022;13: 100284. doi:[10.1016/j.lanepe.2021.100284](https://doi.org/10.1016/j.lanepe.2021.100284)

435.	Pagen DM, Brinkhues S, Dukers-Muijrers NH, Heijer CD den, Bouwmeester-Vincken N, Hanssen DA, et al. Exposure Factors Associated With SARS-CoV-2 Seroprevalence During the First Eight Months of the COVID-19 Pandemic in the Netherlands: A Cross-sectional Study. Research Square. 2021. doi:[10.21203/rs.3.rs-1159143/v1](https://doi.org/10.21203/rs.3.rs-1159143/v1)

436.	Rotee ILM, Ong DSY, Koeleman JGM, Vos ERA, Tramper-Stranders GA. Trends in SARS-CoV-2 seroprevalence amongst urban paediatric patients compared with a nationwide cohort in the Netherlands. Journal of Clinical Virology Plus. 2021;1: 100045. doi:[10.1016/j.jcvp.2021.100045](https://doi.org/10.1016/j.jcvp.2021.100045)

437.	Coyer L, Boyd A, Schinkel J, Agyemang C, Galenkamp H, Koopman ADM, et al. SARS-CoV-2 antibody prevalence and correlates of six ethnic groups living in Amsterdam, the Netherlands: A population-based cross-sectional study, June 2020. BMJ Open. 2022;12: e052752. doi:[10.1136/bmjopen-2021-052752](https://doi.org/10.1136/bmjopen-2021-052752)

438.	Keuning MW, Grobben M, de Groen A-EC, Berman-de Jong EP, Bijlsma MW, Cohen S, et al. Saliva SARS-CoV-2 Antibody Prevalence in Children. Microbiology Spectrum. 2021;9: e00731–21. doi:[10.1128/Spectrum.00731-21](https://doi.org/10.1128/Spectrum.00731-21)

439.	Slot E, Hogema BM, Reusken CBEM, Reimerink JH, Molier M, Karregat JHM, et al. Low SARS-CoV-2 seroprevalence in blood donors in the early COVID-19 epidemic in the Netherlands. Nature Communications. 2020;11: 5744. doi:[10.1038/s41467-020-19481-7](https://doi.org/10.1038/s41467-020-19481-7)

440.	Eskild A, Mørkrid L, Mortensen SB, Leegaard TM. Prevalence of antibodies against SARS-CoV-2 among pregnant women in Norway during the period December 2019 through December 2020. Epidemiology and Infection. 2022; 1–9. doi:[10.1017/S0950268822000073](https://doi.org/10.1017/S0950268822000073)

441.	Zejda JE, Kowalska M, Brożek GM, Barański K, Kaleta-Pilarska A. Infection Fatality Rate (IFR) during the course of COVID-19 pandemic in Upper Silesia Metropolitan Area (Poland) in 2020. Medycyna Pracy. 2021. doi:[10.13075/mp.5893.01179](https://doi.org/10.13075/mp.5893.01179)

442.	Olariu TR, Lighezan R, Ursoniu S, Craciun AC, Mihu AG, Lupu MA. High SARS-CoV-2 seroprevalence in blood donors from Romania after the third COVID-19 pandemic wave. Infectious Diseases. 2022; 1–4. doi:[10.1080/23744235.2022.2036811](https://doi.org/10.1080/23744235.2022.2036811)

443.	Olariu TR, Craciun AC, Vlad DC, Dumitrascu V, Pop LL, Horhat F, et al. SARS-CoV-2 Seroprevalence in Children from Western Romania, March to June 2021. Vector-Borne and Zoonotic Diseases. 2022;22. doi:[10.1089/vbz.2022.0003](https://doi.org/10.1089/vbz.2022.0003)

444.	Olariu TR, Lighezan R, Ursoniu S, Craciun AC, Paduraru AA, Lupu MA. Seroprevalence of SARS-CoV-2 antibodies in 2115 blood donors from Romania. Clinical Microbiology and Infection: The Official Publication of the European Society of Clinical Microbiology and Infectious Diseases. 2021;27: 817–819. doi:[10.1016/j.cmi.2020.12.027](https://doi.org/10.1016/j.cmi.2020.12.027)

445.	Karachaliou M, Moncunill G, Espinosa A, Castaño-Vinyals G, Rubio R, Vidal M, et al. SARS-CoV-2 infection, vaccination and antibody response trajectories in adults: A cohort study in Catalonia. In Review; 2022 Apr. doi:[10.21203/rs.3.rs-1536936/v1](https://doi.org/10.21203/rs.3.rs-1536936/v1)

446.	Soriano V, Ganado-Pinilla P, Sanchez-Santos M, Gómez-Gallego F, Barreiro P, de Mendoza C, et al. Main differences between the first and second waves of COVID-19 in Madrid, Spain. International Journal of Infectious Diseases. 2021;105: 374–376. doi:[10.1016/j.ijid.2021.02.115](https://doi.org/10.1016/j.ijid.2021.02.115)

447.	Kogevinas M, Casta ño-VG, Karachaliou M, Espinosa A, de CR, Garcia -AJ, et al. Ambient Air Pollution in Relation to SARS-CoV-2 Infection, Antibody Response, and COVID-19 Disease: A Cohort Study in Catalonia, Spain (COVICAT Study). Environmental Health Perspectives. 2021;129: 117003. doi:[10.1289/EHP9726](https://doi.org/10.1289/EHP9726)

448.	Viñuela MC, De León-Luis JA, Alonso R, Catalán P, Lizarraga S, Muñoz P, et al. SARS-CoV-2 screening of asymptomatic women admitted for delivery must be performed with a combination of microbiological techniques: An observational study. Revista Española de Quimioterapia. 2020;33: 415–421. doi:[10.37201/req/088.2020](https://doi.org/10.37201/req/088.2020)

449.	Crovetto F, Crispi F, Llurba E, Pascal R, Larroya M, Trilla C, et al. Impact of SARS-CoV-2 Infection on Pregnancy Outcomes: A Population-Based Study. Clinical Infectious Diseases. 2021. doi:[10.1093/cid/ciab104](https://doi.org/10.1093/cid/ciab104)

450.	Alenda R, Gonzalez-Diez R, Richart A, Montero A, Ruiz-Tovar M, Rodriguez Gambarte J, et al. P-216 Anti SARS-CoV-2 antibodies prevalence in Madrid blood donors prior to first outbreak. Vox Sanguinis. 2021;116: 5–188.

451.	Lundkvist Å, Hanson S, Olsen B. Pronounced difference in Covid-19 antibody prevalence indicates cluster transmission in Stockholm, Sweden. Infection Ecology & Epidemiology. 2020;10: 1806505. doi:[10.1080/20008686.2020.1806505](https://doi.org/10.1080/20008686.2020.1806505)

452.	El May E, Anker D, Epure A, Wagner C, Magnin J-L, Schmid A, et al. Corona Immunitas Fribourg - Immunité de la population Episode 3/2021. Corona Immunitas; 2021 Nov.

453.	Stringhini S, Zaballa M-E, Pullen N, Perez-Saez J, de Mestral C, Loizeau A, et al. Seroprevalence of anti-SARS-CoV-2 antibodies six months into the vaccination campaign in Geneva, Switzerland. medRxiv. 2021. doi:[10.1101/2021.08.12.21261929](https://doi.org/10.1101/2021.08.12.21261929)

454.	Giuliano E, Gennaro V, Milani GP, Bianchetti M, Kocher C, Buehrer T, et al. Seroprevalence of anti-SARS-CoV-2 IgG among adolescents at military fitness-for-duty evaluation. BMJ Military Health. 2021. doi:[10.1136/bmjmilitary-2021-001828](https://doi.org/10.1136/bmjmilitary-2021-001828)

455.	Emmenegger M, De Cecco E, Lamparter D, Jacquat RPB, Ebner D, Schneider MM, et al. Early peak and rapid decline of SARS-CoV-2 seroprevalence in a Swiss metropolitan region. medRxiv. 2020. doi:[10.1101/2020.05.31.20118554](https://doi.org/10.1101/2020.05.31.20118554)

456.	Ulyte A, Radtke T, Abela IA, Haile SR, Blankenberger J, Jung R, et al. Variation in SARS-CoV-2 seroprevalence across districts, schools and classes: Baseline measurements from a cohort of primary and secondary school children in Switzerland. BMJ Open. 2021;11: e047483. doi:[10.1136/bmjopen-2020-047483](https://doi.org/10.1136/bmjopen-2020-047483)

457.	Baggio S, Vernaz N, Spechbach H, Salamun J, Jacquerioz F, Stringhini S, et al. Vulnerable patients forgo health care during the first wave of the Covid-19 pandemic. Preventive Medicine. 2021;150: 106696. doi:[10.1016/j.ypmed.2021.106696](https://doi.org/10.1016/j.ypmed.2021.106696)

458.	Milani GP, Bianchetti MG, Togni G, Schoenenberger AW, Muggli F. SARS-CoV-2 Ig G among Healthcare Workers and the General Population. Pathogens. 2021;10: 465. doi:[10.3390/pathogens10040465](https://doi.org/10.3390/pathogens10040465)

459.	Major-Smith D, Matthews S, Breeze T, Crawford M, Woodward H, Wells N, et al. The Avon Longitudinal Study of Parents and Children - A resource for COVID-19 research: Antibody testing results, April June 2021. Wellcome Open Research. 2022;6: 283. doi:[10.12688/wellcomeopenres.17294.2](https://doi.org/10.12688/wellcomeopenres.17294.2)

460.	Beale S, Patel P, Rodger A, Braithwaite I, Byrne T, Fong WLE, et al. Occupation, work-related contact and SARS-CoV-2 anti-nucleocapsid serological status: Findings from the Virus Watch prospective cohort study. Occupational and Environmental Medicine. 2022; oemed-2021-107920. doi:[10.1136/oemed-2021-107920](https://doi.org/10.1136/oemed-2021-107920)

461.	Gaskell KM, Johnson M, Gould V, Hunt A, Stone NR, Waites W, et al. SARS-CoV-2 seroprevalence in a strictly-Orthodox Jewish community in the UK: A retrospective cohort study. The Lancet Regional Health - Europe. 2021;6: 100127. doi:[10.1016/j.lanepe.2021.100127](https://doi.org/10.1016/j.lanepe.2021.100127)

462.	Oakey J, Haslam S, Brown AP, Eglin J, Houghton B, Singleton D. EXPRESS: Comparison of four high throughput, automated immunoassays for the detection of SARS-CoV-2 antibodies. Annals of Clinical Biochemistry: International Journal of Laboratory Medicine. 2021; 000456322110157. doi:[10.1177/00045632211015711](https://doi.org/10.1177/00045632211015711)

463.	Abo-Leyah H, Gallant S, Cassidy D, Giam YH, Killick J, Marshall B, et al. The Protective Effect of SARS-CoV-2 Antibodies in Scottish Healthcare Workers. SSRN Electronic Journal. 2021. doi:[10.2139/ssrn.3770674](https://doi.org/10.2139/ssrn.3770674)

464.	Wells PM, Doores KM, Couvreur S, Martin Martinez R, Seow J, Graham C, et al. Estimates of the rate of infection and asymptomatic COVID-19 disease in a population sample from SE England. medRxiv. 2020; 2020.07.29.20162701. doi:[10.1101/2020.07.29.20162701](https://doi.org/10.1101/2020.07.29.20162701)

465.	Lumley SF, Eyre DW, McNaughton AL, Howarth A, Hoosdally S, Hatch SB, et al. SARS-CoV-2 antibody prevalence, titres and neutralising activity in an antenatal cohort, United Kingdom, 14 April to 15 June 2020. Eurosurveillance. 2020;25: 2001721. doi:[10.2807/1560-7917.ES.2020.25.41.2001721](https://doi.org/10.2807/1560-7917.ES.2020.25.41.2001721)

466.	Glennon K, Donnelly J, Knowles S, McAuliffe FM, O’Reilly A, Corcoran S, et al. Immunological assessment of SARS-CoV-2 infection in pregnancy from diagnosis to delivery: A multicentre prospective study. PLoS One. 2021;16: e0253090. doi:[10.1371/journal.pone.0253090](https://doi.org/10.1371/journal.pone.0253090)

467.	Amirthalingam G, Whitaker H, Brooks T, Brown K, Hoschler K, Linley E, et al. Seroprevalence of SARS-CoV-2 among Blood Donors and Changes after Introduction of Public Health and Social Measures, London, UK. Emerging Infectious Diseases. 2021;27: 1795–1801. doi:[10.3201/eid2707.203167](https://doi.org/10.3201/eid2707.203167)

468.	Bird PW, Sandhu K, Fletcher O, Ames B, Toovey O, Tang JW-T. Retrospective SARS-CoV-2 IgG screening during the first wave (March 2020) of the COVID-19 pandemic in the United Kingdom. Journal of Medical Virology. 2021;n/a. doi:[10.1002/jmv.27162](https://doi.org/10.1002/jmv.27162)

469.	Hughes EC, Amat JAR, Haney J, Parr YA, Logan N, Palmateer N, et al. Severe Acute Respiratory Syndrome Coronavirus 2 Serosurveillance in a Patient Population Reveals Differences in Virus Exposure and Antibody-Mediated Immunity According to Host Demography and Healthcare Setting. The Journal of Infectious Diseases. 2021;223: 971–980. doi:[10.1093/infdis/jiaa788](https://doi.org/10.1093/infdis/jiaa788)

470.	Bhuiyan TR, Hulse JD, Hegde ST, Akhtar M, Islam T, Khan ZH, et al. SARS-CoV-2 Seroprevalence before Delta Variant Surge, Chattogram, Bangladesh, March 2021. Emerging Infectious Diseases. 2022;28: 429–431. doi:[10.3201/eid2802.211689](https://doi.org/10.3201/eid2802.211689)

471.	Ahmed ZB, Razu MH, Akter F, Rabby MRI, Karmaker P, Khan M. Seropositivity of SARS-CoV-2 IgG Antibody among People in Dhaka City during the Prevaccination Period. BioMed Research International. 2022;2022: e4451144. doi:[10.1155/2022/4451144](https://doi.org/10.1155/2022/4451144)

472.	Shirin T, Flora MS, Salimuzzaman M, Alamgir A, Alam AN, Rahman M, et al. Population-based seroepidemiological investigation for coronavirus 2019 (COVID-19) infection in Cox’s Bazar Rohingya camps. Zenodo; 2021. doi:[10.5281/ZENODO.5646776](https://doi.org/10.5281/ZENODO.5646776)

473.	Dayanand D, Irudhayanathan I, Kundu D, Manesh A, Abraham V, Abhilash KP, et al. Community seroprevalence and risk factors for SARS-CoV-2 infection in different subpopulations in Vellore, India, and their implications for future prevention. International Journal of Infectious Diseases. 2022;116: 138–146. doi:[10.1016/j.ijid.2021.12.356](https://doi.org/10.1016/j.ijid.2021.12.356)

474.	Dash P, Mansingh A, Nayak SR, Sahoo D, Bhattacharya D, Kanungo S, et al. Infection, cases due to SARS-CoV-2 in rural areas during early COVID-19 vaccination: Findings from sero-survey study in a rural cohort of eastern India. Epidemiology and Infection. 2022; 1–27. doi:[10.1017/S0950268822000346](https://doi.org/10.1017/S0950268822000346)

475.	Hitchings MDT, Patel EU, Khan R, Srikrishnan AK, Anderson M, Kumar KS, et al. A mixture model to estimate SARS-CoV-2 seroprevalence in Chennai, India. Epidemiology; 2022 Feb. doi:[10.1101/2022.02.24.22271002](https://doi.org/10.1101/2022.02.24.22271002)

476.	Prakash O, Solanki B, Sheth JK, Nayak M, Kadam M, Vyas S, et al. COVID-Kavach-Based Seropositivity in the General Population of Ahmedabad: Just Before the Start of the Vaccination for the Elderly in India. Cureus. 2022. doi:[10.7759/cureus.22759](https://doi.org/10.7759/cureus.22759)

477.	Laxmaiah A, Rao NM, Arlappa N, Babu J, Kumar PU, Singh P, et al. SARS-CoV-2 seroprevalence in the city of Hyderabad, India in early 2021. medRxiv. 2021. doi:[10.1101/2021.07.18.21260555](https://doi.org/10.1101/2021.07.18.21260555)

478.	Kumar D, Burma A, Mandal AK, Joshy V. A Comparative Analysis of COVID-19 IgG Antibody Level and Socio-Demographic Status in Symptomatic and Symptomatic Population of South Andaman, India. Cureus. 2022;14: e22398. doi:[10.7759/cureus.22398](https://doi.org/10.7759/cureus.22398)

479.	Ramaswamy S, Athotra A, Kasar P, Tiwari R, Tomar SP, Kuraria R, et al. Cross-sectional Study on Sero-Prevalence of SARS-CoV-2 Infection in Jabalpur, Madhya Pradesh, India. Journal of Communicable Diseases. 2021;53: 82–88. doi:[10.24321/0019.5138.202114](https://doi.org/10.24321/0019.5138.202114)

480.	Mahapatra S. SARS COV-2- IgG antibodies in blood donors in pandemic a game changer for policy makers. Transfusion Clinique et Biologique. 2022;29: 11–15. doi:[10.1016/j.tracli.2021.10.004](https://doi.org/10.1016/j.tracli.2021.10.004)

481.	Goyal A, Gupta Y, Kalaivani M, Tandon N. Mild and Asymptomatic SARS-CoV-2 Infection is Not Associated with Progression of Thyroid Dysfunction or Thyroid Autoimmunity. Clinical Endocrinology. 2022;n/a. doi:[10.1111/cen.14731](https://doi.org/10.1111/cen.14731)

482.	Isaac R, Paul B, Finkel M, Moorthy M, Venkateswaran S, Bachmann T, et al. The Burden of COVID-19 Infection in a Rural Tamil Nadu Community. 2021. doi:[10.21203/rs.3.rs-289385/v1](https://doi.org/10.21203/rs.3.rs-289385/v1)

483.	Prakash O, Solanki B, Sheth J, Kharadi A, Kadam M, Vyas S, et al. Reassessing the population based Seroprevalence for IgG antibodies against SARS COV2 in Ahmedabad. Asian Journal of Medical Sciences. 2021;12: 1–6. doi:[10.3126/ajms.v12i5.35099](https://doi.org/10.3126/ajms.v12i5.35099)

484.	Ayub T, Raja MW, Khan SMS, Haq I, Qureshi MA, Majid S, et al. Seroprevalence of SARS CoV 2 specific Ig G antibodies in District Srinagar, Kashmir: A population based study. International Journal Of Community Medicine And Public Health. 2021;8: 1792–1797. doi:[10.18203/2394-6040.ijcmph20211236](https://doi.org/10.18203/2394-6040.ijcmph20211236)

485.	Banerjee A, Gaikwad B, Desale A, Jadhav SL, Rathod H, Srivastava K. Severe acute respiratory syndrome-coronavirus-2 seroprevalence study in Pimpri-Chinchwad, Maharashtra, India coinciding with falling trend - Do the results suggest imminent herd immunity? Indian Journal of Public Health. 2021 Jul-Sep;65: 256–260. doi:[10.4103/ijph.IJPH_122_21](https://doi.org/10.4103/ijph.IJPH_122_21)

486.	Kumar MS, Thangaraj JWV, Saravanakumar V, Selvaraju S, Kumar CPG, Sabarinathan R, et al. Monitoring the trend of SARS-CoV-2 seroprevalence in Chennai, India, July and October 2020. Transactions of The Royal Society of Tropical Medicine and Hygiene. 2021;115: 1350–1352. doi:[10.1093/trstmh/trab136](https://doi.org/10.1093/trstmh/trab136)

487.	Kshatri JS, Bhattacharya D, Praharaj I, Mansingh A, Parai D, Kanungo S, et al. Seroprevalence of SARS-CoV-2 in Bhubaneswar, India: Findings from 3 rounds of community surveys. Epidemiology & Infection. undefined/ed; 1–29. doi:[10.1017/S0950268821000972](https://doi.org/10.1017/S0950268821000972)

488.	Prakash Om, Solanki Bhavin, Sheth Jay, Oza Divyang, Kadam Mina, Vyas Sheetal, et al. Population-based seropositivity for IgG antibodies against SARS-CoV-2 in Ahmedabad city. Journal of Family Medicine and Primary Care. 2021;10: 2363–2368. doi:[10.4103/jfmpc.jfmpc_2062_20](https://doi.org/10.4103/jfmpc.jfmpc_2062_20)

489.	Kolthur-Seetharam U, Shah D, Shastri J, Juneja S, Kang G, Malani A, et al. SARS-CoV2 Serological Survey in Mumbai by NITI-BMC-TIFR - Preliminary Report of Round-2. NITI-BMC-TIFR; 2020 Aug.

490.	Sakalle S, Saroshe S, Shukla H, Mutha A, Vaze A, Arora A, et al. Seroprevalence of anti-SARS-CoV-2 antibodies in Indore, Madhya Pradesh: A community-based cross-sectional study, August 2020. Journal of family medicine and primary care. 2021;10: 1479–1484. doi:[10.4103/jfmpc.jfmpc_2015_20](https://doi.org/10.4103/jfmpc.jfmpc_2015_20)

491.	Kshatri JS, Bhattacharya D, Kanungo S, Giri S, Palo SK, Parai D, et al. Serological surveys to inform SARS-CoV-2 epidemic curve: A cross-sectional study from Odisha, India. Scientific Reports. 2021;11: 10551. doi:[10.1038/s41598-021-89877-y](https://doi.org/10.1038/s41598-021-89877-y)

492.	Saple P, Gosavi S, Pawar T, Chaudhari G, Mahale H, Deshmukh P, et al. Seroprevalence of anti-SARS-CoV-2 of IgG antibody by ELISA: Community-based, cross-sectional study from urban area of Malegaon, Maharashtra. Journal of Family Medicine and Primary Care. 2021;10: 1453. doi:[10.4103/jfmpc.jfmpc_2191_20](https://doi.org/10.4103/jfmpc.jfmpc_2191_20)

493.	Selvaraju S, Kumar MS, Thangaraj JWV, Bhatnagar T, Saravanakumar V, Kumar CPG, et al. Population-Based Serosurvey for Severe Acute Respiratory Syndrome Coronavirus 2 Transmission, Chennai, India. Emerging Infectious Diseases. 2021;27: 586–589. doi:[10.3201/eid2702.203938](https://doi.org/10.3201/eid2702.203938)

494.	Malani A, Shah D, Kang G, Lobo GN, Shastri J, Mohanan M, et al. Seroprevalence of SARS-CoV-2 in slums versus non-slums in Mumbai, India. The Lancet Global Health. 2021;9: e110–e111. doi:[10.1016/S2214-109X(20)30467-8](https://doi.org/10.1016/S2214-109X(20)30467-8)

495.	Prakash O, Solanki B, Sheth JK, Joshi B, Kadam M, Vyas S, et al. Assessing seropositivity for IgG antibodies against SARS-CoV-2 in Ahmedabad city of India: A cross-sectional study. BMJ Open. 2021;11: e044101. doi:[10.1136/bmjopen-2020-044101](https://doi.org/10.1136/bmjopen-2020-044101)

496.	Pandey HC, Dhiman Y, C S C, Coshic P, Jain P. Seroprevalence of SARS-Coronavirus 2 among asymptomatic healthy blood donors from healthcare and non-healthcare settings: Implications for safety of blood donors and blood collection staff during blood donation. Transfusion and Apheresis Science. 2021;60: 103118. doi:[10.1016/j.transci.2021.103118](https://doi.org/10.1016/j.transci.2021.103118)

497.	Chandrasingh S, George CE, Inbaraj LR. Are hospitals epicentres of COVID19 transmission? Findings of serial serosurveys among healthcare workers from a tertiary hospital in South India. Indian Journal of Medical Microbiology. 2021. doi:[10.1016/j.ijmmb.2021.10.006](https://doi.org/10.1016/j.ijmmb.2021.10.006)

498.	Billa V, Kothari J, Bichu S, Kumar R, Usulumarty D, Tilve P, et al. From infection to immunity - Impact of COVID-19 across nine hemodialysis centres in Mumbai. Indian Journal of Nephrology. 2021;31: 544. doi:[10.4103/ijn.IJN_361_20](https://doi.org/10.4103/ijn.IJN_361_20)

499.	Abdul-Raheem R, Moosa S, Waheed F, Aboobakuru M, Ahmed IN, Rafeeg FN, et al. A sero-epidemiological study after two waves of the COVID-19 epidemic. Asian Pacific Journal of Allergy and Immunology. 2021. doi:[10.12932/AP-040721-1177](https://doi.org/10.12932/AP-040721-1177)

500.	Machalek DA, Vette KM, Downes M, Carlin JB, Nicholson S, Hirani R, et al. Serological Testing of Blood Donors to Characterise the Impact of Covid-19 in Melbourne, Australia, 2020. medRxiv. 2022; 2022.03.11.22272185. doi:[10.1101/2022.03.11.22272185](https://doi.org/10.1101/2022.03.11.22272185)

501.	Machalek D, Vette K, Downes M, Carlin J, Hirani R, Irving D, et al. Serological testing of blood donors to characterise the impact of COVID-19 in Melbourne, 2020. Zenodo; 2021. doi:[10.5281/ZENODO.5644235](https://doi.org/10.5281/ZENODO.5644235)

502.	Gidding HF, Machalek DA, Hendry AJ, Quinn HE, Vette K, Beard FH, et al. Seroprevalence of SARS-CoV-2-specific antibodies in Sydney after the first epidemic wave of 2020. Medical Journal of Australia. 2021;214: 179–185. doi:[10.5694/mja2.50940](https://doi.org/10.5694/mja2.50940)

503.	Ho H-L, Wang F-Y, Lee H-R, Huang Y-L, Lai C-L, Jen W-C, et al. Seroprevalence of COVID-19 in Taiwan revealed by testing anti-SARS-CoV-2 serological antibodies on 14,765 hospital patients. The Lancet Regional Health - Western Pacific. 2020;3: 100041. doi:[10.1016/j.lanwpc.2020.100041](https://doi.org/10.1016/j.lanwpc.2020.100041)

504.	He L, Zeng Y, Zeng C, Zhou Y, Li Y, Xie X, et al. Positive Rate of Serology and RT-PCR for COVID-19 between Community Residents and Healthcare Workers in Wuhan, China. Japanese Journal of Infectious Diseases. 2020;advpub. doi:[10.7883/yoken.JJID.2020.691](https://doi.org/10.7883/yoken.JJID.2020.691)

505.	Zhang Z, Bi Q, Fang S, Wei L, Wang X, He J, et al. Insight into the practical performance of RT-PCR testing for SARS-CoV-2 using serological data: A cohort study. The Lancet Microbe. 2021;2: e79–e87. doi:[10.1016/S2666-5247(20)30200-7](https://doi.org/10.1016/S2666-5247(20)30200-7)

506.	Xu R, Huang J, Duan C, Liao Q, Shan Z, Wang M, et al. Low prevalence of antibodies against SARS-CoV-2 among voluntary blood donors in Guangzhou, China. Journal of Medical Virology. 2021;93: 1743–1747. doi:[10.1002/jmv.26445](https://doi.org/10.1002/jmv.26445)

507.	Zhang J, Liu J, Li N, Liu Y, Ye R, Qin X, et al. Serological detection of 2019-nCoV respond to the epidemic: A useful complement to nucleic acid testing. medRxiv. 2020; 2020.03.04.20030916. doi:[10.1101/2020.03.04.20030916](https://doi.org/10.1101/2020.03.04.20030916)

508.	Mori K, Imaki S, Ohyama Y, Satoh K, Abe T, Takeuchi I. Rapid screening for severe acute respiratory syndrome coronavirus 2 infection with a combined point-of-care antigen test and an immunoglobulin G antibody test. PLoS One. 2022年2月1日;17: e0263327. doi:[10.1371/journal.pone.0263327](https://doi.org/10.1371/journal.pone.0263327)

509.	Mizumoto K, Shimakawa Y, Aizawa Y, Butcher C, Chibana N, Collins M, et al. SARS-CoV-2 IgG seroprevalence in the Okinawa Main Island and remote islands in Okinawa, Japan, 2020-2021. medRxiv; 2022. p. 2022.03.02.22271759. doi:[10.1101/2022.03.02.22271759](https://doi.org/10.1101/2022.03.02.22271759)

510.	Morikane K, Satoh N, Hatano K, Kanouchi K, Kakehata S, Satoh S, et al. Seroepidemiology of SARS-CoV-2, Yamagata, Japan, June 2020. Western Pacific Surveillance and Response Journal : WPSAR. 2021;12: 69–71. doi:[10.5365/wpsar.2020.11.3.012](https://doi.org/10.5365/wpsar.2020.11.3.012)

511.	Lee K, Jo S, Lee J. Seroprevalence of SARS-CoV-2 antibodies in South Korea. Journal of the Korean Statistical Society. 2021. doi:[10.1007/s42952-021-00131-7](https://doi.org/10.1007/s42952-021-00131-7)

512.	Jeong HW, Chang H-H, Kim EJ, Kim YK, Kim S-M, Kim E-H, et al. Differences in seroprevalence between epicenter and non-epicenter areas of the COVID-19 outbreak in South Korea. Journal of Microbiology. 2021;59: 530–533. doi:[10.1007/s12275-021-1095-7](https://doi.org/10.1007/s12275-021-1095-7)

513.	Clapham HE, Chia WN, Tan LWL, Kumar V, Lim JM, Shankar N, et al. Contrasting SARS-CoV-2 epidemics in Singapore: Cohort studies in migrant workers and the general population. In Review; 2021 Apr. doi:[10.21203/rs.3.rs-441527/v1](https://doi.org/10.21203/rs.3.rs-441527/v1)

### S6 PROSPERO Protocol Registration

The following protocol is pulled from our most recent PROSPERO protocol registration (October 2021 version) CRD42020183634.

**Review question**

Our objective is to provide ongoing up-to-date synthesis of SARS-CoV-2 seroprevalence surveys from studies aligned with the WHO Unity sero-epidemiological protocol around the world.

**Searches**

We will search four electronic peer-review literature databases (MEDLINE, EMBASE, Web of Science, and Cochrane Database of Systematic Reviews) from January 1, 2020 until the end of the SARS-CoV-2 pandemic (according to WHO) using a comprehensive set of search terms developed in consultation with a health sciences librarian.

We will search grey literature in two ways. First, we will search for pre-print articles through Europe PMC electronic databases. When we encounter reports of potentially eligible government studies we will conduct a targeted search to locate and include the government study.

We will conduct two types of secondary searches. First, we will consult with international experts via e-mail to identify additional literature after all other sources have been searched. Second, we will invite submission of seroprevalence study results on our live dashboard SeroTracker.com. Third, we will coordinate with WHO regional offices to identify seroepidemiological investigators and studies that are receiving support from WHO including financial support, equipment (e.g., serological assays), or expertise.

**Types of study to be included**

We will include cross-sectional studies (single time point or repeated/panel design) and cohort studies. All other study designs will be excluded.

Studies will also be excluded if they have unclear sampling frames, sampling methods other than convenience or random sampling, and studies deliberately excluding cases of COVID-19. These criteria mirror the standards of the WHO sero-epidemiological Unity protocol.

**Condition or domain being studied**

Severe acute respiratory syndrome coronavirus 2.

**Participants/population**

Humans, any age. We will include studies whose participants fall into the category of “general population” as defined by the WHO Unity sero-epidemiological studies: Household and community samples, residual sera, outpatients, samples drawn from general population surveys or registries, outpatients, pregnant and parturient women.

**Intervention(s), exposure(s)**

We will include studies conducting serological testing – defined as the collection and testing of serum (or proxy such as oral fluid) specimens from a sample of a defined population over a specified period of time – to estimate the prevalence of antibodies against SARS-CoV-2 in the general population.

Studies will be excluded if they do not meet the serological assay performance threshold of 90% sensitivity and 97% specificity, as reported by the manufacturer (or author-evaluated in-house test, if manufacturer data is not present). An exception will be made for studies in a humanitarian country, as data from these settings may be sparse and their inclusion is essential for a global understanding of SARS-CoV-2 seroprevalence. These criteria mirror the standards for the WHO sero-epidemiological Unity protocol.

**Comparator(s)/control**

None

**Context**

All settings are eligible. Exceptions to the inclusion criteria will be made for studies in humanitarian settings to maximize inclusion of data from those resource limited settings.

**Main outcome(s)**

The outcome of interest was seroprevalence (the proportion of the population with detectable antibodies).

***Measures of effect***

Proportions

**Additional outcome(s)**

There are two additional outcomes of interest: the ratio of confirmed cases (according to nucleic acid testing) to seroprevalence (according to serology testing); and the prevalence ratios between subgroups, e.g. age, sex, and vaccination status.

***Measures of effect***

Ratio

Prevalence ratio

**Data extraction (selection and coding)**

References will be uploaded to Covidence, de-duplicated, and screened (first by title and abstract, then by reading full-text) by two independent researchers. Discrepancies in screening and extraction will be resolved by arbitration with a third author. We will contact study investigators in order to obtain complete data. The research team has members proficient in reading sources written in English, French, Spanish, Portuguese, Croatian, Serbian, Romanian. Other non-English language data sources will be read via Google Translate.

Included articles will be uploaded into a purpose-built Airtable database for data extraction. Data will be extracted on the characteristics of the study, sample (age, sex, vaccination subgroups), antibody test name, and the prevalence estimates. We will extract sub-group data based on the stratification of one variable (e.g., age) but not two variables (e.g., age and ethnicity). To contextualize prevalence estimates we will extract the number of total confirmed cases from the Johns Hopkins COVID-19 tracker (country level data) with population denominators from the UN Population Division estimates. We will extract the number of confirmed cases at the median date and end date for each prevalence study. We will also extract the number of total people vaccinated from the Our World in Data tracker (country level data) at the median date and end date for each prevalence study.

Data from unpublished studies will be included from investigators participating in the WHO Unity Studies collaboration. These data will be submitted using standardized templates, extracted directly into R, and analyzed as part of this study. Results from these unpublished studies will be housed temporarily on an open-access database to ensure reproducibility until authors have published their results, then the published version will replace any unpublished data in our database.

**Risk of bias (quality) assessment**

We will assess risk of bias for each study using a modified version of the Joanna Briggs Critical Appraisal Checklist for Prevalence Studies.

Two reviewers will provide independent assessments for two of the JBI items (item 1, item 5), while the other seven items will be automatically completed based on the other data elements extracted for the review . These nine items will be synthesized to generate a summary risk of bias assessment (low, moderate, high, unclear), determined using a decision rule (SeroTracker-ROB).

**Strategy for data synthesis**

The purpose of the analysis is to gather, synthesize, critically appraise, and present the best available estimates of seroprevalence in the general population to inform decision making in different regions and populations. We will summarize findings from all included studies. We will synthesize prevalence estimates with 95% confidence intervals for all WHO regions, time periods, and particular groups for which data is available (e.g., age, sex, vacacinated vs non-vaccinated individuals, symptomatic vs asymptomatic individuals). Where there are multiple estimates within each country and time period, we will aggregate these studies with random-effects meta-analysis where deemed appropriate.

We will compare seroprevalence estimates to the cumulative incidence of confirmed cases to understand the gap between these two figures. We will use meta-regression to identify the effects of study- and geographic-level factors on seroprevalence estimates, such as overall risk of bias, geographic scope, WHO region, income class, and severity of public health and social measures (PHSM).

**Analysis of subgroups or subsets**

We will conduct analyses overall and grouped by WHO geographical region. We will stratify the EURO and PAHO WHO regions by World Bank income level (high-income countries (HIC) vs. low-, lower-middle, and upper-middle-income countries (LMIC)).

We will analyze the seroprevalence ratio between different subpopulations within the same study. Variables of interest for this analysis include sex groups and age groups.

Sensitivity analyses will be conducted by stratifying studies based on risk of bias. To compare antibody kinetics and infection- and vaccination-induced seroprevalence in countries using S-based vaccines we will also stratify seroprevalence estimates for studies measuring Spike protein antibodies (anti-S) and studies measuring Nucleocapsid protein antibodies (anti-N)

**Contact details for further information**

Dr Isabel Bergeri

**Organisational affiliation of the review**

World Health Organization, https://www.who.int/

Epiconcept, https://www.epiconcept.fr/en/

University of Calgary, https://www.ucalgary.ca/

University of Toronto, https://www.utoronto.ca/

University of Oxford, https://www.ox.ac.uk/

McGill University, https://www.mcgill.ca/

**Review team members and their organisational affiliations**

Dr Niklas Bobrovitz. University of Toronto

Mr Rahul Arora. University of Oxford

Ms Emily Boucher. University of Calgary

Ms Tingting Yan. University of Toronto

Ms Hannah Rahim. University of Calgary

Mr Jordan Van Wyk. University of Waterloo

Mr Nathan Duarte. University of Waterloo

Mr Austin Atmaja. University of Waterloo

Ms Simona Rocco. University of Waterloo

Mr Abel Joseph. University of Waterloo

Mr Ewan May. University of Calgary

Mr Michael Liu. Harvard University

Ms Claire Donnici. University of Calgary

Mr Christian Cao. University of Calgary

Ms Natasha Illincic. University of Toronto

Mr Lucas Penny. University of Toronto

Mr Mitchell Segal. University of Toronto

Assistant/Associate Professor Tyler Williamson. University of Calgary

Professor Jim Kellner. University of Calgary

Assistant/Associate Professor Cedric Yansouni. McGill University

Assistant/Associate Professor Jesse Papenburg. McGill University

Professor Timothy Evans. McGill University

Assistant/Associate Professor Jonathan Chevrier. McGill University

Assistant/Associate Professor Matthew Cheng. McGill University

Ms Mairead Whelan. University of Calgary

Miss Harriet Ware. University of Calgary

Miss Xiaomeng Ma. University of Toronto

Mr Zihan Li. University of Waterloo

Dr Isabel Bergeri. World Health Organization

Miss Brianna Cheng. World Health Organization

Mrs Hannah Lewis. World Health Organization

Dr Anthony Nardone. Epiconcept

Dr Marta Valenciano. Epiconcept

Dr Lorenzo Subissi. World Health Organization

**Collaborators**

Miss Lubna Al Ariqi. World Health Organization, Regional Office for the Eastern Mediterranean

Dr Andrea Vicari. Pan American Health Organization

Dr Joseph Okeibunor. World Health Organization, Regional Office for Africa

Dr Tasnim Azim. World Health Organization, Regional Office for South-East Asia

Dr Pushpa Wijesinghe. World Health Organization, Regional Office for South-East Asia

Eman Abdelkreem. World Health Organization, Regional Office for the Eastern Mediterranean

Dr Aisling Vaughan. World Health Organization, Regional Office for Europe

Dr Richard Pebody. World Health Organization, Regional Office for Europe

Dr Linh-Vi Ve. World Health Organization, Regional Office for the Western Pacific

**Type and method of review**

Epidemiologic, Meta-analysis, Systematic review

**Anticipated or actual start date**

16 April 2020

**Anticipated completion date**

30 June 2022

**Funding sources/sponsors**

Public Health Agency of Canada 2021-HQ-000056 - operational funding for review and for SeroTracker.com, plus other non-review activities.

WHO Unity Studies, with funding to World Health Organization by the COVID-19 Solidarity Response Fund and the German Federal Ministry of Health (BMG) COVID-19 Research and development (Ref:2021/1114309-0).

***Grant number(s)***

***State the funder, grant or award number and the date of award***

Public Health Agency of Canada 2021-HQ-000056

World Health Organization Ref:2021/1114309-0

**Conflicts of interest**

**Language**

English

**Country**

Canada, Congo, Denmark, Egypt, England, India, Namibia, Philippines, Switzerland, United States of America

**Stage of review**

Review Completed published being updated

**Details of final report/publication(s) or preprints if available**

Niklas Bobrovitz, Rahul Krishan Arora, Christian Cao, Emily Boucher, Michael Liu, Claire Donnici, Mercedes Yanes-Lane, Mairead Whelan, Sara Perlman-Arrow, Judy Chen, Hannah Rahim, Natasha Ilincic, Mitchell Segal, Nathan Duarte, Jordan Van Wyk, Tingting Yan, Austin Atmaja, Simona Rocco, Abel Joseph, Lucas Penny, David A. Clifton, Tyler Williamson, Cedric P. Yansouni, Timothy Grant Evans, Jonathan Chevrier, Jesse Papenburg, Matthew P. Cheng. Global seroprevalence of SARS-CoV-2 antibodies: A systematic review and meta-analysis. PLOS One. Published: June 23, 2021. https://doi.org/10.1371/journal.pone.0252617

<https://journals.plos.org/plosone/article?id=10.1371/journal.pone.0252617>

**Subject index terms status**

Subject indexing assigned by CRD

**Subject index terms**

COVID-19; Humans; SARS Virus; Seroepidemiologic Studies; severe acute respiratory syndrome coronavirus 2

**Date of registration in PROSPERO**

07 May 2020

**Date of first submission**

02 May 2020

**Details of any existing review of the same topic by the same authors**

**Stage of review at time of this submission**

| **Stage** | **Started** | **Completed** |
| --- | --- | --- |
| Preliminary searches | Yes | Yes |
| Piloting of the study selection process | Yes | Yes |
| Formal screening of search results against eligibility criteria | Yes | No |
| Data extraction | No | No |
| Risk of bias (quality) assessment | No | No |
| Data analysis | No | No |

**Revision note**

Our previous review focused on published studies and used primarily descriptive analysis. This updated protocol includes the following changes to expand the robustness of our analysis and data sources: - Focuses on studies whose design aligns with the WHO Unity protocols, thereby updating inclusion criteria - Uses an expanded search strategy to incorporate ongoing, unpublished data accessible through WHO networks and stakeholders which will be made available in an open-access platform to assure reproducibility - New analysis of seroprevalence data including global meta-analysis and seroprevalence over time - New approach to automated critical appraisal and risk of bias assessment using a validated tool - Addition of team members and collaborators with expertise in global health, infectious diseases epidemiology, and biostatistics

*The record owner confirms that the information they have supplied for this submission is accurate and complete and they understand that deliberate provision of inaccurate information or omission of data may be construed as scientific misconduct.*

*The record owner confirms that they will update the status of the review when it is completed and will add publication details in due course.*

### S7 References cited in supplementary file

1.	Page MJ, McKenzie JE, Bossuyt PM, et al. The PRISMA 2020 statement: an updated guideline for reporting systematic reviews. *BMJ*. Published online March 29, 2021:n71. doi:10.1136/bmj.n71

2.	Bobrovitz N, Arora RK, Cao C, et al. Global seroprevalence of SARS-CoV-2 antibodies: A systematic review and meta-analysis. Khudyakov YE, ed. *PLOS ONE*. 2021;16(6):e0252617. doi:10.1371/journal.pone.0252617

3.	Arora RK, Joseph A, Van Wyk J, et al. SeroTracker: a global SARS-CoV-2 seroprevalence dashboard. *Lancet Infect Dis*. 2021;21(4):e75-e76. doi:10.1016/S1473-3099(20)30631-9

4.	Yazdani S, Asli A, Asli A, Sarbakhsh P, Salemi S. Final Sensitivity and Specificity of Different Test Combinations in Screening. *JMED Res*. Published online April 23, 2014:1-10. doi:10.5171/2014.558523

5.	NRL. WHO COVID Evaluations Summary of Results. nrlquality. Published November 22, 2021. Accessed November 22, 2021. https://www.nrlquality.org.au/who-covid-evaluations-summary-of-results

6.	Aus AGD of HTG. Post market review of COVID-19 point-of-care tests. Therapeutic Goods Administration (TGA). Published March 24, 2021. Accessed November 22, 2021. https://www.tga.gov.au/post-market-review-covid-19-point-care-tests

7.	Beld MJC van den, Murk JL, Kluytmans J, Koopmans MPG, Reimerink J, Loo IHM van, et al. Increasing the Efficiency of a National Laboratory Response to COVID-19: a Nationwide Multicenter Evaluation of 47 Commercial SARS-CoV-2 Immunoassays by 41 Laboratories. *J Clin Microbiol*. 2021;59(9):e00767-21.

8.	Bewley KR, Coombes NS, Gagnon L, et al. Quantification of SARS-CoV-2 neutralizing antibody by wild-type plaque reduction neutralization, microneutralization and pseudotyped virus neutralization assays. *Nat Protoc*. 2021;16(6):3114-3140. doi:10.1038/s41596-021-00536-y

9.	Mohit E, Rostami Z, Vahidi H. A comparative review of immunoassays for COVID-19 detection. *Expert Rev Clin Immunol*. 2021;17(6):573-599. doi:10.1080/1744666X.2021.1908886

10.	WHO Unity Global SARS-CoV-2 Seroepidemiological Investigations. Zenodo Community. Published October 26, 2021. Accessed November 5, 2021. https://zenodo.org/communities/unity-sero-2021?page=1&size=20

11.	Ware H, Whelan M, Li Z, et al. WHO Unity Sero-Epidemiological Studies Early Results Submission Template - Blank. Published online November 23, 2021. doi:10.5281/ZENODO.5719533

12.	World Health Organization. Guidelines on the Use of Serosurveys in Support of Measles and Rubella Elimination. Published 2013. Accessed November 21, 2021. https://www.who.int/immunization/monitoring_surveillance/burden/laboratory/Serosurvey_Manual_chapter_1.pdf

13.	Clapham H, Hay J, Routledge I, et al. Seroepidemiologic Study Designs for Determining SARS-COV-2 Transmission and Immunity. *Emerg Infect Dis J*. 2020;26(9):1978. doi:10.3201/eid2609.201840

14.	Bergeri I, Lewis HC, Subissi L, et al. Early epidemiological investigations: World Health Organization UNITY protocols provide a standardized and timely international investigation framework during the COVID-19 pandemic. *Influenza Other Respir Viruses*. 2021;n/a(n/a). doi:10.1111/irv.12915

16.	Bobrovitz N, Noel KC, Li Z, et al. *SeroTracker-ROB: Reproducible Decision Rules for Risk of Bias Assessment of Seroprevalence Studies*. Epidemiology; 2021. doi:10.1101/2021.11.17.21266471

17.	Migliavaca CB, Stein C, Colpani V, Munn Z, Falavigna M. Quality assessment of prevalence studies: a systematic review. *J Clin Epidemiol*. 2020;127:59-68. doi:10.1016/j.jclinepi.2020.06.039

18.	Borges Migliavaca C, Stein C, Colpani V, et al. How are systematic reviews of prevalence conducted? A methodological study. *BMC Med Res Methodol*. 2020;20(1):96. doi:10.1186/s12874-020-00975-3

19.	Ritchie H, Mathieu E, Rodés-Guirao L, et al. Coronavirus Pandemic (COVID-19). Our World in Data. Published 2020. Accessed November 4, 2021. https://ourworldindata.org/coronavirus

20.	World Health Organization. WHO Coronavirus Disease (COVID-19) Dashboard. Published June 11, 2021. Accessed June 11, 2021. https://covid19.who.int/

21.	WHO Headquarters (HQ). WHO COVID-19 Case definition. Published online December 16, 2020. Accessed November 5, 2021. https://www.who.int/publications-detail-redirect/WHO-2019-nCoV-Surveillance_Case_Definition-2020.2

22.	Institute for Health metrics and Evaluation (IHME). *COVID-19: Estimating the Historical Time Series of Infections*. IHME, University of Washington; 2021. http://www.healthdata.org/special-analysis/covid-19-estimating-historical-infections-time-series

23.	Bates D, Mächler M, Bolker B, Walker S. Fitting Linear Mixed-Effects Models Using **lme4**. *J Stat Softw*. 2015;67(1). doi:10.18637/jss.v067.i01

24.	Wiersinga WJ, Rhodes A, Cheng AC, Peacock SJ, Prescott HC. Pathophysiology, Transmission, Diagnosis, and Treatment of Coronavirus Disease 2019 (COVID-19): A Review. *JAMA*. 2020 Aug 25;324(8):782–93.

25.	Wilson N, Kvalsvig A, Barnard LT, Baker M. Case-Fatality Risk Estimates for COVID-19 Calculated by Using a Lag Time for Fatality. *Emerging Infectious Disease journal*. 2020;26(6):1339.
